# Deep learning cardiac motion analysis reveals the dynamic pathophysiology and genetic architecture of heart failure with preserved ejection fraction

**DOI:** 10.64898/2026.09.28.26364208

**Authors:** Kirsten R. Steffner, Nicolas Quach, Shriya G. Reddy, Roger Xia, Larissa Kiwakyou, Bruna Gomes, Euan A. Ashley

## Abstract

**Background:** Heart failure with preserved ejection fraction (HFpEF) accounts for approximately half of the more than 64 million heart failure cases globally, yet its pathophysiology is incompletely understood, its molecular determinants are poorly defined, and disease-specific therapies remain limited. Comprehensive characterization of the cardiac motion abnormalities central to HFpEF has not been feasible at the population level.

**Methods:** We developed a deep learning framework integrating image segmentation with optical flow motion analysis and applied it to standard cine cardiac magnetic resonance images from 83,569 UK Biobank participants, deriving 32 myocardial and inner cavity velocity phenotypes spanning the cardiac cycle, including mid-diastolic velocities not previously quantified at population scale. We examined the prognostic relevance, genetic architecture, and candidate causal mediators of the optical flow velocity phenotypes.

**Results:** In a pragmatically defined HFpEF subcohort, mid-diastolic and late-diastolic optical flow velocities were reduced relative to healthy reference participants, while higher systolic left ventricular myocardial velocity was associated with lower all-cause mortality (HR 0.61 per SD; 95% CI 0.45-0.82). Genome-wide association analyses identified 12 risk loci associated with optical flow velocity phenotypes. Phospholamban (*PLN*), the canonical regulator of sarcoplasmic reticulum calcium reuptake, demonstrated the broadest pleiotropy across all cardiac phases. *SOX5*, a transcription factor involved in extracellular matrix development, showed significant genome-wide association exclusively with mid-diastolic velocities. Mendelian randomization implicated RABGAP1L as a candidate causal mediator of early diastolic velocity (IVW β = −0.255 per NPX for early diastolic right ventricular inner circumferential velocity; p = 5.49×10⁻²²), nominating a calcium-handling pathway in diastolic dysfunction.

**Conclusion:** Genetic and causal-inference evidence implicates intracellular calcium handling and extracellular matrix remodeling as candidate mechanisms underlying diastolic dysfunction. Together, this work establishes deep-learning-enabled cardiac motion phenotyping as a scalable approach to interrogate the molecular basis of HFpEF and nominates candidate targets for therapeutic development.

**CLINICAL PERSPECTIVE:** *What Is New?:* ● Population-scale optical flow motion analysis of 83,569 cardiac magnetic resonance images from the UK Biobank allows for the extraction of 32 biventricular myocardial and inner cavity velocity phenotypes, including mid-diastolic (L-wave) velocities. ● In HFpEF participants, mid-diastolic and late-diastolic optical flow velocities are reduced relative to healthy reference participants, while higher systolic left ventricular myocardial velocity is associated with lower all-cause mortality. ● Genetic (*PLN*, *SOX5*) and causal-inference (*RABGAP1L*) signals independently nominate intracellular calcium handling and extracellular matrix remodeling as candidate mechanistic pathways underlying diastolic motion abnormalities.

*What Are the Clinical Implications?:* ● Optical flow velocity phenotypes require prospective external validation and establishment of population-based reference ranges before clinical application. ● Optical flow velocity phenotypes offer candidate objective metrics for HFpEF identification and risk-stratification for a syndrome that is inconsistently defined in current clinical practice. ● Convergent genomic and causal-inference evidence implicates intracellular calcium handling and extracellular matrix remodeling as possible mechanistic axes for therapeutic development for the diastolic motion abnormalities central to HFpEF.

## INTRODUCTION

Heart failure with preserved ejection fraction (HFpEF) accounts for approximately half of the 64 million heart failure cases across the globe^1,2^, and this proportion will continue to grow with the aging population and the rising burden of cardiometabolic comorbidities^2,3^. Diastolic dysfunction is a key contributor to the abnormal hemodynamics of HFpEF. However, the underlying pathophysiology of HFpEF remains incompletely understood, and the genetic architecture and molecular drivers of the cardiac motion abnormalities that characterize HFpEF are poorly defined. While models of metabolic dysregulation and systemic inflammation have been proposed^2^, HFpEF remains a heterogeneous syndrome without a single unifying biomarker or imaging feature^2,3^, making diagnosis, risk stratification, and disease-specific drug target identification challenging.

Comprehensive characterization of cardiac motion in HFpEF has been hindered, in part, by the constraints of conventional metrics for assessing diastolic dysfunction. The quantitative assessment of diastolic function relies on echocardiography-based parameters such as mitral annular diastolic velocity by pulsed wave tissue Doppler (e’) that carry inherent shortcomings, such as Doppler-angle-dependence and confounding by conditions that alter annular motion independent of diastolic function^4^. Moreover, poor concordance among echocardiographic measures creates significant variability in the definition of diastolic dysfunction and affects its reported prevalence^5,6^. Together, the heterogeneity of clinically defined HFpEF and the limitations of echocardiographic diastolic function assessment have precluded systematic phenotyping of the cardiac motion abnormalities relevant to HFpEF at population scale.

We present a deep learning framework for cardiac motion phenotyping that integrates image segmentation with optical flow motion analysis. Optical flow is a computer vision strategy that quantifies motion across multi-frame images, yielding pixel velocities across the entire image^7–9^. We apply the pipeline to standard cine CMR images from 83,569 UK Biobank participants, extracting 32 optical flow velocity phenotypes, enabling characterization and multi-omic analyses of biventricular myocardial and inner cavity motion, including mid-diastolic motion not previously quantified in large cohort studies. In a pragmatically defined HFpEF subcohort, we evaluate their prognostic relevance. We then perform GWAS, proteome-wide association studies, and Mendelian randomization to identify heritable pathways and prioritize circulating proteins as candidate molecular mediators of cardiac motion.

## METHODS

Detailed methods are provided in Supplemental Detailed Methods and are summarized here.

### Study Population and Cardiac Magnetic Resonance Imaging Data

The United Kingdom (UK) Biobank is a prospective study of more than 500,000 individuals between the ages of 40 and 69 years enrolled between 2006-2010 from across the United Kingdom, with comprehensive clinical phenotyping, imaging, biomarker, and genetic data.^10^ We restricted our analyses to standard cine cardiac magnetic resonance images (CMRs) from the first imaging visit, identifying 83,569 UK Biobank participants with available four-chamber long-axis standard cine CMR data.

### Automated Optical Flow Cardiac Motion Phenotyping at Population Scale

We developed an automated pipeline that integrates deep learning image segmentation with optical flow motion tracking and applied it to the 83,569 four-chamber long-axis standard cine CMR images from the UK Biobank (**Figure 1**). We trained an nnU-Net deep learning segmentation model^11^ to identify four key cardiac structures from four-chamber long-axis standard cine CMR sequences: the left ventricular (LV) inner cavity, the LV myocardium, the right ventricular (RV) inner cavity, and the RV myocardial free wall. Model performance was evaluated by five-fold cross-validation. Model weights from the best-performing fold were applied to the standard cine CMR images. Application of the Dual TV-L1^7,8^ optical flow algorithm to the standard cine CMR images generated pixel-level velocity vector fields between consecutive imaging frames, which were then filtered by the segmentation masks to isolate structure-specific velocity vectors (**Supplemental Figure 1).** For each of the four key cardiac structures, we plotted frame-to-frame median velocity magnitudes as continuous waveforms across the cardiac cycle. LV and RV inner cavity velocity vectors were additionally decomposed into radial and circumferential components. To extract quantitative phenotypes, we developed an automated peak detection algorithm that identified four motion events per velocity waveform, including: the systolic (S-wave) peak, the early diastolic (E-wave) peak, the mid-diastolic (L-wave) peak, and the late diastolic (A-wave) peak (**Figure 1C-D; Supplemental Data 1**). This pipeline yielded 32 distinct optical flow velocity phenotypes per participant. Ten conventional cardiac parameters were also extracted from the UK Biobank cardiac imaging database for comparison against optical flow velocity phenotypes (**Supplemental Table 1**).

**Figure 1.**
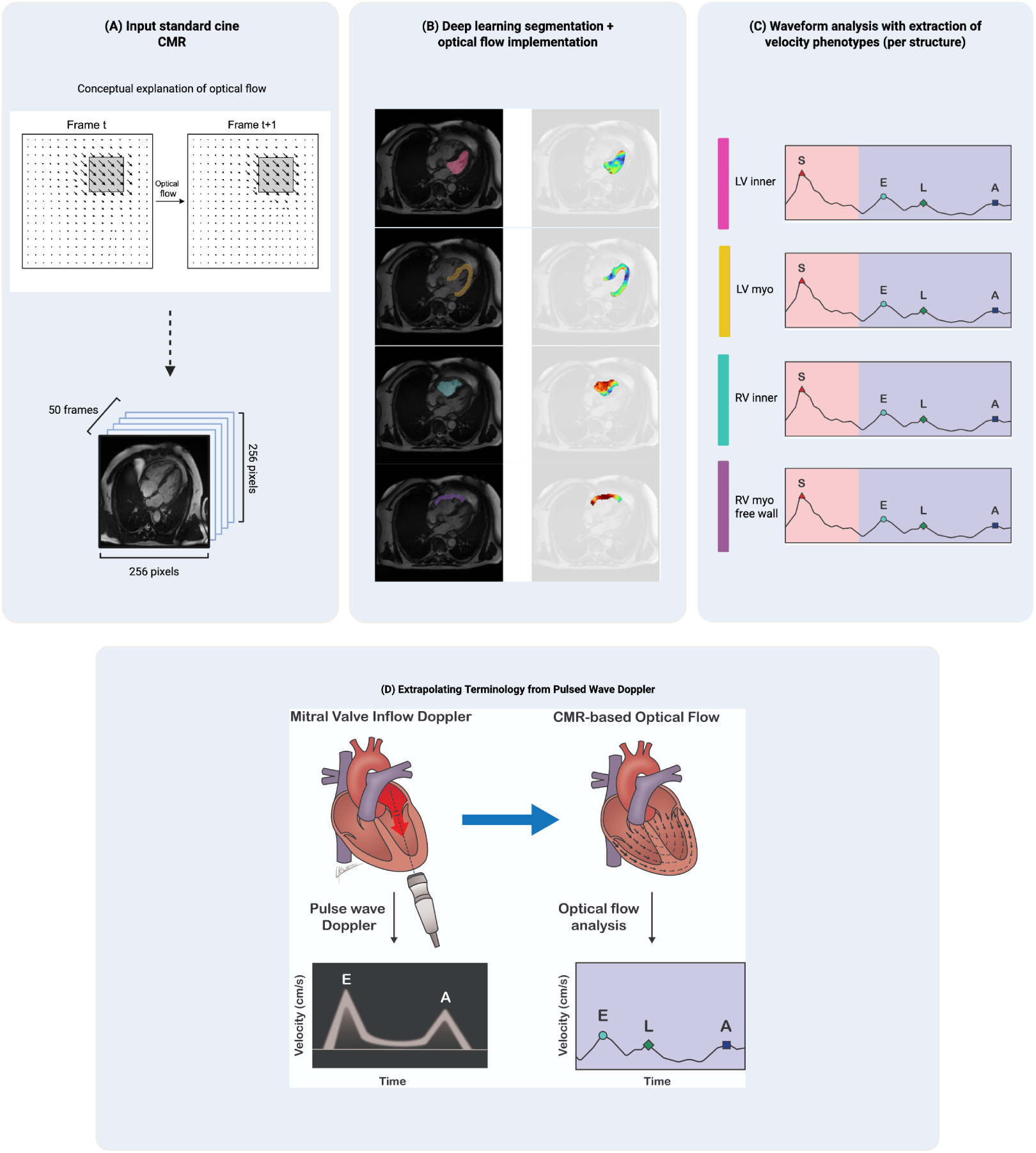
Automated pipeline for the extraction of cardiac motion phenotypes from standard cine CMR using optical flow. (A) Optical flow estimates a motion vector at every pixel between consecutive frames, illustrated conceptually for a two-frame sequence. Four-chamber long-axis standard cine CMR images (50 frames, 256×256 pixels) from 83,569 UK Biobank participants served as input. (B) Deep learning segmentation identified four cardiac structures per frame: LV inner cavity (pink), LV myocardium (yellow), RV inner cavity (teal), and RV myocardial free wall (purple). Optical flow was implemented to generate velocity vector fields (right column, color-coded by magnitude), anatomically filtered using the segmentation masks. (C) Frame-by-frame median velocities were plotted across the cardiac cycle to yield continuous waveforms per structure, from which four velocity peaks were identified: systolic S-wave (red triangle), early diastolic E-wave (cyan circle), mid-diastolic L-wave (green diamond), and late diastolic A-wave (navy square). Pink shading = systole; blue shading = diastole. Waveforms shown are schematic representations for illustrative purposes only. (D) The E-, L-, and A-wave nomenclature is adopted from echocardiography-based pulsed wave Doppler of mitral valve inflow (left), which captures two diastolic velocity peaks, the E-and A-waves; the mid-diastolic L-wave is not routinely visualized. CMR-based optical flow waveforms (right) identify these same diastolic phases and consistently capture the L-wave as a distinct motion event. Diastolic phases only are shown for clarity, as the echocardiographic S-wave is derived from tissue Doppler of the mitral annulus rather than pulsed wave Doppler of mitral inflow. CMR = cardiac magnetic resonance; LV = left ventricular; myo = myocardial; RV = right ventricular; UK = United Kingdom.

### HFpEF and Healthy Reference Subcohort Definition

Among the 83,569 participants with successful optical flow CMR motion analysis, we pragmatically defined a HFpEF subcohort and a healthy reference subcohort. The HFpEF subcohort was defined as the individuals with an LVEF > 50% and an NT-proBNP > the sex-specific 85th percentile. This yielded 690 HFpEF participants. The healthy reference subcohort was defined as the individuals with an LVEF > 50% and an NT-proBNP < the sex-specific 50th percentile. This yielded 2,483 healthy reference participants.

### Statistical Analysis

Baseline cohort and subcohort characteristics were summarized using median and interquartile range (IQR) for continuous variables and frequencies with percentages for categorical variables. HFpEF and healthy reference subcohorts were compared using Mann-Whitney U tests and chi-square or Fisher exact tests. Optical flow phenotypes were compared between subcohorts using Mann-Whitney U tests on age-and sex-adjusted residuals, with cluster-adjusted Benjamini-Hochberg false discovery rate (FDR) correction. Phenotypes reaching FDR significance were confirmed by analysis of covariance (ANCOVA) sensitivity analysis on raw values, adjusting for age and sex. Pearson correlations were computed between all 32 optical flow phenotypes and 10 conventional cardiac parameters, with cluster-adjusted Bonferroni correction.

### Survival Analyses

We assessed all-cause mortality using Cox proportional hazards models across 42 candidate parameters (32 optical flow, 10 conventional). Cluster-adjusted FDR-significant univariate predictors were advanced, after correlation pruning and events-per-variable capping, to multivariable models (Conventional-only, Optical-flow-only, and Combined), which were compared by concordance index, likelihood-ratio tests, and bootstrap resampling. Kaplan-Meier survival curves with log-rank tests compared the highest and lowest quartiles of the strongest univariate predictors.

### Proteomic and Genomic Analyses

We performed proteome-wide association analyses, regressing 1,463 Olink Explore proteins against age-and sex-adjusted residuals of the 32 optical flow phenotypes and of the 10 conventional cardiac parameters, with cluster-adjusted Bonferroni correction. Proteins uniquely associated with optical flow phenotypes were submitted to STRING-DB for protein–protein interaction network analysis.

Single nucleotide polymorphism (SNP)-based heritability for the 32 optical flow phenotypes was estimated using linkage disequilibrium score regression (LDSC) with 1000 Genomes Phase 3 European LD scores^12^, with standard Bonferroni correction. Pairwise genetic correlations were computed for all 496 phenotype pairs using bivariate LDSC with the same reference panel.

Genome-Wide Association Studies (GWAS) were performed for the 32 optical flow phenotypes using PLINK2 in up to 73,978 UK Biobank participants of European ancestry after standard variant-and sample-level quality control, with a hierarchical clustering-based Bonferroni correction. Independent loci were defined by merging genome-wide significant SNPs within 1 Mb of one another into a single locus, with the lead SNP defined as the most significant variant within each locus. Gene mapping and functional annotation were performed using FUMA on the full-cohort GWAS.

### Mendelian Randomization and Causal Inference

We performed two-sample Mendelian randomization (MR) using cis-protein quantitative trait loci (cis-pQTLs) from the UK Biobank Pharma Proteomics Project as genetic instruments using the inverse-variance weighted (IVW) method, with standard Bonferroni correction. To satisfy the independence assumption of two-sample MR, we excluded 9,480 participants who contributed to both the CMR dataset and the proteomics dataset. Re-running the outcome GWAS in the non-overlapping participants yielded 65,162–65,496 participants per phenotype, which were used for MR and colocalization analyses. Horizontal pleiotropy was assessed using MR-PRESSO.

For protein–phenotype pairs passing MR-PRESSO, colocalization between the cis-pQTL signal and the optical flow GWAS signal was assessed using coloc (PP.H4 > 0.8). Expression QTL colocalization was additionally performed for each MR protein hit against GTEx v10 summary statistics in four bulk cardiac-relevant tissues. MR-PRESSO-validated proteins were submitted to STRING-DB for protein–protein interaction network analysis.

## RESULTS

### Optical Flow for Comprehensive Cardiac Motion Phenotyping

Model performance of the nnU-Net deep learning segmentation model was evaluated by five-fold cross-validation, achieving Dice similarity coefficients of 0.949 ± 0.008 (LV inner cavity), 0.906 ± 0.003 (LV myocardium), 0.923 ± 0.007 (RV inner cavity), and 0.830 ± 0.008 (RV myocardial free wall). Model weights from the best-performing fold were applied to the 83,569 baseline four-chamber long-axis standard cine CMR images. Among the 83,569 participants for whom optical flow velocities were successfully calculated, 83,127 (99.47%) had complete data for all 32 optical flow velocity metrics (**Supplemental Table 2; Supplemental Figure 2**).

### Optical Flow Velocity Phenotypes Distinguish HFpEF from Healthy Reference Participants

Baseline study cohort characteristics are summarized in **Table 1A** and **Supplemental Data 2**. The differences in conventional cardiac parameters between the HFpEF subcohort and the healthy reference subcohort were clinically negligible, even where statistically significant (**Table 1A**). When comparing age-and sex-adjusted optical flow residual values between the HFpEF and healthy reference subcohorts, 10 parameters showed statistically significant differences (**Table 1B**), each confirmed by ANCOVA sensitivity analysis. All 10 optical flow parameters were lower in HFpEF, and all significant findings involved mid-diastolic (L-wave) or late diastolic (A-wave) velocity phenotypes. Late diastolic (A-wave) LV inner total velocity showed the most significant difference (p = 1.67×10⁻¹⁶, r = 0.147; **Table 1B**).

**Table 1.** (A) **Baseline characteristics of the full cohort, HFpEF subcohort, and healthy reference subcohort.** Baseline demographic, clinical, and conventional cardiac parameters are shown for the full cohort (n = 83,569), the HFpEF subcohort (n = 690), and the healthy reference subcohort (n = 2,483). Values are reported as median [IQR] for continuous variables and counts with percentages for categorical variables. For conventional cardiac parameters, denominators reflect participants with valid measurements and are shown in parentheses when less than the full cohort size. P-values are from Mann-Whitney U tests for continuous variables and chi-square or Fisher’s exact tests for categorical variables, comparing the HFpEF and healthy reference subcohorts; full cohort values are shown for descriptive context. Effect sizes (r) are reported for significant differences. HFpEF = heart failure with preserved ejection fraction; IQR = interquartile range; LV = left ventricular; MRI = magnetic resonance imaging; NSTEMI = non-ST elevation myocardial infarction; RV = right ventricular; STEMI = ST elevation myocardial infarction.

| Characteristic | Full cohort<br>(n=83,569) | HFpEF<br>(n=690) | Healthy<br>(n=2,483) | p-value |
| --- | --- | --- | --- | --- |
| **DEMOGRAPHICS** |  |  |  |  |
| Age years | 67.0<br>[60.0-72.0] | 68.0<br>[61.0-72.0] | 61.0<br>[55.0-67.0] | <0.001<br>(r=0.277) |
| Female sex n (%) | 43430 (52.0%) | 369 (53.5%) | 1336 (53.8%) | 0.913 |
| Median follow-up years | 4.7 [1.3-6.3]<br>(n=83526) | 6.3 [5.2-7.8] | 6.0 [5.3-7.5]<br>(n=2482) | 0.197 |
| **CLINICAL EVENTS** |  |  |  |  |
| All cause dementia n (%) | 210 (0.3%) | 6 (0.9%) | 11 (0.4%) | 0.288 |
| Alzheimer's disease n (%) | 83 (0.1%) | 2 (0.3%) | 4 (0.2%) | 0.617 |
| End stage renal disease n (%) | 65 (0.1%) | 0 (0.0%) | 1 (0.0%) | 1.000 |
| Frontotemporal dementia n (%) | 7 (0.0%) | 0 (0.0%) | 1 (0.0%) | 1.000 |
| Intracerebral haemorrhage n (%) | 143 (0.2%) | 0 (0.0%) | 5 (0.2%) | 0.592 |
| Ischaemic stroke n (%) | 856 (1.0%) | 16 (2.3%) | 24 (1.0%) | 0.009 |
| Myocardial infarction n (%) | 2688 (3.2%) | 50 (7.2%) | 47 (1.9%) | <0.001 |
| NSTEMI n (%) | 1192 (1.4%) | 27 (3.9%) | 20 (0.8%) | <0.001 |
| STEMI n (%) | 853 (1.0%) | 14 (2.0%) | 19 (0.8%) | 0.007 |
| Stroke n (%) | 1413 (1.7%) | 22 (3.2%) | 39 (1.6%) | 0.010 |
| Subarachnoid haemorrhage n (%) | 142 (0.2%) | 0 (0.0%) | 7 (0.3%) | 0.358 |
| Vascular dementia n (%) | 40 (0.0%) | 0 (0.0%) | 3 (0.1%) | 1.000 |
| **CONVENTIONAL CARDIAC PARAMETERS** |  |  |  |  |
| Left ventricular cardiac output (BSA-indexed) L/min/m<sup>2</sup> | 2.8 [2.5-3.2]<br>(n=38439) | 2.8 [2.5-3.2]<br>(n=669) | 2.9 [2.5-3.2]<br>(n=2410) | 0.065 |
| Left ventricular ejection fraction % | 59.7<br>[55.9-63.7]<br>(n=39120) | 60.2<br>[56.8-64.1] | 59.4<br>[56.1-63.3] | 0.001<br>(r=0.057) |
| Left ventricular end-diastolic volume (BSA-indexed) mL/m<sup>2</sup> | 77.7<br>[69.5-87.1]<br>(n=38439) | 78.6<br>[71.4-88.1]<br>(n=669) | 77.2<br>[68.9-86.2]<br>(n=2410) | <0.001<br>(r=0.062) |
| Left ventricular end-systolic volume (BSA-indexed) mL/m<sup>2</sup> | 31.0<br>[26.4-36.7]<br>(n=38439) | 31.3<br>[26.7-36.2]<br>(n=669) | 30.7<br>[26.6-35.9]<br>(n=2410) | 0.340 |
| Left ventricular circumferential strain (global) % | -22.3<br>[-24.5--20.2]<br>(n=39052) | -22.8<br>[-24.9--20.7]<br>(n=689) | -22.3<br>[-24.3--20.3]<br>(n=2477) | <0.001<br>(r=0.069) |
| **Left ventricular longitudinal strain (global) %** | -18.4<br>[-20.2--16.8]<br>(n=38003) | -18.8<br>[-20.4--17.1]<br>(n=667) | -18.5<br>[-20.1--16.9]<br>(n=2409) | 0.083 |
| **Left ventricular radial strain (global) %** | 44.8<br>[39.5-50.3]<br>(n=39052) | 46.0<br>[40.6-51.2]<br>(n=689) | 44.4<br>[39.5-49.8]<br>(n=2477) | <0.001<br>(r=0.070) |
| **Right ventricular ejection fraction %** | 57.6<br>[53.5-61.3]<br>(n=39120) | 58.4<br>[54.1-62.2] | 57.2<br>[53.7-60.9] | <0.001<br>(r=0.064) |
| **Right ventricular end-diastolic volume (BSA-indexed) mL/m<sup>2</sup>** | 82.3<br>[72.9-92.9]<br>(n=38439) | 82.9<br>[73.6-94.1]<br>(n=669) | 81.6<br>[72.7-92.3]<br>(n=2410) | 0.077 |
| **Right ventricular end-systolic volume (BSA-indexed) mL/m<sup>2</sup>** | 34.9<br>[29.2-41.4]<br>(n=38439) | 34.8<br>[28.9-41.4]<br>(n=669) | 34.8<br>[29.3-41.3]<br>(n=2410) | 0.601 |

**Table 1.** (B) **Optical flow velocity phenotypes differ significantly between HFpEF and healthy reference participants.** Ten optical flow velocity phenotypes showed statistically significant differences between the HFpEF subcohort (n = 690) and the healthy reference subcohort (n = 2,483) after cluster-adjusted Benjamini-Hochberg false discovery rate (FDR) correction (q < 0.05) and ANCOVA sensitivity analysis. Residual values reflect age-and sex-adjusted velocity residuals; raw values reflect unadjusted velocity measurements. P-values are from Mann-Whitney U tests comparing the HFpEF and healthy subcohorts. All 10 phenotypes were lower in HFpEF. Effect size r = Z/√N where Z is the Mann-Whitney standardized statistic. ANCOVA = analysis of covariance; FDR = false discovery rate; HFpEF = heart failure with preserved ejection fraction; IQR = interquartile range; LV = left ventricular; RV = right ventricular.

| Parameter | HFpEF Residual Median [IQR] (cm/s) | Healthy Residual Median [IQR] (cm/s) | HFpEF Raw Median [IQR] (cm/s) | Healthy Raw Median [IQR] (cm/s) | p-value | FDR q-value | Effect Size (r) |
| --- | --- | --- | --- | --- | --- | --- | --- |
| Late Diastolic (A-Wave) LV Inner Total Velocity | -48.39 [-125.00 to 28.22] | -18.03 [-91.91 to 55.85] | 183.52 [141.93 to 245.12] | 198.69 [153.55 to 259.81] | 1.67E-16 | 4.79E-15 | 0.147 |
| Late Diastolic (A-Wave) LV Inner Circumferential Velocity | -25.33 [-69.15 to 18.50] | -8.40 [-50.31 to 33.51] | 101.21 [71.87 to 135.31] | 109.91 [81.04 to 144.09] | 6.25E-15 | 8.95E-14 | 0.139 |
| Late Diastolic (A-Wave) LV Inner Radial Velocity | -31.61 [-85.75 to 22.54] | -14.81 [-67.48 to 37.85] | 128.52 [98.97 to 176.38] | 139.23 [106.75 to 184.71] | 2.17E-14 | 2.07E-13 | 0.136 |
| Late Diastolic (A-Wave) RV Inner Circumferential Velocity | -22.28 [-69.16 to 24.60] | -10.64 [-59.34 to 38.07] | 136.83 [108.27 to 172.34] | 145.50 [114.60 to 181.99] | 1.32E-09 | 9.46E-09 | 0.108 |
| Late Diastolic (A-Wave) RV Inner Total Velocity | -34.98 [-115.18 to 45.22] | -18.44 [-97.90 to 61.03] | 212.72 [170.44 to 265.33] | 226.04 [180.07 to 283.90] | 5.13E-09 | 2.94E-08 | 0.104 |
| Mid-Diastolic (L-Wave) LV Inner Circumferential Velocity | -29.79 [-80.82 to 21.24] | -17.44 [-74.39 to 39.51] | 123.71 [93.66 to 169.09] | 135.60 [101.27 to 187.81] | 6.59E-06 | 3.04E-05 | 0.080 |
| Late Diastolic (A-Wave) RV Inner Radial Velocity | -21.42 [-71.69 to 28.86] | -11.75 [-60.99 to 37.50] | 127.32 [97.06 to 162.75] | 133.26 [103.56 to 171.93] | 7.44E-06 | 3.04E-05 | 0.080 |
| Mid-Diastolic (L-Wave) RV Inner Circumferential Velocity | -20.17 [-77.27 to 36.92] | -12.10 [-68.59 to 44.39] | 140.95 [102.17 to 188.66] | 150.29 [112.55 to 198.09] | 9.98E-04 | 0.004 | 0.059 |
| Mid-Diastolic (L-Wave) RV Myocardial Free Wall Velocity | -0.08 [-2.60 to 2.44] | 0.42 [-2.01 to 2.85] | 44.74 [42.65 to 46.89] | 45.24 [42.89 to 47.25] | 0.002 | 0.005 | 0.056 |
| Mid-Diastolic (L-Wave) LV Inner Total Velocity | -40.70 [-143.16 to 61.76] | -21.97 [-131.48 to 87.55] | 235.89 [161.99 to 325.91] | 250.13 [172.66 to 344.28] | 0.006 | 0.017 | 0.049 |

We computed Pearson correlation coefficients between the 32 age-and sex-adjusted optical flow velocity phenotypes and the 10 age-and sex-adjusted conventional cardiac parameters. Of 320 pairwise comparisons, 278 reached cluster-adjusted Bonferroni significance (**Supplemental Figure 3**). The strongest associations involved mid-diastolic (L-wave) and late diastolic (A-wave) velocities, which correlated positively with LV cardiac output and negatively with ventricular volumes. The strongest association was between mid-diastolic (L-wave) LV inner circumferential velocity and LV cardiac output indexed to body surface area (BSA) (r = 0.394, R² = 0.155, p < 1×10⁻¹⁰). However, conventional cardiac parameters explained at most 15.5% of the variance in any individual optical flow phenotype.

### Optical Flow Velocity Phenotypes Are Associated with All-Cause Mortality

In the full cohort, we performed univariate survival analysis using Cox proportional hazards models, adjusting for age and sex. Eight conventional cardiac parameters and 13 optical flow velocity parameters were significantly associated with mortality (**Supplemental Table 3, Supplemental Figure 4**). Higher early diastolic (E-wave) velocities were associated with decreased risk; higher mid-diastolic (L-wave) and higher late diastolic (A-wave) velocities were associated with increased risk. Kaplan-Meier analyses of the strongest optical flow (**Supplemental Figure 5A**) and conventional (**Supplemental Figure 5B**) univariate predictors were concordant with these associations. Among the 21 FDR-significant univariate parameters, we advanced 10 parameters (4 conventional, 6 optical flow) to multivariable modeling (**Supplemental Data 3**). In multivariable Cox models restricted to a common cohort with complete data for all included variables (n = 37,043; 1,050 deaths; death rate 2.8%), the Combined model (conventional plus optical flow parameters) achieved concordance index [C-index] 0.732, modestly exceeding both the Conventional-only model (C-index 0.729) and the Optical Flow-only model (C-index 0.726). Optical flow parameters added significant incremental prognostic value to conventional parameters (likelihood ratio test χ² = 13.1, df = 6, p = 0.042) and vice versa (χ² = 51.6, df = 4, p = 1.65×10⁻¹⁰).

In the HFpEF subcohort, we performed univariate survival analysis using Cox proportional hazards models, adjusting for age and sex. No conventional cardiac parameters were significantly associated with mortality. One optical flow velocity parameter, systolic LV myocardial velocity, was significantly associated with survival (HR 0.61 per SD, 95% CI 0.45-0.82, q = 0.04996), with higher systolic velocities conferring lower mortality risk (**Figure 2**). Kaplan-Meier analysis confirmed this finding (**Supplemental Figure 5C**).

**Figure 2.**
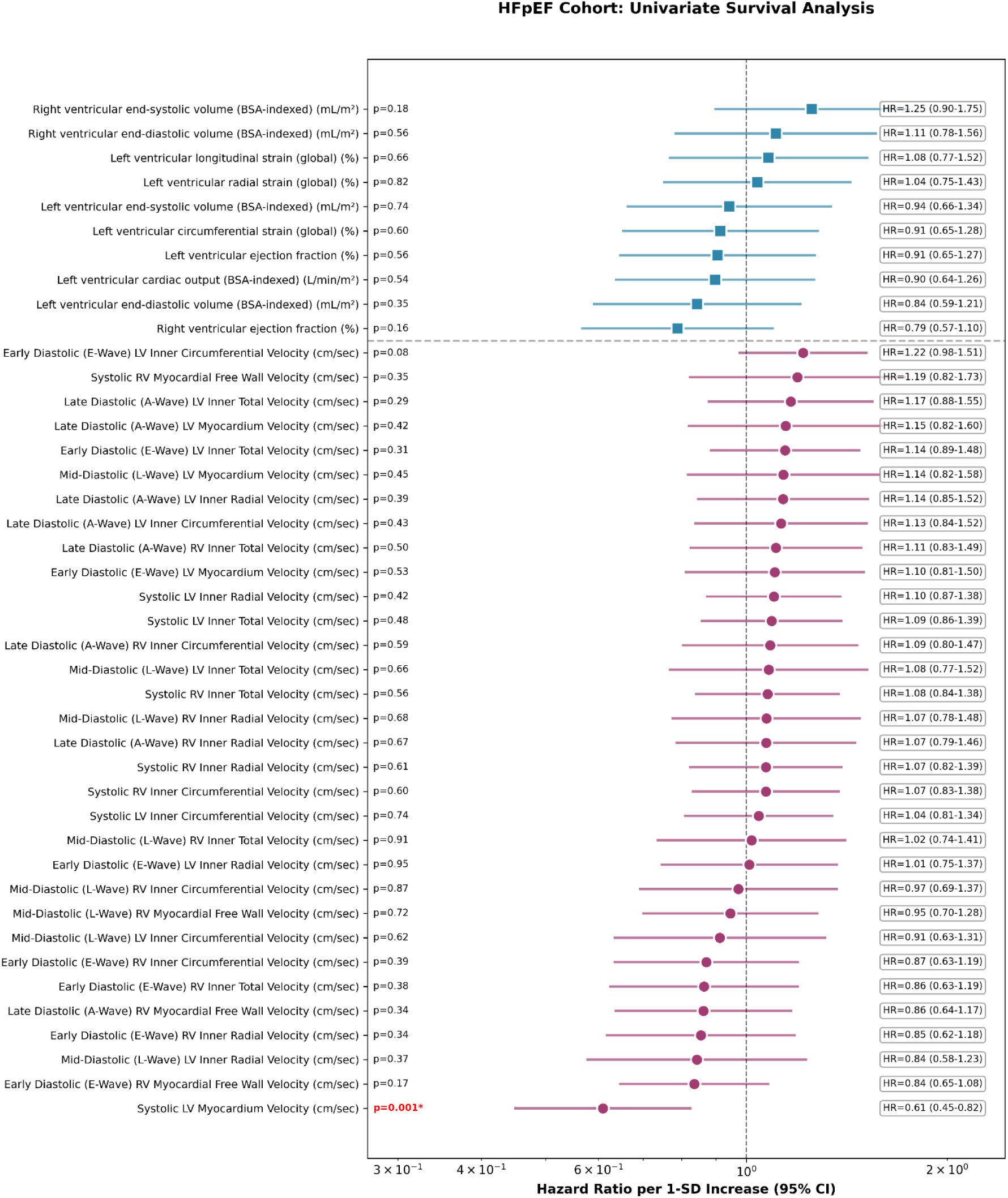
HFpEF subcohort univariate survival analysis forest plot. Sample sizes varied by parameter availability (Supplemental Table 1, Supplemental Table 2), with univariate Cox complete-case samples ranging from 667 to 690 participants with 34 to 38 deaths (death rate 5.1-5.5%), over median follow-up of 6.3 years (IQR 5.2-7.8 years). Hazard ratios per 1-SD increase (95% confidence intervals) from Cox proportional hazards models adjusted for age and sex. All 42 candidate parameters (10 conventional cardiac parameters [blue squares], 32 optical flow phenotypes [magenta circles]) are shown. Only one parameter, systolic LV myocardial velocity (HR 0.61, 95% CI 0.45-0.82, q = 0.04996), reached significance after cluster-adjusted FDR correction (red, asterisk). No conventional cardiac parameters were significantly associated with mortality in the HFpEF subcohort. FDR = false discovery rate; HFpEF = heart failure with preserved ejection fraction; LV = left ventricular; RV = right ventricular; SD = standard deviation.

### Proteome-Wide Association Analyses Identify a Diastolic-Specific Proteomic Signature

Proteome-wide association analysis of optical flow velocity phenotypes identified 344 significant protein–phenotype associations spanning 129 unique proteins (**Supplemental Table 4**). Significant associations were concentrated in diastolic velocities: 323 of 344 associations involved late diastolic (A-wave) velocities, 16 mid-diastolic (L-wave) velocities, and 5 systolic (S-wave) velocities. N-terminal pro-B-type natriuretic peptide (NT-proBNP) demonstrated the most significant association (with late diastolic [A-wave] LV inner velocity, r = −0.117, p = 7.29×10⁻²⁶). Higher NT-proBNP levels were associated with reduced late diastolic (A-wave) LV inner velocities, reduced mid-diastolic (L-wave) LV inner velocities, and reduced late diastolic (A-wave) RV inner velocities.

Proteome-wide association analysis of conventional parameters identified 1,141 significant protein–parameter associations across 396 unique proteins (**Supplemental Table 5**). The strongest associations were with proteins strongly linked to metabolic syndrome and systemic inflammation, including FABP4 (r = −0.250, p = 2.94×10⁻⁶³), leptin (r = −0.244, p = 1.66×10⁻⁶⁰), and IL1RN (r = −0.217, p = 7.07×10⁻⁴⁷).

Comparing the two proteome-wide association analyses, we observed broad overlap (**Supplemental Table 6**) but also a distinct optical-flow-specific signature that included a DCN-ERBB3 interaction hub (**Supplemental Data 4**). The 12 proteins uniquely associated with optical flow parameters were exclusively associated with mid-diastolic (L-wave) and late diastolic (A-wave) velocity phenotypes. The most significant optical-flow-unique association overall was with ACE2 (angiotensin-converting enzyme 2; r = 0.068, p = 9.18×10⁻¹⁰), an important enzyme in the renin-angiotensin-aldosterone system^13^.

### Genome-Wide Association Studies Identify 12 Genomic Risk Loci

We performed GWAS for all 32 optical flow velocity phenotypes in up to 73,978 European-ancestry UK Biobank participants and estimated SNP-based heritability using LD score regression. Twenty-five of the 32 optical flow velocity phenotypes demonstrated significant heritability after standard Bonferroni correction (p < 1.56 × 10⁻³), with h² estimates ranging from 0.036 to 0.10 (**Supplemental Table 7; Supplemental Figure 6A**). Heritability was highest for diastolic velocity phenotypes, with the strongest estimates observed for mid-diastolic (L-wave) RV inner cavity circumferential velocity (h² = 0.100, SE = 0.013, p = 6.41 × 10⁻¹⁵) and mid-diastolic (L-wave) LV inner cavity circumferential velocity (h² = 0.096, SE = 0.013, p = 3.36 × 10⁻¹³). All 24 inner cavity velocity phenotypes (LV inner and RV inner) were significantly heritable, whereas myocardial phenotypes showed lower and largely non-significant heritability.

Pairwise genetic correlations revealed that the genetic architecture of optical flow velocity phenotypes was organized by cardiac structure (left ventricle vs. right ventricle) and cardiac cycle phase (**Supplemental Table 8; Supplemental Figure 6B**). Genetic correlations between the left and right ventricles within the same cardiac phase were notably strong for mid-diastolic (L-wave) and late diastolic (A-wave) velocity phenotypes. LV inner cavity mid-diastolic (L-wave) velocity and RV inner cavity mid-diastolic (L-wave) velocity showed near-complete genetic correlation (mean rg = 0.93). Late diastolic (A-wave) motion demonstrated strong cross-ventricle correlation (mean rg = 0.74).

Across all 32 optical flow velocity phenotypes, we identified 2,599 SNP–phenotype associations at conventional genome-wide significance (p < 5×10⁻⁸); 1,212 remained significant after cluster-corrected Bonferroni correction (p < 3.85×10⁻⁹; **Supplemental Table 9**), including 1,206 autosomal associations across 17 phenotypes. Manhattan plots for the 17 phenotypes with autosomal significant associations are shown in **Supplemental Figure 7**. By merging significant SNP–phenotype associations within 1 Mb into independent loci, the 1,206 significant autosomal associations mapped to 12 distinct genomic loci (**Table 2**; **Figure 3A**; **Figure 3B**). Genes at each locus were prioritized using the FUMA platform^14^ for positional mapping. The *PLN* locus (chr6q22.31) showed the strongest overall association (p = 4.1×10⁻²² for late diastolic [A-wave] LV inner circumferential velocity) and the broadest pleiotropy, reaching cluster-corrected Bonferroni significance for 13 phenotypes spanning systolic (S-wave), early diastolic (E-wave), mid-diastolic (L-wave), and late diastolic (A-wave) velocities. *PLN* encodes phospholamban, a critical regulator of SERCA2a calcium reuptake into the sarcoplasmic reticulum during diastole.^15^ The second strongest association was at *SOX5* (chr12p12.1; p = 6.6×10⁻¹⁸), which was associated exclusively with mid-diastolic (L-wave) phenotypes. *SOX5* encodes a transcription factor implicated in extracellular matrix development.^16–18^ A GWAS catalog search (FUMA^14^) showed that all 12 risk loci had previously reported associations with cardiovascular traits (**Supplemental Table 10**).

**Figure 3.**
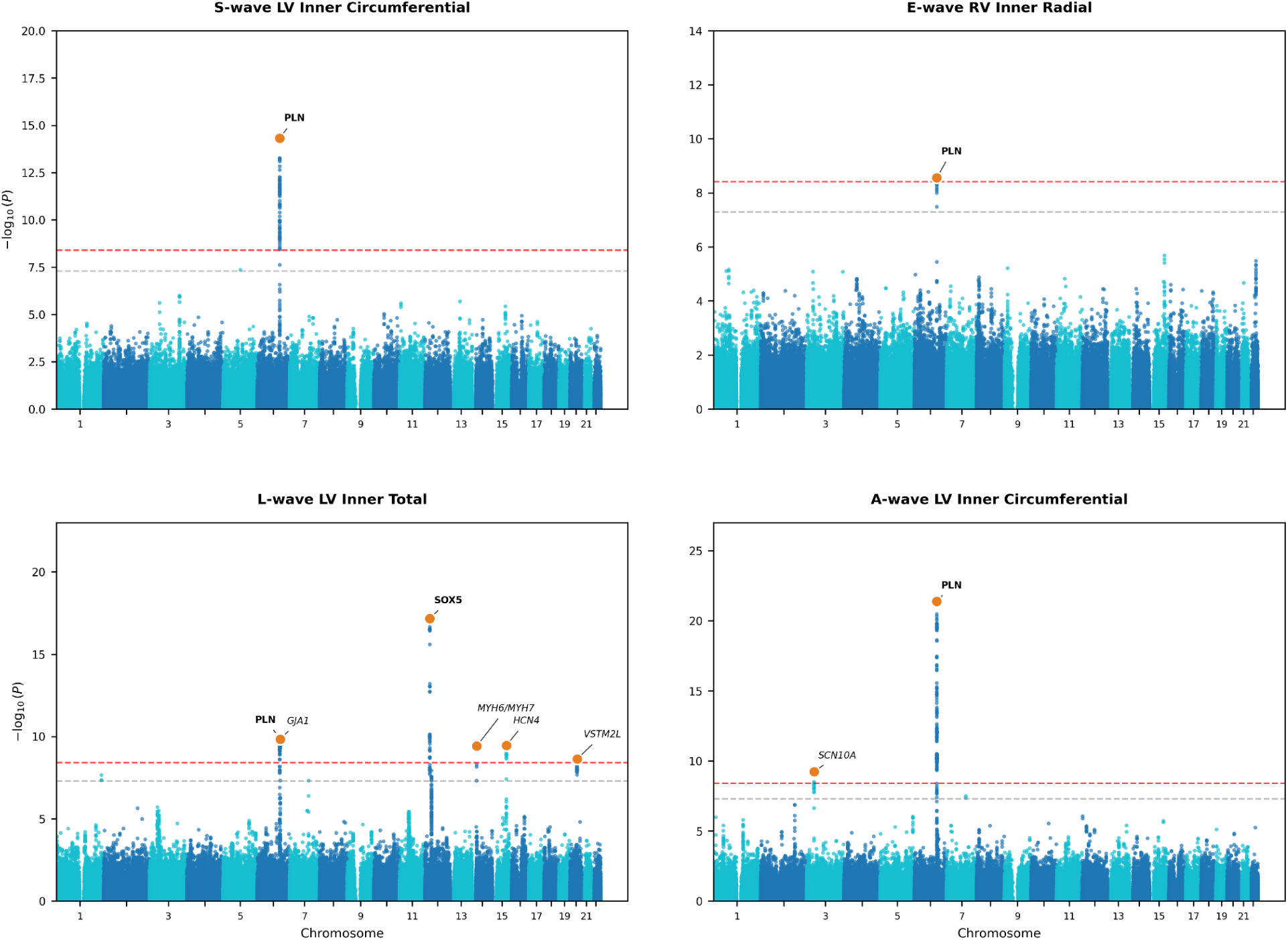
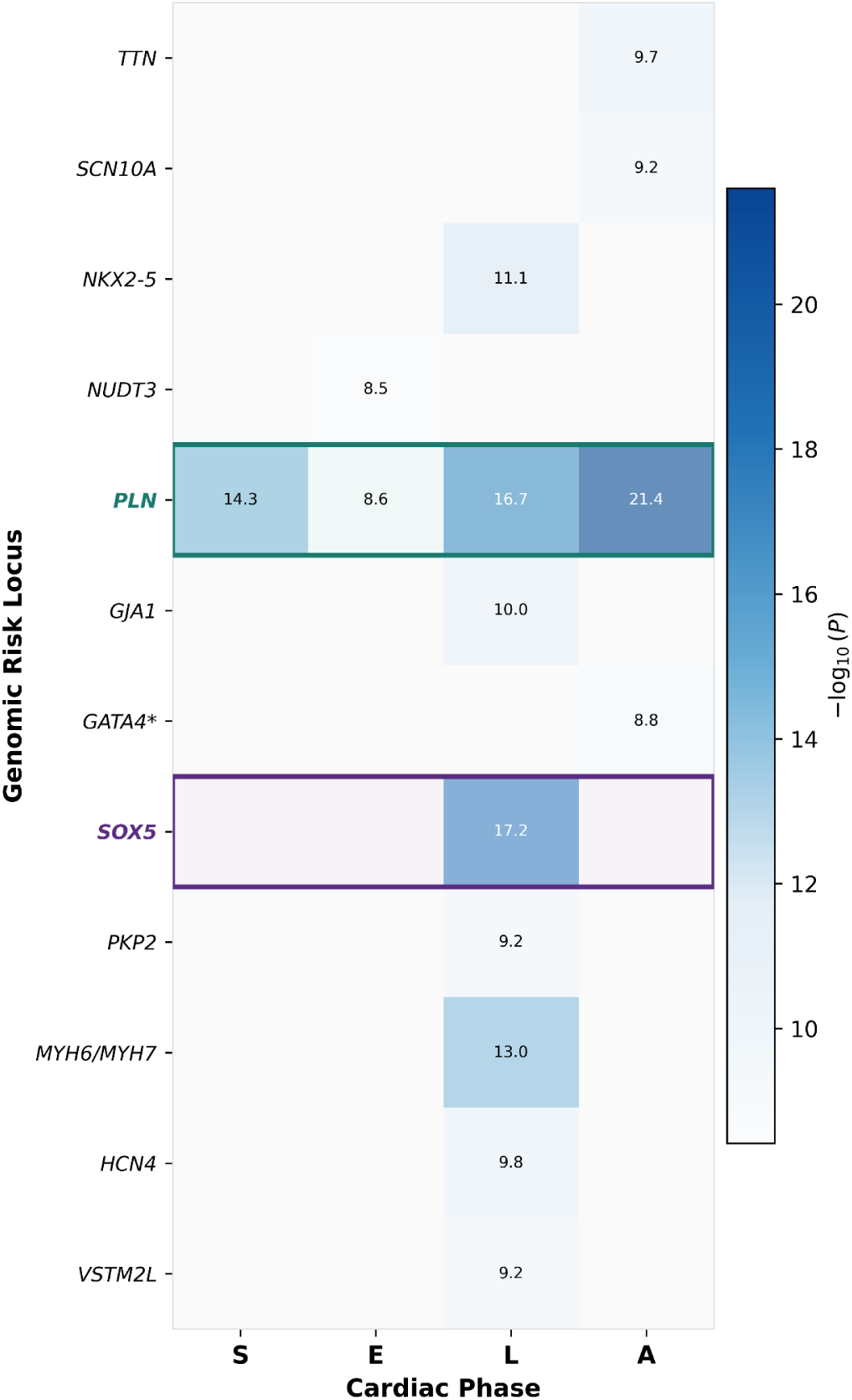
Locus-level architecture of optical flow velocity phenotypes across the cardiac cycle. (A) Representative Manhattan plots, one per cardiac phase. Each panel shows genome-wide association results for the optical flow velocity phenotype with the strongest lead-SNP association within each cardiac phase: S-wave LV Inner Circumferential, E-wave RV Inner Radial, L-wave LV Inner Total, and A-wave LV Inner Circumferential. Horizontal dashed lines indicate conventional genome-wide significance (gray, P = 5×10⁻⁸) and the cluster-corrected Bonferroni threshold applied throughout this study (red, P = 3.85×10⁻⁹). Orange markers denote lead SNPs at genome-wide significant loci. At chromosome 6q22 in the L-wave panel, the visible peak corresponds to two independent genomic risk loci, PLN (6q22.31) and GJA1 (6q22.33). **(B) Genomic locus × cardiac phase map for optical flow velocity phenotypes.** Each row represents one of 12 genomic risk loci identified by merging genome-wide significant SNP-phenotype associations within 1 Mb (Table 2). Each column represents a cardiac phase: S = systolic, E = early diastolic (E-wave), L = mid-diastolic (L-wave), A = late diastolic (A-wave). Cell color and annotation indicate −log₁₀(P) of the most significant SNP at that locus across all phenotypes within that cardiac phase; only associations reaching cluster-corrected Bonferroni significance (p < 3.85×10⁻⁹) are colored. Gray cells indicate no association at this threshold. PLN (green border) demonstrates broad pleiotropy across all four cardiac phases; SOX5 (purple border) is exclusive to mid-diastolic (L-wave) phenotypes. SNP = single nucleotide polymorphism.

**Table 2.**
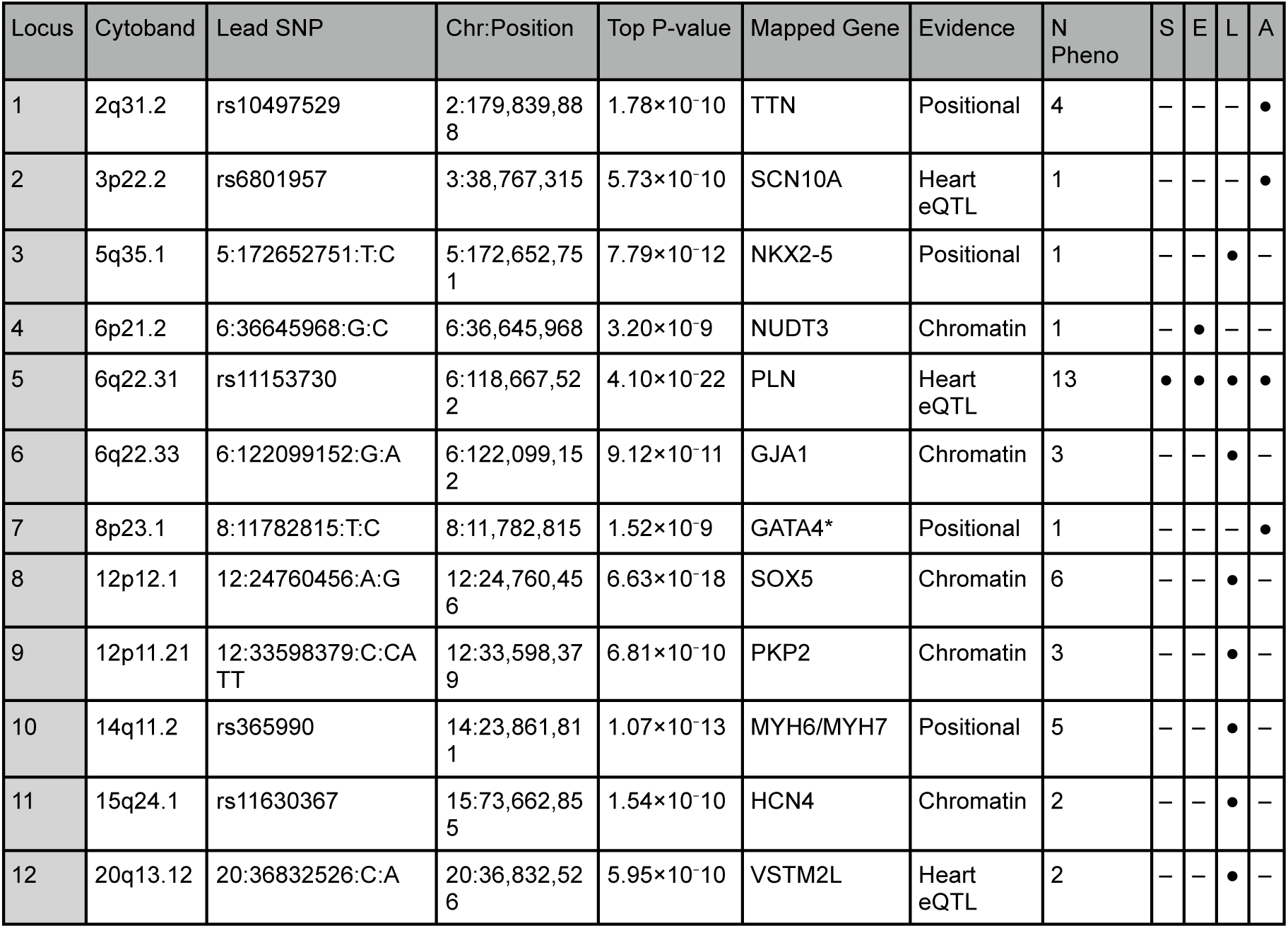
Genomic risk loci for optical flow velocity phenotypes. Loci were identified by merging the 1,206 SNP-phenotype autosomal associations reaching cluster-corrected Bonferroni significance (p < 3.85×10⁻⁹; **Supplemental Detailed Methods**) within 1 Mb into independent loci, across 17 phenotypes with autosomal hits reaching this threshold. Lead SNP = SNP with the lowest p-value at each locus across all phenotypes. Chr:Position = genomic coordinates of the lead SNP (GRCh37/hg19). Mapped Gene = genes at each locus were prioritized using the FUMA platform for positional mapping. Evidence = primary gene mapping evidence (Heart eQTL: SNPs at this locus are associated with the mapped gene’s expression in cardiac tissue; Chromatin: chromatin interaction links the locus to the mapped gene in cardiac tissue; Positional: gene prioritized by proximity and biological plausibility). Phase columns = ● indicates at least one association with cluster-corrected significance; – indicates no associations with cluster-corrected Bonferroni significance; S indicates systolic; E indicates early diastolic (E-wave); L indicates mid-diastolic (L-wave); A indicates late diastolic (A-wave). *GATA4 is ∼160kb downstream, nearest mapped transcript is AF131215.9. eQTL = expression quantitative trait loci; LV = left ventricular; RV = right ventricular; SNP = single nucleotide polymorphism.

| Locus | Cytoband | Lead SNP | Chr:Position | Top P-value | Mapped Gene | Evidence | N Pheno | S | E | L | A |
| --- | --- | --- | --- | --- | --- | --- | --- | --- | --- | --- | --- |
| 1 | 2q31.2 | rs10497529 | 2:179,839,888 | $1.78 \times 10^{-10}$ | TTN | Positional | 4 | – | – | – | • |
| 2 | 3p22.2 | rs6801957 | 3:38,767,315 | $5.73 \times 10^{-10}$ | SCN10A | Heart eQTL | 1 | – | – | – | • |
| 3 | 5q35.1 | 5:172652751:T:C | 5:172,652,751 | $7.79 \times 10^{-12}$ | NKX2-5 | Positional | 1 | – | – | • | – |
| 4 | 6p21.2 | 6:36645968:G:C | 6:36,645,968 | $3.20 \times 10^{-9}$ | NUDT3 | Chromatin | 1 | – | • | – | – |
| 5 | 6q22.31 | rs11153730 | 6:118,667,522 | $4.10 \times 10^{-22}$ | PLN | Heart eQTL | 13 | • | • | • | • |
| 6 | 6q22.33 | 6:122099152:G:A | 6:122,099,152 | $9.12 \times 10^{-11}$ | GJA1 | Chromatin | 3 | – | – | • | – |
| 7 | 8p23.1 | 8:11782815:T:C | 8:11,782,815 | $1.52 \times 10^{-9}$ | GATA4* | Positional | 1 | – | – | – | • |
| 8 | 12p12.1 | 12:24760456:A:G | 12:24,760,456 | $6.63 \times 10^{-18}$ | SOX5 | Chromatin | 6 | – | – | • | – |
| 9 | 12p11.21 | 12:33598379:C:CA TT | 12:33,598,379 | $6.81 \times 10^{-10}$ | PKP2 | Chromatin | 3 | – | – | • | – |
| 10 | 14q11.2 | rs365990 | 14:23,861,811 | $1.07 \times 10^{-13}$ | MYH6/MYH7 | Positional | 5 | – | – | • | – |
| 11 | 15q24.1 | rs11630367 | 15:73,662,855 | $1.54 \times 10^{-10}$ | HCN4 | Chromatin | 2 | – | – | • | – |
| 12 | 20q13.12 | 20:36832526:C:A | 20:36,832,526 | $5.95 \times 10^{-10}$ | VSTM2L | Heart eQTL | 2 | – | – | • | – |

### RABGAP1L and C2 Are Candidate Causal Mediators of Optical Flow Velocity Phenotypes

We performed two-sample Mendelian randomization using cis-protein quantitative trait loci (cis-pQTLs) from the UK Biobank Pharma Proteomics Project^19^ as genetic instruments. Across 26,886 protein–phenotype pairs, inverse-variance-weighted (IVW) MR with Bonferroni correction (p < 1.86×10⁻⁶) identified 88 significant associations across 43 unique proteins. After applying MR-PRESSO, 81 protein–phenotype associations across 38 unique proteins were carried forward to cis-pQTL colocalization (**Supplemental Table 11; Supplemental Figure 8**).

RABGAP1L showed the most significant MR associations overall, across 8 diastolic velocity phenotypes and both ventricles (**Supplemental Figure 8**). Higher genetically predicted RABGAP1L levels were associated with decreased early diastolic (E-wave) and decreased mid-diastolic (L-wave) velocities. RABGAP1L’s strongest association was with early diastolic RV inner circumferential velocity (IVW p = 5.49×10⁻²², β = −0.255 per unit Normalized Protein eXpression [NPX], 15 instruments), which showed consistent effect direction across weighted median (β = −0.261) and MR-Egger (β = −0.265) methods and colocalized with high confidence (PP.H4 = 0.937) (**Supplemental Figure 9A**). RABGAP1L encodes a Rab GTPase-activating protein that interacts with Ankyrin-B, a scaffold protein critical for localizing calcium handling machinery at the cardiomyocyte membrane.^20,21^ C2 showed significant MR association with systolic LV inner velocity (IVW p = 5.05×10⁻⁷, β = 0.034 per unit NPX, 39 instruments) and colocalized with high confidence (PP.H4 = 0.966). Higher genetically predicted C2 levels were associated with increased systolic velocity (**Supplemental Figure 9B**).

eQTL colocalization results in the context of bulk cardiovascular tissue are shown in **Supplemental Table 12** and **Supplemental Data 5**. STRING-DB analysis on the 38 proteins with MR-PRESSO-validated causal associations revealed a significantly enriched interaction network converging on two biological themes: vesicular trafficking and endosomal processing, and extracellular matrix remodeling (**Supplemental Data 6; Supplemental Table 13; Supplemental Figure 10**).

## DISCUSSION

Disease-specific therapies remain limited in HFpEF in part because the cardiac motion abnormalities central to its pathophysiology have not been comprehensively characterized at population scale. We address this gap with a deep-learning-enabled pipeline for extracting cardiac motion phenotypes from standard cine CMR. By integrating optical flow motion analysis with nnU-Net segmentation and applying this framework to CMR images from 83,569 UK Biobank participants, we derived 32 biventricular myocardial and inner cavity velocity phenotypes across the cardiac cycle, including the first population-level quantification of mid-diastolic (L-wave) motion. Our work provides a coherent diagnostic and molecular framework for the cardiac motion abnormalities relevant to HFpEF pathophysiology.

Optical flow velocity phenotypes captured features of cardiac motion not fully reflected by conventional cardiac parameters. Ten optical flow velocity phenotypes were significantly reduced in HFpEF, with all significant findings concentrated in mid-diastolic (L-wave) and late diastolic (A-wave) phases across both ventricles. In contrast, differences in conventional cardiac parameters between HFpEF and healthy reference subcohorts were clinically negligible, despite some reaching statistical significance. Moreover, conventional cardiac parameters explained at most 15.5% of the variance in any individual optical flow phenotype. Together, these findings position optical flow velocity phenotypes as candidate metrics for objective HFpEF identification.

Survival analyses revealed that optical flow velocity phenotypes are associated with all-cause mortality. In the full cohort, optical flow and conventional parameters contributed statistically significant bidirectional incremental prognostic value, indicating they capture complementary rather than redundant information. In the HFpEF subcohort, systolic LV myocardial velocity was the only significant predictor of survival (HR 0.61 per SD, 95% CI 0.45-0.82). The protective association of systolic LV myocardial velocity suggests that optical flow may capture subtle variation in contractile function that conventional parameters do not detect in HFpEF.

Proteomic associations with optical flow phenotypes and conventional cardiac parameters shared a broad landscape of metabolic and inflammatory proteins. NT-proBNP, the clinical gold standard biomarker for heart failure^22^, was the most significant protein association for optical flow velocity phenotypes. Higher circulating NT-proBNP levels were associated with reduced mid-diastolic (L-wave) and late diastolic (A-wave) velocities, supporting the biologic validity of the optical flow velocity phenotypes. The 12 proteins uniquely associated with optical flow velocity phenotypes, concentrated in mid-diastolic (L-wave) and late diastolic (A-wave) phases, revealed a distinct biological signal that included a DCN-ERBB3 interaction hub and ACE2 as the top individual association, pointing toward extracellular matrix remodeling and neurohumoral regulation as biological themes captured by spatiotemporal diastolic phenotypes.

We also established the heritability of optical flow velocity phenotypes, supporting their use as genetically informative traits. Our h² estimates ranged from 0.036 to 0.10, consistent with prior GWAS studies showing that velocity-and flow-related features are approximately 10% heritable or less,^23^ as such features are significantly influenced by environmental factors such as cardiac loading conditions.

GWAS identified 12 risk loci associated with optical flow velocity phenotypes and highlighted the relationship among intracellular calcium handling pathways, extracellular matrix development, and cardiac motion. The PLN locus at chr6q22.31 showed the strongest association and broadest pleiotropy, reaching cluster-corrected Bonferroni significance for 13 phenotypes across all cardiac phases. PLN encodes phospholamban, the canonical regulator of the SERCA2a calcium pump^15^. The activity of SERCA2a determines the rate of removal of >70% of cytosolic calcium in human cardiomyocytes^15^ and governs both the rate of myocardial relaxation and the strength of the subsequent contraction. In an independent GWAS of diastolic function, Thanaj et al. independently identified PLN as significantly associated with peak early diastolic strain rate^24^. Our data extend this work by showing that PLN is associated with velocity phenotypes across systole (S-wave), early diastole (E-wave), mid-diastole (L-wave), and late diastole (A-wave), consistent with PLN’s established influence spanning both cardiac relaxation and contractility.

The second strongest association in our GWAS was at SOX5, which reached cluster-corrected Bonferroni significance exclusively with mid-diastolic (L-wave) velocity phenotypes. SOX5 is a transcription factor involved in extracellular matrix development.^16,17^ Direct functional evidence for a cardiac role comes from a transgenic Drosophila model in which silencing of the highly conserved SOX5 ortholog Sox102F led to significant decreases in heart rate, chamber size, and cardiac wall velocities, as well as significant increases in cardiac wall thickness associated with disrupted myofibril structure.^18^ The specificity of the association between SOX5 and L-wave velocities in our study suggests a relationship between cardiac extracellular matrix composition and the physiology of mid-diastolic (L-wave) filling.

Mendelian randomization identified RABGAP1L as a candidate causal mediator of early diastolic velocity. RABGAP1L showed the most significant MR association with early diastolic (E-wave) RV inner circumferential velocity (IVW p = 5.49×10⁻²², 15 cis-pQTL instruments), colocalizing with high confidence (PP.H4 = 0.937). Higher genetically predicted RABGAP1L levels were associated with decreased early-and mid-diastolic (E-and L-wave) velocities across eight phenotypes spanning both ventricles, though robust colocalization support was specific to the early-diastolic signal. We hypothesize the causal link between RABGAP1L and impaired early diastolic (E-wave) motion is mediated through the ankyrin-B (AnkB) scaffolding complex and AnkB’s role in intracellular calcium handling, based on prior data showing AnkB-RABGAP1L interactions as part of a coordinated sequence of endosomal trafficking of specialized membrane proteins (**Supplemental Data 7**). We hypothesize that elevated RABGAP1L levels may influence diastolic velocities through effects on endosomal transport and membrane assembly of the AnkB/NCX1/Na/K-ATPase complex, potentially affecting cytosolic calcium extrusion during diastole (**Figure 4)**. This hypothesized mechanism links our MR findings with the PLN signal at the GWAS level, supporting a potential role for intracellular calcium handling in cardiac motion phenotypes across independent layers of multi-omic evidence.

**Figure 4.**
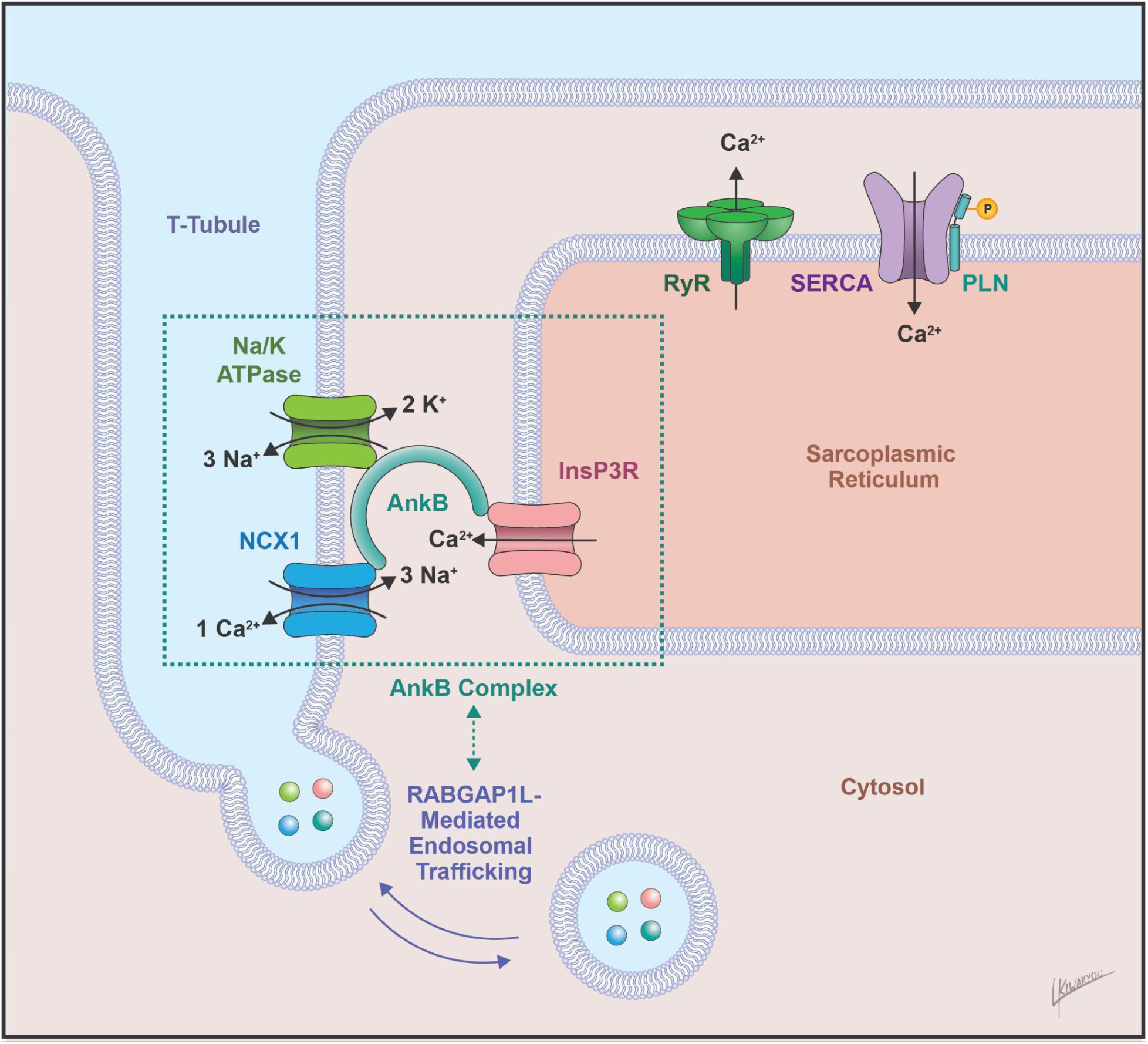
Proposed mechanism linking RABGAP1L to diastolic velocity via the AnkB/NCX1/Na-K-ATPase complex. Schematic of the cardiomyocyte T-tubule membrane illustrating the AnkB-based complex (comprising Na/K-ATPase, NCX1, and InsP3R), which is involved in the regulation of cytosolic calcium. RABGAP1L interacts with AnkB in endosomal trafficking processes and polarized membrane assembly of specialized membrane proteins. We hypothesize that elevated RABGAP1L levels impair diastolic velocities through disruption of this endosomal trafficking pathway, affecting membrane localization of the AnkB/NCX1/Na-K-ATPase complex and impairing cytosolic calcium regulation. The PLN GWAS signal implicates the SERCA/PLN axis as a convergent, independent layer of genomic evidence for intracellular calcium handling as a determinant of cardiac motion phenotypes. AnkB = ankyrin-B; InsP3R = inositol 1,4,5-trisphosphate receptor; NCX1 = sodium-calcium exchanger 1; PLN = phospholamban; RABGAP1L = Rab GTPase-activating protein 1-like; RyR = ryanodine receptor; SERCA = sarco/endoplasmic reticulum Ca²⁺-ATPase.

We acknowledge the limitations of our study. The UK Biobank cohort demonstrates healthy-volunteer bias^25^ and is predominantly of European ancestry, limiting generalizability of our findings. Our HFpEF and healthy reference subcohorts were defined using objective biomarker and imaging criteria rather than symptom assessment, an approach necessitated by the absence of contemporaneously documented clinical symptom data in the UK Biobank imaging cohort. Event counts in the HFpEF subgroup were modest, constraining statistical power. We were unable to validate our MR findings in the deCODE cohort, as its proteomics resource is based on the SomaScan platform, which does not provide an independent cis-pQTL instrument for RABGAP1L. Mechanistic validation of the prioritized causal proteins is required.

Our findings have implications for the mechanistic understanding of HFpEF pathophysiology and nominate optical flow velocities as candidate objective motion phenotypes for HFpEF identification and prognosis. We identify two distinct biological axes, intracellular calcium handling and extracellular matrix composition, as potential pathways underlying reduced diastolic velocity phenotypes at the population level. This work establishes a scalable framework for deriving motion phenotypes from cardiac imaging databases and linking them to population genomics, molecular mechanisms, and, ultimately, precision cardiovascular medicine.

## Data Availability

DATA AVAILABILITY
UK Biobank data are available to approved researchers through the UK Biobank Access Management System (https://www.ukbiobank.ac.uk/enable-your-research/apply-for-access); this research was conducted under application number 22282. The UK Biobank Pharma Proteomics Project proteomic data and the conventional cardiac MRI parameters are available within the UK Biobank resource under the same application. The Genotype-Tissue Expression (GTEx) v10 datasets are available at https://www.gtexportal.org/home/datasets. The Heart Cell Atlas v2 is available at https://www.heartcellatlas.org/. The STRING database (v12.0) is available at https://string-db.org/. The 1000 Genomes Phase 3 European reference panel is publicly available. Aggregate and summary-level data supporting the findings of this study are provided in the article and its supplementary tables. The individual-level optical flow velocity phenotypes derived in this study, and any other individual-level UK Biobank data, are subject to UK Biobank access restrictions and are available to approved researchers through the process described above.
CODE AVAILABILITY
The custom software for cardiac motion phenotyping developed in this study, including trained nnU-Net segmentation weights, optical flow computation (Dual TV-L1), segmentation-masked velocity waveform extraction, and peak detection, is available at https://github.com/KRSteffner/cardiac-motion-phenotyping. The repository provides source code, a Dockerfile, and instructions to build a self-contained Docker image for processing user-supplied four-chamber long-axis standard cine CMR DICOM data; no participant-level UK Biobank data are included. Segmentation used an nnU-Net model trained on manually annotated CMR frames; optical flow was computed using the Dual TV-L1 algorithm; and downstream genetic analyses used PLINK2, FUMA, LD score regression (LDSC), coloc, and the TwoSampleMR (v0.6.14), MendelianRandomization (v0.10.0), and MR-PRESSO R packages, as described in Methods.

https://github.com/KRSteffner/cardiac-motion-phenotyping

## ACKNOWLEDGEMENTS

This research was conducted using the UK Biobank Resource under application number 22282. We thank the UK Biobank participants and the UK Biobank team.

## DATA AVAILABILITY

UK Biobank data are available to approved researchers through the UK Biobank Access Management System (https://www.ukbiobank.ac.uk/enable-your-research/apply-for-access); this research was conducted under application number 22282. The UK Biobank Pharma Proteomics Project proteomic data and the conventional cardiac MRI parameters are available within the UK Biobank resource under the same application. The Genotype-Tissue Expression (GTEx) v10 datasets are available at https://www.gtexportal.org/home/datasets. The Heart Cell Atlas v2 is available at https://www.heartcellatlas.org/. The STRING database (v12.0) is available at https://string-db.org/. The 1000 Genomes Phase 3 European reference panel is publicly available. Aggregate and summary-level data supporting the findings of this study are provided in the article and its supplementary tables. The individual-level optical flow velocity phenotypes derived in this study, and any other individual-level UK Biobank data, are subject to UK Biobank access restrictions and are available to approved researchers through the process described above.

## CODE AVAILABILITY

The custom software for cardiac motion phenotyping developed in this study, including trained nnU-Net segmentation weights, optical flow computation (Dual TV-L1), segmentation-masked velocity waveform extraction, and peak detection, is available at https://github.com/KRSteffner/cardiac-motion-phenotyping. The repository provides source code, a Dockerfile, and instructions to build a self-contained Docker image for processing user-supplied four-chamber long-axis standard cine CMR DICOM data; no participant-level UK Biobank data are included. Segmentation used an nnU-Net model trained on manually annotated CMR frames; optical flow was computed using the Dual TV-L1 algorithm; and downstream genetic analyses used PLINK2, FUMA, LD score regression (LDSC), coloc, and the TwoSampleMR (v0.6.14), MendelianRandomization (v0.10.0), and MR-PRESSO R packages, as described in Methods.

## AUTHOR CONTRIBUTIONS

K.R.S., B.G., and E.A.A. designed the study. K.R.S. trained, validated, and tested the deep learning segmentation model and developed the optical flow integration, with contribution of N.Q.; conducted the genetic and proteomic analyses, including the genome-wide association, proteome-wide association, heritability, Mendelian randomization, and colocalization analyses, with contribution of S.G.R. and R.X.; conducted the epidemiological and survival analyses; and prepared the supplementary material and tables. K.R.S. designed the figures, with contribution of L.K. K.R.S. wrote the manuscript, with contributions of B.G. and E.A.A. B.G. and E.A.A. jointly supervised the work.

## SUPPLEMENTAL DETAILED METHODS

### Identification of the UK Biobank Cohort with CMR Data

The UK Biobank is a prospective study of more than 500,000 individuals between the ages of 40 and 69 years enrolled between 2006-2010 from across the United Kingdom, with comprehensive clinical phenotyping, imaging, biomarker, and genetic data.^10^ The UK Biobank received ethical approval from the National Health Service’s National Research Ethics Service North West. The most recent release of the UK Biobank CMR dataset includes imaging data from >80,000 participants.^26–28^ In the UK Biobank data structure, participant assessments are indexed as follows: Instance 0 (initial baseline assessment, 2006-2010); Instance 1 (first repeat assessment, 2012-2013); Instance 2 (first imaging visit, beginning 2014); and Instance 3 (repeat imaging visit, beginning 2019). For this study, we restricted our analyses to standard cine cardiac magnetic resonance images (CMRs) from the first imaging visit (Instance 2), which represents the baseline CMR acquisition within the cohort. We selected for only four-chamber long-axis standard cine view acquisitions (identified by SeriesDescription tags containing “4ch,” “4-ch,” “four-chamber,” or equivalent terms), which provides simultaneous visualization of right-and left-sided cardiac structures. We identified 83,569 UK Biobank participants with available Instance 2 four-chamber long-axis standard cine CMR data.

### Automated Optical Flow-Based Cardiac Motion Phenotyping at Population Scale Optical Flow Analysis Paired with Deep Learning Segmentation

We trained an nnU-Net deep learning segmentation model^11^ to segment four key cardiac structures from four-chamber long-axis standard cine CMR sequences: the left ventricular (LV) inner cavity, the LV myocardium, the right ventricular (RV) inner cavity, and the RV myocardial free wall. Training data comprised 156 frames manually annotated by an expert clinician board-certified in cardiac imaging, randomly sampled across the cardiac cycle from randomly selected UK Biobank participants to ensure representation of diverse cycle phases and cardiac morphologies. Model performance was evaluated by five-fold cross-validation, achieving Dice similarity coefficients of 0.949 ± 0.008 (LV inner cavity), 0.906 ± 0.003 (LV myocardium), 0.923 ± 0.007 (RV inner cavity), and 0.830 ± 0.008 (RV myocardial free wall). Model weights from the best-performing fold (fold 4) were applied to all 83,569 Instance 2 four-chamber long-axis standard cine CMR images.

We applied optical flow-based motion analysis using the Dual TV-L1^7,8^ algorithm to all 83,569 Instance 2 four-chamber long-axis standard cine CMR images. The Dual TV-L1 optical flow method quantifies motion across the entire CMR image and yields frame-to-frame displacement vectors for every pixel, ultimately generating velocity vector fields between consecutive imaging frames. Segmentation masks were used to filter the resulting velocity fields, isolating motion within each of the four anatomically defined key cardiac structures (**Supplemental Figure 1).**

### Velocity Waveform Plotting

For each of the four key cardiac structures, we computed median velocity magnitudes between each consecutive CMR frame. For the myocardial regions – specifically, the LV myocardium and the RV myocardial free wall – velocity vectors exceeding 50 cm/s were excluded to remove contamination from adjacent high-velocity intracavitary motion. For the LV and RV inner cavities, velocity vectors were additionally decomposed into radial components (which reflect chamber expansion and contraction relative to the chamber centroid) and circumferential components (which reflect rotational motion relative to the chamber centroid). Frame-by-frame median velocities were plotted as continuous waveforms across the cardiac cycle.

### Waveform Peak Detection and Phenotype Extraction

To extract quantitative phenotypes from the velocity waveforms, we developed an automated peak detection algorithm that identified physiologically relevant motion events across systolic and diastolic phases of the cardiac cycle, including: the systolic (S-wave) peak, the early diastolic (E-wave) peak, the mid-diastolic (L-wave) peak, and the late diastolic (A-wave) peak (Figure 1C). We use uppercase S-/E-/L-/A-wave nomenclature to denote phases of the cardiac cycle and apply this notation to both inner cavity and myocardial velocity phenotypes. This phase-based usage is adapted from echocardiographic convention, in which uppercase letters refer to atrioventricular inflow (blood flow) velocities and lowercase letters with a prime (e.g. e’, a’) refer to tissue Doppler (myocardial) velocities^29,30^. This yielded 32 distinct velocity metrics per participant across the four cardiac structures and three velocity types: total vector, radial component, and circumferential component velocities for the LV and RV inner cavities; and total vector velocities only for the LV myocardium and the RV myocardial free wall. All 32 optical flow-derived velocity phenotypes were carried forward for downstream analyses.

### Selection of Ten Conventional Cardiac Parameters

Conventional cardiac parameters were extracted from the UK Biobank cardiac imaging database (Instance 2) for comparison against optical flow velocity phenotypes. Twenty-three CMR measurement-related fields were initially identified, spanning functional, volumetric, and strain measurements derived from three UK Biobank pipelines: the inlineVF automated analysis (Category 133), the Bai et al. 2020 deep learning pipeline (Category 157)^27^, and the Petersen et al. 2017/Aung et al. 2022 pipeline (Category 162)^31,32^. These 23 fields included the following physiologic concepts: cardiac output, LV and RV ejection fractions, LV and RV end-diastolic and end-systolic volumes, and LV circumferential, radial, and longitudinal strain. When the same physiologic concept was available from multiple pipelines (e.g. LV ejection fraction from Category 133 and Category 157), we prioritized the Category 157 measurement^27^ because of its transparent methodology using manual annotations for training a convolutional neural network-based segmentation model and rigorous quality control; and when possible, we discarded Category 133 measurements, which the UK Biobank Showcase states have known quality issues^33^. Stroke volume parameters and cardiac index were excluded as mathematically redundant with end-diastolic minus end-systolic volume and LV cardiac output indexed to BSA, respectively, to avoid adding collinear variables that would inflate multiple-testing burden without contributing independent physiologic information. Ultimately, this yielded a final panel of 10 conventional cardiac parameters for downstream analyses: LV cardiac output, LV ejection fraction, LV end-diastolic volume, LV end-systolic volume, LV circumferential strain (global), LV longitudinal strain (global), LV radial strain (global), RV ejection fraction, RV end-diastolic volume, and RV end-systolic volume. Volumetric parameters and LV cardiac output were BSA-indexed prior to statistical analyses. Among 83,569 participants with successful optical flow CMR motion analysis, conventional cardiac parameters were available in 45.5–46.8% of participants (**Supplemental Table 2**).

### Age and Sex Adjustment

For each of the 32 optical flow velocity phenotypes and 10 conventional cardiac parameters, age-and sex-adjusted residual values were generated using linear regression. Conventional volumetric parameters (LV and RV end-diastolic and end-systolic volumes) and LV cardiac output were first BSA-indexed by dividing each raw value by the participant’s BSA. All parameters were then winsorized at ±3 standard deviations from the mean to limit the influence of extreme outliers. Each parameter was modeled as a function of age and sex using ordinary least squares linear regression, fit on participants with complete data for the parameter and both covariates. Residuals (observed minus predicted) were computed and used as the age-and sex-adjusted values for downstream analyses (correlation analysis, HFpEF vs healthy comparison, and proteome-wide association analyses).

### Multiple Testing Correction

We applied multiple testing correction methods tailored to the structure of each analysis and the correlation properties of the phenotype panels tested. To account for correlation among the 32 optical flow velocity phenotypes and among the 10 conventional cardiac parameters, we estimated the effective number of independent tests using the Nyholt-Gao eigenvalue method^34^:

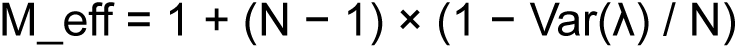

where λ denotes the eigenvalues of the Pearson correlation matrix of the phenotype panel and Var(λ) is the population variance across eigenvalues. For the 32 optical flow phenotypes, M_eff was computed on age-and sex-adjusted residuals (n = 83,127), yielding M_eff = 28.7 effective independent tests. For the 10 conventional cardiac parameters, M_eff was computed on age-and sex-adjusted residuals (n = 37,244), yielding M_eff = 8.5 effective independent tests. For the combined 42-parameter panel (32 optical flow plus 10 conventional cardiac metrics) used in survival analysis, M_eff was computed on raw values processed parallel to the survival analysis preprocessing (cardiac output and volumes body surface area-indexed, 1st/99th percentile winsorized; n = 37,048; this set differs slightly from the full cohort multivariable risk set [n = 37,043] owing to differing variable-completeness requirements), yielding M_eff = 38.8 effective independent tests.

Cluster-adjusted Bonferroni correction was used for analyses with high-dimensional test grids involving the optical flow or conventional cardiac metrics panels. For the correlation analysis (32 optical flow phenotypes × 10 conventional cardiac parameters), correction accounted for correlation along both dimensions: α / (M_eff_OF × M_eff_conventional) = 0.05 / (28.666 × 8.458) = 2.06×10⁻⁴. For the proteome-wide association studies, the threshold was α / (number of proteins × M_eff): 0.05 / (1,463 × 28.666) = 1.19×10⁻⁶ for the optical flow PWAS and 0.05 / (1,463 × 8.458) = 4.04×10⁻⁶ for the conventional cardiac parameters PWAS.

Cluster-adjusted Benjamini-Hochberg false discovery rate (BH-FDR) at q < 0.05 was used for discovery analyses involving the phenotype panels. For the HFpEF vs. healthy subcohort comparison, BH-FDR was applied across 32 optical flow phenotypes using M_eff = 28.7 as the denominator. Findings reaching cluster-adjusted FDR significance in the HFpEF vs. healthy comparison were additionally subjected to ANCOVA sensitivity testing. For univariate survival analyses (full cohort and HFpEF subcohort), BH-FDR was applied across 42 imaging parameters using M_eff = 38.8 as the denominator.

Hierarchical clustering-based Bonferroni correction was applied to the genome-wide association study (GWAS) analyses, where the dominant multiplicity arises from the SNP dimension (∼7 million imputed variants). Hierarchical clustering at distance threshold 0.5 (distance = 1 − |r|) yielded 13 phenotype clusters, giving a cluster-corrected threshold of 5×10⁻⁸ / 13 = 3.85×10⁻⁹.

Standard Bonferroni correction was applied to the SNP-based heritability analysis across 32 phenotypes (α = 0.05/32 = 1.56×10⁻³). Standard Bonferroni correction was also applied to the Mendelian randomization analysis across 26,886 unique protein–phenotype pairs (α = 0.05 / 26,886 = 1.86×10⁻⁶), providing a conservative threshold appropriate for causal inference claims.

Baseline cohort characteristics (Table 1) are reported with raw p-values from Mann-Whitney U tests (continuous variables) and chi-square or Fisher’s exact tests (categorical variables); no multiple testing correction was applied for this descriptive cohort characterization.

### HFpEF and Healthy Reference Subcohort Definition

Among the 83,569 participants with successful optical flow CMR motion analysis, we pragmatically defined a HFpEF subcohort and a healthy reference subcohort. According to consensus guidelines among international professional societies, HFpEF diagnosis requires: (1) clinical signs or symptoms of heart failure (e.g. shortness of breath, lower extremity edema, or elevated jugular venous pressure); (2) at least one objective marker of cardiopulmonary congestion (e.g. elevated BNP or NT-proBNP, pulmonary edema on chest radiograph, or elevated filling pressures on cardiac catheterization); and (3) the demonstration of preserved LVEF by echocardiography or CMR.^2^ We operationalized the definition of HFpEF based on the objective data points available within the UK Biobank. The HFpEF subcohort was defined as the individuals with an LVEF > 50% and an NT-proBNP > the sex-specific 85th percentile. The 85th percentile was selected as the NT-proBNP elevation threshold after sensitivity analysis at the 90th percentile yielded a subcohort with insufficient events-per-variable ratio for multivariable Cox modeling. This yielded 690 HFpEF participants. This pragmatic HFpEF definition relies on objective biomarker and imaging criteria rather than symptom assessment, an approach necessitated by the absence of contemporaneously documented clinical symptom data in the UK Biobank imaging cohort. We therefore interpret findings as reflective of HFpEF-related pathophysiology rather than clinically adjudicated HFpEF. The healthy reference subcohort was defined as the individuals with an LVEF > 50% and an NT-proBNP < the sex-specific 50th percentile. Given that lower NT-proBNP levels have established utility for excluding heart failure,^35^ participants below the sex-specific median NT-proBNP in the healthy-volunteer-biased UK Biobank imaging cohort^25,36^ represent those least likely to have subclinical cardiopulmonary congestion. This yielded 2,483 healthy reference participants. Sex-specific NT-proBNP thresholds were utilized because reference values have been shown to differ significantly between men and women.^37^

### Cohort Characterization and Between-Group Comparisons

Baseline cohort and subcohort characteristics were summarized using median and IQR for continuous variables and frequencies with percentages for categorical variables. Between-group comparisons (HFpEF vs. healthy reference) for baseline characteristics were performed using Mann-Whitney U tests for continuous variables and chi-square tests for categorical variables (or Fisher’s exact test when expected cell counts were fewer than five), with effect sizes reported as r (= Z/√N) for significant differences.

For optical flow velocity phenotype comparisons between subcohorts, Mann-Whitney U tests were applied to age-and sex-adjusted residuals across all 32 phenotypes, with cluster-adjusted Benjamini-Hochberg false discovery rate correction (M_eff = 28.7; see Multiple Testing Correction) and a significance threshold of q < 0.05. As a sensitivity analysis, ANCOVA was performed on unadjusted (raw) phenotype values with group (HFpEF vs. healthy), age, and sex as covariates to confirm concordance with the residual-based approach.

### Correlation Analyses

Pearson correlation coefficients and coefficients of determination (R²) were computed between all pairwise combinations of the 32 age-and sex-adjusted optical flow velocity phenotypes and 10 age-and sex-adjusted conventional cardiac parameters (320 total comparisons), with significance determined after cluster-adjusted Bonferroni correction (α = 0.05 / (M_eff_OF × M_eff_conventional) = 2.06×10⁻⁴; see Multiple Testing Correction). A minimum of 10 overlapping observations was required per correlation pair.

### Survival Analyses

To assess the prognostic value of optical flow velocity phenotypes relative to conventional cardiac parameters, we performed Cox proportional hazards survival analyses with all-cause mortality as the primary endpoint. Survival analyses used the 10 conventional cardiac parameters described above (see Selection of Ten Conventional Cardiac Parameters) and the 32 optical flow velocity phenotypes, for a total of 42 candidate parameters. Sample sizes varied by parameter availability (**Supplemental Table 1, Supplemental Table 2**).

All parameters were winsorized at the 1st and 99th percentiles prior to analysis to limit the influence of extreme values. Volumes and LV cardiac output were BSA-indexed prior to winsorization. Survival time was defined as days from the Instance 2 assessment centre attendance date to death or last follow-up. All-cause mortality was ascertained from UK Biobank death records. Participants without a recorded death date were censored as alive at the administratively censored date, defined as the maximum observed follow-up date in the dataset. Hazard ratios are reported per 1-SD increase in the winsorized parameter value.

Univariate Cox proportional hazards models were fitted for each of the 42 candidate parameters (10 conventional, 32 optical flow), with age and sex included as covariates, restricted to parameters with at least 100 complete cases. Cluster-adjusted Benjamini-Hochberg false discovery rate (FDR) correction was applied across all univariate tests (M_eff = 38.8; see Multiple Testing Correction; q < 0.05). Cluster-adjusted FDR-significant parameters were advanced to multivariable modeling after correlation pruning (|r| > 0.5, retaining the stronger univariate predictor) and events-per-variable capping at N_events/10.

Three multivariable Cox proportional hazards models were constructed on a common risk set of participants with complete data for all included variables, with age and sex included as covariates in all models: Conventional parameters only, Optical Flow parameters only, and Combined. Model discrimination was assessed using the concordance index (C-index). Incremental prognostic value of optical flow parameters over conventional parameters (and vice versa) was assessed using likelihood ratio tests for nested model comparisons. Bootstrap resampling (1,000 samples) was used to derive confidence intervals for C-index differences, including for the non-nested comparison between Conventional-only and Optical Flow-only models. Proportional hazards assumptions were assessed for all multivariable models. Kaplan-Meier survival curves were constructed for the strongest univariate conventional and optical flow predictors in the full cohort, and for the single FDR-significant optical flow finding in the HFpEF subcohort, comparing the highest vs. lowest quartiles of each parameter; between-group differences were assessed using the log-rank test, presented for descriptive purposes.

In the full cohort, we first performed univariate survival analysis. Among the 21 FDR-significant univariate parameters, we advanced 10 parameters to multivariable modeling: LV cardiac output (BSA-indexed), LV circumferential strain global, LV longitudinal strain global, RV end-diastolic volume (BSA-indexed), LV myocardial early diastolic (E-wave) velocity, RV free wall myocardial early diastolic (E-wave) velocity, LV inner circumferential late diastolic (A-wave) velocity, LV myocardial late diastolic (A-wave) velocity, LV myocardial mid-diastolic (L-wave) velocity, and RV inner radial mid-diastolic (L-wave) velocity.

In the HFpEF subcohort, we again performed univariate survival analysis. Only one parameter was cluster-adjusted FDR-significant on univariate analysis, precluding multivariable modeling.

### Proteome-Wide Association Analyses

We performed proteome-wide association analyses in up to 8,116 UK Biobank participants with concurrent proteomic and optical flow velocity data. Age-and sex-adjusted optical flow velocity residuals (winsorized at ±3 SD; see Age and Sex Adjustment) were used as outcomes; 1,463 individual Olink Explore protein levels served as predictors in simple linear regression models. We tested 1,463 proteins against all 32 optical flow phenotypes, requiring a minimum of 10 complete observations per protein–phenotype pair. Cluster-adjusted Bonferroni correction was applied using the effective number of independent tests for the 32 optical flow phenotype panel (M_eff = 28.7; see Multiple Testing Correction), yielding a significance threshold of p < 0.05 / (1,463 × 28.666) = 1.19×10⁻⁶.

An equivalent proteome-wide association analysis was performed in up to 4,511 UK Biobank participants with concurrent proteomic and conventional cardiac parameter data. Age-and sex-adjusted conventional cardiac parameter residuals (winsorized at ±3 SD) were used as outcomes; 1,463 Olink Explore protein levels served as predictors in identical simple linear regression models. The effective number of independent tests for the 10 conventional cardiac parameter panel was M_eff = 8.5 (see Multiple Testing Correction), yielding a cluster-adjusted Bonferroni threshold of p < 0.05 / (1,463 × 8.458) = 4.04×10⁻⁶.

Proteins significantly associated with optical flow velocity phenotypes were compared against those significantly associated with conventional cardiac parameters to identify overlapping and unique proteomic signatures. Proteins uniquely associated with optical flow velocity phenotypes after comparison against conventional cardiac parameter associations were submitted to STRING-DB (version 12.0) for protein–protein interaction network analysis using a minimum interaction confidence score of 0.4 (medium confidence). The network was assessed for enrichment of protein–protein interactions relative to a random set of proteins of equivalent size drawn from the human proteome (PPI enrichment test).

### Genome-Wide Association Studies Phenotype Preparation

Prior to genetic association testing, all 32 optical flow velocity phenotypes were winsorized at ±3 SD to limit the influence of extreme values. Residuals were computed by regressing each winsorized phenotype on sex, year of birth, genotyping batch, and the first five genetic principal components (PC1-PC5). Residuals were then rank-based inverse normal transformed using qnorm((rank − 0.5)/n) to ensure normality for association testing.

### SNP Heritability and Genetic Correlation

SNP-based heritability (h²) for all 32 optical flow phenotypes was estimated using linkage disequilibrium score regression (LDSC) with 1000 Genomes Phase 3 European LD scores, restricted to HapMap3 SNPs, with the LD score regression intercept freely estimated.^12^ Heritability significance for individual phenotypes was determined using standard Bonferroni correction across the 32 phenotypes (p < 1.56×10⁻³; see Multiple Testing Correction). Pairwise genetic correlations (r_g) were computed for all 496 phenotype pairs using bivariate LDSC with the same reference panel.

### Genome-Wide Association Studies

GWAS was performed for all 32 optical flow velocity phenotypes using PLINK2 on imputed genotype data from up to 73,978 UK Biobank participants of European ancestry (depending on phenotype availability), excluding first-and second-degree relatives and individuals with genotype missingness > 10%. Variants were filtered for minor allele frequency > 0.01, minor allele count > 20, genotype missingness < 0.1, and Hardy-Weinberg equilibrium P > 1×10⁻¹⁵, retaining approximately 7 million variants per phenotype. Pre-adjusted, inverse normal-transformed phenotypes were supplied directly to PLINK2 with no additional covariates in the association model (--glm hide-covar). Genomic inflation was assessed using the median-based lambda (λ = observed median χ² / 0.4549364) across all autosomal variants.

To account for correlation among the 32 optical flow velocity phenotypes, hierarchical clustering was performed on the phenotype correlation matrix using average linkage and a distance threshold of 0.5 (distance = 1 − |r|), yielding 13 clusters. The cluster-corrected Bonferroni significance threshold was set at p < 5×10⁻⁸ / 13 = 3.85×10⁻⁹. Independent loci were defined by merging genome-wide significant SNPs within 1 Mb of one another into a single locus, with the lead SNP defined as the most significant variant within each locus. Benjamini-Hochberg FDR q-values were computed across all tests to validate the cluster-corrected threshold. Gene mapping and functional annotation were performed using FUMA on the full-cohort GWAS results, with lead SNPs identified at p < 5×10⁻⁸ and defined as independent using a secondary r² threshold of 0.1; candidate SNPs within each locus were defined at r² ≥ 0.6 with a lead SNP and p < 0.05. LD was calculated using the 1000 Genomes Phase 3 European reference panel, and LD blocks within 250 kb were merged into a single locus. Minimum MAF was set at 0.01 and per-SNP sample size was used. GWAS summary statistics were lifted from hg19 to hg38 using UCSC liftOver prior to Mendelian randomization. All locus discovery and functional annotation were performed using the full-cohort GWAS.

### Two-Sample Mendelian Randomization

To test for causal effects of circulating proteins on optical flow velocity phenotypes, we performed two-sample Mendelian randomization using cis-protein quantitative trait loci (cis-pQTLs) from the UK Biobank Pharma Proteomics Project (UKB-PPP) as genetic instruments. Genetic variants located within 100 kb of the protein-coding gene were used as cis-pQTL instruments. To ensure instrument independence, linkage disequilibrium clumping was performed retaining only variants with r² < 0.01, using index variants with p < 5×10⁻⁵ as independent lead signals. Data formatting and harmonization were performed using the TwoSampleMR package (version 0.6.14); inverse-variance weighted (IVW) MR was performed using fixed-effects models in the MendelianRandomization package (version 0.10.0), requiring a minimum of three harmonized SNPs per instrument.

To satisfy the independence assumption of two-sample MR, we identified 9,480 participants who contributed to both the Instance 2 CMR dataset and the UKB-PPP proteomics dataset. Of these, 8,544 were present in the European-ancestry, unrelated, quality-control-passing GWAS analysis set and were removed; the remainder had already been excluded by ancestry and relatedness filtering. Re-running the outcome GWAS in the non-overlapping participants yielded 65,162–65,496 participants per phenotype (varying by phenotype availability), used exclusively for MR and colocalization analyses. After deduplication retaining the instrument with the greatest number of SNPs per unique protein–phenotype pair, Bonferroni correction was applied across 26,886 unique pairs spanning 1,651 proteins and 27 of 32 optical flow phenotypes (α = 0.05/26,886 = 1.86×10⁻⁶); five phenotypes (the three L-wave LV inner cavity velocities [total, radial, and circumferential], L-wave LV myocardial velocity, and L-wave RV myocardial free wall velocity) yielded no protein–phenotype pairs with sufficient SNP overlap after harmonization and were excluded from MR analysis.

Bonferroni-significant associations were carried forward to MR-PRESSO (1,000 permutations), which tested for global horizontal pleiotropy, identified outlier instruments, and provided outlier-corrected causal estimates. A minimum of four harmonized SNPs was required for MR-PRESSO analysis. Associations were retained if the global test was non-significant (p ≥ 0.05) or, where significant pleiotropy was detected, if the outlier-corrected estimate remained significant (p < 0.05). Sensitivity analyses using weighted median and MR-Egger methods were performed on all MR-PRESSO-passing associations and are reported in **Supplemental Table 11**; directional concordance with IVW estimates is noted. Given the limited number of cis-pQTL instruments available for most proteins, weighted median and MR-Egger methods reduced statistical power relative to IVW and were therefore used for directional concordance assessment only rather than independent filtering. Colocalization between cis-pQTL and optical flow GWAS signals was performed using coloc; a minimum of 15 shared SNPs was required at each locus.

### Colocalization

For protein–phenotype pairs passing MR-PRESSO, colocalization between the cis-pQTL signal and the optical flow GWAS signal was assessed using the coloc.abf Bayesian colocalization framework, requiring a minimum of 15 shared SNPs. The split-sample GWAS was used as the outcome dataset to maintain sample independence with the MR analysis. A posterior probability of shared causal variant (PP.H4) > 0.8 was considered strong evidence for colocalization.

To triangulate biological mechanism, eQTL colocalization was additionally performed for each MR protein hit against GTEx v10 summary statistics in four cardiac-relevant tissues: Heart Left Ventricle (N≈899), Heart Atrial Appendage (N≈917), Artery Aorta (N≈943), and Artery Coronary (N≈535), using identical coloc.abf parameters (minimum 15 shared SNPs, PP.H4 > 0.8). A uniform approximate sample size of 700 was used in the coloc.abf prior variance calculation; as this falls within the range of true tissue-specific sample sizes, the effect on posterior probability estimates is expected to be minimal.

### Protein-Protein Interaction Network Analysis

To assess whether MR-significant proteins converge on shared biological pathways, the 38 proteins with MR-PRESSO-validated causal associations were submitted to STRING-DB (version 12.0) for protein-protein interaction network and functional enrichment analysis using a minimum interaction confidence score of 0.4 (medium confidence). One protein (HCG22) was excluded as a non-coding RNA gene, leaving 37 proteins for network analysis. The network was assessed for enrichment of protein-protein interactions relative to a random set of proteins of equivalent size drawn from the human proteome (PPI enrichment test).

## SUPPLEMENTAL FIGURES

## SUPPLEMENTAL TABLES

**Supplemental Table 1.** Full conventional cardiac parameter list.

**Supplemental Table 2.** Descriptive statistics and data completeness for 32 optical flow velocity phenotypes.

**Supplemental Table 3.** Full cohort univariate survival analysis: FDR-significant associations with all-cause mortality.

**Supplemental Table 4.** Proteome-wide association analysis of circulating proteins with optical flow velocity phenotypes.

**Supplemental Table 5.** Proteome-wide association analysis of circulating proteins with conventional cardiac parameters.

**Supplemental Table 6.** Proteins significantly associated with both optical flow velocity phenotypes and conventional cardiac parameters.

**Supplemental Table 7.** SNP-based heritability of optical flow velocity phenotypes.

**Supplemental Table 8.** Pairwise genetic correlation matrix for optical flow-derived cardiac velocity phenotypes.

**Supplemental Table 9.** GWAS summary for all 32 optical flow phenotypes.

**Supplemental Table 10.** Previously reported GWAS Catalog associations at genomic risk loci identified in the optical flow velocity phenotype GWAS.

**Supplemental Table 11.** Mendelian randomization sensitivity analyses for 81 protein-phenotype associations.

**Supplemental Table 12.** eQTL colocalization of MR-significant proteins with optical flow GWAS signals in cardiovascular tissues.

**Supplemental Table 13 (A-E).** STRING-DB protein-protein interaction and functional enrichment analysis of MR-significant proteins.

## SUPPLEMENTAL DATA

### Supplemental Data 1: Systolic and diastolic cardiac cycle events

We use uppercase S-/E-/L-/A-wave nomenclature to denote phases of the cardiac cycle and apply this notation to both inner cavity and myocardial velocity phenotypes. We adapted the S-, E-, L-, and A-wave nomenclature from echocardiography-based tissue and pulsed wave Doppler metrics of systolic and diastolic function^29,30^. The S-wave corresponds to systole, the contractile phase of the cardiac cycle. Diastole comprises a complex sequence of distinct ventricular filling events^30^. The E-wave corresponds to early diastole, representing active ventricular relaxation immediately after mitral valve opening. The A-wave corresponds to late diastole, reflecting the contribution of atrial contraction just before mitral valve closure. The L-wave corresponds to mid-diastole, a phase influenced by complex atrioventricular interactions that remain incompletely understood, and one not previously quantified or genetically interrogated at population scale^30,38^.

### Supplemental Data 2: HFpEF subcohort participants vs. healthy subcohort participants

HFpEF participants were older than healthy reference participants (median age 68.0 vs. 61.0 years, p < 0.001), while the sex distribution was similar between groups (HFpEF 53.5% female vs. healthy reference 53.8% female, p = 0.913). Over median follow-up of 6.3 years [IQR 5.2-7.8 years] in the HFpEF subcohort and 6.0 years [IQR 5.3-7.5 years] in the healthy reference subcohort, HFpEF participants experienced significantly higher rates of adverse cardiovascular events compared to healthy reference participants, including myocardial infarction (7.2% vs. 1.9%, p < 0.001), NSTEMI (3.9% vs. 0.8%, p < 0.001), STEMI (2.0% vs. 0.8%, p = 0.007), and stroke (3.2% vs. 1.6%, p = 0.010).

### Supplemental Data 3: Full cohort multivariable survival analysis using Cox proportional hazards

We restricted multivariable model analyses to a common cohort with complete data for all included variables (n = 37,043; 1,050 deaths; death rate 2.8%). Three multivariable models were constructed, with age and sex included as covariates: Conventional parameters only, Optical Flow parameters only, and Combined parameters. The Conventional-only and Optical Flow-only models achieved similar discrimination (concordance index [C-index] 0.729 vs. 0.726; bootstrap ΔC = −0.003, 95% CI −0.008 to 0.002). The Combined model achieved C-index 0.732, modestly exceeding both the Conventional-only model (bootstrap ΔC = 0.003, 95% CI 0.001 to 0.006) and the Optical Flow-only model (bootstrap ΔC = 0.006, 95% CI 0.002 to 0.010). The addition of optical flow parameters to conventional parameters provided statistically significant incremental value (likelihood ratio test χ² = 13.1, df = 6, p = 0.042); the addition of conventional parameters to optical flow parameters provided statistically significant incremental value (χ² = 51.6, df = 4, p = 1.65×10⁻¹⁰).

In the full cohort, the C-indices achieved by the Conventional-only, Optical flow-only, and Combined models (0.729, 0.726, and 0.732, respectively) were consistent with or exceeded prior CMR-based survival analyses in the UK Biobank, including models incorporating novel imaging-derived phenotypes that achieved C-indices of 0.547-0.614 in a similarly low-risk population with healthy-volunteer bias.^25,36^ Nested model comparisons demonstrated that optical flow velocity metrics and conventional cardiac parameters provide statistically significant bidirectional incremental value, confirming that optical flow velocity phenotypes and conventional cardiac parameters capture complementary rather than redundant prognostic information.

### Supplemental Data 4: STRING-DB analysis of the optical-flow-unique protein associations

When we compared the proteins significantly associated with optical flow velocity phenotypes with the proteins significantly associated with conventional cardiac parameters, we observed broad overlap (**Supplemental Table 6**) but also a distinct optical-flow-specific proteomic signature concentrated in diastolic velocity phenotypes. The 12 proteins uniquely associated with optical flow parameters were exclusively associated with mid-diastolic (L-wave) and late diastolic (A-wave) velocity phenotypes. STRING-DB network analysis of these 12 proteins revealed significantly more interactions than expected by chance (4 observed vs. 1 expected edge, PPI enrichment p = 0.017), with the network organizing primarily into a subcluster anchored by a DCN (decorin)-ERBB3 (Erb-B2 Receptor Tyrosine Kinase 3) interaction hub. DCN plays a role in collagen fibril assembly,^39^ while ERBB3 is a key component of growth factor receptor signaling.^40^ Notably, the most significant optical-flow-unique association overall was with ACE2 (angiotensin-converting enzyme 2), an important counter-regulatory enzyme in the renin-angiotensin-aldosterone system and a key contributor to the regulation of cardiovascular and renal function^13^ (r = 0.068, p = 9.18×10⁻¹⁰). Functional enrichment analysis identified signaling receptor binding, receptor ligand activity, and extracellular region localization as the dominant biological themes. The protein set also showed significant overlap with a published plasma biomarker profile of HFpEF^41^ (4 of 12 proteins, FDR = 0.027).

### Supplemental Data 5: eQTL colocalization from GTEx v10 across four bulk cardiovascular tissues

To further characterize the regulatory context of the MR-significant associations, we performed eQTL colocalization between optical flow GWAS signals and gene expression data from GTEx v10 across four bulk cardiovascular tissues (Heart Atrial Appendage, Heart Left Ventricle, Artery Aorta, Artery Coronary)^42^. Of the 38 proteins with MR-PRESSO-validated associations, 25 were testable for eQTL colocalization in at least one bulk cardiovascular tissue (9 lacked eQTL data in GTEx cardiovascular tissues; 4 had insufficient shared SNPs for colocalization testing). WWP2 showed strong eQTL colocalization in coronary artery tissue (PP.H4 = 0.893 for late diastolic [A-wave] LV inner velocity, PP.H4 = 0.878 for late diastolic [A-wave] LV inner radial velocity), suggesting a transcriptional mechanism at this locus. Neither of the two proteins with the strongest cis-pQTL colocalization (namely, RABGAP1L and C2) showed evidence of strong eQTL colocalization in the context of bulk cardiovascular tissue (**Supplemental Table 12**).

### Supplemental Data 6: STRING-DB analysis of the MR-PRESSO-validated causal associations

To assess whether MR-significant proteins converge on shared biological pathways, we performed protein-protein interaction network and functional enrichment analysis using STRING-DB on the 38 proteins with MR-PRESSO-validated causal associations. Of these, 37 proteins mapped to STRING (HCG22 was excluded as a non-coding RNA gene), revealing significantly more interactions than expected by chance (19 observed vs. 7 expected edges, PPI enrichment p = 1.18×10⁻⁴). Gene Ontology cellular component analysis identified RABGAP1L within both the vesicle and cytoplasmic vesicle compartments, consistent with its established role in endosomal trafficking. RABGAP1L is known to interact with Ankyrin-B, a scaffold protein that localizes calcium handling machinery to the cardiomyocyte membrane.^20,21^ Gene Ontology cellular component analysis also identified enrichment for collagen-containing extracellular matrix (7 proteins, FDR = 2.6×10⁻³), supporting extracellular matrix remodeling as a convergent biological theme (**Supplemental Table 13, Supplemental Figure 10**). Collectively, these analyses reveal that proteins causally associated with cardiac motion phenotypes converge on two biological themes: vesicular trafficking and endosomal processing, and extracellular matrix remodeling.

### Supplemental Data 7: AnkB-RABGAP1L interaction data

An AnkB-based macromolecular complex consisting of Na/K-ATPase, Na/Ca exchanger 1, and InsP3 receptor has been identified in cardiomyocyte T-tubules^20^. Mutations in the AnkB gene cause significant alterations in intracellular calcium handling in cardiomyocytes and a spectrum of pathologic cardiac phenotypes, from subclinical electrocardiographic abnormalities to life-threatening arrhythmias.^43,44^ It has also been shown that AnkB recruits RABGAP1L as part of a coordinated sequence of endosomal trafficking and polarized transport of specialized membrane proteins.^21^ While this AnkB-RABGAP1L interaction was initially characterized in fibroblasts, RABGAP1L is highly expressed in adult ventricular and atrial cardiomyocytes^45–47^. Moreover, single-cell RNA-sequencing with spatial transcriptomics of the developing human heart localizes RABGAP1L expression to compact ventricular cardiomyocytes near the epicardium^48^.

---

**Supplemental Figure 1.**
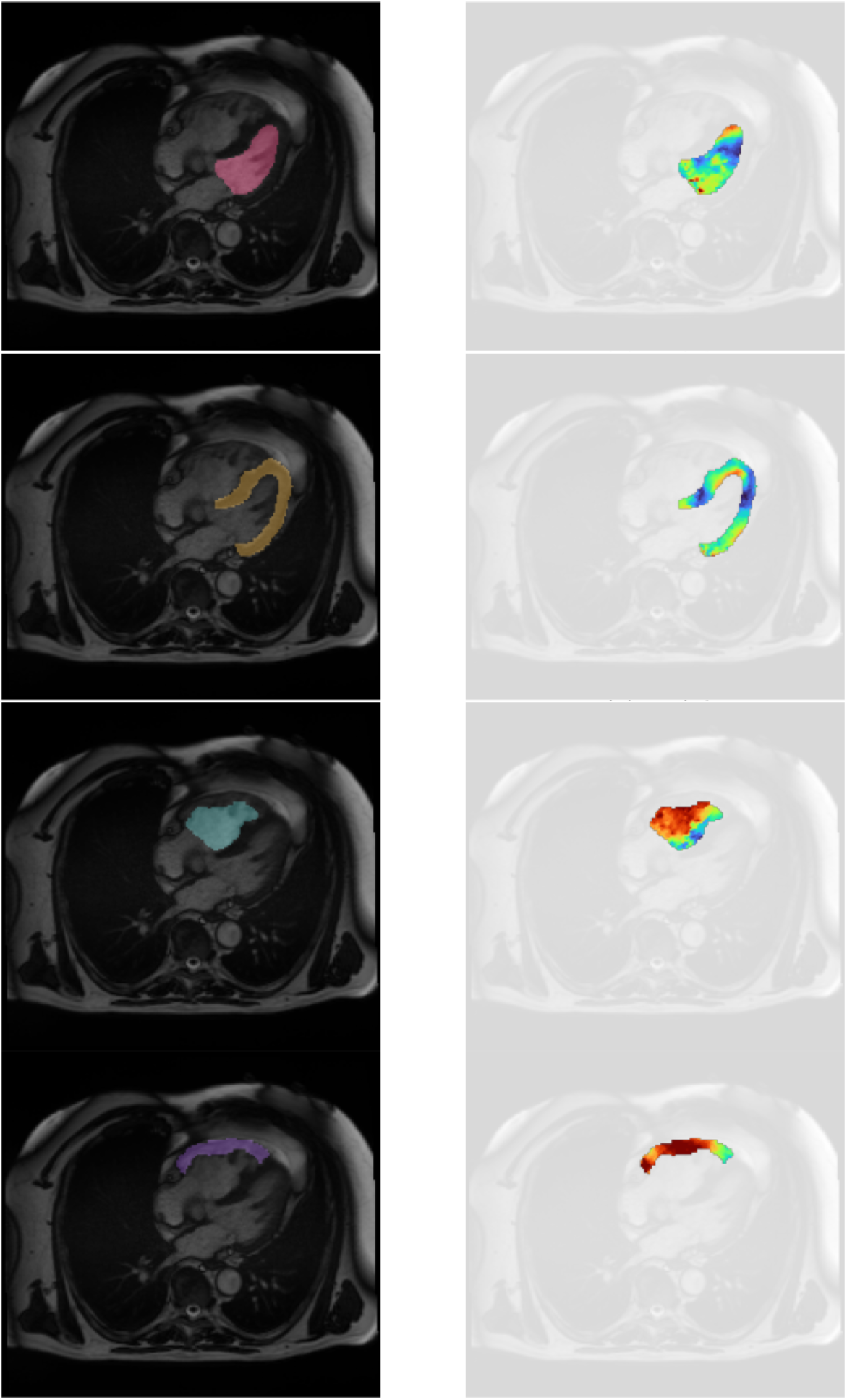
Deep learning segmentation and anatomically masked optical flow velocity fields for four key cardiac structures. Left column: representative four-chamber long-axis standard cine CMR frame with segmentation overlays for the LV inner cavity (pink), LV myocardium (yellow), RV inner cavity (teal), and RV myocardial free wall (purple). Right column: corresponding velocity color maps derived from optical flow, with color indicating velocity vector magnitude. CMR = cardiac magnetic resonance; LV = left ventricular; RV = right ventricular.

**Supplemental Figure 2.**
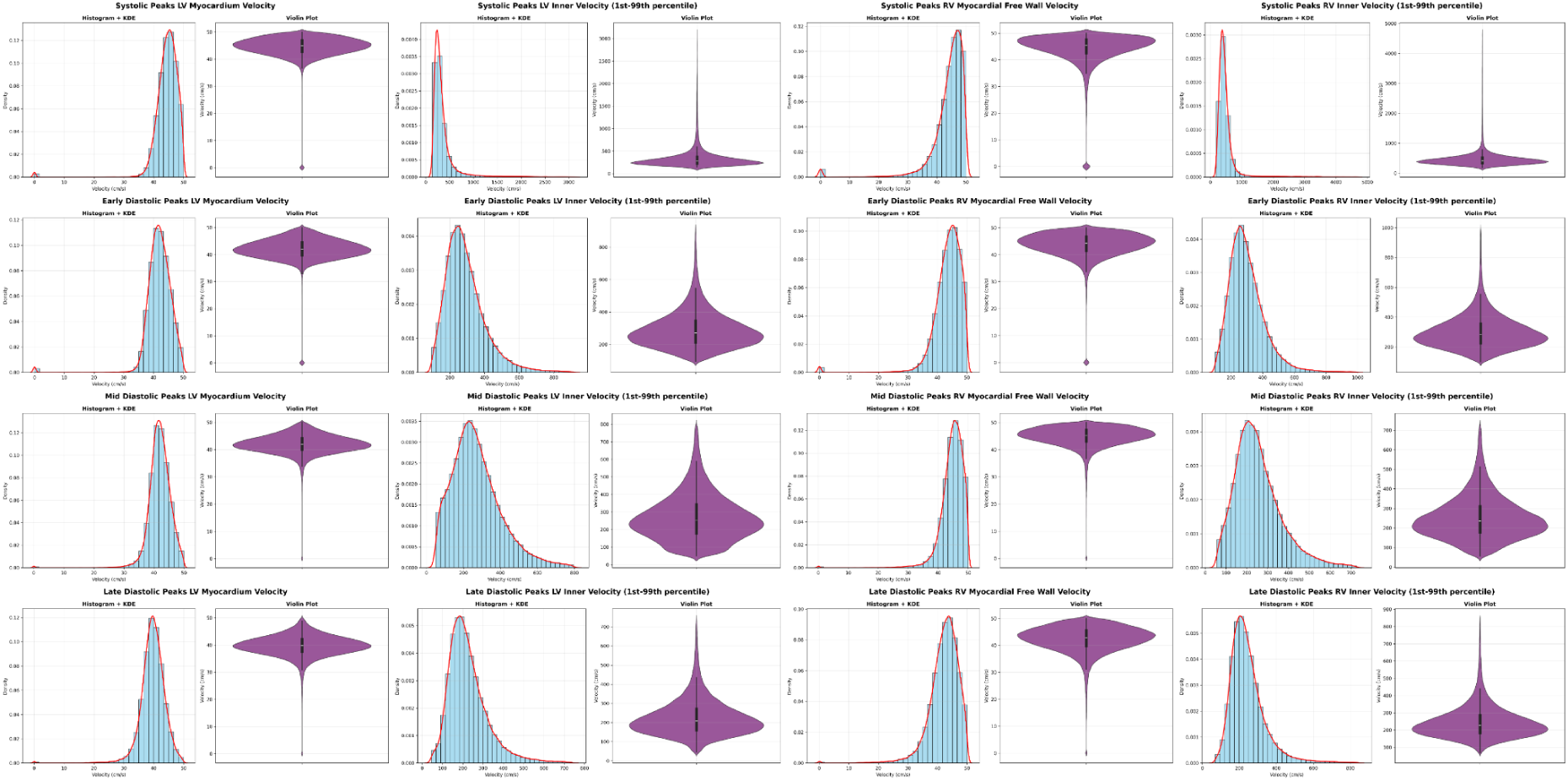
Distribution of optical flow velocity phenotypes. Histograms with kernel density estimates (left) and violin plots (right) for 16 total vector velocity phenotypes across four cardiac structures (LV myocardium, LV inner cavity, RV myocardial free wall, RV inner cavity) and four cardiac cycle phases (systolic, early diastolic, mid-diastolic, late diastolic). For visualization purposes, velocity distributions are truncated to the 1st-99th percentile range to reduce the influence of extreme outliers on axis scaling. Distributions for radial and circumferential component velocities (16 additional phenotypes) not shown. LV = left ventricular; RV = right ventricular.

**Supplemental Figure 3.**
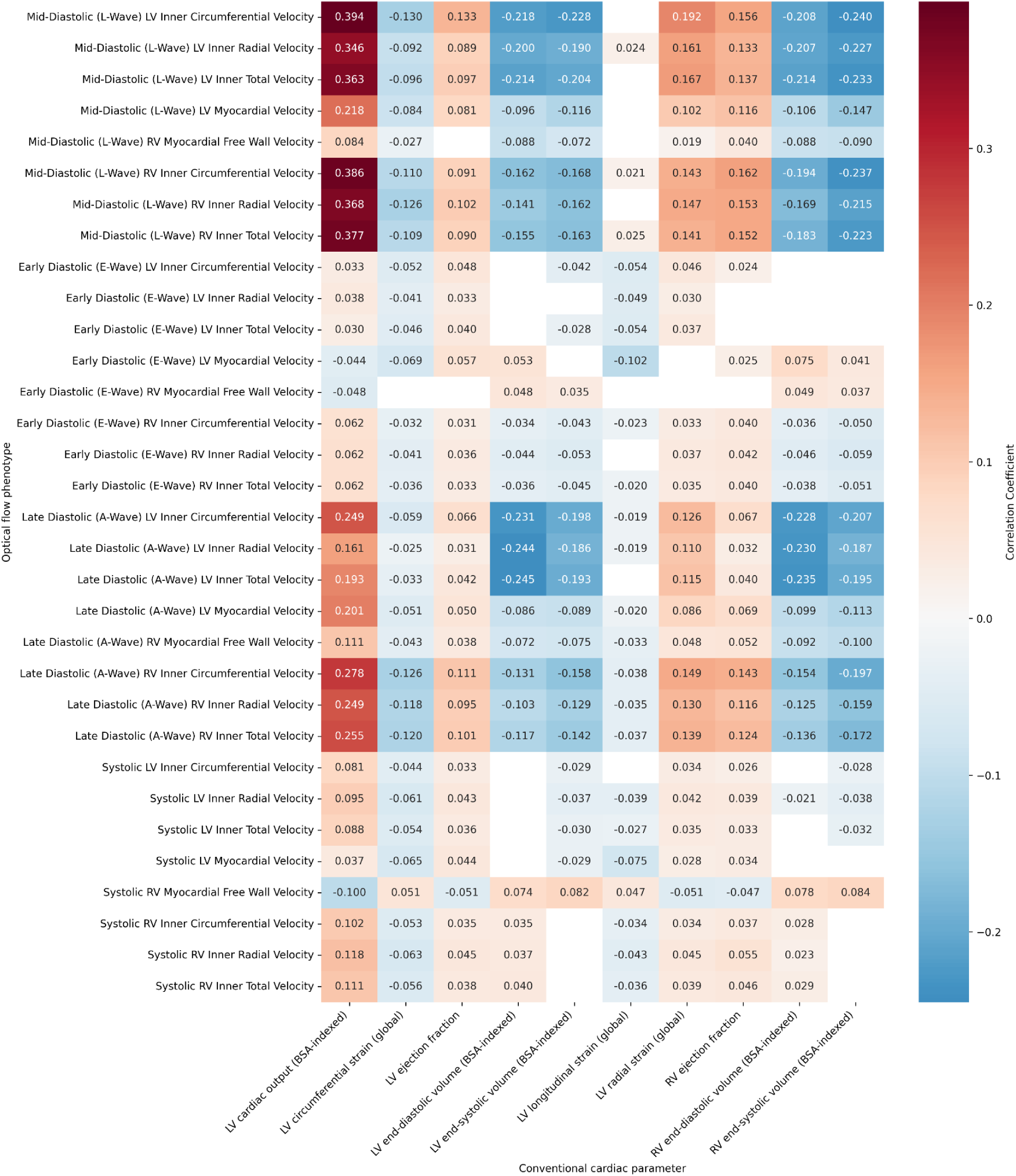
Correlation between optical flow velocity phenotypes and conventional cardiac parameters. Heatmap showing cluster-adjusted Bonferroni-significant Pearson correlation coefficients between the 32 age-and sex-adjusted optical flow phenotypes and 10 age-and sex-adjusted conventional cardiac parameters. Non-significant cells are left blank. The strongest correlations were observed between mid-diastolic (L-wave) and late diastolic (A-wave) velocities and LV cardiac output (positive) and ventricular volumes (negative). BSA = body surface area; LV = left ventricular; RV = right ventricular.

**Supplemental Figure 4.**
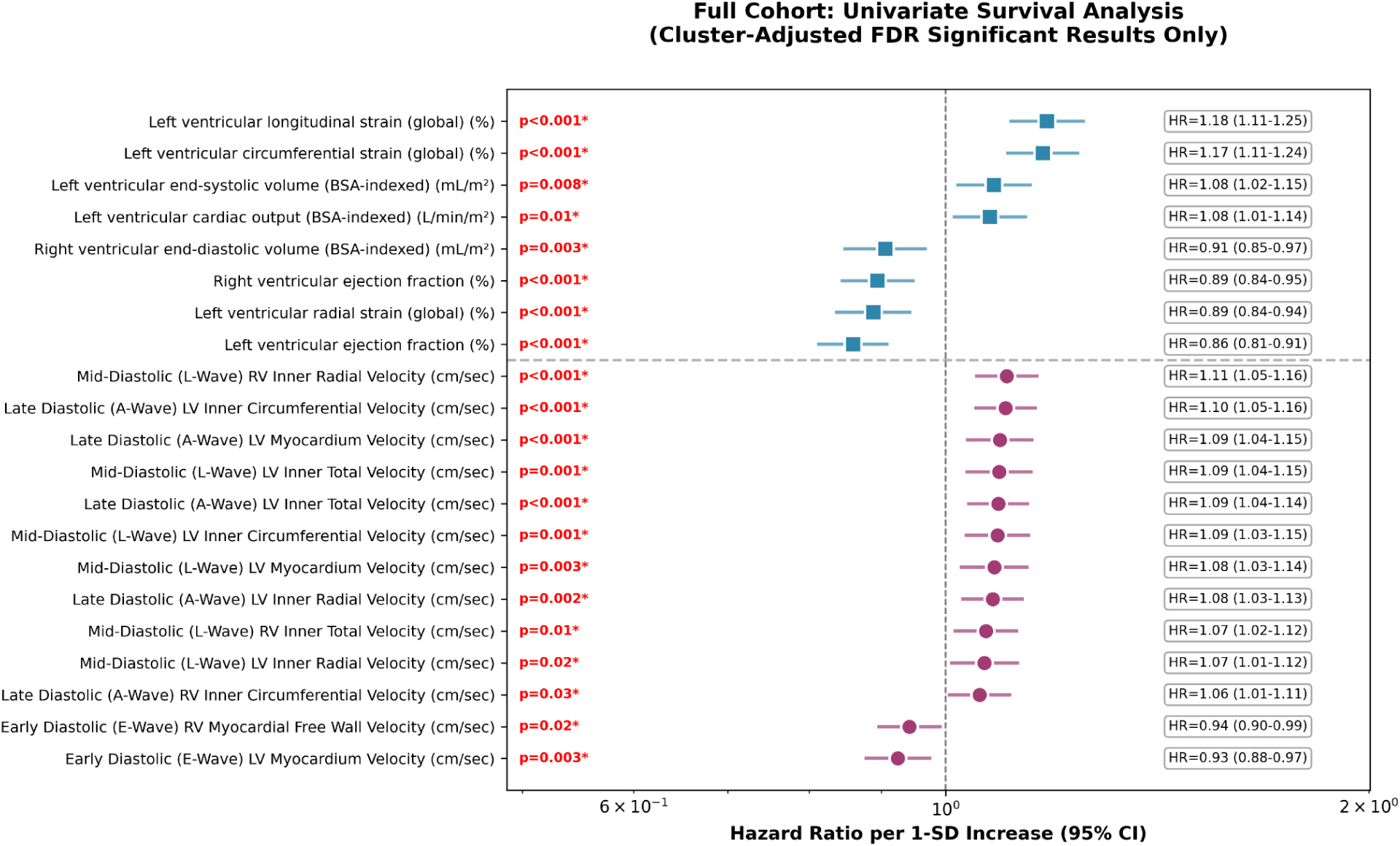
Full cohort univariate survival analysis forest plot: FDR-significant associations with all-cause mortality. Hazard ratios per standard deviation increase (95% confidence intervals) from Cox proportional hazards models adjusted for age and sex. Twenty-one parameters (8 conventional, 13 optical flow) reached significance after cluster-adjusted FDR correction (q < 0.05). Conventional parameters (blue squares) and optical flow parameters (magenta circles) are separated by a dashed line and ordered by hazard ratio within each group. BSA = body surface area; FDR = false discovery rate; LV = left ventricular; RV = right ventricular; SD = standard deviation.

**Supplemental Figure 5.**
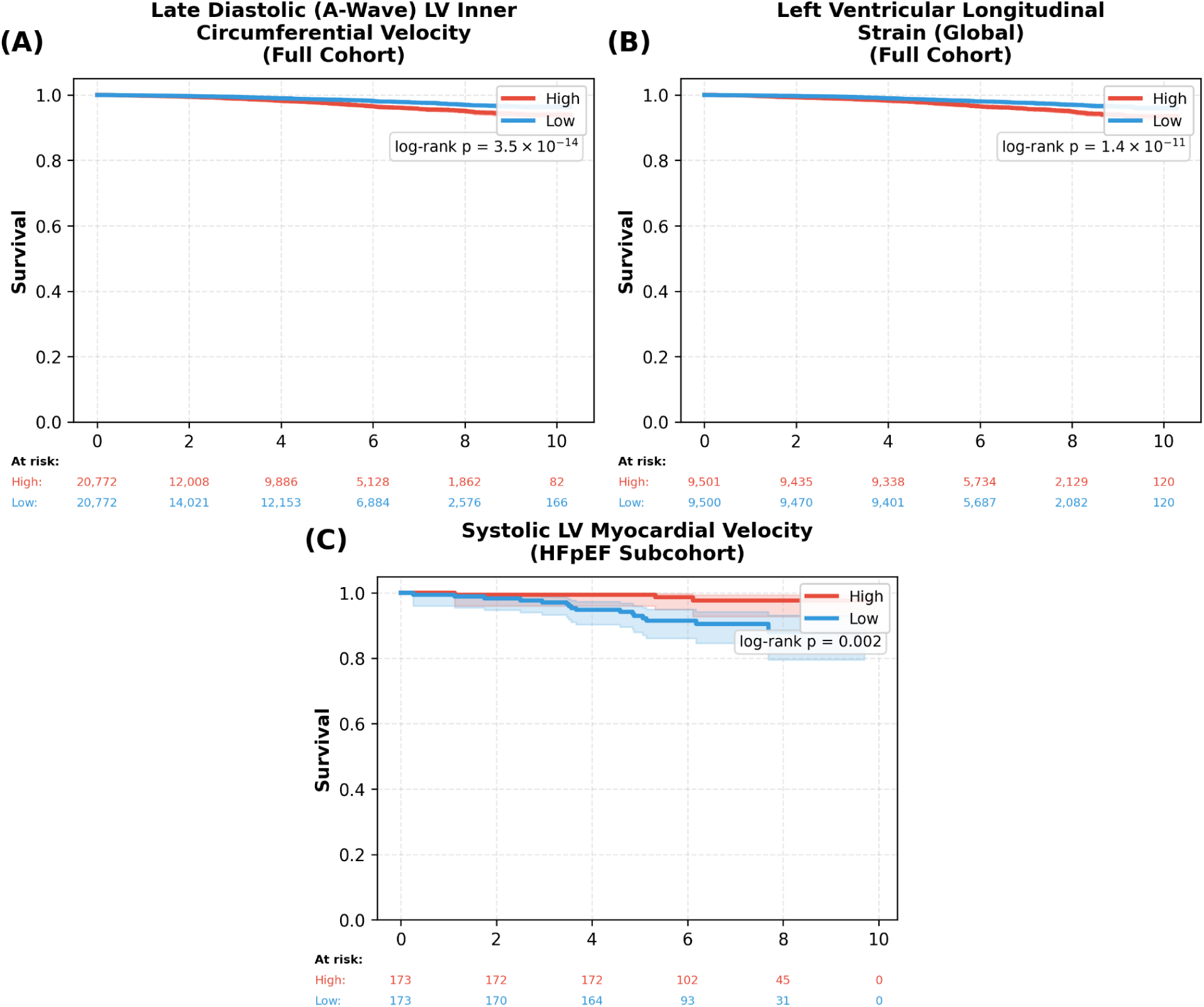
Kaplan-Meier survival curves for the strongest optical flow and conventional cardiac parameter predictors of all-cause mortality. Survival curves compare participants in the highest (Q4, red) vs. lowest (Q1, blue) quartiles of each parameter. Panels A and B show the strongest optical flow and conventional univariate predictors in the full cohort; Panel C shows the single FDR-significant predictor in the HFpEF subcohort. All parameters were identified through univariate Cox regression with age and sex adjustment and cluster-adjusted FDR correction (q < 0.05). Shaded regions indicate 95% confidence intervals. **(A)** Late diastolic (A-Wave) LV inner circumferential velocity in the full cohort (Q4: n = 20,772, 416 deaths; Q1: n = 20,772, 289 deaths; log-rank p = 3.5×10⁻¹⁴); higher velocity was associated with worse survival. **(B)** Left ventricular longitudinal strain (global) in the full cohort (Q4: n = 9,501, 394 deaths; Q1: n = 9,500, 225 deaths; log-rank p = 1.4×10⁻¹¹); worse longitudinal strain (less negative values) was associated with worse survival. **(C)** Systolic LV myocardial velocity in the HFpEF subcohort (Q4: n = 173, 3 deaths; Q1: n = 173, 16 deaths; log-rank p = 0.002); higher velocity was associated with better survival. FDR = false discovery rate; HFpEF = heart failure with preserved ejection fraction; LV = left ventricular; Q1 = first quartile; Q4 = fourth quartile.

**Supplemental Figure 6.**
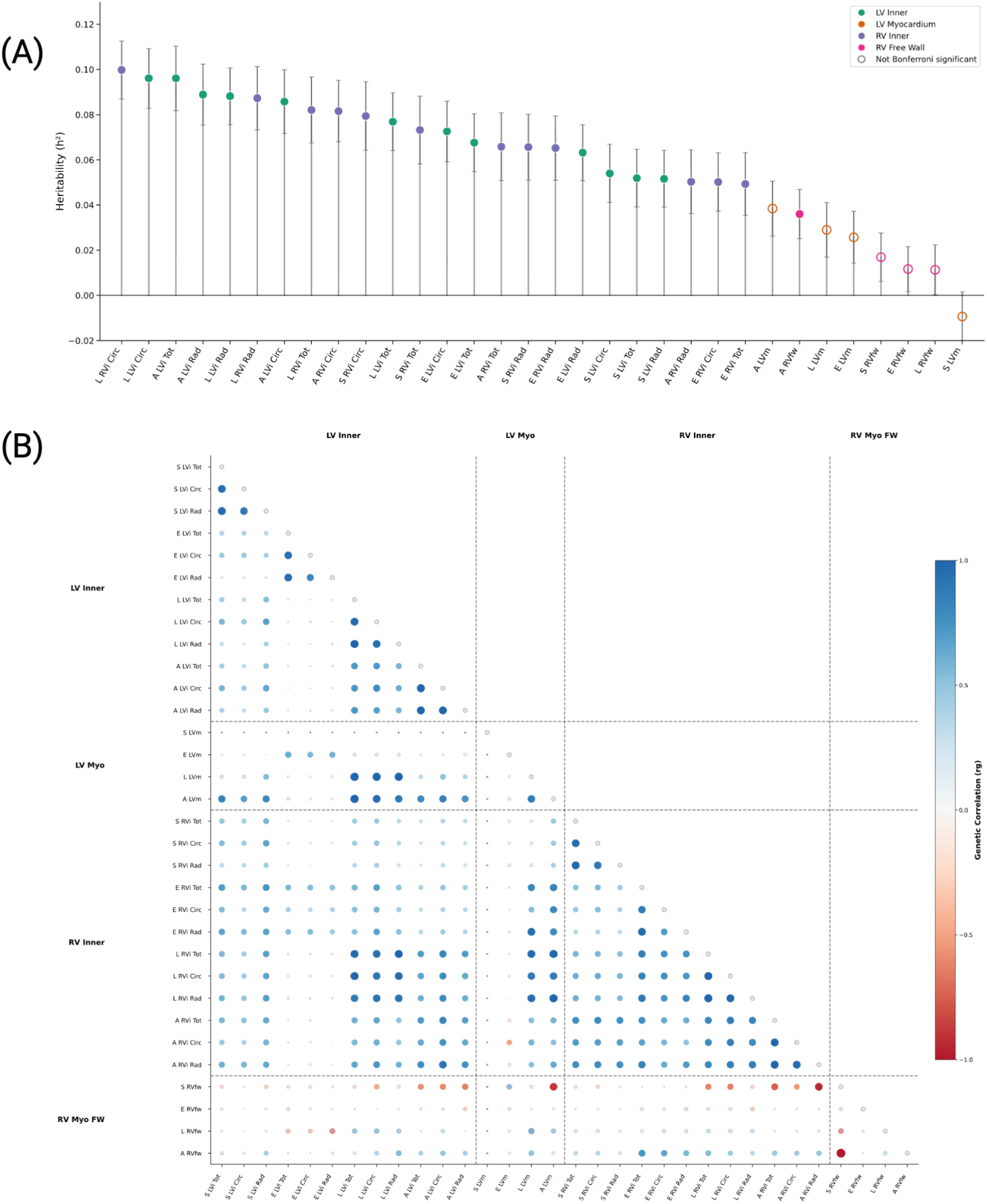
Heritability and genetic correlation structure of optical flow-derived cardiac velocity phenotypes. (A) SNP-based heritability (h²_SNP) estimated by LD score regression for all 32 optical flow-derived velocity phenotypes, ordered by decreasing heritability. Points are colored by cardiac structure: LV inner cavity (green), RV inner cavity (purple), LV myocardium (orange), and RV myocardial free wall (pink). Filled points indicate Bonferroni significance (p < 1.56 × 10⁻³; 0.05/32); open points indicate non-significant estimates. Error bars represent standard errors. (B) Pairwise genetic correlations (rg) estimated by cross-trait LD score regression. Circle size reflects the absolute magnitude of the genetic correlation; color indicates direction and strength (blue = positive, red = negative). Phenotypes are grouped by cardiac structure (LV inner cavity, LV myocardium, RV inner cavity, RV myocardial free wall) and ordered by cardiac phase within each group. Genetic correlations involving systolic LV myocardium velocity could not be estimated due to non-significant heritability (h² ≈ 0). GWAS summary statistics were derived from up to 73,978 European-ancestry UK Biobank participants. Phenotype abbreviations: S = systolic, E = early diastolic (E-wave), L = mid-diastolic (L-wave), A = late diastolic (A-wave); LVi = LV inner cavity, LVm = LV myocardium, RVi = RV inner cavity, RVfw = RV myocardial free wall; Tot = total velocity, Circ = circumferential velocity, Rad = radial velocity.

**Supplemental Figure 7.**
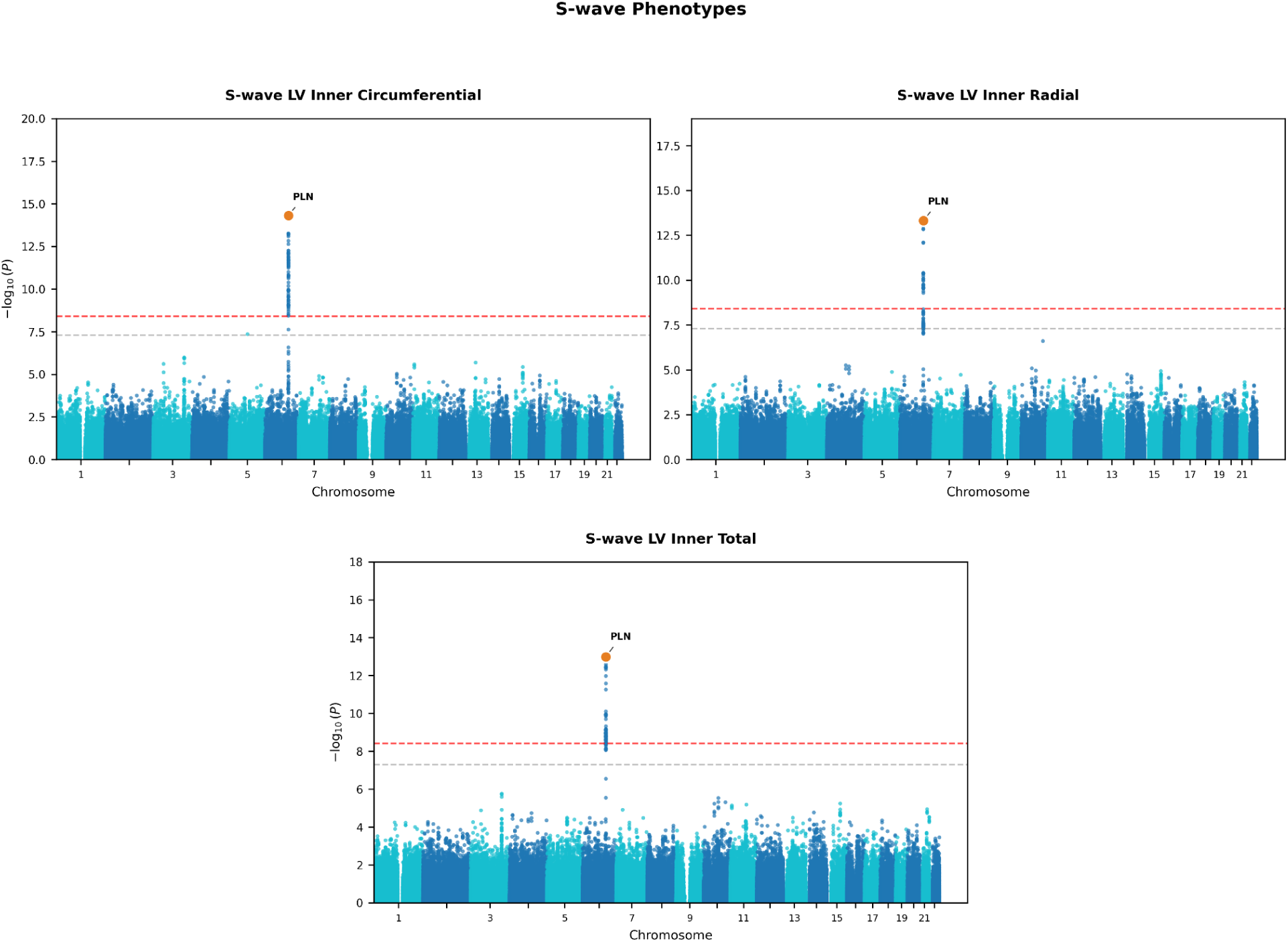
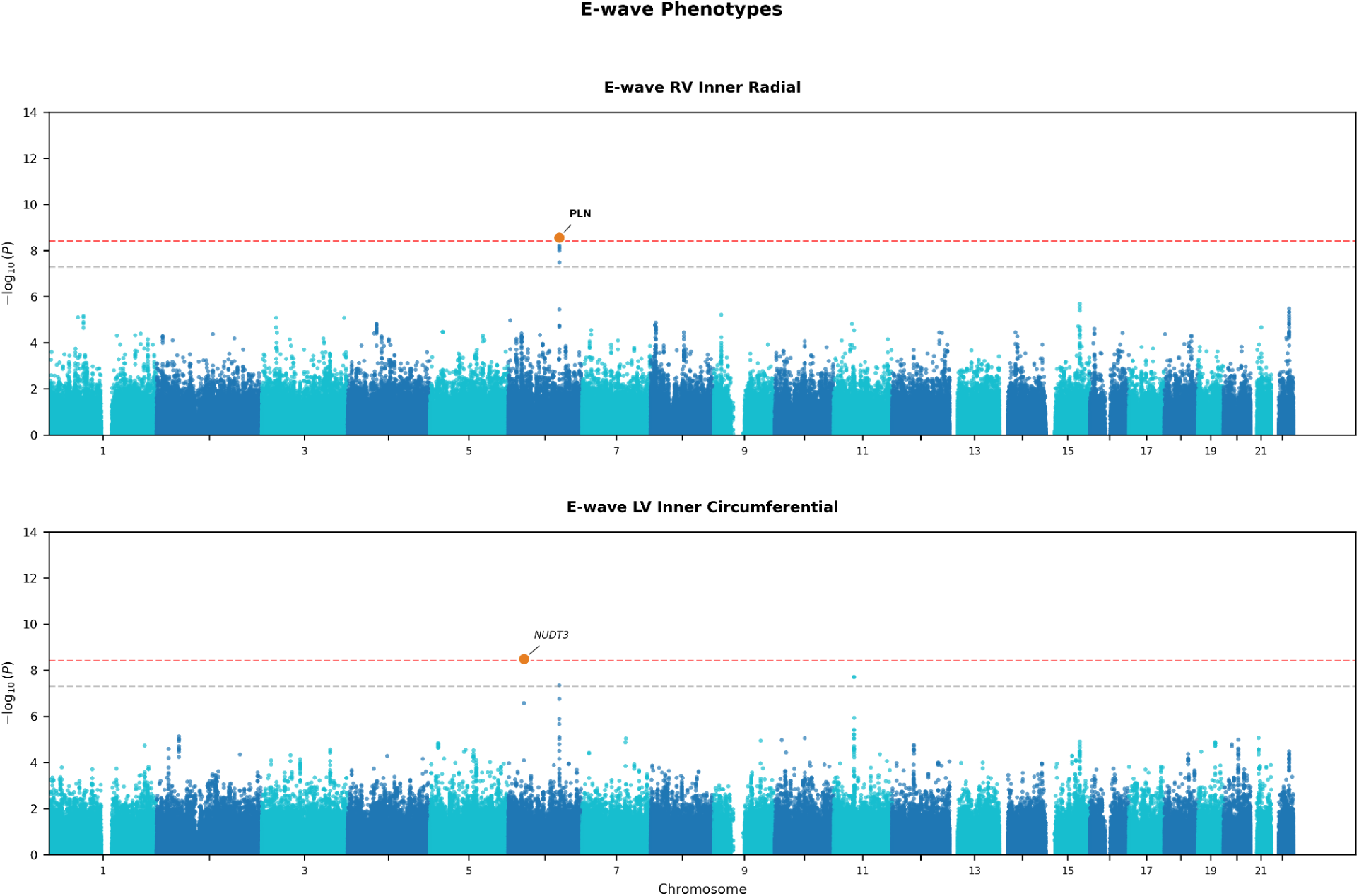
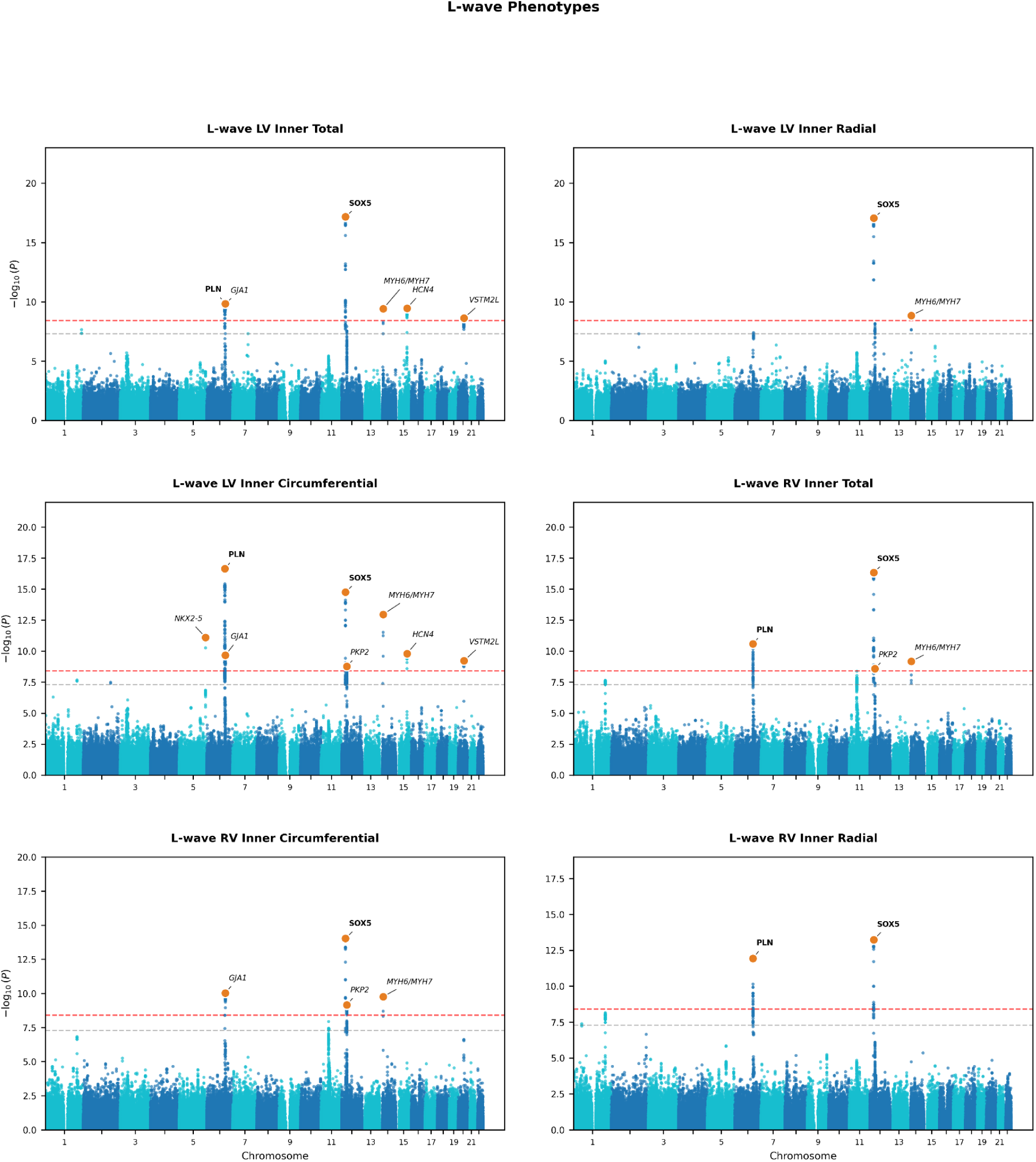
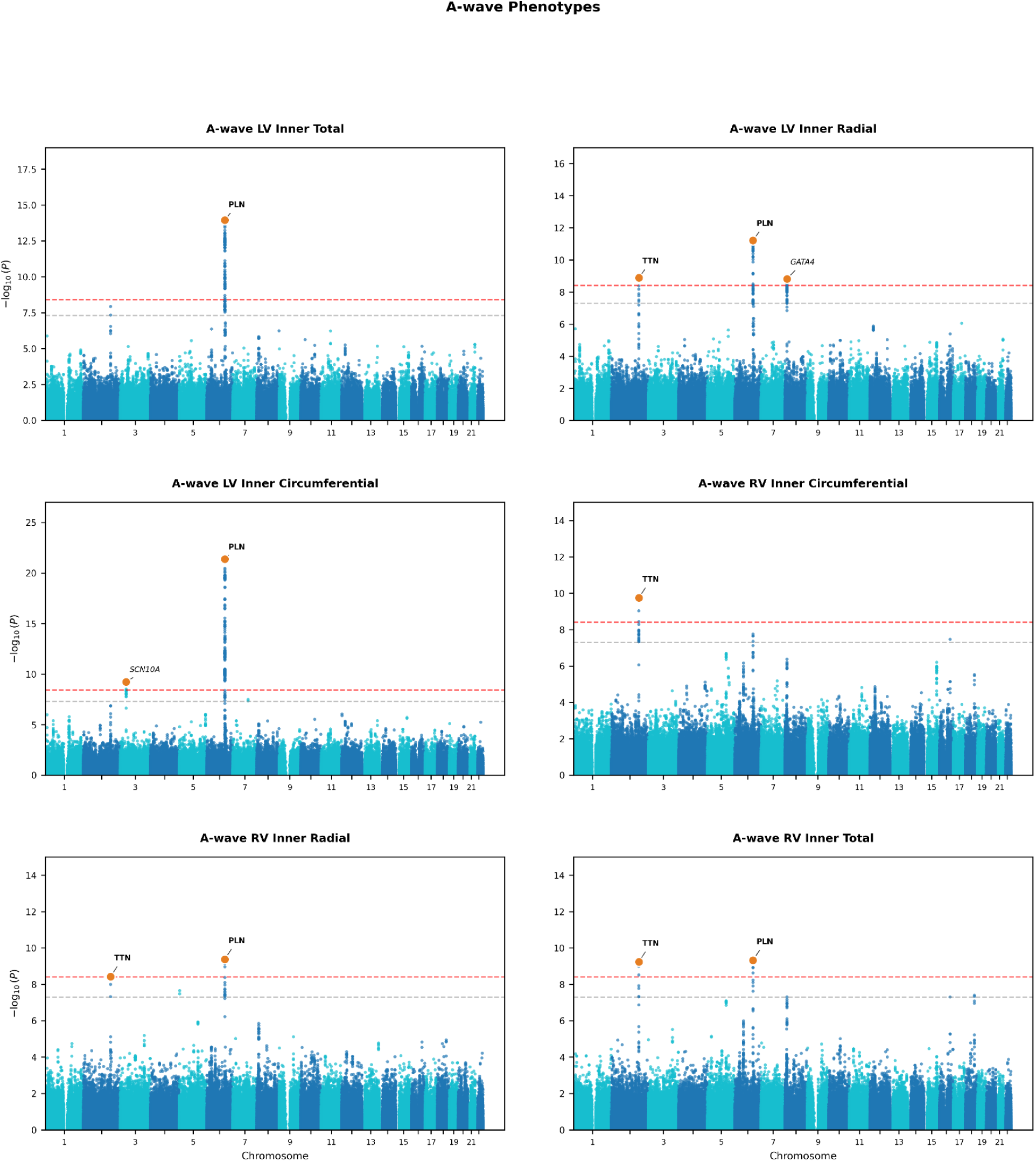
Locus architecture of optical flow velocity phenotypes by cardiac phase. Manhattan plots for the 17 optical flow velocity phenotypes with autosomal hits reaching the cluster-corrected Bonferroni significance threshold (p < 3.85×10⁻⁹), organized by cardiac phase: (A) systolic (S-wave, 3 phenotypes), (B) early diastolic (E-wave, 2 phenotypes), (C) mid-diastolic (L-wave, 6 phenotypes), and (D) late diastolic (A-wave, 6 phenotypes). Red dashed lines indicate the cluster-corrected Bonferroni threshold (p < 3.85×10⁻⁹); gray dashed lines indicate conventional genome-wide significance (p < 5×10⁻⁸). Lead SNPs at the genomic risk loci are shown as orange circles, labeled with the mapped gene. PLN, SOX5, and TTN are highlighted (bold) as the top loci, based on statistical significance and pleiotropy across phenotypes. LV = left ventricular; RV = right ventricular; SNP = single nucleotide polymorphism.

**Supplemental Figure 8.**
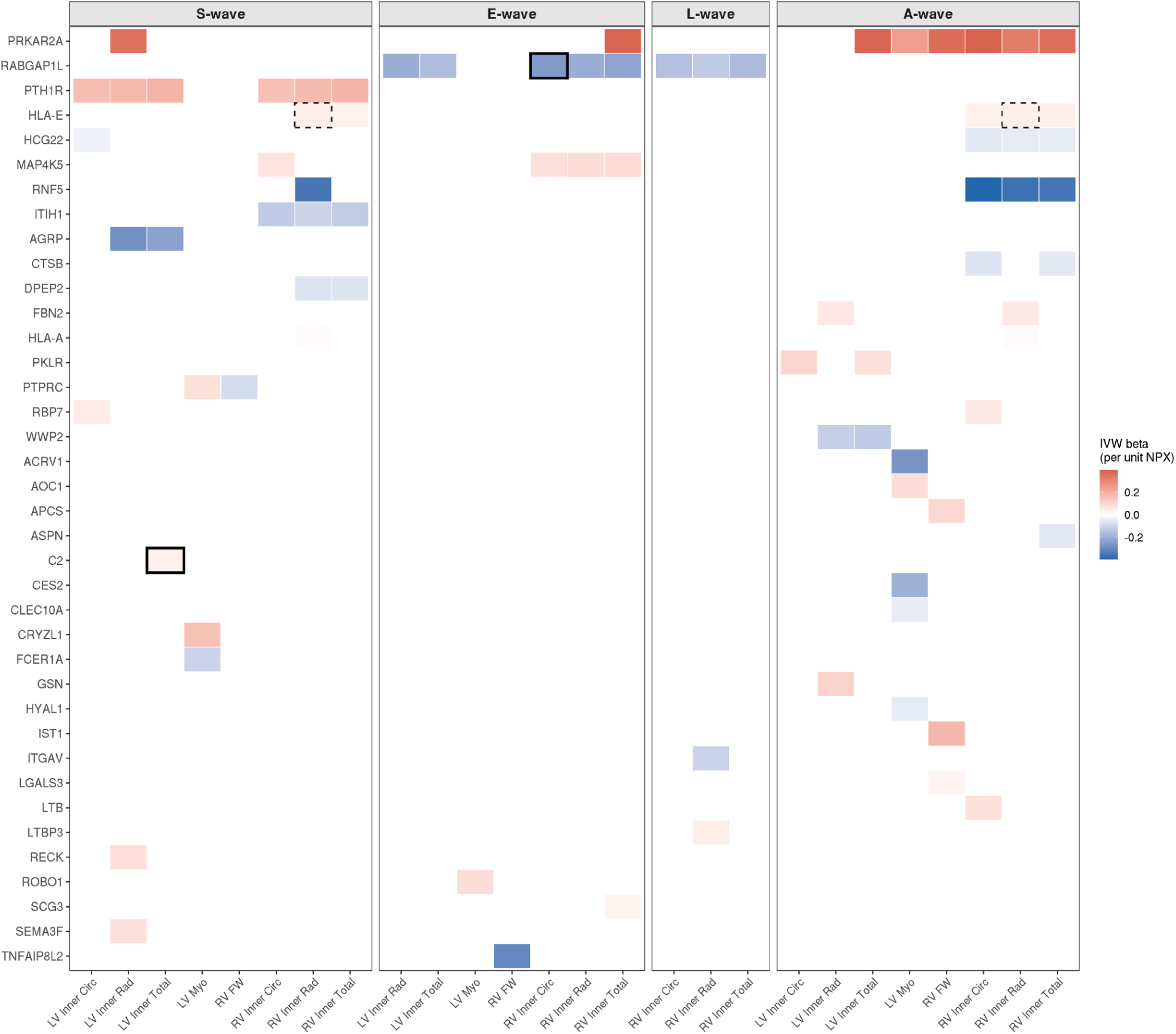
Causal protein-phenotype associations identified by Mendelian randomization. Heatmap displaying 81 protein-phenotype associations reaching IVW significance at Bonferroni-corrected threshold (p < 1.86×10⁻⁶) and passing MR-PRESSO outlier correction. Color indicates the IVW effect estimate (β per unit NPX), with red denoting positive and blue denoting negative associations with velocity. Columns represent the 26 optical flow velocity phenotypes with significant MR associations, grouped by cardiac phase (S-wave, E-wave, L-wave, A-wave); rows represent 38 unique proteins ordered by the number of associated phenotypes. Black solid outlines indicate strong colocalization evidence (PP.H4 > 0.8); black dashed outlines indicate moderate colocalization evidence (PP.H4 > 0.5). Two associations demonstrated strong colocalization: RABGAP1L with early diastolic RV inner circumferential velocity (PP.H4 = 0.937) and C2 with systolic LV inner velocity (PP.H4 = 0.966). Circ = circumferential; FW = free wall; IVW = inverse-variance-weighted; LV = left ventricular; Myo = myocardial; Rad = radial; RV = right ventricular; Total = total velocity magnitude.

**Supplemental Figure 9.**
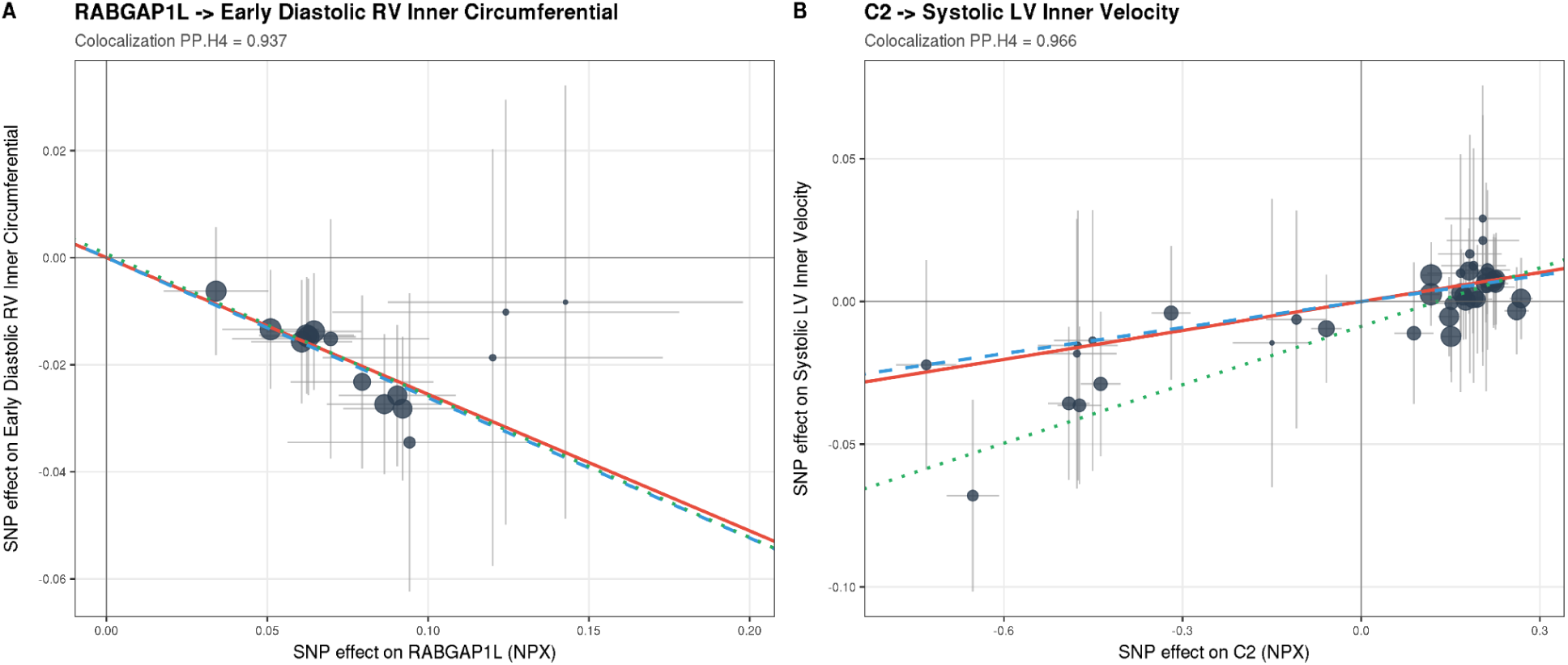
Mendelian randomization scatter plots for colocalized protein-phenotype associations. Each point represents a cis-pQTL instrument SNP, with horizontal and vertical bars indicating 95% confidence intervals for SNP effects on protein levels (NPX) and optical flow velocity phenotype, respectively. Point size is proportional to instrument precision (inverse of outcome SE). Lines represent causal effect estimates from fixed-effects IVW (red, solid), weighted median (blue, dashed), and MR-Egger (green, dotted) methods; weighted median and MR-Egger estimates with sensitivity statistics are reported in Supplemental Table 11. β values are per unit NPX (log₂-normalized protein expression). (A) RABGAP1L and early diastolic RV inner circumferential velocity. Higher genetically predicted RABGAP1L levels were associated with reduced early diastolic velocity (IVW β = −0.255, p = 5.49×10⁻²²), with directionally consistent estimates from weighted median (β = −0.261) and MR-Egger (β = −0.265) methods and no evidence of directional pleiotropy (MR-Egger intercept p = 0.92). Bayesian colocalization confirmed a shared causal variant (PP.H4 = 0.937). (B) C2 and systolic LV inner velocity. Higher genetically predicted C2 levels were associated with increased systolic velocity (IVW β = 0.034, p = 5.05×10⁻⁷; weighted median β = 0.034; PP.H4 = 0.966); the MR-Egger intercept was nominally significant (p = 0.015), and the MR-PRESSO global test was non-significant (p = 0.40). cis-pQTL = cis-protein quantitative trait locus; IVW = inverse variance weighted; LV = left ventricular; MR = Mendelian randomization; NPX = normalized protein expression; RV = right ventricular; SE = standard error; SNP = single nucleotide polymorphism.

**Supplemental Figure 10.**
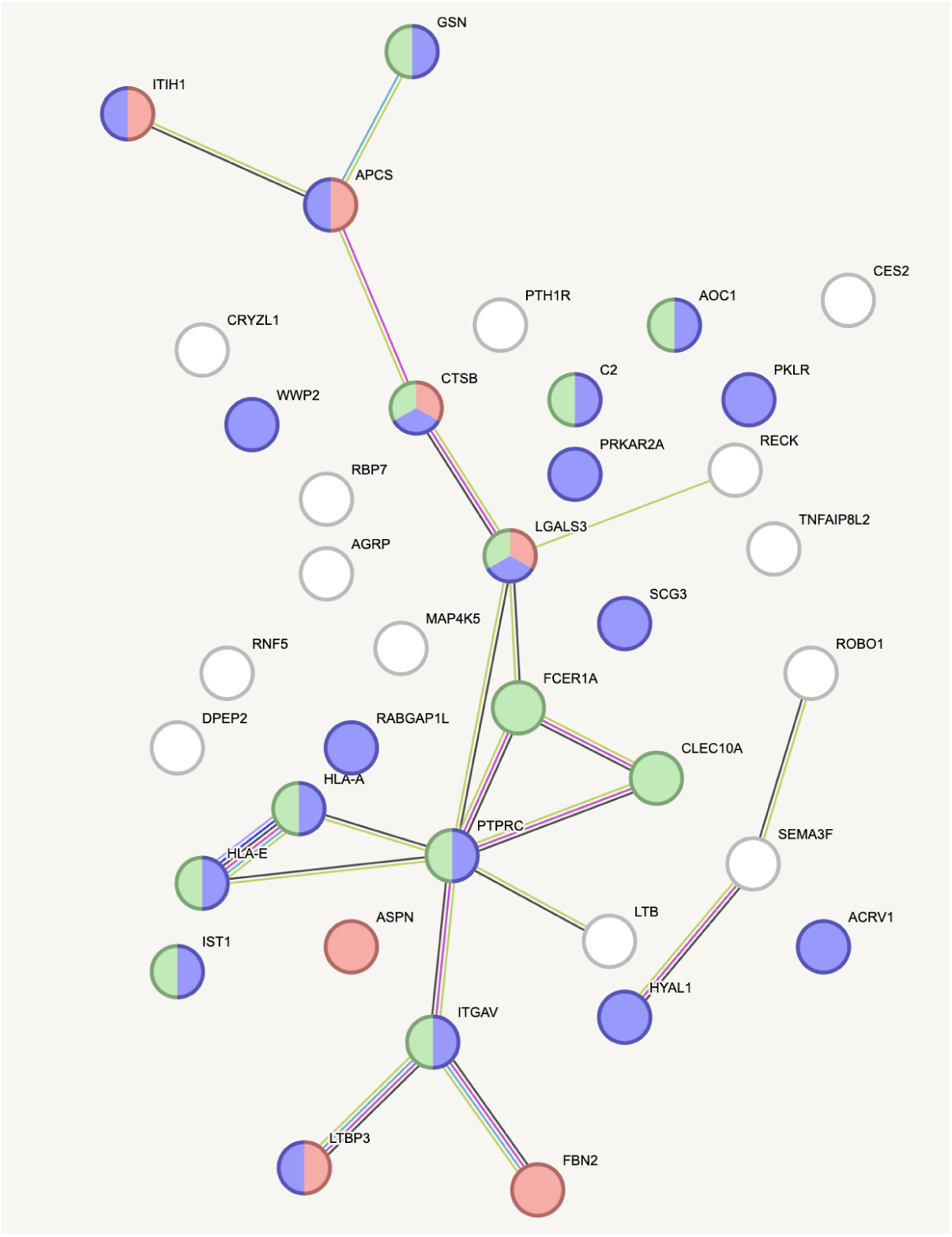
Protein-protein interaction network of MR-significant proteins. STRING-DB network for 37 mapped proteins (of 38 MR-PRESSO-validated) showing known and predicted protein-protein interactions. The network showed significantly more interactions than expected by chance (19 edges observed vs. 7 expected, PPI enrichment p = 1.18×10⁻⁴). Nodes are colored by functional enrichment: green, innate immune system (Reactome HSA-168249, 12 proteins, FDR = 6.1×10⁻⁴); pink, collagen-containing extracellular matrix (GO:0062023, 7 proteins, FDR = 2.6×10⁻³); purple, vesicle (GO:0031982, 20 proteins, FDR = 1.5×10⁻³). Proteins belonging to multiple categories are shown with split coloring. White nodes are not annotated to any of the three highlighted terms. Edge colors represent interaction evidence types (green: text mining; pink: experimentally determined; blue: curated databases; yellow: co-expression). FDR = false discovery rate; GO = Gene Ontology; MR = Mendelian randomization; PPI = protein-protein interaction.

