## Supplemental Tables for "Deep learning cardiac motion analysis reveals the dynamic pathophysiology and genetic architecture of heart failure with preserved ejection fraction"

| Parameter | Units | n | Valid % |
|---|---|---|---|
| Left ventricular cardiac output (BSA-indexed) | L/min/m² | 38,439 | 46.00% |
| Left ventricular ejection fraction | % | 39,120 | 46.80% |
| Left ventricular end-diastolic volume (BSA-indexed) | mL/m² | 38,439 | 46.00% |
| Left ventricular end-systolic volume (BSA-indexed) | mL/m² | 38,439 | 46.00% |
| Left ventricular circumferential strain (global) | % | 39,052 | 46.70% |
| Left ventricular longitudinal strain (global) | % | 38,003 | 45.50% |
| Left ventricular radial strain (global) | % | 39,052 | 46.70% |
| Right ventricular ejection fraction | % | 39,120 | 46.80% |
| Right ventricular end-diastolic volume (BSA-indexed) | mL/m² | 38,439 | 46.00% |
| Right ventricular end-systolic volume (BSA-indexed) | mL/m² | 38,439 | 46.00% |

**Supplemental Table 1. Full conventional cardiac parameter list.** Ten conventional cardiac parameters derived from UK Biobank cardiac MRI data (Instance 2) were extracted for comparison against optical flow velocity phenotypes. Volume and cardiac output parameters were body surface area (BSA)-indexed prior to analyses by dividing raw values by BSA (Body surface area | Instance 2); for these parameters, the reported n reflects participants with both a non-missing raw value and a valid BSA measurement. Among the 83,569 participants with successful optical flow analysis, conventional cardiac parameter availability ranged from 45.5% to 46.8%. BSA = body surface area; CMR = cardiac magnetic resonance.

| Parameter | n | Median (IQR) (cm/s) |
|---|---|---|
| LV Inner | | |
| Systolic LV Inner Total Velocity | 83566 | 280.2 (153.8) |
| Systolic LV Inner Circumferential Velocity | 83566 | 133.7 (83.1) |
| Systolic LV Inner Radial Velocity | 83566 | 222.4 (115.5) |
| Early Diastolic (E-Wave) LV Inner Total Velocity | 83544 | 270.3 (137.2) |
| Early Diastolic (E-Wave) LV Inner Circumferential Velocity | 83544 | 147.7 (81.9) |
| Early Diastolic (E-Wave) LV Inner Radial Velocity | 83544 | 186.9 (101.2) |
| Mid-Diastolic (L-Wave) LV Inner Total Velocity | 83127 | 251.9 (168.0) |
| Mid-Diastolic (L-Wave) LV Inner Circumferential Velocity | 83127 | 137.0 (85.0) |
| Mid-Diastolic (L-Wave) LV Inner Radial Velocity | 83127 | 176.0 (124.5) |
| Late Diastolic (A-Wave) LV Inner Total Velocity | 83130 | 208.4 (112.6) |
| Late Diastolic (A-Wave) LV Inner Circumferential Velocity | 83130 | 113.2 (67.1) |
| Late Diastolic (A-Wave) LV Inner Radial Velocity | 83130 | 146.0 (82.1) |
| LV Myocardium | | |
| Systolic LV Myocardium Velocity | 83566 | 44.9 (4.1) |
| Early Diastolic (E-Wave) LV Myocardium Velocity | 83544 | 42.0 (4.7) |
| Mid-Diastolic (L-Wave) LV Myocardium Velocity | 83127 | 42.0 (4.1) |
| Late Diastolic (A-Wave) LV Myocardium Velocity | 83130 | 39.8 (4.5) |
| RV Inner | | |
| Systolic RV Inner Total Velocity | 83566 | 416.9 (184.0) |
| Systolic RV Inner Circumferential Velocity | 83566 | 224.6 (105.9) |
| Systolic RV Inner Radial Velocity | 83566 | 283.5 (138.8) |
| Early Diastolic (E-Wave) RV Inner Total Velocity | 83544 | 282.9 (134.6) |
| Early Diastolic (E-Wave) RV Inner Circumferential Velocity | 83544 | 175.7 (88.2) |
| Early Diastolic (E-Wave) RV Inner Radial Velocity | 83544 | 179.1 (93.2) |
| Mid-Diastolic (L-Wave) RV Inner Total Velocity | 83127 | 236.1 (137.4) |
| Mid-Diastolic (L-Wave) RV Inner Circumferential Velocity | 83127 | 147.9 (87.1) |
| Mid-Diastolic (L-Wave) RV Inner Radial Velocity | 83127 | 146.4 (90.0) |
| Late Diastolic (A-Wave) RV Inner Total Velocity | 83130 | 228.0 (104.8) |
| Late Diastolic (A-Wave) RV Inner Circumferential Velocity | 83130 | 146.9 (70.3) |

| | | |
|---|---|---|
| Late Diastolic (A-Wave) RV Inner Radial Velocity | 83130 | 133.3 (68.6) |
| RV Myocardial Free Wall | | |
| Systolic RV Myocardial Free Wall Velocity | 83566 | 45.4 (5.1) |
| Early Diastolic (E-Wave) RV Myocardial Free Wall Velocity | 83544 | 44.2 (5.2) |
| Mid-Diastolic (L-Wave) RV Myocardial Free Wall Velocity | 83127 | 45.2 (4.2) |
| Late Diastolic (A-Wave) RV Myocardial Free Wall Velocity | 83130 | 42.8 (5.8) |

**Supplemental Table 2. Descriptive statistics and data completeness for 32 optical flow velocity phenotypes.** Velocity phenotypes represent peak values extracted from the optical flow velocity waveforms across the four key cardiac structures (LV inner cavity, LV myocardium, RV inner cavity, RV myocardial free wall) and across the entire cardiac cycle. Inner cavity velocities include total, radial, and circumferential vector components; myocardial velocities include total velocity only. The n column reflects the number of participants with a successfully detected peak for each optical flow velocity phenotype. Among the 83,569 participants for whom optical flow velocities were successfully calculated, 442 participants (0.53%) were missing one or more diastolic velocity metrics, reflecting physiologic variability in the presence and detectability of early (E-wave), mid-diastolic (L-wave), and late diastolic (A-wave) peaks. Only 3 participants (0.004%) had missing systolic (S-wave) velocity metrics. LV = left ventricular; RV = right ventricular; IQR = interquartile range.

| Parameter | Type | n | Deaths | HR per SD (95% CI) | p-value | FDR q-value |
|---|---|---|---|---|---|---|
| LV longitudinal strain (global) (%) | Conventional | 37,998 | 1,100 | 1.18 (1.11-1.25) | 3.63E-08 | 8.41E-07 |
| LV circumferential strain (global) (%) | Conventional | 39,047 | 1,149 | 1.17 (1.11-1.24) | 4.34E-08 | 8.41E-07 |
| LV ejection fraction (%) | Conventional | 39,115 | 1,152 | 0.86 (0.81-0.91) | 1.14E-07 | 1.48E-06 |
| LV radial strain (global) (%) | Conventional | 39,047 | 1,149 | 0.89 (0.84-0.94) | 1.03E-04 | 6.68E-04 |
| RV ejection fraction (%) | Conventional | 39,115 | 1,152 | 0.89 (0.84-0.95) | 1.61E-04 | 8.91E-04 |
| RV end-diastolic volume (BSA-indexed) (mL/m²) | Conventional | 38,434 | 1,114 | 0.91 (0.85-0.97) | 0.003 | 0.009 |
| LV end-systolic volume (BSA-indexed) (mL/m²) | Conventional | 38,434 | 1,114 | 1.08 (1.02-1.15) | 0.008 | 0.02 |
| LV cardiac output (BSA-indexed) (L/min/m²) | Conventional | 38,434 | 1,114 | 1.08 (1.01-1.14) | 0.014 | 0.031 |
| Late Diastolic (A-Wave) LV Inner Circumferential Velocity (cm/sec) | Optical Flow | 83,087 | 1,408 | 1.10 (1.05-1.16) | 6.99E-05 | 5.63E-04 |
| Mid-Diastolic (L-Wave) RV Inner Radial Velocity (cm/sec) | Optical Flow | 83,084 | 1,408 | 1.11 (1.05-1.16) | 7.25E-05 | 5.63E-04 |
| Late Diastolic (A-Wave) LV Inner Total Velocity (cm/sec) | Optical Flow | 83,087 | 1,408 | 1.09 (1.04-1.14) | 4.24E-04 | 0.002 |
| Late Diastolic (A-Wave) LV Myocardium Velocity (cm/sec) | Optical Flow | 83,087 | 1,408 | 1.09 (1.04-1.15) | 9.41E-04 | 0.004 |
| Mid-Diastolic (L-Wave) LV Inner Total Velocity (cm/sec) | Optical Flow | 83,084 | 1,408 | 1.09 (1.04-1.15) | 0.001 | 0.004 |
| Mid-Diastolic (L-Wave) LV Inner Circumferential Velocity (cm/sec) | Optical Flow | 83,084 | 1,408 | 1.09 (1.03-1.15) | 0.001 | 0.004 |
| Late Diastolic (A-Wave) LV Inner Radial Velocity (cm/sec) | Optical Flow | 83,087 | 1,408 | 1.08 (1.03-1.13) | 0.002 | 0.006 |
| Mid-Diastolic (L-Wave) LV Myocardium Velocity (cm/sec) | Optical Flow | 83,084 | 1,408 | 1.08 (1.03-1.14) | 0.003 | 0.009 |
| Early Diastolic (E-Wave) LV Myocardium Velocity (cm/sec) | Optical Flow | 83,501 | 1,418 | 0.93 (0.88-0.97) | 0.003 | 0.009 |
| Mid-Diastolic (L-Wave) RV Inner Total Velocity (cm/sec) | Optical Flow | 83,084 | 1,408 | 1.07 (1.02-1.12) | 0.01 | 0.023 |
| Early Diastolic (E-Wave) RV Myocardial Free Wall Velocity (cm/sec) | Optical Flow | 83,501 | 1,418 | 0.94 (0.90-0.99) | 0.02 | 0.041 |
| Mid-Diastolic (L-Wave) LV Inner Radial Velocity (cm/sec) | Optical Flow | 83,084 | 1,408 | 1.07 (1.01-1.12) | 0.021 | 0.041 |
| Late Diastolic (A-Wave) RV Inner Circumferential Velocity (cm/sec) | Optical Flow | 83,087 | 1,408 | 1.06 (1.01-1.11) | 0.025 | 0.046 |

**Supplemental Table 3. Full cohort univariate survival analysis: FDR-significant associations with all-cause mortality.** Sample sizes varied by parameter availability (Supplemental Table 1, Supplemental Table 2), with univariate Cox complete-case samples ranging from 37,998 to 83,501 participants with 1,100 to 1,418 deaths (death rate 1.7-2.9%), over median follow-up of 4.7 years (IQR 1.3 to 6.3 years). Twenty-one parameters (8 conventional, 13 optical flow) were significantly associated with all-cause mortality after cluster-adjusted FDR correction ($q < 0.05$) in univariate Cox proportional hazards models adjusted for age and sex. Hazard ratios are reported per standard deviation increase. Conventional parameters are ordered by p-value, followed by optical flow parameters ordered by p-value. FDR = false discovery rate; LV = left ventricular; RV = right ventricular.

| protein | velocity | beta | intercept | r_value | p_value | std_err | pearson_r | pearson_p | n_observations | fdr_bh_p | fdr_bh_sig | bonferroni_standard_p | bonferroni_standard_sig | bonferroni_cluster_p | bonferroni_cluster_sig | nominally_significant | beta_numeric |
|---|---|---|---|---|---|---|---|---|---|---|---|---|---|---|---|---|---|
| ace2 | late_diastolic_peaks_lv_inner_velocity_corrected | 12.3439631 | -3.146147565 | 0.06504522545 | 4.97E-09 | 2.108446035 | 0.06504522545 | 4.97E-09 | 8069 | 1.33E-06 | TRUE | 0.0002327146457 | TRUE | 0.0002084686885 | TRUE | FALSE | 12.3439631 |
| ace2 | late_diastolic_peaks_lv_inner_circumferential_velocity_corrected | 7.260391906 | -1.89823004 | 0.06809675439 | 9.18E-10 | 1.184318006 | 0.06809675439 | 9.18E-10 | 8069 | 2.90E-07 | TRUE | 4.30E-05 | TRUE | 3.85E-05 | TRUE | FALSE | 7.260391906 |
| acp5 | late_diastolic_peaks_lv_inner_velocity_corrected | 21.10213692 | -1.337427032 | 0.07519867528 | 1.35E-11 | 3.115507492 | 0.07519867528 | 1.35E-11 | 8069 | 8.19E-09 | TRUE | 6.31E-07 | TRUE | 5.65E-07 | TRUE | FALSE | 21.10213692 |
| acp5 | late_diastolic_peaks_lv_inner_radial_velocity_corrected | 14.63709236 | -0.5894619187 | 0.07381809208 | 3.16E-11 | 2.201656053 | 0.07381809208 | 3.16E-11 | 8069 | 1.59E-08 | TRUE | 1.48E-06 | TRUE | 1.33E-06 | TRUE | FALSE | 14.63709236 |
| acp5 | late_diastolic_peaks_rv_inner_circumferential_velocity_corrected | 10.43269841 | -0.9444136616 | 0.05815336195 | 1.72E-07 | 1.994023173 | 0.05815336195 | 1.72E-07 | 8069 | 3.03E-05 | TRUE | 0.008048488271 | TRUE | 0.007209936399 | TRUE | FALSE | 10.43269841 |
| acp5 | late_diastolic_peaks_lv_inner_circumferential_velocity_corrected | 10.78416796 | -0.9718561396 | 0.06840312414 | 7.71E-10 | 1.751202077 | 0.06840312414 | 7.71E-10 | 8069 | 2.52E-07 | TRUE | 3.61E-05 | TRUE | 3.23E-05 | TRUE | FALSE | 10.78416796 |
| acy1 | late_diastolic_peaks_lv_inner_velocity_corrected | 14.38596926 | -2.828120781 | 0.09932090672 | 3.80E-19 | 1.604684688 | 0.09932090672 | 3.80E-19 | 8069 | 2.97E-15 | TRUE | 1.78E-14 | TRUE | 1.60E-14 | TRUE | FALSE | 14.38596926 |
| acy1 | late_diastolic_peaks_lv_inner_radial_velocity_corrected | 9.25928952 | -1.638035 | 0.09046976474 | 3.88E-16 | 1.134837188 | 0.09046976474 | 3.88E-16 | 8069 | 9.88E-13 | TRUE | 1.82E-11 | TRUE | 1.63E-11 | TRUE | FALSE | 9.25928952 |
| acy1 | late_diastolic_peaks_lv_inner_circumferential_velocity_corrected | 7.487487014 | -1.730920679 | 0.09201172452 | 1.22E-16 | 0.9021747213 | 0.09201172452 | 1.22E-16 | 8069 | 3.35E-13 | TRUE | 5.70E-12 | TRUE | 5.10E-12 | TRUE | FALSE | 7.487487014 |
| adh4 | late_diastolic_peaks_lv_inner_velocity_corrected | 9.160630407 | -2.554854209 | 0.08898784177 | 1.16E-15 | 1.141595534 | 0.08898784177 | 1.16E-15 | 8069 | 2.59E-12 | TRUE | 5.45E-11 | TRUE | 4.88E-11 | TRUE | FALSE | 9.160630407 |
| adh4 | late_diastolic_peaks_lv_inner_radial_velocity_corrected | 6.087535809 | -1.450345208 | 0.08368952265 | 5.09E-14 | 0.8070272015 | 0.08368952265 | 5.09E-14 | 8069 | 6.61E-11 | TRUE | 2.38E-09 | TRUE | 2.13E-09 | TRUE | FALSE | 6.087535809 |
| adh4 | late_diastolic_peaks_lv_inner_circumferential_velocity_corrected | 4.587400615 | -1.599821044 | 0.07931905319 | 9.67E-13 | 0.6418929683 | 0.07931905319 | 9.67E-13 | 8069 | 7.67E-10 | TRUE | 4.53E-08 | TRUE | 4.05E-08 | TRUE | FALSE | 4.587400615 |
| agrn | late_diastolic_peaks_lv_inner_velocity_corrected | 18.35829382 | -1.871468512 | 0.06046122725 | 6.62E-08 | 3.395783219 | 0.06046122725 | 6.62E-08 | 7968 | 1.29E-05 | TRUE | 0.003101051696 | TRUE | 0.002777960873 | TRUE | FALSE | 18.35829382 |
| agrn | late_diastolic_peaks_lv_inner_circumferential_velocity_corrected | 9.525174668 | -1.227342528 | 0.05581991703 | 6.17E-07 | 1.908912199 | 0.05581991703 | 6.17E-07 | 7968 | 9.00E-05 | TRUE | 0.02888894586 | TRUE | 0.02587907882 | TRUE | FALSE | 9.525174668 |
| agrp | late_diastolic_peaks_lv_inner_velocity_corrected | 9.968150641 | -2.444610011 | 0.05643763889 | 4.12E-07 | 1.967129853 | 0.05643763889 | 4.12E-07 | 8038 | 6.46E-05 | TRUE | 0.0193052906 | TRUE | 0.01729392063 | TRUE | FALSE | 9.968150641 |
| agrp | late_diastolic_peaks_lv_inner_radial_velocity_corrected | 7.13180844 | -1.348662776 | 0.05714547544 | 2.95E-07 | 1.389912874 | 0.05714547544 | 2.95E-07 | 8038 | 4.83E-05 | TRUE | 0.01379968076 | TRUE | 0.01236192652 | TRUE | FALSE | 7.13180844 |
| aldh1a1 | late_diastolic_peaks_lv_inner_velocity_corrected | 11.18545026 | -3.223856956 | 0.06422182745 | 9.70E-09 | 1.948002618 | 0.06422182745 | 9.70E-09 | 7963 | 2.39E-06 | TRUE | 0.0004541014453 | TRUE | 0.0004067897509 | TRUE | FALSE | 11.18545026 |
| aldh1a1 | late_diastolic_peaks_lv_inner_radial_velocity_corrected | 6.951369006 | -1.861794381 | 0.0564485946 | 4.64E-07 | 1.377971912 | 0.0564485946 | 4.64E-07 | 7963 | 7.15E-05 | TRUE | 0.02174259173 | TRUE | 0.01947728546 | TRUE | FALSE | 6.951369006 |
| aldh1a1 | late_diastolic_peaks_lv_inner_circumferential_velocity_corrected | 6.30481581 | -1.956262676 | 0.06450340952 | 8.36E-09 | 1.093202342 | 0.06450340952 | 8.36E-09 | 7963 | 2.09E-06 | TRUE | 0.000391230804 | TRUE | 0.0003504694446 | TRUE | FALSE | 6.30481581 |
| ambp | late_diastolic_peaks_lv_inner_velocity_corrected | 28.69437736 | -1.427128309 | 0.06048547056 | 5.49E-08 | 5.274839814 | 0.06048547056 | 5.49E-08 | 8061 | 1.11E-05 | TRUE | 0.002569593646 | TRUE | 0.002301874108 | TRUE | FALSE | 28.69437736 |
| ambp | late_diastolic_peaks_lv_inner_radial_velocity_corrected | 19.46208636 | -0.6746032141 | 0.05805302112 | 1.83E-07 | 3.728129044 | 0.05805302112 | 1.83E-07 | 8061 | 3.19E-05 | TRUE | 0.00856995257 | TRUE | 0.007677070636 | TRUE | FALSE | 19.46208636 |
| ang | late_diastolic_peaks_lv_inner_velocity_corrected | 19.03260978 | -2.287400111 | 0.06984073056 | 4.42E-10 | 3.046991616 | 0.06984073056 | 4.42E-10 | 7962 | 1.60E-07 | TRUE | 2.07E-05 | TRUE | 1.85E-05 | TRUE | FALSE | 19.03260978 |
| ang | late_diastolic_peaks_lv_inner_radial_velocity_corrected | 11.98808797 | -1.306055535 | 0.06222824379 | 2.74E-08 | 2.155079717 | 0.06222824379 | 2.74E-08 | 7962 | 6.03E-06 | TRUE | 0.001283818206 | TRUE | 0.001150060397 | TRUE | FALSE | 11.98808797 |
| ang | late_diastolic_peaks_lv_inner_circumferential_velocity_corrected | 10.35844993 | -1.376903991 | 0.06756289895 | 1.59E-09 | 1.714495977 | 0.06756289895 | 1.59E-09 | 7962 | 4.75E-07 | TRUE | 7.46E-05 | TRUE | 6.68E-05 | TRUE | FALSE | 10.35844993 |
| angptl2 | late_diastolic_peaks_lv_inner_velocity_corrected | 19.73413015 | -1.927766567 | 0.08355763229 | 8.45E-14 | 2.639531722 | 0.08355763229 | 8.45E-14 | 7952 | 9.83E-11 | TRUE | 3.96E-09 | TRUE | 3.55E-09 | TRUE | FALSE | 19.73413015 |
| angptl2 | late_diastolic_peaks_lv_inner_radial_velocity_corrected | 12.29688077 | -1.052830937 | 0.07366007125 | 4.82E-11 | 1.867230172 | 0.07366007125 | 4.82E-11 | 7952 | 2.28E-08 | TRUE | 2.26E-06 | TRUE | 2.02E-06 | TRUE | FALSE | 12.29688077 |
| angptl2 | late_diastolic_peaks_lv_inner_circumferential_velocity_corrected | 8.326398403 | -1.351142856 | 0.06271700276 | 2.18E-08 | 1.486046884 | 0.06271700276 | 2.18E-08 | 7952 | 4.92E-06 | TRUE | 0.00101892574 | TRUE | 0.0009127664146 | TRUE | FALSE | 8.326398403 |
| asgr1 | late_diastolic_peaks_lv_inner_velocity_corrected | 25.62505681 | -1.383776989 | 0.0822277261 | 1.39E-13 | 3.457938958 | 0.0822277261 | 1.39E-13 | 8069 | 1.44E-10 | TRUE | 6.49E-09 | TRUE | 5.81E-09 | TRUE | FALSE | 25.62505681 |
| asgr1 | late_diastolic_peaks_lv_inner_radial_velocity_corrected | 16.42196564 | -0.7132290289 | 0.07457664337 | 1.98E-11 | 2.44486681 | 0.07457664337 | 1.98E-11 | 8069 | 1.12E-08 | TRUE | 9.28E-07 | TRUE | 8.31E-07 | TRUE | FALSE | 16.42196564 |
| asgr1 | late_diastolic_peaks_lv_inner_circumferential_velocity_corrected | 13.982663 | -0.9354473142 | 0.07986359857 | 6.75E-13 | 1.943101755 | 0.07986359857 | 6.75E-13 | 8069 | 5.75E-10 | TRUE | 3.16E-08 | TRUE | 2.83E-08 | TRUE | FALSE | 13.982663 |
| baiap2 | late_diastolic_peaks_lv_inner_velocity_corrected | 10.42734278 | -2.714671762 | 0.06872451704 | 6.42E-10 | 1.685302621 | 0.06872451704 | 6.42E-10 | 8069 | 2.16E-07 | TRUE | 3.01E-05 | TRUE | 2.69E-05 | TRUE | FALSE | 10.42734278 |
| baiap2 | late_diastolic_peaks_lv_inner_radial_velocity_corrected | 6.408303871 | -1.576790174 | 0.0597730467 | 7.73E-08 | 1.191528567 | 0.0597730467 | 7.73E-08 | 8069 | 1.49E-05 | TRUE | 0.003620427326 | TRUE | 0.003243224054 | TRUE | FALSE | 6.408303871 |
| baiap2 | late_diastolic_peaks_lv_inner_circumferential_velocity_corrected | 5.898205264 | -1.65357257 | 0.06919320852 | 4.91E-10 | 0.9468000058 | 0.06919320852 | 4.91E-10 | 8069 | 1.75E-07 | TRUE | 2.30E-05 | TRUE | 2.06E-05 | TRUE | FALSE | 5.898205264 |
| c19orf12 | late_diastolic_peaks_lv_inner_velocity_corrected | 9.474829574 | -3.56023048 | 0.06189958612 | 3.74E-08 | 1.720151111 | 0.06189958612 | 3.74E-08 | 7890 | 8.00E-06 | TRUE | 0.001751222838 | TRUE | 0.001568767309 | TRUE | FALSE | 9.474829574 |
| c19orf12 | late_diastolic_peaks_lv_inner_radial_velocity_corrected | 6.586843962 | -2.14355064 | 0.06088733065 | 6.21E-08 | 1.215795331 | 0.06088733065 | 6.21E-08 | 7890 | 1.23E-05 | TRUE | 0.002909279546 | TRUE | 0.002606168984 | TRUE | FALSE | 6.586843962 |
| ca14 | diastolic_peaks_lwave_lv_inner_velocity_corrected | -21.53179787 | -0.04818473834 | -0.06295477373 | 1.52E-08 | 3.800670218 | -0.06295477373 | 1.52E-08 | 8068 | 3.52E-06 | TRUE | 0.0007108823584 | TRUE | 0.0006368173026 | TRUE | FALSE | -21.53179787 |
| ca14 | diastolic_peaks_lwave_lv_inner_radial_velocity_corrected | -15.98359277 | 0.9935112841 | -0.06529415711 | 4.35E-09 | 2.719839199 | -0.06529415711 | 4.35E-09 | 8068 | 1.18E-06 | TRUE | 0.0002037628535 | TRUE | 0.0001825333112 | TRUE | FALSE | -15.98359277 |
| ca14 | diastolic_peaks_lwave_lv_inner_circumferential_velocity_corrected | -10.66445576 | -0.4331384624 | -0.05928583473 | 9.87E-08 | 1.999374152 | -0.05928583473 | 9.87E-08 | 8068 | 1.85E-05 | TRUE | 0.004622362507 | TRUE | 0.004140770113 | TRUE | FALSE | -10.66445576 |
| ca14 | late_diastolic_peaks_lv_inner_velocity_corrected | -19.78044388 | -1.357772691 | -0.07734275605 | 3.48E-12 | 2.83894845 | -0.07734275605 | 3.48E-12 | 8069 | 2.40E-09 | TRUE | 1.63E-07 | TRUE | 1.46E-07 | TRUE | FALSE | -19.78044388 |
| ca14 | late_diastolic_peaks_lv_inner_radial_velocity_corrected | -13.09157065 | -0.6595824987 | -0.0724435316 | 7.27E-11 | 2.00675109 | -0.0724435316 | 7.27E-11 | 8069 | 3.12E-08 | TRUE | 3.40E-06 | TRUE | 3.05E-06 | TRUE | FALSE | -13.09157065 |
| ca14 | late_diastolic_peaks_lv_inner_circumferential_velocity_corrected | -10.70439546 | -0.9291922901 | -0.07449914636 | 2.08E-11 | 1.595314341 | -0.07449914636 | 2.08E-11 | 8069 | 1.15E-08 | TRUE | 9.73E-07 | TRUE | 8.72E-07 | TRUE | FALSE | -10.70439546 |
| ca5a | late_diastolic_peaks_lv_inner_velocity_corrected | 7.082665708 | -2.797359488 | 0.06972286505 | 3.62E-10 | 1.128255096 | 0.06972286505 | 3.62E-10 | 8069 | 1.34E-07 | TRUE | 1.69E-05 | TRUE | 1.52E-05 | TRUE | FALSE | 7.082665708 |
| ca5a | late_diastolic_peaks_lv_inner_radial_velocity_corrected | 4.574825004 | -1.617499228 | 0.06373488642 | 1.00E-08 | 0.7975489791 | 0.06373488642 | 1.00E-08 | 8069 | 2.45E-06 | TRUE | 0.0004699216883 | TRUE | 0.0004209617224 | TRUE | FALSE | 4.574825004 |
| ca5a | late_diastolic_peaks_lv_inner_circumferential_velocity_corrected | 3.312322047 | -1.731935603 | 0.05803855306 | 1.82E-07 | 0.634347541 | 0.05803855306 | 1.82E-07 | 8069 | 3.19E-05 | TRUE | 0.008510707366 | TRUE | 0.007623998042 | TRUE | FALSE | 3.312322047 |
| ccl16 | late_diastolic_peaks_lv_inner_velocity_corrected | 13.1112102 | -1.053258111 | 0.07430475727 | 2.34E-11 | 1.959150941 | 0.07430475727 | 2.34E-11 | 8069 | 1.25E-08 | TRUE | 1.10E-06 | TRUE | 9.83E-07 | TRUE | FALSE | 13.1112102 |
| ccl16 | late_diastolic_peaks_lv_inner_radial_velocity_corrected | 8.694469307 | -0.4553789175 | 0.06973345255 | 3.60E-10 | 1.384801014 | 0.06973345255 | 3.60E-10 | 8069 | 1.34E-07 | TRUE | 1.68E-05 | TRUE | 1.51E-05 | TRUE | FALSE | 8.694469307 |
| ccl16 | late_diastolic_peaks_lv_inner_circumferential_velocity_corrected | 6.332946319 | -0.8845541917 | 0.06388298301 | 9.28E-09 | 1.10147994 | 0.06388298301 | 9.28E-09 | 8069 | 2.30E-06 | TRUE | 0.000434338802 | TRUE | 0.0003890861281 | TRUE | FALSE | 6.332946319 |
| ccl17 | late_diastolic_peaks_rv_inner_radial_velocity_corrected | 3.812840773 | -0.5063184436 | 0.05595512859 | 6.01E-07 | 0.7633227459 | 0.05595512859 | 6.01E-07 | 7946 | 8.87E-05 | TRUE | 0.02812342872 | TRUE | 0.02519331899 | TRUE | FALSE | 3.812840773 |
| ccl18 | late_diastolic_peaks_lv_inner_velocity_corrected | 7.809547566 | -2.381301653 | 0.05879749163 | 1.83E-07 | 1.495940527 | 0.05879749163 | 1.83E-07 | 7858 | 3.19E-05 | TRUE | 0.008568144184 | TRUE | 0.007675450661 | TRUE | FALSE | 7.809547566 |
| ccl18 | late_diastolic_peaks_lv_inner_radial_velocity_corrected | 5.374079339 | -1.403775479 | 0.05730649602 | 3.71E-07 | 1.056294915 | 0.05730649602 | 3.71E-07 | 7858 | 5.91E-05 | TRUE | 0.01736337601 | TRUE | 0.01555432927 | TRUE | FALSE | 5.374079339 |
| ccl18 | late_diastolic_peaks_lv_inner_circumferential_velocity_corrected | 4.099368361 | -1.42487403 | 0.05488859449 | 1.12E-06 | 0.8413542597 | 0.05488859449 | 1.12E-06 | 7858 | 0.0001534688033 | TRUE | 0.05263979953 | FALSE | 0.04715539042 | TRUE | FALSE | 4.099368361 |
| ccl22 | diastolic_peaks_lwave_lv_inner_velocity_corrected | 15.41681672 | -0.03555877466 | 0.0619239877 | 3.92E-08 | 2.80314467 | 0.0619239877 | 3.92E-08 | 7860 | 8.34E-06 | TRUE | 0.00183566293 | TRUE | 0.001644409799 | TRUE | FALSE | 15.41681672 |
| ccl22 | diastolic_peaks_lwave_lv_inner_circumferential_velocity_corrected | 7.841792038 | -0.3981760297 | 0.05988561393 | 1.08E-07 | 1.474540313 | 0.05988561393 | 1.08E-07 | 7860 | 1.99E-05 | TRUE | 0.005043372956 | TRUE | 0.004517916536 | TRUE | FALSE | 7.841792038 |
| ccl22 | late_diastolic_peaks_rv_inner_velocity_corrected | 11.7502561 | -0.9831010855 | 0.06050785151 | 7.92E-08 | 2.186530184 | 0.06050785151 | 7.92E-08 | 7861 | 1.52E-05 | TRUE | 0.003710116898 | TRUE | 0.003323569094 | TRUE | FALSE | 11.7502561 |
| ccl22 | late_diastolic_peaks_lv_inner_velocity_corrected | 14.65814454 | -1.582894713 | 0.07887427107 | 2.51E-12 | 2.089800705 | 0.07887427107 | 2.51E-12 | 7861 | 1.78E-09 | TRUE | 1.17E-07 | TRUE | 1.05E-07 | TRUE | FALSE | 14.65814454 |
| ccl22 | late_diastolic_peaks_rv_inner_radial_velocity_corrected | 7.148676302 | -0.3114146416 | 0.05939277568 | 1.37E-07 | 1.355317432 | 0.05939277568 | 1.37E-07 | 7861 | 2.45E-05 | TRUE | 0.006396787499 | TRUE | 0.005730322202 | TRUE | FALSE | 7.148676302 |
| ccl22 | late_diastolic_peaks_lv_inner_radial_velocity_corrected | 10.02046756 | -0.7481060149 | 0.07622183485 | 1.32E-11 | 1.478630597 | 0.07622183485 | 1.32E-11 | 7861 | 8.11E-09 | TRUE | 6.17E-07 | TRUE | 5.52E-07 | TRUE | FALSE | 10.02046756 |
| ccl22 | late_diastolic_peaks_rv_inner_circumferential_velocity_corrected | 7.397383123 | -1.035465911 | 0.06220324173 | 3.40E-08 | 1.338873741 | 0.06220324173 | 3.40E-08 | 7861 | 7.33E-06 | TRUE | 0.0015911351 | TRUE | 0.001425358712 | TRUE | FALSE | 7.397383123 |
| ccl22 | late_diastolic_peaks_lv_inner_circumferential_velocity_corrected | 7.911202987 | -1.040037792 | 0.07578979893 | 1.72E-11 | 1.174078725 | 0.07578979893 | 1.72E-11 | 7861 | 1.01E-08 | TRUE | 8.04E-07 | TRUE | 7.20E-07 | TRUE | FALSE | 7.911202987 |
| ccl7 | late_diastolic_peaks_lv_inner_velocity_corrected | 8.992170992 | -2.496808741 | 0.05622973198 | 5.05E-07 | 1.788245901 | 0.05622973198 | 5.05E-07 | 7974 | 7.73E-05 | TRUE | 0.02364880105 | TRUE | 0.02118489159 | TRUE | FALSE | 8.992170992 |
| cd163 | late_diastolic_peaks_lv_inner_velocity_corrected | 13.9285439 | -2.163813799 | 0.06234474018 | 2.49E-08 | 2.496238048 | 0.06234474018 | 2.49E-08 | 7981 | 5.52E-06 | TRUE | 0.001164121777 | TRUE | 0.001042834839 | TRUE | FALSE | 13.9285439 |
| cd163 | late_diastolic_peaks_lv_inner_radial_velocity_corrected | 9.550244748 | -1.1703879 | 0.06047843229 | 6.41E-08 | 1.764590925 | 0.06047843229 | 6.41E-08 | 7981 | 1.26E-05 | TRUE | 0.002999873152 | TRUE | 0.002687323868 | TRUE | FALSE | 9.550244748 |
| cd163 | late_diastolic_peaks_rv_inner_circumferential_velocity_corrected | 7.866399587 | -1.285609409 | 0.05497397222 | 8.92E-07 | 1.599511658 | 0.05497397222 | 8.92E-07 | 7981 | 0.000124256627 | TRUE | 0.04175022666 | TRUE | 0.03740037492 | TRUE | FALSE | 7.866399587 |
| cd4 | late_diastolic_peaks_lv_inner_circumferential_velocity_corrected | 10.94850098 | -1.29859916 | 0.05584107278 | 6.13E-07 | 2.193598351 | 0.05584107278 | 6.13E-07 | 7966 | 9.00E-05 | TRUE | 0.02870030129 | TRUE | 0.02571008865 | TRUE | FALSE | 10.94850098 |

| | | | | | | | | | | | | | | | | | |
|---|---|---|---|---|---|---|---|---|---|---|---|---|---|---|---|---|---|
| **cdcp1** | late_diastolic_peaks_lv_inner_velocity_corrected | 10.43277702 | -1.512080957 | 0.05672354122 | 3.42E-07 | 2.044470038 | 0.05672354122 | 3.42E-07 | 8069 | 5.50E-05 | TRUE | 0.01601450698 | TRUE | 0.01434599553 | TRUE | FALSE | 10.43277702 |
| **cdh2** | late_diastolic_peaks_lv_inner_velocity_corrected | 14.68802237 | -1.796104091 | 0.05774771915 | 2.09E-07 | 2.827138655 | 0.05774771915 | 2.09E-07 | 8069 | 3.58E-05 | TRUE | 0.009799337695 | TRUE | 0.008778369199 | TRUE | FALSE | 14.68802237 |
| **cdh2** | late_diastolic_peaks_lv_inner_circumferential_velocity_corrected | 7.87715089 | -1.172836881 | 0.05512469704 | 7.24E-07 | 1.588568928 | 0.05512469704 | 7.24E-07 | 8069 | 0.000103703095 | TRUE | 0.03391091207 | TRUE | 0.03037781892 | TRUE | FALSE | 7.87715089 |
| **cdhr2** | late_diastolic_peaks_lv_inner_velocity_corrected | 7.876019982 | -1.420870214 | 0.07790944242 | 2.42E-12 | 1.12211821 | 0.07790944242 | 2.42E-12 | 8069 | 1.77E-09 | TRUE | 1.13E-07 | TRUE | 1.01E-07 | TRUE | FALSE | 7.876019982 |
| **cdhr2** | late_diastolic_peaks_lv_inner_radial_velocity_corrected | 4.905746131 | -0.76755449 | 0.0686772041 | 6.60E-10 | 0.7934322055 | 0.0686772041 | 6.60E-10 | 8069 | 2.21E-07 | TRUE | 3.09E-05 | TRUE | 2.77E-05 | TRUE | FALSE | 4.905746131 |
| **cdhr2** | late_diastolic_peaks_lv_inner_circumferential_velocity_corrected | 4.513614784 | -0.909104593 | 0.07947184927 | 8.74E-13 | 0.6303464939 | 0.07947184927 | 8.74E-13 | 8069 | 7.31E-10 | TRUE | 4.09E-08 | TRUE | 3.67E-08 | TRUE | FALSE | 4.513614784 |
| **cdhr5** | late_diastolic_peaks_lv_inner_velocity_corrected | 13.55914543 | -2.025667024 | 0.05765275831 | 2.19E-07 | 2.614166441 | 0.05765275831 | 2.19E-07 | 8069 | 3.72E-05 | TRUE | 0.01025950877 | TRUE | 0.009190596199 | TRUE | FALSE | 13.55914543 |
| **cdhr5** | late_diastolic_peaks_lv_inner_radial_velocity_corrected | 10.36500759 | -0.989385239 | 0.0623708475 | 2.06E-08 | 1.846654182 | 0.0623708475 | 2.06E-08 | 8069 | 4.70E-06 | TRUE | 0.0009625708957 | TRUE | 0.0008622830405 | TRUE | FALSE | 10.36500759 |
| **ces1** | late_diastolic_peaks_lv_inner_velocity_corrected | 9.311409732 | -3.084392956 | 0.07821141492 | 2.47E-12 | 1.32715675 | 0.07821141492 | 2.47E-12 | 8000 | 1.78E-09 | TRUE | 1.16E-07 | TRUE | 1.04E-07 | TRUE | FALSE | 9.311409732 |
| **ces1** | late_diastolic_peaks_lv_inner_radial_velocity_corrected | 5.930788111 | -1.845097592 | 0.07057837994 | 2.62E-10 | 0.9372719992 | 0.07057837994 | 2.62E-10 | 8000 | 1.01E-07 | TRUE | 1.23E-05 | TRUE | 1.10E-05 | TRUE | FALSE | 5.930788111 |
| **ces1** | late_diastolic_peaks_lv_inner_circumferential_velocity_corrected | 4.792533543 | -1.860592126 | 0.07154112658 | 1.49E-10 | 0.7471442254 | 0.07154112658 | 1.49E-10 | 8000 | 5.98E-08 | TRUE | 6.99E-06 | TRUE | 6.26E-06 | TRUE | FALSE | 4.792533543 |
| **cntn3** | late_diastolic_peaks_lv_inner_velocity_corrected | 18.72955327 | -2.372296213 | 0.07028560853 | 3.41E-10 | 2.979214803 | 0.07028560853 | 3.41E-10 | 7963 | 1.30E-07 | TRUE | 1.60E-05 | TRUE | 1.43E-05 | TRUE | FALSE | 18.72955327 |
| **cntn3** | late_diastolic_peaks_lv_inner_radial_velocity_corrected | 13.38775125 | -1.261833999 | 0.07105591348 | 2.18E-10 | 2.106320498 | 0.07105591348 | 2.18E-10 | 7963 | 8.60E-08 | TRUE | 1.02E-05 | TRUE | 9.16E-06 | TRUE | FALSE | 13.38775125 |
| **cntn3** | late_diastolic_peaks_lv_inner_circumferential_velocity_corrected | 8.594580842 | -1.555699409 | 0.0574704729 | 2.87E-07 | 1.673315081 | 0.0574704729 | 2.87E-07 | 7963 | 4.71E-05 | TRUE | 0.01343181262 | TRUE | 0.01203238564 | TRUE | FALSE | 8.594580842 |
| **col4a1** | late_diastolic_peaks_lv_inner_velocity_corrected | -20.5061452 | -2.909622587 | -0.08337540766 | 6.32E-14 | 2.72882238 | -0.08337540766 | 6.32E-14 | 8069 | 7.78E-11 | TRUE | 2.96E-09 | TRUE | 2.65E-09 | TRUE | FALSE | -20.5061452 |
| **col4a1** | late_diastolic_peaks_lv_inner_radial_velocity_corrected | -15.39839625 | -1.667946546 | -0.08860407235 | 1.54E-15 | 1.927322089 | -0.08860407235 | 1.54E-15 | 8069 | 3.28E-12 | TRUE | 7.22E-11 | TRUE | 6.47E-11 | TRUE | FALSE | -15.39839625 |
| **col4a1** | late_diastolic_peaks_lv_inner_circumferential_velocity_corrected | -9.050311525 | -1.789968385 | -0.06549724181 | 3.89E-09 | 1.53515046 | -0.06549724181 | 3.89E-09 | 8069 | 1.06E-06 | TRUE | 0.0001820384326 | TRUE | 0.0001630723034 | TRUE | FALSE | -9.050311525 |
| **cpm** | late_diastolic_peaks_lv_inner_velocity_corrected | 22.25908364 | -1.643031801 | 0.08679220395 | 7.07E-15 | 2.854395557 | 0.08679220395 | 7.07E-15 | 8014 | 1.14E-11 | TRUE | 3.31E-10 | TRUE | 2.97E-10 | TRUE | FALSE | 22.25908364 |
| **cpm** | late_diastolic_peaks_lv_inner_radial_velocity_corrected | 14.53403363 | -0.8511299418 | 0.08018556668 | 6.54E-13 | 2.018453658 | 0.08018556668 | 6.54E-13 | 8014 | 5.67E-10 | TRUE | 3.06E-08 | TRUE | 2.74E-08 | TRUE | FALSE | 14.53403363 |
| **cpm** | late_diastolic_peaks_lv_inner_circumferential_velocity_corrected | 10.75880542 | -1.180094444 | 0.0747021404 | 2.15E-11 | 1.604520879 | 0.0747021404 | 2.15E-11 | 8014 | 1.17E-08 | TRUE | 1.00E-06 | TRUE | 9.00E-07 | TRUE | FALSE | 10.75880542 |
| **crhbp** | late_diastolic_peaks_lv_inner_velocity_corrected | 21.18110058 | -1.894328087 | 0.06364741223 | 1.41E-08 | 3.729962817 | 0.06364741223 | 1.41E-08 | 7930 | 3.31E-06 | TRUE | 0.0006580837385 | TRUE | 0.000589519639 | TRUE | FALSE | 21.18110058 |
| **crhbp** | late_diastolic_peaks_lv_inner_radial_velocity_corrected | 14.35820277 | -0.9884523963 | 0.06099552531 | 5.45E-08 | 2.63882684 | 0.06099552531 | 5.45E-08 | 7930 | 1.11E-05 | TRUE | 0.002552573159 | TRUE | 0.002286626943 | TRUE | FALSE | 14.35820277 |
| **crip2** | late_diastolic_peaks_lv_inner_velocity_corrected | 14.99544339 | -2.055311733 | 0.05952554117 | 8.75E-08 | 2.799813792 | 0.05952554117 | 8.75E-08 | 8069 | 1.65E-05 | TRUE | 0.004096018407 | TRUE | 0.003669264489 | TRUE | FALSE | 14.99544339 |
| **crip2** | late_diastolic_peaks_lv_inner_circumferential_velocity_corrected | 7.827941246 | -1.327045862 | 0.0553091697 | 6.65E-07 | 1.57336359 | 0.0553091697 | 6.65E-07 | 8069 | 9.64E-05 | TRUE | 0.03113095811 | TRUE | 0.02788750141 | TRUE | FALSE | 7.827941246 |
| **ctsd** | late_diastolic_peaks_lv_inner_velocity_corrected | 18.64387027 | -2.412762052 | 0.0734126392 | 1.85E-10 | 2.921255197 | 0.0734126392 | 1.85E-10 | 7519 | 7.34E-08 | TRUE | 8.66E-06 | TRUE | 7.76E-06 | TRUE | FALSE | 18.64387027 |
| **ctsd** | late_diastolic_peaks_lv_inner_radial_velocity_corrected | 11.93267873 | -1.263510376 | 0.06659137972 | 7.48E-09 | 2.062208621 | 0.06659137972 | 7.48E-09 | 7519 | 1.90E-06 | TRUE | 0.0003502620936 | TRUE | 0.0003137691618 | TRUE | FALSE | 11.93267873 |
| **ctsd** | late_diastolic_peaks_lv_inner_circumferential_velocity_corrected | 9.162658384 | -1.585047692 | 0.06417635297 | 2.55E-08 | 1.643342156 | 0.06417635297 | 2.55E-08 | 7519 | 5.64E-06 | TRUE | 0.001194730137 | TRUE | 0.001070254191 | TRUE | FALSE | 9.162658384 |
| **cxcl10** | late_diastolic_peaks_lv_inner_velocity_corrected | 7.861303065 | -2.69383402 | 0.05604010037 | 5.57E-07 | 1.569251171 | 0.05604010037 | 5.57E-07 | 7968 | 8.36E-05 | TRUE | 0.0260855297 | TRUE | 0.02336774358 | TRUE | FALSE | 7.861303065 |
| **cxcl11** | late_diastolic_peaks_lv_inner_velocity_corrected | 7.713696526 | -2.861706618 | 0.06031551543 | 6.12E-08 | 1.423066158 | 0.06031551543 | 6.12E-08 | 8049 | 1.21E-05 | TRUE | 0.002863363155 | TRUE | 0.002565036506 | TRUE | FALSE | 7.713696526 |
| **cxcl11** | late_diastolic_peaks_lv_inner_radial_velocity_corrected | 5.385668677 | -1.67153968 | 0.05960492473 | 8.72E-08 | 1.005466591 | 0.05960492473 | 8.72E-08 | 8049 | 1.65E-05 | TRUE | 0.004084664643 | TRUE | 0.003659093646 | TRUE | FALSE | 5.385668677 |
| **dcn** | diastolic_peaks_lwave_lv_inner_radial_velocity_corrected | -22.99608904 | -0.9335681777 | -0.05642174138 | 3.95E-07 | 4.530916041 | -0.05642174138 | 3.95E-07 | 8068 | 6.25E-05 | TRUE | 0.01850991704 | TRUE | 0.01658141506 | TRUE | FALSE | -22.99608904 |
| **dcxr** | late_diastolic_peaks_lv_inner_velocity_corrected | 12.39597886 | -3.067412048 | 0.07511479312 | 1.74E-11 | 1.840192002 | 0.07511479312 | 1.74E-11 | 7999 | 1.01E-08 | TRUE | 8.14E-07 | TRUE | 7.29E-07 | TRUE | FALSE | 12.39597886 |
| **dcxr** | late_diastolic_peaks_lv_inner_radial_velocity_corrected | 8.12278663 | -1.811603558 | 0.06962274365 | 4.56E-10 | 1.301473173 | 0.06962274365 | 4.56E-10 | 7999 | 1.64E-07 | TRUE | 2.14E-05 | TRUE | 1.91E-05 | TRUE | FALSE | 8.12278663 |
| **dcxr** | late_diastolic_peaks_lv_inner_circumferential_velocity_corrected | 6.490509811 | -1.821802541 | 0.07000357943 | 3.67E-10 | 1.034256525 | 0.07000357943 | 3.67E-10 | 7999 | 1.35E-07 | TRUE | 1.72E-05 | TRUE | 1.54E-05 | TRUE | FALSE | 6.490509811 |
| **defb4a_defb4b** | late_diastolic_peaks_lv_inner_velocity_corrected | 4.561546512 | -2.606002395 | 0.07353943488 | 4.96E-11 | 0.6930971907 | 0.07353943488 | 4.96E-11 | 7968 | 2.32E-08 | TRUE | 2.32E-06 | TRUE | 2.08E-06 | TRUE | FALSE | 4.561546512 |
| **defb4a_defb4b** | late_diastolic_peaks_lv_inner_radial_velocity_corrected | 3.210908947 | -1.464007513 | 0.07324544526 | 5.91E-11 | 0.4898454827 | 0.07324544526 | 5.91E-11 | 7968 | 2.69E-08 | TRUE | 2.77E-06 | TRUE | 2.48E-06 | TRUE | FALSE | 3.210908947 |
| **defb4a_defb4b** | late_diastolic_peaks_lv_inner_circumferential_velocity_corrected | 2.429682377 | -1.588948218 | 0.06961416924 | 4.95E-10 | 0.3901007862 | 0.06961416924 | 4.95E-10 | 7968 | 1.76E-07 | TRUE | 2.32E-05 | TRUE | 2.08E-05 | TRUE | FALSE | 2.429682377 |
| **dkk3** | late_diastolic_peaks_lv_inner_velocity_corrected | -18.13460074 | -3.435112405 | -0.05670109724 | 3.46E-07 | 3.555177187 | -0.05670109724 | 3.46E-07 | 8069 | 5.54E-05 | TRUE | 0.01618631423 | TRUE | 0.01449990261 | TRUE | FALSE | -18.13460074 |
| **dkk3** | late_diastolic_peaks_lv_inner_radial_velocity_corrected | -12.72133465 | -2.046961051 | -0.05629118507 | 4.20E-07 | 2.512158812 | -0.05629118507 | 4.20E-07 | 8069 | 6.55E-05 | TRUE | 0.01965558662 | TRUE | 0.01760772019 | TRUE | FALSE | -12.72133465 |
| **dkk3** | late_diastolic_peaks_lv_inner_circumferential_velocity_corrected | -10.24079532 | -2.060792665 | -0.05699315401 | 3.01E-07 | 1.997323795 | -0.05699315401 | 3.01E-07 | 8069 | 4.90E-05 | TRUE | 0.01408338559 | TRUE | 0.01261607286 | TRUE | FALSE | -10.24079532 |
| **dsg2** | late_diastolic_peaks_lv_inner_velocity_corrected | -25.36449244 | -2.682083493 | -0.07193443405 | 1.31E-10 | 3.94165831 | -0.07193443405 | 1.31E-10 | 7963 | 5.32E-08 | TRUE | 6.12E-06 | TRUE | 5.48E-06 | TRUE | FALSE | -25.36449244 |
| **dsg2** | late_diastolic_peaks_lv_inner_radial_velocity_corrected | -17.31267206 | -1.497717881 | -0.06944298164 | 5.53E-10 | 2.787418916 | -0.06944298164 | 5.53E-10 | 7963 | 1.92E-07 | TRUE | 2.59E-05 | TRUE | 2.32E-05 | TRUE | FALSE | -17.31267206 |
| **dsg2** | late_diastolic_peaks_lv_inner_circumferential_velocity_corrected | -13.54993649 | -1.664087644 | -0.06847451956 | 9.56E-10 | 2.212605096 | -0.06847451956 | 9.56E-10 | 7963 | 3.00E-07 | TRUE | 4.48E-05 | TRUE | 4.01E-05 | TRUE | FALSE | -13.54993649 |
| **erbb2** | diastolic_peaks_lwave_lv_inner_velocity_corrected | 27.77371459 | -0.8134133635 | 0.05635148635 | 4.59E-07 | 5.503025862 | 0.05635148635 | 4.59E-07 | 7998 | 7.09E-05 | TRUE | 0.02147552365 | TRUE | 0.01923804253 | TRUE | FALSE | 27.77371459 |
| **erbb2** | diastolic_peaks_lwave_lv_inner_circumferential_velocity_corrected | 14.42634794 | -0.7946166151 | 0.05564691432 | 6.37E-07 | 2.894712695 | 0.05564691432 | 6.37E-07 | 7998 | 9.26E-05 | TRUE | 0.02981844357 | TRUE | 0.02671173448 | TRUE | FALSE | 14.42634794 |
| **erbb2** | late_diastolic_peaks_lv_inner_velocity_corrected | 30.90314919 | -2.055858975 | 0.083912814 | 5.60E-14 | 4.103708319 | 0.083912814 | 5.60E-14 | 7999 | 7.09E-11 | TRUE | 2.62E-09 | TRUE | 2.35E-09 | TRUE | FALSE | 30.90314919 |
| **erbb2** | late_diastolic_peaks_lv_inner_radial_velocity_corrected | 19.40132095 | -1.174871405 | 0.07451750183 | 2.51E-11 | 2.903356176 | 0.07451750183 | 2.51E-11 | 7999 | 1.32E-08 | TRUE | 1.17E-06 | TRUE | 1.05E-06 | TRUE | FALSE | 19.40132095 |
| **erbb2** | late_diastolic_peaks_lv_inner_circumferential_velocity_corrected | 17.68557797 | -1.245858429 | 0.08547548856 | 1.90E-14 | 2.305270023 | 0.08547548856 | 1.90E-14 | 7999 | 2.86E-11 | TRUE | 8.88E-10 | TRUE | 7.95E-10 | TRUE | FALSE | 17.68557797 |
| **erbb3** | late_diastolic_peaks_lv_inner_radial_velocity_corrected | 19.34593297 | -1.651412954 | 0.05573396567 | 6.83E-07 | 3.892354107 | 0.05573396567 | 6.83E-07 | 7930 | 9.86E-05 | TRUE | 0.03196186318 | TRUE | 0.02863183656 | TRUE | FALSE | 19.34593297 |
| **esm1** | late_diastolic_peaks_lv_inner_velocity_corrected | -15.87010606 | -2.83738728 | -0.05686129802 | 4.04E-07 | 3.129522077 | -0.05686129802 | 4.04E-07 | 7930 | 6.36E-05 | TRUE | 0.01893675482 | TRUE | 0.01696378167 | TRUE | FALSE | -15.87010606 |
| **esm1** | late_diastolic_peaks_lv_inner_radial_velocity_corrected | -10.93362462 | -1.625421656 | -0.05538176151 | 8.02E-07 | 2.21385172 | -0.05538176151 | 8.02E-07 | 7930 | 0.000113488002 | TRUE | 0.03756452866 | TRUE | 0.03365077433 | TRUE | FALSE | -10.93362462 |
| **esm1** | late_diastolic_peaks_lv_inner_circumferential_velocity_corrected | -9.580047777 | -1.678172664 | -0.06108876606 | 5.20E-08 | 1.757974727 | -0.06108876606 | 5.20E-08 | 7930 | 1.07E-05 | TRUE | 0.002436035319 | TRUE | 0.002182230889 | TRUE | FALSE | -9.580047777 |
| **f7** | late_diastolic_peaks_lv_inner_velocity_corrected | 26.28069308 | -1.587843249 | 0.08303328935 | 1.09E-13 | 3.531083819 | 0.08303328935 | 1.09E-13 | 7981 | 1.21E-10 | TRUE | 5.10E-09 | TRUE | 4.57E-09 | TRUE | FALSE | 26.28069308 |
| **f7** | late_diastolic_peaks_lv_inner_radial_velocity_corrected | 18.09718623 | -0.7711695603 | 0.08089439014 | 4.57E-13 | 2.496276898 | 0.08089439014 | 4.57E-13 | 7981 | 4.11E-10 | TRUE | 2.14E-08 | TRUE | 1.92E-08 | TRUE | FALSE | 18.09718623 |
| **f7** | late_diastolic_peaks_lv_inner_circumferential_velocity_corrected | 12.44448658 | -1.096348985 | 0.0698884215 | 4.10E-10 | 1.988539806 | 0.0698884215 | 4.10E-10 | 7981 | 1.50E-07 | TRUE | 1.92E-05 | TRUE | 1.72E-05 | TRUE | FALSE | 12.44448658 |
| **f9** | late_diastolic_peaks_lv_inner_velocity_corrected | 43.54281906 | -1.207910371 | 0.08686338925 | 7.84E-15 | 5.59308584 | 0.08686338925 | 7.84E-15 | 7974 | 1.22E-11 | TRUE | 3.67E-10 | TRUE | 3.29E-10 | TRUE | FALSE | 43.54281906 |
| **f9** | late_diastolic_peaks_lv_inner_radial_velocity_corrected | 27.94500994 | -0.5942854702 | 0.07887769094 | 1.75E-12 | 3.955589283 | 0.07887769094 | 1.75E-12 | 7974 | 1.30E-09 | TRUE | 8.17E-08 | TRUE | 7.32E-08 | TRUE | FALSE | 27.94500994 |
| **f9** | late_diastolic_peaks_lv_inner_circumferential_velocity_corrected | 22.53462308 | -0.8510121699 | 0.07991033822 | 8.93E-13 | 3.148274628 | 0.07991033822 | 8.93E-13 | 7974 | 7.33E-10 | TRUE | 4.18E-08 | TRUE | 3.74E-08 | TRUE | FALSE | 22.53462308 |
| **fabp4** | systolic_peaks_rv_inner_velocity_corrected | -64.71727485 | -6.024428768 | -0.06199791271 | 2.27E-08 | 11.56616505 | -0.06199791271 | 2.27E-08 | 8116 | 5.09E-06 | TRUE | 0.001064043653 | TRUE | 0.0009531836048 | TRUE | FALSE | -64.71727485 |
| **fabp4** | systolic_peaks_rv_inner_circumferential_velocity_corrected | -38.34634173 | -2.825760766 | -0.05818765255 | 1.56E-07 | 7.303636233 | -0.05818765255 | 1.56E-07 | 8116 | 2.76E-05 | TRUE | 0.007288957896 | TRUE | 0.006529539595 | TRUE | FALSE | -38.34634173 |
| **fabp4** | late_diastolic_peaks_lv_inner_velocity_corrected | 11.09258839 | -1.361812003 | 0.07198992649 | 9.54E-11 | 1.711105695 | 0.07198992649 | 9.54E-11 | 8069 | 3.95E-08 | TRUE | 4.47E-06 | TRUE | 4.00E-06 | TRUE | FALSE | 11.09258839 |
| **fabp4** | late_diastolic_peaks_lv_inner_radial_velocity_corrected | 7.86975334 | -0.5785480762 | 0.07228100829 | 8.01E-11 | 1.209047659 | 0.07228100829 | 8.01E-11 | 8069 | 3.41E-08 | TRUE | 3.75E-06 | TRUE | 3.36E-06 | TRUE | FALSE | 7.86975334 |
| **fabp4** | late_diastolic_peaks_lv_inner_circumferential_velocity_corrected | 5.412157075 | -1.024995052 | 0.0625193948 | 1.90E-08 | 0.9619426434 | 0.0625193948 | 1.90E-08 | 8069 | 4.37E-06 | TRUE | 0.000890904859 | TRUE | 0.000798083709 | TRUE | FALSE | 5.412157075 |
| **fcgr2b** | late_diastolic_peaks_lv_inner_velocity_corrected | 9.832021959 | -3.14123022 | 0.05799300695 | 1.86E-07 | 1.884428422 | 0.05799300695 | 1.86E-07 | 8069 | 3.22E-05 | TRUE | 0.00870109547 | TRUE | 0.007794550086 | TRUE | FALSE | 9.832021959 |
| **fcgr2b** | late_diastolic_peaks_lv_inner_radial_velocity_corrected | 6.474779822 | -1.839882503 | 0.05404830041 | 1.19E-06 | 1.331838826 | 0.05404830041 | 1.19E-06 | 8069 | 0.0001615154732 | TRUE | 0.05556132278 | FALSE | 0.04977252747 | TRUE | FALSE | 6.474779822 |

| | | | | | | | | | | | | | | | | | |
|---|---|---|---|---|---|---|---|---|---|---|---|---|---|---|---|---|---|
| **fcgr2b** | late_diastolic_peaks_lv_inner_circumferential_velocity_corrected | 5.310822669 | -1.89430755 | 0.05575709375 | 5.40E-07 | 1.058837984 | 0.05575709375 | 5.40E-07 | 8069 | 8.19E-05 | TRUE | 0.02526447355 | TRUE | 0.02263223121 | TRUE | FALSE | 5.310822669 |
| **fgf21** | late_diastolic_peaks_lv_inner_velocity_corrected | 5.975532798 | -2.076568664 | 0.07450882015 | 2.07E-11 | 0.8904387557 | 0.07450882015 | 2.07E-11 | 8069 | 1.15E-08 | TRUE | 9.68E-07 | TRUE | 8.67E-07 | TRUE | FALSE | 5.975532798 |
| **fgf21** | late_diastolic_peaks_lv_inner_radial_velocity_corrected | 4.127524225 | -1.105171713 | 0.07283580629 | 5.74E-11 | 0.6292650709 | 0.07283580629 | 5.74E-11 | 8069 | 2.63E-08 | TRUE | 2.69E-06 | TRUE | 2.41E-06 | TRUE | FALSE | 4.127524225 |
| **fgf21** | late_diastolic_peaks_lv_inner_circumferential_velocity_corrected | 2.961248084 | -1.365744774 | 0.06572209295 | 3.44E-09 | 0.5005729794 | 0.06572209295 | 3.44E-09 | 8069 | 9.47E-07 | TRUE | 0.0001610061165 | TRUE | 0.0001442312918 | TRUE | FALSE | 2.961248084 |
| **fgfr2** | late_diastolic_peaks_lv_inner_velocity_corrected | 24.46956573 | -1.978394486 | 0.05532418622 | 7.38E-07 | 4.938351781 | 0.05532418622 | 7.38E-07 | 7999 | 0.0001053534586 | TRUE | 0.03455593442 | TRUE | 0.030955638 | TRUE | FALSE | 24.46956573 |
| **furin** | late_diastolic_peaks_lv_inner_velocity_corrected | 21.89807727 | -1.311309078 | 0.08460284208 | 3.48E-14 | 2.884015779 | 0.08460284208 | 3.48E-14 | 7999 | 4.79E-11 | TRUE | 1.63E-09 | TRUE | 1.46E-09 | TRUE | FALSE | 21.89807727 |
| **furin** | late_diastolic_peaks_lv_inner_radial_velocity_corrected | 14.92102736 | -0.6166070039 | 0.08154158247 | 2.80E-13 | 2.039423426 | 0.08154158247 | 2.80E-13 | 7999 | 2.62E-10 | TRUE | 1.31E-08 | TRUE | 1.17E-08 | TRUE | FALSE | 14.92102736 |
| **furin** | late_diastolic_peaks_lv_inner_circumferential_velocity_corrected | 9.954564424 | -1.01930983 | 0.06845387195 | 8.87E-10 | 1.622335855 | 0.06845387195 | 8.87E-10 | 7999 | 2.82E-07 | TRUE | 4.15E-05 | TRUE | 3.72E-05 | TRUE | FALSE | 9.954564424 |
| **furin** | late_diastolic_peaks_rv_freewall_velocity_filtered_50_corrected | 0.6228483516 | 0.08048174041 | 0.05721010109 | 3.05E-07 | 0.1215441802 | 0.05721010109 | 3.05E-07 | 7999 | 4.95E-05 | TRUE | 0.01429629082 | TRUE | 0.01280679602 | TRUE | FALSE | 0.6228483516 |
| **gfra1** | late_diastolic_peaks_lv_inner_velocity_corrected | 19.5781545 | -1.883568607 | 0.05463574864 | 9.07E-07 | 3.983727803 | 0.05463574864 | 9.07E-07 | 8069 | 0.0001260681989 | TRUE | 0.04248498301 | TRUE | 0.03805857885 | TRUE | FALSE | 19.5781545 |
| **ggt1** | late_diastolic_peaks_lv_inner_velocity_corrected | 16.28688979 | -3.052144738 | 0.08807629482 | 3.57E-15 | 2.0656173 | 0.08807629482 | 3.57E-15 | 7954 | 6.69E-12 | TRUE | 1.67E-10 | TRUE | 1.50E-10 | TRUE | FALSE | 16.28688979 |
| **ggt1** | late_diastolic_peaks_lv_inner_radial_velocity_corrected | 10.37427568 | -1.768353792 | 0.07933773218 | 1.38E-12 | 1.461735076 | 0.07933773218 | 1.38E-12 | 7954 | 1.06E-09 | TRUE | 6.47E-08 | TRUE | 5.80E-08 | TRUE | FALSE | 10.37427568 |
| **ggt1** | late_diastolic_peaks_lv_inner_circumferential_velocity_corrected | 9.228569289 | -1.806932773 | 0.08885227136 | 2.04E-15 | 1.160129677 | 0.08885227136 | 2.04E-15 | 7954 | 4.16E-12 | TRUE | 9.56E-11 | TRUE | 8.56E-11 | TRUE | FALSE | 9.228569289 |
| **gsta1** | late_diastolic_peaks_lv_inner_velocity_corrected | 9.873529595 | -2.737844621 | 0.09346923618 | 3.99E-17 | 1.170960561 | 0.09346923618 | 3.99E-17 | 8069 | 1.44E-13 | TRUE | 1.87E-12 | TRUE | 1.67E-12 | TRUE | FALSE | 9.873529595 |
| **gsta1** | late_diastolic_peaks_lv_inner_radial_velocity_corrected | 6.476130238 | -1.575242067 | 0.08676330462 | 5.84E-15 | 0.8279090318 | 0.08676330462 | 5.84E-15 | 8069 | 9.77E-12 | TRUE | 2.74E-10 | TRUE | 2.45E-10 | TRUE | FALSE | 6.476130238 |
| **gsta1** | late_diastolic_peaks_lv_inner_circumferential_velocity_corrected | 5.178171049 | -1.682415061 | 0.08725256328 | 4.11E-15 | 0.6582376211 | 0.08725256328 | 4.11E-15 | 8069 | 7.40E-12 | TRUE | 1.93E-10 | TRUE | 1.72E-10 | TRUE | FALSE | 5.178171049 |
| **gsta3** | late_diastolic_peaks_lv_inner_velocity_corrected | 9.06243284 | -3.401598842 | 0.07408600973 | 2.68E-11 | 1.358180283 | 0.07408600973 | 2.68E-11 | 8069 | 1.39E-08 | TRUE | 1.26E-06 | TRUE | 1.12E-06 | TRUE | FALSE | 9.06243284 |
| **gsta3** | late_diastolic_peaks_lv_inner_radial_velocity_corrected | 5.959588445 | -2.011084864 | 0.06894962715 | 5.65E-10 | 0.9600492842 | 0.06894962715 | 5.65E-10 | 8069 | 1.94E-07 | TRUE | 2.64E-05 | TRUE | 2.37E-05 | TRUE | FALSE | 5.959588445 |
| **gsta3** | late_diastolic_peaks_lv_inner_circumferential_velocity_corrected | 4.750093789 | -2.030436965 | 0.06911926776 | 5.12E-10 | 0.7633208522 | 0.06911926776 | 5.12E-10 | 8069 | 1.80E-07 | TRUE | 2.40E-05 | TRUE | 2.15E-05 | TRUE | FALSE | 4.750093789 |
| **guca2a** | late_diastolic_peaks_lv_inner_radial_velocity_corrected | -11.71199 | -2.018972062 | -0.05491116128 | 9.88E-07 | 2.39123893 | -0.05491116128 | 9.88E-07 | 7934 | 0.0001364359737 | TRUE | 0.04625179508 | TRUE | 0.04143293618 | TRUE | FALSE | -11.71199 |
| **gusb** | late_diastolic_peaks_lv_inner_velocity_corrected | 11.30625637 | -2.392255104 | 0.07633752158 | 6.59E-12 | 1.644203755 | 0.07633752158 | 6.59E-12 | 8069 | 4.34E-09 | TRUE | 3.09E-07 | TRUE | 2.77E-07 | TRUE | FALSE | 11.30625637 |
| **gusb** | late_diastolic_peaks_lv_inner_radial_velocity_corrected | 7.078420514 | -1.370280621 | 0.0676362991 | 1.19E-09 | 1.162531794 | 0.0676362991 | 1.19E-09 | 8069 | 3.71E-07 | TRUE | 5.57E-05 | TRUE | 4.99E-05 | TRUE | FALSE | 7.078420514 |
| **gusb** | late_diastolic_peaks_lv_inner_circumferential_velocity_corrected | 5.958298799 | -1.499321729 | 0.07160560958 | 1.20E-10 | 0.9240659058 | 0.07160560958 | 1.20E-10 | 8069 | 4.92E-08 | TRUE | 5.61E-06 | TRUE | 5.03E-06 | TRUE | FALSE | 5.958298799 |
| **hao1** | late_diastolic_peaks_lv_inner_velocity_corrected | 4.603046947 | -3.088524607 | 0.06424558783 | 7.65E-09 | 0.7960639179 | 0.06424558783 | 7.65E-09 | 8069 | 1.93E-06 | TRUE | 0.0003579149576 | TRUE | 0.0003206246929 | TRUE | FALSE | 4.603046947 |
| **hao1** | late_diastolic_peaks_lv_inner_radial_velocity_corrected | 3.044916628 | -1.80508124 | 0.06014472702 | 6.42E-08 | 0.5626455512 | 0.06014472702 | 6.42E-08 | 8069 | 1.26E-05 | TRUE | 0.00300517338 | TRUE | 0.002692071878 | TRUE | FALSE | 3.044916628 |
| **hao1** | late_diastolic_peaks_lv_inner_circumferential_velocity_corrected | 2.323835674 | -1.86694171 | 0.05773097045 | 2.11E-07 | 0.4474202096 | 0.05773097045 | 2.11E-07 | 8069 | 3.59E-05 | TRUE | 0.009879025219 | TRUE | 0.008849754279 | TRUE | FALSE | 2.323835674 |
| **hgf** | diastolic_peaks_lwave_lv_inner_velocity_corrected | 23.84437106 | 0.2882383256 | 0.06679634727 | 2.05E-09 | 3.973463173 | 0.06679634727 | 2.05E-09 | 8037 | 5.99E-07 | TRUE | 9.58E-05 | TRUE | 8.59E-05 | TRUE | FALSE | 23.84437106 |
| **hgf** | diastolic_peaks_lwave_lv_inner_circumferential_velocity_corrected | 12.89597673 | -0.2160236336 | 0.06870569599 | 7.02E-10 | 2.089012758 | 0.06870569599 | 7.02E-10 | 8037 | 2.31E-07 | TRUE | 3.28E-05 | TRUE | 2.94E-05 | TRUE | FALSE | 12.89597673 |
| **hgf** | late_diastolic_peaks_lv_inner_velocity_corrected | 25.45888654 | -0.8383659542 | 0.09528747256 | 1.12E-17 | 2.966900598 | 0.09528747256 | 1.12E-17 | 8038 | 5.81E-14 | TRUE | 5.23E-13 | TRUE | 4.69E-13 | TRUE | FALSE | 25.45888654 |
| **hgf** | late_diastolic_peaks_lv_inner_radial_velocity_corrected | 15.94050636 | -0.3936068608 | 0.08443576682 | 3.39E-14 | 2.098466414 | 0.08443576682 | 3.39E-14 | 8038 | 4.79E-11 | TRUE | 1.59E-09 | TRUE | 1.42E-09 | TRUE | FALSE | 15.94050636 |
| **hgf** | late_diastolic_peaks_rv_inner_circumferential_velocity_corrected | 10.39315711 | -0.936850959 | 0.06089218821 | 4.67E-08 | 1.900462478 | 0.06089218821 | 4.67E-08 | 8038 | 9.76E-06 | TRUE | 0.002185500441 | TRUE | 0.001957798614 | TRUE | FALSE | 10.39315711 |
| **hgf** | late_diastolic_peaks_lv_inner_circumferential_velocity_corrected | 14.66687878 | -0.5985763017 | 0.09774040021 | 1.60E-18 | 1.665939423 | 0.09774040021 | 1.60E-18 | 8038 | 1.07E-14 | TRUE | 7.48E-14 | TRUE | 6.70E-14 | TRUE | FALSE | 14.66687878 |
| **hnmt** | late_diastolic_peaks_lv_inner_velocity_corrected | 11.95341354 | -1.887813631 | 0.05917255482 | 1.08E-07 | 2.248122427 | 0.05917255482 | 1.08E-07 | 8048 | 1.99E-05 | TRUE | 0.005069554581 | TRUE | 0.004541370364 | TRUE | FALSE | 11.95341354 |
| **ifnlr1** | late_diastolic_peaks_lv_inner_velocity_corrected | 16.2390863 | -2.861128134 | 0.06755569823 | 1.56E-09 | 2.686103105 | 0.06755569823 | 1.56E-09 | 7974 | 4.67E-07 | TRUE | 7.28E-05 | TRUE | 6.52E-05 | TRUE | FALSE | 16.2390863 |
| **ifnlr1** | late_diastolic_peaks_lv_inner_radial_velocity_corrected | 10.22057648 | -1.696690075 | 0.06018499375 | 7.52E-08 | 1.898521595 | 0.06018499375 | 7.52E-08 | 7974 | 1.45E-05 | TRUE | 0.003518491254 | TRUE | 0.003151908446 | TRUE | FALSE | 10.22057648 |
| **igfbp1** | late_diastolic_peaks_lv_inner_velocity_corrected | -6.658115119 | -3.158857928 | -0.09551250337 | 8.66E-18 | 0.7732540003 | -0.09551250337 | 8.66E-18 | 8055 | 5.07E-14 | TRUE | 4.05E-13 | TRUE | 3.63E-13 | TRUE | FALSE | -6.658115119 |
| **igfbp1** | late_diastolic_peaks_lv_inner_radial_velocity_corrected | -4.436215153 | -1.847222031 | -0.09006519529 | 5.56E-16 | 0.5466484631 | -0.09006519529 | 5.56E-16 | 8055 | 1.30E-12 | TRUE | 2.60E-11 | TRUE | 2.33E-11 | TRUE | FALSE | -4.436215153 |
| **igfbp1** | late_diastolic_peaks_lv_inner_circumferential_velocity_corrected | -3.614780956 | -1.882112716 | -0.09231191137 | 1.03E-16 | 0.4344972636 | -0.09231191137 | 1.03E-16 | 8055 | 3.01E-13 | TRUE | 4.82E-12 | TRUE | 4.31E-12 | TRUE | FALSE | -3.614780956 |
| **igfbp1** | late_diastolic_peaks_rv_freewall_velocity_filtered_50_corrected | -0.1781421448 | 0.03132968366 | -0.06072969863 | 4.91E-08 | 0.03262754228 | -0.06072969863 | 4.91E-08 | 8055 | 1.02E-05 | TRUE | 0.002297126659 | TRUE | 0.002057794775 | TRUE | FALSE | -0.1781421448 |
| **igfbp2** | late_diastolic_peaks_lv_inner_velocity_corrected | -11.2005695 | -3.913166663 | -0.0809576575 | 3.47E-13 | 1.537033417 | -0.0809576575 | 3.47E-13 | 8051 | 3.18E-10 | TRUE | 1.62E-08 | TRUE | 1.45E-08 | TRUE | FALSE | -11.2005695 |
| **igfbp2** | late_diastolic_peaks_lv_inner_radial_velocity_corrected | -8.068870962 | -2.396497106 | -0.08255662944 | 1.18E-13 | 1.085687402 | -0.08255662944 | 1.18E-13 | 8051 | 1.25E-10 | TRUE | 5.52E-09 | TRUE | 4.95E-09 | TRUE | FALSE | -8.068870962 |
| **igfbp2** | late_diastolic_peaks_lv_inner_circumferential_velocity_corrected | -5.81218574 | -2.301804765 | -0.07477086175 | 1.85E-11 | 0.8640099174 | -0.07477086175 | 1.85E-11 | 8051 | 1.06E-08 | TRUE | 8.66E-07 | TRUE | 7.76E-07 | TRUE | FALSE | -5.81218574 |
| **igsf3** | late_diastolic_peaks_lv_inner_velocity_corrected | 19.22864591 | -2.570767122 | 0.0686322959 | 6.77E-10 | 3.111994932 | 0.0686322959 | 6.77E-10 | 8069 | 2.25E-07 | TRUE | 3.17E-05 | TRUE | 2.84E-05 | TRUE | FALSE | 19.22864591 |
| **igsf3** | late_diastolic_peaks_lv_inner_radial_velocity_corrected | 12.22662451 | -1.476664149 | 0.06176047815 | 2.82E-08 | 2.199935947 | 0.06176047815 | 2.82E-08 | 8069 | 6.17E-06 | TRUE | 0.001320381254 | TRUE | 0.001182814032 | TRUE | FALSE | 12.22662451 |
| **igsf3** | late_diastolic_peaks_lv_inner_circumferential_velocity_corrected | 9.374450692 | -1.615063172 | 0.05955677808 | 8.61E-08 | 1.749391504 | 0.05955677808 | 8.61E-08 | 8069 | 1.64E-05 | TRUE | 0.004032817454 | TRUE | 0.003612648285 | TRUE | FALSE | 9.374450692 |
| **il10rb** | late_diastolic_peaks_lv_inner_velocity_corrected | 19.67629627 | -1.956543432 | 0.05528100443 | 7.91E-07 | 3.981829511 | 0.05528100443 | 7.91E-07 | 7968 | 0.0001122101893 | TRUE | 0.03702936246 | TRUE | 0.03317136576 | TRUE | FALSE | 19.67629627 |
| **il18** | late_diastolic_peaks_lv_inner_velocity_corrected | 11.35467371 | -2.329811571 | 0.05610086123 | 6.02E-07 | 2.273419562 | 0.05610086123 | 6.02E-07 | 7903 | 8.87E-05 | TRUE | 0.02820260788 | TRUE | 0.02526424867 | TRUE | FALSE | 11.35467371 |
| **il18r1** | late_diastolic_peaks_lv_inner_velocity_corrected | 21.14014443 | -1.688995522 | 0.07418771993 | 2.75E-11 | 3.169987301 | 0.07418771993 | 2.75E-11 | 8038 | 1.41E-08 | TRUE | 1.29E-06 | TRUE | 1.15E-06 | TRUE | FALSE | 21.14014443 |
| **il18r1** | late_diastolic_peaks_lv_inner_radial_velocity_corrected | 13.03168462 | -0.9390193207 | 0.06472209577 | 6.33E-09 | 2.24138518 | 0.06472209577 | 6.33E-09 | 8038 | 1.65E-06 | TRUE | 0.0002962703357 | TRUE | 0.0002654026701 | TRUE | FALSE | 13.03168462 |
| **il18r1** | late_diastolic_peaks_lv_inner_circumferential_velocity_corrected | 11.80901672 | -1.111763913 | 0.07378665444 | 3.51E-11 | 1.78045283 | 0.07378665444 | 3.51E-11 | 8038 | 1.73E-08 | TRUE | 1.64E-06 | TRUE | 1.47E-06 | TRUE | FALSE | 11.80901672 |
| **il1rn** | late_diastolic_peaks_lv_inner_velocity_corrected | 13.03194227 | -2.097438842 | 0.07664283866 | 8.01E-12 | 1.902841906 | 0.07664283866 | 8.01E-12 | 7940 | 5.00E-09 | TRUE | 3.75E-07 | TRUE | 3.36E-07 | TRUE | FALSE | 13.03194227 |
| **il1rn** | late_diastolic_peaks_lv_inner_radial_velocity_corrected | 8.067918378 | -1.15679189 | 0.06708867438 | 2.18E-09 | 1.346719585 | 0.06708867438 | 2.18E-09 | 7940 | 6.26E-07 | TRUE | 0.000102023448 | TRUE | 9.14E-05 | TRUE | FALSE | 8.067918378 |
| **il1rn** | late_diastolic_peaks_lv_inner_circumferential_velocity_corrected | 7.046230733 | -1.30752659 | 0.07374709494 | 4.73E-11 | 1.06947882 | 0.07374709494 | 4.73E-11 | 7940 | 2.26E-08 | TRUE | 2.21E-06 | TRUE | 1.98E-06 | TRUE | FALSE | 7.046230733 |
| **il6** | late_diastolic_peaks_lv_inner_velocity_corrected | 8.871957885 | -2.372053428 | 0.06655130028 | 2.18E-09 | 1.48095888 | 0.06655130028 | 2.18E-09 | 8069 | 6.26E-07 | TRUE | 0.0001020216522 | TRUE | 9.14E-05 | TRUE | FALSE | 8.871957885 |
| **il6** | late_diastolic_peaks_lv_inner_radial_velocity_corrected | 5.99935532 | -1.320132477 | 0.06368921667 | 1.03E-08 | 1.046646533 | 0.06368921667 | 1.03E-08 | 8069 | 2.49E-06 | TRUE | 0.0004814550691 | TRUE | 0.0004312934691 | TRUE | FALSE | 5.99935532 |
| **il6** | late_diastolic_peaks_lv_inner_circumferential_velocity_corrected | 4.341046185 | -1.516858455 | 0.05796115361 | 1.89E-07 | 0.8324738955 | 0.05796115361 | 1.89E-07 | 8069 | 3.26E-05 | TRUE | 0.008836685313 | TRUE | 0.007916013162 | TRUE | FALSE | 4.341046185 |
| **inhbc** | late_diastolic_peaks_lv_inner_velocity_corrected | 16.32665475 | -0.8749036325 | 0.07693585469 | 6.22E-12 | 2.37134899 | 0.07693585469 | 6.22E-12 | 7963 | 4.22E-09 | TRUE | 2.91E-07 | TRUE | 2.61E-07 | TRUE | FALSE | 16.32665475 |
| **inhbc** | late_diastolic_peaks_lv_inner_radial_velocity_corrected | 10.96806789 | -0.2885001704 | 0.07309965139 | 6.54E-11 | 1.677133222 | 0.07309965139 | 6.54E-11 | 7963 | 2.94E-08 | TRUE | 3.06E-06 | TRUE | 2.74E-06 | TRUE | FALSE | 10.96806789 |
| **inhbc** | late_diastolic_peaks_lv_inner_circumferential_velocity_corrected | 7.769515056 | -0.8302341218 | 0.06523889035 | 5.65E-09 | 1.331917606 | 0.06523889035 | 5.65E-09 | 7963 | 1.49E-06 | TRUE | 0.0002643085498 | TRUE | 0.0002367709028 | TRUE | FALSE | 7.769515056 |
| **itga5** | late_diastolic_peaks_lv_inner_velocity_corrected | 23.68195392 | -1.886046913 | 0.06266535317 | 2.19E-08 | 4.227190538 | 0.06266535317 | 2.19E-08 | 7963 | 4.92E-06 | TRUE | 0.001023344979 | TRUE | 0.0009167252236 | TRUE | FALSE | 23.68195392 |
| **itga5** | late_diastolic_peaks_lv_inner_radial_velocity_corrected | 15.386206 | -0.9952579317 | 0.0575831084 | 2.72E-07 | 2.989726147 | 0.0575831084 | 2.72E-07 | 7963 | 4.51E-05 | TRUE | 0.01273099186 | TRUE | 0.01140458164 | TRUE | FALSE | 15.386206 |
| **kdr** | late_diastolic_peaks_lv_inner_velocity_corrected | 27.49400936 | -2.419141423 | 0.07403812774 | 3.36E-11 | 4.141193293 | 0.07403812774 | 3.36E-11 | 7999 | 1.67E-08 | TRUE | 1.57E-06 | TRUE | 1.41E-06 | TRUE | FALSE | 27.49400936 |
| **kdr** | late_diastolic_peaks_lv_inner_radial_velocity_corrected | 19.126479 | -1.363081032 | 0.07285407015 | 6.87E-11 | 2.927939071 | 0.07285407015 | 6.87E-11 | 7999 | 3.01E-08 | TRUE | 3.22E-06 | TRUE | 2.88E-06 | TRUE | FALSE | 19.126479 |
| **kdr** | late_diastolic_peaks_lv_inner_circumferential_velocity_corrected | 13.85192584 | -1.493991581 | 0.06639330639 | 2.79E-09 | 2.327892902 | 0.06639330639 | 2.79E-09 | 7999 | 7.81E-07 | TRUE | 0.0001304912485 | TRUE | 0.0001168956915 | TRUE | FALSE | 13.85192584 |
| **krt18** | late_diastolic_peaks_lv_inner_velocity_corrected | 8.237707463 | -2.816575832 | 0.07253302376 | 6.89E-11 | 1.261156894 | 0.07253302376 | 6.89E-11 | 8069 | 3.01E-08 | TRUE | 3.23E-06 | TRUE | 2.89E-06 | TRUE | FALSE | 8.237707463 |

| | | | | | | | | | | | | | | | | | |
|---|---|---|---|---|---|---|---|---|---|---|---|---|---|---|---|---|---|
| krt18 | late_diastolic_peaks_lv_inner_radial_velocity_corrected | 5.241584409 | -1.632830282 | 0.06531549926 | 4.29E-09 | 0.8915833467 | 0.06531549926 | 4.29E-09 | 8069 | 1.17E-06 | TRUE | 0.0002009709153 | TRUE | 0.0001800322581 | TRUE | FALSE | 5.241584409 |
| krt18 | late_diastolic_peaks_lv_inner_circumferential_velocity_corrected | 4.303662778 | -1.724316675 | 0.06744863922 | 1.32E-09 | 0.708792131 | 0.06744863922 | 1.32E-09 | 8069 | 4.07E-07 | TRUE | 6.19E-05 | TRUE | 5.54E-05 | TRUE | FALSE | 4.303662778 |
| kynu | late_diastolic_peaks_lv_inner_velocity_corrected | 14.8835867 | -2.395986338 | 0.07240688002 | 8.08E-11 | 2.28700102 | 0.07240688002 | 8.08E-11 | 8038 | 3.41E-08 | TRUE | 3.78E-06 | TRUE | 3.39E-06 | TRUE | FALSE | 14.8835867 |
| kynu | late_diastolic_peaks_lv_inner_radial_velocity_corrected | 9.417310583 | -1.364809706 | 0.06483761979 | 5.95E-09 | 1.616832659 | 0.06483761979 | 5.95E-09 | 8038 | 1.56E-06 | TRUE | 0.0002784364765 | TRUE | 0.0002494268761 | TRUE | FALSE | 9.417310583 |
| kynu | late_diastolic_peaks_lv_inner_circumferential_velocity_corrected | 7.04654518 | -1.559127163 | 0.06103633975 | 4.34E-08 | 1.285456298 | 0.06103633975 | 4.34E-08 | 8038 | 9.15E-06 | TRUE | 0.002031415115 | TRUE | 0.001819767053 | TRUE | FALSE | 7.04654518 |
| lamp2 | late_diastolic_peaks_lv_inner_velocity_corrected | 27.8125356 | -3.152783365 | 0.05710914349 | 5.14E-07 | 5.534433547 | 0.05710914349 | 5.14E-07 | 7720 | 7.83E-05 | TRUE | 0.02405282597 | TRUE | 0.02154682216 | TRUE | FALSE | 27.8125356 |
| lamp2 | late_diastolic_peaks_lv_inner_radial_velocity_corrected | 22.28414792 | -1.722443509 | 0.06474785816 | 1.24E-08 | 3.909362854 | 0.06474785816 | 1.24E-08 | 7720 | 2.98E-06 | TRUE | 0.000580873062 | TRUE | 0.0005203533499 | TRUE | FALSE | 22.28414792 |
| ldlr | late_diastolic_peaks_lv_inner_velocity_corrected | 15.42288211 | -1.753480431 | 0.09074106127 | 4.01E-16 | 1.891136774 | 0.09074106127 | 4.01E-16 | 8013 | 9.88E-13 | TRUE | 1.88E-11 | TRUE | 1.68E-11 | TRUE | FALSE | 15.42288211 |
| ldlr | late_diastolic_peaks_lv_inner_radial_velocity_corrected | 11.17980667 | -0.8989708084 | 0.09313734164 | 6.59E-17 | 1.335288825 | 0.09313734164 | 6.59E-17 | 8013 | 2.06E-13 | TRUE | 3.09E-12 | TRUE | 2.77E-12 | TRUE | FALSE | 11.17980667 |
| ldlr | late_diastolic_peaks_lv_inner_circumferential_velocity_corrected | 7.455498736 | -1.18033118 | 0.07797089474 | 2.77E-12 | 1.065066316 | 0.07797089474 | 2.77E-12 | 8013 | 1.93E-09 | TRUE | 1.29E-07 | TRUE | 1.16E-07 | TRUE | FALSE | 7.455498736 |
| ldlr | late_diastolic_peaks_rv_freewall_velocity_filtered_50_corrected | 0.4007396946 | 0.061428929 | 0.05594843667 | 5.40E-07 | 0.07990063941 | 0.05594843667 | 5.40E-07 | 8013 | 8.19E-05 | TRUE | 0.02529874713 | TRUE | 0.02266293391 | TRUE | FALSE | 0.4007396946 |
| lep | systolic_peaks_rv_inner_velocity_corrected | -36.53696228 | -7.296395837 | -0.06421725231 | 7.02E-09 | 6.303269564 | -0.06421725231 | 7.02E-09 | 8116 | 1.80E-06 | TRUE | 0.0003288470799 | TRUE | 0.0002945853247 | TRUE | FALSE | -36.53696228 |
| lep | systolic_peaks_rv_inner_radial_velocity_corrected | -19.62201012 | -3.460819153 | -0.05506757392 | 6.91E-07 | 3.9497587 | -0.05506757392 | 6.91E-07 | 8116 | 9.95E-05 | TRUE | 0.03233312506 | TRUE | 0.02896441759 | TRUE | FALSE | -19.62201012 |
| lep | systolic_peaks_rv_inner_circumferential_velocity_corrected | -21.84555504 | -3.641221982 | -0.06081803972 | 4.18E-08 | 3.980232883 | -0.06081803972 | 4.18E-08 | 8116 | 8.85E-06 | TRUE | 0.001954853723 | TRUE | 0.0017511824 | TRUE | FALSE | -21.84555504 |
| lep | late_diastolic_peaks_lv_inner_velocity_corrected | 6.067328977 | -1.206297215 | 0.07220304911 | 8.40E-11 | 0.9331489385 | 0.07220304911 | 8.40E-11 | 8069 | 3.51E-08 | TRUE | 3.93E-06 | TRUE | 3.52E-06 | TRUE | FALSE | 6.067328977 |
| lep | late_diastolic_peaks_lv_inner_radial_velocity_corrected | 4.534558663 | -0.3956715077 | 0.07636902899 | 6.46E-12 | 0.6591610967 | 0.07636902899 | 6.46E-12 | 8069 | 4.32E-09 | TRUE | 3.03E-07 | TRUE | 2.71E-07 | TRUE | FALSE | 4.534558663 |
| lep | late_diastolic_peaks_lv_inner_circumferential_velocity_corrected | 2.953039122 | -0.9514066352 | 0.06255078089 | 1.87E-08 | 0.5246010548 | 0.06255078089 | 1.87E-08 | 8069 | 4.32E-06 | TRUE | 0.0008764397218 | TRUE | 0.0007851256583 | TRUE | FALSE | 2.953039122 |
| lgals9 | late_diastolic_peaks_lv_inner_velocity_corrected | 20.2953096 | -1.294308747 | 0.06879015337 | 8.22E-10 | 3.301078413 | 0.06879015337 | 8.22E-10 | 7952 | 2.65E-07 | TRUE | 3.85E-05 | TRUE | 3.45E-05 | TRUE | FALSE | 20.2953096 |
| lgals9 | late_diastolic_peaks_lv_inner_radial_velocity_corrected | 13.08557628 | -0.6357509943 | 0.06271570578 | 2.18E-08 | 2.335485604 | 0.06271570578 | 2.18E-08 | 7952 | 4.92E-06 | TRUE | 0.001019607378 | TRUE | 0.0009133770347 | TRUE | FALSE | 13.08557628 |
| lilra5 | late_diastolic_peaks_rv_inner_velocity_corrected | 16.94859859 | -0.7526966013 | 0.05520085413 | 7.07E-07 | 3.414741153 | 0.05520085413 | 7.07E-07 | 8062 | 0.0001015347652 | TRUE | 0.03310033344 | TRUE | 0.02965169245 | TRUE | FALSE | 16.94859859 |
| lilra5 | late_diastolic_peaks_lv_inner_velocity_corrected | 23.86681696 | -1.082356635 | 0.08124631566 | 2.75E-13 | 3.261258115 | 0.08124631566 | 2.75E-13 | 8062 | 2.62E-10 | TRUE | 1.29E-08 | TRUE | 1.15E-08 | TRUE | FALSE | 23.86681696 |
| lilra5 | late_diastolic_peaks_lv_inner_radial_velocity_corrected | 15.36511035 | -0.5129265722 | 0.07402978731 | 2.83E-11 | 2.305517018 | 0.07402978731 | 2.83E-11 | 8062 | 1.44E-08 | TRUE | 1.33E-06 | TRUE | 1.19E-06 | TRUE | FALSE | 15.36511035 |
| lilra5 | late_diastolic_peaks_rv_inner_circumferential_velocity_corrected | 11.10141517 | -0.8997983327 | 0.05911410057 | 1.09E-07 | 2.088139903 | 0.05911410057 | 1.09E-07 | 8062 | 1.99E-05 | TRUE | 0.005087439158 | TRUE | 0.004557391591 | TRUE | FALSE | 11.10141517 |
| lilra5 | late_diastolic_peaks_lv_inner_circumferential_velocity_corrected | 12.57184406 | -0.8250731412 | 0.07617867189 | 7.44E-12 | 1.832880921 | 0.07617867189 | 7.44E-12 | 8062 | 4.77E-09 | TRUE | 3.48E-07 | TRUE | 3.12E-07 | TRUE | FALSE | 12.57184406 |
| lpl | late_diastolic_peaks_lv_inner_velocity_corrected | -9.764487486 | -3.271474635 | -0.05469870866 | 8.82E-07 | 1.984566627 | -0.05469870866 | 8.82E-07 | 8069 | 0.0001232056946 | TRUE | 0.04127390769 | TRUE | 0.03697368243 | TRUE | FALSE | -9.764487486 |
| lpl | late_diastolic_peaks_lv_inner_circumferential_velocity_corrected | -6.090764946 | -1.977343626 | -0.06073024046 | 4.78E-08 | 1.114572839 | -0.06073024046 | 4.78E-08 | 8069 | 9.94E-06 | TRUE | 0.002236029374 | TRUE | 0.002003063063 | TRUE | FALSE | -6.090764946 |
| ltbp2 | late_diastolic_peaks_lv_inner_circumferential_velocity_corrected | -8.665128961 | -2.50583613 | -0.05552622547 | 6.01E-07 | 1.734803604 | -0.05552622547 | 6.01E-07 | 8069 | 8.87E-05 | TRUE | 0.02814072338 | TRUE | 0.02520881177 | TRUE | FALSE | -8.665128961 |
| marco | late_diastolic_peaks_lv_inner_velocity_corrected | 20.34090425 | -1.963523405 | 0.05737635094 | 2.50E-07 | 3.940626421 | 0.05737635094 | 2.50E-07 | 8069 | 4.22E-05 | TRUE | 0.01172090976 | TRUE | 0.01049973747 | TRUE | FALSE | 20.34090425 |
| marco | late_diastolic_peaks_lv_inner_radial_velocity_corrected | 14.58850407 | -0.9964893173 | 0.05823682355 | 1.65E-07 | 2.784321242 | 0.05823682355 | 1.65E-07 | 8069 | 2.92E-05 | TRUE | 0.007727804574 | TRUE | 0.006922663935 | TRUE | FALSE | 14.58850407 |
| mfge8 | late_diastolic_peaks_lv_inner_velocity_corrected | 12.88994408 | -2.949668882 | 0.05760031273 | 2.70E-07 | 2.50392163 | 0.05760031273 | 2.70E-07 | 7963 | 4.49E-05 | TRUE | 0.012627104 | TRUE | 0.0113115176 | TRUE | FALSE | 12.88994408 |
| mfge8 | late_diastolic_peaks_lv_inner_radial_velocity_corrected | 9.799152176 | -1.666329787 | 0.06193211513 | 3.18E-08 | 1.769922659 | 0.06193211513 | 3.18E-08 | 7963 | 6.90E-06 | TRUE | 0.001490719271 | TRUE | 0.001335404957 | TRUE | FALSE | 9.799152176 |
| mmp9 | diastolic_peaks_lwave_lv_inner_velocity_corrected | 11.46447029 | -0.7308583665 | 0.05482047492 | 1.01E-06 | 2.342520216 | 0.05482047492 | 1.01E-06 | 7948 | 0.000138269794 | TRUE | 0.04714999975 | TRUE | 0.04223755915 | TRUE | FALSE | 11.46447029 |
| mmp9 | diastolic_peaks_lwave_lv_inner_circumferential_velocity_corrected | 6.093333135 | -0.7976819416 | 0.05538185788 | 7.80E-07 | 1.232384066 | 0.05538185788 | 7.80E-07 | 7948 | 0.0001109423929 | TRUE | 0.03650004727 | TRUE | 0.03269719859 | TRUE | FALSE | 6.093333135 |
| mvk | late_diastolic_peaks_lv_inner_velocity_corrected | 7.731183631 | -2.61518421 | 0.06201538142 | 3.49E-08 | 1.400518274 | 0.06201538142 | 3.49E-08 | 7895 | 7.50E-06 | TRUE | 0.001635274287 | TRUE | 0.001464899147 | TRUE | FALSE | 7.731183631 |
| mvk | late_diastolic_peaks_lv_inner_radial_velocity_corrected | 5.131211919 | -1.46299607 | 0.05820321646 | 2.28E-07 | 0.990638163 | 0.05820321646 | 2.28E-07 | 7895 | 3.85E-05 | TRUE | 0.01066153715 | TRUE | 0.00955073825 | TRUE | FALSE | 5.131211919 |
| mvk | late_diastolic_peaks_lv_inner_circumferential_velocity_corrected | 3.980452288 | -1.680683198 | 0.05685756806 | 4.30E-07 | 0.7867195892 | 0.05685756806 | 4.30E-07 | 7895 | 6.66E-05 | TRUE | 0.02011738466 | TRUE | 0.01802140464 | TRUE | FALSE | 3.980452288 |
| nomo1 | late_diastolic_peaks_lv_inner_velocity_corrected | 26.76974612 | -1.926417819 | 0.06879443397 | 7.98E-10 | 4.350879066 | 0.06879443397 | 7.98E-10 | 7963 | 2.60E-07 | TRUE | 3.74E-05 | TRUE | 3.35E-05 | TRUE | FALSE | 26.76974612 |
| nomo1 | late_diastolic_peaks_lv_inner_radial_velocity_corrected | 17.85884331 | -1.000507531 | 0.06491065301 | 6.73E-09 | 3.077065048 | 0.06491065301 | 6.73E-09 | 7963 | 1.74E-06 | TRUE | 0.0003150301877 | TRUE | 0.00028220798 | TRUE | FALSE | 17.85884331 |
| nomo1 | late_diastolic_peaks_lv_inner_circumferential_velocity_corrected | 12.96005169 | -1.32069396 | 0.05934673925 | 1.16E-07 | 2.443205005 | 0.05934673925 | 1.16E-07 | 7963 | 2.10E-05 | TRUE | 0.005430799568 | TRUE | 0.004864978138 | TRUE | FALSE | 12.96005169 |
| nos3 | late_diastolic_peaks_lv_inner_velocity_corrected | 13.22484913 | -2.306758911 | 0.07133090802 | 2.25E-10 | 2.082197094 | 0.07133090802 | 2.25E-10 | 7890 | 8.79E-08 | TRUE | 1.05E-05 | TRUE | 9.44E-06 | TRUE | FALSE | 13.22484913 |
| nos3 | late_diastolic_peaks_lv_inner_radial_velocity_corrected | 8.359179925 | -1.319867666 | 0.06379460023 | 1.42E-08 | 1.472348993 | 0.06379460023 | 1.42E-08 | 7890 | 3.31E-06 | TRUE | 0.0006627788712 | TRUE | 0.0005937255976 | TRUE | FALSE | 8.359179925 |
| nos3 | late_diastolic_peaks_rv_inner_circumferential_velocity_corrected | 6.670262846 | -1.446396003 | 0.05636622297 | 5.44E-07 | 1.330299889 | 0.05636622297 | 5.44E-07 | 7890 | 8.22E-05 | TRUE | 0.02548760082 | TRUE | 0.02283211141 | TRUE | FALSE | 6.670262846 |
| nos3 | late_diastolic_peaks_lv_inner_circumferential_velocity_corrected | 6.873487152 | -1.421672418 | 0.06585846255 | 4.76E-09 | 1.17256843 | 0.06585846255 | 4.76E-09 | 7890 | 1.28E-06 | TRUE | 0.000222843698 | TRUE | 0.0001996261702 | TRUE | FALSE | 6.873487152 |
| notch3 | late_diastolic_peaks_lv_inner_velocity_corrected | -16.28643801 | -3.607647644 | -0.05848736179 | 1.46E-07 | 3.095023606 | -0.05848736179 | 1.46E-07 | 8069 | 2.60E-05 | TRUE | 0.006837627916 | TRUE | 0.006125232558 | TRUE | FALSE | -16.28643801 |
| notch3 | late_diastolic_peaks_lv_inner_radial_velocity_corrected | -13.22629738 | -2.221957205 | -0.06721999157 | 1.50E-09 | 2.18574952 | -0.06721999157 | 1.50E-09 | 8069 | 4.57E-07 | TRUE | 7.03E-05 | TRUE | 6.30E-05 | TRUE | FALSE | -13.22629738 |
| notch3 | late_diastolic_peaks_lv_inner_circumferential_velocity_corrected | -9.158379855 | -2.157064631 | -0.05854099698 | 1.42E-07 | 1.738829732 | -0.05854099698 | 1.42E-07 | 8069 | 2.54E-05 | TRUE | 0.006660379352 | TRUE | 0.005966451078 | TRUE | FALSE | -9.158379855 |
| nppb | late_diastolic_peaks_rv_inner_velocity_corrected | -4.203832783 | -3.010369252 | -0.05470295996 | 8.80E-07 | 0.8543342335 | -0.05470295996 | 8.80E-07 | 8069 | 0.0001232056946 | TRUE | 0.04119334042 | TRUE | 0.03690150926 | TRUE | FALSE | -4.203832783 |
| nppb | late_diastolic_peaks_lv_inner_velocity_corrected | -5.498197192 | -4.221611031 | -0.0747805953 | 1.75E-11 | 0.8163147426 | -0.0747805953 | 1.75E-11 | 8069 | 1.01E-08 | TRUE | 8.18E-07 | TRUE | 7.33E-07 | TRUE | FALSE | -5.498197192 |
| nppb | late_diastolic_peaks_lv_inner_radial_velocity_corrected | -3.965008026 | -2.620339061 | -0.07631974781 | 6.67E-12 | 0.5767433548 | -0.07631974781 | 6.67E-12 | 8069 | 4.34E-09 | TRUE | 3.12E-07 | TRUE | 2.80E-07 | TRUE | FALSE | -3.965008026 |
| nppb | late_diastolic_peaks_rv_inner_circumferential_velocity_corrected | -3.104382024 | -2.447706416 | -0.06604473277 | 2.88E-09 | 0.5221937822 | -0.06604473277 | 2.88E-09 | 8069 | 8.03E-07 | TRUE | 0.0001349044961 | TRUE | 0.0001208491339 | TRUE | FALSE | -3.104382024 |
| nppb | late_diastolic_peaks_lv_inner_circumferential_velocity_corrected | -3.261301182 | -2.536281561 | -0.07895234886 | 1.23E-12 | 0.4584711439 | -0.07895234886 | 1.23E-12 | 8069 | 9.59E-10 | TRUE | 5.75E-08 | TRUE | 5.15E-08 | TRUE | FALSE | -3.261301182 |
| npy | late_diastolic_peaks_lv_inner_velocity_corrected | 9.226388365 | -2.086350595 | 0.06462869646 | 6.22E-09 | 1.586139613 | 0.06462869646 | 6.22E-09 | 8069 | 1.63E-06 | TRUE | 0.0002913966816 | TRUE | 0.0002610367899 | TRUE | FALSE | 9.226388365 |
| npy | late_diastolic_peaks_lv_inner_radial_velocity_corrected | 6.67012588 | -1.078649475 | 0.06612290794 | 2.76E-09 | 1.120661767 | 0.06612290794 | 2.76E-09 | 8069 | 7.78E-07 | TRUE | 0.000129228935 | TRUE | 0.0001157648954 | TRUE | FALSE | 6.67012588 |
| npy | late_diastolic_peaks_lv_inner_circumferential_velocity_corrected | 4.638685447 | -1.363150971 | 0.05783545139 | 2.01E-07 | 0.8914914941 | 0.05783545139 | 2.01E-07 | 8069 | 3.45E-05 | TRUE | 0.009391965394 | TRUE | 0.008413439999 | TRUE | FALSE | 4.638685447 |
| ntprobnp | diastolic_peaks_lwave_lv_inner_circumferential_velocity_corrected | -4.165948349 | -1.890278742 | -0.06035094975 | 5.80E-08 | 0.7671995115 | -0.06035094975 | 5.80E-08 | 8068 | 1.16E-05 | TRUE | 0.002713945329 | TRUE | 0.00243118615 | TRUE | FALSE | -4.165948349 |
| ntprobnp | late_diastolic_peaks_rv_inner_velocity_corrected | -6.664080177 | -2.979340659 | -0.06496404271 | 5.19E-09 | 1.139705831 | -0.06496404271 | 5.19E-09 | 8069 | 1.38E-06 | TRUE | 0.000243167319 | TRUE | 0.0002178323239 | TRUE | FALSE | -6.664080177 |
| ntprobnp | late_diastolic_peaks_lv_inner_velocity_corrected | -11.45213741 | -4.514185644 | -0.1166870317 | 7.29E-26 | 1.085252935 | -0.1166870317 | 7.29E-26 | 8069 | 1.78E-21 | TRUE | 3.41E-21 | TRUE | 3.06E-21 | TRUE | FALSE | -11.45213741 |
| ntprobnp | late_diastolic_peaks_lv_inner_radial_velocity_corrected | -7.959869361 | -2.794945963 | -0.114780003 | 4.50E-25 | 0.7670150274 | -0.114780003 | 4.50E-25 | 8069 | 7.02E-21 | TRUE | 2.11E-20 | TRUE | 1.89E-20 | TRUE | FALSE | -7.959869361 |
| ntprobnp | late_diastolic_peaks_rv_inner_circumferential_velocity_corrected | -5.205887885 | -2.459457939 | -0.08297079931 | 8.34E-14 | 0.6961670503 | -0.08297079931 | 8.34E-14 | 8069 | 9.83E-11 | TRUE | 3.91E-09 | TRUE | 3.50E-09 | TRUE | FALSE | -5.205887885 |
| ntprobnp | late_diastolic_peaks_lv_inner_circumferential_velocity_corrected | -6.431637365 | -2.665833656 | -0.1166441956 | 7.59E-26 | 0.6097160516 | -0.1166441956 | 7.59E-26 | 8069 | 1.78E-21 | TRUE | 3.56E-21 | TRUE | 3.19E-21 | TRUE | FALSE | -6.431637365 |
| osm | diastolic_peaks_lwave_lv_inner_circumferential_velocity_corrected | 5.298006798 | -0.7589748979 | 0.05599770958 | 5.85E-07 | 1.059574091 | 0.05599770958 | 5.85E-07 | 7950 | 8.72E-05 | TRUE | 0.02739562971 | TRUE | 0.02454134754 | TRUE | FALSE | 5.298006798 |
| oxt | late_diastolic_peaks_lv_inner_velocity_corrected | 4.519528347 | -3.158298168 | 0.06982267208 | 3.59E-10 | 0.7198109063 | 0.06982267208 | 3.59E-10 | 8049 | 1.34E-07 | TRUE | 1.68E-05 | TRUE | 1.51E-05 | TRUE | FALSE | 4.519528347 |
| oxt | late_diastolic_peaks_lv_inner_radial_velocity_corrected | 2.887822228 | -1.876956538 | 0.06314657822 | 1.43E-08 | 0.5087870264 | 0.06314657822 | 1.43E-08 | 8049 | 3.32E-06 | TRUE | 0.0006683205277 | TRUE | 0.0005986898827 | TRUE | FALSE | 2.887822228 |
| oxt | late_diastolic_peaks_lv_inner_circumferential_velocity_corrected | 2.459857043 | -1.875221436 | 0.06764852672 | 1.24E-09 | 0.4044254786 | 0.06764852672 | 1.24E-09 | 8049 | 3.84E-07 | TRUE | 5.80E-05 | TRUE | 5.19E-05 | TRUE | FALSE | 2.459857043 |
| oxt | late_diastolic_peaks_rv_freewall_velocity_filtered_50_corrected | 0.1613854889 | 0.03038420921 | 0.05925467809 | 1.04E-07 | 0.03030826441 | 0.05925467809 | 1.04E-07 | 8049 | 1.92E-05 | TRUE | 0.004859179198 | TRUE | 0.004352913466 | TRUE | FALSE | 0.1613854889 |

| | | | | | | | | | | | | | | | | |
|---|---|---|---|---|---|---|---|---|---|---|---|---|---|---|---|---|
| **p4hb** | late_diastolic_peaks_lv_inner_radial_velocity_corrected | 9.749973199 | -1.491005052 | 0.05612635546 | 5.47E-07 | 1.944849315 | 0.05612635546 | 5.47E-07 | 7955 | 8.23E-05 | TRUE | 0.02559916522 | TRUE | 0.0229320522 | TRUE | FALSE | 9.749973199 |
| **pamr1** | late_diastolic_peaks_lv_inner_velocity_corrected | 28.53101938 | -1.520822852 | 0.08765151353 | 4.67E-15 | 3.634120465 | 0.08765151353 | 4.67E-15 | 7963 | 8.10E-12 | TRUE | 2.19E-10 | TRUE | 1.96E-10 | TRUE | FALSE | 28.53101938 |
| **pamr1** | late_diastolic_peaks_lv_inner_radial_velocity_corrected | 18.92779049 | -0.7359062054 | 0.08224237684 | 1.98E-13 | 2.57067233 | 0.08224237684 | 1.98E-13 | 7963 | 1.93E-10 | TRUE | 9.26E-09 | TRUE | 8.30E-09 | TRUE | FALSE | 18.92779049 |
| **pamr1** | late_diastolic_peaks_rv_inner_circumferential_velocity_corrected | 11.6258972 | -1.15713125 | 0.05584226093 | 6.16E-07 | 2.329707899 | 0.05584226093 | 6.16E-07 | 7963 | 9.00E-05 | TRUE | 0.02882428353 | TRUE | 0.02582115349 | TRUE | FALSE | 11.6258972 |
| **pamr1** | late_diastolic_peaks_lv_inner_circumferential_velocity_corrected | 14.46376552 | -1.087607056 | 0.0791780101 | 1.49E-12 | 2.04092649 | 0.0791780101 | 1.49E-12 | 7963 | 1.12E-09 | TRUE | 6.97E-08 | TRUE | 6.25E-08 | TRUE | FALSE | 14.46376552 |
| **pbld** | late_diastolic_peaks_lv_inner_velocity_corrected | 10.26258927 | -3.190275602 | 0.05550006551 | 8.10E-07 | 2.078787514 | 0.05550006551 | 8.10E-07 | 7890 | 0.000114272954 | TRUE | 0.03793862072 | TRUE | 0.03398589067 | TRUE | FALSE | 10.26258927 |
| **plat** | late_diastolic_peaks_lv_inner_velocity_corrected | 14.76271513 | -0.9030052723 | 0.08752576549 | 3.38E-15 | 1.870700989 | 0.08752576549 | 3.38E-15 | 8069 | 6.59E-12 | TRUE | 1.58E-10 | TRUE | 1.42E-10 | TRUE | FALSE | 14.76271513 |
| **plat** | late_diastolic_peaks_lv_inner_radial_velocity_corrected | 9.771108464 | -0.358523455 | 0.08198563446 | 1.63E-13 | 1.322469089 | 0.08198563446 | 1.63E-13 | 8069 | 1.66E-10 | TRUE | 7.64E-09 | TRUE | 6.85E-09 | TRUE | FALSE | 9.771108464 |
| **plat** | late_diastolic_peaks_lv_inner_circumferential_velocity_corrected | 6.872223132 | -0.8507851579 | 0.07252245062 | 6.93E-11 | 1.052261429 | 0.07252245062 | 6.93E-11 | 8069 | 3.01E-08 | TRUE | 3.25E-06 | TRUE | 2.91E-06 | TRUE | FALSE | 6.872223132 |
| **plin1** | late_diastolic_peaks_lv_inner_velocity_corrected | 15.82603132 | -2.848089275 | 0.05973513161 | 1.10E-07 | 2.977710798 | 0.05973513161 | 1.10E-07 | 7890 | 2.00E-05 | TRUE | 0.005134627894 | TRUE | 0.004599663851 | TRUE | FALSE | 15.82603132 |
| **plin1** | late_diastolic_peaks_lv_inner_radial_velocity_corrected | 11.42734237 | -1.642702963 | 0.06102900151 | 5.79E-08 | 2.104336754 | 0.06102900151 | 5.79E-08 | 7890 | 1.16E-05 | TRUE | 0.002711069968 | TRUE | 0.002428610366 | TRUE | FALSE | 11.42734237 |
| **pltp** | late_diastolic_peaks_lv_inner_velocity_corrected | -16.62924084 | -3.200546975 | -0.06395985256 | 9.25E-09 | 2.892037127 | -0.06395985256 | 9.25E-09 | 8051 | 2.30E-06 | TRUE | 0.0004330427782 | TRUE | 0.0003879251338 | TRUE | FALSE | -16.62924084 |
| **pltp** | late_diastolic_peaks_lv_inner_radial_velocity_corrected | -11.6873604 | -1.881529926 | -0.06363159689 | 1.10E-08 | 2.043109488 | -0.06363159689 | 1.10E-08 | 8051 | 2.66E-06 | TRUE | 0.0005154047951 | TRUE | 0.000461706058 | TRUE | FALSE | -11.6873604 |
| **pltp** | late_diastolic_peaks_lv_inner_circumferential_velocity_corrected | -9.231436893 | -1.935303458 | -0.06319451851 | 1.39E-08 | 1.624987439 | -0.06319451851 | 1.39E-08 | 8051 | 3.29E-06 | TRUE | 0.0006490202175 | TRUE | 0.0005814004236 | TRUE | FALSE | -9.231436893 |
| **pon3** | late_diastolic_peaks_lv_inner_velocity_corrected | -22.32647538 | -2.806342466 | -0.06725396903 | 1.86E-09 | 3.711057566 | -0.06725396903 | 1.86E-09 | 7968 | 5.49E-07 | TRUE | 8.73E-05 | TRUE | 7.82E-05 | TRUE | FALSE | -22.32647538 |
| **pon3** | late_diastolic_peaks_lv_inner_radial_velocity_corrected | -14.93846836 | -1.591087222 | -0.06363386284 | 1.31E-08 | 2.624922954 | -0.06363386284 | 1.31E-08 | 7968 | 3.12E-06 | TRUE | 0.0006121906227 | TRUE | 0.0005484080122 | TRUE | FALSE | -14.93846836 |
| **pon3** | late_diastolic_peaks_lv_inner_circumferential_velocity_corrected | -11.08307686 | -1.716645113 | -0.05940591554 | 1.12E-07 | 2.086617373 | -0.05940591554 | 1.12E-07 | 7968 | 2.03E-05 | TRUE | 0.005227568529 | TRUE | 0.004682921233 | TRUE | FALSE | -11.08307686 |
| **prcp** | late_diastolic_peaks_lv_inner_velocity_corrected | 24.10181582 | -1.400088324 | 0.08299184443 | 8.61E-14 | 3.22483669 | 0.08299184443 | 8.61E-14 | 8056 | 9.83E-11 | TRUE | 4.03E-09 | TRUE | 3.61E-09 | TRUE | FALSE | 24.10181582 |
| **prcp** | late_diastolic_peaks_lv_inner_radial_velocity_corrected | 16.44592268 | -0.6475786932 | 0.08013981546 | 5.87E-13 | 2.279318374 | 0.08013981546 | 5.87E-13 | 8056 | 5.19E-10 | TRUE | 2.75E-08 | TRUE | 2.46E-08 | TRUE | FALSE | 16.44592268 |
| **prcp** | late_diastolic_peaks_lv_inner_circumferential_velocity_corrected | 11.48187829 | -1.057343014 | 0.07036991418 | 2.57E-10 | 1.81360263 | 0.07036991418 | 2.57E-10 | 8056 | 9.93E-08 | TRUE | 1.20E-05 | TRUE | 1.08E-05 | TRUE | FALSE | 11.48187829 |
| **proc** | late_diastolic_peaks_lv_inner_velocity_corrected | 24.21780083 | -2.364588381 | 0.06481794709 | 6.90E-09 | 4.175817878 | 0.06481794709 | 6.90E-09 | 7974 | 1.78E-06 | TRUE | 0.0003232049112 | TRUE | 0.0002895309995 | TRUE | FALSE | 24.21780083 |
| **proc** | late_diastolic_peaks_lv_inner_radial_velocity_corrected | 16.9383036 | -1.299389339 | 0.06414461075 | 9.87E-09 | 2.951418476 | 0.06414461075 | 9.87E-09 | 7974 | 2.42E-06 | TRUE | 0.0004620842785 | TRUE | 0.0004139408728 | TRUE | FALSE | 16.9383036 |
| **proc** | late_diastolic_peaks_lv_inner_circumferential_velocity_corrected | 13.12404984 | -1.433869367 | 0.06243971071 | 2.40E-08 | 2.349498074 | 0.06243971071 | 2.40E-08 | 7974 | 5.35E-06 | TRUE | 0.001124291245 | TRUE | 0.001007154151 | TRUE | FALSE | 13.12404984 |
| **prss8** | late_diastolic_peaks_lv_inner_velocity_corrected | 18.07650447 | -1.297467332 | 0.08008864407 | 9.30E-13 | 2.527399927 | 0.08008864407 | 9.30E-13 | 7926 | 7.51E-10 | TRUE | 4.35E-08 | TRUE | 3.90E-08 | TRUE | FALSE | 18.07650447 |
| **prss8** | late_diastolic_peaks_lv_inner_radial_velocity_corrected | 11.80390369 | -0.5790701752 | 0.07389594685 | 4.49E-11 | 1.789549846 | 0.07389594685 | 4.49E-11 | 7926 | 2.19E-08 | TRUE | 2.10E-06 | TRUE | 1.88E-06 | TRUE | FALSE | 11.80390369 |
| **prss8** | late_diastolic_peaks_lv_inner_circumferential_velocity_corrected | 8.594570237 | -1.00544329 | 0.06779272035 | 1.53E-09 | 1.420916617 | 0.06779272035 | 1.53E-09 | 7926 | 4.61E-07 | TRUE | 7.15E-05 | TRUE | 6.41E-05 | TRUE | FALSE | 8.594570237 |
| **pts** | late_diastolic_peaks_lv_inner_velocity_corrected | 12.04106821 | -2.814974269 | 0.06904724868 | 8.82E-10 | 1.962069509 | 0.06904724868 | 8.82E-10 | 7864 | 2.82E-07 | TRUE | 4.13E-05 | TRUE | 3.70E-05 | TRUE | FALSE | 12.04106821 |
| **pts** | late_diastolic_peaks_lv_inner_radial_velocity_corrected | 7.525010556 | -1.623589345 | 0.06105252893 | 6.02E-08 | 1.387477747 | 0.06105252893 | 6.02E-08 | 7864 | 1.20E-05 | TRUE | 0.002817108254 | TRUE | 0.002523600787 | TRUE | FALSE | 7.525010556 |
| **pts** | late_diastolic_peaks_lv_inner_circumferential_velocity_corrected | 6.661663148 | -1.694278636 | 0.06786926155 | 1.70E-09 | 1.104435817 | 0.06786926155 | 1.70E-09 | 7864 | 5.02E-07 | TRUE | 7.94E-05 | TRUE | 7.11E-05 | TRUE | FALSE | 6.661663148 |
| **rarres2** | late_diastolic_peaks_lv_inner_velocity_corrected | 17.86819377 | -0.6820097385 | 0.09376590337 | 4.44E-17 | 2.122219805 | 0.09376590337 | 4.44E-17 | 7994 | 1.48E-13 | TRUE | 2.08E-12 | TRUE | 1.86E-12 | TRUE | FALSE | 17.86819377 |
| **rarres2** | late_diastolic_peaks_lv_inner_radial_velocity_corrected | 12.73999564 | -0.0777858795 | 0.09458917905 | 2.35E-17 | 1.499852326 | 0.09458917905 | 2.35E-17 | 7994 | 1.00E-13 | TRUE | 1.10E-12 | TRUE | 9.87E-13 | TRUE | FALSE | 12.73999564 |
| **rarres2** | late_diastolic_peaks_rv_inner_circumferential_velocity_corrected | 7.412305485 | -0.80802228 | 0.0607782805 | 5.38E-08 | 1.361675226 | 0.0607782805 | 5.38E-08 | 7994 | 1.09E-05 | TRUE | 0.002518102144 | TRUE | 0.002255747377 | TRUE | FALSE | 7.412305485 |
| **rarres2** | late_diastolic_peaks_lv_inner_circumferential_velocity_corrected | 8.805847586 | -0.6739161489 | 0.08223100313 | 1.79E-13 | 1.193808084 | 0.08223100313 | 1.79E-13 | 7994 | 1.78E-10 | TRUE | 8.39E-09 | TRUE | 7.51E-09 | TRUE | FALSE | 8.805847586 |
| **rarres2** | late_diastolic_peaks_rv_freewall_velocity_filtered_50_corrected | 0.487954591 | 0.0973105136 | 0.06081945413 | 5.27E-08 | 0.08957863383 | 0.06081945413 | 5.27E-08 | 7994 | 1.08E-05 | TRUE | 0.002466468055 | TRUE | 0.002209492914 | TRUE | FALSE | 0.487954591 |
| **rbks** | late_diastolic_peaks_lv_inner_velocity_corrected | 10.61039564 | -2.868397426 | 0.05696425893 | 3.71E-07 | 2.085642122 | 0.05696425893 | 3.71E-07 | 7952 | 5.91E-05 | TRUE | 0.01738765929 | TRUE | 0.01557608254 | TRUE | FALSE | 10.61039564 |
| **rbks** | late_diastolic_peaks_lv_inner_circumferential_velocity_corrected | 6.030388079 | -1.75501151 | 0.05768834402 | 2.64E-07 | 1.170441644 | 0.05768834402 | 2.64E-07 | 7952 | 4.41E-05 | TRUE | 0.01234023571 | TRUE | 0.0110545374 | TRUE | FALSE | 6.030388079 |
| **rbp5** | late_diastolic_peaks_lv_inner_velocity_corrected | 15.49746653 | -2.309484579 | 0.09380347335 | 3.08E-17 | 1.831329978 | 0.09380347335 | 3.08E-17 | 8069 | 1.20E-13 | TRUE | 1.44E-12 | TRUE | 1.29E-12 | TRUE | FALSE | 15.49746653 |
| **rbp5** | late_diastolic_peaks_lv_inner_radial_velocity_corrected | 9.902286732 | -1.308008878 | 0.08482384594 | 2.31E-14 | 1.295070673 | 0.08482384594 | 2.31E-14 | 8069 | 3.38E-11 | TRUE | 1.08E-09 | TRUE | 9.69E-10 | TRUE | FALSE | 9.902286732 |
| **rbp5** | late_diastolic_peaks_lv_inner_circumferential_velocity_corrected | 7.769254957 | -1.476500191 | 0.08370340531 | 5.04E-14 | 1.02980139 | 0.08370340531 | 5.04E-14 | 8069 | 6.61E-11 | TRUE | 2.36E-09 | TRUE | 2.11E-09 | TRUE | FALSE | 7.769254957 |
| **rgma** | late_diastolic_peaks_lv_inner_velocity_corrected | -17.60541359 | -2.573160544 | -0.05705778264 | 3.02E-07 | 3.434047084 | -0.05705778264 | 3.02E-07 | 8049 | 4.90E-05 | TRUE | 0.01412403683 | TRUE | 0.01265248874 | TRUE | FALSE | -17.60541359 |
| **rgma** | late_diastolic_peaks_lv_inner_radial_velocity_corrected | -12.46404378 | -1.464634905 | -0.05717466658 | 2.85E-07 | 2.42620403 | -0.05717466658 | 2.85E-07 | 8049 | 4.70E-05 | TRUE | 0.0133573499 | TRUE | 0.011965681 | TRUE | FALSE | -12.46404378 |
| **rgmb** | diastolic_peaks_lwave_lv_inner_radial_velocity_corrected | -20.29364407 | -0.3224105859 | -0.05634710129 | 4.23E-07 | 4.008740164 | -0.05634710129 | 4.23E-07 | 8048 | 6.58E-05 | TRUE | 0.01981733648 | TRUE | 0.01775261773 | TRUE | FALSE | -20.29364407 |
| **rtn4r** | late_diastolic_peaks_lv_inner_velocity_corrected | 14.67376134 | -2.354136228 | 0.0543909648 | 1.02E-06 | 2.999268213 | 0.0543909648 | 1.02E-06 | 8069 | 0.0001389672457 | TRUE | 0.04752679803 | TRUE | 0.04257509976 | TRUE | FALSE | 14.67376134 |
| **scly** | late_diastolic_peaks_lv_inner_velocity_corrected | 14.28597009 | -2.932970345 | 0.07438522571 | 2.28E-11 | 2.13342621 | 0.07438522571 | 2.28E-11 | 8061 | 1.23E-08 | TRUE | 1.07E-06 | TRUE | 9.57E-07 | TRUE | FALSE | 14.28597009 |
| **scly** | late_diastolic_peaks_lv_inner_radial_velocity_corrected | 9.031297522 | -1.704157957 | 0.06654390304 | 2.23E-09 | 1.508473269 | 0.06654390304 | 2.23E-09 | 8061 | 6.36E-07 | TRUE | 0.0001043241471 | TRUE | 9.35E-05 | TRUE | FALSE | 9.031297522 |
| **scly** | late_diastolic_peaks_lv_inner_circumferential_velocity_corrected | 7.418458534 | -1.794233734 | 0.06877466958 | 6.36E-10 | 1.198712543 | 0.06877466958 | 6.36E-10 | 8061 | 2.16E-07 | TRUE | 2.98E-05 | TRUE | 2.67E-05 | TRUE | FALSE | 7.418458534 |
| **sdc4** | late_diastolic_peaks_lv_inner_velocity_corrected | 12.12303641 | -2.739001624 | 0.05677112009 | 3.34E-07 | 2.373706018 | 0.05677112009 | 3.34E-07 | 8069 | 5.40E-05 | TRUE | 0.01565609467 | TRUE | 0.01402492531 | TRUE | FALSE | 12.12303641 |
| **sdc4** | late_diastolic_peaks_lv_inner_radial_velocity_corrected | 9.186039583 | -1.537228376 | 0.06087915495 | 4.43E-08 | 1.676862168 | 0.06087915495 | 4.43E-08 | 8069 | 9.30E-06 | TRUE | 0.002073170893 | TRUE | 0.0018571724 | TRUE | FALSE | 9.186039583 |
| **sele** | late_diastolic_peaks_lv_inner_velocity_corrected | 15.04049316 | -1.276247046 | 0.08249817585 | 1.15E-13 | 2.022920228 | 0.08249817585 | 1.15E-13 | 8069 | 1.25E-10 | TRUE | 5.40E-09 | TRUE | 4.83E-09 | TRUE | FALSE | 15.04049316 |
| **sele** | late_diastolic_peaks_lv_inner_radial_velocity_corrected | 9.401548867 | -0.6733963397 | 0.07298034742 | 5.26E-11 | 1.430466881 | 0.07298034742 | 5.26E-11 | 8069 | 2.44E-08 | TRUE | 2.46E-06 | TRUE | 2.21E-06 | TRUE | FALSE | 9.401548867 |
| **sele** | late_diastolic_peaks_lv_inner_circumferential_velocity_corrected | 7.309894912 | -0.9867374161 | 0.0713672469 | 1.38E-10 | 1.13748936 | 0.0713672469 | 1.38E-10 | 8069 | 5.57E-08 | TRUE | 6.47E-06 | TRUE | 5.79E-06 | TRUE | FALSE | 7.309894912 |
| **sema3f** | late_diastolic_peaks_lv_inner_velocity_corrected | 23.76054768 | -2.102152043 | 0.06628068052 | 2.53E-09 | 3.982514836 | 0.06628068052 | 2.53E-09 | 8069 | 7.18E-07 | TRUE | 0.0001184738118 | TRUE | 0.0001061303216 | TRUE | FALSE | 23.76054768 |
| **sema3f** | late_diastolic_peaks_lv_inner_circumferential_velocity_corrected | 13.54612272 | -1.302561504 | 0.06725910104 | 1.47E-09 | 2.237295569 | 0.06725910104 | 1.47E-09 | 8069 | 4.50E-07 | TRUE | 6.88E-05 | TRUE | 6.16E-05 | TRUE | FALSE | 13.54612272 |
| **slc39a5** | late_diastolic_peaks_lv_inner_velocity_corrected | 10.85830649 | -2.416478722 | 0.06455373394 | 7.95E-09 | 1.879967319 | 0.06455373394 | 7.95E-09 | 7974 | 2.00E-06 | TRUE | 0.0003720309985 | TRUE | 0.0003332700188 | TRUE | FALSE | 10.85830649 |
| **slc39a5** | late_diastolic_peaks_lv_inner_radial_velocity_corrected | 7.078494326 | -1.399178339 | 0.05956794126 | 1.02E-07 | 1.328534296 | 0.05956794126 | 1.02E-07 | 7974 | 1.90E-05 | TRUE | 0.004774639602 | TRUE | 0.004277181839 | TRUE | FALSE | 7.078494326 |
| **slc39a5** | late_diastolic_peaks_lv_inner_circumferential_velocity_corrected | 5.17151774 | -1.52800267 | 0.05475420751 | 9.97E-07 | 1.056245534 | 0.05475420751 | 9.97E-07 | 7974 | 0.000137229153 | TRUE | 0.04665791202 | TRUE | 0.04179674081 | TRUE | FALSE | 5.17151774 |
| **smoc1** | late_diastolic_peaks_lv_inner_velocity_corrected | 16.6001269 | -1.562228515 | 0.05929627352 | 9.80E-08 | 3.111452197 | 0.05929627352 | 9.80E-08 | 8069 | 1.84E-05 | TRUE | 0.004590143584 | TRUE | 0.004111907999 | TRUE | FALSE | 16.6001269 |
| **smoc1** | late_diastolic_peaks_lv_inner_circumferential_velocity_corrected | 9.082333956 | -1.030546753 | 0.05774561359 | 2.10E-07 | 1.748224243 | 0.05774561359 | 2.10E-07 | 8069 | 3.58E-05 | TRUE | 0.009809321362 | TRUE | 0.008787312693 | TRUE | FALSE | 9.082333956 |
| **sord** | late_diastolic_peaks_lv_inner_velocity_corrected | 9.471305482 | -2.576513273 | 0.06890742799 | 6.01E-10 | 1.528218207 | 0.06890742799 | 6.01E-10 | 8053 | 2.06E-07 | TRUE | 2.82E-05 | TRUE | 2.52E-05 | TRUE | FALSE | 9.471305482 |
| **sord** | late_diastolic_peaks_lv_inner_radial_velocity_corrected | 6.136826685 | -1.469651452 | 0.06316247317 | 1.40E-08 | 1.080666719 | 0.06316247317 | 1.40E-08 | 8053 | 3.31E-06 | TRUE | 0.0006573323065 | TRUE | 0.0005888464968 | TRUE | FALSE | 6.136826685 |
| **sord** | late_diastolic_peaks_lv_inner_circumferential_velocity_corrected | 5.33229628 | -1.586198503 | 0.06909170735 | 5.41E-10 | 0.8580732516 | 0.06909170735 | 5.41E-10 | 8053 | 1.89E-07 | TRUE | 2.53E-05 | TRUE | 2.27E-05 | TRUE | FALSE | 5.33229628 |
| **sort1** | late_diastolic_peaks_lv_inner_velocity_corrected | 14.70616755 | -2.612023627 | 0.05567682925 | 5.67E-07 | 2.937536773 | 0.05567682925 | 5.67E-07 | 8062 | 8.48E-05 | TRUE | 0.02652805268 | TRUE | 0.02376416119 | TRUE | FALSE | 14.70616755 |
| **sort1** | late_diastolic_peaks_lv_inner_radial_velocity_corrected | 10.23768823 | -1.472852407 | 0.05485781802 | 8.28E-07 | 2.075589121 | 0.05485781802 | 8.28E-07 | 8062 | 0.0001164627601 | TRUE | 0.03878209913 | TRUE | 0.03474148917 | TRUE | FALSE | 10.23768823 |
| **spon1** | late_diastolic_peaks_rv_inner_radial_velocity_corrected | 11.87712938 | -0.1281947767 | 0.0611363677 | 4.93E-08 | 2.175600226 | 0.0611363677 | 4.93E-08 | 7946 | 1.02E-05 | TRUE | 0.00230615612 | TRUE | 0.00206588348 | TRUE | FALSE | 11.87712938 |
| **ssc4d** | late_diastolic_peaks_lv_inner_velocity_corrected | 7.332102236 | -3.870901094 | 0.1018882347 | 4.69E-20 | 0.7973902462 | 0.1018882347 | 4.69E-20 | 8062 | 5.49E-16 | TRUE | 2.20E-15 | TRUE | 1.97E-15 | TRUE | FALSE | 7.332102236 |

| | | | | | | | | | | | | | | | | | |
|---|---|---|---|---|---|---|---|---|---|---|---|---|---|---|---|---|---|
| **ssc4d** | late_diastolic_peaks_lv_inner_radial_velocity_corrected | 4.828324029 | -2.319705775 | 0.09496281465 | 1.29E-17 | 0.5637781052 | 0.09496281465 | 1.29E-17 | 8062 | 6.04E-14 | TRUE | 6.04E-13 | TRUE | 5.41E-13 | TRUE | FALSE | 4.828324029 |
| **ssc4d** | late_diastolic_peaks_lv_inner_circumferential_velocity_corrected | 4.02857068 | -2.311736354 | 0.09964872614 | 3.01E-19 | 0.4480687904 | 0.09964872614 | 3.01E-19 | 8062 | 2.82E-15 | TRUE | 1.41E-14 | TRUE | 1.26E-14 | TRUE | FALSE | 4.02857068 |
| **ssc5d** | late_diastolic_peaks_lv_inner_velocity_corrected | 16.18007781 | -1.753813195 | 0.07649160111 | 7.79E-12 | 2.361121682 | 0.07649160111 | 7.79E-12 | 7981 | 4.93E-09 | TRUE | 3.64E-07 | TRUE | 3.27E-07 | TRUE | FALSE | 16.18007781 |
| **ssc5d** | late_diastolic_peaks_lv_inner_radial_velocity_corrected | 10.90232457 | -0.9045556785 | 0.07291958266 | 6.94E-11 | 1.669333044 | 0.07291958266 | 6.94E-11 | 7981 | 3.01E-08 | TRUE | 3.25E-06 | TRUE | 2.91E-06 | TRUE | FALSE | 10.90232457 |
| **ssc5d** | late_diastolic_peaks_lv_inner_circumferential_velocity_corrected | 7.869860816 | -1.158332269 | 0.06613220703 | 3.35E-09 | 1.329315391 | 0.06613220703 | 3.35E-09 | 7981 | 9.27E-07 | TRUE | 0.0001567314468 | TRUE | 0.0001404019892 | TRUE | FALSE | 7.869860816 |
| **stc1** | late_diastolic_peaks_lv_inner_velocity_corrected | 12.36959123 | -2.503351124 | 0.05602759491 | 4.91E-07 | 2.457274366 | 0.05602759491 | 4.91E-07 | 8049 | 7.54E-05 | TRUE | 0.02299176085 | TRUE | 0.02059630677 | TRUE | FALSE | 12.36959123 |
| **sult2a1** | late_diastolic_peaks_lv_inner_velocity_corrected | 10.92123843 | -3.551325855 | 0.06195725656 | 3.08E-08 | 1.970429482 | 0.06195725656 | 3.08E-08 | 7974 | 6.70E-06 | TRUE | 0.001439973966 | TRUE | 0.001289946678 | TRUE | FALSE | 10.92123843 |
| **tfpi** | late_diastolic_peaks_lv_inner_velocity_corrected | 17.13685289 | -2.152697201 | 0.05687458051 | 3.81E-07 | 3.371729919 | 0.05687458051 | 3.81E-07 | 7962 | 6.05E-05 | TRUE | 0.01783490972 | TRUE | 0.01597673506 | TRUE | FALSE | 17.13685289 |
| **tfpi** | late_diastolic_peaks_lv_inner_radial_velocity_corrected | 12.08656339 | -1.157497328 | 0.05674363654 | 4.05E-07 | 2.383575032 | 0.05674363654 | 4.05E-07 | 7962 | 6.36E-05 | TRUE | 0.01896727965 | TRUE | 0.01699112621 | TRUE | FALSE | 12.08656339 |
| **thop1** | late_diastolic_peaks_lv_inner_velocity_corrected | 21.98015231 | -2.510046486 | 0.07332210103 | 4.62E-11 | 3.334450086 | 0.07332210103 | 4.62E-11 | 8041 | 2.23E-08 | TRUE | 2.16E-06 | TRUE | 1.94E-06 | TRUE | FALSE | 21.98015231 |
| **thop1** | late_diastolic_peaks_lv_inner_radial_velocity_corrected | 14.13257482 | -1.429712927 | 0.06673552321 | 2.10E-09 | 2.356643858 | 0.06673552321 | 2.10E-09 | 8041 | 6.10E-07 | TRUE | 9.82E-05 | TRUE | 8.80E-05 | TRUE | FALSE | 14.13257482 |
| **thop1** | late_diastolic_peaks_lv_inner_circumferential_velocity_corrected | 12.65383525 | -1.592678154 | 0.07513774588 | 1.52E-11 | 1.872981904 | 0.07513774588 | 1.52E-11 | 8041 | 9.11E-09 | TRUE | 7.10E-07 | TRUE | 6.36E-07 | TRUE | FALSE | 12.65383525 |
| **tnfrsf11b** | diastolic_peaks_lwave_rv_inner_radial_velocity_corrected | 13.64282733 | 0.9276671148 | 0.05764123496 | 2.80E-07 | 2.653957727 | 0.05764123496 | 2.80E-07 | 7929 | 4.64E-05 | TRUE | 0.01313034514 | TRUE | 0.01176232731 | TRUE | FALSE | 13.64282733 |
| **tpp1** | late_diastolic_peaks_lv_inner_velocity_corrected | 12.92862417 | -2.532634442 | 0.05990690798 | 1.04E-07 | 2.428019417 | 0.05990690798 | 1.04E-07 | 7874 | 1.92E-05 | TRUE | 0.00486311744 | TRUE | 0.004356441392 | TRUE | FALSE | 12.92862417 |
| **tpp1** | late_diastolic_peaks_lv_inner_radial_velocity_corrected | 8.868474569 | -1.43069319 | 0.05806750641 | 2.52E-07 | 1.718461409 | 0.05806750641 | 2.52E-07 | 7874 | 4.23E-05 | TRUE | 0.01179909003 | TRUE | 0.01056977234 | TRUE | FALSE | 8.868474569 |
| **vegfd** | late_diastolic_peaks_lv_inner_velocity_corrected | -15.35696234 | -2.938452275 | -0.05980614218 | 8.28E-08 | 2.861983943 | -0.05980614218 | 8.28E-08 | 8023 | 1.58E-05 | TRUE | 0.003877356585 | TRUE | 0.003473384496 | TRUE | FALSE | -15.35696234 |
| **vegfd** | late_diastolic_peaks_lv_inner_radial_velocity_corrected | -10.89488437 | -1.697964448 | -0.0600478129 | 7.34E-08 | 2.02221246 | -0.0600478129 | 7.34E-08 | 8023 | 1.43E-05 | TRUE | 0.003437616163 | TRUE | 0.003079459529 | TRUE | FALSE | -10.89488437 |
| **vegfd** | late_diastolic_peaks_lv_inner_circumferential_velocity_corrected | -8.50673905 | -1.811470189 | -0.05899420004 | 1.24E-07 | 1.607246457 | -0.05899420004 | 1.24E-07 | 8023 | 2.23E-05 | TRUE | 0.005790462323 | TRUE | 0.00518716853 | TRUE | FALSE | -8.50673905 |
| **vmo1** | late_diastolic_peaks_lv_inner_velocity_corrected | 7.675342177 | -1.984755418 | 0.05445675881 | 9.85E-07 | 1.566913467 | 0.05445675881 | 9.85E-07 | 8069 | 0.0001364359737 | TRUE | 0.04611774345 | TRUE | 0.04131285105 | TRUE | FALSE | 7.675342177 |
| **vwa1** | late_diastolic_peaks_lv_inner_velocity_corrected | 12.36173418 | -2.624858157 | 0.05552102274 | 6.03E-07 | 2.475115707 | 0.05552102274 | 6.03E-07 | 8069 | 8.87E-05 | TRUE | 0.02820904489 | TRUE | 0.02527001502 | TRUE | FALSE | 12.36173418 |

**Supplemental Table 4. Proteome-wide association analysis of circulating proteins with optical flow velocity phenotypes.** Significant associations between 1,463 Olink Explore protein levels and 32 optical flow-derived velocity phenotypes in up to 8,116 UK Biobank participants, after cluster-adjusted Bonferroni correction ($p < 1.19\times10^{-6}$; M_eff = 28.7). Three hundred forty-four significant associations across 129 unique proteins and 14 distinct velocity phenotypes are shown, sorted by p-value.

<table>
<tr><th>protein</th><th>traditional_parameter</th><th>beta</th><th>intercept</th><th>r_value</th><th>p_value</th><th>std_err</th><th>pearson_r</th><th>pearson_p</th><th>n_observations</th><th>fdr_bh_p</th><th>fdr_bh_sig</th><th>bonferroni_standard_p</th><th>bonferroni_standard_sig</th><th>bonferroni_cluster_p</th><th>bonferroni_cluster_sig</th><th>nominally_significant</th><th>beta_numeric</th></tr>
<tr><td>acaa1</td><td>LV end diastolic volume | Instance 2(participant - p24100_i2)_BSA_indexed_corrected</td><td>-1.053350248</td><td>0.3072250099</td><td>-0.07735772485</td><td>3.01E-07</td><td>0.2052938856</td><td>-0.07735772485</td><td>3.01E-07</td><td>4375</td><td>4.76E-06</td><td>TRUE</td><td>0.004400630723</td><td>TRUE</td><td>0.003722053465</td><td>TRUE</td><td>FALSE</td><td>-1.053350248</td></tr>
<tr><td>acaa1</td><td>RV end diastolic volume | Instance 2_BSA_indexed_corrected</td><td>-1.340933763</td><td>0.1851508846</td><td>-0.09153571044</td><td>1.32E-09</td><td>0.2205971693</td><td>-0.09153571044</td><td>1.32E-09</td><td>4375</td><td>3.16E-08</td><td>TRUE</td><td>1.92E-05</td><td>TRUE</td><td>1.63E-05</td><td>TRUE</td><td>FALSE</td><td>-1.340933763</td></tr>
<tr><td>acaa1</td><td>RV end systolic volume | Instance 2_BSA_indexed_corrected</td><td>-0.7315401311</td><td>0.1853206972</td><td>-0.08397180456</td><td>2.66E-08</td><td>0.1312738869</td><td>-0.08397180456</td><td>2.66E-08</td><td>4375</td><td>5.09E-07</td><td>TRUE</td><td>0.0003891736494</td><td>TRUE</td><td>0.0003291630727</td><td>TRUE</td><td>FALSE</td><td>-0.7315401311</td></tr>
<tr><td>acp5</td><td>LV end diastolic volume | Instance 2(participant - p24100_i2)_BSA_indexed_corrected</td><td>-4.326534069</td><td>-0.03857705414</td><td>-0.1333872902</td><td>8.01E-19</td><td>0.4861134478</td><td>-0.1333872902</td><td>8.01E-19</td><td>4375</td><td>9.09E-17</td><td>TRUE</td><td>1.17E-14</td><td>TRUE</td><td>9.92E-15</td><td>TRUE</td><td>FALSE</td><td>-4.326534069</td></tr>
<tr><td>acp5</td><td>LV end systolic volume | Instance 2(participant - p24101_i2)_BSA_indexed_corrected</td><td>-2.085065775</td><td>0.123193663</td><td>-0.1107396362</td><td>2.05E-13</td><td>0.2829746498</td><td>-0.1107396362</td><td>2.05E-13</td><td>4375</td><td>9.92E-12</td><td>TRUE</td><td>3.00E-09</td><td>TRUE</td><td>2.54E-09</td><td>TRUE</td><td>FALSE</td><td>-2.085065775</td></tr>
<tr><td>acp5</td><td>RV end diastolic volume | Instance 2_BSA_indexed_corrected</td><td>-5.299041164</td><td>-0.2361707324</td><td>-0.1518532038</td><td>5.53E-24</td><td>0.5215760915</td><td>-0.1518532038</td><td>5.53E-24</td><td>4375</td><td>1.51E-21</td><td>TRUE</td><td>8.09E-20</td><td>TRUE</td><td>6.84E-20</td><td>TRUE</td><td>FALSE</td><td>-5.299041164</td></tr>
<tr><td>acp5</td><td>RV end systolic volume | Instance 2_BSA_indexed_corrected</td><td>-2.726199251</td><td>-0.02962534591</td><td>-0.1313700484</td><td>2.67E-18</td><td>0.3110937129</td><td>-0.1313700484</td><td>2.67E-18</td><td>4375</td><td>2.73E-16</td><td>TRUE</td><td>3.91E-14</td><td>TRUE</td><td>3.30E-14</td><td>TRUE</td><td>FALSE</td><td>-2.726199251</td></tr>
<tr><td>acvrl1</td><td>LV end diastolic volume | Instance 2(participant - p24100_i2)_BSA_indexed_corrected</td><td>-4.421461663</td><td>0.08231076044</td><td>-0.09345609395</td><td>6.05E-10</td><td>0.71279023</td><td>-0.09345609395</td><td>6.05E-10</td><td>4369</td><td>1.57E-08</td><td>TRUE</td><td>8.85E-06</td><td>TRUE</td><td>7.49E-06</td><td>TRUE</td><td>FALSE</td><td>-4.421461663</td></tr>
<tr><td>acvrl1</td><td>LV end systolic volume | Instance 2(participant - p24101_i2)_BSA_indexed_corrected</td><td>-2.430779625</td><td>0.1674363724</td><td>-0.08852054428</td><td>4.60E-09</td><td>0.4139059534</td><td>-0.08852054428</td><td>4.60E-09</td><td>4369</td><td>9.99E-08</td><td>TRUE</td><td>6.74E-05</td><td>TRUE</td><td>5.70E-05</td><td>TRUE</td><td>FALSE</td><td>-2.430779625</td></tr>
<tr><td>acvrl1</td><td>RV end diastolic volume | Instance 2_BSA_indexed_corrected</td><td>-4.937016526</td><td>-0.0623540655</td><td>-0.0969932855</td><td>1.32E-10</td><td>0.7666176058</td><td>-0.0969932855</td><td>1.32E-10</td><td>4369</td><td>3.87E-09</td><td>TRUE</td><td>1.94E-06</td><td>TRUE</td><td>1.64E-06</td><td>TRUE</td><td>FALSE</td><td>-4.937016526</td></tr>
<tr><td>acvrl1</td><td>RV end systolic volume | Instance 2_BSA_indexed_corrected</td><td>-2.967103097</td><td>0.0320370397</td><td>-0.09804773124</td><td>8.33E-11</td><td>0.455728175</td><td>-0.09804773124</td><td>8.33E-11</td><td>4369</td><td>2.52E-09</td><td>TRUE</td><td>1.22E-06</td><td>TRUE</td><td>1.03E-06</td><td>TRUE</td><td>FALSE</td><td>-2.967103097</td></tr>
<tr><td>acy1</td><td>LV end diastolic volume | Instance 2(participant - p24100_i2)_BSA_indexed_corrected</td><td>-2.320872396</td><td>0.292342826</td><td>-0.1411937806</td><td>6.38E-21</td><td>0.2460782592</td><td>-0.1411937806</td><td>6.38E-21</td><td>4375</td><td>1.03E-18</td><td>TRUE</td><td>9.34E-17</td><td>TRUE</td><td>7.90E-17</td><td>TRUE</td><td>FALSE</td><td>-2.320872396</td></tr>
<tr><td>acy1</td><td>LV end systolic volume | Instance 2(participant - p24101_i2)_BSA_indexed_corrected</td><td>-1.069565518</td><td>0.2839511931</td><td>-0.1120935718</td><td>1.04E-13</td><td>0.1433808702</td><td>-0.1120935718</td><td>1.04E-13</td><td>4375</td><td>5.25E-12</td><td>TRUE</td><td>1.52E-09</td><td>TRUE</td><td>1.29E-09</td><td>TRUE</td><td>FALSE</td><td>-1.069565518</td></tr>
<tr><td>acy1</td><td>RV end diastolic volume | Instance 2_BSA_indexed_corrected</td><td>-2.388702667</td><td>0.1809969901</td><td>-0.1350761714</td><td>2.88E-19</td><td>0.2649689883</td><td>-0.1350761714</td><td>2.88E-19</td><td>4375</td><td>3.52E-17</td><td>TRUE</td><td>4.22E-15</td><td>TRUE</td><td>3.57E-15</td><td>TRUE</td><td>FALSE</td><td>-2.388702667</td></tr>
<tr><td>acy1</td><td>RV end systolic volume | Instance 2_BSA_indexed_corrected</td><td>-1.167934181</td><td>0.1865892551</td><td>-0.1110573015</td><td>1.75E-13</td><td>0.1580471344</td><td>-0.1110573015</td><td>1.75E-13</td><td>4375</td><td>8.52E-12</td><td>TRUE</td><td>2.56E-09</td><td>TRUE</td><td>2.17E-09</td><td>TRUE</td><td>FALSE</td><td>-1.167934181</td></tr>
<tr><td>adam22</td><td>LV cardiac output | Instance 2_BSA_indexed_corrected</td><td>-0.1266079399</td><td>-0.02182833746</td><td>-0.08328398263</td><td>3.45E-08</td><td>0.02290858629</td><td>-0.08328398263</td><td>3.45E-08</td><td>4375</td><td>6.48E-07</td><td>TRUE</td><td>0.0005052711653</td><td>TRUE</td><td>0.0004273583516</td><td>TRUE</td><td>FALSE</td><td>-0.1266079399</td></tr>
<tr><td>adam22</td><td>LV end diastolic volume | Instance 2(participant - p24100_i2)_BSA_indexed_corrected</td><td>-2.966143362</td><td>0.3042525228</td><td>-0.08708245972</td><td>7.96E-09</td><td>0.5131197506</td><td>-0.08708245972</td><td>7.96E-09</td><td>4375</td><td>1.65E-07</td><td>TRUE</td><td>0.0001164711404</td><td>TRUE</td><td>9.85E-05</td><td>TRUE</td><td>FALSE</td><td>-2.966143362</td></tr>
<tr><td>adam22</td><td>RV end diastolic volume | Instance 2_BSA_indexed_corrected</td><td>-3.221825044</td><td>0.1904765721</td><td>-0.08792104612</td><td>5.71E-09</td><td>0.5519938943</td><td>-0.08792104612</td><td>5.71E-09</td><td>4375</td><td>1.21E-07</td><td>TRUE</td><td>8.35E-05</td><td>TRUE</td><td>7.06E-05</td><td>TRUE</td><td>FALSE</td><td>-3.221825044</td></tr>
<tr><td>adam8</td><td>RV end diastolic volume | Instance 2_BSA_indexed_corrected</td><td>-3.424475715</td><td>0.09144558956</td><td>-0.07956777496</td><td>1.44E-07</td><td>0.6500306061</td><td>-0.07956777496</td><td>1.44E-07</td><td>4358</td><td>2.42E-06</td><td>TRUE</td><td>0.002113132309</td><td>TRUE</td><td>0.001787287307</td><td>TRUE</td><td>FALSE</td><td>-3.424475715</td></tr>
<tr><td>adamts15</td><td>LV end diastolic volume | Instance 2(participant - p24100_i2)_BSA_indexed_corrected</td><td>-3.976251292</td><td>0.06571346621</td><td>-0.159356521</td><td>2.82E-26</td><td>0.3725020556</td><td>-0.159356521</td><td>2.82E-26</td><td>4375</td><td>1.06E-23</td><td>TRUE</td><td>4.12E-22</td><td>TRUE</td><td>3.49E-22</td><td>TRUE</td><td>FALSE</td><td>-3.976251292</td></tr>
<tr><td>adamts15</td><td>LV end systolic volume | Instance 2(participant - p24101_i2)_BSA_indexed_corrected</td><td>-1.77713159</td><td>0.1835062184</td><td>-0.1226943641</td><td>3.83E-16</td><td>0.217375936</td><td>-0.1226943641</td><td>3.83E-16</td><td>4375</td><td>2.99E-14</td><td>TRUE</td><td>5.60E-12</td><td>TRUE</td><td>4.73E-12</td><td>TRUE</td><td>FALSE</td><td>-1.77713159</td></tr>
<tr><td>adamts15</td><td>RV end diastolic volume | Instance 2_BSA_indexed_corrected</td><td>-3.941358854</td><td>-0.0413380189</td><td>-0.146823083</td><td>1.65E-22</td><td>0.4015407836</td><td>-0.146823083</td><td>1.65E-22</td><td>4375</td><td>3.21E-20</td><td>TRUE</td><td>2.41E-18</td><td>TRUE</td><td>2.04E-18</td><td>TRUE</td><td>FALSE</td><td>-3.941358854</td></tr>
<tr><td>adamts15</td><td>RV end systolic volume | Instance 2_BSA_indexed_corrected</td><td>-1.796608171</td><td>0.08730856552</td><td>-0.1125417933</td><td>8.30E-14</td><td>0.2398733073</td><td>-0.1125417933</td><td>8.30E-14</td><td>4375</td><td>4.29E-12</td><td>TRUE</td><td>1.21E-09</td><td>TRUE</td><td>1.03E-09</td><td>TRUE</td><td>FALSE</td><td>-1.796608171</td></tr>
<tr><td>adamts8</td><td>RV end diastolic volume | Instance 2_BSA_indexed_corrected</td><td>2.16397253</td><td>0.2100160718</td><td>0.08069360301</td><td>1.02E-07</td><td>0.4058348085</td><td>0.08069360301</td><td>1.02E-07</td><td>4340</td><td>1.76E-06</td><td>TRUE</td><td>0.001492143548</td><td>TRUE</td><td>0.001262055013</td><td>TRUE</td><td>FALSE</td><td>2.16397253</td></tr>
<tr><td>adgrg1</td><td>LV end diastolic volume | Instance 2(participant - p24100_i2)_BSA_indexed_corrected</td><td>-1.961612804</td><td>0.3724123739</td><td>-0.09939599498</td><td>4.43E-11</td><td>0.2969604932</td><td>-0.09939599498</td><td>4.43E-11</td><td>4375</td><td>1.43E-09</td><td>TRUE</td><td>6.48E-07</td><td>TRUE</td><td>5.48E-07</td><td>TRUE</td><td>FALSE</td><td>-1.961612804</td></tr>
<tr><td>adgrg1</td><td>RV end diastolic volume | Instance 2_BSA_indexed_corrected</td><td>-2.39176898</td><td>0.267093333</td><td>-0.1126489559</td><td>7.86E-14</td><td>0.3190282445</td><td>-0.1126489559</td><td>7.86E-14</td><td>4375</td><td>4.09E-12</td><td>TRUE</td><td>1.15E-09</td><td>TRUE</td><td>9.72E-10</td><td>TRUE</td><td>FALSE</td><td>-2.39176898</td></tr>
<tr><td>adgrg1</td><td>RV end systolic volume | Instance 2_BSA_indexed_corrected</td><td>-0.90177496</td><td>0.2260385432</td><td>-0.07141973157</td><td>2.26E-06</td><td>0.1904494686</td><td>-0.07141973157</td><td>2.26E-06</td><td>4375</td><td>3.02E-05</td><td>TRUE</td><td>0.03306468373</td><td>TRUE</td><td>0.0279661095</td><td>TRUE</td><td>FALSE</td><td>-0.90177496</td></tr>
<tr><td>adgrg2</td><td>RV end diastolic volume | Instance 2_BSA_indexed_corrected</td><td>3.10975141</td><td>0.192589348</td><td>0.07727857751</td><td>3.09E-07</td><td>0.6067029813</td><td>0.07727857751</td><td>3.09E-07</td><td>4375</td><td>4.87E-06</td><td>TRUE</td><td>0.004525041004</td><td>TRUE</td><td>0.003827279681</td><td>TRUE</td><td>FALSE</td><td>3.10975141</td></tr>
<tr><td>adh4</td><td>LV end diastolic volume | Instance 2(participant - p24100_i2)_BSA_indexed_corrected</td><td>-1.617084598</td><td>0.2539503628</td><td>-0.1366138297</td><td>1.12E-19</td><td>0.177319847</td><td>-0.1366138297</td><td>1.12E-19</td><td>4375</td><td>1.54E-17</td><td>TRUE</td><td>1.65E-15</td><td>TRUE</td><td>1.39E-15</td><td>TRUE</td><td>FALSE</td><td>-1.617084598</td></tr>
<tr><td>adh4</td><td>LV end systolic volume | Instance 2(participant - p24101_i2)_BSA_indexed_corrected</td><td>-0.741209462</td><td>0.2665043175</td><td>-0.1078727849</td><td>8.43E-13</td><td>0.1032993684</td><td>-0.1078727849</td><td>8.43E-13</td><td>4375</td><td>3.68E-11</td><td>TRUE</td><td>1.23E-08</td><td>TRUE</td><td>1.04E-08</td><td>TRUE</td><td>FALSE</td><td>-0.741209462</td></tr>
<tr><td>adh4</td><td>RV end diastolic volume | Instance 2_BSA_indexed_corrected</td><td>-1.743409771</td><td>0.1366389013</td><td>-0.1369032459</td><td>9.41E-20</td><td>0.190760076</td><td>-0.1369032459</td><td>9.41E-20</td><td>4375</td><td>1.34E-17</td><td>TRUE</td><td>1.38E-15</td><td>TRUE</td><td>1.16E-15</td><td>TRUE</td><td>FALSE</td><td>-1.743409771</td></tr>
<tr><td>adh4</td><td>RV end systolic volume | Instance 2_BSA_indexed_corrected</td><td>-0.849490197</td><td>0.1650805142</td><td>-0.1121720717</td><td>1.00E-13</td><td>0.11379791</td><td>-0.1121720717</td><td>1.00E-13</td><td>4375</td><td>5.10E-12</td><td>TRUE</td><td>1.46E-09</td><td>TRUE</td><td>1.24E-09</td><td>TRUE</td><td>FALSE</td><td>-0.849490197</td></tr>
<tr><td>adm</td><td>LV end diastolic volume | Instance 2(participant - p24100_i2)_BSA_indexed_corrected</td><td>-6.018760128</td><td>-0.297100512</td><td>-0.1648086314</td><td>5.15E-28</td><td>0.5447003226</td><td>-0.1648086314</td><td>5.15E-28</td><td>4375</td><td>2.28E-25</td><td>TRUE</td><td>7.53E-24</td><td>TRUE</td><td>6.37E-24</td><td>TRUE</td><td>FALSE</td><td>-6.018760128</td></tr>
<tr><td>adm</td><td>LV end systolic volume | Instance 2(participant - p24101_i2)_BSA_indexed_corrected</td><td>-2.813962329</td><td>0.007962003702</td><td>-0.1327395092</td><td>1.18E-18</td><td>0.3177372543</td><td>-0.1327395092</td><td>1.18E-18</td><td>4375</td><td>1.32E-16</td><td>TRUE</td><td>1.73E-14</td><td>TRUE</td><td>1.46E-14</td><td>TRUE</td><td>FALSE</td><td>-2.813962329</td></tr>
<tr><td>adm</td><td>RV end diastolic volume | Instance 2_BSA_indexed_corrected</td><td>-6.672152054</td><td>-0.477249172</td><td>-0.1698209229</td><td>1.15E-29</td><td>0.5855049672</td><td>-0.1698209229</td><td>1.15E-29</td><td>4375</td><td>6.00E-27</td><td>TRUE</td><td>1.68E-25</td><td>TRUE</td><td>1.42E-25</td><td>TRUE</td><td>FALSE</td><td>-6.672152054</td></tr>
<tr><td>adm</td><td>RV end systolic volume | Instance 2_BSA_indexed_corrected</td><td>-3.145410957</td><td>-0.1226297607</td><td>-0.1346216028</td><td>3.80E-19</td><td>0.3501075277</td><td>-0.1346216028</td><td>3.80E-19</td><td>4375</td><td>4.56E-17</td><td>TRUE</td><td>5.56E-15</td><td>TRUE</td><td>4.70E-15</td><td>TRUE</td><td>FALSE</td><td>-3.145410957</td></tr>
<tr><td>agr3</td><td>LV end diastolic volume | Instance 2(participant - p24100_i2)_BSA_indexed_corrected</td><td>-1.749757928</td><td>0.3088633814</td><td>-0.09721329482</td><td>1.22E-10</td><td>0.2711739982</td><td>-0.09721329482</td><td>1.22E-10</td><td>4366</td><td>3.58E-09</td><td>TRUE</td><td>1.78E-06</td><td>TRUE</td><td>1.51E-06</td><td>TRUE</td><td>FALSE</td><td>-1.749757928</td></tr>
<tr><td>agr3</td><td>RV end diastolic volume | Instance 2_BSA_indexed_corrected</td><td>-1.736036169</td><td>0.1982817938</td><td>-0.08961668454</td><td>3.00E-09</td><td>0.2920630006</td><td>-0.08961668454</td><td>3.00E-09</td><td>4366</td><td>6.70E-08</td><td>TRUE</td><td>4.39E-05</td><td>TRUE</td><td>3.71E-05</td><td>TRUE</td><td>FALSE</td><td>-1.736036169</td></tr>
<tr><td>agrn</td><td>LV end diastolic volume | Instance 2(participant - p24100_i2)_BSA_indexed_corrected</td><td>-5.477545773</td><td>-0.01440284642</td><td>-0.1539162087</td><td>2.52E-24</td><td>0.5349997872</td><td>-0.1539162087</td><td>2.52E-24</td><td>4322</td><td>7.24E-22</td><td>TRUE</td><td>3.69E-20</td><td>TRUE</td><td>3.12E-20</td><td>TRUE</td><td>FALSE</td><td>-5.477545773</td></tr>
<tr><td>agrn</td><td>LV end systolic volume | Instance 2(participant - p24101_i2)_BSA_indexed_corrected</td><td>-2.172695784</td><td>0.1615925172</td><td>-0.1052850495</td><td>3.95E-12</td><td>0.3122265554</td><td>-0.1052850495</td><td>3.95E-12</td><td>4322</td><td>1.54E-10</td><td>TRUE</td><td>5.78E-08</td><td>TRUE</td><td>4.89E-08</td><td>TRUE</td><td>FALSE</td><td>-2.172695784</td></tr>
<tr><td>agrn</td><td>RV end diastolic volume | Instance 2_BSA_indexed_corrected</td><td>-6.546657121</td><td>-0.1973102069</td><td>-0.1710873011</td><td>9.54E-30</td><td>0.573599784</td><td>-0.1710873011</td><td>9.54E-30</td><td>4322</td><td>5.17E-27</td><td>TRUE</td><td>1.40E-25</td><td>TRUE</td><td>1.18E-25</td><td>TRUE</td><td>FALSE</td><td>-6.546657121</td></tr>
<tr><td>agrn</td><td>RV end systolic volume | Instance 2_BSA_indexed_corrected</td><td>-2.784149097</td><td>0.02531904969</td><td>-0.1222934028</td><td>7.16E-16</td><td>0.3437759353</td><td>-0.1222934028</td><td>7.16E-16</td><td>4322</td><td>5.40E-14</td><td>TRUE</td><td>1.05E-11</td><td>TRUE</td><td>8.86E-12</td><td>TRUE</td><td>FALSE</td><td>-2.784149097</td></tr>
<tr><td>agrp</td><td>LV end diastolic volume | Instance 2(participant - p24100_i2)_BSA_indexed_corrected</td><td>-1.503150518</td><td>0.2637852543</td><td>-0.07383007871</td><td>1.06E-06</td><td>0.3075309455</td><td>-0.07383007871</td><td>1.06E-06</td><td>4361</td><td>1.50E-05</td><td>TRUE</td><td>0.01545397638</td><td>TRUE</td><td>0.01307097322</td><td>TRUE</td><td>FALSE</td><td>-1.503150518</td></tr>
<tr><td>agxt</td><td>LV end diastolic volume | Instance 2(participant - p24100_i2)_BSA_indexed_corrected</td><td>-1.038174771</td><td>0.4004011892</td><td>-0.07253881766</td><td>1.56E-06</td><td>0.2158562561</td><td>-0.07253881766</td><td>1.56E-06</td><td>4375</td><td>2.14E-05</td><td>TRUE</td><td>0.02287340787</td><td>TRUE</td><td>0.01934632838</td><td>TRUE</td><td>FALSE</td><td>-1.038174771</td></tr>
<tr><td>agxt</td><td>RV end diastolic volume | Instance 2_BSA_indexed_corrected</td><td>-1.160514054</td><td>0.2964125874</td><td>-0.07537073495</td><td>6.01E-07</td><td>0.2321778791</td><td>-0.07537073495</td><td>6.01E-07</td><td>4375</td><td>8.96E-06</td><td>TRUE</td><td>0.008788739422</td><td>TRUE</td><td>0.007433515803</td><td>TRUE</td><td>FALSE</td><td>-1.160514054</td></tr>
<tr><td>aifm1</td><td>RV end diastolic volume | Instance 2_BSA_indexed_corrected</td><td>-0.8602022938</td><td>0.1964616844</td><td>-0.07569675619</td><td>5.43E-07</td><td>0.1714290715</td><td>-0.07569675619</td><td>5.43E-07</td><td>4371</td><td>8.18E-06</td><td>TRUE</td><td>0.007949118817</td><td>TRUE</td><td>0.006723364696</td><td>TRUE</td><td>FALSE</td><td>-0.8602022938</td></tr>
<tr><td>aldh1a1</td><td>LV end diastolic volume | Instance 2(participant - p24100_i2)_BSA_indexed_corrected</td><td>-2.071367815</td><td>0.3179485535</td><td>-0.1033575516</td><td>1.03E-11</td><td>0.3036648664</td><td>-0.1033575516</td><td>1.03E-11</td><td>4311</td><td>3.70E-10</td><td>TRUE</td><td>1.50E-07</td><td>TRUE</td><td>1.27E-07</td><td>TRUE</td><td>FALSE</td><td>-2.071367815</td></tr>
<tr><td>aldh1a1</td><td>LV end systolic volume | Instance 2(participant - p24101_i2)_BSA_indexed_corrected</td><td>-0.8829319196</td><td>0.3021112738</td><td>-0.07583984722</td><td>6.19E-07</td><td>0.1768434397</td><td>-0.07583984722</td><td>6.19E-07</td><td>4311</td><td>9.19E-06</td><td>TRUE</td><td>0.009053540154</td><td>TRUE</td><td>0.007657484263</td><td>TRUE</td><td>FALSE</td><td>-0.8829319196</td></tr>
<tr><td>aldh1a1</td><td>RV end diastolic volume | Instance 2_BSA_indexed_corrected</td><td>-2.127332287</td><td>0.2388673305</td><td>-0.09860671536</td><td>8.68E-11</td><td>0.3270535563</td><td>-0.09860671536</td><td>8.68E-11</td><td>4311</td><td>2.61E-09</td><td>TRUE</td><td>1.27E-06</td><td>TRUE</td><td>1.07E-06</td><td>TRUE</td><td>FALSE</td><td>-2.127332287</td></tr>
<tr><td>aldh1a1</td><td>RV end systolic volume | Instance 2_BSA_indexed_corrected</td><td>-1.00372057</td><td>0.2146763826</td><td>-0.07824232776</td><td>2.69E-07</td><td>0.194827108</td><td>-0.07824232776</td><td>2.69E-07</td><td>4311</td><td>4.30E-06</td><td>TRUE</td><td>0.003942124941</td><td>TRUE</td><td>0.003334249275</td><td>TRUE</td><td>FALSE</td><td>-1.00372057</td></tr>
<tr><td>ambp</td><td>LV end diastolic volume | Instance 2(participant - p24100_i2)_BSA_indexed_corrected</td><td>-5.77788577</td><td>-0.01863674048</td><td>-0.1052996892</td><td>2.94E-12</td><td>0.8253350274</td><td>-0.1052996892</td><td>2.94E-12</td><td>4373</td><td>1.17E-10</td><td>TRUE</td><td>4.30E-08</td><td>TRUE</td><td>3.63E-08</td><td>TRUE</td><td>FALSE</td><td>-5.77788577</td></tr>
<tr><td>ambp</td><td>LV end systolic volume | Instance 2(participant - p24101_i2)_BSA_indexed_corrected</td><td>-2.683886046</td><td>0.1431155397</td><td>-0.08427575472</td><td>2.39E-08</td><td>0.4799801183</td><td>-0.08427575472</td><td>2.39E-08</td><td>4373</td><td>4.62E-07</td><td>TRUE</td><td>0.000349096025</td><td>TRUE</td><td>0.000295265418</td><td>TRUE</td><td>FALSE</td><td>-2.683886046</td></tr>
<tr><td>ambp</td><td>RV end diastolic volume | Instance 2_BSA_indexed_corrected</td><td>-7.829671586</td><td>-0.2591669356</td><td>-0.1326436099</td><td>1.27E-18</td><td>0.8849367701</td><td>-0.1326436099</td><td>1.27E-18</td><td>4373</td><td>1.41E-16</td><td>TRUE</td><td>1.86E-14</td><td>TRUE</td><td>1.58E-14</td><td>TRUE</td><td>FALSE</td><td>-7.829671586</td></tr>
<tr><td>ambp</td><td>RV end systolic volume | Instance 2_BSA_indexed_corrected</td><td>-3.615107811</td><td>-0.01283877417</td><td>-0.1029693743</td><td>8.77E-12</td><td>0.5282118483</td><td>-0.1029693743</td><td>8.77E-12</td><td>4373</td><td>3.22E-10</td><td>TRUE</td><td>1.28E-07</td><td>TRUE</td><td>1.08E-07</td><td>TRUE</td><td>FALSE</td><td>-3.615107811</td></tr>
<tr><td>ang</td><td>LV end diastolic volume | Instance 2(participant - p24100_i2)_BSA_indexed_corrected</td><td>-3.283106697</td><td>0.2316574412</td><td>-0.1024693875</td><td>1.53E-11</td><td>0.4854111522</td><td>-0.1024693875</td><td>1.53E-11</td><td>4313</td><td>5.33E-10</td><td>TRUE</td><td>2.23E-07</td><td>TRUE</td><td>1.89E-07</td><td>TRUE</td><td>FALSE</td><td>-3.283106697</td></tr>
<tr><td>ang</td><td>LV end systolic volume | Instance 2(participant - p24101_i2)_BSA_indexed_corrected</td><td>-1.476661034</td><td>0.2568607519</td><td>-0.07931070841</td><td>1.84E-07</td><td>0.2826765377</td><td>-0.07931070841</td><td>1.84E-07</td><td>4313</td><td>3.02E-06</td><td>TRUE</td><td>0.002684702718</td><td>TRUE</td><td>0.002270721559</td><td>TRUE</td><td>FALSE</td><td>-1.476661034</td></tr>
<tr><td>ang</td><td>RV end diastolic volume | Instance 2_BSA_indexed_corrected</td><td>-3.299364259</td><td>0.106582146</td><td>-0.09578659898</td><td>2.91E-10</td><td>0.5221977097</td><td>-0.09578659898</td><td>2.91E-10</td><td>4313</td><td>8.09E-09</td><td>TRUE</td><td>4.26E-06</td><td>TRUE</td><td>3.61E-06</td><td>TRUE</td><td>FALSE</td><td>-3.299364259</td></tr>
<tr><td>angptl2</td><td>LV end diastolic volume | Instance 2(participant - p24100_i2)_BSA_indexed_corrected</td><td>-2.470850772</td><td>0.1933558756</td><td>-0.08968597548</td><td>3.66E-09</td><td>0.4180036086</td><td>-0.08968597548</td><td>3.66E-09</td><td>4311</td><td>8.08E-08</td><td>TRUE</td><td>5.36E-05</td><td>TRUE</td><td>4.53E-05</td><td>TRUE</td><td>FALSE</td><td>-2.470850772</td></tr>
<tr><td>angptl2</td><td>RV end diastolic volume | Instance 2_BSA_indexed_corrected</td><td>-2.618401707</td><td>0.07351539718</td><td>-0.08842390594</td><td>6.05E-09</td><td>0.4493387088</td><td>-0.08842390594</td><td>6.05E-09</td><td>4311</td><td>1.28E-07</td><td>TRUE</td><td>8.85E-05</td><td>TRUE</td><td>7.48E-05</td><td>TRUE</td><td>FALSE</td><td>-2.618401707</td></tr>
<tr><td>angptl4</td><td>LV end diastolic volume | Instance 2(participant - p24100_i2)_BSA_indexed_corrected</td><td>-2.39326746</td><td>0.1128567726</td><td>-0.09872689425</td><td>8.19E-11</td><td>0.3674431954</td><td>-0.09872689425</td><td>8.19E-11</td><td>4312</td><td>2.49E-09</td><td>TRUE</td><td>1.20E-06</td><td>TRUE</td><td>1.01E-06</td><td>TRUE</td><td>FALSE</td><td>-2.39326746</td></tr>
<tr><td>angptl4</td><td>LV end systolic volume | Instance 2(participant - p24101_i2)_BSA_indexed_corrected</td><td>-1.000329822</td><td>0.2088251936</td><td>-0.07118344374</td><td>2.88E-06</td><td>0.2135120973</td><td>-0.07118344374</td><td>2.88E-06</td><td>4312</td><td>3.77E-05</td><td>TRUE</td><td>0.04219555608</td><td>TRUE</td><td>0.03568900134</td><td>TRUE</td><td>FALSE</td><td>-1.000329822</td></tr>
<tr><td>angptl4</td><td>RV end diastolic volume | Instance 2_BSA_indexed_corrected</td><td>-2.15773556</td><td>0.02885386716</td><td>-0.08286211333</td><td>5.07E-08</td><td>0.3952823976</td><td>-0.08286211333</td><td>5.07E-08</td><td>4312</td><td>9.19E-07</td><td>TRUE</td><td>0.0007410507524</td><td>TRUE</td><td>0.0006267807264</td><td>TRUE</td><td>FALSE</td><td>-2.15773556</td></tr>
<tr><td>apbb1ip</td><td>LV end diastolic volume | Instance 2(participant - p24100_i2)_BSA_indexed_corrected</td><td>-2.460283969</td><td>0.2165757649</td><td>-0.1064457476</td><td>1.68E-12</td><td>0.3475303251</td><td>-0.1064457476</td><td>1.68E-12</td><td>4375</td><td>6.88E-11</td><td>TRUE</td><td>2.46E-08</td><td>TRUE</td><td>2.08E-08</td><td>TRUE</td><td>FALSE</td><td>-2.460283969</td></tr>
<tr><td>apbb1ip</td><td>LV end systolic volume | Instance 2(participant - p24101_i2)_BSA_indexed_corrected</td><td>-1.170488082</td><td>0.2470003317</td><td>-0.0872406889</td><td>7.48E-09</td><td>0.2021152836</td><td>-0.0872406889</td><td>7.48E-09</td><td>4375</td><td>1.56E-07</td><td>TRUE</td><td>0.0001094154373</td><td>TRUE</td><td>9.25E-05</td><td>TRUE</td><td>FALSE</td><td>-1.170488082</td></tr>
<tr><td>apbb1ip</td><td>RV end diastolic volume | Instance 2_BSA_indexed_corrected</td><td>-2.59920072</td><td>0.09929927899</td><td>-0.1045286262</td><td>4.18E-12</td><td>0.3739634685</td><td>-0.1045286262</td><td>4.18E-12</td><td>4375</td><td>1.63E-10</td><td>TRUE</td><td>6.12E-08</td><td>TRUE</td><td>5.17E-08</td><td>TRUE</td><td>FALSE</td><td>-2.59920072</td></tr>
<tr><td>apbb1ip</td><td>RV end systolic volume | Instance 2_BSA_indexed_corrected</td><td>-1.350136031</td><td>0.1422472592</td><td>-0.09130300068</td><td>1.45E-09</td><td>0.2226819211</td><td>-0.09130300068</td><td>1.45E-09</td><td>4375</td><td>3.42E-08</td><td>TRUE</td><td>2.12E-05</td><td>TRUE</td><td>1.79E-05</td><td>TRUE</td><td>FALSE</td><td>-1.350136031</td></tr>
<tr><td>apex1</td><td>LV end diastolic volume | Instance 2(participant - p24100_i2)_BSA_indexed_corrected</td><td>-1.923412156</td><td>0.1699773103</td><td>-0.1037384323</td><td>6.33E-12</td><td>0.2791200416</td><td>-0.1037384323</td><td>6.33E-12</td><td>4367</td><td>2.39E-10</td><td>TRUE</td><td>9.26E-08</td><td>TRUE</td><td>7.84E-08</td><td>TRUE</td><td>FALSE</td><td>-1.923412156</td></tr>
</table>

| | | | | | | | | | | | | | | | | | |
|---|---|---|---|---|---|---|---|---|---|---|---|---|---|---|---|---|---|
| **apex1** | LV end systolic volume \| Instance 2(participant - p24101_i2)_BSA_indexed_corrected | -0.8285859197 | 0.2330296626 | -0.07697126461 | 3.53E-07 | 0.1624525271 | -0.07697126461 | 3.53E-07 | 4367 | 5.50E-06 | TRUE | 0.005166743449 | TRUE | 0.004370031609 | TRUE | FALSE | -0.8285859197 |
| **apex1** | RV end diastolic volume \| Instance 2_BSA_indexed_corrected | -1.836962378 | 0.07596964582 | -0.09206138445 | 1.09E-09 | 0.300733913 | -0.09206138445 | 1.09E-09 | 4367 | 2.68E-08 | TRUE | 1.60E-05 | TRUE | 1.35E-05 | TRUE | FALSE | -1.836962378 |
| **apex1** | RV end systolic volume \| Instance 2_BSA_indexed_corrected | -0.8813883214 | 0.1386813897 | -0.07426882477 | 8.96E-07 | 0.1791296788 | -0.07426882477 | 8.96E-07 | 4367 | 1.29E-05 | TRUE | 0.01310145841 | TRUE | 0.01108121352 | TRUE | FALSE | -0.8813883214 |
| **aplp1** | LV cardiac output \| Instance 2_BSA_indexed_corrected | -0.08060392079 | -0.01361502208 | -0.08690973237 | 8.52E-09 | 0.01397177593 | -0.08690973237 | 8.52E-09 | 4375 | 1.76E-07 | TRUE | 0.0001246782418 | TRUE | 0.000105452857 | TRUE | FALSE | -0.08060392079 |
| **areg** | RV end diastolic volume \| Instance 2_BSA_indexed_corrected | -1.890151112 | 0.2616920479 | -0.06972708212 | 3.91E-06 | 0.4089281241 | -0.06972708212 | 3.91E-06 | 4375 | 5.01E-05 | TRUE | 0.05714705133 | FALSE | 0.04833497601 | TRUE | FALSE | -1.890151112 |
| **arsa** | LV end diastolic volume \| Instance 2(participant - p24100_i2)_BSA_indexed_corrected | -2.063589573 | 0.3274638792 | -0.08273237901 | 5.33E-08 | 0.3786763295 | -0.08273237901 | 5.33E-08 | 4311 | 9.63E-07 | TRUE | 0.0007803197864 | TRUE | 0.0006599944753 | TRUE | FALSE | -2.063589573 |
| **arsa** | RV end diastolic volume \| Instance 2_BSA_indexed_corrected | -2.078966938 | 0.2489848507 | -0.07742584306 | 3.58E-07 | 0.4078192574 | -0.07742584306 | 3.58E-07 | 4311 | 5.58E-06 | TRUE | 0.005243740217 | TRUE | 0.004435155475 | TRUE | FALSE | -2.078966938 |
| **art3** | LV cardiac output \| Instance 2_BSA_indexed_corrected | -0.1161516275 | -0.01412937298 | -0.08254866336 | 5.47E-08 | 0.02133233297 | -0.08254866336 | 5.47E-08 | 4323 | 9.86E-07 | TRUE | 0.0008005765717 | TRUE | 0.0006771276643 | TRUE | FALSE | -0.1161516275 |
| **asah2** | LV end diastolic volume \| Instance 2(participant - p24100_i2)_BSA_indexed_corrected | -1.724469288 | 0.1935848317 | -0.08465685868 | 2.05E-08 | 0.3069316862 | -0.08465685868 | 2.05E-08 | 4375 | 4.00E-07 | TRUE | 0.0002994545182 | TRUE | 0.0002532786315 | TRUE | FALSE | -1.724469288 |
| **asah2** | RV end diastolic volume \| Instance 2_BSA_indexed_corrected | -1.705327577 | 0.08578189995 | -0.07781563116 | 2.56E-07 | 0.3303941501 | -0.07781563116 | 2.56E-07 | 4375 | 4.10E-06 | TRUE | 0.003743140259 | TRUE | 0.003165948031 | TRUE | FALSE | -1.705327577 |
| **asgr1** | LV end diastolic volume \| Instance 2(participant - p24100_i2)_BSA_indexed_corrected | -5.221834438 | -0.02447151844 | -0.1490956315 | 3.60E-23 | 0.5237052218 | -0.1490956315 | 3.60E-23 | 4375 | 8.08E-21 | TRUE | 5.27E-19 | TRUE | 4.46E-19 | TRUE | FALSE | -5.221834438 |
| **asgr1** | LV end systolic volume \| Instance 2(participant - p24101_i2)_BSA_indexed_corrected | -2.543843159 | 0.1280172656 | -0.1251241565 | 9.86E-17 | 0.3050231959 | -0.1251241565 | 9.86E-17 | 4375 | 8.48E-15 | TRUE | 1.44E-12 | TRUE | 1.22E-12 | TRUE | FALSE | -2.543843159 |
| **asgr1** | RV end diastolic volume \| Instance 2_BSA_indexed_corrected | -6.597109665 | -0.2334628286 | -0.1750845677 | 1.86E-31 | 0.5609903261 | -0.1750845677 | 1.86E-31 | 4375 | 1.30E-28 | TRUE | 2.73E-27 | TRUE | 2.31E-27 | TRUE | FALSE | -6.597109665 |
| **asgr1** | RV end systolic volume \| Instance 2_BSA_indexed_corrected | -3.326790891 | -0.02337243711 | -0.1484676273 | 5.50E-23 | 0.3350919211 | -0.1484676273 | 5.50E-23 | 4375 | 1.18E-20 | TRUE | 8.04E-19 | TRUE | 6.80E-19 | TRUE | FALSE | -3.326790891 |
| **azu1** | LV end diastolic volume \| Instance 2(participant - p24100_i2)_BSA_indexed_corrected | -1.262262082 | 0.3829775571 | -0.08515972064 | 2.59E-08 | 0.2262981271 | -0.08515972064 | 2.59E-08 | 4261 | 4.97E-07 | TRUE | 0.0003782874625 | TRUE | 0.0003199555358 | TRUE | FALSE | -1.262262082 |
| **azu1** | RV end diastolic volume \| Instance 2_BSA_indexed_corrected | -1.23550325 | 0.2990655183 | -0.0774381027 | 4.17E-07 | 0.2437413937 | -0.0774381027 | 4.17E-07 | 4261 | 6.42E-06 | TRUE | 0.006101851792 | TRUE | 0.005160946246 | TRUE | FALSE | -1.23550325 |
| **baiap2** | LV end diastolic volume \| Instance 2(participant - p24100_i2)_BSA_indexed_corrected | -1.672093151 | 0.2596514101 | -0.09445279584 | 3.86E-10 | 0.2665078318 | -0.09445279584 | 3.86E-10 | 4375 | 1.05E-08 | TRUE | 5.64E-06 | TRUE | 4.77E-06 | TRUE | FALSE | -1.672093151 |
| **baiap2** | RV end diastolic volume \| Instance 2_BSA_indexed_corrected | -1.962915961 | 0.1338402945 | -0.1030643338 | 8.30E-12 | 0.2864736197 | -0.1030643338 | 8.30E-12 | 4375 | 3.07E-10 | TRUE | 1.21E-07 | TRUE | 1.03E-07 | TRUE | FALSE | -1.962915961 |
| **baiap2** | RV end systolic volume \| Instance 2_BSA_indexed_corrected | -0.908049306 | 0.1664191862 | -0.08017301656 | 1.10E-07 | 0.1707226733 | -0.08017301656 | 1.10E-07 | 4375 | 1.89E-06 | TRUE | 0.001603991113 | TRUE | 0.001356655683 | TRUE | FALSE | -0.908049306 |
| **bcan** | LV cardiac output \| Instance 2_BSA_indexed_corrected | -0.134662751 | -0.01106668898 | -0.0931356055 | 6.74E-10 | 0.02176958735 | -0.0931356055 | 6.74E-10 | 4375 | 1.74E-08 | TRUE | 9.87E-06 | TRUE | 8.34E-06 | TRUE | FALSE | -0.134662751 |
| **bid** | LV end diastolic volume \| Instance 2(participant - p24100_i2)_BSA_indexed_corrected | -1.241438079 | 0.3271766915 | -0.07264061545 | 1.70E-06 | 0.2590316032 | -0.07264061545 | 1.70E-06 | 4332 | 2.33E-05 | TRUE | 0.02489136849 | TRUE | 0.02105311947 | TRUE | FALSE | -1.241438079 |
| **bid** | RV end diastolic volume \| Instance 2_BSA_indexed_corrected | -1.443356132 | 0.2165995953 | -0.07851537289 | 2.29E-07 | 0.2785044078 | -0.07851537289 | 2.29E-07 | 4332 | 3.70E-06 | TRUE | 0.003348351181 | TRUE | 0.002832035429 | TRUE | FALSE | -1.443356132 |
| **bid** | RV end systolic volume \| Instance 2_BSA_indexed_corrected | -0.783419962 | 0.2040984764 | -0.07158703592 | 2.40E-06 | 0.1658825214 | -0.07158703592 | 2.40E-06 | 4332 | 3.18E-05 | TRUE | 0.03511950198 | TRUE | 0.02970407478 | TRUE | FALSE | -0.783419962 |
| **bsg** | RV end diastolic volume \| Instance 2_BSA_indexed_corrected | -4.786799189 | 0.1416392904 | -0.07484151912 | 7.66E-07 | 0.9668040377 | -0.07484151912 | 7.66E-07 | 4354 | 1.12E-05 | TRUE | 0.01120161526 | TRUE | 0.009474326191 | TRUE | FALSE | -4.786799189 |
| **bst2** | RV end diastolic volume \| Instance 2_BSA_indexed_corrected | -1.752163017 | 0.2326657966 | -0.07218912655 | 1.81E-06 | 0.3665853518 | -0.07218912655 | 1.81E-06 | 4363 | 2.47E-05 | TRUE | 0.02653118073 | TRUE | 0.02244007266 | TRUE | FALSE | -1.752163017 |
| **btn2a1** | RV end diastolic volume \| Instance 2_BSA_indexed_corrected | -3.737786634 | 0.1034877343 | -0.07344360626 | 1.34E-06 | 0.7722255827 | -0.07344360626 | 1.34E-06 | 4322 | 1.86E-05 | TRUE | 0.01963103242 | TRUE | 0.01660392722 | TRUE | FALSE | -3.737786634 |
| **c19orf12** | LV end diastolic volume \| Instance 2(participant - p24100_i2)_BSA_indexed_corrected | -1.961313344 | 0.4547130154 | -0.1111062142 | 3.22E-13 | 0.2683767628 | -0.1111062142 | 3.22E-13 | 4275 | 1.52E-11 | TRUE | 4.71E-09 | TRUE | 3.98E-09 | TRUE | FALSE | -1.961313344 |
| **c19orf12** | LV end systolic volume \| Instance 2(participant - p24101_i2)_BSA_indexed_corrected | -0.8432151511 | 0.3400645571 | -0.08245184302 | 6.72E-08 | 0.1559158666 | -0.08245184302 | 6.72E-08 | 4275 | 1.19E-06 | TRUE | 0.0009825606283 | TRUE | 0.0008310497794 | TRUE | FALSE | -0.8432151511 |
| **c19orf12** | RV end diastolic volume \| Instance 2_BSA_indexed_corrected | -2.023115036 | 0.3402605834 | -0.1065821107 | 2.82E-12 | 0.2887280379 | -0.1065821107 | 2.82E-12 | 4275 | 1.13E-10 | TRUE | 4.12E-08 | TRUE | 3.49E-08 | TRUE | FALSE | -2.023115036 |
| **c19orf12** | RV end systolic volume \| Instance 2_BSA_indexed_corrected | -0.8563274307 | 0.2461604309 | -0.07599161033 | 6.55E-07 | 0.1718898219 | -0.07599161033 | 6.55E-07 | 4275 | 9.68E-06 | TRUE | 0.009578461099 | TRUE | 0.008101462398 | TRUE | FALSE | -0.8563274307 |
| **c2** | LV end diastolic volume \| Instance 2(participant - p24100_i2)_BSA_indexed_corrected | -4.170078506 | 0.1460199227 | -0.09224625881 | 1.23E-09 | 0.6848538516 | -0.09224625881 | 1.23E-09 | 4322 | 2.99E-08 | TRUE | 1.81E-05 | TRUE | 1.53E-05 | TRUE | FALSE | -4.170078506 |
| **c2** | RV end diastolic volume \| Instance 2_BSA_indexed_corrected | -4.371680112 | 0.02571356546 | -0.08996448557 | 3.13E-09 | 0.7363270342 | -0.08996448557 | 3.13E-09 | 4322 | 6.98E-08 | TRUE | 4.58E-05 | TRUE | 3.87E-05 | TRUE | FALSE | -4.371680112 |
| **c2** | RV end systolic volume \| Instance 2_BSA_indexed_corrected | -2.1924969 | 0.1192487083 | -0.07587981232 | 5.91E-07 | 0.4383459262 | -0.07587981232 | 5.91E-07 | 4322 | 8.83E-06 | TRUE | 0.00864151311 | TRUE | 0.007308991788 | TRUE | FALSE | -2.1924969 |
| **ca14** | LV end diastolic volume \| Instance 2(participant - p24100_i2)_BSA_indexed_corrected | 3.442069593 | 0.02665577473 | 0.1172411963 | 7.29E-15 | 0.4409041294 | 0.1172411963 | 7.29E-15 | 4375 | 4.54E-13 | TRUE | 1.07E-10 | TRUE | 9.02E-11 | TRUE | FALSE | 3.442069593 |
| **ca14** | LV end systolic volume \| Instance 2(participant - p24101_i2)_BSA_indexed_corrected | 1.783776328 | 0.1427832975 | 0.1046670542 | 3.92E-12 | 0.2563000199 | 0.1046670542 | 3.92E-12 | 4375 | 1.53E-10 | TRUE | 5.73E-08 | TRUE | 4.85E-08 | TRUE | FALSE | 1.783776328 |
| **ca14** | RV end diastolic volume \| Instance 2_BSA_indexed_corrected | 4.317502696 | -0.1659208967 | 0.1366927736 | 1.07E-19 | 0.4731529592 | 0.1366927736 | 1.07E-19 | 4375 | 1.49E-17 | TRUE | 1.57E-15 | TRUE | 1.33E-15 | TRUE | FALSE | 4.317502696 |
| **ca14** | RV end systolic volume \| Instance 2_BSA_indexed_corrected | 2.137856751 | 0.01442083248 | 0.1138160842 | 4.33E-14 | 0.2821979778 | 0.1138160842 | 4.33E-14 | 4375 | 2.38E-12 | TRUE | 6.34E-10 | TRUE | 5.36E-10 | TRUE | FALSE | 2.137856751 |
| **ca5a** | LV end diastolic volume \| Instance 2(participant - p24100_i2)_BSA_indexed_corrected | -1.456863958 | 0.2754231804 | -0.1250655947 | 1.02E-16 | 0.1747704793 | -0.1250655947 | 1.02E-16 | 4375 | 8.72E-15 | TRUE | 1.49E-12 | TRUE | 1.26E-12 | TRUE | FALSE | -1.456863958 |
| **ca5a** | LV end systolic volume \| Instance 2(participant - p24101_i2)_BSA_indexed_corrected | -0.6192799062 | 0.2789292172 | -0.09158299727 | 1.29E-09 | 0.1018247624 | -0.09158299727 | 1.29E-09 | 4375 | 3.10E-08 | TRUE | 1.89E-05 | TRUE | 1.60E-05 | TRUE | FALSE | -0.6192799062 |
| **ca5a** | RV end diastolic volume \| Instance 2_BSA_indexed_corrected | -1.504423136 | 0.163317584 | -0.1200442053 | 1.63E-15 | 0.1881425706 | -0.1200442053 | 1.63E-15 | 4375 | 1.14E-13 | TRUE | 2.38E-11 | TRUE | 2.02E-11 | TRUE | FALSE | -1.504423136 |
| **ca5a** | RV end systolic volume \| Instance 2_BSA_indexed_corrected | -0.7993216967 | 0.1745498914 | -0.1072518763 | 1.14E-12 | 0.1120507195 | -0.1072518763 | 1.14E-12 | 4375 | 4.80E-11 | TRUE | 1.67E-08 | TRUE | 1.41E-08 | TRUE | FALSE | -0.7993216967 |
| **ca6** | LV end diastolic volume \| Instance 2(participant - p24100_i2)_BSA_indexed_corrected | 1.271658002 | 0.2029488703 | 0.07634213321 | 5.28E-07 | 0.2531338337 | 0.07634213321 | 5.28E-07 | 4307 | 7.98E-06 | TRUE | 0.007717404823 | TRUE | 0.006527381 | TRUE | FALSE | 1.271658002 |
| **ca6** | RV end diastolic volume \| Instance 2_BSA_indexed_corrected | 1.743260095 | 0.07614116872 | 0.09722681203 | 1.62E-10 | 0.2719738173 | 0.09722681203 | 1.62E-10 | 4307 | 4.63E-09 | TRUE | 2.36E-06 | TRUE | 2.00E-06 | TRUE | FALSE | 1.743260095 |
| **calb1** | RV end diastolic volume \| Instance 2_BSA_indexed_corrected | -2.139906721 | 0.1619774292 | -0.07283855912 | 1.42E-06 | 0.4430866101 | -0.07283855912 | 1.42E-06 | 4375 | 1.95E-05 | TRUE | 0.02070478493 | TRUE | 0.01751210709 | TRUE | FALSE | -2.139906721 |
| **calb2** | LV end diastolic volume \| Instance 2(participant - p24100_i2)_BSA_indexed_corrected | -5.127022414 | 0.2170743202 | -0.1436828598 | 3.70E-21 | 0.5402116315 | -0.1436828598 | 3.70E-21 | 4275 | 6.22E-19 | TRUE | 5.41E-17 | TRUE | 4.58E-17 | TRUE | FALSE | -5.127022414 |
| **calb2** | LV end systolic volume \| Instance 2(participant - p24101_i2)_BSA_indexed_corrected | -2.324564935 | 0.2349006208 | -0.1124480761 | 1.66E-13 | 0.3142389147 | -0.1124480761 | 1.66E-13 | 4275 | 8.13E-12 | TRUE | 2.43E-09 | TRUE | 2.06E-09 | TRUE | FALSE | -2.324564935 |
| **calb2** | RV end diastolic volume \| Instance 2_BSA_indexed_corrected | -5.792047291 | 0.08259300938 | -0.1509538585 | 3.28E-23 | 0.5802511865 | -0.1509538585 | 3.28E-23 | 4275 | 7.49E-21 | TRUE | 4.79E-19 | TRUE | 4.05E-19 | TRUE | FALSE | -5.792047291 |
| **calb2** | RV end systolic volume \| Instance 2_BSA_indexed_corrected | -2.601533297 | 0.1333626317 | -0.11420985 | 6.90E-14 | 0.3461850439 | -0.11420985 | 6.90E-14 | 4275 | 3.66E-12 | TRUE | 1.01E-09 | TRUE | 8.54E-10 | TRUE | FALSE | -2.601533297 |
| **cant1** | LV end diastolic volume \| Instance 2(participant - p24100_i2)_BSA_indexed_corrected | -4.204222442 | 0.2257797815 | -0.07412915264 | 9.18E-07 | 0.855284041 | -0.07412915264 | 9.18E-07 | 4375 | 1.32E-05 | TRUE | 0.01342433068 | TRUE | 0.01135429889 | TRUE | FALSE | -4.204222442 |
| **cant1** | RV end diastolic volume \| Instance 2_BSA_indexed_corrected | -5.124110756 | 0.08836794849 | -0.08397967742 | 2.65E-08 | 0.9194279292 | -0.08397967742 | 2.65E-08 | 4375 | 5.09E-07 | TRUE | 0.0003880078309 | TRUE | 0.0003281770234 | TRUE | FALSE | -5.124110756 |
| **capg** | LV end diastolic volume \| Instance 2(participant - p24100_i2)_BSA_indexed_corrected | -1.70684325 | 0.0932758885 | -0.09189331492 | 1.54E-09 | 0.2819895779 | -0.09189331492 | 1.54E-09 | 4304 | 3.63E-08 | TRUE | 2.26E-05 | TRUE | 1.91E-05 | TRUE | FALSE | -1.70684325 |
| **capg** | RV end diastolic volume \| Instance 2_BSA_indexed_corrected | -1.883252296 | -0.0199531396 | -0.09420736213 | 5.92E-10 | 0.3034259138 | -0.09420736213 | 5.92E-10 | 4304 | 1.55E-08 | TRUE | 8.67E-06 | TRUE | 7.33E-06 | TRUE | FALSE | -1.883252296 |
| **cbln4** | LV end diastolic volume \| Instance 2(participant - p24100_i2)_BSA_indexed_corrected | -3.580376296 | 0.3527036817 | -0.1016722775 | 1.58E-11 | 0.5297610131 | -0.1016722775 | 1.58E-11 | 4375 | 5.50E-10 | TRUE | 2.31E-07 | TRUE | 1.95E-07 | TRUE | FALSE | -3.580376296 |
| **cbln4** | LV end systolic volume \| Instance 2(participant - p24101_i2)_BSA_indexed_corrected | -2.044209288 | 0.311733987 | -0.100001899 | 3.37E-11 | 0.3075706465 | -0.100001899 | 3.37E-11 | 4375 | 1.11E-09 | TRUE | 4.94E-07 | TRUE | 4.18E-07 | TRUE | FALSE | -2.044209288 |
| **cbln4** | RV end diastolic volume \| Instance 2_BSA_indexed_corrected | -3.434987262 | 0.2431437095 | -0.09066744009 | 1.88E-09 | 0.5705472065 | -0.09066744009 | 1.88E-09 | 4375 | 4.34E-08 | TRUE | 2.75E-05 | TRUE | 2.33E-05 | TRUE | FALSE | -3.434987262 |
| **cbln4** | RV end systolic volume \| Instance 2_BSA_indexed_corrected | -1.823648094 | 0.2169627852 | -0.08094301179 | 8.27E-08 | 0.3395819191 | -0.08094301179 | 8.27E-08 | 4375 | 1.45E-06 | TRUE | 0.001209841556 | TRUE | 0.001023283988 | TRUE | FALSE | -1.823648094 |
| **ccdc80** | RV end diastolic volume \| Instance 2_BSA_indexed_corrected | -2.966881207 | -0.06068867188 | -0.08871379528 | 4.16E-09 | 0.5037366164 | -0.08871379528 | 4.16E-09 | 4375 | 9.08E-08 | TRUE | 6.08E-05 | TRUE | 5.15E-05 | TRUE | FALSE | -2.966881207 |
| **ccl16** | LV end diastolic volume \| Instance 2(participant - p24100_i2)_BSA_indexed_corrected | -2.34148826 | -0.05851323713 | -0.1157163456 | 1.62E-14 | 0.3039346627 | -0.1157163456 | 1.62E-14 | 4375 | 9.65E-13 | TRUE | 2.37E-10 | TRUE | 2.01E-10 | TRUE | FALSE | -2.34148826 |
| **ccl16** | LV end systolic volume \| Instance 2(participant - p24101_i2)_BSA_indexed_corrected | -1.101316536 | 0.1183498681 | -0.09376135957 | 5.18E-10 | 0.1768402424 | -0.09376135957 | 5.18E-10 | 4375 | 1.37E-08 | TRUE | 7.57E-06 | TRUE | 6.41E-06 | TRUE | FALSE | -1.101316536 |
| **ccl16** | RV end diastolic volume \| Instance 2_BSA_indexed_corrected | -2.452026009 | -0.187513418 | -0.112636758 | 7.91E-14 | 0.3271015568 | -0.112636758 | 7.91E-14 | 4375 | 4.10E-12 | TRUE | 1.16E-09 | TRUE | 9.78E-10 | TRUE | FALSE | -2.452026009 |
| **ccl16** | RV end systolic volume \| Instance 2_BSA_indexed_corrected | -1.076799147 | 0.02786853799 | -0.08317673558 | 3.60E-08 | 0.1950902506 | -0.08317673558 | 3.60E-08 | 4375 | 6.71E-07 | TRUE | 0.0005261659715 | TRUE | 0.0004450311787 | TRUE | FALSE | -1.076799147 |
| **ccl18** | LV end diastolic volume \| Instance 2(participant - p24100_i2)_BSA_indexed_corrected | -1.235794426 | 0.3046152565 | -0.07945206365 | 2.14E-07 | 0.2378606325 | -0.07945206365 | 2.14E-07 | 4251 | 3.48E-06 | TRUE | 0.003127770617 | TRUE | 0.002645468388 | TRUE | FALSE | -1.235794426 |
| **ccl18** | RV end diastolic volume \| Instance 2_BSA_indexed_corrected | -1.426109164 | 0.1185945754 | -0.08533779762 | 2.51E-08 | 0.2554352354 | -0.08533779762 | 2.51E-08 | 4251 | 4.85E-07 | TRUE | 0.0003673113927 | TRUE | 0.000310671976 | TRUE | FALSE | -1.426109164 |
| **ccl19** | RV end diastolic volume \| Instance 2_BSA_indexed_corrected | -0.7933347708 | 0.3992430275 | -0.07957320621 | 1.42E-07 | 0.150493334 | -0.07957320621 | 1.42E-07 | 4363 | 2.38E-06 | TRUE | 0.002074654346 | TRUE | 0.001754742645 | TRUE | FALSE | -0.7933347708 |
| **ccl19** | RV end systolic volume \| Instance 2_BSA_indexed_corrected | -0.4633786109 | 0.3100617022 | -0.07821518664 | 2.31E-07 | 0.08943743813 | -0.07821518664 | 2.31E-07 | 4363 | 3.73E-06 | TRUE | 0.00337396506 | TRUE | 0.002853699647 | TRUE | FALSE | -0.4633786109 |
| **ccl20** | LV end diastolic volume \| Instance 2(participant - p24100_i2)_BSA_indexed_corrected | -0.9527279715 | 0.3044334524 | -0.07285173724 | 1.46E-06 | 0.1975513573 | -0.07285173724 | 1.46E-06 | 4361 | 2.01E-05 | TRUE | 0.02142836516 | TRUE | 0.01812411125 | TRUE | FALSE | -0.9527279715 |
| **ccl20** | RV end diastolic volume \| Instance 2_BSA_indexed_corrected | -1.513167263 | 0.1586942488 | -0.1075273214 | 1.08E-12 | 0.2119089815 | -0.1075273214 | 1.08E-12 | 4361 | 4.61E-11 | TRUE | 1.58E-08 | TRUE | 1.34E-08 | TRUE | FALSE | -1.513167263 |
| **ccl20** | RV end systolic volume \| Instance 2_BSA_indexed_corrected | -0.6927691472 | 0.1762540873 | -0.08276629884 | 4.41E-08 | 0.1263423455 | -0.08276629884 | 4.41E-08 | 4361 | 8.11E-07 | TRUE | 0.0006453931657 | TRUE | 0.0005458735396 | TRUE | FALSE | -0.6927691472 |

| | | | | | | | | | | | | | | | | | |
|---|---|---|---|---|---|---|---|---|---|---|---|---|---|---|---|---|---|
| **ccl22** | LV end diastolic volume \| Instance 2(participant - p24100_i2)_BSA_indexed_corrected | -2.478455667 | 0.09876638719 | -0.1158465668 | 3.55E-14 | 0.3260023129 | -0.1158465668 | 3.55E-14 | 4251 | 1.97E-12 | TRUE | 5.20E-10 | TRUE | 4.39E-10 | TRUE | FALSE | -2.478455667 |
| **ccl22** | LV end systolic volume \| Instance 2(participant - p24101_i2)_BSA_indexed_corrected | -1.012217463 | 0.1878502404 | -0.08169418294 | 9.62E-08 | 0.1894459134 | -0.08169418294 | 9.62E-08 | 4251 | 1.67E-06 | TRUE | 0.00140719834 | TRUE | 0.001190208356 | TRUE | FALSE | -1.012217463 |
| **ccl22** | RV end diastolic volume \| Instance 2_BSA_indexed_corrected | -3.019599875 | -0.05740714525 | -0.131388651 | 7.88E-18 | 0.3495158202 | -0.131388651 | 7.88E-18 | 4251 | 7.59E-16 | TRUE | 1.15E-13 | TRUE | 9.75E-14 | TRUE | FALSE | -3.019599875 |
| **ccl22** | RV end systolic volume \| Instance 2_BSA_indexed_corrected | -1.23890243 | 0.06171893661 | -0.09070533457 | 3.13E-09 | 0.2086733316 | -0.09070533457 | 3.13E-09 | 4251 | 6.98E-08 | TRUE | 4.58E-05 | TRUE | 3.88E-05 | TRUE | FALSE | -1.23890243 |
| **ccl23** | LV end diastolic volume \| Instance 2(participant - p24100_i2)_BSA_indexed_corrected | -1.915148113 | 0.2277230908 | -0.07126857914 | 2.73E-06 | 0.4078088497 | -0.07126857914 | 2.73E-06 | 4322 | 3.59E-05 | TRUE | 0.03998119021 | TRUE | 0.03381609068 | TRUE | FALSE | -1.915148113 |
| **ccl23** | RV end diastolic volume \| Instance 2_BSA_indexed_corrected | -2.307433482 | 0.09088783135 | -0.07985916441 | 1.46E-07 | 0.4382009016 | -0.07985916441 | 1.46E-07 | 4322 | 2.44E-06 | TRUE | 0.002142496026 | TRUE | 0.001812123139 | TRUE | FALSE | -2.307433482 |
| **ccl23** | RV end systolic volume \| Instance 2_BSA_indexed_corrected | -1.3380491 | 0.1249906222 | -0.07783590558 | 3.00E-07 | 0.2607537855 | -0.07783590558 | 3.00E-07 | 4322 | 4.75E-06 | TRUE | 0.004390665179 | TRUE | 0.003713624608 | TRUE | FALSE | -1.3380491 |
| **ccl27** | LV end diastolic volume \| Instance 2(participant - p24100_i2)_BSA_indexed_corrected | -1.338359172 | 0.1563329132 | -0.0779847517 | 2.41E-07 | 0.2587310838 | -0.0779847517 | 2.41E-07 | 4375 | 3.87E-06 | TRUE | 0.003525172055 | TRUE | 0.002981590524 | TRUE | FALSE | -1.338359172 |
| **ccl27** | RV end diastolic volume \| Instance 2_BSA_indexed_corrected | -1.44424258 | 0.0311999164 | -0.07822210577 | 2.21E-07 | 0.2783480372 | -0.07822210577 | 2.21E-07 | 4375 | 3.60E-06 | TRUE | 0.003239825271 | TRUE | 0.002740244214 | TRUE | FALSE | -1.44424258 |
| **ccl3** | RV end diastolic volume \| Instance 2_BSA_indexed_corrected | -1.718223799 | 0.1198471193 | -0.07851510649 | 2.29E-07 | 0.3315429399 | -0.07851510649 | 2.29E-07 | 4332 | 3.70E-06 | TRUE | 0.003348666966 | TRUE | 0.00283230252 | TRUE | FALSE | -1.718223799 |
| **ccl7** | LV end diastolic volume \| Instance 2(participant - p24100_i2)_BSA_indexed_corrected | -2.156465181 | 0.172187754 | -0.1155655673 | 2.35E-14 | 0.2816763719 | -0.1155655673 | 2.35E-14 | 4332 | 1.37E-12 | TRUE | 3.44E-10 | TRUE | 2.91E-10 | TRUE | FALSE | -2.156465181 |
| **ccl7** | LV end systolic volume \| Instance 2(participant - p24101_i2)_BSA_indexed_corrected | -0.912056291 | 0.2390842116 | -0.08425941738 | 2.79E-08 | 0.1639125004 | -0.08425941738 | 2.79E-08 | 4332 | 5.32E-07 | TRUE | 0.0004083404117 | TRUE | 0.0003453743202 | TRUE | FALSE | -0.912056291 |
| **ccl7** | RV end diastolic volume \| Instance 2_BSA_indexed_corrected | -2.506819384 | 0.0364372984 | -0.1248923372 | 1.58E-16 | 0.3026422282 | -0.1248923372 | 1.58E-16 | 4332 | 1.33E-14 | TRUE | 2.32E-12 | TRUE | 1.96E-12 | TRUE | FALSE | -2.506819384 |
| **ccl7** | RV end systolic volume \| Instance 2_BSA_indexed_corrected | -1.056668704 | 0.133755353 | -0.088432142 | 5.54E-09 | 0.1808755784 | -0.088432142 | 5.54E-09 | 4332 | 1.17E-07 | TRUE | 8.10E-05 | TRUE | 6.85E-05 | TRUE | FALSE | -1.056668704 |
| **ccn1** | LV end diastolic volume \| Instance 2(participant - p24100_i2)_BSA_indexed_corrected | -1.080595734 | 0.1246660056 | -0.07584653667 | 5.13E-07 | 0.2148742142 | -0.07584653667 | 5.13E-07 | 4373 | 7.78E-06 | TRUE | 0.007503458067 | TRUE | 0.006346424833 | TRUE | FALSE | -1.080595734 |
| **ccn5** | LV end diastolic volume \| Instance 2(participant - p24100_i2)_BSA_indexed_corrected | -1.849238602 | 0.2477565165 | -0.07189075663 | 2.30E-06 | 0.3908467103 | -0.07189075663 | 2.30E-06 | 4311 | 3.07E-05 | TRUE | 0.03367190853 | TRUE | 0.02847970023 | TRUE | FALSE | -1.849238602 |
| **ccn5** | RV end diastolic volume \| Instance 2_BSA_indexed_corrected | -2.041828879 | 0.1592708364 | -0.07373702609 | 1.26E-06 | 0.4206894236 | -0.07373702609 | 1.26E-06 | 4311 | 1.76E-05 | TRUE | 0.01837131251 | TRUE | 0.01553845612 | TRUE | FALSE | -2.041828879 |
| **cd14** | RV end diastolic volume \| Instance 2_BSA_indexed_corrected | -2.14271473 | 0.1302361265 | -0.07182513899 | 2.73E-06 | 0.4562648988 | -0.07182513899 | 2.73E-06 | 4255 | 3.59E-05 | TRUE | 0.03999705892 | TRUE | 0.03382951243 | TRUE | FALSE | -2.14271473 |
| **cd163** | LV end diastolic volume \| Instance 2(participant - p24100_i2)_BSA_indexed_corrected | -2.097493799 | 0.2343521216 | -0.08085420332 | 1.02E-07 | 0.3933523153 | -0.08085420332 | 1.02E-07 | 4323 | 1.76E-06 | TRUE | 0.00149077739 | TRUE | 0.001260899517 | TRUE | FALSE | -2.097493799 |
| **cd163** | RV end diastolic volume \| Instance 2_BSA_indexed_corrected | -2.539558307 | 0.09469600566 | -0.091075095 | 1.99E-09 | 0.4224329105 | -0.091075095 | 1.99E-09 | 4323 | 4.58E-08 | TRUE | 2.91E-05 | TRUE | 2.46E-05 | TRUE | FALSE | -2.539558307 |
| **cd22** | RV end diastolic volume \| Instance 2_BSA_indexed_corrected | -2.262546443 | 0.2102674335 | -0.07808229768 | 2.43E-07 | 0.4375457987 | -0.07808229768 | 2.43E-07 | 4361 | 3.91E-06 | TRUE | 0.003559351336 | TRUE | 0.00301049936 | TRUE | FALSE | -2.262546443 |
| **cd300e** | LV end diastolic volume \| Instance 2(participant - p24100_i2)_BSA_indexed_corrected | -2.863934985 | 0.1009371106 | -0.100914519 | 2.43E-11 | 0.4277529485 | -0.100914519 | 2.43E-11 | 4359 | 8.20E-10 | TRUE | 3.55E-07 | TRUE | 3.00E-07 | TRUE | FALSE | -2.863934985 |
| **cd300e** | LV end systolic volume \| Instance 2(participant - p24101_i2)_BSA_indexed_corrected | -1.229303259 | 0.2124663033 | -0.07462787588 | 8.11E-07 | 0.2488578988 | -0.07462787588 | 8.11E-07 | 4359 | 1.18E-05 | TRUE | 0.01187100693 | TRUE | 0.01004049766 | TRUE | FALSE | -1.229303259 |
| **cd300e** | RV end diastolic volume \| Instance 2_BSA_indexed_corrected | -3.676779709 | -0.07921218566 | -0.1205438784 | 1.40E-15 | 0.4587226574 | -0.1205438784 | 1.40E-15 | 4359 | 9.92E-14 | TRUE | 2.05E-11 | TRUE | 1.73E-11 | TRUE | FALSE | -3.676779709 |
| **cd300e** | RV end systolic volume \| Instance 2_BSA_indexed_corrected | -1.810488564 | 0.06228988884 | -0.09980318324 | 4.01E-11 | 0.2734537196 | -0.09980318324 | 4.01E-11 | 4359 | 1.31E-09 | TRUE | 5.86E-07 | TRUE | 4.96E-07 | TRUE | FALSE | -1.810488564 |
| **cd300lg** | LV end diastolic volume \| Instance 2(participant - p24100_i2)_BSA_indexed_corrected | 2.260103789 | 0.3803234132 | 0.0755330754 | 6.87E-07 | 0.4545278962 | 0.0755330754 | 6.87E-07 | 4311 | 1.01E-05 | TRUE | 0.01004984535 | TRUE | 0.008500159196 | TRUE | FALSE | 2.260103789 |
| **cd302** | LV end diastolic volume \| Instance 2(participant - p24100_i2)_BSA_indexed_corrected | -4.262163358 | 0.06600771367 | -0.1074832402 | 1.02E-12 | 0.596178587 | -0.1074832402 | 1.02E-12 | 4375 | 4.37E-11 | TRUE | 1.49E-08 | TRUE | 1.26E-08 | TRUE | FALSE | -4.262163358 |
| **cd302** | LV end systolic volume \| Instance 2(participant - p24101_i2)_BSA_indexed_corrected | -1.790460145 | 0.1913450125 | -0.07778293536 | 2.59E-07 | 0.3470346343 | -0.07778293536 | 2.59E-07 | 4375 | 4.14E-06 | TRUE | 0.003786755256 | TRUE | 0.003202837596 | TRUE | FALSE | -1.790460145 |
| **cd302** | RV end diastolic volume \| Instance 2_BSA_indexed_corrected | -5.007076962 | -0.09372621048 | -0.1173673423 | 6.82E-15 | 0.6406712582 | -0.1173673423 | 6.82E-15 | 4375 | 4.26E-13 | TRUE | 9.97E-11 | TRUE | 8.44E-11 | TRUE | FALSE | -5.007076962 |
| **cd302** | RV end systolic volume \| Instance 2_BSA_indexed_corrected | -2.111566012 | 0.07493186542 | -0.08322997539 | 3.52E-08 | 0.382318836 | -0.08322997539 | 3.52E-08 | 4375 | 6.59E-07 | TRUE | 0.0005156907148 | TRUE | 0.0004361712066 | TRUE | FALSE | -2.111566012 |
| **cd34** | RV end diastolic volume \| Instance 2_BSA_indexed_corrected | 3.749037272 | 0.252779451 | 0.07470692438 | 8.62E-07 | 0.7607643146 | 0.07470692438 | 8.62E-07 | 4329 | 1.25E-05 | TRUE | 0.01260912096 | TRUE | 0.01066479451 | TRUE | FALSE | 3.749037272 |
| **cd38** | LV cardiac output \| Instance 2_BSA_indexed_corrected | -0.09415531577 | -0.02163665028 | -0.0720510549 | 1.84E-06 | 0.01970990528 | -0.0720510549 | 1.84E-06 | 4375 | 2.50E-05 | TRUE | 0.0268762959 | TRUE | 0.02273197108 | TRUE | FALSE | -0.09415531577 |
| **cd38** | LV end diastolic volume \| Instance 2(participant - p24100_i2)_BSA_indexed_corrected | -3.468500901 | 0.2834017438 | -0.1184609792 | 3.81E-15 | 0.439650877 | -0.1184609792 | 3.81E-15 | 4375 | 2.51E-13 | TRUE | 5.58E-11 | TRUE | 4.72E-11 | TRUE | FALSE | -3.468500901 |
| **cd38** | LV end systolic volume \| Instance 2(participant - p24101_i2)_BSA_indexed_corrected | -1.583504995 | 0.2801305943 | -0.09316696642 | 6.65E-10 | 0.2559025773 | -0.09316696642 | 6.65E-10 | 4375 | 1.72E-08 | TRUE | 9.74E-06 | TRUE | 8.23E-06 | TRUE | FALSE | -1.583504995 |
| **cd38** | RV end diastolic volume \| Instance 2_BSA_indexed_corrected | -3.157185888 | 0.1800772654 | -0.1002272936 | 3.05E-11 | 0.4739494521 | -0.1002272936 | 3.05E-11 | 4375 | 1.01E-09 | TRUE | 4.46E-07 | TRUE | 3.77E-07 | TRUE | FALSE | -3.157185888 |
| **cd38** | RV end systolic volume \| Instance 2_BSA_indexed_corrected | -1.487568426 | 0.1872656711 | -0.07940994629 | 1.45E-07 | 0.2823830716 | -0.07940994629 | 1.45E-07 | 4375 | 2.42E-06 | TRUE | 0.002115798812 | TRUE | 0.001789542635 | TRUE | FALSE | -1.487568426 |
| **cd4** | LV end diastolic volume \| Instance 2(participant - p24100_i2)_BSA_indexed_corrected | -4.992854939 | 0.04412415133 | -0.1222352644 | 7.00E-16 | 0.6162969309 | -0.1222352644 | 7.00E-16 | 4329 | 5.31E-14 | TRUE | 1.02E-11 | TRUE | 8.67E-12 | TRUE | FALSE | -4.992854939 |
| **cd4** | LV end systolic volume \| Instance 2(participant - p24101_i2)_BSA_indexed_corrected | -2.117715328 | 0.1836918989 | -0.08941074979 | 3.79E-09 | 0.3586257615 | -0.08941074979 | 3.79E-09 | 4329 | 8.35E-08 | TRUE | 5.55E-05 | TRUE | 4.70E-05 | TRUE | FALSE | -2.117715328 |
| **cd4** | RV end diastolic volume \| Instance 2_BSA_indexed_corrected | -5.156008681 | -0.07511412354 | -0.1173803445 | 9.36E-15 | 0.663150534 | -0.1173803445 | 9.36E-15 | 4329 | 5.68E-13 | TRUE | 1.37E-10 | TRUE | 1.16E-10 | TRUE | FALSE | -5.156008681 |
| **cd4** | RV end systolic volume \| Instance 2_BSA_indexed_corrected | -2.193307003 | 0.08111473153 | -0.08391758155 | 3.21E-08 | 0.3959300118 | -0.08391758155 | 3.21E-08 | 4329 | 6.05E-07 | TRUE | 0.0004697559813 | TRUE | 0.000397319609 | TRUE | FALSE | -2.193307003 |
| **cd5** | LV end diastolic volume \| Instance 2(participant - p24100_i2)_BSA_indexed_corrected | -2.439160336 | 0.2428172544 | -0.07428522698 | 8.75E-07 | 0.4952744112 | -0.07428522698 | 8.75E-07 | 4373 | 1.26E-05 | TRUE | 0.01280621741 | TRUE | 0.01083149869 | TRUE | FALSE | -2.439160336 |
| **cd5** | RV end diastolic volume \| Instance 2_BSA_indexed_corrected | -2.920862127 | 0.112459714 | -0.08269091774 | 4.35E-08 | 0.5324427722 | -0.08269091774 | 4.35E-08 | 4373 | 8.01E-07 | TRUE | 0.0006362718357 | TRUE | 0.0005381587186 | TRUE | FALSE | -2.920862127 |
| **cd59** | LV cardiac output \| Instance 2_BSA_indexed_corrected | -0.1807442299 | -0.02768968763 | -0.07997211235 | 2.06E-07 | 0.034741586 | -0.07997211235 | 2.06E-07 | 4207 | 3.36E-06 | TRUE | 0.003013116903 | TRUE | 0.002548494276 | TRUE | FALSE | -0.1807442299 |
| **cd59** | LV end diastolic volume \| Instance 2(participant - p24100_i2)_BSA_indexed_corrected | -4.821483611 | 0.2132221334 | -0.09478381399 | 7.28E-10 | 0.78091516 | -0.09478381399 | 7.28E-10 | 4207 | 1.86E-08 | TRUE | 1.07E-05 | TRUE | 9.01E-06 | TRUE | FALSE | -4.821483611 |
| **cd59** | LV end systolic volume \| Instance 2(participant - p24101_i2)_BSA_indexed_corrected | -2.227080043 | 0.2557684826 | -0.0752189768 | 1.04E-06 | 0.45529546 | -0.0752189768 | 1.04E-06 | 4207 | 1.48E-05 | TRUE | 0.01518719621 | TRUE | 0.01284533055 | TRUE | FALSE | -2.227080043 |
| **cd59** | RV end diastolic volume \| Instance 2_BSA_indexed_corrected | -5.231859498 | 0.06929695042 | -0.09546598168 | 5.49E-10 | 0.8412717971 | -0.09546598168 | 5.49E-10 | 4207 | 1.45E-08 | TRUE | 8.03E-06 | TRUE | 6.79E-06 | TRUE | FALSE | -5.231859498 |
| **cd59** | RV end systolic volume \| Instance 2_BSA_indexed_corrected | -2.391210297 | 0.134543613 | -0.07341295028 | 1.87E-06 | 0.5009433581 | -0.07341295028 | 1.87E-06 | 4207 | 2.53E-05 | TRUE | 0.02739948893 | TRUE | 0.0231744885 | TRUE | FALSE | -2.391210297 |
| **cd6** | LV end diastolic volume \| Instance 2(participant - p24100_i2)_BSA_indexed_corrected | -1.491497411 | 0.3195943574 | -0.07228914646 | 1.80E-06 | 0.3119372487 | -0.07228914646 | 1.80E-06 | 4354 | 2.45E-05 | TRUE | 0.02630864068 | TRUE | 0.02225184829 | TRUE | FALSE | -1.491497411 |
| **cd74** | LV end diastolic volume \| Instance 2(participant - p24100_i2)_BSA_indexed_corrected | -2.705018122 | 0.1184420437 | -0.08241932681 | 4.99E-08 | 0.4952995922 | -0.08241932681 | 4.99E-08 | 4363 | 9.06E-07 | TRUE | 0.0007296462769 | TRUE | 0.000617134821 | TRUE | FALSE | -2.705018122 |
| **cd74** | RV end diastolic volume \| Instance 2_BSA_indexed_corrected | -3.35570602 | -0.05361625637 | -0.09496479114 | 3.28E-10 | 0.5326734626 | -0.09496479114 | 3.28E-10 | 4363 | 9.01E-09 | TRUE | 4.79E-06 | TRUE | 4.05E-06 | TRUE | FALSE | -3.35570602 |
| **cd74** | RV end systolic volume \| Instance 2_BSA_indexed_corrected | -1.534051869 | 0.08426442611 | -0.07305727836 | 1.36E-06 | 0.3171183794 | -0.07305727836 | 1.36E-06 | 4363 | 1.88E-05 | TRUE | 0.01990204621 | TRUE | 0.01683315068 | TRUE | FALSE | -1.534051869 |
| **cd83** | LV end diastolic volume \| Instance 2(participant - p24100_i2)_BSA_indexed_corrected | -3.340461362 | 0.1606264826 | -0.09379282079 | 6.21E-10 | 0.5388578211 | -0.09379282079 | 6.21E-10 | 4332 | 1.61E-08 | TRUE | 9.08E-06 | TRUE | 7.68E-06 | TRUE | FALSE | -3.340461362 |
| **cd83** | RV end diastolic volume \| Instance 2_BSA_indexed_corrected | -4.761953952 | -0.03126353434 | -0.1243011308 | 2.20E-16 | 0.5776767412 | -0.1243011308 | 2.20E-16 | 4332 | 1.82E-14 | TRUE | 3.22E-12 | TRUE | 2.72E-12 | TRUE | FALSE | -4.761953952 |
| **cd83** | RV end systolic volume \| Instance 2_BSA_indexed_corrected | -2.145368853 | 0.09668980142 | -0.09406984107 | 5.52E-10 | 0.3450463953 | -0.09406984107 | 5.52E-10 | 4332 | 1.45E-08 | TRUE | 8.08E-06 | TRUE | 6.84E-06 | TRUE | FALSE | -2.145368853 |
| **cd8a** | LV end diastolic volume \| Instance 2(participant - p24100_i2)_BSA_indexed_corrected | -1.773552996 | 0.3222603987 | -0.07740285784 | 3.07E-07 | 0.3459308111 | -0.07740285784 | 3.07E-07 | 4363 | 4.85E-06 | TRUE | 0.004495957454 | TRUE | 0.003802680815 | TRUE | FALSE | -1.773552996 |
| **cd8a** | RV end diastolic volume \| Instance 2_BSA_indexed_corrected | -1.904741841 | 0.2062288423 | -0.07720915046 | 3.29E-07 | 0.372456846 | -0.07720915046 | 3.29E-07 | 4363 | 5.15E-06 | TRUE | 0.004812509112 | TRUE | 0.004070420207 | TRUE | FALSE | -1.904741841 |
| **cd99l2** | LV end diastolic volume \| Instance 2(participant - p24100_i2)_BSA_indexed_corrected | -5.363915523 | 0.1374469428 | -0.1123509366 | 1.38E-13 | 0.7227008193 | -0.1123509366 | 1.38E-13 | 4311 | 6.85E-12 | TRUE | 2.02E-09 | TRUE | 1.71E-09 | TRUE | FALSE | -5.363915523 |
| **cd99l2** | LV end systolic volume \| Instance 2(participant - p24101_i2)_BSA_indexed_corrected | -2.315244372 | 0.2240547508 | -0.08347892505 | 4.04E-08 | 0.4210301571 | -0.08347892505 | 4.04E-08 | 4311 | 7.46E-07 | TRUE | 0.0005911344359 | TRUE | 0.0004999815059 | TRUE | FALSE | -2.315244372 |
| **cd99l2** | RV end diastolic volume \| Instance 2_BSA_indexed_corrected | -6.943255648 | -0.002042089356 | -0.1350964995 | 5.17E-19 | 0.7757665199 | -0.1350964995 | 5.17E-19 | 4311 | 6.05E-17 | TRUE | 7.57E-15 | TRUE | 6.40E-15 | TRUE | FALSE | -6.943255648 |
| **cd99l2** | RV end systolic volume \| Instance 2_BSA_indexed_corrected | -3.337644632 | 0.0986227868 | -0.1092141251 | 6.47E-13 | 0.4627723395 | -0.1092141251 | 6.47E-13 | 4311 | 2.88E-11 | TRUE | 9.46E-09 | TRUE | 8.00E-09 | TRUE | FALSE | -3.337644632 |
| **cdcp1** | LV end diastolic volume \| Instance 2(participant - p24100_i2)_BSA_indexed_corrected | -2.010489018 | 0.009518634759 | -0.09485150487 | 3.25E-10 | 0.3190841763 | -0.09485150487 | 3.25E-10 | 4375 | 8.98E-09 | TRUE | 4.76E-06 | TRUE | 4.03E-06 | TRUE | FALSE | -2.010489018 |
| **cdcp1** | RV end diastolic volume \| Instance 2_BSA_indexed_corrected | -2.280919781 | -0.1462576683 | -0.1000241515 | 3.34E-11 | 0.3431088614 | -0.1000241515 | 3.34E-11 | 4375 | 1.10E-09 | TRUE | 4.89E-07 | TRUE | 4.13E-07 | TRUE | FALSE | -2.280919781 |
| **cdh15** | LV end diastolic volume \| Instance 2(participant - p24100_i2)_BSA_indexed_corrected | -1.568953408 | 0.5000434819 | -0.08843053918 | 4.89E-09 | 0.2676147049 | -0.08843053918 | 4.89E-09 | 4363 | 1.05E-07 | TRUE | 7.15E-05 | TRUE | 6.05E-05 | TRUE | FALSE | -1.568953408 |
| **cdh15** | RV end diastolic volume \| Instance 2_BSA_indexed_corrected | -1.83497128 | 0.4101395388 | -0.09605972163 | 2.05E-10 | 0.2879266621 | -0.09605972163 | 2.05E-10 | 4363 | 5.79E-09 | TRUE | 2.99E-06 | TRUE | 2.53E-06 | TRUE | FALSE | -1.83497128 |
| **cdh15** | RV end systolic volume \| Instance 2_BSA_indexed_corrected | -0.869465226 | 0.2989182347 | -0.07659651393 | 4.07E-07 | 0.1713848422 | -0.07659651393 | 4.07E-07 | 4363 | 6.28E-06 | TRUE | 0.005961639559 | TRUE | 0.005042354739 | TRUE | FALSE | -0.869465226 |
| **cdh2** | LV end diastolic volume \| Instance 2(participant - p24100_i2)_BSA_indexed_corrected | -2.091842869 | 0.1453552008 | -0.07165473665 | 2.09E-06 | 0.4403283247 | -0.07165473665 | 2.09E-06 | 4375 | 2.82E-05 | TRUE | 0.03061618896 | TRUE | 0.02589517263 | TRUE | FALSE | -2.091842869 |
| **cdh2** | RV end diastolic volume \| Instance 2_BSA_indexed_corrected | -2.556050705 | -0.01029991156 | -0.0813837419 | 7.03E-08 | 0.4733681719 | -0.0813837419 | 7.03E-08 | 4375 | 1.24E-06 | TRUE | 0.001028312635 | TRUE | 0.0008697468268 | TRUE | FALSE | -2.556050705 |
| **cdhr2** | LV end diastolic volume \| Instance 2(participant - p24100_i2)_BSA_indexed_corrected | -1.439483924 | -0.002722549944 | -0.1242021299 | 1.65E-16 | 0.1739050449 | -0.1242021299 | 1.65E-16 | 4375 | 1.38E-14 | TRUE | 2.42E-12 | TRUE | 2.05E-12 | TRUE | FALSE | -1.439483924 |

| | | | | | | | | | | | | | | | | | |
|---|---|---|---|---|---|---|---|---|---|---|---|---|---|---|---|---|---|
| **cdhr2** | LV end systolic volume \| Instance 2(participant - p24101_i2)_BSA_indexed_corrected | -0.7126524532 | 0.135794984 | -0.1059275725 | 2.15E-12 | 0.1011646352 | -0.1059275725 | 2.15E-12 | 4375 | 8.77E-11 | TRUE | 3.15E-08 | TRUE | 2.66E-08 | TRUE | FALSE | -0.7126524532 |
| **cdhr2** | RV end diastolic volume \| Instance 2_BSA_indexed_corrected | -1.499912604 | -0.1272288208 | -0.1202930463 | 1.42E-15 | 0.1871847765 | -0.1202930463 | 1.42E-15 | 4375 | 1.00E-13 | TRUE | 2.08E-11 | TRUE | 1.76E-11 | TRUE | FALSE | -1.499912604 |
| **cdhr2** | RV end systolic volume \| Instance 2_BSA_indexed_corrected | -0.7858326364 | 0.02291968464 | -0.1059782466 | 2.10E-12 | 0.1114989887 | -0.1059782466 | 2.10E-12 | 4375 | 8.59E-11 | TRUE | 3.07E-08 | TRUE | 2.60E-08 | TRUE | FALSE | -0.7858326364 |
| **cdhr5** | LV end diastolic volume \| Instance 2(participant - p24100_i2)_BSA_indexed_corrected | -3.910520838 | 0.01828038769 | -0.1435992594 | 1.36E-21 | 0.4075380053 | -0.1435992594 | 1.36E-21 | 4375 | 2.40E-19 | TRUE | 1.99E-17 | TRUE | 1.68E-17 | TRUE | FALSE | -3.910520838 |
| **cdhr5** | LV end systolic volume \| Instance 2(participant - p24101_i2)_BSA_indexed_corrected | -1.751986585 | 0.1619443662 | -0.1108300127 | 1.96E-13 | 0.237574517 | -0.1108300127 | 1.96E-13 | 4375 | 9.51E-12 | TRUE | 2.87E-09 | TRUE | 2.43E-09 | TRUE | FALSE | -1.751986585 |
| **cdhr5** | RV end diastolic volume \| Instance 2_BSA_indexed_corrected | -3.752209099 | -0.07774098313 | -0.1280728199 | 1.83E-17 | 0.4393888148 | -0.1280728199 | 1.83E-17 | 4375 | 1.71E-15 | TRUE | 2.68E-13 | TRUE | 2.27E-13 | TRUE | FALSE | -3.752209099 |
| **cdhr5** | RV end systolic volume \| Instance 2_BSA_indexed_corrected | -1.48702954 | 0.08983390507 | -0.0853496286 | 1.57E-08 | 0.2625068317 | -0.0853496286 | 1.57E-08 | 4375 | 3.13E-07 | TRUE | 0.000229263448 | TRUE | 0.0001939110244 | TRUE | FALSE | -1.48702954 |
| **cdon** | LV end diastolic volume \| Instance 2(participant - p24100_i2)_BSA_indexed_corrected | -3.078337887 | 0.3130544998 | -0.07414977017 | 1.10E-06 | 0.630843789 | -0.07414977017 | 1.10E-06 | 4309 | 1.56E-05 | TRUE | 0.01610454523 | TRUE | 0.01362122435 | TRUE | FALSE | -3.078337887 |
| **ceacam8** | LV end diastolic volume \| Instance 2(participant - p24100_i2)_BSA_indexed_corrected | -2.149721007 | 0.1745553808 | -0.09217896343 | 1.01E-09 | 0.3511620425 | -0.09217896343 | 1.01E-09 | 4375 | 2.49E-08 | TRUE | 1.47E-05 | TRUE | 1.25E-05 | TRUE | FALSE | -2.149721007 |
| **ceacam8** | RV end diastolic volume \| Instance 2_BSA_indexed_corrected | -2.157874557 | 0.06430589473 | -0.08600590264 | 1.21E-08 | 0.3780036435 | -0.08600590264 | 1.21E-08 | 4375 | 2.46E-07 | TRUE | 0.0001776694201 | TRUE | 0.0001502727955 | TRUE | FALSE | -2.157874557 |
| **ceacam8** | RV end systolic volume \| Instance 2_BSA_indexed_corrected | -1.096744776 | 0.1260748158 | -0.0735054834 | 1.13E-06 | 0.2250192423 | -0.0735054834 | 1.13E-06 | 4375 | 1.60E-05 | TRUE | 0.01656594157 | TRUE | 0.01401147338 | TRUE | FALSE | -1.096744776 |
| **cebpb** | LV end diastolic volume \| Instance 2(participant - p24100_i2)_BSA_indexed_corrected | -2.426079626 | 0.2718924316 | -0.08467509181 | 2.03E-08 | 0.4317149715 | -0.08467509181 | 2.03E-08 | 4375 | 3.98E-07 | TRUE | 0.0002973648091 | TRUE | 0.0002515111556 | TRUE | FALSE | -2.426079626 |
| **cebpb** | LV end systolic volume \| Instance 2(participant - p24101_i2)_BSA_indexed_corrected | -1.214674876 | 0.2712957817 | -0.07303309154 | 1.33E-06 | 0.2508356717 | -0.07303309154 | 1.33E-06 | 4375 | 1.85E-05 | TRUE | 0.01940462537 | TRUE | 0.01641243214 | TRUE | FALSE | -1.214674876 |
| **ces1** | LV end diastolic volume \| Instance 2(participant - p24100_i2)_BSA_indexed_corrected | -2.24789591 | 0.3258656445 | -0.1621866775 | 5.59E-27 | 0.2076001833 | -0.1621866775 | 5.59E-27 | 4342 | 2.15E-24 | TRUE | 8.18E-23 | TRUE | 6.92E-23 | TRUE | FALSE | -2.24789591 |
| **ces1** | LV end systolic volume \| Instance 2(participant - p24101_i2)_BSA_indexed_corrected | -0.9742725679 | 0.2979677973 | -0.1208886781 | 1.32E-15 | 0.1214375953 | -0.1208886781 | 1.32E-15 | 4342 | 9.41E-14 | TRUE | 1.93E-11 | TRUE | 1.63E-11 | TRUE | FALSE | -0.9742725679 |
| **ces1** | RV end diastolic volume \| Instance 2_BSA_indexed_corrected | -2.483473646 | 0.220360272 | -0.1664690572 | 2.36E-28 | 0.2232948187 | -0.1664690572 | 2.36E-28 | 4342 | 1.12E-25 | TRUE | 3.46E-24 | TRUE | 2.92E-24 | TRUE | FALSE | -2.483473646 |
| **ces1** | RV end systolic volume \| Instance 2_BSA_indexed_corrected | -1.150326413 | 0.2054923073 | -0.1295139333 | 1.05E-17 | 0.133686158 | -0.1295139333 | 1.05E-17 | 4342 | 1.00E-15 | TRUE | 1.54E-13 | TRUE | 1.30E-13 | TRUE | FALSE | -1.150326413 |
| **ces2** | LV end diastolic volume \| Instance 2(participant - p24100_i2)_BSA_indexed_corrected | -1.64901934 | 0.3390004997 | -0.08209434072 | 5.40E-08 | 0.3027292449 | -0.08209434072 | 5.40E-08 | 4375 | 9.74E-07 | TRUE | 0.0007898047977 | TRUE | 0.0006680168979 | TRUE | FALSE | -1.64901934 |
| **ces2** | RV end diastolic volume \| Instance 2_BSA_indexed_corrected | -1.700451707 | 0.2289907729 | -0.07868720457 | 1.88E-07 | 0.3257780069 | -0.07868720457 | 1.88E-07 | 4375 | 3.08E-06 | TRUE | 0.002743985619 | TRUE | 0.002320863036 | TRUE | FALSE | -1.700451707 |
| **cfc1** | LV end diastolic volume \| Instance 2(participant - p24100_i2)_BSA_indexed_corrected | -2.10622219 | 0.3479660122 | -0.0919081166 | 1.13E-09 | 0.3450789936 | -0.0919081166 | 1.13E-09 | 4375 | 2.75E-08 | TRUE | 1.65E-05 | TRUE | 1.39E-05 | TRUE | FALSE | -2.10622219 |
| **cfc1** | RV end diastolic volume \| Instance 2_BSA_indexed_corrected | -2.45422611 | 0.2375591564 | -0.09954434626 | 4.15E-11 | 0.3709759658 | -0.09954434626 | 4.15E-11 | 4375 | 1.35E-09 | TRUE | 6.07E-07 | TRUE | 5.13E-07 | TRUE | FALSE | -2.45422611 |
| **cfc1** | RV end systolic volume \| Instance 2_BSA_indexed_corrected | -1.047428702 | 0.2146104824 | -0.07143953138 | 2.25E-06 | 0.2211490423 | -0.07143953138 | 2.25E-06 | 4375 | 3.00E-05 | TRUE | 0.03285134691 | TRUE | 0.02778566922 | TRUE | FALSE | -1.047428702 |
| **cgref1** | RV end diastolic volume \| Instance 2_BSA_indexed_corrected | -2.046777235 | 0.1665702976 | -0.07047293027 | 3.07E-06 | 0.438104158 | -0.07047293027 | 3.07E-06 | 4375 | 4.00E-05 | TRUE | 0.04497187661 | TRUE | 0.03803721324 | TRUE | FALSE | -2.046777235 |
| **chi3l1** | RV end diastolic volume \| Instance 2_BSA_indexed_corrected | -1.287203465 | 0.136259353 | -0.07709971496 | 4.66E-07 | 0.2550022464 | -0.07709971496 | 4.66E-07 | 4263 | 7.09E-06 | TRUE | 0.006810920646 | TRUE | 0.005760676683 | TRUE | FALSE | -1.287203465 |
| **chrdl1** | LV end diastolic volume \| Instance 2(participant - p24100_i2)_BSA_indexed_corrected | -2.660286365 | 0.1238241461 | -0.0777101259 | 3.24E-07 | 0.5198724976 | -0.0777101259 | 3.24E-07 | 4312 | 5.07E-06 | TRUE | 0.004734512987 | TRUE | 0.004004451084 | TRUE | FALSE | -2.660286365 |
| **chrdl1** | RV end diastolic volume \| Instance 2_BSA_indexed_corrected | -3.38247961 | -0.04797728387 | -0.09198096435 | 1.44E-09 | 0.5577680123 | -0.09198096435 | 1.44E-09 | 4312 | 3.41E-08 | TRUE | 2.10E-05 | TRUE | 1.78E-05 | TRUE | FALSE | -3.38247961 |
| **chrdl1** | RV end systolic volume \| Instance 2_BSA_indexed_corrected | -1.566028282 | 0.07676650407 | -0.07157027729 | 2.55E-06 | 0.3324398772 | -0.07157027729 | 2.55E-06 | 4312 | 3.36E-05 | TRUE | 0.0372514544 | TRUE | 0.03150728013 | TRUE | FALSE | -1.566028282 |
| **ckap4** | RV end diastolic volume \| Instance 2_BSA_indexed_corrected | -3.61986306 | 0.1466183202 | -0.07142054559 | 2.39E-06 | 0.766326872 | -0.07142054559 | 2.39E-06 | 4354 | 3.17E-05 | TRUE | 0.03495719778 | TRUE | 0.02956679788 | TRUE | FALSE | -3.61986306 |
| **clec4a** | LV end diastolic volume \| Instance 2(participant - p24100_i2)_BSA_indexed_corrected | 3.934948654 | 0.3473867049 | 0.1254919038 | 1.13E-16 | 0.4727510681 | 0.1254919038 | 1.13E-16 | 4332 | 9.63E-15 | TRUE | 1.66E-12 | TRUE | 1.40E-12 | TRUE | FALSE | 3.934948654 |
| **clec4a** | LV end systolic volume \| Instance 2(participant - p24101_i2)_BSA_indexed_corrected | 1.870736577 | 0.3121596261 | 0.1028490013 | 1.16E-11 | 0.2749535999 | 0.1028490013 | 1.16E-11 | 4332 | 4.14E-10 | TRUE | 1.69E-07 | TRUE | 1.43E-07 | TRUE | FALSE | 1.870736577 |
| **clec4a** | RV end diastolic volume \| Instance 2_BSA_indexed_corrected | 4.307178843 | 0.2414238072 | 0.1277015846 | 3.25E-17 | 0.5083728019 | 0.1277015846 | 3.25E-17 | 4332 | 2.95E-15 | TRUE | 4.76E-13 | TRUE | 4.02E-13 | TRUE | FALSE | 4.307178843 |
| **clec4a** | RV end systolic volume \| Instance 2_BSA_indexed_corrected | 1.97795647 | 0.2193559544 | 0.09850962418 | 8.16E-11 | 0.3036523172 | 0.09850962418 | 8.16E-11 | 4332 | 2.49E-09 | TRUE | 1.19E-06 | TRUE | 1.01E-06 | TRUE | FALSE | 1.97795647 |
| **clec4d** | LV end diastolic volume \| Instance 2(participant - p24100_i2)_BSA_indexed_corrected | -1.742630294 | 0.1689106843 | -0.09328795081 | 7.67E-10 | 0.2826426735 | -0.09328795081 | 7.67E-10 | 4332 | 1.95E-08 | TRUE | 1.12E-05 | TRUE | 9.49E-06 | TRUE | FALSE | -1.742630294 |
| **clec4d** | LV end systolic volume \| Instance 2(participant - p24101_i2)_BSA_indexed_corrected | -0.8386685202 | 0.2261511235 | -0.07739652093 | 3.39E-07 | 0.164180012 | -0.07739652093 | 3.39E-07 | 4332 | 5.30E-06 | TRUE | 0.004962303488 | TRUE | 0.004197116291 | TRUE | FALSE | -0.8386685202 |
| **clec4d** | RV end diastolic volume \| Instance 2_BSA_indexed_corrected | -1.843771137 | 0.05330206448 | -0.09176013618 | 1.44E-09 | 0.3040695356 | -0.09176013618 | 1.44E-09 | 4332 | 3.41E-08 | TRUE | 2.11E-05 | TRUE | 1.79E-05 | TRUE | FALSE | -1.843771137 |
| **clec4d** | RV end systolic volume \| Instance 2_BSA_indexed_corrected | -0.8656541566 | 0.1308130247 | -0.07236857268 | 1.86E-06 | 0.181305187 | -0.07236857268 | 1.86E-06 | 4332 | 2.52E-05 | TRUE | 0.02721740574 | TRUE | 0.02302048178 | TRUE | FALSE | -0.8656541566 |
| **clec4g** | LV end diastolic volume \| Instance 2(participant - p24100_i2)_BSA_indexed_corrected | -3.644854205 | 0.214763543 | -0.1086472265 | 7.49E-13 | 0.5068028706 | -0.1086472265 | 7.49E-13 | 4332 | 3.30E-11 | TRUE | 1.10E-08 | TRUE | 9.27E-09 | TRUE | FALSE | -3.644854205 |
| **clec4g** | LV end systolic volume \| Instance 2(participant - p24101_i2)_BSA_indexed_corrected | -1.925058722 | 0.2410851413 | -0.09892208654 | 6.80E-11 | 0.2942872107 | -0.09892208654 | 6.80E-11 | 4332 | 2.09E-09 | TRUE | 9.95E-07 | TRUE | 8.42E-07 | TRUE | FALSE | -1.925058722 |
| **clec4g** | RV end diastolic volume \| Instance 2_BSA_indexed_corrected | -3.661777289 | 0.1099390165 | -0.1014745545 | 2.17E-11 | 0.545560916 | -0.1014745545 | 2.17E-11 | 4332 | 7.43E-10 | TRUE | 3.17E-07 | TRUE | 2.68E-07 | TRUE | FALSE | -3.661777289 |
| **clec4g** | RV end systolic volume \| Instance 2_BSA_indexed_corrected | -2.231432393 | 0.1360258304 | -0.1038741567 | 7.21E-12 | 0.3246957156 | -0.1038741567 | 7.21E-12 | 4332 | 2.69E-10 | TRUE | 1.06E-07 | TRUE | 8.93E-08 | TRUE | FALSE | -2.231432393 |
| **clec6a** | LV end diastolic volume \| Instance 2(participant - p24100_i2)_BSA_indexed_corrected | -1.789265746 | 0.2110219894 | -0.07312211676 | 1.29E-06 | 0.3690389336 | -0.07312211676 | 1.29E-06 | 4375 | 1.80E-05 | TRUE | 0.01883615169 | TRUE | 0.0159316171 | TRUE | FALSE | -1.789265746 |
| **clec6a** | RV end diastolic volume \| Instance 2_BSA_indexed_corrected | -1.880333138 | 0.09422241935 | -0.07142677658 | 2.25E-06 | 0.3970757236 | -0.07142677658 | 2.25E-06 | 4375 | 3.01E-05 | TRUE | 0.03298862404 | TRUE | 0.02790177821 | TRUE | FALSE | -1.880333138 |
| **clmp** | LV end diastolic volume \| Instance 2(participant - p24100_i2)_BSA_indexed_corrected | -7.849058528 | -0.06353204194 | -0.1575098205 | 1.06E-25 | 0.7441576627 | -0.1575098205 | 1.06E-25 | 4375 | 3.52E-23 | TRUE | 1.55E-21 | TRUE | 1.31E-21 | TRUE | FALSE | -7.849058528 |
| **clmp** | LV end systolic volume \| Instance 2(participant - p24101_i2)_BSA_indexed_corrected | -3.625815662 | 0.1194908565 | -0.1253443609 | 8.71E-17 | 0.4339827339 | -0.1253443609 | 8.71E-17 | 4375 | 7.54E-15 | TRUE | 1.27E-12 | TRUE | 1.08E-12 | TRUE | FALSE | -3.625815662 |
| **clmp** | RV end diastolic volume \| Instance 2_BSA_indexed_corrected | -9.190857122 | -0.2443134962 | -0.17143458 | 3.29E-30 | 0.7987121121 | -0.17143458 | 3.29E-30 | 4375 | 2.01E-27 | TRUE | 4.82E-26 | TRUE | 4.08E-26 | TRUE | FALSE | -9.190857122 |
| **clmp** | RV end systolic volume \| Instance 2_BSA_indexed_corrected | -4.153235394 | -0.00328953032 | -0.130268994 | 5.11E-18 | 0.4780123662 | -0.130268994 | 5.11E-18 | 4375 | 5.05E-16 | TRUE | 7.47E-14 | TRUE | 6.32E-14 | TRUE | FALSE | -4.153235394 |
| **cntn3** | LV end diastolic volume \| Instance 2(participant - p24100_i2)_BSA_indexed_corrected | -3.826699935 | 0.1772706005 | -0.1240896851 | 2.92E-16 | 0.4661554663 | -0.1240896851 | 2.92E-16 | 4311 | 2.34E-14 | TRUE | 4.27E-12 | TRUE | 3.61E-12 | TRUE | FALSE | -3.826699935 |
| **cntn3** | LV end systolic volume \| Instance 2(participant - p24101_i2)_BSA_indexed_corrected | -1.674894045 | 0.2402280827 | -0.09349402454 | 7.73E-10 | 0.2717119751 | -0.09349402454 | 7.73E-10 | 4311 | 1.96E-08 | TRUE | 1.13E-05 | TRUE | 9.57E-06 | TRUE | FALSE | -1.674894045 |
| **cntn3** | RV end diastolic volume \| Instance 2_BSA_indexed_corrected | -3.653470601 | 0.1065204834 | -0.1100532982 | 4.30E-13 | 0.5026529344 | -0.1100532982 | 4.30E-13 | 4311 | 2.01E-11 | TRUE | 6.29E-09 | TRUE | 5.32E-09 | TRUE | FALSE | -3.653470601 |
| **cntn3** | RV end systolic volume \| Instance 2_BSA_indexed_corrected | -1.677765769 | 0.1542506384 | -0.08499366398 | 2.28E-08 | 0.299627853 | -0.08499366398 | 2.28E-08 | 4311 | 4.43E-07 | TRUE | 0.0003340667258 | TRUE | 0.0002825536367 | TRUE | FALSE | -1.677765769 |
| **cntn4** | LV end diastolic volume \| Instance 2(participant - p24100_i2)_BSA_indexed_corrected | 3.68238181 | 0.3709338154 | 0.08061345846 | 1.16E-07 | 0.6936130779 | 0.08061345846 | 1.16E-07 | 4311 | 1.97E-06 | TRUE | 0.001693940184 | TRUE | 0.001432734608 | TRUE | FALSE | 3.68238181 |
| **cntn4** | LV end systolic volume \| Instance 2(participant - p24101_i2)_BSA_indexed_corrected | 1.949355514 | 0.3273602896 | 0.07346069464 | 1.38E-06 | 0.4031556684 | 0.07346069464 | 1.38E-06 | 4311 | 1.90E-05 | TRUE | 0.02013356961 | TRUE | 0.01702897317 | TRUE | FALSE | 1.949355514 |
| **cntn4** | RV end diastolic volume \| Instance 2_BSA_indexed_corrected | 4.092065325 | 0.2954601719 | 0.08321612085 | 4.46E-08 | 0.7465138771 | 0.08321612085 | 4.46E-08 | 4311 | 8.17E-07 | TRUE | 0.0006520095837 | TRUE | 0.0005514697059 | TRUE | FALSE | 4.092065325 |
| **col18a1** | LV end diastolic volume \| Instance 2(participant - p24100_i2)_BSA_indexed_corrected | -4.665720933 | 0.03191689122 | -0.0964594128 | 1.74E-10 | 0.7292906496 | -0.0964594128 | 1.74E-10 | 4360 | 4.97E-09 | TRUE | 2.55E-06 | TRUE | 2.16E-06 | TRUE | FALSE | -4.665720933 |
| **col18a1** | LV end systolic volume \| Instance 2(participant - p24101_i2)_BSA_indexed_corrected | -2.573703506 | 0.1260033472 | -0.09163540179 | 1.35E-09 | 0.4236629818 | -0.09163540179 | 1.35E-09 | 4360 | 3.22E-08 | TRUE | 1.97E-05 | TRUE | 1.67E-05 | TRUE | FALSE | -2.573703506 |
| **col18a1** | RV end diastolic volume \| Instance 2_BSA_indexed_corrected | -5.060562782 | -0.1012167067 | -0.09725640701 | 1.23E-10 | 0.7844644245 | -0.09725640701 | 1.23E-10 | 4360 | 3.61E-09 | TRUE | 1.80E-06 | TRUE | 1.52E-06 | TRUE | FALSE | -5.060562782 |
| **col18a1** | RV end systolic volume \| Instance 2_BSA_indexed_corrected | -2.473923946 | 0.04816144054 | -0.07993917617 | 1.25E-07 | 0.4672948347 | -0.07993917617 | 1.25E-07 | 4360 | 2.11E-06 | TRUE | 0.001835511313 | TRUE | 0.001552475469 | TRUE | FALSE | -2.473923946 |
| **col4a1** | LV end diastolic volume \| Instance 2(participant - p24100_i2)_BSA_indexed_corrected | 3.957849867 | 0.3087431968 | 0.1402863793 | 1.14E-20 | 0.4224136494 | 0.1402863793 | 1.14E-20 | 4375 | 1.75E-18 | TRUE | 1.66E-16 | TRUE | 1.41E-16 | TRUE | FALSE | 3.957849867 |
| **col4a1** | LV end systolic volume \| Instance 2(participant - p24101_i2)_BSA_indexed_corrected | 1.778063698 | 0.2920226388 | 0.1085706696 | 6.00E-13 | 0.2461898886 | 0.1085706696 | 6.00E-13 | 4375 | 2.70E-11 | TRUE | 8.78E-09 | TRUE | 7.42E-09 | TRUE | FALSE | 1.778063698 |
| **col4a1** | RV end diastolic volume \| Instance 2_BSA_indexed_corrected | 4.160648692 | 0.1969021047 | 0.1370785752 | 8.44E-20 | 0.4546556487 | 0.1370785752 | 8.44E-20 | 4375 | 1.22E-17 | TRUE | 1.24E-15 | TRUE | 1.04E-15 | TRUE | FALSE | 4.160648692 |
| **col4a1** | RV end systolic volume \| Instance 2_BSA_indexed_corrected | 1.839361163 | 0.1965465908 | 0.1019031588 | 1.42E-11 | 0.2715331923 | 0.1019031588 | 1.42E-11 | 4375 | 5.00E-10 | TRUE | 2.08E-07 | TRUE | 1.76E-07 | TRUE | FALSE | 1.839361163 |
| **col6a3** | LV cardiac output \| Instance 2_BSA_indexed_corrected | -0.1107538754 | -0.02890942621 | -0.0710835449 | 2.52E-06 | 0.02350174538 | -0.0710835449 | 2.52E-06 | 4375 | 3.33E-05 | TRUE | 0.03689649427 | TRUE | 0.03120705485 | TRUE | FALSE | -0.1107538754 |
| **col6a3** | LV end diastolic volume \| Instance 2(participant - p24100_i2)_BSA_indexed_corrected | -3.937252308 | 0.02729262329 | -0.1127824175 | 7.34E-14 | 0.5245444611 | -0.1127824175 | 7.34E-14 | 4375 | 3.86E-12 | TRUE | 1.07E-09 | TRUE | 9.09E-10 | TRUE | FALSE | -3.937252308 |
| **col6a3** | LV end systolic volume \| Instance 2(participant - p24101_i2)_BSA_indexed_corrected | -1.808911834 | 0.1622634551 | -0.08926363414 | 3.33E-09 | 0.3052220738 | -0.08926363414 | 3.33E-09 | 4375 | 7.40E-08 | TRUE | 4.87E-05 | TRUE | 4.12E-05 | TRUE | FALSE | -1.808911834 |
| **col6a3** | RV end diastolic volume \| Instance 2_BSA_indexed_corrected | -4.284161348 | -0.1109792115 | -0.1140686538 | 3.81E-14 | 0.5642424746 | -0.1140686538 | 3.81E-14 | 4375 | 2.10E-12 | TRUE | 5.57E-10 | TRUE | 4.71E-10 | TRUE | FALSE | -4.284161348 |
| **col6a3** | RV end systolic volume \| Instance 2_BSA_indexed_corrected | -1.933057982 | 0.05720266274 | -0.08654808803 | 9.82E-09 | 0.336484439 | -0.08654808803 | 9.82E-09 | 4375 | 2.02E-07 | TRUE | 0.000143722309 | TRUE | 0.000121560329 | TRUE | FALSE | -1.933057982 |
| **colec12** | LV end diastolic volume \| Instance 2(participant - p24100_i2)_BSA_indexed_corrected | -5.800814294 | -0.1068758121 | -0.1343372465 | 9.02E-19 | 0.6526867204 | -0.1343372465 | 9.02E-19 | 4300 | 1.01E-16 | TRUE | 1.32E-14 | TRUE | 1.12E-14 | TRUE | FALSE | -5.800814294 |
| **colec12** | LV end systolic volume \| Instance 2(participant - p24101_i2)_BSA_indexed_corrected | -2.570571073 | 0.09701898288 | -0.102732379 | 1.45E-11 | 0.3796515057 | -0.102732379 | 1.45E-11 | 4300 | 5.10E-10 | TRUE | 2.13E-07 | TRUE | 1.80E-07 | TRUE | FALSE | -2.570571073 |

| | | | | | | | | | | | | | | | | | |
|---|---|---|---|---|---|---|---|---|---|---|---|---|---|---|---|---|---|
| **colec12** | RV end diastolic volume \| Instance 2_BSA_indexed_corrected | -7.063988951 | -0.3204832548 | -0.1522261762 | 1.04E-23 | 0.6995783124 | -0.1522261762 | 1.04E-23 | 4300 | 2.62E-21 | TRUE | 1.52E-19 | TRUE | 1.29E-19 | TRUE | FALSE | -7.063988951 |
| **colec12** | RV end systolic volume \| Instance 2_BSA_indexed_corrected | -3.358971239 | -0.07046299062 | -0.1218479086 | 1.08E-15 | 0.4173561923 | -0.1218479086 | 1.08E-15 | 4300 | 8.00E-14 | TRUE | 1.58E-11 | TRUE | 1.33E-11 | TRUE | FALSE | -3.358971239 |
| **cpm** | LV end diastolic volume \| Instance 2(participant - p24100_i2)_BSA_indexed_corrected | -4.942799093 | 0.016137633 | -0.1692599493 | 2.67E-29 | 0.4366283924 | -0.1692599493 | 2.67E-29 | 4347 | 1.35E-26 | TRUE | 3.90E-25 | TRUE | 3.30E-25 | TRUE | FALSE | -4.942799093 |
| **cpm** | LV end systolic volume \| Instance 2(participant - p24101_i2)_BSA_indexed_corrected | -2.218798751 | 0.1562310748 | -0.1310084189 | 4.23E-18 | 0.2547208648 | -0.1310084189 | 4.23E-18 | 4347 | 4.23E-16 | TRUE | 6.18E-14 | TRUE | 5.23E-14 | TRUE | FALSE | -2.218798751 |
| **cpm** | RV end diastolic volume \| Instance 2_BSA_indexed_corrected | -5.392670539 | -0.1329034218 | -0.1715057256 | 4.75E-30 | 0.4699453299 | -0.1715057256 | 4.75E-30 | 4347 | 2.78E-27 | TRUE | 6.95E-26 | TRUE | 5.88E-26 | TRUE | FALSE | -5.392670539 |
| **cpm** | RV end systolic volume \| Instance 2_BSA_indexed_corrected | -2.427892312 | 0.0488187128 | -0.1299471199 | 7.86E-18 | 0.2810410475 | -0.1299471199 | 7.86E-18 | 4347 | 7.59E-16 | TRUE | 1.15E-13 | TRUE | 9.72E-14 | TRUE | FALSE | -2.427892312 |
| **creg1** | LV end diastolic volume \| Instance 2(participant - p24100_i2)_BSA_indexed_corrected | -3.025047326 | 0.2950833158 | -0.1010141337 | 3.73E-11 | 0.4561550458 | -0.1010141337 | 3.73E-11 | 4268 | 1.23E-09 | TRUE | 5.46E-07 | TRUE | 4.62E-07 | TRUE | FALSE | -3.025047326 |
| **creg1** | LV end systolic volume \| Instance 2(participant - p24101_i2)_BSA_indexed_corrected | -1.412153668 | 0.2930951968 | -0.0813046428 | 1.04E-07 | 0.265042782 | -0.0813046428 | 1.04E-07 | 4268 | 1.80E-06 | TRUE | 0.001527504249 | TRUE | 0.001291963094 | TRUE | FALSE | -1.412153668 |
| **creg1** | RV end diastolic volume \| Instance 2_BSA_indexed_corrected | -3.067477482 | 0.1912510185 | -0.09559859984 | 3.90E-10 | 0.4890190454 | -0.09559859984 | 3.90E-10 | 4268 | 1.05E-08 | TRUE | 5.71E-06 | TRUE | 4.83E-06 | TRUE | FALSE | -3.067477482 |
| **crhbp** | LV end diastolic volume \| Instance 2(participant - p24100_i2)_BSA_indexed_corrected | -4.157049568 | 0.1162915568 | -0.1073809799 | 1.67E-12 | 0.5870920954 | -0.1073809799 | 1.67E-12 | 4300 | 6.85E-11 | TRUE | 2.44E-08 | TRUE | 2.06E-08 | TRUE | FALSE | -4.157049568 |
| **crhbp** | LV end systolic volume \| Instance 2(participant - p24101_i2)_BSA_indexed_corrected | -1.923539041 | 0.1912075098 | -0.08574591006 | 1.79E-08 | 0.3409195783 | -0.08574591006 | 1.79E-08 | 4300 | 3.53E-07 | TRUE | 0.0002614024868 | TRUE | 0.0002210942234 | TRUE | FALSE | -1.923539041 |
| **crhbp** | RV end diastolic volume \| Instance 2_BSA_indexed_corrected | -5.00123148 | -0.04518941628 | -0.1202128692 | 2.59E-15 | 0.629987408 | -0.1202128692 | 2.59E-15 | 4300 | 1.75E-13 | TRUE | 3.79E-11 | TRUE | 3.20E-11 | TRUE | FALSE | -5.00123148 |
| **crhbp** | RV end systolic volume \| Instance 2_BSA_indexed_corrected | -2.387364318 | 0.05990627728 | -0.09659734111 | 2.19E-10 | 0.375218806 | -0.09659734111 | 2.19E-10 | 4300 | 6.18E-09 | TRUE | 3.21E-06 | TRUE | 2.71E-06 | TRUE | FALSE | -2.387364318 |
| **crip2** | LV end diastolic volume \| Instance 2(participant - p24100_i2)_BSA_indexed_corrected | -3.018532034 | 0.1091711549 | -0.1021264444 | 1.28E-11 | 0.4446221973 | -0.1021264444 | 1.28E-11 | 4375 | 4.53E-10 | TRUE | 1.87E-07 | TRUE | 1.58E-07 | TRUE | FALSE | -3.018532034 |
| **crip2** | LV end systolic volume \| Instance 2(participant - p24101_i2)_BSA_indexed_corrected | -1.311073632 | 0.2060002501 | -0.07641498224 | 4.19E-07 | 0.2586944637 | -0.07641498224 | 4.19E-07 | 4375 | 6.43E-06 | TRUE | 0.006123064558 | TRUE | 0.005178888003 | TRUE | FALSE | -1.311073632 |
| **crip2** | RV end diastolic volume \| Instance 2_BSA_indexed_corrected | -2.886339755 | 0.01027733253 | -0.09076997542 | 1.80E-09 | 0.4788714731 | -0.09076997542 | 1.80E-09 | 4375 | 4.18E-08 | TRUE | 2.64E-05 | TRUE | 2.23E-05 | TRUE | FALSE | -2.886339755 |
| **crip2** | RV end systolic volume \| Instance 2_BSA_indexed_corrected | -1.513002594 | 0.09489758261 | -0.08001041797 | 1.16E-07 | 0.2850419772 | -0.08001041797 | 1.16E-07 | 4375 | 1.98E-06 | TRUE | 0.001701852459 | TRUE | 0.001439426809 | TRUE | FALSE | -1.513002594 |
| **crlf1** | LV cardiac output \| Instance 2_BSA_indexed_corrected | -0.1609781183 | -0.01976358173 | -0.08690827952 | 1.05E-08 | 0.02807485175 | -0.08690827952 | 1.05E-08 | 4322 | 2.15E-07 | TRUE | 0.0001533921729 | TRUE | 0.0001297390998 | TRUE | FALSE | -0.1609781183 |
| **crlf1** | LV end diastolic volume \| Instance 2(participant - p24100_i2)_BSA_indexed_corrected | -3.986473404 | 0.3264661465 | -0.09588269357 | 2.68E-10 | 0.6296528845 | -0.09588269357 | 2.68E-10 | 4322 | 7.45E-09 | TRUE | 3.92E-06 | TRUE | 3.32E-06 | TRUE | FALSE | -3.986473404 |
| **crlf1** | RV end diastolic volume \| Instance 2_BSA_indexed_corrected | -3.63691692 | 0.2169218702 | -0.08135506065 | 8.52E-08 | 0.6778989102 | -0.08135506065 | 8.52E-08 | 4322 | 1.49E-06 | TRUE | 0.001246370208 | TRUE | 0.001054179922 | TRUE | FALSE | -3.63691692 |
| **crlf1** | RV end systolic volume \| Instance 2_BSA_indexed_corrected | -2.006839769 | 0.1986949848 | -0.0754529549 | 6.83E-07 | 0.4035104524 | -0.0754529549 | 6.83E-07 | 4322 | 1.01E-05 | TRUE | 0.009995837471 | TRUE | 0.008454479333 | TRUE | FALSE | -2.006839769 |
| **csf1** | LV end diastolic volume \| Instance 2(participant - p24100_i2)_BSA_indexed_corrected | -4.138094031 | 0.1517616478 | -0.09336509163 | 7.76E-10 | 0.6713867625 | -0.09336509163 | 7.76E-10 | 4322 | 1.97E-08 | TRUE | 1.14E-05 | TRUE | 9.61E-06 | TRUE | FALSE | -4.138094031 |
| **csf1** | RV end diastolic volume \| Instance 2_BSA_indexed_corrected | -4.850801471 | 0.005850145112 | -0.101788218 | 1.98E-11 | 0.7212943853 | -0.101788218 | 1.98E-11 | 4322 | 6.86E-10 | TRUE | 2.90E-07 | TRUE | 2.45E-07 | TRUE | FALSE | -4.850801471 |
| **csf1** | RV end systolic volume \| Instance 2_BSA_indexed_corrected | -2.190908241 | 0.1055690748 | -0.07727171665 | 3.65E-07 | 0.4300919365 | -0.07727171665 | 3.65E-07 | 4322 | 5.67E-06 | TRUE | 0.005346519157 | TRUE | 0.004522085903 | TRUE | FALSE | -2.190908241 |
| **cst3** | LV cardiac output \| Instance 2_BSA_indexed_corrected | -0.136133495 | -0.03103895655 | -0.07667272581 | 4.57E-07 | 0.0269496655 | -0.07667272581 | 4.57E-07 | 4317 | 6.97E-06 | TRUE | 0.006681596868 | TRUE | 0.005651294631 | TRUE | FALSE | -0.136133495 |
| **cst3** | LV end diastolic volume \| Instance 2(participant - p24100_i2)_BSA_indexed_corrected | -5.390293056 | -0.06981863627 | -0.135298287 | 4.33E-19 | 0.6009216212 | -0.135298287 | 4.33E-19 | 4317 | 5.15E-17 | TRUE | 6.34E-15 | TRUE | 5.36E-15 | TRUE | FALSE | -5.390293056 |
| **cst3** | LV end systolic volume \| Instance 2(participant - p24101_i2)_BSA_indexed_corrected | -2.227190546 | 0.1419847699 | -0.0962556523 | 2.34E-10 | 0.3506060878 | -0.0962556523 | 2.34E-10 | 4317 | 6.54E-09 | TRUE | 3.42E-06 | TRUE | 2.89E-06 | TRUE | FALSE | -2.227190546 |
| **cst3** | RV end diastolic volume \| Instance 2_BSA_indexed_corrected | -5.824054414 | -0.2152868898 | -0.1360772625 | 2.71E-19 | 0.6454919732 | -0.1360772625 | 2.71E-19 | 4317 | 3.33E-17 | TRUE | 3.96E-15 | TRUE | 3.35E-15 | TRUE | FALSE | -5.824054414 |
| **cst3** | RV end systolic volume \| Instance 2_BSA_indexed_corrected | -2.426748426 | 0.04327035486 | -0.09541468121 | 3.35E-10 | 0.3854190699 | -0.09541468121 | 3.35E-10 | 4317 | 9.18E-09 | TRUE | 4.90E-06 | TRUE | 4.15E-06 | TRUE | FALSE | -2.426748426 |
| **cstb** | LV end diastolic volume \| Instance 2(participant - p24100_i2)_BSA_indexed_corrected | -3.022027986 | 0.04810227537 | -0.1155752529 | 2.65E-14 | 0.3955251758 | -0.1155752529 | 2.65E-14 | 4314 | 1.51E-12 | TRUE | 3.87E-10 | TRUE | 3.28E-10 | TRUE | FALSE | -3.022027986 |
| **cstb** | LV end systolic volume \| Instance 2(participant - p24101_i2)_BSA_indexed_corrected | -1.397600161 | 0.1753774962 | -0.09198735359 | 1.42E-09 | 0.2303934852 | -0.09198735359 | 1.42E-09 | 4314 | 3.38E-08 | TRUE | 2.08E-05 | TRUE | 1.76E-05 | TRUE | FALSE | -1.397600161 |
| **cstb** | RV end diastolic volume \| Instance 2_BSA_indexed_corrected | -3.358883663 | -0.09311656476 | -0.1192303355 | 3.94E-15 | 0.4259510866 | -0.1192303355 | 3.94E-15 | 4314 | 2.58E-13 | TRUE | 5.76E-11 | TRUE | 4.87E-11 | TRUE | FALSE | -3.358883663 |
| **cstb** | RV end systolic volume \| Instance 2_BSA_indexed_corrected | -1.60843684 | 0.05660209733 | -0.09612045171 | 2.51E-10 | 0.2536490576 | -0.09612045171 | 2.51E-10 | 4314 | 7.01E-09 | TRUE | 3.68E-06 | TRUE | 3.11E-06 | TRUE | FALSE | -1.60843684 |
| **ctsb** | LV end diastolic volume \| Instance 2(participant - p24100_i2)_BSA_indexed_corrected | 2.18094063 | 0.2659036021 | 0.1041887322 | 5.77E-12 | 0.3158697943 | 0.1041887322 | 5.77E-12 | 4346 | 2.20E-10 | TRUE | 8.43E-08 | TRUE | 7.13E-08 | TRUE | FALSE | 2.18094063 |
| **ctsb** | LV end systolic volume \| Instance 2(participant - p24101_i2)_BSA_indexed_corrected | 1.190718821 | 0.2616037776 | 0.0979685848 | 9.66E-11 | 0.1835199423 | 0.0979685848 | 9.66E-11 | 4346 | 2.88E-09 | TRUE | 1.41E-06 | TRUE | 1.20E-06 | TRUE | FALSE | 1.190718821 |
| **ctsb** | RV end diastolic volume \| Instance 2_BSA_indexed_corrected | 2.276015361 | 0.1585670057 | 0.1010448747 | 2.45E-11 | 0.3400067248 | 0.1010448747 | 2.45E-11 | 4346 | 8.23E-10 | TRUE | 3.58E-07 | TRUE | 3.03E-07 | TRUE | FALSE | 2.276015361 |
| **ctsb** | RV end systolic volume \| Instance 2_BSA_indexed_corrected | 1.105377695 | 0.1797299745 | 0.08252563005 | 5.09E-08 | 0.2025316982 | 0.08252563005 | 5.09E-08 | 4346 | 9.23E-07 | TRUE | 0.0007445482391 | TRUE | 0.0006297389007 | TRUE | FALSE | 1.105377695 |
| **ctsd** | LV end diastolic volume \| Instance 2(participant - p24100_i2)_BSA_indexed_corrected | -3.989212443 | 0.2132704126 | -0.1368103975 | 1.72E-18 | 0.4524820679 | -0.1368103975 | 1.72E-18 | 4077 | 1.87E-16 | TRUE | 2.52E-14 | TRUE | 2.13E-14 | TRUE | FALSE | -3.989212443 |
| **ctsd** | LV end systolic volume \| Instance 2(participant - p24101_i2)_BSA_indexed_corrected | -1.7092051 | 0.225665745 | -0.1014536432 | 8.43E-11 | 0.2625524541 | -0.1014536432 | 8.43E-11 | 4077 | 2.54E-09 | TRUE | 1.23E-06 | TRUE | 1.04E-06 | TRUE | FALSE | -1.7092051 |
| **ctsd** | RV end diastolic volume \| Instance 2_BSA_indexed_corrected | -4.737532301 | 0.06878722228 | -0.1514883176 | 2.34E-22 | 0.4842480897 | -0.1514883176 | 2.34E-22 | 4077 | 4.44E-20 | TRUE | 3.42E-18 | TRUE | 2.89E-18 | TRUE | FALSE | -4.737532301 |
| **ctsd** | RV end systolic volume \| Instance 2_BSA_indexed_corrected | -2.056316023 | 0.1371198912 | -0.1106386779 | 1.40E-12 | 0.2893640616 | -0.1106386779 | 1.40E-12 | 4077 | 5.85E-11 | TRUE | 2.05E-08 | TRUE | 1.73E-08 | TRUE | FALSE | -2.056316023 |
| **ctso** | LV end diastolic volume \| Instance 2(participant - p24100_i2)_BSA_indexed_corrected | -4.305537748 | 0.1562418403 | -0.1205118379 | 1.40E-15 | 0.5371894571 | -0.1205118379 | 1.40E-15 | 4361 | 9.92E-14 | TRUE | 2.05E-11 | TRUE | 1.74E-11 | TRUE | FALSE | -4.305537748 |
| **ctso** | LV end systolic volume \| Instance 2(participant - p24101_i2)_BSA_indexed_corrected | -1.967435323 | 0.2205261516 | -0.09479808896 | 3.55E-10 | 0.3129299955 | -0.09479808896 | 3.55E-10 | 4361 | 9.64E-09 | TRUE | 5.19E-06 | TRUE | 4.39E-06 | TRUE | FALSE | -1.967435323 |
| **ctso** | RV end diastolic volume \| Instance 2_BSA_indexed_corrected | -3.986630666 | 0.06568840633 | -0.1036977195 | 6.67E-12 | 0.5791561682 | -0.1036977195 | 6.67E-12 | 4361 | 2.51E-10 | TRUE | 9.76E-08 | TRUE | 8.26E-08 | TRUE | FALSE | -3.986630666 |
| **ctso** | RV end systolic volume \| Instance 2_BSA_indexed_corrected | -1.857258789 | 0.1320724423 | -0.08122107458 | 7.83E-08 | 0.345201775 | -0.08122107458 | 7.83E-08 | 4361 | 1.38E-06 | TRUE | 0.001145519909 | TRUE | 0.0009688807387 | TRUE | FALSE | -1.857258789 |
| **ctss** | LV end diastolic volume \| Instance 2(participant - p24100_i2)_BSA_indexed_corrected | -5.417174509 | 0.08988504584 | -0.1064926528 | 4.34E-10 | 0.8654013746 | -0.1064926528 | 4.34E-10 | 3418 | 1.16E-08 | TRUE | 6.34E-06 | TRUE | 5.37E-06 | TRUE | FALSE | -5.417174509 |
| **ctss** | RV end diastolic volume \| Instance 2_BSA_indexed_corrected | -5.702749702 | -0.04915325952 | -0.1038889592 | 1.14E-09 | 0.9341133806 | -0.1038889592 | 1.14E-09 | 3418 | 2.79E-08 | TRUE | 1.67E-05 | TRUE | 1.41E-05 | TRUE | FALSE | -5.702749702 |
| **ctsz** | LV end diastolic volume \| Instance 2(participant - p24100_i2)_BSA_indexed_corrected | -3.492370435 | 0.1279733442 | -0.0995856344 | 5.44E-11 | 0.5311517626 | -0.0995856344 | 5.44E-11 | 4318 | 1.72E-09 | TRUE | 7.96E-07 | TRUE | 6.73E-07 | TRUE | FALSE | -3.492370435 |
| **ctsz** | RV end diastolic volume \| Instance 2_BSA_indexed_corrected | -4.074016025 | -0.016257728 | -0.1080528209 | 1.09E-12 | 0.5705526221 | -0.1080528209 | 1.09E-12 | 4318 | 4.61E-11 | TRUE | 1.59E-08 | TRUE | 1.34E-08 | TRUE | FALSE | -4.074016025 |
| **cxcl10** | LV end diastolic volume \| Instance 2(participant - p24100_i2)_BSA_indexed_corrected | -1.629106863 | 0.2507929808 | -0.09860980739 | 8.21E-11 | 0.2501299748 | -0.09860980739 | 8.21E-11 | 4322 | 2.49E-09 | TRUE | 1.20E-06 | TRUE | 1.02E-06 | TRUE | FALSE | -1.629106863 |
| **cxcl10** | RV end diastolic volume \| Instance 2_BSA_indexed_corrected | -1.72743665 | 0.133105539 | -0.09724616978 | 1.49E-10 | 0.2689827677 | -0.09724616978 | 1.49E-10 | 4322 | 4.31E-09 | TRUE | 2.18E-06 | TRUE | 1.84E-06 | TRUE | FALSE | -1.72743665 |
| **cxcl13** | LV end diastolic volume \| Instance 2(participant - p24100_i2)_BSA_indexed_corrected | -1.583225595 | 0.3214915407 | -0.08422152859 | 2.53E-08 | 0.2836487457 | -0.08422152859 | 2.53E-08 | 4363 | 4.87E-07 | TRUE | 0.0003697178696 | TRUE | 0.0003127073741 | TRUE | FALSE | -1.583225595 |
| **cxcl13** | RV end diastolic volume \| Instance 2_BSA_indexed_corrected | -1.830614427 | 0.2018826973 | -0.09044759517 | 2.17E-09 | 0.3052270897 | -0.09044759517 | 2.17E-09 | 4363 | 4.93E-08 | TRUE | 3.17E-05 | TRUE | 2.68E-05 | TRUE | FALSE | -1.830614427 |
| **cxcl16** | LV end diastolic volume \| Instance 2(participant - p24100_i2)_BSA_indexed_corrected | -4.141646848 | 0.1392240665 | -0.08727343118 | 7.38E-09 | 0.7148929418 | -0.08727343118 | 7.38E-09 | 4375 | 1.55E-07 | TRUE | 0.0001080081849 | TRUE | 9.14E-05 | TRUE | FALSE | -4.141646848 |
| **cxcl16** | RV end diastolic volume \| Instance 2_BSA_indexed_corrected | -5.53637416 | -0.04234410321 | -0.1084393144 | 6.40E-13 | 0.7675034676 | -0.1084393144 | 6.40E-13 | 4375 | 2.87E-11 | TRUE | 9.36E-09 | TRUE | 7.92E-09 | TRUE | FALSE | -5.53637416 |
| **cxcl16** | RV end systolic volume \| Instance 2_BSA_indexed_corrected | -2.325468867 | 0.09708113932 | -0.07659209694 | 3.93E-07 | 0.4577825336 | -0.07659209694 | 3.93E-07 | 4375 | 6.08E-06 | TRUE | 0.005756319165 | TRUE | 0.004868694749 | TRUE | FALSE | -2.325468867 |
| **cxcl17** | LV end diastolic volume \| Instance 2(participant - p24100_i2)_BSA_indexed_corrected | -2.10743625 | 0.1129395737 | -0.09214698895 | 1.08E-09 | 0.3449277843 | -0.09214698895 | 1.08E-09 | 4361 | 2.66E-08 | TRUE | 1.59E-05 | TRUE | 1.34E-05 | TRUE | FALSE | -2.10743625 |
| **cxcl17** | RV end diastolic volume \| Instance 2_BSA_indexed_corrected | -2.685277209 | -0.06434251689 | -0.1091130567 | 5.01E-13 | 0.3705255622 | -0.1091130567 | 5.01E-13 | 4361 | 2.31E-11 | TRUE | 7.33E-09 | TRUE | 6.20E-09 | TRUE | FALSE | -2.685277209 |
| **cxcl17** | RV end systolic volume \| Instance 2_BSA_indexed_corrected | -1.142241203 | 0.08482129431 | -0.07803303972 | 2.48E-07 | 0.2210342516 | -0.07803303972 | 2.48E-07 | 4361 | 3.97E-06 | TRUE | 0.003622019859 | TRUE | 0.003063504397 | TRUE | FALSE | -1.142241203 |
| **dcxr** | LV end diastolic volume \| Instance 2(participant - p24100_i2)_BSA_indexed_corrected | -2.231474851 | 0.3599118439 | -0.1186721423 | 4.38E-15 | 0.2834775045 | -0.1186721423 | 4.38E-15 | 4340 | 2.84E-13 | TRUE | 6.41E-11 | TRUE | 5.42E-11 | TRUE | FALSE | -2.231474851 |
| **dcxr** | LV end systolic volume \| Instance 2(participant - p24101_i2)_BSA_indexed_corrected | -0.9433459371 | 0.3134612207 | -0.08644117098 | 1.17E-08 | 0.1650733387 | -0.08644117098 | 1.17E-08 | 4340 | 2.39E-07 | TRUE | 0.0001715464236 | TRUE | 0.0001450939651 | TRUE | FALSE | -0.9433459371 |
| **dcxr** | RV end diastolic volume \| Instance 2_BSA_indexed_corrected | -2.26237446 | 0.2554208858 | -0.1118655665 | 1.46E-13 | 0.3051327223 | -0.1118655665 | 1.46E-13 | 4340 | 7.20E-12 | TRUE | 2.14E-09 | TRUE | 1.81E-09 | TRUE | FALSE | -2.26237446 |
| **dcxr** | RV end systolic volume \| Instance 2_BSA_indexed_corrected | -1.028986722 | 0.2157815884 | -0.08552637995 | 1.67E-08 | 0.181999707 | -0.08552637995 | 1.67E-08 | 4340 | 3.32E-07 | TRUE | 0.000244366852 | TRUE | 0.0002066854834 | TRUE | FALSE | -1.028986722 |
| **ddah1** | LV end diastolic volume \| Instance 2(participant - p24100_i2)_BSA_indexed_corrected | -2.453430446 | 0.2446268331 | -0.09576117884 | 2.20E-10 | 0.3856506406 | -0.09576117884 | 2.20E-10 | 4375 | 6.19E-09 | TRUE | 3.22E-06 | TRUE | 2.72E-06 | TRUE | FALSE | -2.453430446 |
| **ddah1** | RV end diastolic volume \| Instance 2_BSA_indexed_corrected | -2.993620182 | 0.1111895844 | -0.10860874 | 5.89E-13 | 0.4143482593 | -0.10860874 | 5.89E-13 | 4375 | 2.66E-11 | TRUE | 8.61E-09 | TRUE | 7.29E-09 | TRUE | FALSE | -2.993620182 |
| **ddah1** | RV end systolic volume \| Instance 2_BSA_indexed_corrected | -1.44940774 | 0.1530891075 | -0.08842412142 | 4.67E-09 | 0.24690255 | -0.08842412142 | 4.67E-09 | 4375 | 1.01E-07 | TRUE | 6.83E-05 | TRUE | 5.78E-05 | TRUE | FALSE | -1.44940774 |
| **ddc** | LV end diastolic volume \| Instance 2(participant - p24100_i2)_BSA_indexed_corrected | 1.815145536 | 0.3014131724 | 0.07669505218 | 3.81E-07 | 0.3568806228 | 0.07669505218 | 3.81E-07 | 4374 | 5.90E-06 | TRUE | 0.005569969249 | TRUE | 0.004711079991 | TRUE | FALSE | 1.815145536 |
| **ddc** | LV end systolic volume \| Instance 2(participant - p24101_i2)_BSA_indexed_corrected | 0.9711777311 | 0.2831227727 | 0.070682221 | 2.88E-06 | 0.2072816561 | 0.070682221 | 2.88E-06 | 4374 | 3.77E-05 | TRUE | 0.04213963997 | TRUE | 0.03564170748 | TRUE | FALSE | 0.9711777311 |

| | | | | | | | | | | | | | | | | | |
|---|---|---|---|---|---|---|---|---|---|---|---|---|---|---|---|---|---|
| ddc | RV end diastolic volume \| Instance 2_BSA_indexed_corrected | 2.086028831 | 0.1832407794 | 0.08192328763 | 5.77E-08 | 0.3838049553 | 0.08192328763 | 5.77E-08 | 4374 | 1.04E-06 | TRUE | 0.0008447012934 | TRUE | 0.000714448354 | TRUE | FALSE | 2.086028831 |
| defb4a_defb4b | LV end diastolic volume \| Instance 2(participant - p24100_i2)_BSA_indexed_corrected | -0.5277651599 | 0.2862373887 | -0.07407023257 | 1.11E-06 | 0.1081963583 | -0.07407023257 | 1.11E-06 | 4315 | 1.57E-05 | TRUE | 0.01625621477 | TRUE | 0.01374950645 | TRUE | FALSE | -0.5277651599 |
| dkk3 | LV cardiac output \| Instance 2_BSA_indexed_corrected | -0.1316117897 | -0.0229435804 | -0.07917990339 | 1.57E-07 | 0.02505673085 | -0.07917990339 | 1.57E-07 | 4375 | 2.61E-06 | TRUE | 0.002298890655 | TRUE | 0.001944401716 | TRUE | FALSE | -0.1316117897 |
| dlk1 | LV end diastolic volume \| Instance 2(participant - p24100_i2)_BSA_indexed_corrected | -1.356580773 | 0.2812152828 | -0.06989924748 | 3.70E-06 | 0.2927654731 | -0.06989924748 | 3.70E-06 | 4375 | 4.76E-05 | TRUE | 0.05408375644 | FALSE | 0.04574404119 | TRUE | FALSE | -1.356580773 |
| dpt | LV end diastolic volume \| Instance 2(participant - p24100_i2)_BSA_indexed_corrected | -4.284474061 | -0.1180821922 | -0.1163024641 | 1.19E-14 | 0.5533012185 | -0.1163024641 | 1.19E-14 | 4375 | 7.16E-13 | TRUE | 1.75E-10 | TRUE | 1.48E-10 | TRUE | FALSE | -4.284474061 |
| dpt | LV end systolic volume \| Instance 2(participant - p24101_i2)_BSA_indexed_corrected | -1.694245268 | 0.1256218593 | -0.07922763471 | 1.54E-07 | 0.3223610184 | -0.07922763471 | 1.54E-07 | 4375 | 2.57E-06 | TRUE | 0.002259684316 | TRUE | 0.001911240995 | TRUE | FALSE | -1.694245268 |
| dpt | RV end diastolic volume \| Instance 2_BSA_indexed_corrected | -4.505338073 | -0.2519398042 | -0.1136766136 | 4.65E-14 | 0.5954457135 | -0.1136766136 | 4.65E-14 | 4375 | 2.55E-12 | TRUE | 6.81E-10 | TRUE | 5.76E-10 | TRUE | FALSE | -4.505338073 |
| dpt | RV end systolic volume \| Instance 2_BSA_indexed_corrected | -1.868127516 | 0.01171233684 | -0.07926151323 | 1.53E-07 | 0.3552923959 | -0.07926151323 | 1.53E-07 | 4375 | 2.54E-06 | TRUE | 0.002232249665 | TRUE | 0.001888036766 | TRUE | FALSE | -1.868127516 |
| dpy30 | LV end diastolic volume \| Instance 2(participant - p24100_i2)_BSA_indexed_corrected | -1.788354154 | 0.1808306885 | -0.09946495119 | 4.48E-11 | 0.2707898207 | -0.09946495119 | 4.48E-11 | 4367 | 1.45E-09 | TRUE | 6.55E-07 | TRUE | 5.54E-07 | TRUE | FALSE | -1.788354154 |
| dpy30 | LV end systolic volume \| Instance 2(participant - p24101_i2)_BSA_indexed_corrected | -0.7872426362 | 0.2362333634 | -0.07541346605 | 6.06E-07 | 0.1575538784 | -0.07541346605 | 6.06E-07 | 4367 | 9.02E-06 | TRUE | 0.008867933596 | TRUE | 0.007500498236 | TRUE | FALSE | -0.7872426362 |
| dpy30 | RV end diastolic volume \| Instance 2_BSA_indexed_corrected | -1.928853999 | 0.06702833356 | -0.09968408287 | 4.06E-11 | 0.2914156345 | -0.09968408287 | 4.06E-11 | 4367 | 1.32E-09 | TRUE | 5.93E-07 | TRUE | 5.02E-07 | TRUE | FALSE | -1.928853999 |
| dpy30 | RV end systolic volume \| Instance 2_BSA_indexed_corrected | -0.93883394 | 0.1332239029 | -0.08157879071 | 6.72E-08 | 0.1736078775 | -0.08157879071 | 6.72E-08 | 4367 | 1.19E-06 | TRUE | 0.0009834137292 | TRUE | 0.0008317713322 | TRUE | FALSE | -0.93883394 |
| dsg2 | LV end diastolic volume \| Instance 2(participant - p24100_i2)_BSA_indexed_corrected | 3.827506723 | 0.2596022556 | 0.09411197453 | 5.97E-10 | 0.6168093877 | 0.09411197453 | 5.97E-10 | 4311 | 1.55E-08 | TRUE | 8.74E-06 | TRUE | 7.39E-06 | TRUE | FALSE | 3.827506723 |
| dsg2 | LV end systolic volume \| Instance 2(participant - p24101_i2)_BSA_indexed_corrected | 1.736250007 | 0.2749008271 | 0.07348969188 | 1.36E-06 | 0.358939831 | 0.07348969188 | 1.36E-06 | 4311 | 1.89E-05 | TRUE | 0.01994127552 | TRUE | 0.01686633083 | TRUE | FALSE | 1.736250007 |
| dsg2 | RV end diastolic volume \| Instance 2_BSA_indexed_corrected | 4.111217254 | 0.1749170891 | 0.09390430925 | 6.51E-10 | 0.664008069 | 0.09390430925 | 6.51E-10 | 4311 | 1.69E-08 | TRUE | 9.53E-06 | TRUE | 8.06E-06 | TRUE | FALSE | 4.111217254 |
| dtx3 | LV end diastolic volume \| Instance 2(participant - p24100_i2)_BSA_indexed_corrected | -3.209181065 | 0.1727687586 | -0.06991003744 | 3.68E-06 | 0.6924701504 | -0.06991003744 | 3.68E-06 | 4375 | 4.75E-05 | TRUE | 0.05389710798 | FALSE | 0.04558617393 | TRUE | FALSE | -3.209181065 |
| efna1 | LV cardiac output \| Instance 2_BSA_indexed_corrected | -0.1438330134 | -0.02602019013 | -0.07207971425 | 2.18E-06 | 0.03033034735 | -0.07207971425 | 2.18E-06 | 4308 | 2.92E-05 | TRUE | 0.0319270314 | TRUE | 0.02700388316 | TRUE | FALSE | -0.1438330134 |
| efna1 | LV end diastolic volume \| Instance 2(participant - p24100_i2)_BSA_indexed_corrected | -3.878189707 | 0.1760722621 | -0.0868787789 | 1.12E-08 | 0.6776931581 | -0.0868787789 | 1.12E-08 | 4308 | 2.29E-07 | TRUE | 0.0001638660894 | TRUE | 0.0001385979385 | TRUE | FALSE | -3.878189707 |
| efna1 | RV end diastolic volume \| Instance 2_BSA_indexed_corrected | -4.896663057 | 0.05262955609 | -0.1019214057 | 2.01E-11 | 0.7283334995 | -0.1019214057 | 2.01E-11 | 4308 | 6.94E-10 | TRUE | 2.94E-07 | TRUE | 2.49E-07 | TRUE | FALSE | -4.896663057 |
| efna4 | LV end diastolic volume \| Instance 2(participant - p24100_i2)_BSA_indexed_corrected | -4.552903954 | 0.07146215043 | -0.1099286322 | 3.19E-13 | 0.6229397029 | -0.1099286322 | 3.19E-13 | 4369 | 1.51E-11 | TRUE | 4.67E-09 | TRUE | 3.95E-09 | TRUE | FALSE | -4.552903954 |
| efna4 | LV end systolic volume \| Instance 2(participant - p24101_i2)_BSA_indexed_corrected | -2.013168104 | 0.1921917725 | -0.08374504163 | 2.96E-08 | 0.362494209 | -0.08374504163 | 2.96E-08 | 4369 | 5.62E-07 | TRUE | 0.0004335835723 | TRUE | 0.0003667249855 | TRUE | FALSE | -2.013168104 |
| efna4 | RV end diastolic volume \| Instance 2_BSA_indexed_corrected | -5.378247822 | -0.0929331565 | -0.1206975552 | 1.20E-15 | 0.669366685 | -0.1206975552 | 1.20E-15 | 4369 | 8.70E-14 | TRUE | 1.75E-11 | TRUE | 1.48E-11 | TRUE | FALSE | -5.378247822 |
| efna4 | RV end systolic volume \| Instance 2_BSA_indexed_corrected | -2.543398955 | 0.05685839883 | -0.09600635335 | 2.03E-10 | 0.3990360598 | -0.09600635335 | 2.03E-10 | 4369 | 5.78E-09 | TRUE | 2.98E-06 | TRUE | 2.52E-06 | TRUE | FALSE | -2.543398955 |
| egln1 | LV end diastolic volume \| Instance 2(participant - p24100_i2)_BSA_indexed_corrected | -2.853373714 | 0.1690056005 | -0.1326413304 | 1.84E-18 | 0.3240271516 | -0.1326413304 | 1.84E-18 | 4332 | 1.98E-16 | TRUE | 2.70E-14 | TRUE | 2.28E-14 | TRUE | FALSE | -2.853373714 |
| egln1 | LV end systolic volume \| Instance 2(participant - p24101_i2)_BSA_indexed_corrected | -1.384607243 | 0.2254078905 | -0.1109577433 | 2.42E-13 | 0.1884668654 | -0.1109577433 | 2.42E-13 | 4332 | 1.15E-11 | TRUE | 3.53E-09 | TRUE | 2.99E-09 | TRUE | FALSE | -1.384607243 |
| egln1 | LV longitudinal strain global \| Instance 2_corrected | -0.3262963174 | -0.021656414 | -0.07036533484 | 3.42E-06 | 0.07017472121 | -0.07036533484 | 3.42E-06 | 4347 | 4.43E-05 | TRUE | 0.05005911468 | FALSE | 0.0423399992 | TRUE | FALSE | -0.3262963174 |
| egln1 | RV end diastolic volume \| Instance 2_BSA_indexed_corrected | -2.527954004 | 0.08745527671 | -0.1092486614 | 5.59E-13 | 0.3495439533 | -0.1092486614 | 5.59E-13 | 4332 | 2.54E-11 | TRUE | 8.18E-09 | TRUE | 6.92E-09 | TRUE | FALSE | -2.527954004 |
| egln1 | RV end systolic volume \| Instance 2_BSA_indexed_corrected | -1.268676906 | 0.1411753557 | -0.09209934515 | 1.26E-09 | 0.2084494637 | -0.09209934515 | 1.26E-09 | 4332 | 3.03E-08 | TRUE | 1.84E-05 | TRUE | 1.55E-05 | TRUE | FALSE | -1.268676906 |
| eloa | LV end diastolic volume \| Instance 2(participant - p24100_i2)_BSA_indexed_corrected | -2.719926137 | 0.2535878769 | -0.1287385845 | 1.25E-17 | 0.3168323863 | -0.1287385845 | 1.25E-17 | 4375 | 1.18E-15 | TRUE | 1.83E-13 | TRUE | 1.54E-13 | TRUE | FALSE | -2.719926137 |
| eloa | LV end systolic volume \| Instance 2(participant - p24101_i2)_BSA_indexed_corrected | -1.239862947 | 0.2665884502 | -0.1010960789 | 2.05E-11 | 0.1845095615 | -0.1010960789 | 2.05E-11 | 4375 | 7.07E-10 | TRUE | 3.00E-07 | TRUE | 2.54E-07 | TRUE | FALSE | -1.239862947 |
| eloa | RV end diastolic volume \| Instance 2_BSA_indexed_corrected | -2.568791353 | 0.1495400085 | -0.1130141424 | 6.53E-14 | 0.3415190919 | -0.1130141424 | 6.53E-14 | 4375 | 3.50E-12 | TRUE | 9.55E-10 | TRUE | 8.07E-10 | TRUE | FALSE | -2.568791353 |
| eloa | RV end systolic volume \| Instance 2_BSA_indexed_corrected | -1.251445258 | 0.1713746732 | -0.09258229028 | 8.51E-10 | 0.2035283454 | -0.09258229028 | 8.51E-10 | 4375 | 2.13E-08 | TRUE | 1.24E-05 | TRUE | 1.05E-05 | TRUE | FALSE | -1.251445258 |
| eng | LV longitudinal strain global \| Instance 2_corrected | 1.015583018 | 0.003399681199 | 0.07422388903 | 8.51E-07 | 0.2059865212 | 0.07422388903 | 8.51E-07 | 4390 | 1.23E-05 | TRUE | 0.01245290594 | TRUE | 0.01053266785 | TRUE | FALSE | 1.015583018 |
| enpp7 | LV end diastolic volume \| Instance 2(participant - p24100_i2)_BSA_indexed_corrected | -1.1231939 | 0.1320050019 | -0.08627998993 | 2.19E-08 | 0.2003140774 | -0.08627998993 | 2.19E-08 | 4194 | 4.26E-07 | TRUE | 0.0003202500666 | TRUE | 0.0002708675063 | TRUE | FALSE | -1.1231939 |
| enpp7 | RV end diastolic volume \| Instance 2_BSA_indexed_corrected | -1.098538655 | 0.06630996311 | -0.07847375729 | 3.61E-07 | 0.2155454719 | -0.07847375729 | 3.61E-07 | 4194 | 5.61E-06 | TRUE | 0.005283410138 | TRUE | 0.004468708295 | TRUE | FALSE | -1.098538655 |
| epha1 | LV end diastolic volume \| Instance 2(participant - p24100_i2)_BSA_indexed_corrected | -3.32189145 | 0.08658797202 | -0.1084747711 | 9.19E-13 | 0.4637114128 | -0.1084747711 | 9.19E-13 | 4312 | 3.98E-11 | TRUE | 1.34E-08 | TRUE | 1.14E-08 | TRUE | FALSE | -3.32189145 |
| epha1 | LV end systolic volume \| Instance 2(participant - p24101_i2)_BSA_indexed_corrected | -1.432932403 | 0.1995728353 | -0.08073449266 | 1.10E-07 | 0.2694684577 | -0.08073449266 | 1.10E-07 | 4312 | 1.90E-06 | TRUE | 0.001615945222 | TRUE | 0.001366766468 | TRUE | FALSE | -1.432932403 |
| epha1 | RV end diastolic volume \| Instance 2_BSA_indexed_corrected | -3.891987118 | -0.06352823627 | -0.11823895 | 6.74E-15 | 0.4978688536 | -0.11823895 | 6.74E-15 | 4312 | 4.23E-13 | TRUE | 9.85E-11 | TRUE | 8.33E-11 | TRUE | FALSE | -3.891987118 |
| epha1 | RV end systolic volume \| Instance 2_BSA_indexed_corrected | -1.808510659 | 0.07807149621 | -0.09241721057 | 1.20E-09 | 0.2968021033 | -0.09241721057 | 1.20E-09 | 4312 | 2.92E-08 | TRUE | 1.76E-05 | TRUE | 1.49E-05 | TRUE | FALSE | -1.808510659 |
| epha2 | RV end diastolic volume \| Instance 2_BSA_indexed_corrected | -3.668458908 | -0.06578404324 | -0.08572514898 | 2.90E-08 | 0.6600085377 | -0.08572514898 | 2.90E-08 | 4175 | 5.50E-07 | TRUE | 0.0004236597078 | TRUE | 0.0003583313809 | TRUE | FALSE | -3.668458908 |
| epha2 | RV end systolic volume \| Instance 2_BSA_indexed_corrected | -1.847242711 | 0.05064338602 | -0.07254306242 | 2.70E-06 | 0.3931498641 | -0.07254306242 | 2.70E-06 | 4175 | 3.56E-05 | TRUE | 0.03956287475 | TRUE | 0.03346227946 | TRUE | FALSE | -1.847242711 |
| ephb6 | LV end diastolic volume \| Instance 2(participant - p24100_i2)_BSA_indexed_corrected | -2.677764466 | 0.3036471332 | -0.0706353931 | 2.92E-06 | 0.5718394745 | -0.0706353931 | 2.92E-06 | 4375 | 3.81E-05 | TRUE | 0.04267159377 | TRUE | 0.03609163401 | TRUE | FALSE | -2.677764466 |
| ephb6 | RV end diastolic volume \| Instance 2_BSA_indexed_corrected | -3.342841453 | 0.1818129898 | -0.08196305084 | 5.67E-08 | 0.6146731183 | -0.08196305084 | 5.67E-08 | 4375 | 1.02E-06 | TRUE | 0.0008294034247 | TRUE | 0.0007015094166 | TRUE | FALSE | -3.342841453 |
| ephb6 | RV end systolic volume \| Instance 2_BSA_indexed_corrected | -1.711722454 | 0.1855636449 | -0.07057449576 | 2.97E-06 | 0.3658571648 | -0.07057449576 | 2.97E-06 | 4375 | 3.88E-05 | TRUE | 0.04352028544 | TRUE | 0.03680945742 | TRUE | FALSE | -1.711722454 |
| erbb2 | LV end diastolic volume \| Instance 2(participant - p24100_i2)_BSA_indexed_corrected | -6.011899403 | 0.1581961464 | -0.1423898124 | 4.27E-21 | 0.634512438 | -0.1423898124 | 4.27E-21 | 4340 | 7.10E-19 | TRUE | 6.24E-17 | TRUE | 5.28E-17 | TRUE | FALSE | -6.011899403 |
| erbb2 | LV end systolic volume \| Instance 2(participant - p24101_i2)_BSA_indexed_corrected | -2.901677499 | 0.2161607666 | -0.1184156838 | 5.02E-15 | 0.3694270507 | -0.1184156838 | 5.02E-15 | 4340 | 3.24E-13 | TRUE | 7.35E-11 | TRUE | 6.21E-11 | TRUE | FALSE | -2.901677499 |
| erbb2 | RV end diastolic volume \| Instance 2_BSA_indexed_corrected | -5.561203762 | 0.06874012806 | -0.1224647702 | 5.68E-16 | 0.6842763057 | -0.1224647702 | 5.68E-16 | 4340 | 4.35E-14 | TRUE | 8.31E-12 | TRUE | 7.03E-12 | TRUE | FALSE | -5.561203762 |
| erbb2 | RV end systolic volume \| Instance 2_BSA_indexed_corrected | -2.388741037 | 0.1355701999 | -0.08842393077 | 5.38E-09 | 0.4085544812 | -0.08842393077 | 5.38E-09 | 4340 | 1.14E-07 | TRUE | 7.87E-05 | TRUE | 6.66E-05 | TRUE | FALSE | -2.388741037 |
| erp44 | LV end diastolic volume \| Instance 2(participant - p24100_i2)_BSA_indexed_corrected | -3.101174721 | 0.275232587 | -0.07313011835 | 1.29E-06 | 0.6396982058 | -0.07313011835 | 1.29E-06 | 4373 | 1.80E-05 | TRUE | 0.01889086913 | TRUE | 0.01597789711 | TRUE | FALSE | -3.101174721 |
| erp44 | RV end diastolic volume \| Instance 2_BSA_indexed_corrected | -3.374347602 | 0.1597568384 | -0.07396810772 | 9.74E-07 | 0.6881190342 | -0.07396810772 | 9.74E-07 | 4373 | 1.39E-05 | TRUE | 0.01425669734 | TRUE | 0.01205831461 | TRUE | FALSE | -3.374347602 |
| esm1 | LV end diastolic volume \| Instance 2(participant - p24100_i2)_BSA_indexed_corrected | 3.863477282 | 0.298079749 | 0.1197833533 | 3.25E-15 | 0.4884391679 | 0.1197833533 | 3.25E-15 | 4300 | 2.16E-13 | TRUE | 4.76E-11 | TRUE | 4.02E-11 | TRUE | FALSE | 3.863477282 |
| esm1 | LV end systolic volume \| Instance 2(participant - p24101_i2)_BSA_indexed_corrected | 1.960648261 | 0.272701837 | 0.1049030323 | 5.35E-12 | 0.2835146904 | 0.1049030323 | 5.35E-12 | 4300 | 2.05E-10 | TRUE | 7.83E-08 | TRUE | 6.62E-08 | TRUE | FALSE | 1.960648261 |
| esm1 | RV end diastolic volume \| Instance 2_BSA_indexed_corrected | 3.998209172 | 0.183367546 | 0.1153494325 | 3.27E-14 | 0.5251800241 | 0.1153494325 | 3.27E-14 | 4300 | 1.83E-12 | TRUE | 4.79E-10 | TRUE | 4.05E-10 | TRUE | FALSE | 3.998209172 |
| esm1 | RV end systolic volume \| Instance 2_BSA_indexed_corrected | 1.943178011 | 0.1684843792 | 0.09437022828 | 5.64E-10 | 0.3126812624 | 0.09437022828 | 5.64E-10 | 4300 | 1.48E-08 | TRUE | 8.24E-06 | TRUE | 6.97E-06 | TRUE | FALSE | 1.943178011 |
| ezr | LV end diastolic volume \| Instance 2(participant - p24100_i2)_BSA_indexed_corrected | -3.866559848 | 0.1514370547 | -0.08586634029 | 1.34E-08 | 0.6793628212 | -0.08586634029 | 1.34E-08 | 4363 | 2.70E-07 | TRUE | 0.0001963699334 | TRUE | 0.0001660896897 | TRUE | FALSE | -3.866559848 |
| ezr | RV end diastolic volume \| Instance 2_BSA_indexed_corrected | -3.92047972 | 0.03554516535 | -0.08086437878 | 8.87E-08 | 0.7317526683 | -0.08086437878 | 8.87E-08 | 4363 | 1.55E-06 | TRUE | 0.001297008798 | TRUE | 0.001097010041 | TRUE | FALSE | -3.92047972 |
| f7 | LV end diastolic volume \| Instance 2(participant - p24100_i2)_BSA_indexed_corrected | -6.038699506 | 0.01982540834 | -0.1628672258 | 4.40E-27 | 0.5565187807 | -0.1628672258 | 4.40E-27 | 4323 | 1.74E-24 | TRUE | 6.44E-23 | TRUE | 5.45E-23 | TRUE | FALSE | -6.038699506 |
| f7 | LV end systolic volume \| Instance 2(participant - p24101_i2)_BSA_indexed_corrected | -2.68126976 | 0.16719866 | -0.1245370749 | 2.07E-16 | 0.3249791397 | -0.1245370749 | 2.07E-16 | 4323 | 1.72E-14 | TRUE | 3.03E-12 | TRUE | 2.56E-12 | TRUE | FALSE | -2.68126976 |
| f7 | RV end diastolic volume \| Instance 2_BSA_indexed_corrected | -6.817693435 | -0.1353125793 | -0.1710674085 | 9.54E-30 | 0.5973496509 | -0.1710674085 | 9.54E-30 | 4323 | 5.17E-27 | TRUE | 1.40E-25 | TRUE | 1.18E-25 | TRUE | FALSE | -6.817693435 |
| f7 | RV end systolic volume \| Instance 2_BSA_indexed_corrected | -3.038684715 | 0.06163474159 | -0.1282326708 | 2.59E-17 | 0.3575151505 | -0.1282326708 | 2.59E-17 | 4323 | 2.38E-15 | TRUE | 3.79E-13 | TRUE | 3.20E-13 | TRUE | FALSE | -3.038684715 |
| f9 | LV end diastolic volume \| Instance 2(participant - p24100_i2)_BSA_indexed_corrected | -8.966424725 | -0.04583249827 | -0.1551530318 | 1.07E-24 | 0.8686122562 | -0.1551530318 | 1.07E-24 | 4322 | 3.28E-22 | TRUE | 1.57E-20 | TRUE | 1.33E-20 | TRUE | FALSE | -8.966424725 |
| f9 | LV end systolic volume \| Instance 2(participant - p24101_i2)_BSA_indexed_corrected | -4.130627878 | 0.1299272786 | -0.1230752854 | 4.67E-16 | 0.5067443755 | -0.1230752854 | 4.67E-16 | 4322 | 3.64E-14 | TRUE | 6.83E-12 | TRUE | 5.78E-12 | TRUE | FALSE | -4.130627878 |
| f9 | RV end diastolic volume \| Instance 2_BSA_indexed_corrected | -9.893578447 | -0.1992204694 | -0.1592621432 | 5.99E-26 | 0.9330826219 | -0.1592621432 | 5.99E-26 | 4322 | 2.09E-23 | TRUE | 8.77E-22 | TRUE | 7.41E-22 | TRUE | FALSE | -9.893578447 |
| f9 | RV end systolic volume \| Instance 2_BSA_indexed_corrected | -4.377883387 | 0.03460015327 | -0.1185189726 | 5.41E-15 | 0.5580364918 | -0.1185189726 | 5.41E-15 | 4322 | 3.47E-13 | TRUE | 7.92E-11 | TRUE | 6.70E-11 | TRUE | FALSE | -4.377883387 |
| fabp1 | RV end diastolic volume \| Instance 2_BSA_indexed_corrected | -0.9503072472 | 0.08192469103 | -0.07682674907 | 4.56E-07 | 0.1881191046 | -0.07682674907 | 4.56E-07 | 4300 | 6.97E-06 | TRUE | 0.006674768303 | TRUE | 0.005645518802 | TRUE | FALSE | -0.9503072472 |
| fabp1 | RV end systolic volume \| Instance 2_BSA_indexed_corrected | -0.5306938849 | 0.10744333 | -0.07222124412 | 2.13E-06 | 0.1117918976 | -0.07222124412 | 2.13E-06 | 4300 | 2.87E-05 | TRUE | 0.03116250546 | TRUE | 0.02635724712 | TRUE | FALSE | -0.5306938849 |
| fabp4 | LV end diastolic volume \| Instance 2(participant - p24100_i2)_BSA_indexed_corrected | -4.168137269 | -0.3618648185 | -0.2356031333 | 3.01E-56 | 0.2599980928 | -0.2356031333 | 3.01E-56 | 4375 | 1.47E-52 | TRUE | 4.40E-52 | TRUE | 3.72E-52 | TRUE | FALSE | -4.168137269 |
| fabp4 | LV end systolic volume \| Instance 2(participant - p24101_i2)_BSA_indexed_corrected | -1.871860292 | -0.009132181819 | -0.182272643 | 5.42E-34 | 0.152695165 | -0.182272643 | 5.42E-34 | 4375 | 5.28E-31 | TRUE | 7.92E-30 | TRUE | 6.70E-30 | TRUE | FALSE | -1.871860292 |

| | | | | | | | | | | | | | | | | | |
|---|---|---|---|---|---|---|---|---|---|---|---|---|---|---|---|---|---|
| **fabp4** | RV end diastolic volume \| Instance 2_BSA_indexed_corrected | -4.755588111 | -0.5721915052 | -0.2498593612 | 2.94E-63 | 0.2786896173 | -0.2498593612 | 2.94E-63 | 4375 | 4.31E-59 | TRUE | 4.31E-59 | TRUE | 3.64E-59 | TRUE | FALSE | -4.755588111 |
| **fabp4** | RV end systolic volume \| Instance 2_BSA_indexed_corrected | -2.100780293 | -0.1431847821 | -0.1856026306 | 3.33E-35 | 0.1681878144 | -0.1856026306 | 3.33E-35 | 4375 | 4.42E-32 | TRUE | 4.87E-31 | TRUE | 4.12E-31 | TRUE | FALSE | -2.100780293 |
| **fabp5** | LV end diastolic volume \| Instance 2(participant - p24100_i2)_BSA_indexed_corrected | -1.6239455 | 0.1995975383 | -0.09903569636 | 5.91E-11 | 0.2474540773 | -0.09903569636 | 5.91E-11 | 4350 | 1.85E-09 | TRUE | 8.64E-07 | TRUE | 7.31E-07 | TRUE | FALSE | -1.6239455 |
| **fabp5** | LV end systolic volume \| Instance 2(participant - p24101_i2)_BSA_indexed_corrected | -0.7522054675 | 0.2313228114 | -0.07918603061 | 1.70E-07 | 0.143607543 | -0.07918603061 | 1.70E-07 | 4350 | 2.81E-06 | TRUE | 0.002488137622 | TRUE | 0.002104466801 | TRUE | FALSE | -0.7522054675 |
| **fabp5** | RV end diastolic volume \| Instance 2_BSA_indexed_corrected | -1.794503608 | 0.07901366442 | -0.1015452242 | 1.91E-11 | 0.2666179237 | -0.1015452242 | 1.91E-11 | 4350 | 6.62E-10 | TRUE | 2.79E-07 | TRUE | 2.36E-07 | TRUE | FALSE | -1.794503608 |
| **fabp5** | RV end systolic volume \| Instance 2_BSA_indexed_corrected | -0.8220748965 | 0.1408312562 | -0.07826673068 | 2.36E-07 | 0.1588017047 | -0.07826673068 | 2.36E-07 | 4350 | 3.81E-06 | TRUE | 0.003452551369 | TRUE | 0.002920167948 | TRUE | FALSE | -0.8220748965 |
| **fam3c** | RV end diastolic volume \| Instance 2_BSA_indexed_corrected | -3.090198948 | 0.06324757256 | -0.07430124522 | 8.68E-07 | 0.6272608047 | -0.07430124522 | 8.68E-07 | 4374 | 1.25E-05 | TRUE | 0.01270029445 | TRUE | 0.01074190905 | TRUE | FALSE | -3.090198948 |
| **fas** | LV end diastolic volume \| Instance 2(participant - p24100_i2)_BSA_indexed_corrected | -2.440926923 | 0.2289702066 | -0.07621513986 | 4.49E-07 | 0.482901725 | -0.07621513986 | 4.49E-07 | 4375 | 6.86E-06 | TRUE | 0.006563937101 | TRUE | 0.005551778 | TRUE | FALSE | -2.440926923 |
| **fas** | RV end diastolic volume \| Instance 2_BSA_indexed_corrected | -2.679981184 | 0.10724915 | -0.07778046984 | 2.59E-07 | 0.5194619677 | -0.07778046984 | 2.59E-07 | 4375 | 4.14E-06 | TRUE | 0.003790063991 | TRUE | 0.003205636124 | TRUE | FALSE | -2.679981184 |
| **fbp1** | LV end diastolic volume \| Instance 2(participant - p24100_i2)_BSA_indexed_corrected | -1.084621352 | 0.272571074 | -0.07529591254 | 6.16E-07 | 0.2172112907 | -0.07529591254 | 6.16E-07 | 4375 | 9.16E-06 | TRUE | 0.009017676079 | TRUE | 0.007627150428 | TRUE | FALSE | -1.084621352 |
| **fcar** | LV end diastolic volume \| Instance 2(participant - p24100_i2)_BSA_indexed_corrected | -1.722795329 | 0.2817251208 | -0.07231363364 | 1.89E-06 | 0.3611029644 | -0.07231363364 | 1.89E-06 | 4332 | 2.56E-05 | TRUE | 0.02771184328 | TRUE | 0.02343867704 | TRUE | FALSE | -1.722795329 |
| **fcar** | RV end diastolic volume \| Instance 2_BSA_indexed_corrected | -2.087861522 | 0.1595607372 | -0.08147322976 | 7.88E-08 | 0.388147332 | -0.08147322976 | 7.88E-08 | 4332 | 1.39E-06 | TRUE | 0.001153320939 | TRUE | 0.0009754788502 | TRUE | FALSE | -2.087861522 |
| **fcgr2b** | LV end diastolic volume \| Instance 2(participant - p24100_i2)_BSA_indexed_corrected | -1.366778378 | 0.3551973252 | -0.07026892185 | 3.28E-06 | 0.2934068186 | -0.07026892185 | 3.28E-06 | 4375 | 4.27E-05 | TRUE | 0.0480291333 | TRUE | 0.04062304094 | TRUE | FALSE | -1.366778378 |
| **fcgr2b** | RV end systolic volume \| Instance 2_BSA_indexed_corrected | -0.9055685481 | 0.2185672574 | -0.07276976371 | 1.45E-06 | 0.1876841862 | -0.07276976371 | 1.45E-06 | 4375 | 2.00E-05 | TRUE | 0.02118431455 | TRUE | 0.01791769324 | TRUE | FALSE | -0.9055685481 |
| **fcn2** | LV end diastolic volume \| Instance 2(participant - p24100_i2)_BSA_indexed_corrected | -2.681411233 | 0.1895183562 | -0.098849176 | 8.59E-11 | 0.4121255221 | -0.098849176 | 8.59E-11 | 4292 | 2.58E-09 | TRUE | 1.26E-06 | TRUE | 1.06E-06 | TRUE | FALSE | -2.681411233 |
| **fcn2** | LV end systolic volume \| Instance 2(participant - p24101_i2)_BSA_indexed_corrected | -1.201093677 | 0.2473906671 | -0.0761821872 | 5.83E-07 | 0.2400108435 | -0.0761821872 | 5.83E-07 | 4292 | 8.73E-06 | TRUE | 0.008529538863 | TRUE | 0.00721428397 | TRUE | FALSE | -1.201093677 |
| **fcn2** | RV end diastolic volume \| Instance 2_BSA_indexed_corrected | -2.789225955 | 0.05894984739 | -0.09565511502 | 3.40E-10 | 0.4431500801 | -0.09565511502 | 3.40E-10 | 4292 | 9.30E-09 | TRUE | 4.98E-06 | TRUE | 4.21E-06 | TRUE | FALSE | -2.789225955 |
| **fcn2** | RV end systolic volume \| Instance 2_BSA_indexed_corrected | -1.310827872 | 0.142413873 | -0.0756333472 | 7.03E-07 | 0.2638505259 | -0.0756333472 | 7.03E-07 | 4292 | 1.03E-05 | TRUE | 0.0102783035 | TRUE | 0.008693389098 | TRUE | FALSE | -1.310827872 |
| **fen1** | LV end diastolic volume \| Instance 2(participant - p24100_i2)_BSA_indexed_corrected | -1.447983459 | 0.4362939129 | -0.08579870799 | 1.32E-08 | 0.2542662334 | -0.08579870799 | 1.32E-08 | 4375 | 2.66E-07 | TRUE | 0.0001926003895 | TRUE | 0.0001629014094 | TRUE | FALSE | -1.447983459 |
| **fetub** | LV end diastolic volume \| Instance 2(participant - p24100_i2)_BSA_indexed_corrected | -2.427211713 | 0.2637961119 | -0.08314047943 | 4.41E-08 | 0.4426363277 | -0.08314047943 | 4.41E-08 | 4322 | 8.11E-07 | TRUE | 0.0006447513272 | TRUE | 0.0005453306726 | TRUE | FALSE | -2.427211713 |
| **fetub** | RV end diastolic volume \| Instance 2_BSA_indexed_corrected | -2.786516545 | 0.1369458836 | -0.0887942178 | 4.99E-09 | 0.4755719192 | -0.0887942178 | 4.99E-09 | 4322 | 1.07E-07 | TRUE | 7.31E-05 | TRUE | 6.18E-05 | TRUE | FALSE | -2.786516545 |
| **fgf21** | LV end diastolic volume \| Instance 2(participant - p24100_i2)_BSA_indexed_corrected | -1.330739288 | 0.08723344925 | -0.1429440157 | 2.08E-21 | 0.1393331118 | -0.1429440157 | 2.08E-21 | 4375 | 3.58E-19 | TRUE | 3.04E-17 | TRUE | 2.57E-17 | TRUE | FALSE | -1.330739288 |
| **fgf21** | LV end systolic volume \| Instance 2(participant - p24101_i2)_BSA_indexed_corrected | -0.6788489887 | 0.1763288041 | -0.1256189578 | 7.45E-17 | 0.08107264028 | -0.1256189578 | 7.45E-17 | 4375 | 6.49E-15 | TRUE | 1.09E-12 | TRUE | 9.22E-13 | TRUE | FALSE | -0.6788489887 |
| **fgf21** | RV end diastolic volume \| Instance 2_BSA_indexed_corrected | -1.475946388 | -0.05134062497 | -0.1473655354 | 1.15E-22 | 0.1498018785 | -0.1473655354 | 1.15E-22 | 4375 | 2.33E-20 | TRUE | 1.68E-18 | TRUE | 1.42E-18 | TRUE | FALSE | -1.475946388 |
| **fgf21** | RV end systolic volume \| Instance 2_BSA_indexed_corrected | -0.6876050319 | 0.07978962083 | -0.115445362 | 1.87E-14 | 0.08946626373 | -0.115445362 | 1.87E-14 | 4375 | 1.09E-12 | TRUE | 2.73E-10 | TRUE | 2.31E-10 | TRUE | FALSE | -0.6876050319 |
| **fgfr2** | LV end diastolic volume \| Instance 2(participant - p24100_i2)_BSA_indexed_corrected | -4.847664516 | 0.1292756756 | -0.09521143961 | 3.28E-10 | 0.7695222601 | -0.09521143961 | 3.28E-10 | 4340 | 9.01E-09 | TRUE | 4.80E-06 | TRUE | 4.06E-06 | TRUE | FALSE | -4.847664516 |
| **fgfr2** | RV end diastolic volume \| Instance 2_BSA_indexed_corrected | -5.547288455 | -0.008373116532 | -0.1013007069 | 2.25E-11 | 0.8271488283 | -0.1013007069 | 2.25E-11 | 4340 | 7.69E-10 | TRUE | 3.29E-07 | TRUE | 2.78E-07 | TRUE | FALSE | -5.547288455 |
| **fgfr2** | RV end systolic volume \| Instance 2_BSA_indexed_corrected | -2.57023258 | 0.09356602036 | -0.0788973712 | 1.95E-07 | 0.4930706998 | -0.0788973712 | 1.95E-07 | 4340 | 3.19E-06 | TRUE | 0.00284933397 | TRUE | 0.002409966672 | TRUE | FALSE | -2.57023258 |
| **fgr** | LV end diastolic volume \| Instance 2(participant - p24100_i2)_BSA_indexed_corrected | -1.977283045 | 0.3440635877 | -0.1040105695 | 9.29E-12 | 0.289243082 | -0.1040105695 | 9.29E-12 | 4275 | 3.39E-10 | TRUE | 1.36E-07 | TRUE | 1.15E-07 | TRUE | FALSE | -1.977283045 |
| **fgr** | LV end systolic volume \| Instance 2(participant - p24101_i2)_BSA_indexed_corrected | -1.031744214 | 0.292427757 | -0.09368094726 | 8.42E-10 | 0.1677413756 | -0.09368094726 | 8.42E-10 | 4275 | 2.12E-08 | TRUE | 1.23E-05 | TRUE | 1.04E-05 | TRUE | FALSE | -1.031744214 |
| **fgr** | RV end diastolic volume \| Instance 2_BSA_indexed_corrected | -1.934359625 | 0.2261627742 | -0.09462768658 | 5.68E-10 | 0.311314514 | -0.09462768658 | 5.68E-10 | 4275 | 1.49E-08 | TRUE | 8.30E-06 | TRUE | 7.02E-06 | TRUE | FALSE | -1.934359625 |
| **fgr** | RV end systolic volume \| Instance 2_BSA_indexed_corrected | -1.082409778 | 0.197770287 | -0.08919381991 | 5.17E-09 | 0.184908155 | -0.08919381991 | 5.17E-09 | 4275 | 1.10E-07 | TRUE | 7.56E-05 | TRUE | 6.39E-05 | TRUE | FALSE | -1.082409778 |
| **flt4** | LV end diastolic volume \| Instance 2(participant - p24100_i2)_BSA_indexed_corrected | -2.688294863 | 0.1652110925 | -0.08380291165 | 3.22E-08 | 0.4853361387 | -0.08380291165 | 3.22E-08 | 4340 | 6.07E-07 | TRUE | 0.0004712775202 | TRUE | 0.0003986065266 | TRUE | FALSE | -2.688294863 |
| **flt4** | RV end diastolic volume \| Instance 2_BSA_indexed_corrected | -3.005127179 | 0.03787547743 | -0.08710041285 | 9.07E-09 | 0.5218482179 | -0.08710041285 | 9.07E-09 | 4340 | 1.87E-07 | TRUE | 0.0001326394234 | TRUE | 0.0001121864243 | TRUE | FALSE | -3.005127179 |
| **flt4** | RV end systolic volume \| Instance 2_BSA_indexed_corrected | -1.560110403 | 0.1029069692 | -0.07601005947 | 5.35E-07 | 0.3107286326 | -0.07601005947 | 5.35E-07 | 4340 | 8.06E-06 | TRUE | 0.007829598798 | TRUE | 0.006622274663 | TRUE | FALSE | -1.560110403 |
| **fmnl1** | LV end diastolic volume \| Instance 2(participant - p24100_i2)_BSA_indexed_corrected | -1.510558121 | 0.3011501603 | -0.08707992813 | 1.18E-08 | 0.2643626204 | -0.08707992813 | 1.18E-08 | 4275 | 2.40E-07 | TRUE | 0.0001724635132 | TRUE | 0.0001458696395 | TRUE | FALSE | -1.510558121 |
| **fmnl1** | LV end systolic volume \| Instance 2(participant - p24101_i2)_BSA_indexed_corrected | -0.784768657 | 0.2701349088 | -0.07808954112 | 3.19E-07 | 0.1532689279 | -0.07808954112 | 3.19E-07 | 4275 | 5.00E-06 | TRUE | 0.004661555324 | TRUE | 0.003942743493 | TRUE | FALSE | -0.784768657 |
| **fmnl1** | RV end diastolic volume \| Instance 2_BSA_indexed_corrected | -1.34088886 | 0.1881345581 | -0.07188624132 | 2.54E-06 | 0.2846135114 | -0.07188624132 | 2.54E-06 | 4275 | 3.35E-05 | TRUE | 0.03716134822 | TRUE | 0.03143106832 | TRUE | FALSE | -1.34088886 |
| **folr2** | RV end diastolic volume \| Instance 2_BSA_indexed_corrected | -3.105839495 | 0.09536101563 | -0.07640685835 | 4.35E-07 | 0.6137368496 | -0.07640685835 | 4.35E-07 | 4363 | 6.67E-06 | TRUE | 0.006368132844 | TRUE | 0.00538616676 | TRUE | FALSE | -3.105839495 |
| **folr2** | RV end systolic volume \| Instance 2_BSA_indexed_corrected | -1.779102968 | 0.1343245059 | -0.07365447759 | 1.11E-06 | 0.364777179 | -0.07365447759 | 1.11E-06 | 4363 | 1.57E-05 | TRUE | 0.01630025525 | TRUE | 0.01378675589 | TRUE | FALSE | -1.779102968 |
| **fstl3** | LV end diastolic volume \| Instance 2(participant - p24100_i2)_BSA_indexed_corrected | -4.722083252 | -0.01993838181 | -0.1276979344 | 4.47E-17 | 0.559821732 | -0.1276979344 | 4.47E-17 | 4294 | 3.99E-15 | TRUE | 6.54E-13 | TRUE | 5.53E-13 | TRUE | FALSE | -4.722083252 |
| **fstl3** | LV end systolic volume \| Instance 2(participant - p24101_i2)_BSA_indexed_corrected | -1.83435882 | 0.1689287425 | -0.08558492074 | 1.94E-08 | 0.3259573463 | -0.08558492074 | 1.94E-08 | 4294 | 3.82E-07 | TRUE | 0.0002843667963 | TRUE | 0.0002405174363 | TRUE | FALSE | -1.83435882 |
| **fstl3** | RV end diastolic volume \| Instance 2_BSA_indexed_corrected | -5.915083371 | -0.2092665298 | -0.1487470759 | 1.14E-22 | 0.6002388405 | -0.1487470759 | 1.14E-22 | 4294 | 2.33E-20 | TRUE | 1.67E-18 | TRUE | 1.41E-18 | TRUE | FALSE | -5.915083371 |
| **fstl3** | RV end systolic volume \| Instance 2_BSA_indexed_corrected | -2.945026948 | -0.01127783899 | -0.1245748525 | 2.55E-16 | 0.358040972 | -0.1245748525 | 2.55E-16 | 4294 | 2.07E-14 | TRUE | 3.73E-12 | TRUE | 3.16E-12 | TRUE | FALSE | -2.945026948 |
| **furin** | LV end diastolic volume \| Instance 2(participant - p24100_i2)_BSA_indexed_corrected | -4.927683633 | -0.08912306915 | -0.1650724095 | 6.88E-28 | 0.4470173897 | -0.1650724095 | 6.88E-28 | 4340 | 2.96E-25 | TRUE | 1.01E-23 | TRUE | 8.52E-24 | TRUE | FALSE | -4.927683633 |
| **furin** | LV end systolic volume \| Instance 2(participant - p24101_i2)_BSA_indexed_corrected | -2.096619511 | 0.122409558 | -0.1210163831 | 1.25E-15 | 0.2611121319 | -0.1210163831 | 1.25E-15 | 4340 | 8.96E-14 | TRUE | 1.83E-11 | TRUE | 1.55E-11 | TRUE | FALSE | -2.096619511 |
| **furin** | RV end diastolic volume \| Instance 2_BSA_indexed_corrected | -5.640375624 | -0.2584297393 | -0.1756769777 | 2.02E-31 | 0.4798893627 | -0.1756769777 | 2.02E-31 | 4340 | 1.34E-28 | TRUE | 2.96E-27 | TRUE | 2.50E-27 | TRUE | FALSE | -5.640375624 |
| **furin** | RV end systolic volume \| Instance 2_BSA_indexed_corrected | -2.291948949 | 0.006931777735 | -0.1199969331 | 2.16E-15 | 0.287899247 | -0.1199969331 | 2.16E-15 | 4340 | 1.49E-13 | TRUE | 3.16E-11 | TRUE | 2.68E-11 | TRUE | FALSE | -2.291948949 |
| **fus** | LV end diastolic volume \| Instance 2(participant - p24100_i2)_BSA_indexed_corrected | -2.366498063 | 0.2771700776 | -0.08350121219 | 3.26E-08 | 0.4274173582 | -0.08350121219 | 3.26E-08 | 4368 | 6.13E-07 | TRUE | 0.0004772833899 | TRUE | 0.0004036862912 | TRUE | FALSE | -2.366498063 |
| **fus** | RV end diastolic volume \| Instance 2_BSA_indexed_corrected | -2.393255661 | 0.1668847042 | -0.0784905081 | 2.06E-07 | 0.4600321234 | -0.0784905081 | 2.06E-07 | 4368 | 3.36E-06 | TRUE | 0.003010645793 | TRUE | 0.002546404212 | TRUE | FALSE | -2.393255661 |
| **gcg** | LV end diastolic volume \| Instance 2(participant - p24100_i2)_BSA_indexed_corrected | -0.7370473328 | 0.3247232702 | -0.08379447596 | 2.85E-08 | 0.1325440298 | -0.08379447596 | 2.85E-08 | 4375 | 5.41E-07 | TRUE | 0.0004163512047 | TRUE | 0.000352149849 | TRUE | FALSE | -0.7370473328 |
| **gcg** | RV end diastolic volume \| Instance 2_BSA_indexed_corrected | -0.8372922704 | 0.2113027188 | -0.08848088063 | 4.57E-09 | 0.1425381772 | -0.08848088063 | 4.57E-09 | 4375 | 9.92E-08 | TRUE | 6.68E-05 | TRUE | 5.65E-05 | TRUE | FALSE | -0.8372922704 |
| **gdf15** | LV end diastolic volume \| Instance 2(participant - p24100_i2)_BSA_indexed_corrected | -2.574469917 | 0.00474139963 | -0.1002738873 | 2.98E-11 | 0.3862920736 | -0.1002738873 | 2.98E-11 | 4375 | 9.92E-10 | TRUE | 4.37E-07 | TRUE | 3.69E-07 | TRUE | FALSE | -2.574469917 |
| **gdf15** | LV end systolic volume \| Instance 2(participant - p24101_i2)_BSA_indexed_corrected | -1.142090736 | 0.1574100142 | -0.07663190987 | 3.88E-07 | 0.2247099356 | -0.07663190987 | 3.88E-07 | 4375 | 6.00E-06 | TRUE | 0.005676846152 | TRUE | 0.004801476476 | TRUE | FALSE | -1.142090736 |
| **gdf15** | RV end diastolic volume \| Instance 2_BSA_indexed_corrected | -3.22956307 | -0.1934520617 | -0.1169219754 | 8.62E-15 | 0.4148287462 | -0.1169219754 | 8.62E-15 | 4375 | 5.27E-13 | TRUE | 1.26E-10 | TRUE | 1.07E-10 | TRUE | FALSE | -3.22956307 |
| **gdf15** | RV end systolic volume \| Instance 2_BSA_indexed_corrected | -1.495757164 | 0.01477572032 | -0.09105956377 | 1.60E-09 | 0.2473647033 | -0.09105956377 | 1.60E-09 | 4375 | 3.74E-08 | TRUE | 2.34E-05 | TRUE | 1.98E-05 | TRUE | FALSE | -1.495757164 |
| **gfra1** | LV end diastolic volume \| Instance 2(participant - p24100_i2)_BSA_indexed_corrected | -4.189291264 | 0.06853313362 | -0.100879211 | 2.27E-11 | 0.6247812611 | -0.100879211 | 2.27E-11 | 4375 | 7.73E-10 | TRUE | 3.32E-07 | TRUE | 2.81E-07 | TRUE | FALSE | -4.189291264 |
| **gfra1** | LV end systolic volume \| Instance 2(participant - p24101_i2)_BSA_indexed_corrected | -1.870973255 | 0.1848597116 | -0.0776135791 | 2.75E-07 | 0.363436137 | -0.0776135791 | 2.75E-07 | 4375 | 4.38E-06 | TRUE | 0.00402064028 | TRUE | 0.003400657549 | TRUE | FALSE | -1.870973255 |
| **gfra1** | RV end diastolic volume \| Instance 2_BSA_indexed_corrected | -5.964349967 | -0.161578283 | -0.1334985607 | 7.49E-19 | 0.6695637409 | -0.1334985607 | 7.49E-19 | 4375 | 8.63E-17 | TRUE | 1.10E-14 | TRUE | 9.27E-15 | TRUE | FALSE | -5.964349967 |
| **gfra1** | RV end systolic volume \| Instance 2_BSA_indexed_corrected | -2.869140431 | 0.02228685156 | -0.1079884656 | 7.97E-13 | 0.3994271222 | -0.1079884656 | 7.97E-13 | 4375 | 3.50E-11 | TRUE | 1.17E-08 | TRUE | 9.86E-09 | TRUE | FALSE | -2.869140431 |
| **ggh** | LV end diastolic volume \| Instance 2(participant - p24100_i2)_BSA_indexed_corrected | -4.42209043 | 0.05214086255 | -0.1372780058 | 2.42E-19 | 0.489384106 | -0.1372780058 | 2.42E-19 | 4253 | 3.02E-17 | TRUE | 3.53E-15 | TRUE | 2.99E-15 | TRUE | FALSE | -4.42209043 |
| **ggh** | LV end systolic volume \| Instance 2(participant - p24101_i2)_BSA_indexed_corrected | -2.042338871 | 0.1693336345 | -0.1092463058 | 9.08E-13 | 0.2850153841 | -0.1092463058 | 9.08E-13 | 4253 | 3.94E-11 | TRUE | 1.33E-08 | TRUE | 1.12E-08 | TRUE | FALSE | -2.042338871 |
| **ggh** | RV end diastolic volume \| Instance 2_BSA_indexed_corrected | -5.154889386 | -0.1160446275 | -0.14903046 | 1.50E-22 | 0.5245918443 | -0.14903046 | 1.50E-22 | 4253 | 2.98E-20 | TRUE | 2.20E-18 | TRUE | 1.86E-18 | TRUE | FALSE | -5.154889386 |
| **ggh** | RV end systolic volume \| Instance 2_BSA_indexed_corrected | -2.072895724 | 0.07275115305 | -0.1007568399 | 4.53E-11 | 0.3139363707 | -0.1007568399 | 4.53E-11 | 4253 | 1.46E-09 | TRUE | 6.62E-07 | TRUE | 5.60E-07 | TRUE | FALSE | -2.072895724 |
| **ggt1** | LV end diastolic volume \| Instance 2(participant - p24100_i2)_BSA_indexed_corrected | -2.190456463 | 0.3101365895 | -0.1032016499 | 1.12E-11 | 0.3217257514 | -0.1032016499 | 1.12E-11 | 4308 | 4.03E-10 | TRUE | 1.64E-07 | TRUE | 1.39E-07 | TRUE | FALSE | -2.190456463 |
| **ggt1** | LV end systolic volume \| Instance 2(participant - p24101_i2)_BSA_indexed_corrected | -0.9276181359 | 0.3001148208 | -0.07523643146 | 7.66E-07 | 0.1873576245 | -0.07523643146 | 7.66E-07 | 4308 | 1.12E-05 | TRUE | 0.01121185356 | TRUE | 0.009482985743 | TRUE | FALSE | -0.9276181359 |
| **ggt1** | RV end diastolic volume \| Instance 2_BSA_indexed_corrected | -2.354672542 | 0.2283607811 | -0.103077306 | 1.19E-11 | 0.3462668597 | -0.103077306 | 1.19E-11 | 4308 | 4.24E-10 | TRUE | 1.74E-07 | TRUE | 1.47E-07 | TRUE | FALSE | -2.354672542 |
| **ggt1** | RV end systolic volume \| Instance 2_BSA_indexed_corrected | -1.185598629 | 0.2083779983 | -0.08731656046 | 9.45E-09 | 0.2061304458 | -0.08731656046 | 9.45E-09 | 4308 | 1.94E-07 | TRUE | 0.0001382205495 | TRUE | 0.0001169069407 | TRUE | FALSE | -1.185598629 |

| | | | | | | | | | | | | | | | | | |
|---|---|---|---|---|---|---|---|---|---|---|---|---|---|---|---|---|---|
| **gh1** | LV end diastolic volume \| Instance 2(participant - p24100_i2)_BSA_indexed_corrected | 0.404581576 | 0.2427475168 | 0.07305662299 | 1.32E-06 | 0.08353036026 | 0.07305662299 | 1.32E-06 | 4374 | 1.84E-05 | TRUE | 0.01930641637 | TRUE | 0.01632936696 | TRUE | FALSE | 0.404581576 |
| **gh1** | RV end diastolic volume \| Instance 2_BSA_indexed_corrected | 0.4859939962 | 0.1099292903 | 0.08156720224 | 6.59E-08 | 0.0898102092 | 0.08156720224 | 6.59E-08 | 4374 | 1.17E-06 | TRUE | 0.0009641043094 | TRUE | 0.0008154394249 | TRUE | FALSE | 0.4859939962 |
| **glb1** | LV end diastolic volume \| Instance 2(participant - p24100_i2)_BSA_indexed_corrected | -1.764392363 | 0.3254748028 | -0.0701314382 | 3.43E-06 | 0.3795089561 | -0.0701314382 | 3.43E-06 | 4375 | 4.43E-05 | TRUE | 0.05020079332 | FALSE | 0.04245983099 | TRUE | FALSE | -1.764392363 |
| **glb1** | RV end diastolic volume \| Instance 2_BSA_indexed_corrected | -1.994795563 | 0.2123058395 | -0.07370014008 | 1.06E-06 | 0.408185568 | -0.07370014008 | 1.06E-06 | 4375 | 1.51E-05 | TRUE | 0.01551638619 | TRUE | 0.01312375944 | TRUE | FALSE | -1.994795563 |
| **golm2** | LV end diastolic volume \| Instance 2(participant - p24100_i2)_BSA_indexed_corrected | -3.860726322 | 0.1782775402 | -0.0795518557 | 1.43E-07 | 0.7325665356 | -0.0795518557 | 1.43E-07 | 4363 | 2.39E-06 | TRUE | 0.002090705348 | TRUE | 0.001768318584 | TRUE | FALSE | -3.860726322 |
| **golm2** | RV end diastolic volume \| Instance 2_BSA_indexed_corrected | -4.746719233 | 0.02266965448 | -0.09084366953 | 1.84E-09 | 0.787963995 | -0.09084366953 | 1.84E-09 | 4363 | 4.26E-08 | TRUE | 2.69E-05 | TRUE | 2.28E-05 | TRUE | FALSE | -4.746719233 |
| **gpkow** | LV end diastolic volume \| Instance 2(participant - p24100_i2)_BSA_indexed_corrected | -4.294529853 | 0.2571745889 | -0.1078352127 | 1.56E-12 | 0.6056876272 | -0.1078352127 | 1.56E-12 | 4275 | 6.48E-11 | TRUE | 2.28E-08 | TRUE | 1.93E-08 | TRUE | FALSE | -4.294529853 |
| **gpkow** | LV end systolic volume \| Instance 2(participant - p24101_i2)_BSA_indexed_corrected | -1.98261038 | 0.2523578203 | -0.0859316099 | 1.83E-08 | 0.3516483337 | -0.0859316099 | 1.83E-08 | 4275 | 3.61E-07 | TRUE | 0.0002678427086 | TRUE | 0.000226541363 | TRUE | FALSE | -1.98261038 |
| **gpkow** | RV end diastolic volume \| Instance 2_BSA_indexed_corrected | -3.97569382 | 0.1457623499 | -0.09283896193 | 1.19E-09 | 0.6522836433 | -0.09283896193 | 1.19E-09 | 4275 | 2.90E-08 | TRUE | 1.74E-05 | TRUE | 1.47E-05 | TRUE | FALSE | -3.97569382 |
| **gpkow** | RV end systolic volume \| Instance 2_BSA_indexed_corrected | -1.852103028 | 0.1603810668 | -0.07285251347 | 1.86E-06 | 0.3878810653 | -0.07285251347 | 1.86E-06 | 4275 | 2.52E-05 | TRUE | 0.02718130454 | TRUE | 0.02298994738 | TRUE | FALSE | -1.852103028 |
| **gpr37** | LV end diastolic volume \| Instance 2(participant - p24100_i2)_BSA_indexed_corrected | -1.82031338 | 0.1037794492 | -0.1057947203 | 2.29E-12 | 0.2587308999 | -0.1057947203 | 2.29E-12 | 4375 | 9.32E-11 | TRUE | 3.36E-08 | TRUE | 2.84E-08 | TRUE | FALSE | -1.82031338 |
| **gpr37** | LV end systolic volume \| Instance 2(participant - p24101_i2)_BSA_indexed_corrected | -0.8397502649 | 0.1969339459 | -0.08407695867 | 2.56E-08 | 0.1505022409 | -0.08407695867 | 2.56E-08 | 4375 | 4.92E-07 | TRUE | 0.0003738789853 | TRUE | 0.0003162268458 | TRUE | FALSE | -0.8397502649 |
| **gpr37** | RV end diastolic volume \| Instance 2_BSA_indexed_corrected | -1.673115971 | 0.01436096537 | -0.0903849708 | 2.11E-09 | 0.2787782106 | -0.0903849708 | 2.11E-09 | 4375 | 4.83E-08 | TRUE | 3.09E-05 | TRUE | 2.61E-05 | TRUE | FALSE | -1.673115971 |
| **gsta1** | LV end diastolic volume \| Instance 2(participant - p24100_i2)_BSA_indexed_corrected | -1.835039678 | 0.2745483771 | -0.1508126628 | 1.13E-23 | 0.1818955118 | -0.1508126628 | 1.13E-23 | 4375 | 2.79E-21 | TRUE | 1.65E-19 | TRUE | 1.39E-19 | TRUE | FALSE | -1.835039678 |
| **gsta1** | LV end systolic volume \| Instance 2(participant - p24101_i2)_BSA_indexed_corrected | -0.8117072643 | 0.277202996 | -0.1149213694 | 2.45E-14 | 0.1061016039 | -0.1149213694 | 2.45E-14 | 4375 | 1.42E-12 | TRUE | 3.59E-10 | TRUE | 3.03E-10 | TRUE | FALSE | -0.8117072643 |
| **gsta1** | RV end diastolic volume \| Instance 2_BSA_indexed_corrected | -1.887467525 | 0.1627339341 | -0.1441863609 | 9.30E-22 | 0.1958860701 | -0.1441863609 | 9.30E-22 | 4375 | 1.70E-19 | TRUE | 1.36E-17 | TRUE | 1.15E-17 | TRUE | FALSE | -1.887467525 |
| **gsta1** | RV end systolic volume \| Instance 2_BSA_indexed_corrected | -0.8904463423 | 0.1790457115 | -0.114383807 | 3.24E-14 | 0.1169482263 | -0.114383807 | 3.24E-14 | 4375 | 1.81E-12 | TRUE | 4.73E-10 | TRUE | 4.00E-10 | TRUE | FALSE | -0.8904463423 |
| **gsta3** | LV end diastolic volume \| Instance 2(participant - p24100_i2)_BSA_indexed_corrected | -2.1277873 | 0.4236973697 | -0.1518399702 | 5.58E-24 | 0.2094533629 | -0.1518399702 | 5.58E-24 | 4375 | 1.51E-21 | TRUE | 8.16E-20 | TRUE | 6.91E-20 | TRUE | FALSE | -2.1277873 |
| **gsta3** | LV end systolic volume \| Instance 2(participant - p24101_i2)_BSA_indexed_corrected | -0.9996216676 | 0.3451178929 | -0.1228860457 | 3.44E-16 | 0.122078506 | -0.1228860457 | 3.44E-16 | 4375 | 2.74E-14 | TRUE | 5.03E-12 | TRUE | 4.26E-12 | TRUE | FALSE | -0.9996216676 |
| **gsta3** | RV end diastolic volume \| Instance 2_BSA_indexed_corrected | -2.129599305 | 0.3141849453 | -0.1412564013 | 6.13E-21 | 0.2256957544 | -0.1412564013 | 6.13E-21 | 4375 | 9.97E-19 | TRUE | 8.97E-17 | TRUE | 7.59E-17 | TRUE | FALSE | -2.129599305 |
| **gsta3** | RV end systolic volume \| Instance 2_BSA_indexed_corrected | -0.9856513618 | 0.2498634333 | -0.1099374521 | 3.06E-13 | 0.1347557907 | -0.1099374521 | 3.06E-13 | 4375 | 1.45E-11 | TRUE | 4.48E-09 | TRUE | 3.79E-09 | TRUE | FALSE | -0.9856513618 |
| **gusb** | LV end diastolic volume \| Instance 2(participant - p24100_i2)_BSA_indexed_corrected | -2.668259573 | 0.1729779625 | -0.1567566036 | 1.81E-25 | 0.2542201441 | -0.1567566036 | 1.81E-25 | 4375 | 5.88E-23 | TRUE | 2.64E-21 | TRUE | 2.24E-21 | TRUE | FALSE | -2.668259573 |
| **gusb** | LV end systolic volume \| Instance 2(participant - p24101_i2)_BSA_indexed_corrected | -1.192532277 | 0.2314473154 | -0.1206915659 | 1.15E-15 | 0.1483259549 | -0.1206915659 | 1.15E-15 | 4375 | 8.43E-14 | TRUE | 1.68E-11 | TRUE | 1.42E-11 | TRUE | FALSE | -1.192532277 |
| **gusb** | RV end diastolic volume \| Instance 2_BSA_indexed_corrected | -2.655437241 | 0.06427051471 | -0.1450060607 | 5.45E-22 | 0.2739968432 | -0.1450060607 | 5.45E-22 | 4375 | 1.01E-19 | TRUE | 7.97E-18 | TRUE | 6.74E-18 | TRUE | FALSE | -2.655437241 |
| **gusb** | RV end systolic volume \| Instance 2_BSA_indexed_corrected | -1.094621373 | 0.1432633332 | -0.1005139029 | 2.68E-11 | 0.1638487162 | -0.1005139029 | 2.68E-11 | 4375 | 8.96E-10 | TRUE | 3.92E-07 | TRUE | 3.31E-07 | TRUE | FALSE | -1.094621373 |
| **gzma** | LV end diastolic volume \| Instance 2(participant - p24100_i2)_BSA_indexed_corrected | -2.423572225 | 0.2894064892 | -0.07815724102 | 2.37E-07 | 0.4682338299 | -0.07815724102 | 2.37E-07 | 4361 | 3.82E-06 | TRUE | 0.00346600865 | TRUE | 0.002931550116 | TRUE | FALSE | -2.423572225 |
| **gzma** | RV end diastolic volume \| Instance 2_BSA_indexed_corrected | -2.845889175 | 0.1686826658 | -0.08528882502 | 1.69E-08 | 0.5035551893 | -0.08528882502 | 1.69E-08 | 4361 | 3.36E-07 | TRUE | 0.0002474030544 | TRUE | 0.0002092535035 | TRUE | FALSE | -2.845889175 |
| **gzma** | RV end systolic volume \| Instance 2_BSA_indexed_corrected | -1.498327838 | 0.1742337555 | -0.07549428952 | 6.00E-07 | 0.2997492913 | -0.07549428952 | 6.00E-07 | 4361 | 8.96E-06 | TRUE | 0.008779852277 | TRUE | 0.007425999056 | TRUE | FALSE | -1.498327838 |
| **hao1** | LV end diastolic volume \| Instance 2(participant - p24100_i2)_BSA_indexed_corrected | -0.7787581819 | 0.3442180417 | -0.09428349624 | 4.15E-10 | 0.1243478471 | -0.09428349624 | 4.15E-10 | 4375 | 1.12E-08 | TRUE | 6.07E-06 | TRUE | 5.13E-06 | TRUE | FALSE | -0.7787581819 |
| **hao1** | RV end diastolic volume \| Instance 2_BSA_indexed_corrected | -0.8824667245 | 0.2334739094 | -0.09930788635 | 4.61E-11 | 0.1337127168 | -0.09930788635 | 4.61E-11 | 4375 | 1.48E-09 | TRUE | 6.75E-07 | TRUE | 5.71E-07 | TRUE | FALSE | -0.8824667245 |
| **hao1** | RV end systolic volume \| Instance 2_BSA_indexed_corrected | -0.4439759966 | 0.2121061361 | -0.08401503079 | 2.62E-08 | 0.07962960966 | -0.08401503079 | 2.62E-08 | 4375 | 5.02E-07 | TRUE | 0.0003828143088 | TRUE | 0.0003237843423 | TRUE | FALSE | -0.4439759966 |
| **havcr1** | LV end diastolic volume \| Instance 2(participant - p24100_i2)_BSA_indexed_corrected | -1.403819306 | 0.1107439058 | -0.08002752132 | 1.16E-07 | 0.2644760017 | -0.08002752132 | 1.16E-07 | 4373 | 1.98E-06 | TRUE | 0.001702560094 | TRUE | 0.001440025328 | TRUE | FALSE | -1.403819306 |
| **havcr1** | RV end diastolic volume \| Instance 2_BSA_indexed_corrected | -1.829173401 | -0.07123048909 | -0.09693211332 | 1.33E-10 | 0.2840840612 | -0.09693211332 | 1.33E-10 | 4373 | 3.89E-09 | TRUE | 1.95E-06 | TRUE | 1.65E-06 | TRUE | FALSE | -1.829173401 |
| **havcr1** | RV end systolic volume \| Instance 2_BSA_indexed_corrected | -0.8992231405 | 0.06451236811 | -0.08011693235 | 1.13E-07 | 0.1692210786 | -0.08011693235 | 1.13E-07 | 4373 | 1.93E-06 | TRUE | 0.001648044377 | TRUE | 0.001393915934 | TRUE | FALSE | -0.8992231405 |
| **havcr2** | LV end diastolic volume \| Instance 2(participant - p24100_i2)_BSA_indexed_corrected | -2.467710728 | 0.1238072423 | -0.07401140335 | 1.15E-06 | 0.5065410294 | -0.07401140335 | 1.15E-06 | 4311 | 1.61E-05 | TRUE | 0.01676880363 | TRUE | 0.01418305411 | TRUE | FALSE | -2.467710728 |
| **havcr2** | RV end diastolic volume \| Instance 2_BSA_indexed_corrected | -3.581832056 | -0.05445031407 | -0.09979201242 | 5.15E-11 | 0.544061385 | -0.09979201242 | 5.15E-11 | 4311 | 1.63E-09 | TRUE | 7.53E-07 | TRUE | 6.37E-07 | TRUE | FALSE | -3.581832056 |
| **havcr2** | RV end systolic volume \| Instance 2_BSA_indexed_corrected | -1.585192597 | 0.08568020303 | -0.07427289881 | 1.05E-06 | 0.3242367317 | -0.07427289881 | 1.05E-06 | 4311 | 1.49E-05 | TRUE | 0.01536724995 | TRUE | 0.01299762001 | TRUE | FALSE | -1.585192597 |
| **hgf** | LV end diastolic volume \| Instance 2(participant - p24100_i2)_BSA_indexed_corrected | -5.637929822 | -0.1878992814 | -0.1812312303 | 1.62E-33 | 0.4633842701 | -0.1812312303 | 1.62E-33 | 4361 | 1.32E-30 | TRUE | 2.37E-29 | TRUE | 2.01E-29 | TRUE | FALSE | -5.637929822 |
| **hgf** | LV end systolic volume \| Instance 2(participant - p24101_i2)_BSA_indexed_corrected | -2.73714058 | 0.04731742767 | -0.1514632551 | 8.51E-24 | 0.2705557397 | -0.1514632551 | 8.51E-24 | 4361 | 2.22E-21 | TRUE | 1.25E-19 | TRUE | 1.05E-19 | TRUE | FALSE | -2.73714058 |
| **hgf** | RV end diastolic volume \| Instance 2_BSA_indexed_corrected | -6.348644527 | -0.3648508646 | -0.1896510615 | 1.35E-36 | 0.4978266368 | -0.1896510615 | 1.35E-36 | 4361 | 2.19E-33 | TRUE | 1.97E-32 | TRUE | 1.67E-32 | TRUE | FALSE | -6.348644527 |
| **hgf** | RV end systolic volume \| Instance 2_BSA_indexed_corrected | -3.019119654 | -0.07459882827 | -0.1516308951 | 7.59E-24 | 0.2980905909 | -0.1516308951 | 7.59E-24 | 4361 | 2.02E-21 | TRUE | 1.11E-19 | TRUE | 9.39E-20 | TRUE | FALSE | -3.019119654 |
| **hnmt** | LV end diastolic volume \| Instance 2(participant - p24100_i2)_BSA_indexed_corrected | -3.290270558 | -0.02173797204 | -0.140965235 | 8.17E-21 | 0.3498392724 | -0.140965235 | 8.17E-21 | 4365 | 1.30E-18 | TRUE | 1.20E-16 | TRUE | 1.01E-16 | TRUE | FALSE | -3.290270558 |
| **hnmt** | LV end systolic volume \| Instance 2(participant - p24101_i2)_BSA_indexed_corrected | -1.361436849 | 0.1621106141 | -0.1005239527 | 2.81E-11 | 0.203999869 | -0.1005239527 | 2.81E-11 | 4365 | 9.37E-10 | TRUE | 4.11E-07 | TRUE | 3.47E-07 | TRUE | FALSE | -1.361436849 |
| **hnmt** | RV end diastolic volume \| Instance 2_BSA_indexed_corrected | -3.597628322 | -0.1687423811 | -0.1432537691 | 1.89E-21 | 0.3762833935 | -0.1432537691 | 1.89E-21 | 4365 | 3.29E-19 | TRUE | 2.77E-17 | TRUE | 2.34E-17 | TRUE | FALSE | -3.597628322 |
| **hnmt** | RV end systolic volume \| Instance 2_BSA_indexed_corrected | -1.833683152 | 0.005338391119 | -0.1228387109 | 3.82E-16 | 0.2242822245 | -0.1228387109 | 3.82E-16 | 4365 | 2.99E-14 | TRUE | 5.58E-12 | TRUE | 4.72E-12 | TRUE | FALSE | -1.833683152 |
| **hsd11b1** | LV end diastolic volume \| Instance 2(participant - p24100_i2)_BSA_indexed_corrected | 3.510055069 | 0.269283039 | 0.1240448887 | 2.53E-16 | 0.4267011972 | 0.1240448887 | 2.53E-16 | 4332 | 2.07E-14 | TRUE | 3.71E-12 | TRUE | 3.13E-12 | TRUE | FALSE | 3.510055069 |
| **hsd11b1** | LV end systolic volume \| Instance 2(participant - p24101_i2)_BSA_indexed_corrected | 1.68659411 | 0.2745313167 | 0.1027510637 | 1.21E-11 | 0.2481278564 | 0.1027510637 | 1.21E-11 | 4332 | 4.30E-10 | TRUE | 1.77E-07 | TRUE | 1.50E-07 | TRUE | FALSE | 1.68659411 |
| **hsd11b1** | RV end diastolic volume \| Instance 2_BSA_indexed_corrected | 3.755533591 | 0.1583387876 | 0.1233852763 | 3.65E-16 | 0.4590215858 | 0.1233852763 | 3.65E-16 | 4332 | 2.88E-14 | TRUE | 5.33E-12 | TRUE | 4.51E-12 | TRUE | FALSE | 3.755533591 |
| **hsd11b1** | RV end systolic volume \| Instance 2_BSA_indexed_corrected | 1.687306995 | 0.1822391778 | 0.09312029546 | 8.22E-10 | 0.2741666552 | 0.09312029546 | 8.22E-10 | 4332 | 2.08E-08 | TRUE | 1.20E-05 | TRUE | 1.02E-05 | TRUE | FALSE | 1.687306995 |
| **hspb6** | LV end diastolic volume \| Instance 2(participant - p24100_i2)_BSA_indexed_corrected | -1.929419193 | 0.2025856005 | -0.07595811806 | 4.91E-07 | 0.3830065232 | -0.07595811806 | 4.91E-07 | 4375 | 7.46E-06 | TRUE | 0.007176120979 | TRUE | 0.006069563124 | TRUE | FALSE | -1.929419193 |
| **hspb6** | RV end diastolic volume \| Instance 2_BSA_indexed_corrected | -2.062075774 | 0.08267025487 | -0.07545787416 | 5.83E-07 | 0.4120694038 | -0.07545787416 | 5.83E-07 | 4375 | 8.73E-06 | TRUE | 0.008529173357 | TRUE | 0.007213974825 | TRUE | FALSE | -2.062075774 |
| **hspg2** | LV cardiac output \| Instance 2_BSA_indexed_corrected | -0.1549231906 | -0.02603954287 | -0.07571664342 | 6.71E-07 | 0.03112751517 | -0.07571664342 | 6.71E-07 | 4298 | 9.89E-06 | TRUE | 0.00981500846 | TRUE | 0.008301534155 | TRUE | FALSE | -0.1549231906 |
| **hspg2** | LV end diastolic volume \| Instance 2(participant - p24100_i2)_BSA_indexed_corrected | -5.198005614 | 0.151785358 | -0.1132461657 | 9.61E-14 | 0.6957900962 | -0.1132461657 | 9.61E-14 | 4298 | 4.92E-12 | TRUE | 1.41E-09 | TRUE | 1.19E-09 | TRUE | FALSE | -5.198005614 |
| **hspg2** | LV end systolic volume \| Instance 2(participant - p24101_i2)_BSA_indexed_corrected | -2.277002279 | 0.2250561593 | -0.08541727833 | 2.04E-08 | 0.4052242819 | -0.08541727833 | 2.04E-08 | 4298 | 3.99E-07 | TRUE | 0.000298569886 | TRUE | 0.0002525304096 | TRUE | FALSE | -2.277002279 |
| **hspg2** | RV end diastolic volume \| Instance 2_BSA_indexed_corrected | -5.70017165 | 0.003576388461 | -0.1154976243 | 3.07E-14 | 0.7479396781 | -0.1154976243 | 3.07E-14 | 4298 | 1.73E-12 | TRUE | 4.50E-10 | TRUE | 3.80E-10 | TRUE | FALSE | -5.70017165 |
| **hspg2** | RV end systolic volume \| Instance 2_BSA_indexed_corrected | -2.616634111 | 0.128524888 | -0.08919581593 | 4.70E-09 | 0.4457915892 | -0.08919581593 | 4.70E-09 | 4298 | 1.01E-07 | TRUE | 6.87E-05 | TRUE | 5.81E-05 | TRUE | FALSE | -2.616634111 |
| **icam1** | LV end diastolic volume \| Instance 2(participant - p24100_i2)_BSA_indexed_corrected | -3.794041553 | 0.1562200772 | -0.09538874248 | 3.55E-10 | 0.6034391908 | -0.09538874248 | 3.55E-10 | 4307 | 9.64E-09 | TRUE | 5.19E-06 | TRUE | 4.39E-06 | TRUE | FALSE | -3.794041553 |
| **icam1** | RV end diastolic volume \| Instance 2_BSA_indexed_corrected | -4.926824171 | -0.04403211051 | -0.1152323102 | 3.31E-14 | 0.6472968829 | -0.1152323102 | 3.31E-14 | 4307 | 1.84E-12 | TRUE | 4.84E-10 | TRUE | 4.10E-10 | TRUE | FALSE | -4.926824171 |
| **icam1** | RV end systolic volume \| Instance 2_BSA_indexed_corrected | -1.985870645 | 0.1179038479 | -0.07816514276 | 2.80E-07 | 0.386029469 | -0.07816514276 | 2.80E-07 | 4307 | 4.46E-06 | TRUE | 0.004102122743 | TRUE | 0.003469575416 | TRUE | FALSE | -1.985870645 |
| **idua** | LV end diastolic volume \| Instance 2(participant - p24100_i2)_BSA_indexed_corrected | -2.165592265 | 0.1401447774 | -0.09298315931 | 7.19E-10 | 0.35066875 | -0.09298315931 | 7.19E-10 | 4375 | 1.84E-08 | TRUE | 1.05E-05 | TRUE | 8.90E-06 | TRUE | FALSE | -2.165592265 |
| **idua** | LV end systolic volume \| Instance 2(participant - p24101_i2)_BSA_indexed_corrected | -1.202083462 | 0.1937510919 | -0.08891422345 | 3.84E-09 | 0.2036339158 | -0.08891422345 | 3.84E-09 | 4375 | 8.43E-08 | TRUE | 5.61E-05 | TRUE | 4.75E-05 | TRUE | FALSE | -1.202083462 |
| **idua** | RV end diastolic volume \| Instance 2_BSA_indexed_corrected | -2.373599628 | 0.0101257499 | -0.09472997353 | 3.43E-10 | 0.3772010437 | -0.09472997353 | 3.43E-10 | 4375 | 9.35E-09 | TRUE | 5.01E-06 | TRUE | 4.24E-06 | TRUE | FALSE | -2.373599628 |
| **idua** | RV end systolic volume \| Instance 2_BSA_indexed_corrected | -1.050130085 | 0.1138972036 | -0.0704750108 | 3.07E-06 | 0.22476931 | -0.0704750108 | 3.07E-06 | 4375 | 4.00E-05 | TRUE | 0.0449416809 | TRUE | 0.0380116737 | TRUE | FALSE | -1.050130085 |
| **ifngr2** | LV end diastolic volume \| Instance 2(participant - p24100_i2)_BSA_indexed_corrected | -1.899942809 | 0.5181267261 | -0.08816020498 | 5.19E-09 | 0.3246264399 | -0.08816020498 | 5.19E-09 | 4375 | 1.11E-07 | TRUE | 7.59E-05 | TRUE | 6.42E-05 | TRUE | FALSE | -1.899942809 |
| **ifngr2** | RV end diastolic volume \| Instance 2_BSA_indexed_corrected | -1.692293644 | 0.3905077821 | -0.07298945659 | 1.35E-06 | 0.3496760747 | -0.07298945659 | 1.35E-06 | 4375 | 1.87E-05 | TRUE | 0.01968924684 | TRUE | 0.01665316498 | TRUE | FALSE | -1.692293644 |
| **ifnlr1** | LV end diastolic volume \| Instance 2(participant - p24100_i2)_BSA_indexed_corrected | -2.353770186 | 0.299688176 | -0.08657987322 | 1.15E-08 | 0.4115942324 | -0.08657987322 | 1.15E-08 | 4332 | 2.34E-07 | TRUE | 0.0001676473555 | TRUE | 0.0001417961332 | TRUE | FALSE | -2.353770186 |
| **ifnlr1** | LV end systolic volume \| Instance 2(participant - p24101_i2)_BSA_indexed_corrected | -1.190182682 | 0.2874513118 | -0.07547051576 | 6.59E-07 | 0.23897474 | -0.07547051576 | 6.59E-07 | 4332 | 9.74E-06 | TRUE | 0.009646128546 | TRUE | 0.008158695524 | TRUE | FALSE | -1.190182682 |
| **ifnlr1** | RV end diastolic volume \| Instance 2_BSA_indexed_corrected | -2.705552706 | 0.1855267127 | -0.09252000127 | 1.06E-09 | 0.4424963061 | -0.09252000127 | 1.06E-09 | 4332 | 2.60E-08 | TRUE | 1.54E-05 | TRUE | 1.31E-05 | TRUE | FALSE | -2.705552706 |

| | | | | | | | | | | | | | | | | | |
|---|---|---|---|---|---|---|---|---|---|---|---|---|---|---|---|---|---|
| **igf2r** | LV end diastolic volume \| Instance 2(participant - p24100_i2)_BSA_indexed_corrected | -3.59376934 | 0.2139229148 | -0.07612149568 | 5.62E-07 | 0.7171214679 | -0.07612149568 | 5.62E-07 | 4311 | 8.44E-06 | TRUE | 0.008223147138 | TRUE | 0.006955137849 | TRUE | FALSE | -3.59376934 |
| **igf2r** | RV end diastolic volume \| Instance 2_BSA_indexed_corrected | -3.767450752 | 0.1292356474 | -0.07412946567 | 1.10E-06 | 0.7720970869 | -0.07412946567 | 1.10E-06 | 4311 | 1.56E-05 | TRUE | 0.01612142912 | TRUE | 0.01363550475 | TRUE | FALSE | -3.767450752 |
| **igfbp1** | LV end diastolic volume \| Instance 2(participant - p24100_i2)_BSA_indexed_corrected | 1.251396699 | 0.365206677 | 0.1533829127 | 2.07E-24 | 0.1219991205 | 0.1533829127 | 2.07E-24 | 4369 | 6.18E-22 | TRUE | 3.03E-20 | TRUE | 2.56E-20 | TRUE | FALSE | 1.251396699 |
| **igfbp1** | LV end systolic volume \| Instance 2(participant - p24101_i2)_BSA_indexed_corrected | 0.5524001755 | 0.3183239526 | 0.1165936848 | 1.07E-14 | 0.0712057422 | 0.1165936848 | 1.07E-14 | 4369 | 6.46E-13 | TRUE | 1.56E-10 | TRUE | 1.32E-10 | TRUE | FALSE | 0.5524001755 |
| **igfbp1** | RV end diastolic volume \| Instance 2_BSA_indexed_corrected | 1.303619245 | 0.2600920365 | 0.1485020247 | 5.75E-23 | 0.1313664563 | 0.1485020247 | 5.75E-23 | 4369 | 1.22E-20 | TRUE | 8.41E-19 | TRUE | 7.11E-19 | TRUE | FALSE | 1.303619245 |
| **igfbp1** | RV end systolic volume \| Instance 2_BSA_indexed_corrected | 0.5506517938 | 0.2267260542 | 0.1054775141 | 2.76E-12 | 0.07855903937 | 0.1054775141 | 2.76E-12 | 4369 | 1.11E-10 | TRUE | 4.04E-08 | TRUE | 3.42E-08 | TRUE | FALSE | 0.5506517938 |
| **igfbp2** | LV end diastolic volume \| Instance 2(participant - p24100_i2)_BSA_indexed_corrected | 2.977104467 | 0.5946447124 | 0.1853416114 | 4.69E-35 | 0.2388849591 | 0.1853416114 | 4.69E-35 | 4368 | 5.72E-32 | TRUE | 6.86E-31 | TRUE | 5.80E-31 | TRUE | FALSE | 2.977104467 |
| **igfbp2** | LV end systolic volume \| Instance 2(participant - p24101_i2)_BSA_indexed_corrected | 1.371566644 | 0.4177301537 | 0.1470695851 | 1.51E-22 | 0.1396059295 | 0.1470695851 | 1.51E-22 | 4368 | 2.98E-20 | TRUE | 2.21E-18 | TRUE | 1.87E-18 | TRUE | FALSE | 1.371566644 |
| **igfbp2** | RV end diastolic volume \| Instance 2_BSA_indexed_corrected | 3.136130488 | 0.499356737 | 0.1815191245 | 1.14E-33 | 0.257131046 | 0.1815191245 | 1.14E-33 | 4368 | 9.79E-31 | TRUE | 1.66E-29 | TRUE | 1.41E-29 | TRUE | FALSE | 3.136130488 |
| **igfbp2** | RV end systolic volume \| Instance 2_BSA_indexed_corrected | 1.5381859 | 0.3413684182 | 0.149721577 | 2.56E-23 | 0.1537303045 | 0.149721577 | 2.56E-23 | 4368 | 6.04E-21 | TRUE | 3.74E-19 | TRUE | 3.17E-19 | TRUE | FALSE | 1.5381859 |
| **igfbp4** | LV end diastolic volume \| Instance 2(participant - p24100_i2)_BSA_indexed_corrected | -2.395438021 | 0.08325510293 | -0.08126046228 | 1.08E-07 | 0.4500495085 | -0.08126046228 | 1.08E-07 | 4264 | 1.85E-06 | TRUE | 0.001573430227 | TRUE | 0.001330807286 | TRUE | FALSE | -2.395438021 |
| **igfbp4** | RV end diastolic volume \| Instance 2_BSA_indexed_corrected | -2.97454129 | -0.05645150551 | -0.09377261934 | 8.52E-10 | 0.4837481814 | -0.09377261934 | 8.52E-10 | 4264 | 2.13E-08 | TRUE | 1.25E-05 | TRUE | 1.05E-05 | TRUE | FALSE | -2.97454129 |
| **igfbp6** | LV cardiac output \| Instance 2_BSA_indexed_corrected | -0.1777936652 | -0.0234368277 | -0.1095942493 | 5.44E-13 | 0.02457067897 | -0.1095942493 | 5.44E-13 | 4309 | 2.48E-11 | TRUE | 7.96E-09 | TRUE | 6.74E-09 | TRUE | FALSE | -0.1777936652 |
| **igfbp6** | LV end diastolic volume \| Instance 2(participant - p24100_i2)_BSA_indexed_corrected | -4.003642527 | 0.2976755946 | -0.1098697568 | 4.76E-13 | 0.5518899452 | -0.1098697568 | 4.76E-13 | 4309 | 2.21E-11 | TRUE | 6.96E-09 | TRUE | 5.89E-09 | TRUE | FALSE | -4.003642527 |
| **igfbp6** | LV end systolic volume \| Instance 2(participant - p24101_i2)_BSA_indexed_corrected | -1.494918556 | 0.2950631288 | -0.07059915338 | 3.50E-06 | 0.3218441232 | -0.07059915338 | 3.50E-06 | 4309 | 4.53E-05 | TRUE | 0.05127340266 | FALSE | 0.04336704397 | TRUE | FALSE | -1.494918556 |
| **igfbp6** | RV end diastolic volume \| Instance 2_BSA_indexed_corrected | -4.067904311 | 0.1694941616 | -0.1038681074 | 8.22E-12 | 0.5935340438 | -0.1038681074 | 8.22E-12 | 4309 | 3.04E-10 | TRUE | 1.20E-07 | TRUE | 1.02E-07 | TRUE | FALSE | -4.067904311 |
| **igsf3** | LV end diastolic volume \| Instance 2(participant - p24100_i2)_BSA_indexed_corrected | -3.523846913 | 0.2255640937 | -0.1117379322 | 1.25E-13 | 0.4739127954 | -0.1117379322 | 1.25E-13 | 4375 | 6.24E-12 | TRUE | 1.82E-09 | TRUE | 1.54E-09 | TRUE | FALSE | -3.523846913 |
| **igsf3** | LV end systolic volume \| Instance 2(participant - p24101_i2)_BSA_indexed_corrected | -1.318377121 | 0.2642285303 | -0.07201654223 | 1.86E-06 | 0.2761140552 | -0.07201654223 | 1.86E-06 | 4375 | 2.52E-05 | TRUE | 0.02718368655 | TRUE | 0.02299196208 | TRUE | FALSE | -1.318377121 |
| **igsf3** | RV end diastolic volume \| Instance 2_BSA_indexed_corrected | -3.637426087 | 0.1118835297 | -0.1072087218 | 1.16E-12 | 0.5101102367 | -0.1072087218 | 1.16E-12 | 4375 | 4.89E-11 | TRUE | 1.70E-08 | TRUE | 1.44E-08 | TRUE | FALSE | -3.637426087 |
| **igsf8** | LV end diastolic volume \| Instance 2(participant - p24100_i2)_BSA_indexed_corrected | -3.826029492 | 0.1331670012 | -0.09846693539 | 6.71E-11 | 0.5847260972 | -0.09846693539 | 6.71E-11 | 4375 | 2.07E-09 | TRUE | 9.82E-07 | TRUE | 8.30E-07 | TRUE | FALSE | -3.826029492 |
| **igsf8** | LV end systolic volume \| Instance 2(participant - p24101_i2)_BSA_indexed_corrected | -1.584194519 | 0.2208821292 | -0.07023597655 | 3.32E-06 | 0.340239933 | -0.07023597655 | 3.32E-06 | 4375 | 4.30E-05 | TRUE | 0.04854117172 | TRUE | 0.04105612304 | TRUE | FALSE | -1.584194519 |
| **igsf8** | RV end diastolic volume \| Instance 2_BSA_indexed_corrected | -4.061714568 | 0.01005164783 | -0.09716365712 | 1.19E-10 | 0.6291526531 | -0.09716365712 | 1.19E-10 | 4375 | 3.51E-09 | TRUE | 1.75E-06 | TRUE | 1.48E-06 | TRUE | FALSE | -4.061714568 |
| **il10rb** | LV end diastolic volume \| Instance 2(participant - p24100_i2)_BSA_indexed_corrected | -3.91476085 | 0.1345299896 | -0.09462843315 | 4.57E-10 | 0.6265972753 | -0.09462843315 | 4.57E-10 | 4322 | 1.22E-08 | TRUE | 6.68E-06 | TRUE | 5.65E-06 | TRUE | FALSE | -3.91476085 |
| **il10rb** | RV end diastolic volume \| Instance 2_BSA_indexed_corrected | -4.821087073 | -0.02716000976 | -0.1083830145 | 9.05E-13 | 0.6727844939 | -0.1083830145 | 9.05E-13 | 4322 | 3.94E-11 | TRUE | 1.32E-08 | TRUE | 1.12E-08 | TRUE | FALSE | -4.821087073 |
| **il10rb** | RV end systolic volume \| Instance 2_BSA_indexed_corrected | -2.15432087 | 0.09193850237 | -0.08140273565 | 8.37E-08 | 0.4013153957 | -0.08140273565 | 8.37E-08 | 4322 | 1.47E-06 | TRUE | 0.001224842989 | TRUE | 0.0010359722 | TRUE | FALSE | -2.15432087 |
| **il12a_il12b** | RV end diastolic volume \| Instance 2_BSA_indexed_corrected | -1.209786717 | 0.1868797414 | -0.0759976145 | 4.84E-07 | 0.2400276722 | -0.0759976145 | 4.84E-07 | 4375 | 7.37E-06 | TRUE | 0.007078591999 | TRUE | 0.005987073112 | TRUE | FALSE | -1.209786717 |
| **il12b** | RV end diastolic volume \| Instance 2_BSA_indexed_corrected | -1.617300482 | 0.1709769303 | -0.07325709086 | 1.28E-06 | 0.3334871716 | -0.07325709086 | 1.28E-06 | 4361 | 1.79E-05 | TRUE | 0.01872369741 | TRUE | 0.01583650327 | TRUE | FALSE | -1.617300482 |
| **il13ra1** | LV end diastolic volume \| Instance 2(participant - p24100_i2)_BSA_indexed_corrected | -4.923643916 | 0.2754390407 | -0.09999263222 | 6.28E-11 | 0.7512575765 | -0.09999263222 | 6.28E-11 | 4255 | 1.96E-09 | TRUE | 9.18E-07 | TRUE | 7.77E-07 | TRUE | FALSE | -4.923643916 |
| **il13ra1** | LV end systolic volume \| Instance 2(participant - p24101_i2)_BSA_indexed_corrected | -2.184179464 | 0.2779706573 | -0.07632140167 | 6.22E-07 | 0.437548016 | -0.07632140167 | 6.22E-07 | 4255 | 9.23E-06 | TRUE | 0.009098799051 | TRUE | 0.007695764237 | TRUE | FALSE | -2.184179464 |
| **il13ra1** | RV end diastolic volume \| Instance 2_BSA_indexed_corrected | -4.469771565 | 0.140428518 | -0.08464452474 | 3.21E-08 | 0.8068213317 | -0.08464452474 | 3.21E-08 | 4255 | 6.05E-07 | TRUE | 0.0004693094777 | TRUE | 0.0003969419562 | TRUE | FALSE | -4.469771565 |
| **il16** | LV end diastolic volume \| Instance 2(participant - p24100_i2)_BSA_indexed_corrected | -3.303297969 | 0.0997057501 | -0.1236454007 | 2.52E-16 | 0.4015419502 | -0.1236454007 | 2.52E-16 | 4361 | 2.07E-14 | TRUE | 3.68E-12 | TRUE | 3.12E-12 | TRUE | FALSE | -3.303297969 |
| **il16** | LV end systolic volume \| Instance 2(participant - p24101_i2)_BSA_indexed_corrected | -1.474687489 | 0.1975492321 | -0.09502262446 | 3.23E-10 | 0.2339968156 | -0.09502262446 | 3.23E-10 | 4361 | 8.92E-09 | TRUE | 4.72E-06 | TRUE | 3.99E-06 | TRUE | FALSE | -1.474687489 |
| **il16** | RV end diastolic volume \| Instance 2_BSA_indexed_corrected | -3.237486578 | -0.001359552787 | -0.1126157378 | 8.75E-14 | 0.4326571806 | -0.1126157378 | 8.75E-14 | 4361 | 4.51E-12 | TRUE | 1.28E-09 | TRUE | 1.08E-09 | TRUE | FALSE | -3.237486578 |
| **il16** | RV end systolic volume \| Instance 2_BSA_indexed_corrected | -1.653110158 | 0.08893184396 | -0.09667763344 | 1.58E-10 | 0.2577764415 | -0.09667763344 | 1.58E-10 | 4361 | 4.55E-09 | TRUE | 2.31E-06 | TRUE | 1.95E-06 | TRUE | FALSE | -1.653110158 |
| **il17rb** | LV end diastolic volume \| Instance 2(participant - p24100_i2)_BSA_indexed_corrected | 1.630665794 | 0.3469222087 | 0.0791985083 | 2.13E-07 | 0.3138426844 | 0.0791985083 | 2.13E-07 | 4279 | 3.48E-06 | TRUE | 0.003120989044 | TRUE | 0.002639732533 | TRUE | FALSE | 1.630665794 |
| **il17rb** | RV end diastolic volume \| Instance 2_BSA_indexed_corrected | 1.987711898 | 0.2637379735 | 0.08957455898 | 4.37E-09 | 0.3379479397 | 0.08957455898 | 4.37E-09 | 4279 | 9.53E-08 | TRUE | 6.39E-05 | TRUE | 5.41E-05 | TRUE | FALSE | 1.987711898 |
| **il18** | LV end diastolic volume \| Instance 2(participant - p24100_i2)_BSA_indexed_corrected | -2.169196763 | 0.2020472171 | -0.09053983913 | 2.90E-09 | 0.3645841629 | -0.09053983913 | 2.90E-09 | 4285 | 6.50E-08 | TRUE | 4.24E-05 | TRUE | 3.59E-05 | TRUE | FALSE | -2.169196763 |
| **il18** | RV end diastolic volume \| Instance 2_BSA_indexed_corrected | -2.078227035 | 0.1005261251 | -0.08074685008 | 1.20E-07 | 0.3919881297 | -0.08074685008 | 1.20E-07 | 4285 | 2.04E-06 | TRUE | 0.001762454556 | TRUE | 0.001490684063 | TRUE | FALSE | -2.078227035 |
| **il18bp** | RV end diastolic volume \| Instance 2_BSA_indexed_corrected | -3.105768677 | 0.09825187077 | -0.07440464079 | 9.71E-07 | 0.6332445636 | -0.07440464079 | 9.71E-07 | 4323 | 1.39E-05 | TRUE | 0.01420443669 | TRUE | 0.01201411255 | TRUE | FALSE | -3.105768677 |
| **il18r1** | LV end diastolic volume \| Instance 2(participant - p24100_i2)_BSA_indexed_corrected | -4.490065216 | 0.05771039755 | -0.1356337855 | 2.34E-19 | 0.496774592 | -0.1356337855 | 2.34E-19 | 4361 | 2.95E-17 | TRUE | 3.42E-15 | TRUE | 2.90E-15 | TRUE | FALSE | -4.490065216 |
| **il18r1** | LV end systolic volume \| Instance 2(participant - p24101_i2)_BSA_indexed_corrected | -1.767463499 | 0.1953495596 | -0.09190994309 | 1.20E-09 | 0.2900363244 | -0.09190994309 | 1.20E-09 | 4361 | 2.91E-08 | TRUE | 1.75E-05 | TRUE | 1.48E-05 | TRUE | FALSE | -1.767463499 |
| **il18r1** | RV end diastolic volume \| Instance 2_BSA_indexed_corrected | -4.634918808 | -0.05887644582 | -0.1301122715 | 6.32E-18 | 0.5349618782 | -0.1301122715 | 6.32E-18 | 4361 | 6.17E-16 | TRUE | 9.25E-14 | TRUE | 7.82E-14 | TRUE | FALSE | -4.634918808 |
| **il18r1** | RV end systolic volume \| Instance 2_BSA_indexed_corrected | -1.611947039 | 0.1122529523 | -0.07607816048 | 4.91E-07 | 0.3199903315 | -0.07607816048 | 4.91E-07 | 4361 | 7.46E-06 | TRUE | 0.007179843622 | TRUE | 0.006072711736 | TRUE | FALSE | -1.611947039 |
| **il1rap** | LV end diastolic volume \| Instance 2(participant - p24100_i2)_BSA_indexed_corrected | 2.100827581 | 0.06379497217 | 0.08782488238 | 7.65E-09 | 0.3629971302 | 0.08782488238 | 7.65E-09 | 4311 | 1.59E-07 | TRUE | 0.0001119721342 | TRUE | 9.47E-05 | TRUE | FALSE | 2.100827581 |
| **il1rap** | LV end systolic volume \| Instance 2(participant - p24101_i2)_BSA_indexed_corrected | 1.044641278 | 0.1738050431 | 0.07517606543 | 7.75E-07 | 0.2110904901 | 0.07517606543 | 7.75E-07 | 4311 | 1.13E-05 | TRUE | 0.01134246172 | TRUE | 0.009593454122 | TRUE | FALSE | 1.044641278 |
| **il1rap** | RV end diastolic volume \| Instance 2_BSA_indexed_corrected | 2.088606102 | -0.01291613302 | 0.08110915573 | 9.67E-08 | 0.3909895494 | 0.08110915573 | 9.67E-08 | 4311 | 1.68E-06 | TRUE | 0.001415449713 | TRUE | 0.001197187367 | TRUE | FALSE | 2.088606102 |
| **il1rn** | LV end diastolic volume \| Instance 2(participant - p24100_i2)_BSA_indexed_corrected | -4.092129782 | -0.0157060179 | -0.2047676749 | 5.48E-42 | 0.2981607928 | -0.2047676749 | 5.48E-42 | 4306 | 1.34E-38 | TRUE | 8.01E-38 | TRUE | 6.78E-38 | TRUE | FALSE | -4.092129782 |
| **il1rn** | LV end systolic volume \| Instance 2(participant - p24101_i2)_BSA_indexed_corrected | -1.723857325 | 0.160928206 | -0.1487245025 | 1.01E-22 | 0.1747131812 | -0.1487245025 | 1.01E-22 | 4306 | 2.11E-20 | TRUE | 1.48E-18 | TRUE | 1.25E-18 | TRUE | FALSE | -1.723857325 |
| **il1rn** | RV end diastolic volume \| Instance 2_BSA_indexed_corrected | -4.656629906 | -0.1660639773 | -0.2165738297 | 7.07E-47 | 0.3199616206 | -0.2165738297 | 7.07E-47 | 4306 | 2.07E-43 | TRUE | 1.03E-42 | TRUE | 8.74E-43 | TRUE | FALSE | -4.656629906 |
| **il1rn** | RV end systolic volume \| Instance 2_BSA_indexed_corrected | -2.104582081 | 0.03877497755 | -0.1645247381 | 1.65E-27 | 0.192326747 | -0.1645247381 | 1.65E-27 | 4306 | 6.72E-25 | TRUE | 2.42E-23 | TRUE | 2.05E-23 | TRUE | FALSE | -2.104582081 |
| **il6** | LV end diastolic volume \| Instance 2(participant - p24100_i2)_BSA_indexed_corrected | -1.733726967 | 0.1852027846 | -0.1127709853 | 7.38E-14 | 0.2310012635 | -0.1127709853 | 7.38E-14 | 4375 | 3.87E-12 | TRUE | 1.08E-09 | TRUE | 9.14E-10 | TRUE | FALSE | -1.733726967 |
| **il6** | LV end systolic volume \| Instance 2(participant - p24101_i2)_BSA_indexed_corrected | -0.7678612668 | 0.2375883328 | -0.08604159297 | 1.20E-08 | 0.1344531635 | -0.08604159297 | 1.20E-08 | 4375 | 2.43E-07 | TRUE | 0.0001752136336 | TRUE | 0.0001481956913 | TRUE | FALSE | -0.7678612668 |
| **il6** | RV end diastolic volume \| Instance 2_BSA_indexed_corrected | -1.913181198 | 0.05826029017 | -0.1156711674 | 1.66E-14 | 0.2484369816 | -0.1156711674 | 1.66E-14 | 4375 | 9.76E-13 | TRUE | 2.43E-10 | TRUE | 2.05E-10 | TRUE | FALSE | -1.913181198 |
| **il6** | RV end systolic volume \| Instance 2_BSA_indexed_corrected | -0.9216077951 | 0.1279163915 | -0.09369733829 | 5.32E-10 | 0.1480861193 | -0.09369733829 | 5.32E-10 | 4375 | 1.41E-08 | TRUE | 7.78E-06 | TRUE | 6.58E-06 | TRUE | FALSE | -0.9216077951 |
| **inhbc** | LV end diastolic volume \| Instance 2(participant - p24100_i2)_BSA_indexed_corrected | -4.599662876 | -0.3487885404 | -0.1863799966 | 5.37E-35 | 0.3693696194 | -0.1863799966 | 5.37E-35 | 4311 | 6.05E-32 | TRUE | 7.86E-31 | TRUE | 6.65E-31 | TRUE | FALSE | -4.599662876 |
| **inhbc** | LV end systolic volume \| Instance 2(participant - p24101_i2)_BSA_indexed_corrected | -2.276765861 | -0.02978007076 | -0.1588096583 | 9.52E-26 | 0.2156286205 | -0.1588096583 | 9.52E-26 | 4311 | 3.24E-23 | TRUE | 1.39E-21 | TRUE | 1.18E-21 | TRUE | FALSE | -2.276765861 |
| **inhbc** | RV end diastolic volume \| Instance 2_BSA_indexed_corrected | -5.000893654 | -0.4876645121 | -0.1882379085 | 1.12E-35 | 0.3974828256 | -0.1882379085 | 1.12E-35 | 4311 | 1.65E-32 | TRUE | 1.65E-31 | TRUE | 1.39E-31 | TRUE | FALSE | -5.000893654 |
| **inhbc** | RV end systolic volume \| Instance 2_BSA_indexed_corrected | -2.315714603 | -0.1215072482 | -0.1465893033 | 3.88E-22 | 0.2380551002 | -0.1465893033 | 3.88E-22 | 4311 | 7.27E-20 | TRUE | 5.67E-18 | TRUE | 4.80E-18 | TRUE | FALSE | -2.315714603 |
| **ism1** | LV end diastolic volume \| Instance 2(participant - p24100_i2)_BSA_indexed_corrected | -2.439065823 | 0.1825492214 | -0.08756719235 | 7.79E-09 | 0.421664115 | -0.08756719235 | 7.79E-09 | 4332 | 1.62E-07 | TRUE | 0.0001139815805 | TRUE | 9.64E-05 | TRUE | FALSE | -2.439065823 |
| **ism1** | RV end diastolic volume \| Instance 2_BSA_indexed_corrected | -2.982618965 | 0.03735069842 | -0.09955026554 | 5.15E-11 | 0.4530526605 | -0.09955026554 | 5.15E-11 | 4332 | 1.63E-09 | TRUE | 7.53E-07 | TRUE | 6.37E-07 | TRUE | FALSE | -2.982618965 |
| **itga11** | LV end diastolic volume \| Instance 2(participant - p24100_i2)_BSA_indexed_corrected | 3.008106972 | 0.3300919731 | 0.09177349277 | 1.44E-09 | 0.4960157042 | 0.09177349277 | 1.44E-09 | 4332 | 3.41E-08 | TRUE | 2.10E-05 | TRUE | 1.78E-05 | TRUE | FALSE | 3.008106972 |
| **itga11** | LV end systolic volume \| Instance 2(participant - p24101_i2)_BSA_indexed_corrected | 1.364034223 | 0.3047455773 | 0.07173978228 | 2.28E-06 | 0.2882045342 | 0.07173978228 | 2.28E-06 | 4332 | 3.05E-05 | TRUE | 0.03341957408 | TRUE | 0.02826627576 | TRUE | FALSE | 1.364034223 |
| **itga11** | RV end diastolic volume \| Instance 2_BSA_indexed_corrected | 3.810858821 | 0.2161544376 | 0.1080870311 | 9.83E-13 | 0.5326641881 | 0.1080870311 | 9.83E-13 | 4332 | 4.23E-11 | TRUE | 1.44E-08 | TRUE | 1.22E-08 | TRUE | FALSE | 3.810858821 |
| **itga11** | RV end systolic volume \| Instance 2_BSA_indexed_corrected | 1.742465696 | 0.2078442594 | 0.08301823882 | 4.45E-08 | 0.3178666807 | 0.08301823882 | 4.45E-08 | 4332 | 8.17E-07 | TRUE | 0.0006511625864 | TRUE | 0.0005507533156 | TRUE | FALSE | 1.742465696 |
| **itga5** | LV end diastolic volume \| Instance 2(participant - p24100_i2)_BSA_indexed_corrected | -5.756593503 | 0.02039591536 | -0.1313676534 | 4.70E-18 | 0.6617725542 | -0.1313676534 | 4.70E-18 | 4311 | 4.68E-16 | TRUE | 6.88E-14 | TRUE | 5.82E-14 | TRUE | FALSE | -5.756593503 |
| **itga5** | LV end systolic volume \| Instance 2(participant - p24101_i2)_BSA_indexed_corrected | -2.318617126 | 0.1829017874 | -0.09108296538 | 2.08E-09 | 0.3861843232 | -0.09108296538 | 2.08E-09 | 4311 | 4.77E-08 | TRUE | 3.05E-05 | TRUE | 2.58E-05 | TRUE | FALSE | -2.318617126 |
| **itga5** | RV end diastolic volume \| Instance 2_BSA_indexed_corrected | -6.440668874 | -0.09653756175 | -0.1365337514 | 2.17E-19 | 0.7118960016 | -0.1365337514 | 2.17E-19 | 4311 | 2.80E-17 | TRUE | 3.18E-15 | TRUE | 2.69E-15 | TRUE | FALSE | -6.440668874 |
| **itga5** | RV end systolic volume \| Instance 2_BSA_indexed_corrected | -3.011173768 | 0.05798601589 | -0.1073501477 | 1.59E-12 | 0.4248425213 | -0.1073501477 | 1.59E-12 | 4311 | 6.53E-11 | TRUE | 2.32E-08 | TRUE | 1.96E-08 | TRUE | FALSE | -3.011173768 |

| | | | | | | | | | | | | | | | | | |
|---|---|---|---|---|---|---|---|---|---|---|---|---|---|---|---|---|---|
| **itgam** | LV end systolic volume \| Instance 2(participant - p24101_i2)_BSA_indexed_corrected | 1.547330406 | 0.3136206862 | 0.07859508842 | 2.38E-07 | 0.2989880885 | 0.07859508842 | 2.38E-07 | 4311 | 3.83E-06 | TRUE | 0.003481881639 | TRUE | 0.00294497549 | TRUE | FALSE | 1.547330406 |
| **itgam** | RV end diastolic volume \| Instance 2_BSA_indexed_corrected | 2.691855458 | 0.2666414347 | 0.07378447854 | 1.24E-06 | 0.5542593972 | 0.07378447854 | 1.24E-06 | 4311 | 1.74E-05 | TRUE | 0.01808401371 | TRUE | 0.01529545879 | TRUE | FALSE | 2.691855458 |
| **itgav** | LV end diastolic volume \| Instance 2(participant - p24100_i2)_BSA_indexed_corrected | 5.834716304 | 0.418907957 | 0.09096276972 | 1.67E-09 | 0.9659666451 | 0.09096276972 | 1.67E-09 | 4375 | 3.88E-08 | TRUE | 2.44E-05 | TRUE | 2.06E-05 | TRUE | FALSE | 5.834716304 |
| **itgav** | LV end systolic volume \| Instance 2(participant - p24101_i2)_BSA_indexed_corrected | 3.488441972 | 0.3513076072 | 0.09368810291 | 5.34E-10 | 0.5605868472 | 0.09368810291 | 5.34E-10 | 4375 | 1.41E-08 | TRUE | 7.81E-06 | TRUE | 6.61E-06 | TRUE | FALSE | 3.488441972 |
| **itgav** | RV end diastolic volume \| Instance 2_BSA_indexed_corrected | 6.796703272 | 0.3201992954 | 0.09849055512 | 6.64E-11 | 1.038477975 | 0.09849055512 | 6.64E-11 | 4375 | 2.05E-09 | TRUE | 9.72E-07 | TRUE | 8.22E-07 | TRUE | FALSE | 6.796703272 |
| **itgav** | RV end systolic volume \| Instance 2_BSA_indexed_corrected | 3.523900611 | 0.2569176659 | 0.08586813412 | 1.28E-08 | 0.6182937475 | 0.08586813412 | 1.28E-08 | 4375 | 2.59E-07 | TRUE | 0.0001874664759 | TRUE | 0.0001585591453 | TRUE | FALSE | 3.523900611 |
| **itgb6** | RV end diastolic volume \| Instance 2_BSA_indexed_corrected | 3.001895851 | 0.2251281316 | 0.07803166755 | 3.16E-07 | 0.5861046441 | 0.07803166755 | 3.16E-07 | 4284 | 4.97E-06 | TRUE | 0.004622782036 | TRUE | 0.003909949046 | TRUE | FALSE | 3.001895851 |
| **itgb6** | RV end systolic volume \| Instance 2_BSA_indexed_corrected | 1.833731922 | 0.1977768541 | 0.08029374977 | 1.42E-07 | 0.347877447 | 0.08029374977 | 1.42E-07 | 4284 | 2.38E-06 | TRUE | 0.002080505183 | TRUE | 0.001759691284 | TRUE | FALSE | 1.833731922 |
| **kdr** | LV end diastolic volume \| Instance 2(participant - p24100_i2)_BSA_indexed_corrected | -4.679942484 | 0.2724985907 | -0.1095139208 | 4.68E-13 | 0.6449207629 | -0.1095139208 | 4.68E-13 | 4340 | 2.18E-11 | TRUE | 6.84E-09 | TRUE | 5.79E-09 | TRUE | FALSE | -4.679942484 |
| **kdr** | LV end systolic volume \| Instance 2(participant - p24101_i2)_BSA_indexed_corrected | -2.420703146 | 0.2683393783 | -0.0976030118 | 1.17E-10 | 0.3747615316 | -0.0976030118 | 1.17E-10 | 4340 | 3.45E-09 | TRUE | 1.71E-06 | TRUE | 1.44E-06 | TRUE | FALSE | -2.420703146 |
| **kdr** | RV end diastolic volume \| Instance 2_BSA_indexed_corrected | -4.736776732 | 0.1669443961 | -0.1030592264 | 1.01E-11 | 0.6941168096 | -0.1030592264 | 1.01E-11 | 4340 | 3.64E-10 | TRUE | 1.47E-07 | TRUE | 1.25E-07 | TRUE | FALSE | -4.736776732 |
| **kdr** | RV end systolic volume \| Instance 2_BSA_indexed_corrected | -2.599520046 | 0.1673195885 | -0.09507263188 | 3.48E-10 | 0.413257938 | -0.09507263188 | 3.48E-10 | 4340 | 9.48E-09 | TRUE | 5.09E-06 | TRUE | 4.31E-06 | TRUE | FALSE | -2.599520046 |
| **klk6** | LV cardiac output \| Instance 2_BSA_indexed_corrected | -0.1505253833 | -0.01944846363 | -0.08913590403 | 4.15E-09 | 0.02555528176 | -0.08913590403 | 4.15E-09 | 4334 | 9.08E-08 | TRUE | 6.07E-05 | TRUE | 5.14E-05 | TRUE | FALSE | -0.1505253833 |
| **klk6** | LV end diastolic volume \| Instance 2(participant - p24100_i2)_BSA_indexed_corrected | -3.777789605 | 0.360334563 | -0.09981532888 | 4.53E-11 | 0.5721659276 | -0.09981532888 | 4.53E-11 | 4334 | 1.46E-09 | TRUE | 6.62E-07 | TRUE | 5.60E-07 | TRUE | FALSE | -3.777789605 |
| **klk6** | LV end systolic volume \| Instance 2(participant - p24101_i2)_BSA_indexed_corrected | -1.73118609 | 0.3133977985 | -0.07882332411 | 2.04E-07 | 0.3326529769 | -0.07882332411 | 2.04E-07 | 4334 | 3.34E-06 | TRUE | 0.002982615219 | TRUE | 0.002522695953 | TRUE | FALSE | -1.73118609 |
| **klk6** | RV end diastolic volume \| Instance 2_BSA_indexed_corrected | -3.366570396 | 0.2560222309 | -0.08270944899 | 4.96E-08 | 0.6163079529 | -0.08270944899 | 4.96E-08 | 4334 | 9.02E-07 | TRUE | 0.0007254174303 | TRUE | 0.0006135580625 | TRUE | FALSE | -3.366570396 |
| **klk8** | LV cardiac output \| Instance 2_BSA_indexed_corrected | -0.1030301555 | -0.02112184505 | -0.0728454837 | 2.56E-06 | 0.02187580931 | -0.0728454837 | 2.56E-06 | 4160 | 3.37E-05 | TRUE | 0.03746421027 | TRUE | 0.03168722905 | TRUE | FALSE | -0.1030301555 |
| **klk8** | LV end diastolic volume \| Instance 2(participant - p24100_i2)_BSA_indexed_corrected | -2.317244321 | 0.3832789349 | -0.0726184835 | 2.75E-06 | 0.4935535328 | -0.0726184835 | 2.75E-06 | 4160 | 3.60E-05 | TRUE | 0.04025711307 | TRUE | 0.03404946623 | TRUE | FALSE | -2.317244321 |
| **klrd1** | LV end diastolic volume \| Instance 2(participant - p24100_i2)_BSA_indexed_corrected | -1.865312095 | 0.2541106648 | -0.07870440075 | 1.95E-07 | 0.3578572159 | -0.07870440075 | 1.95E-07 | 4361 | 3.19E-06 | TRUE | 0.002852654992 | TRUE | 0.002412775592 | TRUE | FALSE | -1.865312095 |
| **klrd1** | RV end diastolic volume \| Instance 2_BSA_indexed_corrected | -2.075207393 | 0.1344630942 | -0.08137104816 | 7.41E-08 | 0.3849954478 | -0.08137104816 | 7.41E-08 | 4361 | 1.31E-06 | TRUE | 0.001083963702 | TRUE | 0.0009168164991 | TRUE | FALSE | -2.075207393 |
| **krt18** | LV end diastolic volume \| Instance 2(participant - p24100_i2)_BSA_indexed_corrected | -1.799593682 | 0.2970677407 | -0.1380139527 | 4.73E-20 | 0.195292533 | -0.1380139527 | 4.73E-20 | 4375 | 6.92E-18 | TRUE | 6.92E-16 | TRUE | 5.85E-16 | TRUE | FALSE | -1.799593682 |
| **krt18** | LV end systolic volume \| Instance 2(participant - p24101_i2)_BSA_indexed_corrected | -0.7502329049 | 0.2885878827 | -0.09911825633 | 5.02E-11 | 0.1138960912 | -0.09911825633 | 5.02E-11 | 4375 | 1.60E-09 | TRUE | 7.34E-07 | TRUE | 6.21E-07 | TRUE | FALSE | -0.7502329049 |
| **krt18** | RV end diastolic volume \| Instance 2_BSA_indexed_corrected | -1.969628273 | 0.1822089408 | -0.1404058322 | 1.05E-20 | 0.2100321741 | -0.1404058322 | 1.05E-20 | 4375 | 1.64E-18 | TRUE | 1.54E-16 | TRUE | 1.30E-16 | TRUE | FALSE | -1.969628273 |
| **krt18** | RV end systolic volume \| Instance 2_BSA_indexed_corrected | -0.8606054872 | 0.1903661396 | -0.1031613672 | 7.93E-12 | 0.1254798409 | -0.1031613672 | 7.93E-12 | 4375 | 2.95E-10 | TRUE | 1.16E-07 | TRUE | 9.81E-08 | TRUE | FALSE | -0.8606054872 |
| **kynu** | LV end diastolic volume \| Instance 2(participant - p24100_i2)_BSA_indexed_corrected | -2.938081467 | 0.2264782771 | -0.1248423643 | 1.29E-16 | 0.353669322 | -0.1248423643 | 1.29E-16 | 4361 | 1.09E-14 | TRUE | 1.89E-12 | TRUE | 1.60E-12 | TRUE | FALSE | -2.938081467 |
| **kynu** | LV end systolic volume \| Instance 2(participant - p24101_i2)_BSA_indexed_corrected | -1.272231104 | 0.2560853028 | -0.09305955458 | 7.41E-10 | 0.2061687807 | -0.09305955458 | 7.41E-10 | 4361 | 1.89E-08 | TRUE | 1.08E-05 | TRUE | 9.17E-06 | TRUE | FALSE | -1.272231104 |
| **kynu** | RV end diastolic volume \| Instance 2_BSA_indexed_corrected | -2.907916204 | 0.121490032 | -0.1148261841 | 2.83E-14 | 0.3810354477 | -0.1148261841 | 2.83E-14 | 4361 | 1.60E-12 | TRUE | 4.14E-10 | TRUE | 3.50E-10 | TRUE | FALSE | -2.907916204 |
| **kynu** | RV end systolic volume \| Instance 2_BSA_indexed_corrected | -1.37113038 | 0.1572604207 | -0.0910271415 | 1.72E-09 | 0.2271996454 | -0.0910271415 | 1.72E-09 | 4361 | 4.00E-08 | TRUE | 2.52E-05 | TRUE | 2.13E-05 | TRUE | FALSE | -1.37113038 |
| **lair1** | LV end diastolic volume \| Instance 2(participant - p24100_i2)_BSA_indexed_corrected | -2.979024327 | 0.07606603516 | -0.09361787356 | 7.72E-10 | 0.4832485259 | -0.09361787356 | 7.72E-10 | 4300 | 1.96E-08 | TRUE | 1.13E-05 | TRUE | 9.55E-06 | TRUE | FALSE | -2.979024327 |
| **lair1** | RV end diastolic volume \| Instance 2_BSA_indexed_corrected | -4.014259826 | -0.1341110013 | -0.1173875528 | 1.14E-14 | 0.5180088304 | -0.1173875528 | 1.14E-14 | 4300 | 6.89E-13 | TRUE | 1.67E-10 | TRUE | 1.42E-10 | TRUE | FALSE | -4.014259826 |
| **lair1** | RV end systolic volume \| Instance 2_BSA_indexed_corrected | -1.841946804 | 0.02445556231 | -0.0906705229 | 2.58E-09 | 0.3085923674 | -0.0906705229 | 2.58E-09 | 4300 | 5.84E-08 | TRUE | 3.78E-05 | TRUE | 3.19E-05 | TRUE | FALSE | -1.841946804 |
| **lamp2** | LV end diastolic volume \| Instance 2(participant - p24100_i2)_BSA_indexed_corrected | -6.268801004 | 0.2917150331 | -0.1121286093 | 3.30E-13 | 0.8581471319 | -0.1121286093 | 3.30E-13 | 4193 | 1.55E-11 | TRUE | 4.83E-09 | TRUE | 4.08E-09 | TRUE | FALSE | -6.268801004 |
| **lamp2** | LV end systolic volume \| Instance 2(participant - p24101_i2)_BSA_indexed_corrected | -2.795294907 | 0.2742521374 | -0.08623773763 | 2.23E-08 | 0.4988276296 | -0.08623773763 | 2.23E-08 | 4193 | 4.34E-07 | TRUE | 0.0003266216767 | TRUE | 0.0002762566141 | TRUE | FALSE | -2.795294907 |
| **lamp2** | RV end diastolic volume \| Instance 2_BSA_indexed_corrected | -7.083572444 | 0.1764601777 | -0.1174669786 | 2.34E-14 | 0.9250400637 | -0.1174669786 | 2.34E-14 | 4193 | 1.36E-12 | TRUE | 3.42E-10 | TRUE | 2.89E-10 | TRUE | FALSE | -7.083572444 |
| **lamp2** | RV end systolic volume \| Instance 2_BSA_indexed_corrected | -3.375481309 | 0.1888440177 | -0.09422645731 | 9.76E-10 | 0.5508932299 | -0.09422645731 | 9.76E-10 | 4193 | 2.42E-08 | TRUE | 1.43E-05 | TRUE | 1.21E-05 | TRUE | FALSE | -3.375481309 |
| **lamp3** | LV end diastolic volume \| Instance 2(participant - p24100_i2)_BSA_indexed_corrected | -1.656919245 | 0.2005310306 | -0.07771901239 | 2.83E-07 | 0.3221914608 | -0.07771901239 | 2.83E-07 | 4354 | 4.50E-06 | TRUE | 0.004137170997 | TRUE | 0.003499219229 | TRUE | FALSE | -1.656919245 |
| **lamp3** | RV end diastolic volume \| Instance 2_BSA_indexed_corrected | -1.816817666 | 0.0742233523 | -0.07921573059 | 1.66E-07 | 0.3465680701 | -0.07921573059 | 1.66E-07 | 4354 | 2.74E-06 | TRUE | 0.002429947984 | TRUE | 0.002055250005 | TRUE | FALSE | -1.816817666 |
| **lbp** | LV end diastolic volume \| Instance 2(participant - p24100_i2)_BSA_indexed_corrected | -1.335172849 | 0.2434123198 | -0.07124776248 | 2.74E-06 | 0.2843603431 | -0.07124776248 | 2.74E-06 | 4323 | 3.60E-05 | TRUE | 0.04014381524 | TRUE | 0.03395363893 | TRUE | FALSE | -1.335172849 |
| **lbp** | RV end diastolic volume \| Instance 2_BSA_indexed_corrected | -1.856671646 | 0.08047842273 | -0.09217398638 | 1.27E-09 | 0.3051278347 | -0.09217398638 | 1.27E-09 | 4323 | 3.05E-08 | TRUE | 1.85E-05 | TRUE | 1.57E-05 | TRUE | FALSE | -1.856671646 |
| **lbr** | LV end diastolic volume \| Instance 2(participant - p24100_i2)_BSA_indexed_corrected | -1.590353663 | 0.3078489538 | -0.09857014003 | 6.81E-11 | 0.2431281032 | -0.09857014003 | 6.81E-11 | 4363 | 2.09E-09 | TRUE | 9.96E-07 | TRUE | 8.42E-07 | TRUE | FALSE | -1.590353663 |
| **lbr** | LV end systolic volume \| Instance 2(participant - p24101_i2)_BSA_indexed_corrected | -0.6760635217 | 0.2924221103 | -0.07224691122 | 1.78E-06 | 0.1413314394 | -0.07224691122 | 1.78E-06 | 4363 | 2.43E-05 | TRUE | 0.02603091267 | TRUE | 0.02201694593 | TRUE | FALSE | -0.6760635217 |
| **lbr** | RV end diastolic volume \| Instance 2_BSA_indexed_corrected | -1.547498478 | 0.1964455908 | -0.08908441182 | 3.76E-09 | 0.2620024234 | -0.08908441182 | 3.76E-09 | 4363 | 8.29E-08 | TRUE | 5.50E-05 | TRUE | 4.66E-05 | TRUE | FALSE | -1.547498478 |
| **lcn2** | LV end diastolic volume \| Instance 2(participant - p24100_i2)_BSA_indexed_corrected | -2.328893287 | 0.2048858053 | -0.07151548867 | 3.32E-06 | 0.5001891147 | -0.07151548867 | 3.32E-06 | 4219 | 4.31E-05 | TRUE | 0.04861328345 | TRUE | 0.04111711514 | TRUE | FALSE | -2.328893287 |
| **lcn2** | RV end diastolic volume \| Instance 2_BSA_indexed_corrected | -2.73793529 | 0.1112133772 | -0.07824382798 | 3.61E-07 | 0.5372022302 | -0.07824382798 | 3.61E-07 | 4219 | 5.61E-06 | TRUE | 0.00527916028 | TRUE | 0.004465113765 | TRUE | FALSE | -2.73793529 |
| **ldlr** | LV end diastolic volume \| Instance 2(participant - p24100_i2)_BSA_indexed_corrected | -3.791067983 | -0.01282051039 | -0.1901795196 | 1.33E-36 | 0.2972376984 | -0.1901795196 | 1.33E-36 | 4337 | 2.19E-33 | TRUE | 1.95E-32 | TRUE | 1.65E-32 | TRUE | FALSE | -3.791067983 |
| **ldlr** | LV end systolic volume \| Instance 2(participant - p24101_i2)_BSA_indexed_corrected | -1.591181016 | 0.1566658862 | -0.1372543594 | 1.09E-19 | 0.1744090765 | -0.1372543594 | 1.09E-19 | 4337 | 1.51E-17 | TRUE | 1.60E-15 | TRUE | 1.35E-15 | TRUE | FALSE | -1.591181016 |
| **ldlr** | RV end diastolic volume \| Instance 2_BSA_indexed_corrected | -3.956785936 | -0.1216980126 | -0.1844091055 | 1.76E-34 | 0.3202968711 | -0.1844091055 | 1.76E-34 | 4337 | 1.84E-31 | TRUE | 2.58E-30 | TRUE | 2.18E-30 | TRUE | FALSE | -3.956785936 |
| **ldlr** | RV end systolic volume \| Instance 2_BSA_indexed_corrected | -1.68538021 | 0.05937152249 | -0.1319391576 | 2.67E-18 | 0.1923163719 | -0.1319391576 | 2.67E-18 | 4337 | 2.73E-16 | TRUE | 3.91E-14 | TRUE | 3.30E-14 | TRUE | FALSE | -1.68538021 |
| **lep** | LV end diastolic volume \| Instance 2(participant - p24100_i2)_BSA_indexed_corrected | -2.238507252 | -0.3977297083 | -0.2293628678 | 2.52E-53 | 0.1436516664 | -0.2293628678 | 2.52E-53 | 4375 | 9.23E-50 | TRUE | 3.69E-49 | TRUE | 3.12E-49 | TRUE | FALSE | -2.238507252 |
| **lep** | LV end systolic volume \| Instance 2(participant - p24101_i2)_BSA_indexed_corrected | -1.030967271 | -0.03385136552 | -0.1819778348 | 6.91E-34 | 0.08424106491 | -0.1819778348 | 6.91E-34 | 4375 | 6.32E-31 | TRUE | 1.01E-29 | TRUE | 8.56E-30 | TRUE | FALSE | -1.030967271 |
| **lep** | RV end diastolic volume \| Instance 2_BSA_indexed_corrected | -2.565890011 | -0.6170991094 | -0.2443739926 | 1.66E-60 | 0.1539650773 | -0.2443739926 | 1.66E-60 | 4375 | 1.21E-56 | TRUE | 2.43E-56 | TRUE | 2.05E-56 | TRUE | FALSE | -2.565890011 |
| **lep** | RV end systolic volume \| Instance 2_BSA_indexed_corrected | -1.229904681 | -0.1953608648 | -0.1969701895 | 1.62E-39 | 0.0925739328 | -0.1969701895 | 1.62E-39 | 4375 | 3.39E-36 | TRUE | 2.37E-35 | TRUE | 2.01E-35 | TRUE | FALSE | -1.229904681 |
| **lepr** | LV end diastolic volume \| Instance 2(participant - p24100_i2)_BSA_indexed_corrected | 2.518103258 | 0.33121673 | 0.07378544915 | 1.03E-06 | 0.5146685471 | 0.07378544915 | 1.03E-06 | 4375 | 1.47E-05 | TRUE | 0.01507685278 | TRUE | 0.01275200208 | TRUE | FALSE | 2.518103258 |
| **lepr** | RV end diastolic volume \| Instance 2_BSA_indexed_corrected | 3.03688141 | 0.2171532315 | 0.08271368457 | 4.28E-08 | 0.5533118829 | 0.08271368457 | 4.28E-08 | 4375 | 7.90E-07 | TRUE | 0.0006264062958 | TRUE | 0.000529814445 | TRUE | FALSE | 3.03688141 |
| **lgals1** | LV end diastolic volume \| Instance 2(participant - p24100_i2)_BSA_indexed_corrected | -4.551607113 | -0.06651061289 | -0.1396743439 | 1.67E-20 | 0.4879554934 | -0.1396743439 | 1.67E-20 | 4375 | 2.50E-18 | TRUE | 2.45E-16 | TRUE | 2.07E-16 | TRUE | FALSE | -4.551607113 |
| **lgals1** | LV end systolic volume \| Instance 2(participant - p24101_i2)_BSA_indexed_corrected | -1.960299564 | 0.1312292165 | -0.1036294819 | 6.38E-12 | 0.2845147633 | -0.1036294819 | 6.38E-12 | 4375 | 2.40E-10 | TRUE | 9.33E-08 | TRUE | 7.89E-08 | TRUE | FALSE | -1.960299564 |
| **lgals1** | RV end diastolic volume \| Instance 2_BSA_indexed_corrected | -5.216649132 | -0.2373807455 | -0.1487975718 | 4.40E-23 | 0.5242569855 | -0.1487975718 | 4.40E-23 | 4375 | 9.62E-21 | TRUE | 6.44E-19 | TRUE | 5.45E-19 | TRUE | FALSE | -5.216649132 |
| **lgals1** | RV end systolic volume \| Instance 2_BSA_indexed_corrected | -2.391467937 | -0.003302341124 | -0.1147046117 | 2.74E-14 | 0.3131972533 | -0.1147046117 | 2.74E-14 | 4375 | 1.55E-12 | TRUE | 4.01E-10 | TRUE | 3.39E-10 | TRUE | FALSE | -2.391467937 |
| **lgals4** | RV end diastolic volume \| Instance 2_BSA_indexed_corrected | -1.706675994 | 0.09151043689 | -0.07366356541 | 1.33E-06 | 0.3524387234 | -0.07366356541 | 1.33E-06 | 4300 | 1.85E-05 | TRUE | 0.01941965305 | TRUE | 0.01642514255 | TRUE | FALSE | -1.706675994 |
| **lgals9** | LV end diastolic volume \| Instance 2(participant - p24100_i2)_BSA_indexed_corrected | -4.385815108 | -0.0739091365 | -0.1268016819 | 6.39E-17 | 0.5225969356 | -0.1268016819 | 6.39E-17 | 4312 | 5.59E-15 | TRUE | 9.34E-13 | TRUE | 7.90E-13 | TRUE | FALSE | -4.385815108 |
| **lgals9** | LV end systolic volume \| Instance 2(participant - p24101_i2)_BSA_indexed_corrected | -1.840845537 | 0.1300048751 | -0.09180864604 | 1.54E-09 | 0.3041284702 | -0.09180864604 | 1.54E-09 | 4312 | 3.63E-08 | TRUE | 2.26E-05 | TRUE | 1.91E-05 | TRUE | FALSE | -1.840845537 |
| **lgals9** | RV end diastolic volume \| Instance 2_BSA_indexed_corrected | -5.082358237 | -0.2507023283 | -0.1367897373 | 1.84E-19 | 0.5606237507 | -0.1367897373 | 1.84E-19 | 4312 | 2.41E-17 | TRUE | 2.70E-15 | TRUE | 2.28E-15 | TRUE | FALSE | -5.082358237 |
| **lgals9** | RV end systolic volume \| Instance 2_BSA_indexed_corrected | -2.441065419 | -0.02576549689 | -0.1104175199 | 3.57E-13 | 0.3346871656 | -0.1104175199 | 3.57E-13 | 4312 | 1.68E-11 | TRUE | 5.23E-09 | TRUE | 4.42E-09 | TRUE | FALSE | -2.441065419 |
| **lilra5** | LV end diastolic volume \| Instance 2(participant - p24100_i2)_BSA_indexed_corrected | -3.102079588 | 0.06493236934 | -0.09094208505 | 1.69E-09 | 0.5137414136 | -0.09094208505 | 1.69E-09 | 4374 | 3.93E-08 | TRUE | 2.47E-05 | TRUE | 2.09E-05 | TRUE | FALSE | -3.102079588 |
| **lilra5** | RV end diastolic volume \| Instance 2_BSA_indexed_corrected | -3.52090269 | -0.08439111418 | -0.09593943665 | 2.04E-10 | 0.5524701547 | -0.09593943665 | 2.04E-10 | 4374 | 5.79E-09 | TRUE | 2.99E-06 | TRUE | 2.53E-06 | TRUE | FALSE | -3.52090269 |
| **lilra5** | RV end systolic volume \| Instance 2_BSA_indexed_corrected | -1.738426486 | 0.05504764464 | -0.07965133074 | 1.33E-07 | 0.3290343007 | -0.07965133074 | 1.33E-07 | 4374 | 2.24E-06 | TRUE | 0.001945248884 | TRUE | 0.001645291506 | TRUE | FALSE | -1.738426486 |
| **lilrb4** | LV end diastolic volume \| Instance 2(participant - p24100_i2)_BSA_indexed_corrected | -2.242654665 | 0.1589039572 | -0.08658701739 | 1.38E-08 | 0.3943230265 | -0.08658701739 | 1.38E-08 | 4284 | 2.77E-07 | TRUE | 0.0002013356182 | TRUE | 0.0001702896659 | TRUE | FALSE | -2.242654665 |
| **lilrb4** | RV end diastolic volume \| Instance 2_BSA_indexed_corrected | -2.50234911 | 0.02261564373 | -0.0896797703 | 4.11E-09 | 0.4246944854 | -0.0896797703 | 4.11E-09 | 4284 | 9.01E-08 | TRUE | 6.01E-05 | TRUE | 5.08E-05 | TRUE | FALSE | -2.50234911 |

| | | | | | | | | | | | | | | | | | |
|---|---|---|---|---|---|---|---|---|---|---|---|---|---|---|---|---|---|
| **lpl** | LV end diastolic volume \| Instance 2(participant - p24100_i2)_BSA_indexed_corrected | 2.688991461 | 0.4071531672 | 0.1308234412 | 3.69E-18 | 0.3081523751 | 0.1308234412 | 3.69E-18 | 4375 | 3.75E-16 | TRUE | 5.39E-14 | TRUE | 4.56E-14 | TRUE | FALSE | 2.688991461 |
| **lpl** | LV end systolic volume \| Instance 2(participant - p24101_i2)_BSA_indexed_corrected | 1.243369704 | 0.3369448284 | 0.1042090502 | 4.86E-12 | 0.1794461278 | 0.1042090502 | 4.86E-12 | 4375 | 1.87E-10 | TRUE | 7.11E-08 | TRUE | 6.01E-08 | TRUE | FALSE | 1.243369704 |
| **lpl** | RV end diastolic volume \| Instance 2_BSA_indexed_corrected | 2.683016177 | 0.2974602485 | 0.1213310011 | 8.09E-16 | 0.3319260502 | 0.1213310011 | 8.09E-16 | 4375 | 6.07E-14 | TRUE | 1.18E-11 | TRUE | 1.00E-11 | TRUE | FALSE | 2.683016177 |
| **lpl** | RV end systolic volume \| Instance 2_BSA_indexed_corrected | 1.240277013 | 0.2420921781 | 0.09431467758 | 4.09E-10 | 0.1979745884 | 0.09431467758 | 4.09E-10 | 4375 | 1.10E-08 | TRUE | 5.99E-06 | TRUE | 5.06E-06 | TRUE | FALSE | 1.240277013 |
| **lrp11** | LV end diastolic volume \| Instance 2(participant - p24100_i2)_BSA_indexed_corrected | -2.366149804 | 0.1911960341 | -0.0731280931 | 1.28E-06 | 0.4879820992 | -0.0731280931 | 1.28E-06 | 4375 | 1.80E-05 | TRUE | 0.01879856868 | TRUE | 0.01589982939 | TRUE | FALSE | -2.366149804 |
| **lrp11** | RV end diastolic volume \| Instance 2_BSA_indexed_corrected | -2.959362038 | 0.04105412066 | -0.08501439226 | 1.78E-08 | 0.5244943161 | -0.08501439226 | 1.78E-08 | 4375 | 3.53E-07 | TRUE | 0.0002609632793 | TRUE | 0.0002207227416 | TRUE | FALSE | -2.959362038 |
| **lsp1** | LV end diastolic volume \| Instance 2(participant - p24100_i2)_BSA_indexed_corrected | -3.586021715 | 0.06595640164 | -0.1326276705 | 1.86E-18 | 0.4072688467 | -0.1326276705 | 1.86E-18 | 4332 | 1.99E-16 | TRUE | 2.72E-14 | TRUE | 2.30E-14 | TRUE | FALSE | -3.586021715 |
| **lsp1** | LV end systolic volume \| Instance 2(participant - p24101_i2)_BSA_indexed_corrected | -1.680082547 | 0.1804417579 | -0.107118048 | 1.57E-12 | 0.2369834456 | -0.107118048 | 1.57E-12 | 4332 | 6.49E-11 | TRUE | 2.29E-08 | TRUE | 1.94E-08 | TRUE | FALSE | -1.680082547 |
| **lsp1** | RV end diastolic volume \| Instance 2_BSA_indexed_corrected | -3.066159933 | 0.005463831 | -0.1054247972 | 3.49E-12 | 0.4395224987 | -0.1054247972 | 3.49E-12 | 4332 | 1.38E-10 | TRUE | 5.11E-08 | TRUE | 4.32E-08 | TRUE | FALSE | -3.066159933 |
| **lsp1** | RV end systolic volume \| Instance 2_BSA_indexed_corrected | -1.681546153 | 0.08804661971 | -0.09712148471 | 1.50E-10 | 0.2618734984 | -0.09712148471 | 1.50E-10 | 4332 | 4.32E-09 | TRUE | 2.19E-06 | TRUE | 1.85E-06 | TRUE | FALSE | -1.681546153 |
| **ltbp2** | LV end systolic volume \| Instance 2(participant - p24101_i2)_BSA_indexed_corrected | 1.332432422 | 0.4198885959 | 0.07134455504 | 2.32E-06 | 0.2816997798 | 0.07134455504 | 2.32E-06 | 4375 | 3.08E-05 | TRUE | 0.03388686186 | TRUE | 0.02866150776 | TRUE | FALSE | 1.332432422 |
| **ltbp3** | LV end diastolic volume \| Instance 2(participant - p24100_i2)_BSA_indexed_corrected | -1.83063936 | 0.2925529664 | -0.0916915541 | 1.23E-09 | 0.3006425004 | -0.0916915541 | 1.23E-09 | 4375 | 2.99E-08 | TRUE | 1.80E-05 | TRUE | 1.53E-05 | TRUE | FALSE | -1.83063936 |
| **ltbp3** | RV end diastolic volume \| Instance 2_BSA_indexed_corrected | -1.88463416 | 0.1811970167 | -0.08774167899 | 6.13E-09 | 0.3235587737 | -0.08774167899 | 6.13E-09 | 4375 | 1.29E-07 | TRUE | 8.97E-05 | TRUE | 7.59E-05 | TRUE | FALSE | -1.88463416 |
| **ly6d** | LV cardiac output \| Instance 2_BSA_indexed_corrected | -0.1176883341 | -0.02472314235 | -0.0808991523 | 1.10E-07 | 0.02212751256 | -0.0808991523 | 1.10E-07 | 4296 | 1.89E-06 | TRUE | 0.001607293527 | TRUE | 0.001359448865 | TRUE | FALSE | -0.1176883341 |
| **ly6d** | LV end diastolic volume \| Instance 2(participant - p24100_i2)_BSA_indexed_corrected | -2.48880442 | 0.22913398 | -0.07634860116 | 5.44E-07 | 0.4960085771 | -0.07634860116 | 5.44E-07 | 4296 | 8.18E-06 | TRUE | 0.007961426525 | TRUE | 0.006733774555 | TRUE | FALSE | -2.48880442 |
| **ly6d** | RV end diastolic volume \| Instance 2_BSA_indexed_corrected | -2.770282905 | 0.1151722358 | -0.07898372902 | 2.18E-07 | 0.5335763441 | -0.07898372902 | 2.18E-07 | 4296 | 3.54E-06 | TRUE | 0.003185959519 | TRUE | 0.002694684561 | TRUE | FALSE | -2.770282905 |
| **ly96** | RV end diastolic volume \| Instance 2_BSA_indexed_corrected | -2.48226483 | 0.1604136754 | -0.07486692489 | 7.14E-07 | 0.4999744758 | -0.07486692489 | 7.14E-07 | 4375 | 1.05E-05 | TRUE | 0.01044536541 | TRUE | 0.008834690061 | TRUE | FALSE | -2.48226483 |
| **ly96** | RV end systolic volume \| Instance 2_BSA_indexed_corrected | -1.376570804 | 0.1710767489 | -0.06981560974 | 3.80E-06 | 0.2974371897 | -0.06981560974 | 3.80E-06 | 4375 | 4.87E-05 | TRUE | 0.05555170848 | FALSE | 0.04698563503 | TRUE | FALSE | -1.376570804 |
| **mad1l1** | LV end diastolic volume \| Instance 2(participant - p24100_i2)_BSA_indexed_corrected | -2.638571518 | 0.1943239301 | -0.1135891685 | 5.06E-14 | 0.3492371053 | -0.1135891685 | 5.06E-14 | 4369 | 2.75E-12 | TRUE | 7.41E-10 | TRUE | 6.26E-10 | TRUE | FALSE | -2.638571518 |
| **mad1l1** | LV end systolic volume \| Instance 2(participant - p24101_i2)_BSA_indexed_corrected | -1.228935821 | 0.2415867832 | -0.09112623708 | 1.60E-09 | 0.2032279577 | -0.09112623708 | 1.60E-09 | 4369 | 3.74E-08 | TRUE | 2.34E-05 | TRUE | 1.98E-05 | TRUE | FALSE | -1.228935821 |
| **mad1l1** | RV end diastolic volume \| Instance 2_BSA_indexed_corrected | -2.722681895 | 0.08213613555 | -0.1089197881 | 5.24E-13 | 0.3760165721 | -0.1089197881 | 5.24E-13 | 4369 | 2.40E-11 | TRUE | 7.67E-09 | TRUE | 6.49E-09 | TRUE | FALSE | -2.722681895 |
| **mad1l1** | RV end systolic volume \| Instance 2_BSA_indexed_corrected | -1.37657772 | 0.1372705165 | -0.09259980702 | 8.67E-10 | 0.2239902177 | -0.09259980702 | 8.67E-10 | 4369 | 2.16E-08 | TRUE | 1.27E-05 | TRUE | 1.07E-05 | TRUE | FALSE | -1.37657772 |
| **megf10** | LV end diastolic volume \| Instance 2(participant - p24100_i2)_BSA_indexed_corrected | 2.816448334 | 0.3629028997 | 0.1016723649 | 2.35E-11 | 0.4203482712 | 0.1016723649 | 2.35E-11 | 4300 | 7.96E-10 | TRUE | 3.44E-07 | TRUE | 2.91E-07 | TRUE | FALSE | 2.816448334 |
| **megf10** | LV end systolic volume \| Instance 2(participant - p24101_i2)_BSA_indexed_corrected | 1.498588513 | 0.3057521355 | 0.09335848279 | 8.59E-10 | 0.2437779942 | 0.09335848279 | 8.59E-10 | 4300 | 2.15E-08 | TRUE | 1.26E-05 | TRUE | 1.06E-05 | TRUE | FALSE | 1.498588513 |
| **megf10** | RV end diastolic volume \| Instance 2_BSA_indexed_corrected | 3.348696525 | 0.251413845 | 0.1124886682 | 1.39E-13 | 0.4511994506 | 0.1124886682 | 1.39E-13 | 4300 | 6.85E-12 | TRUE | 2.03E-09 | TRUE | 1.71E-09 | TRUE | FALSE | 3.348696525 |
| **megf10** | RV end systolic volume \| Instance 2_BSA_indexed_corrected | 1.862434442 | 0.202076707 | 0.1053140817 | 4.42E-12 | 0.2682498551 | 0.1053140817 | 4.42E-12 | 4300 | 1.71E-10 | TRUE | 6.46E-08 | TRUE | 5.47E-08 | TRUE | FALSE | 1.862434442 |
| **megf9** | LV end diastolic volume \| Instance 2(participant - p24100_i2)_BSA_indexed_corrected | -5.120541322 | 0.2515411585 | -0.09565686464 | 2.30E-10 | 0.805775206 | -0.09565686464 | 2.30E-10 | 4375 | 6.45E-09 | TRUE | 3.36E-06 | TRUE | 2.85E-06 | TRUE | FALSE | -5.120541322 |
| **megf9** | LV end systolic volume \| Instance 2(participant - p24101_i2)_BSA_indexed_corrected | -2.724495472 | 0.2579203702 | -0.08767895559 | 6.29E-09 | 0.4680855055 | -0.08767895559 | 6.29E-09 | 4375 | 1.32E-07 | TRUE | 9.20E-05 | TRUE | 7.78E-05 | TRUE | FALSE | -2.724495472 |
| **megf9** | RV end diastolic volume \| Instance 2_BSA_indexed_corrected | -5.461588397 | 0.135210002 | -0.09483563968 | 3.28E-10 | 0.8669535648 | -0.09483563968 | 3.28E-10 | 4375 | 9.01E-09 | TRUE | 4.79E-06 | TRUE | 4.05E-06 | TRUE | FALSE | -5.461588397 |
| **megf9** | RV end systolic volume \| Instance 2_BSA_indexed_corrected | -3.030782746 | 0.1570602822 | -0.08849515956 | 4.54E-09 | 0.5158676468 | -0.08849515956 | 4.54E-09 | 4375 | 9.88E-08 | TRUE | 6.64E-05 | TRUE | 5.62E-05 | TRUE | FALSE | -3.030782746 |
| **mep1b** | LV end diastolic volume \| Instance 2(participant - p24100_i2)_BSA_indexed_corrected | -0.7057334058 | 0.1862665123 | -0.0757335186 | 5.30E-07 | 0.1405121096 | -0.0757335186 | 5.30E-07 | 4375 | 8.00E-06 | TRUE | 0.007755840233 | TRUE | 0.006559889669 | TRUE | FALSE | -0.7057334058 |
| **mep1b** | RV end diastolic volume \| Instance 2_BSA_indexed_corrected | -0.7672935347 | 0.06214865394 | -0.07653522132 | 4.01E-07 | 0.1511592566 | -0.07653522132 | 4.01E-07 | 4375 | 6.19E-06 | TRUE | 0.005871716305 | TRUE | 0.004966297651 | TRUE | FALSE | -0.7672935347 |
| **met** | LV end diastolic volume \| Instance 2(participant - p24100_i2)_BSA_indexed_corrected | 4.345881106 | 0.3291024219 | 0.07954403739 | 1.65E-07 | 0.8288015993 | 0.07954403739 | 1.65E-07 | 4320 | 2.73E-06 | TRUE | 0.002414370822 | TRUE | 0.002042074841 | TRUE | FALSE | 4.345881106 |
| **met** | RV end diastolic volume \| Instance 2_BSA_indexed_corrected | 5.072946707 | 0.2248343143 | 0.08618196126 | 1.40E-08 | 0.8924497334 | 0.08618196126 | 1.40E-08 | 4320 | 2.81E-07 | TRUE | 0.0002048366237 | TRUE | 0.0001732508163 | TRUE | FALSE | 5.072946707 |
| **mfap5** | LV end diastolic volume \| Instance 2(participant - p24100_i2)_BSA_indexed_corrected | -3.129287151 | 0.1004945453 | -0.1213061354 | 8.21E-16 | 0.3872164575 | -0.1213061354 | 8.21E-16 | 4375 | 6.12E-14 | TRUE | 1.20E-11 | TRUE | 1.02E-11 | TRUE | FALSE | -3.129287151 |
| **mfap5** | LV end systolic volume \| Instance 2(participant - p24101_i2)_BSA_indexed_corrected | -1.323950786 | 0.2050744248 | -0.08841342256 | 4.69E-09 | 0.2255588158 | -0.08841342256 | 4.69E-09 | 4375 | 1.01E-07 | TRUE | 6.86E-05 | TRUE | 5.80E-05 | TRUE | FALSE | -1.323950786 |
| **mfap5** | RV end diastolic volume \| Instance 2_BSA_indexed_corrected | -3.112586959 | -0.007730435114 | -0.1121530756 | 1.01E-13 | 0.4170344171 | -0.1121530756 | 1.01E-13 | 4375 | 5.13E-12 | TRUE | 1.48E-09 | TRUE | 1.25E-09 | TRUE | FALSE | -3.112586959 |
| **mfap5** | RV end systolic volume \| Instance 2_BSA_indexed_corrected | -1.304792086 | 0.1118300672 | -0.07905750569 | 1.64E-07 | 0.2487980766 | -0.07905750569 | 1.64E-07 | 4375 | 2.72E-06 | TRUE | 0.002402456429 | TRUE | 0.002031997648 | TRUE | FALSE | -1.304792086 |
| **mfge8** | RV end diastolic volume \| Instance 2_BSA_indexed_corrected | -2.267105415 | 0.2188542119 | -0.08104629893 | 9.90E-08 | 0.4247361664 | -0.08104629893 | 9.90E-08 | 4311 | 1.72E-06 | TRUE | 0.001448141137 | TRUE | 0.001224837774 | TRUE | FALSE | -2.267105415 |
| **mia** | LV cardiac output \| Instance 2_BSA_indexed_corrected | -0.1179856665 | -0.01793504772 | -0.08489625731 | 2.13E-08 | 0.02102447709 | -0.08489625731 | 2.13E-08 | 4340 | 4.15E-07 | TRUE | 0.0003111524587 | TRUE | 0.0002631727496 | TRUE | FALSE | -0.1179856665 |
| **mia** | LV end diastolic volume \| Instance 2(participant - p24100_i2)_BSA_indexed_corrected | -2.560280043 | 0.390236502 | -0.08223858628 | 5.78E-08 | 0.4710786973 | -0.08223858628 | 5.78E-08 | 4340 | 1.04E-06 | TRUE | 0.0008459774345 | TRUE | 0.0007155277141 | TRUE | FALSE | -2.560280043 |
| **mmp7** | RV end diastolic volume \| Instance 2_BSA_indexed_corrected | -1.829698356 | -0.05179506282 | -0.08043592673 | 1.01E-07 | 0.3430667429 | -0.08043592673 | 1.01E-07 | 4370 | 1.76E-06 | TRUE | 0.001481802705 | TRUE | 0.001253308728 | TRUE | FALSE | -1.829698356 |
| **mmp8** | LV end diastolic volume \| Instance 2(participant - p24100_i2)_BSA_indexed_corrected | -1.365094201 | 0.2297380353 | -0.08739136904 | 7.53E-09 | 0.2357682772 | -0.08739136904 | 7.53E-09 | 4358 | 1.57E-07 | TRUE | 0.0001102281026 | TRUE | 9.32E-05 | TRUE | FALSE | -1.365094201 |
| **mmp8** | LV end systolic volume \| Instance 2(participant - p24101_i2)_BSA_indexed_corrected | -0.6677747833 | 0.2545305935 | -0.07368312989 | 1.12E-06 | 0.1369417475 | -0.07368312989 | 1.12E-06 | 4358 | 1.58E-05 | TRUE | 0.01637420405 | TRUE | 0.01384930179 | TRUE | FALSE | -0.6677747833 |
| **mmp8** | RV end diastolic volume \| Instance 2_BSA_indexed_corrected | -1.535358696 | 0.1070830962 | -0.09130290289 | 1.56E-09 | 0.253725173 | -0.09130290289 | 1.56E-09 | 4358 | 3.66E-08 | TRUE | 2.28E-05 | TRUE | 1.93E-05 | TRUE | FALSE | -1.535358696 |
| **mmp8** | RV end systolic volume \| Instance 2_BSA_indexed_corrected | -0.7137388481 | 0.1600587891 | -0.07143678748 | 2.35E-06 | 0.1509949761 | -0.07143678748 | 2.35E-06 | 4358 | 3.12E-05 | TRUE | 0.03440429444 | TRUE | 0.02909915224 | TRUE | FALSE | -0.7137388481 |
| **mmp9** | LV end diastolic volume \| Instance 2(participant - p24100_i2)_BSA_indexed_corrected | -1.853283234 | 0.1345546663 | -0.10162972 | 2.34E-11 | 0.2765867523 | -0.10162972 | 2.34E-11 | 4304 | 7.96E-10 | TRUE | 3.43E-07 | TRUE | 2.90E-07 | TRUE | FALSE | -1.853283234 |
| **mmp9** | LV end systolic volume \| Instance 2(participant - p24101_i2)_BSA_indexed_corrected | -0.8734058759 | 0.2120839313 | -0.0823399002 | 6.32E-08 | 0.1611733856 | -0.0823399002 | 6.32E-08 | 4304 | 1.13E-06 | TRUE | 0.0009245403775 | TRUE | 0.0007819762513 | TRUE | FALSE | -0.8734058759 |
| **mmp9** | RV end diastolic volume \| Instance 2_BSA_indexed_corrected | -2.230008388 | 0.01270082135 | -0.1135916275 | 7.77E-14 | 0.2973752875 | -0.1135916275 | 7.77E-14 | 4304 | 4.06E-12 | TRUE | 1.14E-09 | TRUE | 9.61E-10 | TRUE | FALSE | -2.230008388 |
| **mmp9** | RV end systolic volume \| Instance 2_BSA_indexed_corrected | -1.137270893 | 0.09059509203 | -0.09730750855 | 1.58E-10 | 0.1773439832 | -0.09730750855 | 1.58E-10 | 4304 | 4.55E-09 | TRUE | 2.32E-06 | TRUE | 1.96E-06 | TRUE | FALSE | -1.137270893 |
| **mnda** | LV end diastolic volume \| Instance 2(participant - p24100_i2)_BSA_indexed_corrected | -1.161761196 | 0.3155067688 | -0.1036973852 | 6.18E-12 | 0.1685045611 | -0.1036973852 | 6.18E-12 | 4375 | 2.34E-10 | TRUE | 9.04E-08 | TRUE | 7.64E-08 | TRUE | FALSE | -1.161761196 |
| **mnda** | LV end systolic volume \| Instance 2(participant - p24101_i2)_BSA_indexed_corrected | -0.5082095809 | 0.2955038597 | -0.07814528833 | 2.28E-07 | 0.09804381372 | -0.07814528833 | 2.28E-07 | 4375 | 3.69E-06 | TRUE | 0.003329641001 | TRUE | 0.002816210359 | TRUE | FALSE | -0.5082095809 |
| **mnda** | RV end diastolic volume \| Instance 2_BSA_indexed_corrected | -1.030227132 | 0.2101808247 | -0.08547444043 | 1.49E-08 | 0.1815995229 | -0.08547444043 | 1.49E-08 | 4375 | 2.99E-07 | TRUE | 0.0002184440049 | TRUE | 0.0001847599394 | TRUE | FALSE | -1.030227132 |
| **mog** | LV cardiac output \| Instance 2_BSA_indexed_corrected | -0.1500053968 | -0.02223635324 | -0.09785884787 | 8.79E-11 | 0.0230689351 | -0.09785884787 | 8.79E-11 | 4375 | 2.63E-09 | TRUE | 1.29E-06 | TRUE | 1.09E-06 | TRUE | FALSE | -0.1500053968 |
| **mog** | LV end diastolic volume \| Instance 2(participant - p24100_i2)_BSA_indexed_corrected | -2.618534208 | 0.3095591952 | -0.07624115609 | 4.45E-07 | 0.5178609247 | -0.07624115609 | 4.45E-07 | 4375 | 6.80E-06 | TRUE | 0.006504855748 | TRUE | 0.005501806992 | TRUE | FALSE | -2.618534208 |
| **mpo** | LV end diastolic volume \| Instance 2(participant - p24100_i2)_BSA_indexed_corrected | -2.269744599 | 0.2120320718 | -0.09437112641 | 5.36E-10 | 0.3647598672 | -0.09437112641 | 5.36E-10 | 4311 | 1.41E-08 | TRUE | 7.84E-06 | TRUE | 6.63E-06 | TRUE | FALSE | -2.269744599 |
| **mpo** | LV end systolic volume \| Instance 2(participant - p24101_i2)_BSA_indexed_corrected | -1.056821629 | 0.2517265591 | -0.0756395756 | 6.63E-07 | 0.2122356915 | -0.0756395756 | 6.63E-07 | 4311 | 9.78E-06 | TRUE | 0.00969255487 | TRUE | 0.008197962909 | TRUE | FALSE | -1.056821629 |
| **mpo** | RV end diastolic volume \| Instance 2_BSA_indexed_corrected | -2.338865796 | 0.1296320675 | -0.09033449523 | 2.82E-09 | 0.392811481 | -0.09033449523 | 2.82E-09 | 4311 | 6.33E-08 | TRUE | 4.13E-05 | TRUE | 3.49E-05 | TRUE | FALSE | -2.338865796 |
| **mpo** | RV end systolic volume \| Instance 2_BSA_indexed_corrected | -1.194107269 | 0.1578263377 | -0.07756207846 | 3.42E-07 | 0.2338273892 | -0.07756207846 | 3.42E-07 | 4311 | 5.33E-06 | TRUE | 0.005000937957 | TRUE | 0.004229793324 | TRUE | FALSE | -1.194107269 |
| **msr1** | LV end diastolic volume \| Instance 2(participant - p24100_i2)_BSA_indexed_corrected | -1.861179026 | 0.078242285 | -0.0830531385 | 3.93E-08 | 0.3381707373 | -0.0830531385 | 3.93E-08 | 4363 | 7.28E-07 | TRUE | 0.0005754080836 | TRUE | 0.0004866801571 | TRUE | FALSE | -1.861179026 |
| **msr1** | RV end diastolic volume \| Instance 2_BSA_indexed_corrected | -2.388116955 | -0.1156629553 | -0.09897905826 | 5.67E-11 | 0.3635644263 | -0.09897905826 | 5.67E-11 | 4363 | 1.79E-09 | TRUE | 8.30E-07 | TRUE | 7.02E-07 | TRUE | FALSE | -2.388116955 |
| **msr1** | RV end systolic volume \| Instance 2_BSA_indexed_corrected | -1.026708743 | 0.0658912499 | -0.07161099703 | 2.19E-06 | 0.2165499418 | -0.07161099703 | 2.19E-06 | 4363 | 2.93E-05 | TRUE | 0.03207211569 | TRUE | 0.02712659545 | TRUE | FALSE | -1.026708743 |
| **muc13** | LV end diastolic volume \| Instance 2(participant - p24100_i2)_BSA_indexed_corrected | -2.295446381 | 0.187246247 | -0.09779390631 | 1.48E-10 | 0.3573569213 | -0.09779390631 | 1.48E-10 | 4275 | 4.29E-09 | TRUE | 2.16E-06 | TRUE | 1.83E-06 | TRUE | FALSE | -2.295446381 |
| **muc13** | RV end diastolic volume \| Instance 2_BSA_indexed_corrected | -2.038017749 | 0.08699706402 | -0.0807467359 | 1.25E-07 | 0.3848540815 | -0.0807467359 | 1.25E-07 | 4275 | 2.10E-06 | TRUE | 0.001823126284 | TRUE | 0.001542000211 | TRUE | FALSE | -2.038017749 |
| **mvk** | LV end diastolic volume \| Instance 2(participant - p24100_i2)_BSA_indexed_corrected | -1.312937172 | 0.2750388088 | -0.09087979471 | 2.54E-09 | 0.2198631164 | -0.09087979471 | 2.54E-09 | 4284 | 5.75E-08 | TRUE | 3.72E-05 | TRUE | 3.14E-05 | TRUE | FALSE | -1.312937172 |
| **mvk** | RV end diastolic volume \| Instance 2_BSA_indexed_corrected | -1.469762489 | 0.1518810943 | -0.09443364738 | 5.91E-10 | 0.2367837643 | -0.09443364738 | 5.91E-10 | 4284 | 1.55E-08 | TRUE | 8.64E-06 | TRUE | 7.31E-06 | TRUE | FALSE | -1.469762489 |
| **mvk** | RV end systolic volume \| Instance 2_BSA_indexed_corrected | -0.7287440736 | 0.1642419536 | -0.07887243857 | 2.35E-07 | 0.1407574446 | -0.07887243857 | 2.35E-07 | 4284 | 3.80E-06 | TRUE | 0.003444505837 | TRUE | 0.002913363037 | TRUE | FALSE | -0.7287440736 |
| **mzb1** | LV end diastolic volume \| Instance 2(participant - p24100_i2)_BSA_indexed_corrected | -1.917571384 | 0.2002036174 | -0.08352820728 | 3.85E-08 | 0.3481825627 | -0.08352820728 | 3.85E-08 | 4319 | 7.17E-07 | TRUE | 0.0005638285302 | TRUE | 0.0004768861708 | TRUE | FALSE | -1.917571384 |

| | | | | | | | | | | | | | | | | | |
|---|---|---|---|---|---|---|---|---|---|---|---|---|---|---|---|---|---|
| mzb1 | RV end diastolic volume \| Instance 2_BSA_indexed_corrected | -2.675584229 | 0.03692952807 | -0.1083294471 | 9.45E-13 | 0.3736953767 | -0.1083294471 | 9.45E-13 | 4319 | 4.08E-11 | TRUE | 1.38E-08 | TRUE | 1.17E-08 | TRUE | FALSE | -2.675584229 |
| mzb1 | RV end systolic volume \| Instance 2_BSA_indexed_corrected | -1.274594456 | 0.1077795927 | -0.08683175168 | 1.09E-08 | 0.2225660911 | -0.08683175168 | 1.09E-08 | 4319 | 2.24E-07 | TRUE | 0.000159884306 | TRUE | 0.000135230146 | TRUE | FALSE | -1.274594456 |
| nadk | LV end diastolic volume \| Instance 2(participant - p24100_i2)_BSA_indexed_corrected | -2.293156316 | 0.1343321381 | -0.1123715457 | 9.04E-14 | 0.3066395086 | -0.1123715457 | 9.04E-14 | 4375 | 4.64E-12 | TRUE | 1.32E-09 | TRUE | 1.12E-09 | TRUE | FALSE | -2.293156316 |
| nadk | LV end systolic volume \| Instance 2(participant - p24101_i2)_BSA_indexed_corrected | -1.165945671 | 0.2007233581 | -0.09842604162 | 6.83E-11 | 0.1782644098 | -0.09842604162 | 6.83E-11 | 4375 | 2.10E-09 | TRUE | 1.00E-06 | TRUE | 8.46E-07 | TRUE | FALSE | -1.165945671 |
| nadk | RV end diastolic volume \| Instance 2_BSA_indexed_corrected | -2.129653181 | 0.0403517183 | -0.09700272218 | 1.28E-10 | 0.3304321498 | -0.09700272218 | 1.28E-10 | 4375 | 3.75E-09 | TRUE | 1.87E-06 | TRUE | 1.58E-06 | TRUE | FALSE | -2.129653181 |
| nadk | RV end systolic volume \| Instance 2_BSA_indexed_corrected | -1.288013247 | 0.0942922078 | -0.09865235753 | 6.18E-11 | 0.1964714419 | -0.09865235753 | 6.18E-11 | 4375 | 1.94E-09 | TRUE | 9.04E-07 | TRUE | 7.65E-07 | TRUE | FALSE | -1.288013247 |
| nbl1 | LV cardiac output \| Instance 2_BSA_indexed_corrected | -0.1607287726 | -0.02451673442 | -0.08021006131 | 1.22E-07 | 0.03032623127 | -0.08021006131 | 1.22E-07 | 4340 | 2.06E-06 | TRUE | 0.001778396742 | TRUE | 0.001504167964 | TRUE | FALSE | -0.1607287726 |
| nbl1 | LV end diastolic volume \| Instance 2(participant - p24100_i2)_BSA_indexed_corrected | -3.602944027 | 0.2437329402 | -0.08026414908 | 1.19E-07 | 0.6793407634 | -0.08026414908 | 1.19E-07 | 4340 | 2.02E-06 | TRUE | 0.001743912747 | TRUE | 0.001475001401 | TRUE | FALSE | -3.602944027 |
| nbl1 | RV end diastolic volume \| Instance 2_BSA_indexed_corrected | -4.514684115 | 0.1100771043 | -0.09351184175 | 6.74E-10 | 0.7298082982 | -0.09351184175 | 6.74E-10 | 4340 | 1.74E-08 | TRUE | 9.85E-06 | TRUE | 8.33E-06 | TRUE | FALSE | -4.514684115 |
| nbn | LV end diastolic volume \| Instance 2(participant - p24100_i2)_BSA_indexed_corrected | -1.957228876 | 0.3464993725 | -0.08582253553 | 1.84E-08 | 0.3472255088 | -0.08582253553 | 1.84E-08 | 4284 | 3.63E-07 | TRUE | 0.0002697945374 | TRUE | 0.0002281922197 | TRUE | FALSE | -1.957228876 |
| nbn | RV end diastolic volume \| Instance 2_BSA_indexed_corrected | -1.81298303 | 0.2348866748 | -0.07379206732 | 1.33E-06 | 0.3744338244 | -0.07379206732 | 1.33E-06 | 4284 | 1.85E-05 | TRUE | 0.01947512454 | TRUE | 0.01647206034 | TRUE | FALSE | -1.81298303 |
| ncf2 | LV end diastolic volume \| Instance 2(participant - p24100_i2)_BSA_indexed_corrected | -1.300159634 | 0.2912112699 | -0.09993379908 | 4.41E-11 | 0.1967937644 | -0.09993379908 | 4.41E-11 | 4329 | 1.43E-09 | TRUE | 6.45E-07 | TRUE | 5.45E-07 | TRUE | FALSE | -1.300159634 |
| ncf2 | LV end systolic volume \| Instance 2(participant - p24101_i2)_BSA_indexed_corrected | -0.6230247857 | 0.2811400855 | -0.08260780597 | 5.24E-08 | 0.1142625433 | -0.08260780597 | 5.24E-08 | 4329 | 9.49E-07 | TRUE | 0.0007667908133 | TRUE | 0.0006485516699 | TRUE | FALSE | -0.6230247857 |
| ncf2 | RV end diastolic volume \| Instance 2_BSA_indexed_corrected | -1.220590338 | 0.1972550636 | -0.08716853836 | 9.22E-09 | 0.2120608288 | -0.08716853836 | 9.22E-09 | 4329 | 1.90E-07 | TRUE | 0.0001348413937 | TRUE | 0.0001140488508 | TRUE | FALSE | -1.220590338 |
| ncf2 | RV end systolic volume \| Instance 2_BSA_indexed_corrected | -0.6750880241 | 0.1860500445 | -0.08107221867 | 9.22E-08 | 0.1261719869 | -0.08107221867 | 9.22E-08 | 4329 | 1.61E-06 | TRUE | 0.001349263379 | TRUE | 0.001141206966 | TRUE | FALSE | -0.6750880241 |
| nectin2 | RV end diastolic volume \| Instance 2_BSA_indexed_corrected | -3.348356938 | 0.06161702639 | -0.07274653849 | 1.46E-06 | 0.694267972 | -0.07274653849 | 1.46E-06 | 4374 | 2.01E-05 | TRUE | 0.02140758626 | TRUE | 0.01810653646 | TRUE | FALSE | -3.348356938 |
| nectin4 | LV cardiac output \| Instance 2_BSA_indexed_corrected | -0.1469348411 | -0.02158312748 | -0.08175222908 | 6.92E-08 | 0.02719720353 | -0.08175222908 | 6.92E-08 | 4340 | 1.23E-06 | TRUE | 0.001012562814 | TRUE | 0.0008564256279 | TRUE | FALSE | -0.1469348411 |
| nectin4 | LV end diastolic volume \| Instance 2(participant - p24100_i2)_BSA_indexed_corrected | -2.987385978 | 0.3140916859 | -0.07419848271 | 9.91E-07 | 0.6096109388 | -0.07419848271 | 9.91E-07 | 4340 | 1.42E-05 | TRUE | 0.01449695779 | TRUE | 0.0122615269 | TRUE | FALSE | -2.987385978 |
| nectin4 | RV end diastolic volume \| Instance 2_BSA_indexed_corrected | -3.893261591 | 0.1959904297 | -0.08990677564 | 2.97E-09 | 0.6548080434 | -0.08990677564 | 2.97E-09 | 4340 | 6.64E-08 | TRUE | 4.35E-05 | TRUE | 3.68E-05 | TRUE | FALSE | -3.893261591 |
| nell2 | LV end diastolic volume \| Instance 2(participant - p24100_i2)_BSA_indexed_corrected | -2.765495813 | 0.3928466154 | -0.07068697355 | 3.29E-06 | 0.5937505377 | -0.07068697355 | 3.29E-06 | 4322 | 4.28E-05 | TRUE | 0.04819322946 | TRUE | 0.04076183348 | TRUE | FALSE | -2.765495813 |
| nfkbie | LV end diastolic volume \| Instance 2(participant - p24100_i2)_BSA_indexed_corrected | -1.769143729 | 0.3285838847 | -0.07732120254 | 3.05E-07 | 0.3449631087 | -0.07732120254 | 3.05E-07 | 4375 | 4.81E-06 | TRUE | 0.004457623637 | TRUE | 0.003770258072 | TRUE | FALSE | -1.769143729 |
| nmnat1 | LV end diastolic volume \| Instance 2(participant - p24100_i2)_BSA_indexed_corrected | -1.542174146 | 0.2902245089 | -0.1031169799 | 8.10E-12 | 0.2249532024 | -0.1031169799 | 8.10E-12 | 4375 | 3.01E-10 | TRUE | 1.18E-07 | TRUE | 1.00E-07 | TRUE | FALSE | -1.542174146 |
| nmnat1 | LV end systolic volume \| Instance 2(participant - p24101_i2)_BSA_indexed_corrected | -0.7373262413 | 0.2818910861 | -0.08493084959 | 1.84E-08 | 0.1308074471 | -0.08493084959 | 1.84E-08 | 4375 | 3.63E-07 | TRUE | 0.0002695024913 | TRUE | 0.0002279452071 | TRUE | FALSE | -0.7373262413 |
| nmnat1 | RV end diastolic volume \| Instance 2_BSA_indexed_corrected | -1.362390717 | 0.1879178842 | -0.08467413858 | 2.03E-08 | 0.242436865 | -0.08467413858 | 2.03E-08 | 4375 | 3.98E-07 | TRUE | 0.0002974737077 | TRUE | 0.000251603262 | TRUE | FALSE | -1.362390717 |
| nmnat1 | RV end systolic volume \| Instance 2_BSA_indexed_corrected | -0.7639153885 | 0.1860181083 | -0.07983741553 | 1.24E-07 | 0.1442316331 | -0.07983741553 | 1.24E-07 | 4375 | 2.09E-06 | TRUE | 0.001812312064 | TRUE | 0.001532853544 | TRUE | FALSE | -0.7639153885 |
| nomo1 | LV end diastolic volume \| Instance 2(participant - p24100_i2)_BSA_indexed_corrected | -6.066800955 | 0.04120050401 | -0.1335299662 | 1.32E-18 | 0.6859398544 | -0.1335299662 | 1.32E-18 | 4311 | 1.45E-16 | TRUE | 1.93E-14 | TRUE | 1.63E-14 | TRUE | FALSE | -6.066800955 |
| nomo1 | LV end systolic volume \| Instance 2(participant - p24101_i2)_BSA_indexed_corrected | -2.879710257 | 0.16943358 | -0.1091071041 | 6.81E-13 | 0.3996750232 | -0.1091071041 | 6.81E-13 | 4311 | 3.02E-11 | TRUE | 9.97E-09 | TRUE | 8.43E-09 | TRUE | FALSE | -2.879710257 |
| nomo1 | RV end diastolic volume \| Instance 2_BSA_indexed_corrected | -6.743556017 | -0.07104744188 | -0.137877733 | 9.58E-20 | 0.7379701896 | -0.137877733 | 9.58E-20 | 4311 | 1.35E-17 | TRUE | 1.40E-15 | TRUE | 1.19E-15 | TRUE | FALSE | -6.743556017 |
| nomo1 | RV end systolic volume \| Instance 2_BSA_indexed_corrected | -3.011367702 | 0.07698703355 | -0.1035444271 | 9.43E-12 | 0.4406645436 | -0.1035444271 | 9.43E-12 | 4311 | 3.42E-10 | TRUE | 1.38E-07 | TRUE | 1.17E-07 | TRUE | FALSE | -3.011367702 |
| nos3 | LV end diastolic volume \| Instance 2(participant - p24100_i2)_BSA_indexed_corrected | -2.137161476 | 0.2092858049 | -0.099729415 | 6.36E-11 | 0.3261948275 | -0.099729415 | 6.36E-11 | 4275 | 1.98E-09 | TRUE | 9.30E-07 | TRUE | 7.87E-07 | TRUE | FALSE | -2.137161476 |
| nos3 | LV end systolic volume \| Instance 2(participant - p24101_i2)_BSA_indexed_corrected | -1.056914819 | 0.2257941407 | -0.08513272231 | 2.48E-08 | 0.1892332711 | -0.08513272231 | 2.48E-08 | 4275 | 4.80E-07 | TRUE | 0.0003626175695 | TRUE | 0.0003067019402 | TRUE | FALSE | -1.056914819 |
| nos3 | RV end diastolic volume \| Instance 2_BSA_indexed_corrected | -2.396127072 | 0.07495098759 | -0.1039843909 | 9.40E-12 | 0.3506020905 | -0.1039843909 | 9.40E-12 | 4275 | 3.42E-10 | TRUE | 1.38E-07 | TRUE | 1.16E-07 | TRUE | FALSE | -2.396127072 |
| nos3 | RV end systolic volume \| Instance 2_BSA_indexed_corrected | -1.148850587 | 0.1253263483 | -0.08398155465 | 3.82E-08 | 0.2085337004 | -0.08398155465 | 3.82E-08 | 4275 | 7.11E-07 | TRUE | 0.0005584230518 | TRUE | 0.0004723142172 | TRUE | FALSE | -1.148850587 |
| notch3 | LV end diastolic volume \| Instance 2(participant - p24100_i2)_BSA_indexed_corrected | 3.614874944 | 0.485521391 | 0.1121006222 | 1.04E-13 | 0.4845620896 | 0.1121006222 | 1.04E-13 | 4375 | 5.25E-12 | TRUE | 1.52E-09 | TRUE | 1.28E-09 | TRUE | FALSE | 3.614874944 |
| notch3 | LV end systolic volume \| Instance 2(participant - p24101_i2)_BSA_indexed_corrected | 1.802532935 | 0.3779851543 | 0.09629571565 | 1.74E-10 | 0.2817497465 | 0.09629571565 | 1.74E-10 | 4375 | 4.97E-09 | TRUE | 2.55E-06 | TRUE | 2.16E-06 | TRUE | FALSE | 1.802532935 |
| notch3 | RV end diastolic volume \| Instance 2_BSA_indexed_corrected | 3.644652824 | 0.3770403077 | 0.1050565791 | 3.26E-12 | 0.5217148743 | 0.1050565791 | 3.26E-12 | 4375 | 1.29E-10 | TRUE | 4.77E-08 | TRUE | 4.03E-08 | TRUE | FALSE | 3.644652824 |
| notch3 | RV end systolic volume \| Instance 2_BSA_indexed_corrected | 1.9074414 | 0.287040279 | 0.09245499809 | 8.97E-10 | 0.310646836 | 0.09245499809 | 8.97E-10 | 4375 | 2.22E-08 | TRUE | 1.31E-05 | TRUE | 1.11E-05 | TRUE | FALSE | 1.9074414 |
| npdc1 | LV end diastolic volume \| Instance 2(participant - p24100_i2)_BSA_indexed_corrected | -5.476129991 | 0.05244074854 | -0.1359136229 | 1.73E-19 | 0.6036319992 | -0.1359136229 | 1.73E-19 | 4375 | 2.28E-17 | TRUE | 2.53E-15 | TRUE | 2.14E-15 | TRUE | FALSE | -5.476129991 |
| npdc1 | LV end systolic volume \| Instance 2(participant - p24101_i2)_BSA_indexed_corrected | -2.738654131 | 0.1615922765 | -0.1170942149 | 7.87E-15 | 0.3512481882 | -0.1170942149 | 7.87E-15 | 4375 | 4.88E-13 | TRUE | 1.15E-10 | TRUE | 9.74E-11 | TRUE | FALSE | -2.738654131 |
| npdc1 | RV end diastolic volume \| Instance 2_BSA_indexed_corrected | -5.914097424 | -0.08117106483 | -0.1364363394 | 1.25E-19 | 0.6493643055 | -0.1364363394 | 1.25E-19 | 4375 | 1.70E-17 | TRUE | 1.84E-15 | TRUE | 1.55E-15 | TRUE | FALSE | -5.914097424 |
| npdc1 | RV end systolic volume \| Instance 2_BSA_indexed_corrected | -3.108855937 | 0.04648222154 | -0.1206019366 | 1.20E-15 | 0.3869679652 | -0.1206019366 | 1.20E-15 | 4375 | 8.72E-14 | TRUE | 1.76E-11 | TRUE | 1.49E-11 | TRUE | FALSE | -3.108855937 |
| nppb | LV end diastolic volume \| Instance 2(participant - p24100_i2)_BSA_indexed_corrected | 0.7894323584 | 0.5346435912 | 0.09502301709 | 3.02E-10 | 0.1250624005 | 0.09502301709 | 3.02E-10 | 4375 | 8.37E-09 | TRUE | 4.42E-06 | TRUE | 3.74E-06 | TRUE | FALSE | 0.7894323584 |
| nppb | LV end systolic volume \| Instance 2(participant - p24101_i2)_BSA_indexed_corrected | 0.3970863572 | 0.4032713527 | 0.08233943236 | 4.93E-08 | 0.07267919938 | 0.08233943236 | 4.93E-08 | 4375 | 8.99E-07 | TRUE | 0.0007207269808 | TRUE | 0.0006095918804 | TRUE | FALSE | 0.3970863572 |
| nppc | LV cardiac output \| Instance 2_BSA_indexed_corrected | -0.07141081703 | -0.02036468097 | -0.07056022618 | 3.34E-06 | 0.01534179829 | -0.07056022618 | 3.34E-06 | 4332 | 4.33E-05 | TRUE | 0.04889698691 | TRUE | 0.04135707153 | TRUE | FALSE | -0.07141081703 |
| nppc | LV end diastolic volume \| Instance 2(participant - p24100_i2)_BSA_indexed_corrected | -3.578171215 | 0.4154967621 | -0.1570963703 | 2.44E-25 | 0.3418414691 | -0.1570963703 | 2.44E-25 | 4332 | 7.77E-23 | TRUE | 3.57E-21 | TRUE | 3.02E-21 | TRUE | FALSE | -3.578171215 |
| nppc | LV end systolic volume \| Instance 2(participant - p24101_i2)_BSA_indexed_corrected | -1.6224172 | 0.343471058 | -0.1227943483 | 5.04E-16 | 0.1992695935 | -0.1227943483 | 5.04E-16 | 4332 | 3.90E-14 | TRUE | 7.38E-12 | TRUE | 6.24E-12 | TRUE | FALSE | -1.6224172 |
| nppc | RV end diastolic volume \| Instance 2_BSA_indexed_corrected | -3.447939385 | 0.3096093503 | -0.1407315266 | 1.32E-20 | 0.3686213084 | -0.1407315266 | 1.32E-20 | 4332 | 2.02E-18 | TRUE | 1.94E-16 | TRUE | 1.64E-16 | TRUE | FALSE | -3.447939385 |
| nppc | RV end systolic volume \| Instance 2_BSA_indexed_corrected | -1.57304679 | 0.2505280462 | -0.1078529352 | 1.10E-12 | 0.2203560637 | -0.1078529352 | 1.10E-12 | 4332 | 4.65E-11 | TRUE | 1.61E-08 | TRUE | 1.36E-08 | TRUE | FALSE | -1.57304679 |
| nptxr | LV cardiac output \| Instance 2_BSA_indexed_corrected | -0.1090253222 | -0.01494856522 | -0.07497819421 | 6.87E-07 | 0.02192696261 | -0.07497819421 | 6.87E-07 | 4375 | 1.01E-05 | TRUE | 0.01005543641 | TRUE | 0.008504888113 | TRUE | FALSE | -0.1090253222 |
| npy | LV end diastolic volume \| Instance 2(participant - p24100_i2)_BSA_indexed_corrected | -2.116987861 | 0.08921293239 | -0.1291761724 | 9.68E-18 | 0.2457492616 | -0.1291761724 | 9.68E-18 | 4375 | 9.25E-16 | TRUE | 1.42E-13 | TRUE | 1.20E-13 | TRUE | FALSE | -2.116987861 |
| npy | LV end systolic volume \| Instance 2(participant - p24101_i2)_BSA_indexed_corrected | -0.8409162882 | 0.2071234951 | -0.08839461725 | 4.73E-09 | 0.1432959247 | -0.08839461725 | 4.73E-09 | 4375 | 1.02E-07 | TRUE | 6.91E-05 | TRUE | 5.85E-05 | TRUE | FALSE | -0.8409162882 |
| npy | RV end diastolic volume \| Instance 2_BSA_indexed_corrected | -1.949423324 | 0.0005208714314 | -0.1105662414 | 2.24E-13 | 0.2649859891 | -0.1105662414 | 2.24E-13 | 4375 | 1.07E-11 | TRUE | 3.28E-09 | TRUE | 2.77E-09 | TRUE | FALSE | -1.949423324 |
| npy | RV end systolic volume \| Instance 2_BSA_indexed_corrected | -0.7613493172 | 0.1222481119 | -0.07261267742 | 1.53E-06 | 0.1581371158 | -0.07261267742 | 1.53E-06 | 4375 | 2.09E-05 | TRUE | 0.02231961072 | TRUE | 0.01887792675 | TRUE | FALSE | -0.7613493172 |
| nrcam | LV end diastolic volume \| Instance 2(participant - p24100_i2)_BSA_indexed_corrected | -4.294213904 | 0.3016245212 | -0.1054966468 | 2.64E-12 | 0.6121036658 | -0.1054966468 | 2.64E-12 | 4375 | 1.06E-10 | TRUE | 3.87E-08 | TRUE | 3.27E-08 | TRUE | FALSE | -4.294213904 |
| nrcam | LV end systolic volume \| Instance 2(participant - p24101_i2)_BSA_indexed_corrected | -1.753014743 | 0.2909327562 | -0.07419071584 | 8.99E-07 | 0.3563261796 | -0.07419071584 | 8.99E-07 | 4375 | 1.29E-05 | TRUE | 0.01314736195 | TRUE | 0.01112003874 | TRUE | FALSE | -1.753014743 |
| nrcam | RV end diastolic volume \| Instance 2_BSA_indexed_corrected | -4.052291364 | 0.1949470868 | -0.09253541782 | 8.68E-10 | 0.6593796496 | -0.09253541782 | 8.68E-10 | 4375 | 2.16E-08 | TRUE | 1.27E-05 | TRUE | 1.07E-05 | TRUE | FALSE | -4.052291364 |
| ntprobnp | LV end diastolic volume \| Instance 2(participant - p24100_i2)_BSA_indexed_corrected | 1.494304136 | 0.5607789352 | 0.1318977649 | 1.95E-18 | 0.1698247023 | 0.1318977649 | 1.95E-18 | 4375 | 2.07E-16 | TRUE | 2.86E-14 | TRUE | 2.42E-14 | TRUE | FALSE | 1.494304136 |
| ntprobnp | LV end systolic volume \| Instance 2(participant - p24101_i2)_BSA_indexed_corrected | 0.6888804759 | 0.4076917873 | 0.1047493471 | 3.77E-12 | 0.09890244153 | 0.1047493471 | 3.77E-12 | 4375 | 1.48E-10 | TRUE | 5.51E-08 | TRUE | 4.66E-08 | TRUE | FALSE | 0.6888804759 |
| ntprobnp | RV end diastolic volume \| Instance 2_BSA_indexed_corrected | 0.9644835194 | 0.3775415433 | 0.07913080664 | 1.60E-07 | 0.1837365245 | 0.07913080664 | 1.60E-07 | 4375 | 2.65E-06 | TRUE | 0.00233990433 | TRUE | 0.001979091083 | TRUE | FALSE | 0.9644835194 |
| ntrk3 | LV end diastolic volume \| Instance 2(participant - p24100_i2)_BSA_indexed_corrected | 4.736868174 | 0.3289004199 | 0.09757249138 | 1.06E-10 | 0.7316335805 | 0.09757249138 | 1.06E-10 | 4363 | 3.14E-09 | TRUE | 1.55E-06 | TRUE | 1.31E-06 | TRUE | FALSE | 4.736868174 |
| ntrk3 | LV end systolic volume \| Instance 2(participant - p24101_i2)_BSA_indexed_corrected | 2.281848242 | 0.2993683626 | 0.08104066095 | 8.31E-08 | 0.4249715945 | 0.08104066095 | 8.31E-08 | 4363 | 1.46E-06 | TRUE | 0.001215760751 | TRUE | 0.001028290443 | TRUE | FALSE | 2.281848242 |
| ntrk3 | RV end diastolic volume \| Instance 2_BSA_indexed_corrected | 6.243398681 | 0.2047262859 | 0.1194474796 | 2.45E-15 | 0.785833481 | 0.1194474796 | 2.45E-15 | 4363 | 1.67E-13 | TRUE | 3.59E-11 | TRUE | 3.04E-11 | TRUE | FALSE | 6.243398681 |
| ntrk3 | RV end systolic volume \| Instance 2_BSA_indexed_corrected | 3.10132752 | 0.2005191927 | 0.09985005067 | 3.84E-11 | 0.4679834236 | 0.09985005067 | 3.84E-11 | 4363 | 1.26E-09 | TRUE | 5.62E-07 | TRUE | 4.75E-07 | TRUE | FALSE | 3.10132752 |
| ogn | LV end diastolic volume \| Instance 2(participant - p24100_i2)_BSA_indexed_corrected | -2.785867053 | 0.07924775748 | -0.1014269732 | 2.48E-11 | 0.4162681835 | -0.1014269732 | 2.48E-11 | 4311 | 8.31E-10 | TRUE | 3.62E-07 | TRUE | 3.06E-07 | TRUE | FALSE | -2.785867053 |
| ogn | LV end systolic volume \| Instance 2(participant - p24101_i2)_BSA_indexed_corrected | -1.146585256 | 0.2042637767 | -0.07185957984 | 2.32E-06 | 0.2424427616 | -0.07185957984 | 2.32E-06 | 4311 | 3.09E-05 | TRUE | 0.03401396844 | TRUE | 0.0287690145 | TRUE | FALSE | -1.146585256 |
| ogn | RV end diastolic volume \| Instance 2_BSA_indexed_corrected | -3.446195479 | -0.06211466133 | -0.1165518465 | 1.63E-14 | 0.4473654945 | -0.1165518465 | 1.63E-14 | 4311 | 9.67E-13 | TRUE | 2.39E-10 | TRUE | 2.02E-10 | TRUE | FALSE | -3.446195479 |
| ogn | RV end systolic volume \| Instance 2_BSA_indexed_corrected | -1.371135335 | 0.09698717443 | -0.07798612653 | 2.95E-07 | 0.2670238417 | -0.07798612653 | 2.95E-07 | 4311 | 4.67E-06 | TRUE | 0.004312651541 | TRUE | 0.003647640673 | TRUE | FALSE | -1.371135335 |
| olr1 | LV end diastolic volume \| Instance 2(participant - p24100_i2)_BSA_indexed_corrected | -1.813246084 | 0.2528634214 | -0.09251024492 | 1.07E-09 | 0.2966585816 | -0.09251024492 | 1.07E-09 | 4330 | 2.63E-08 | TRUE | 1.56E-05 | TRUE | 1.32E-05 | TRUE | FALSE | -1.813246084 |

| | | | | | | | | | | | | | | | | | |
|---|---|---|---|---|---|---|---|---|---|---|---|---|---|---|---|---|---|
| **olr1** | LV end systolic volume \| Instance 2(participant - p24101_i2)_BSA_indexed_corrected | -0.9104374916 | 0.2646326293 | -0.0800392951 | 1.34E-07 | 0.1723485641 | -0.0800392951 | 1.34E-07 | 4330 | 2.25E-06 | TRUE | 0.001955544167 | TRUE | 0.001653999256 | TRUE | FALSE | -0.9104374916 |
| **olr1** | RV end diastolic volume \| Instance 2_BSA_indexed_corrected | -2.078481629 | 0.1412736906 | -0.09850376287 | 8.26E-11 | 0.3191776529 | -0.09850376287 | 8.26E-11 | 4330 | 2.50E-09 | TRUE | 1.21E-06 | TRUE | 1.02E-06 | TRUE | FALSE | -2.078481629 |
| **olr1** | RV end systolic volume \| Instance 2_BSA_indexed_corrected | -1.055213606 | 0.173494534 | -0.08410392955 | 2.98E-08 | 0.1900373764 | -0.08410392955 | 2.98E-08 | 4330 | 5.65E-07 | TRUE | 0.0004362612161 | TRUE | 0.0003689897365 | TRUE | FALSE | -1.055213606 |
| **omd** | LV end systolic volume \| Instance 2(participant - p24101_i2)_BSA_indexed_corrected | 1.065639517 | 0.3278460642 | 0.07113445119 | 2.85E-06 | 0.227345771 | 0.07113445119 | 2.85E-06 | 4322 | 3.73E-05 | TRUE | 0.04174657252 | TRUE | 0.03530925104 | TRUE | FALSE | 1.065639517 |
| **osm** | LV end diastolic volume \| Instance 2(participant - p24100_i2)_BSA_indexed_corrected | -1.926394924 | 0.09653975622 | -0.1229657412 | 5.40E-16 | 0.2368451408 | -0.1229657412 | 5.40E-16 | 4311 | 4.16E-14 | TRUE | 7.90E-12 | TRUE | 6.68E-12 | TRUE | FALSE | -1.926394924 |
| **osm** | LV end systolic volume \| Instance 2(participant - p24101_i2)_BSA_indexed_corrected | -0.9679518355 | 0.1815892151 | -0.1065428424 | 2.33E-12 | 0.1376138688 | -0.1065428424 | 2.33E-12 | 4311 | 9.43E-11 | TRUE | 3.40E-08 | TRUE | 2.88E-08 | TRUE | FALSE | -0.9679518355 |
| **osm** | RV end diastolic volume \| Instance 2_BSA_indexed_corrected | -2.292339184 | -0.05924488566 | -0.1360398927 | 2.93E-19 | 0.2543126634 | -0.1360398927 | 2.93E-19 | 4311 | 3.54E-17 | TRUE | 4.29E-15 | TRUE | 3.63E-15 | TRUE | FALSE | -2.292339184 |
| **osm** | RV end systolic volume \| Instance 2_BSA_indexed_corrected | -1.160097114 | 0.06362896272 | -0.1155809802 | 2.70E-14 | 0.1518795542 | -0.1155809802 | 2.70E-14 | 4311 | 1.53E-12 | TRUE | 3.94E-10 | TRUE | 3.34E-10 | TRUE | FALSE | -1.160097114 |
| **oxt** | LV end diastolic volume \| Instance 2(participant - p24100_i2)_BSA_indexed_corrected | -1.203521038 | 0.3326822491 | -0.1594995057 | 2.97E-26 | 0.1127990838 | -0.1594995057 | 2.97E-26 | 4363 | 1.09E-23 | TRUE | 4.34E-22 | TRUE | 3.67E-22 | TRUE | FALSE | -1.203521038 |
| **oxt** | LV end systolic volume \| Instance 2(participant - p24101_i2)_BSA_indexed_corrected | -0.593531231 | 0.3008286947 | -0.1356217526 | 2.31E-19 | 0.06565834724 | -0.1356217526 | 2.31E-19 | 4363 | 2.94E-17 | TRUE | 3.39E-15 | TRUE | 2.86E-15 | TRUE | FALSE | -0.593531231 |
| **oxt** | RV end diastolic volume \| Instance 2_BSA_indexed_corrected | -1.336150777 | 0.2162769677 | -0.1644678253 | 7.84E-28 | 0.1213463761 | -0.1644678253 | 7.84E-28 | 4363 | 3.28E-25 | TRUE | 1.15E-23 | TRUE | 9.70E-24 | TRUE | FALSE | -1.336150777 |
| **oxt** | RV end systolic volume \| Instance 2_BSA_indexed_corrected | -0.636823489 | 0.2069627442 | -0.1319135141 | 2.15E-18 | 0.07246442673 | -0.1319135141 | 2.15E-18 | 4363 | 2.26E-16 | TRUE | 3.15E-14 | TRUE | 2.66E-14 | TRUE | FALSE | -0.636823489 |
| **p4hb** | LV end diastolic volume \| Instance 2(participant - p24100_i2)_BSA_indexed_corrected | -2.528798265 | 0.2604929297 | -0.08969539142 | 3.59E-09 | 0.4275631364 | -0.08969539142 | 3.59E-09 | 4315 | 7.94E-08 | TRUE | 5.25E-05 | TRUE | 4.44E-05 | TRUE | FALSE | -2.528798265 |
| **p4hb** | RV end diastolic volume \| Instance 2_BSA_indexed_corrected | -2.789202716 | 0.1455087366 | -0.09200466967 | 1.41E-09 | 0.4596576954 | -0.09200466967 | 1.41E-09 | 4315 | 3.35E-08 | TRUE | 2.06E-05 | TRUE | 1.74E-05 | TRUE | FALSE | -2.789202716 |
| **padi4** | LV end diastolic volume \| Instance 2(participant - p24100_i2)_BSA_indexed_corrected | -1.149039974 | 0.3753278171 | -0.07549600668 | 7.88E-07 | 0.2323317868 | -0.07549600668 | 7.88E-07 | 4269 | 1.14E-05 | TRUE | 0.01152757919 | TRUE | 0.009750026479 | TRUE | FALSE | -1.149039974 |
| **pag1** | LV end diastolic volume \| Instance 2(participant - p24100_i2)_BSA_indexed_corrected | -2.001193754 | 0.2753917396 | -0.097380758 | 1.08E-10 | 0.309283829 | -0.097380758 | 1.08E-10 | 4375 | 3.21E-09 | TRUE | 1.59E-06 | TRUE | 1.34E-06 | TRUE | FALSE | -2.001193754 |
| **pag1** | LV end systolic volume \| Instance 2(participant - p24101_i2)_BSA_indexed_corrected | -0.9580896063 | 0.2747489025 | -0.08031546011 | 1.04E-07 | 0.1798092283 | -0.08031546011 | 1.04E-07 | 4375 | 1.80E-06 | TRUE | 0.00152275244 | TRUE | 0.001287944014 | TRUE | FALSE | -0.9580896063 |
| **pag1** | RV end diastolic volume \| Instance 2_BSA_indexed_corrected | -1.999271294 | 0.1658936921 | -0.09042907457 | 2.07E-09 | 0.3329591095 | -0.09042907457 | 2.07E-09 | 4375 | 4.76E-08 | TRUE | 3.03E-05 | TRUE | 2.57E-05 | TRUE | FALSE | -1.999271294 |
| **pag1** | RV end systolic volume \| Instance 2_BSA_indexed_corrected | -0.9751256668 | 0.1792977146 | -0.07416671504 | 9.06E-07 | 0.1982732137 | -0.07416671504 | 9.06E-07 | 4375 | 1.30E-05 | TRUE | 0.01325467939 | TRUE | 0.01121080783 | TRUE | FALSE | -0.9751256668 |
| **pamr1** | LV end diastolic volume \| Instance 2(participant - p24100_i2)_BSA_indexed_corrected | -4.831926447 | 0.03577614421 | -0.1284856882 | 2.48E-17 | 0.5681494652 | -0.1284856882 | 2.48E-17 | 4311 | 2.30E-15 | TRUE | 3.63E-13 | TRUE | 3.07E-13 | TRUE | FALSE | -4.831926447 |
| **pamr1** | LV end systolic volume \| Instance 2(participant - p24101_i2)_BSA_indexed_corrected | -2.326652994 | 0.1647400487 | -0.1065004147 | 2.37E-12 | 0.3309139213 | -0.1065004147 | 2.37E-12 | 4311 | 9.59E-11 | TRUE | 3.47E-08 | TRUE | 2.94E-08 | TRUE | FALSE | -2.326652994 |
| **pamr1** | RV end diastolic volume \| Instance 2_BSA_indexed_corrected | -5.441938025 | -0.08162257863 | -0.134423192 | 7.74E-19 | 0.6111270327 | -0.134423192 | 7.74E-19 | 4311 | 8.85E-17 | TRUE | 1.13E-14 | TRUE | 9.58E-15 | TRUE | FALSE | -5.441938025 |
| **pamr1** | RV end systolic volume \| Instance 2_BSA_indexed_corrected | -2.875609372 | 0.04374599742 | -0.1194560856 | 3.57E-15 | 0.3640930631 | -0.1194560856 | 3.57E-15 | 4311 | 2.36E-13 | TRUE | 5.22E-11 | TRUE | 4.42E-11 | TRUE | FALSE | -2.875609372 |
| **parp1** | LV end diastolic volume \| Instance 2(participant - p24100_i2)_BSA_indexed_corrected | -2.353411091 | 0.309395465 | -0.1060484336 | 2.60E-12 | 0.3353464064 | -0.1060484336 | 2.60E-12 | 4332 | 1.05E-10 | TRUE | 3.81E-08 | TRUE | 3.22E-08 | TRUE | FALSE | -2.353411091 |
| **parp1** | LV end systolic volume \| Instance 2(participant - p24101_i2)_BSA_indexed_corrected | -1.054703406 | 0.2956650091 | -0.08193087899 | 6.66E-08 | 0.1949737902 | -0.08193087899 | 6.66E-08 | 4332 | 1.19E-06 | TRUE | 0.0009747125882 | TRUE | 0.0008244119071 | TRUE | FALSE | -1.054703406 |
| **parp1** | RV end diastolic volume \| Instance 2_BSA_indexed_corrected | -2.154083453 | 0.2101466609 | -0.09023930263 | 2.68E-09 | 0.361282852 | -0.09023930263 | 2.68E-09 | 4332 | 6.04E-08 | TRUE | 3.93E-05 | TRUE | 3.32E-05 | TRUE | FALSE | -2.154083453 |
| **pbld** | LV end diastolic volume \| Instance 2(participant - p24100_i2)_BSA_indexed_corrected | -1.638111766 | 0.3652838245 | -0.0757831744 | 7.03E-07 | 0.3297263076 | -0.0757831744 | 7.03E-07 | 4275 | 1.03E-05 | TRUE | 0.01027851692 | TRUE | 0.008693569613 | TRUE | FALSE | -1.638111766 |
| **pbld** | RV end diastolic volume \| Instance 2_BSA_indexed_corrected | -1.97291789 | 0.2515577359 | -0.08488103106 | 2.73E-08 | 0.3542923336 | -0.08488103106 | 2.73E-08 | 4275 | 5.21E-07 | TRUE | 0.0003987088228 | TRUE | 0.0003372279223 | TRUE | FALSE | -1.97291789 |
| **pcsk9** | RV end diastolic volume \| Instance 2_BSA_indexed_corrected | -2.50164793 | 0.06572938897 | -0.08334378486 | 3.38E-08 | 0.4523239911 | -0.08334378486 | 3.38E-08 | 4375 | 6.34E-07 | TRUE | 0.0004939717382 | TRUE | 0.0004178012962 | TRUE | FALSE | -2.50164793 |
| **pdcd5** | LV end diastolic volume \| Instance 2(participant - p24100_i2)_BSA_indexed_corrected | -1.407066044 | 0.2668265112 | -0.07477516942 | 7.63E-07 | 0.2841492312 | -0.07477516942 | 7.63E-07 | 4363 | 1.11E-05 | TRUE | 0.0111608591 | TRUE | 0.009439854625 | TRUE | FALSE | -1.407066044 |
| **pgf** | LV end diastolic volume \| Instance 2(participant - p24100_i2)_BSA_indexed_corrected | -4.023063699 | 0.1575491475 | -0.1002246944 | 3.28E-11 | 0.6049179375 | -0.1002246944 | 3.28E-11 | 4361 | 1.09E-09 | TRUE | 4.80E-07 | TRUE | 4.06E-07 | TRUE | FALSE | -4.023063699 |
| **pgf** | LV end systolic volume \| Instance 2(participant - p24101_i2)_BSA_indexed_corrected | -1.905417597 | 0.2175624863 | -0.0817155697 | 6.52E-08 | 0.3519954706 | -0.0817155697 | 6.52E-08 | 4361 | 1.16E-06 | TRUE | 0.0009544419913 | TRUE | 0.0008072670362 | TRUE | FALSE | -1.905417597 |
| **pgf** | RV end diastolic volume \| Instance 2_BSA_indexed_corrected | -4.13864946 | 0.04493734867 | -0.09581585283 | 2.29E-10 | 0.6512160163 | -0.09581585283 | 2.29E-10 | 4361 | 6.44E-09 | TRUE | 3.36E-06 | TRUE | 2.84E-06 | TRUE | FALSE | -4.13864946 |
| **pi3** | LV end diastolic volume \| Instance 2(participant - p24100_i2)_BSA_indexed_corrected | -1.698034425 | 0.2789019477 | -0.08281963333 | 5.31E-08 | 0.311554409 | -0.08281963333 | 5.31E-08 | 4303 | 9.61E-07 | TRUE | 0.0007772644358 | TRUE | 0.0006574102598 | TRUE | FALSE | -1.698034425 |
| **pi3** | LV end systolic volume \| Instance 2(participant - p24101_i2)_BSA_indexed_corrected | -0.8577620764 | 0.2726379128 | -0.07215260531 | 2.16E-06 | 0.1807992737 | -0.07215260531 | 2.16E-06 | 4303 | 2.90E-05 | TRUE | 0.03160612367 | TRUE | 0.0267324594 | TRUE | FALSE | -0.8577620764 |
| **pi3** | RV end diastolic volume \| Instance 2_BSA_indexed_corrected | -1.999384113 | 0.1395420991 | -0.09055310556 | 2.67E-09 | 0.3352897276 | -0.09055310556 | 2.67E-09 | 4303 | 6.02E-08 | TRUE | 3.91E-05 | TRUE | 3.31E-05 | TRUE | FALSE | -1.999384113 |
| **pi3** | RV end systolic volume \| Instance 2_BSA_indexed_corrected | -0.9473597002 | 0.1685924095 | -0.072125179 | 2.18E-06 | 0.1997610093 | -0.072125179 | 2.18E-06 | 4303 | 2.92E-05 | TRUE | 0.03188886687 | TRUE | 0.0269716036 | TRUE | FALSE | -0.9473597002 |
| **pik3ip1** | LV cardiac output \| Instance 2_BSA_indexed_corrected | -0.1396511905 | -0.02599053151 | -0.07047300662 | 3.63E-06 | 0.03011289142 | -0.07047300662 | 3.63E-06 | 4311 | 4.68E-05 | TRUE | 0.0530982919 | FALSE | 0.04491053529 | TRUE | FALSE | -0.1396511905 |
| **pla2g15** | LV end diastolic volume \| Instance 2(participant - p24100_i2)_BSA_indexed_corrected | -3.096683675 | 0.2437704518 | -0.07113799875 | 2.72E-06 | 0.6592478753 | -0.07113799875 | 2.72E-06 | 4340 | 3.57E-05 | TRUE | 0.03976120393 | TRUE | 0.03363002628 | TRUE | FALSE | -3.096683675 |
| **pla2g15** | RV end diastolic volume \| Instance 2_BSA_indexed_corrected | -3.870071234 | 0.1105049617 | -0.0826606677 | 4.94E-08 | 0.7084129037 | -0.0826606677 | 4.94E-08 | 4340 | 9.01E-07 | TRUE | 0.0007231925734 | TRUE | 0.0006116762786 | TRUE | FALSE | -3.870071234 |
| **pla2g15** | RV end systolic volume \| Instance 2_BSA_indexed_corrected | -2.057559312 | 0.1388121147 | -0.07387384362 | 1.10E-06 | 0.4217242756 | -0.07387384362 | 1.10E-06 | 4340 | 1.56E-05 | TRUE | 0.01616508061 | TRUE | 0.01367242518 | TRUE | FALSE | -2.057559312 |
| **plat** | LV end diastolic volume \| Instance 2(participant - p24100_i2)_BSA_indexed_corrected | -3.507734858 | -0.1971295241 | -0.179615152 | 4.83E-33 | 0.2905179937 | -0.179615152 | 4.83E-33 | 4375 | 3.72E-30 | TRUE | 7.07E-29 | TRUE | 5.98E-29 | TRUE | FALSE | -3.507734858 |
| **plat** | LV end systolic volume \| Instance 2(participant - p24101_i2)_BSA_indexed_corrected | -1.628261006 | 0.0565392823 | -0.1436313508 | 1.33E-21 | 0.1696517895 | -0.1436313508 | 1.33E-21 | 4375 | 2.38E-19 | TRUE | 1.95E-17 | TRUE | 1.65E-17 | TRUE | FALSE | -1.628261006 |
| **plat** | RV end diastolic volume \| Instance 2_BSA_indexed_corrected | -3.609443896 | -0.322653939 | -0.17179434 | 2.49E-30 | 0.312994352 | -0.17179434 | 2.49E-30 | 4375 | 1.58E-27 | TRUE | 3.64E-26 | TRUE | 3.08E-26 | TRUE | FALSE | -3.609443896 |
| **plat** | RV end systolic volume \| Instance 2_BSA_indexed_corrected | -1.590601416 | -0.0323446637 | -0.1273039913 | 2.85E-17 | 0.1874051939 | -0.1273039913 | 2.85E-17 | 4375 | 2.61E-15 | TRUE | 4.18E-13 | TRUE | 3.53E-13 | TRUE | FALSE | -1.590601416 |
| **plaur** | LV end diastolic volume \| Instance 2(participant - p24100_i2)_BSA_indexed_corrected | -4.235726754 | -0.009589584002 | -0.09940749038 | 6.93E-11 | 0.6477801059 | -0.09940749038 | 6.93E-11 | 4286 | 2.12E-09 | TRUE | 1.01E-06 | TRUE | 8.57E-07 | TRUE | FALSE | -4.235726754 |
| **plaur** | RV end diastolic volume \| Instance 2_BSA_indexed_corrected | -5.139458572 | -0.1967987679 | -0.1122816491 | 1.68E-13 | 0.6949104474 | -0.1122816491 | 1.68E-13 | 4286 | 8.20E-12 | TRUE | 2.46E-09 | TRUE | 2.08E-09 | TRUE | FALSE | -5.139458572 |
| **plaur** | RV end systolic volume \| Instance 2_BSA_indexed_corrected | -2.394992501 | -0.01483226017 | -0.08791515943 | 8.16E-09 | 0.4146014263 | -0.08791515943 | 8.16E-09 | 4286 | 1.69E-07 | TRUE | 0.0001194113792 | TRUE | 0.0001009981445 | TRUE | FALSE | -2.394992501 |
| **plin1** | LV end diastolic volume \| Instance 2(participant - p24100_i2)_BSA_indexed_corrected | -3.120280688 | 0.2846642385 | -0.104009981 | 9.29E-12 | 0.4564469249 | -0.104009981 | 9.29E-12 | 4275 | 3.39E-10 | TRUE | 1.36E-07 | TRUE | 1.15E-07 | TRUE | FALSE | -3.120280688 |
| **plin1** | LV end systolic volume \| Instance 2(participant - p24101_i2)_BSA_indexed_corrected | -1.265770398 | 0.2684152799 | -0.07282942618 | 1.87E-06 | 0.2651714471 | -0.07282942618 | 1.87E-06 | 4275 | 2.53E-05 | TRUE | 0.02738642215 | TRUE | 0.02316343585 | TRUE | FALSE | -1.265770398 |
| **plin1** | RV end diastolic volume \| Instance 2_BSA_indexed_corrected | -3.693652364 | 0.1557008462 | -0.1145011225 | 5.96E-14 | 0.4902460357 | -0.1145011225 | 5.96E-14 | 4275 | 3.20E-12 | TRUE | 8.72E-10 | TRUE | 7.37E-10 | TRUE | FALSE | -3.693652364 |
| **plin1** | RV end systolic volume \| Instance 2_BSA_indexed_corrected | -1.46201901 | 0.1699951473 | -0.07634292552 | 5.81E-07 | 0.2921113862 | -0.07634292552 | 5.81E-07 | 4275 | 8.72E-06 | TRUE | 0.008501230926 | TRUE | 0.007190341117 | TRUE | FALSE | -1.46201901 |
| **pltp** | LV end diastolic volume \| Instance 2(participant - p24100_i2)_BSA_indexed_corrected | 4.013161281 | 0.3970312374 | 0.1340516305 | 5.73E-19 | 0.4489883373 | 0.1340516305 | 5.73E-19 | 4368 | 6.65E-17 | TRUE | 8.38E-15 | TRUE | 7.08E-15 | TRUE | FALSE | 4.013161281 |
| **pltp** | LV end systolic volume \| Instance 2(participant - p24101_i2)_BSA_indexed_corrected | 1.829672528 | 0.3265235766 | 0.1052654731 | 3.07E-12 | 0.2615929015 | 0.1052654731 | 3.07E-12 | 4368 | 1.22E-10 | TRUE | 4.49E-08 | TRUE | 3.80E-08 | TRUE | FALSE | 1.829672528 |
| **pltp** | RV end diastolic volume \| Instance 2_BSA_indexed_corrected | 4.214962526 | 0.2910794825 | 0.1308966731 | 3.75E-18 | 0.4831371277 | 0.1308966731 | 3.75E-18 | 4368 | 3.79E-16 | TRUE | 5.49E-14 | TRUE | 4.65E-14 | TRUE | FALSE | 4.214962526 |
| **pltp** | RV end systolic volume \| Instance 2_BSA_indexed_corrected | 1.856192779 | 0.2373998701 | 0.09694050388 | 1.36E-10 | 0.2884201901 | 0.09694050388 | 1.36E-10 | 4368 | 3.96E-09 | TRUE | 1.99E-06 | TRUE | 1.68E-06 | TRUE | FALSE | 1.856192779 |
| **plxnb2** | RV end diastolic volume \| Instance 2_BSA_indexed_corrected | -3.826457943 | 0.04974900972 | -0.07415128601 | 9.27E-07 | 0.7787344743 | -0.07415128601 | 9.27E-07 | 4369 | 1.33E-05 | TRUE | 0.01355499308 | TRUE | 0.01146481315 | TRUE | FALSE | -3.826457943 |
| **polr2f** | LV end diastolic volume \| Instance 2(participant - p24100_i2)_BSA_indexed_corrected | -4.108974314 | 0.1707973702 | -0.1216412911 | 6.83E-16 | 0.5070206139 | -0.1216412911 | 6.83E-16 | 4375 | 5.20E-14 | TRUE | 9.99E-12 | TRUE | 8.45E-12 | TRUE | FALSE | -4.108974314 |
| **polr2f** | LV end systolic volume \| Instance 2(participant - p24101_i2)_BSA_indexed_corrected | -1.771196204 | 0.2333658222 | -0.09032818382 | 2.16E-09 | 0.295307614 | -0.09032818382 | 2.16E-09 | 4375 | 4.93E-08 | TRUE | 3.16E-05 | TRUE | 2.67E-05 | TRUE | FALSE | -1.771196204 |
| **polr2f** | RV end diastolic volume \| Instance 2_BSA_indexed_corrected | -4.046117242 | 0.06401223029 | -0.1113367146 | 1.52E-13 | 0.5461372166 | -0.1113367146 | 1.52E-13 | 4375 | 7.48E-12 | TRUE | 2.23E-09 | TRUE | 1.89E-09 | TRUE | FALSE | -4.046117242 |
| **polr2f** | RV end systolic volume \| Instance 2_BSA_indexed_corrected | -1.873026941 | 0.134059624 | -0.08666745884 | 9.37E-09 | 0.3255824744 | -0.08666745884 | 9.37E-09 | 4375 | 1.93E-07 | TRUE | 0.0001371433307 | TRUE | 0.0001159958291 | TRUE | FALSE | -1.873026941 |
| **pon2** | LV end diastolic volume \| Instance 2(participant - p24100_i2)_BSA_indexed_corrected | -2.79579168 | 0.2260909866 | -0.08823468029 | 5.04E-09 | 0.4772858606 | -0.08823468029 | 5.04E-09 | 4375 | 1.08E-07 | TRUE | 7.37E-05 | TRUE | 6.23E-05 | TRUE | FALSE | -2.79579168 |
| **pon2** | LV end systolic volume \| Instance 2(participant - p24101_i2)_BSA_indexed_corrected | -1.507681073 | 0.2434656732 | -0.08196963514 | 5.66E-08 | 0.2772061121 | -0.08196963514 | 5.66E-08 | 4375 | 1.02E-06 | TRUE | 0.0008273725158 | TRUE | 0.0006997916739 | TRUE | FALSE | -1.507681073 |
| **pon2** | RV end diastolic volume \| Instance 2_BSA_indexed_corrected | -3.104548546 | 0.1025013443 | -0.09107209092 | 1.59E-09 | 0.5133515196 | -0.09107209092 | 1.59E-09 | 4375 | 3.73E-08 | TRUE | 2.33E-05 | TRUE | 1.97E-05 | TRUE | FALSE | -3.104548546 |
| **pon2** | RV end systolic volume \| Instance 2_BSA_indexed_corrected | -1.874584858 | 0.1320184943 | -0.09247068116 | 8.91E-10 | 0.3052435787 | -0.09247068116 | 8.91E-10 | 4375 | 2.21E-08 | TRUE | 1.30E-05 | TRUE | 1.10E-05 | TRUE | FALSE | -1.874584858 |
| **pon3** | LV end diastolic volume \| Instance 2(participant - p24100_i2)_BSA_indexed_corrected | 5.725315133 | 0.2990673772 | 0.152081193 | 8.84E-24 | 0.5661098216 | 0.152081193 | 8.84E-24 | 4322 | 2.27E-21 | TRUE | 1.29E-19 | TRUE | 1.09E-19 | TRUE | FALSE | 5.725315133 |
| **pon3** | LV end systolic volume \| Instance 2(participant - p24101_i2)_BSA_indexed_corrected | 2.582688405 | 0.2831253234 | 0.1183089417 | 6.04E-15 | 0.3298007864 | 0.1183089417 | 6.04E-15 | 4322 | 3.86E-13 | TRUE | 8.84E-11 | TRUE | 7.48E-11 | TRUE | FALSE | 2.582688405 |
| **pon3** | RV end diastolic volume \| Instance 2_BSA_indexed_corrected | 6.133519365 | 0.1837297098 | 0.1515254887 | 1.29E-23 | 0.6087491278 | 0.1515254887 | 1.29E-23 | 4322 | 3.14E-21 | TRUE | 1.89E-19 | TRUE | 1.59E-19 | TRUE | FALSE | 6.133519365 |

| | | | | | | | | | | | | | | | | | |
|---|---|---|---|---|---|---|---|---|---|---|---|---|---|---|---|---|---|
| **pon3** | RV end systolic volume \| Instance 2_BSA_indexed_corrected | 2.737061772 | 0.1862089009 | 0.1136509086 | 6.69E-14 | 0.364038028 | 0.1136509086 | 6.69E-14 | 4322 | 3.57E-12 | TRUE | 9.79E-10 | TRUE | 8.28E-10 | TRUE | FALSE | 2.737061772 |
| **prcp** | LV end diastolic volume \| Instance 2(participant - p24100_i2)_BSA_indexed_corrected | -4.694575716 | -0.02108597653 | -0.1399207104 | 1.53E-20 | 0.5027812205 | -0.1399207104 | 1.53E-20 | 4368 | 2.31E-18 | TRUE | 2.25E-16 | TRUE | 1.90E-16 | TRUE | FALSE | -4.694575716 |
| **prcp** | LV end systolic volume \| Instance 2(participant - p24101_i2)_BSA_indexed_corrected | -2.287978242 | 0.1291506624 | -0.1175250038 | 6.59E-15 | 0.2925900678 | -0.1175250038 | 6.59E-15 | 4368 | 4.16E-13 | TRUE | 9.64E-11 | TRUE | 8.15E-11 | TRUE | FALSE | -2.287978242 |
| **prcp** | RV end diastolic volume \| Instance 2_BSA_indexed_corrected | -5.533151842 | -0.1997994619 | -0.153204736 | 2.37E-24 | 0.5401332973 | -0.153204736 | 2.37E-24 | 4368 | 6.93E-22 | TRUE | 3.47E-20 | TRUE | 2.93E-20 | TRUE | FALSE | -5.533151842 |
| **prcp** | RV end systolic volume \| Instance 2_BSA_indexed_corrected | -2.758474666 | -0.005967935518 | -0.1284692096 | 1.55E-17 | 0.3222658648 | -0.1284692096 | 1.55E-17 | 4368 | 1.45E-15 | TRUE | 2.26E-13 | TRUE | 1.91E-13 | TRUE | FALSE | -2.758474666 |
| **proc** | LV end diastolic volume \| Instance 2(participant - p24100_i2)_BSA_indexed_corrected | -4.54061546 | 0.2376235941 | -0.104055634 | 7.01E-12 | 0.6603029706 | -0.104055634 | 7.01E-12 | 4322 | 2.62E-10 | TRUE | 1.03E-07 | TRUE | 8.67E-08 | TRUE | FALSE | -4.54061546 |
| **proc** | LV end systolic volume \| Instance 2(participant - p24101_i2)_BSA_indexed_corrected | -1.964960131 | 0.2646680793 | -0.07753868634 | 3.33E-07 | 0.3844005028 | -0.07753868634 | 3.33E-07 | 4322 | 5.20E-06 | TRUE | 0.004871551917 | TRUE | 0.004120358612 | TRUE | FALSE | -1.964960131 |
| **proc** | RV end diastolic volume \| Instance 2_BSA_indexed_corrected | -4.965541903 | 0.1150082263 | -0.105860934 | 3.01E-12 | 0.7096461474 | -0.105860934 | 3.01E-12 | 4322 | 1.20E-10 | TRUE | 4.41E-08 | TRUE | 3.73E-08 | TRUE | FALSE | -4.965541903 |
| **proc** | RV end systolic volume \| Instance 2_BSA_indexed_corrected | -2.144098119 | 0.1753886197 | -0.07687378437 | 4.20E-07 | 0.4230945838 | -0.07687378437 | 4.20E-07 | 4322 | 6.43E-06 | TRUE | 0.006138331191 | TRUE | 0.005191800521 | TRUE | FALSE | -2.144098119 |
| **prss2** | RV end diastolic volume \| Instance 2_BSA_indexed_corrected | -1.834591435 | 0.2177508738 | -0.07284451804 | 1.63E-06 | 0.3821158327 | -0.07284451804 | 1.63E-06 | 4323 | 2.23E-05 | TRUE | 0.02386142265 | TRUE | 0.02018199128 | TRUE | FALSE | -1.834591435 |
| **prss8** | LV end diastolic volume \| Instance 2(participant - p24100_i2)_BSA_indexed_corrected | -3.543771171 | -0.004325207614 | -0.1355548312 | 4.40E-19 | 0.3951306999 | -0.1355548312 | 4.40E-19 | 4299 | 5.19E-17 | TRUE | 6.43E-15 | TRUE | 5.44E-15 | TRUE | FALSE | -3.543771171 |
| **prss8** | LV end systolic volume \| Instance 2(participant - p24101_i2)_BSA_indexed_corrected | -1.462837222 | 0.1665300428 | -0.0963489294 | 2.45E-10 | 0.2305373064 | -0.0963489294 | 2.45E-10 | 4299 | 6.84E-09 | TRUE | 3.58E-06 | TRUE | 3.03E-06 | TRUE | FALSE | -1.462837222 |
| **prss8** | RV end diastolic volume \| Instance 2_BSA_indexed_corrected | -4.157787816 | -0.1655876488 | -0.1478119161 | 1.99E-22 | 0.4243979638 | -0.1478119161 | 1.99E-22 | 4299 | 3.83E-20 | TRUE | 2.91E-18 | TRUE | 2.46E-18 | TRUE | FALSE | -4.157787816 |
| **prss8** | RV end systolic volume \| Instance 2_BSA_indexed_corrected | -1.815428375 | 0.03783272957 | -0.1083533572 | 1.05E-12 | 0.2540911744 | -0.1083533572 | 1.05E-12 | 4299 | 4.51E-11 | TRUE | 1.54E-08 | TRUE | 1.30E-08 | TRUE | FALSE | -1.815428375 |
| **psip1** | LV end diastolic volume \| Instance 2(participant - p24100_i2)_BSA_indexed_corrected | -1.48536645 | 0.3221918406 | -0.07971423907 | 1.49E-07 | 0.2822731196 | -0.07971423907 | 1.49E-07 | 4332 | 2.49E-06 | TRUE | 0.002183632937 | TRUE | 0.001846916738 | TRUE | FALSE | -1.48536645 |
| **pspn** | LV end diastolic volume \| Instance 2(participant - p24100_i2)_BSA_indexed_corrected | -0.6138788331 | 0.3089118364 | -0.07309627035 | 1.46E-06 | 0.1272858568 | -0.07309627035 | 1.46E-06 | 4332 | 2.01E-05 | TRUE | 0.02141700781 | TRUE | 0.0181145052 | TRUE | FALSE | -0.6138788331 |
| **ptgds** | LV cardiac output \| Instance 2_BSA_indexed_corrected | -0.1516473464 | -0.02345627823 | -0.08633483776 | 1.48E-08 | 0.02672059358 | -0.08633483776 | 1.48E-08 | 4291 | 2.96E-07 | TRUE | 0.0002159052162 | TRUE | 0.0001826126319 | TRUE | FALSE | -0.1516473464 |
| **ptgds** | LV end diastolic volume \| Instance 2(participant - p24100_i2)_BSA_indexed_corrected | -4.710499021 | 0.2446754275 | -0.119469156 | 4.10E-15 | 0.5977388064 | -0.119469156 | 4.10E-15 | 4291 | 2.67E-13 | TRUE | 6.00E-11 | TRUE | 5.08E-11 | TRUE | FALSE | -4.710499021 |
| **ptgds** | LV end systolic volume \| Instance 2(participant - p24101_i2)_BSA_indexed_corrected | -2.215994105 | 0.2647267671 | -0.09681686 | 2.08E-10 | 0.3478520396 | -0.09681686 | 2.08E-10 | 4291 | 5.88E-09 | TRUE | 3.05E-06 | TRUE | 2.58E-06 | TRUE | FALSE | -2.215994105 |
| **ptgds** | RV end diastolic volume \| Instance 2_BSA_indexed_corrected | -5.132064077 | 0.118487291 | -0.1210243223 | 1.80E-15 | 0.6427429687 | -0.1210243223 | 1.80E-15 | 4291 | 1.24E-13 | TRUE | 2.63E-11 | TRUE | 2.22E-11 | TRUE | FALSE | -5.132064077 |
| **ptgds** | RV end systolic volume \| Instance 2_BSA_indexed_corrected | -2.326623773 | 0.1814444959 | -0.0923191907 | 1.37E-09 | 0.3831753017 | -0.0923191907 | 1.37E-09 | 4291 | 3.28E-08 | TRUE | 2.01E-05 | TRUE | 1.70E-05 | TRUE | FALSE | -2.326623773 |
| **ptprm** | LV end diastolic volume \| Instance 2(participant - p24100_i2)_BSA_indexed_corrected | -4.605532263 | 0.2881701056 | -0.1014256366 | 2.28E-11 | 0.6869001848 | -0.1014256366 | 2.28E-11 | 4327 | 7.75E-10 | TRUE | 3.33E-07 | TRUE | 2.82E-07 | TRUE | FALSE | -4.605532263 |
| **ptprm** | LV end systolic volume \| Instance 2(participant - p24101_i2)_BSA_indexed_corrected | -2.393177421 | 0.2814810532 | -0.09087487793 | 2.12E-09 | 0.3987836095 | -0.09087487793 | 2.12E-09 | 4327 | 4.84E-08 | TRUE | 3.10E-05 | TRUE | 2.62E-05 | TRUE | FALSE | -2.393177421 |
| **ptprm** | RV end diastolic volume \| Instance 2_BSA_indexed_corrected | -3.923663907 | 0.1895587684 | -0.08037867224 | 1.19E-07 | 0.7398615096 | -0.08037867224 | 1.19E-07 | 4327 | 2.02E-06 | TRUE | 0.001747471836 | TRUE | 0.001478011679 | TRUE | FALSE | -3.923663907 |
| **ptprn2** | LV cardiac output \| Instance 2_BSA_indexed_corrected | -0.1831489751 | -0.02180310006 | -0.1117247923 | 1.34E-13 | 0.02466236643 | -0.1117247923 | 1.34E-13 | 4365 | 6.67E-12 | TRUE | 1.95E-09 | TRUE | 1.65E-09 | TRUE | FALSE | -0.1831489751 |
| **ptprn2** | LV end diastolic volume \| Instance 2(participant - p24100_i2)_BSA_indexed_corrected | -3.625092197 | 0.3196496213 | -0.09870384537 | 6.35E-11 | 0.5533073748 | -0.09870384537 | 6.35E-11 | 4365 | 1.98E-09 | TRUE | 9.29E-07 | TRUE | 7.86E-07 | TRUE | FALSE | -3.625092197 |
| **ptprn2** | LV end systolic volume \| Instance 2(participant - p24101_i2)_BSA_indexed_corrected | -1.709771678 | 0.3012432556 | -0.08016430556 | 1.14E-07 | 0.3218581552 | -0.08016430556 | 1.14E-07 | 4365 | 1.95E-06 | TRUE | 0.001663595076 | TRUE | 0.001407068715 | TRUE | FALSE | -1.709771678 |
| **ptprn2** | RV end diastolic volume \| Instance 2_BSA_indexed_corrected | -3.618607782 | 0.208574301 | -0.09162537527 | 1.32E-09 | 0.5953912503 | -0.09162537527 | 1.32E-09 | 4365 | 3.17E-08 | TRUE | 1.94E-05 | TRUE | 1.64E-05 | TRUE | FALSE | -3.618607782 |
| **pts** | LV end diastolic volume \| Instance 2(participant - p24100_i2)_BSA_indexed_corrected | -2.429871307 | 0.2725886196 | -0.1215978101 | 1.66E-15 | 0.3039267881 | -0.1215978101 | 1.66E-15 | 4261 | 1.15E-13 | TRUE | 2.42E-11 | TRUE | 2.05E-11 | TRUE | FALSE | -2.429871307 |
| **pts** | LV end systolic volume \| Instance 2(participant - p24101_i2)_BSA_indexed_corrected | -1.159100915 | 0.2700021928 | -0.1001202593 | 5.75E-11 | 0.1765053125 | -0.1001202593 | 5.75E-11 | 4261 | 1.81E-09 | TRUE | 8.42E-07 | TRUE | 7.12E-07 | TRUE | FALSE | -1.159100915 |
| **pts** | RV end diastolic volume \| Instance 2_BSA_indexed_corrected | -2.35700316 | 0.1578906544 | -0.1097197671 | 6.89E-13 | 0.3271835756 | -0.1097197671 | 6.89E-13 | 4261 | 3.04E-11 | TRUE | 1.01E-08 | TRUE | 8.52E-09 | TRUE | FALSE | -2.35700316 |
| **pts** | RV end systolic volume \| Instance 2_BSA_indexed_corrected | -1.128614034 | 0.1732124862 | -0.08846024996 | 7.29E-09 | 0.1947321042 | -0.08846024996 | 7.29E-09 | 4261 | 1.53E-07 | TRUE | 0.0001067132326 | TRUE | 9.03E-05 | TRUE | FALSE | -1.128614034 |
| **pxn** | LV end diastolic volume \| Instance 2(participant - p24100_i2)_BSA_indexed_corrected | -2.167726401 | 0.2115941977 | -0.1046644283 | 6.88E-12 | 0.3150989959 | -0.1046644283 | 6.88E-12 | 4275 | 2.58E-10 | TRUE | 1.01E-07 | TRUE | 8.51E-08 | TRUE | FALSE | -2.167726401 |
| **pxn** | LV end systolic volume \| Instance 2(participant - p24101_i2)_BSA_indexed_corrected | -0.9792715812 | 0.2326349471 | -0.08161470017 | 9.11E-08 | 0.1829435355 | -0.08161470017 | 9.11E-08 | 4275 | 1.59E-06 | TRUE | 0.001332998875 | TRUE | 0.001127450449 | TRUE | FALSE | -0.9792715812 |
| **pxn** | RV end diastolic volume \| Instance 2_BSA_indexed_corrected | -2.074730781 | 0.09939155064 | -0.09315984577 | 1.04E-09 | 0.3392139854 | -0.09315984577 | 1.04E-09 | 4275 | 2.57E-08 | TRUE | 1.53E-05 | TRUE | 1.29E-05 | TRUE | FALSE | -2.074730781 |
| **pxn** | RV end systolic volume \| Instance 2_BSA_indexed_corrected | -1.006609296 | 0.1363162197 | -0.0761359902 | 6.23E-07 | 0.2016703688 | -0.0761359902 | 6.23E-07 | 4275 | 9.23E-06 | TRUE | 0.009120719226 | TRUE | 0.007714304321 | TRUE | FALSE | -1.006609296 |
| **rarres2** | LV end diastolic volume \| Instance 2(participant - p24100_i2)_BSA_indexed_corrected | -3.589609574 | -0.160290378 | -0.1592590354 | 5.56E-26 | 0.3383149779 | -0.1592590354 | 5.56E-26 | 4328 | 1.98E-23 | TRUE | 8.13E-22 | TRUE | 6.87E-22 | TRUE | FALSE | -3.589609574 |
| **rarres2** | LV end systolic volume \| Instance 2(participant - p24101_i2)_BSA_indexed_corrected | -1.493222432 | 0.1042262083 | -0.1141576041 | 4.97E-14 | 0.1975731608 | -0.1141576041 | 4.97E-14 | 4328 | 2.71E-12 | TRUE | 7.28E-10 | TRUE | 6.15E-10 | TRUE | FALSE | -1.493222432 |
| **rarres2** | RV end diastolic volume \| Instance 2_BSA_indexed_corrected | -4.266365697 | -0.3691962654 | -0.1761006927 | 1.75E-31 | 0.3625877093 | -0.1761006927 | 1.75E-31 | 4328 | 1.28E-28 | TRUE | 2.56E-27 | TRUE | 2.16E-27 | TRUE | FALSE | -4.266365697 |
| **rarres2** | RV end systolic volume \| Instance 2_BSA_indexed_corrected | -1.747361759 | -0.02191226215 | -0.1212924948 | 1.18E-15 | 0.2174138798 | -0.1212924948 | 1.18E-15 | 4328 | 8.61E-14 | TRUE | 1.72E-11 | TRUE | 1.46E-11 | TRUE | FALSE | -1.747361759 |
| **rassf2** | LV end diastolic volume \| Instance 2(participant - p24100_i2)_BSA_indexed_corrected | -1.813014424 | 0.2954011827 | -0.09016123094 | 2.31E-09 | 0.3028442095 | -0.09016123094 | 2.31E-09 | 4375 | 5.25E-08 | TRUE | 3.39E-05 | TRUE | 2.86E-05 | TRUE | FALSE | -1.813014424 |
| **rassf2** | LV end systolic volume \| Instance 2(participant - p24101_i2)_BSA_indexed_corrected | -0.8733062291 | 0.2841598894 | -0.07481594084 | 7.27E-07 | 0.1760207218 | -0.07481594084 | 7.27E-07 | 4375 | 1.06E-05 | TRUE | 0.0106288599 | TRUE | 0.008989889705 | TRUE | FALSE | -0.8733062291 |
| **rassf2** | RV end diastolic volume \| Instance 2_BSA_indexed_corrected | -1.61621704 | 0.1920823841 | -0.07470860005 | 7.54E-07 | 0.3262300548 | -0.07470860005 | 7.54E-07 | 4375 | 1.10E-05 | TRUE | 0.0110253883 | TRUE | 0.009325273426 | TRUE | FALSE | -1.61621704 |
| **rassf2** | RV end systolic volume \| Instance 2_BSA_indexed_corrected | -0.9420436673 | 0.1871851699 | -0.07322410151 | 1.24E-06 | 0.19402595 | -0.07322410151 | 1.24E-06 | 4375 | 1.75E-05 | TRUE | 0.01820460088 | TRUE | 0.01539745142 | TRUE | FALSE | -0.9420436673 |
| **rbks** | LV end diastolic volume \| Instance 2(participant - p24100_i2)_BSA_indexed_corrected | -1.946687704 | 0.3096425708 | -0.08987506756 | 3.47E-09 | 0.3288211844 | -0.08987506756 | 3.47E-09 | 4306 | 7.68E-08 | TRUE | 5.07E-05 | TRUE | 4.29E-05 | TRUE | FALSE | -1.946687704 |
| **rbks** | LV end systolic volume \| Instance 2(participant - p24101_i2)_BSA_indexed_corrected | -0.9187650128 | 0.3013897781 | -0.07315023018 | 1.55E-06 | 0.1909358834 | -0.07315023018 | 1.55E-06 | 4306 | 2.12E-05 | TRUE | 0.02262075538 | TRUE | 0.0191326349 | TRUE | FALSE | -0.9187650128 |
| **rbks** | RV end diastolic volume \| Instance 2_BSA_indexed_corrected | -1.70810899 | 0.2135630733 | -0.07330000298 | 1.47E-06 | 0.3542465268 | -0.07330000298 | 1.47E-06 | 4306 | 2.02E-05 | TRUE | 0.02153142054 | TRUE | 0.0182112755 | TRUE | FALSE | -1.70810899 |
| **rbp5** | LV end diastolic volume \| Instance 2(participant - p24100_i2)_BSA_indexed_corrected | -2.871493298 | 0.1730299378 | -0.150022918 | 1.93E-23 | 0.2861655273 | -0.150022918 | 1.93E-23 | 4375 | 4.62E-21 | TRUE | 2.82E-19 | TRUE | 2.38E-19 | TRUE | FALSE | -2.871493298 |
| **rbp5** | LV end systolic volume \| Instance 2(participant - p24101_i2)_BSA_indexed_corrected | -1.404869098 | 0.2238546119 | -0.1264429652 | 4.67E-17 | 0.1666677934 | -0.1264429652 | 4.67E-17 | 4375 | 4.14E-15 | TRUE | 6.83E-13 | TRUE | 5.78E-13 | TRUE | FALSE | -1.404869098 |
| **rbp5** | RV end diastolic volume \| Instance 2_BSA_indexed_corrected | -3.181895184 | 0.04400133789 | -0.1545211795 | 8.72E-25 | 0.3076524901 | -0.1545211795 | 8.72E-25 | 4375 | 2.71E-22 | TRUE | 1.28E-20 | TRUE | 1.08E-20 | TRUE | FALSE | -3.181895184 |
| **rbp5** | RV end systolic volume \| Instance 2_BSA_indexed_corrected | -1.610178045 | 0.1161954674 | -0.1314886135 | 2.49E-18 | 0.1835730266 | -0.1314886135 | 2.49E-18 | 4375 | 2.58E-16 | TRUE | 3.64E-14 | TRUE | 3.08E-14 | TRUE | FALSE | -1.610178045 |
| **relt** | LV end diastolic volume \| Instance 2(participant - p24100_i2)_BSA_indexed_corrected | -3.853412612 | 0.1594984364 | -0.1004193527 | 3.89E-11 | 0.5816195705 | -0.1004193527 | 3.89E-11 | 4311 | 1.27E-09 | TRUE | 5.69E-07 | TRUE | 4.82E-07 | TRUE | FALSE | -3.853412612 |
| **relt** | LV end systolic volume \| Instance 2(participant - p24101_i2)_BSA_indexed_corrected | -1.819984588 | 0.2260241721 | -0.08164410952 | 7.96E-08 | 0.3384560717 | -0.08164410952 | 7.96E-08 | 4311 | 1.40E-06 | TRUE | 0.00116466855 | TRUE | 0.0009850766596 | TRUE | FALSE | -1.819984588 |
| **relt** | RV end diastolic volume \| Instance 2_BSA_indexed_corrected | -3.804359079 | 0.08351355057 | -0.09209574745 | 1.38E-09 | 0.6266197237 | -0.09209574745 | 1.38E-09 | 4311 | 3.29E-08 | TRUE | 2.02E-05 | TRUE | 1.71E-05 | TRUE | FALSE | -3.804359079 |
| **relt** | RV end systolic volume \| Instance 2_BSA_indexed_corrected | -1.779744116 | 0.1421108961 | -0.07245584374 | 1.91E-06 | 0.3732096868 | -0.07245584374 | 1.91E-06 | 4311 | 2.59E-05 | TRUE | 0.02801539214 | TRUE | 0.02369541868 | TRUE | FALSE | -1.779744116 |
| **ren** | LV end diastolic volume \| Instance 2(participant - p24100_i2)_BSA_indexed_corrected | -1.707370447 | 0.2709854834 | -0.1007181927 | 2.44E-11 | 0.2550445652 | -0.1007181927 | 2.44E-11 | 4375 | 8.22E-10 | TRUE | 3.57E-07 | TRUE | 3.02E-07 | TRUE | FALSE | -1.707370447 |
| **ren** | LV end systolic volume \| Instance 2(participant - p24101_i2)_BSA_indexed_corrected | -0.8469290202 | 0.2712215782 | -0.08606690564 | 1.19E-08 | 0.1482540542 | -0.08606690564 | 1.19E-08 | 4375 | 2.41E-07 | TRUE | 0.0001734919339 | TRUE | 0.0001467394777 | TRUE | FALSE | -0.8469290202 |
| **ren** | RV end diastolic volume \| Instance 2_BSA_indexed_corrected | -1.831114478 | 0.1554676589 | -0.100403304 | 2.81E-11 | 0.2743959026 | -0.100403304 | 2.81E-11 | 4375 | 9.38E-10 | TRUE | 4.12E-07 | TRUE | 3.48E-07 | TRUE | FALSE | -1.831114478 |
| **ren** | RV end systolic volume \| Instance 2_BSA_indexed_corrected | -0.8557416705 | 0.1760077953 | -0.07890186858 | 1.74E-07 | 0.1634969204 | -0.07890186858 | 1.74E-07 | 4375 | 2.86E-06 | TRUE | 0.002540675357 | TRUE | 0.002148903217 | TRUE | FALSE | -0.8557416705 |
| **ret** | LV end diastolic volume \| Instance 2(participant - p24100_i2)_BSA_indexed_corrected | -1.96476169 | 0.3792889208 | -0.06982074826 | 3.79E-06 | 0.4244968542 | -0.06982074826 | 3.79E-06 | 4375 | 4.87E-05 | TRUE | 0.05546043071 | FALSE | 0.04690843229 | TRUE | FALSE | -1.96476169 |
| **retn** | LV end diastolic volume \| Instance 2(participant - p24100_i2)_BSA_indexed_corrected | -2.324199388 | 0.1918393032 | -0.07969018303 | 1.56E-07 | 0.4423780397 | -0.07969018303 | 1.56E-07 | 4321 | 2.59E-06 | TRUE | 0.002283844835 | TRUE | 0.001931675962 | TRUE | FALSE | -2.324199388 |
| **retn** | RV end diastolic volume \| Instance 2_BSA_indexed_corrected | -2.380175952 | 0.1045664785 | -0.07582331746 | 6.04E-07 | 0.4762803443 | -0.07582331746 | 6.04E-07 | 4321 | 9.00E-06 | TRUE | 0.008836422255 | TRUE | 0.007473845943 | TRUE | FALSE | -2.380175952 |
| **rgmb** | LV cardiac output \| Instance 2_BSA_indexed_corrected | -0.1708630363 | -0.01877364985 | -0.08747807372 | 7.14E-09 | 0.02946373326 | -0.08747807372 | 7.14E-09 | 4363 | 1.50E-07 | TRUE | 0.0001044373088 | TRUE | 8.83E-05 | TRUE | FALSE | -0.1708630363 |
| **rnase3** | LV end diastolic volume \| Instance 2(participant - p24100_i2)_BSA_indexed_corrected | -0.7669483508 | 0.3221808735 | -0.07411537721 | 9.22E-07 | 0.1560529413 | -0.07411537721 | 9.22E-07 | 4375 | 1.32E-05 | TRUE | 0.01348706989 | TRUE | 0.01140736372 | TRUE | FALSE | -0.7669483508 |
| **rnase3** | RV end diastolic volume \| Instance 2_BSA_indexed_corrected | -0.7978297305 | 0.2113667094 | -0.07166461389 | 2.09E-06 | 0.167918138 | -0.07166461389 | 2.09E-06 | 4375 | 2.81E-05 | TRUE | 0.03051718877 | TRUE | 0.02581143826 | TRUE | FALSE | -0.7978297305 |
| **robo2** | RV end diastolic volume \| Instance 2_BSA_indexed_corrected | -4.118911702 | 0.1634678853 | -0.07894690941 | 1.78E-07 | 0.7875836202 | -0.07894690941 | 1.78E-07 | 4363 | 2.92E-06 | TRUE | 0.002598808851 | TRUE | 0.002198072526 | TRUE | FALSE | -4.118911702 |
| **robo2** | RV end systolic volume \| Instance 2_BSA_indexed_corrected | -2.323315165 | 0.1741080943 | -0.07493858924 | 7.22E-07 | 0.4681517813 | -0.07493858924 | 7.22E-07 | 4363 | 1.06E-05 | TRUE | 0.01055638566 | TRUE | 0.008928590989 | TRUE | FALSE | -2.323315165 |
| **rtbdn** | LV end diastolic volume \| Instance 2(participant - p24100_i2)_BSA_indexed_corrected | -3.223170223 | 0.3161674955 | -0.08294684545 | 3.92E-08 | 0.5855910823 | -0.08294684545 | 3.92E-08 | 4375 | 7.27E-07 | TRUE | 0.000573815801 | TRUE | 0.0004853334045 | TRUE | FALSE | -3.223170223 |

| | | | | | | | | | | | | | | | | | |
|---|---|---|---|---|---|---|---|---|---|---|---|---|---|---|---|---|---|
| **rtbdn** | LV end systolic volume \| Instance 2(participant - p24101_i2)_BSA_indexed_corrected | -1.669474666 | 0.2928261878 | -0.07401256845 | 9.54E-07 | 0.340166739 | -0.07401256845 | 9.54E-07 | 4375 | 1.37E-05 | TRUE | 0.01396429927 | TRUE | 0.01181100432 | TRUE | FALSE | -1.669474666 |
| **rtbdn** | RV end diastolic volume \| Instance 2_BSA_indexed_corrected | -3.468688188 | 0.2037880948 | -0.08297251174 | 3.88E-08 | 0.6300009135 | -0.08297251174 | 3.88E-08 | 4375 | 7.21E-07 | TRUE | 0.0005682952243 | TRUE | 0.0004806641007 | TRUE | FALSE | -3.468688188 |
| **rtn4r** | LV cardiac output \| Instance 2_BSA_indexed_corrected | -0.1033671553 | -0.02525435531 | -0.07542055798 | 5.91E-07 | 0.02066637797 | -0.07542055798 | 5.91E-07 | 4375 | 8.83E-06 | TRUE | 0.008639411604 | TRUE | 0.007307214335 | TRUE | FALSE | -0.1033671553 |
| **rtn4r** | LV end diastolic volume \| Instance 2(participant - p24100_i2)_BSA_indexed_corrected | -4.690535814 | 0.1031026807 | -0.1527452241 | 2.99E-24 | 0.4589218716 | -0.1527452241 | 2.99E-24 | 4375 | 8.42E-22 | TRUE | 4.38E-20 | TRUE | 3.70E-20 | TRUE | FALSE | -4.690535814 |
| **rtn4r** | LV end systolic volume \| Instance 2(participant - p24101_i2)_BSA_indexed_corrected | -1.99178318 | 0.2057892939 | -0.1117368066 | 1.25E-13 | 0.2678723528 | -0.1117368066 | 1.25E-13 | 4375 | 6.24E-12 | TRUE | 1.82E-09 | TRUE | 1.54E-09 | TRUE | FALSE | -1.99178318 |
| **rtn4r** | RV end diastolic volume \| Instance 2_BSA_indexed_corrected | -5.549999478 | -0.05226227803 | -0.1679927232 | 4.66E-29 | 0.4924888067 | -0.1679927232 | 4.66E-29 | 4375 | 2.27E-26 | TRUE | 6.82E-25 | TRUE | 5.77E-25 | TRUE | FALSE | -5.549999478 |
| **rtn4r** | RV end systolic volume \| Instance 2_BSA_indexed_corrected | -2.30034526 | 0.09455867952 | -0.1170853752 | 7.91E-15 | 0.2950551314 | -0.1170853752 | 7.91E-15 | 4375 | 4.88E-13 | TRUE | 1.16E-10 | TRUE | 9.79E-11 | TRUE | FALSE | -2.30034526 |
| **s100a11** | LV end diastolic volume \| Instance 2(participant - p24100_i2)_BSA_indexed_corrected | -2.664403232 | 0.2378470497 | -0.1086279227 | 6.46E-13 | 0.369434084 | -0.1086279227 | 6.46E-13 | 4358 | 2.88E-11 | TRUE | 9.46E-09 | TRUE | 8.00E-09 | TRUE | FALSE | -2.664403232 |
| **s100a11** | LV end systolic volume \| Instance 2(participant - p24101_i2)_BSA_indexed_corrected | -1.294341156 | 0.252593091 | -0.09090182161 | 1.84E-09 | 0.2148475799 | -0.09090182161 | 1.84E-09 | 4358 | 4.25E-08 | TRUE | 2.69E-05 | TRUE | 2.27E-05 | TRUE | FALSE | -1.294341156 |
| **s100a11** | RV end diastolic volume \| Instance 2_BSA_indexed_corrected | -2.832150139 | 0.1265036805 | -0.1073083044 | 1.23E-12 | 0.397579543 | -0.1073083044 | 1.23E-12 | 4358 | 5.14E-11 | TRUE | 1.79E-08 | TRUE | 1.52E-08 | TRUE | FALSE | -2.832150139 |
| **s100a11** | RV end systolic volume \| Instance 2_BSA_indexed_corrected | -1.462992184 | 0.1573625453 | -0.09318833586 | 7.12E-10 | 0.2368331861 | -0.09318833586 | 7.12E-10 | 4358 | 1.83E-08 | TRUE | 1.04E-05 | TRUE | 8.81E-06 | TRUE | FALSE | -1.462992184 |
| **s100a12** | LV end diastolic volume \| Instance 2(participant - p24100_i2)_BSA_indexed_corrected | -1.120901381 | 0.2164723416 | -0.07798747582 | 2.84E-07 | 0.2179845307 | -0.07798747582 | 2.84E-07 | 4323 | 4.51E-06 | TRUE | 0.00415028072 | TRUE | 0.003510307433 | TRUE | FALSE | -1.120901381 |
| **scarb2** | LV end diastolic volume \| Instance 2(participant - p24100_i2)_BSA_indexed_corrected | -4.826427279 | 0.01271855837 | -0.1214332977 | 1.22E-15 | 0.6008596767 | -0.1214332977 | 1.22E-15 | 4313 | 8.80E-14 | TRUE | 1.79E-11 | TRUE | 1.51E-11 | TRUE | FALSE | -4.826427279 |
| **scarb2** | LV end systolic volume \| Instance 2(participant - p24101_i2)_BSA_indexed_corrected | -2.26010218 | 0.1607670725 | -0.09787951212 | 1.18E-10 | 0.3499910028 | -0.09787951212 | 1.18E-10 | 4313 | 3.48E-09 | TRUE | 1.73E-06 | TRUE | 1.46E-06 | TRUE | FALSE | -2.26010218 |
| **scarb2** | RV end diastolic volume \| Instance 2_BSA_indexed_corrected | -5.594633709 | -0.1661370101 | -0.1310229932 | 5.65E-18 | 0.6447257105 | -0.1310229932 | 5.65E-18 | 4313 | 5.54E-16 | TRUE | 8.26E-14 | TRUE | 6.99E-14 | TRUE | FALSE | -5.594633709 |
| **scarb2** | RV end systolic volume \| Instance 2_BSA_indexed_corrected | -2.786870964 | 0.01224316567 | -0.1097210309 | 4.99E-13 | 0.3845102623 | -0.1097210309 | 4.99E-13 | 4313 | 2.31E-11 | TRUE | 7.30E-09 | TRUE | 6.18E-09 | TRUE | FALSE | -2.786870964 |
| **scg2** | LV cardiac output \| Instance 2_BSA_indexed_corrected | -0.1388896155 | -0.02398445414 | -0.08528725199 | 1.62E-08 | 0.0245420009 | -0.08528725199 | 1.62E-08 | 4373 | 3.22E-07 | TRUE | 0.0002366359298 | TRUE | 0.0002001466695 | TRUE | FALSE | -0.1388896155 |
| **scg2** | LV end diastolic volume \| Instance 2(participant - p24100_i2)_BSA_indexed_corrected | -3.062974753 | 0.2649271847 | -0.08392227068 | 2.73E-08 | 0.5500992085 | -0.08392227068 | 2.73E-08 | 4373 | 5.22E-07 | TRUE | 0.0003994861825 | TRUE | 0.0003378854132 | TRUE | FALSE | -3.062974753 |
| **scg2** | RV end diastolic volume \| Instance 2_BSA_indexed_corrected | -3.669544936 | 0.1388880847 | -0.09346105265 | 5.93E-10 | 0.5912702554 | -0.09346105265 | 5.93E-10 | 4373 | 1.55E-08 | TRUE | 8.68E-06 | TRUE | 7.34E-06 | TRUE | FALSE | -3.669544936 |
| **scg3** | LV cardiac output \| Instance 2_BSA_indexed_corrected | -0.1144047972 | -0.01666281412 | -0.07698997985 | 4.19E-07 | 0.02257520137 | -0.07698997985 | 4.19E-07 | 4309 | 6.43E-06 | TRUE | 0.006137062795 | TRUE | 0.005190727712 | TRUE | FALSE | -0.1144047972 |
| **scgb3a2** | LV end diastolic volume \| Instance 2(participant - p24100_i2)_BSA_indexed_corrected | 0.9558047028 | 0.3262791178 | 0.07161458828 | 2.51E-06 | 0.2027741039 | 0.07161458828 | 2.51E-06 | 4312 | 3.32E-05 | TRUE | 0.03672197184 | TRUE | 0.03105944379 | TRUE | FALSE | 0.9558047028 |
| **scly** | LV end diastolic volume \| Instance 2(participant - p24100_i2)_BSA_indexed_corrected | -2.762944254 | 0.3201776146 | -0.1259356543 | 6.33E-17 | 0.3292012723 | -0.1259356543 | 6.33E-17 | 4373 | 5.58E-15 | TRUE | 9.26E-13 | TRUE | 7.84E-13 | TRUE | FALSE | -2.762944254 |
| **scly** | LV end systolic volume \| Instance 2(participant - p24101_i2)_BSA_indexed_corrected | -1.230873273 | 0.3011176004 | -0.09666524638 | 1.50E-10 | 0.1916964083 | -0.09666524638 | 1.50E-10 | 4373 | 4.32E-09 | TRUE | 2.19E-06 | TRUE | 1.85E-06 | TRUE | FALSE | -1.230873273 |
| **scly** | RV end diastolic volume \| Instance 2_BSA_indexed_corrected | -2.73121434 | 0.2119040199 | -0.1157222884 | 1.64E-14 | 0.3545853279 | -0.1157222884 | 1.64E-14 | 4373 | 9.67E-13 | TRUE | 2.40E-10 | TRUE | 2.03E-10 | TRUE | FALSE | -2.73121434 |
| **scly** | RV end systolic volume \| Instance 2_BSA_indexed_corrected | -1.321856026 | 0.203946854 | -0.09416497993 | 4.40E-10 | 0.2113831145 | -0.09416497993 | 4.40E-10 | 4373 | 1.18E-08 | TRUE | 6.44E-06 | TRUE | 5.45E-06 | TRUE | FALSE | -1.321856026 |
| **sele** | LV end diastolic volume \| Instance 2(participant - p24100_i2)_BSA_indexed_corrected | -2.698915448 | 0.05071686301 | -0.1266836809 | 4.07E-17 | 0.3195697453 | -0.1266836809 | 4.07E-17 | 4375 | 3.68E-15 | TRUE | 5.96E-13 | TRUE | 5.04E-13 | TRUE | FALSE | -2.698915448 |
| **sele** | LV end systolic volume \| Instance 2(participant - p24101_i2)_BSA_indexed_corrected | -1.085493585 | 0.1903285785 | -0.08777435899 | 6.05E-09 | 0.1862903687 | -0.08777435899 | 6.05E-09 | 4375 | 1.28E-07 | TRUE | 8.86E-05 | TRUE | 7.49E-05 | TRUE | FALSE | -1.085493585 |
| **sele** | RV end diastolic volume \| Instance 2_BSA_indexed_corrected | -2.657610167 | -0.05422926118 | -0.1159511379 | 1.43E-14 | 0.3442605411 | -0.1159511379 | 1.43E-14 | 4375 | 8.57E-13 | TRUE | 2.10E-10 | TRUE | 1.78E-10 | TRUE | FALSE | -2.657610167 |
| **sele** | RV end systolic volume \| Instance 2_BSA_indexed_corrected | -1.091372981 | 0.0948796589 | -0.08006985395 | 1.14E-07 | 0.2054554994 | -0.08006985395 | 1.14E-07 | 4375 | 1.95E-06 | TRUE | 0.001665428715 | TRUE | 0.001408619607 | TRUE | FALSE | -1.091372981 |
| **sema3f** | LV end diastolic volume \| Instance 2(participant - p24100_i2)_BSA_indexed_corrected | -5.961030571 | 0.04007125418 | -0.1427403024 | 2.37E-21 | 0.625050308 | -0.1427403024 | 2.37E-21 | 4375 | 4.03E-19 | TRUE | 3.47E-17 | TRUE | 2.93E-17 | TRUE | FALSE | -5.961030571 |
| **sema3f** | LV end systolic volume \| Instance 2(participant - p24101_i2)_BSA_indexed_corrected | -2.703761137 | 0.1699689811 | -0.1115327975 | 1.38E-13 | 0.3642988735 | -0.1115327975 | 1.38E-13 | 4375 | 6.85E-12 | TRUE | 2.02E-09 | TRUE | 1.71E-09 | TRUE | FALSE | -2.703761137 |
| **sema3f** | RV end diastolic volume \| Instance 2_BSA_indexed_corrected | -5.954222441 | -0.06914394758 | -0.1325264799 | 1.34E-18 | 0.6734183554 | -0.1325264799 | 1.34E-18 | 4375 | 1.47E-16 | TRUE | 1.96E-14 | TRUE | 1.66E-14 | TRUE | FALSE | -5.954222441 |
| **sema3f** | RV end systolic volume \| Instance 2_BSA_indexed_corrected | -3.21083625 | 0.04855802831 | -0.1201732661 | 1.52E-15 | 0.4011083777 | -0.1201732661 | 1.52E-15 | 4375 | 1.06E-13 | TRUE | 2.22E-11 | TRUE | 1.88E-11 | TRUE | FALSE | -3.21083625 |
| **serpinb8** | LV end diastolic volume \| Instance 2(participant - p24100_i2)_BSA_indexed_corrected | -1.917743437 | 0.2390658156 | -0.09873577514 | 6.39E-11 | 0.2927488714 | -0.09873577514 | 6.39E-11 | 4361 | 1.98E-09 | TRUE | 9.34E-07 | TRUE | 7.90E-07 | TRUE | FALSE | -1.917743437 |
| **serpinb8** | LV end systolic volume \| Instance 2(participant - p24101_i2)_BSA_indexed_corrected | -0.8784519434 | 0.2582261925 | -0.07785721278 | 2.63E-07 | 0.1703748255 | -0.07785721278 | 2.63E-07 | 4361 | 4.21E-06 | TRUE | 0.003854523016 | TRUE | 0.003260155567 | TRUE | FALSE | -0.8784519434 |
| **serpinb8** | RV end diastolic volume \| Instance 2_BSA_indexed_corrected | -2.122031449 | 0.1185180662 | -0.1015305514 | 1.81E-11 | 0.3149282116 | -0.1015305514 | 1.81E-11 | 4361 | 6.30E-10 | TRUE | 2.65E-07 | TRUE | 2.24E-07 | TRUE | FALSE | -2.122031449 |
| **serpinb8** | RV end systolic volume \| Instance 2_BSA_indexed_corrected | -1.104801628 | 0.1486785501 | -0.08887146527 | 4.13E-09 | 0.1875455063 | -0.08887146527 | 4.13E-09 | 4361 | 9.05E-08 | TRUE | 6.04E-05 | TRUE | 5.11E-05 | TRUE | FALSE | -1.104801628 |
| **sez6l** | LV cardiac output \| Instance 2_BSA_indexed_corrected | -0.1638593292 | -0.01866602021 | -0.09140797419 | 1.61E-09 | 0.02710318543 | -0.09140797419 | 1.61E-09 | 4340 | 3.76E-08 | TRUE | 2.36E-05 | TRUE | 1.99E-05 | TRUE | FALSE | -0.1638593292 |
| **sez6l** | LV end diastolic volume \| Instance 2(participant - p24100_i2)_BSA_indexed_corrected | -3.006756034 | 0.3719900637 | -0.07487554101 | 7.88E-07 | 0.6079846517 | -0.07487554101 | 7.88E-07 | 4340 | 1.14E-05 | TRUE | 0.01153451409 | TRUE | 0.009755892016 | TRUE | FALSE | -3.006756034 |
| **sez6l** | RV end diastolic volume \| Instance 2_BSA_indexed_corrected | -3.323044421 | 0.2688592615 | -0.07694015914 | 3.88E-07 | 0.6538061586 | -0.07694015914 | 3.88E-07 | 4340 | 6.00E-06 | TRUE | 0.005675649434 | TRUE | 0.004800464292 | TRUE | FALSE | -3.323044421 |
| **sfrp1** | LV end diastolic volume \| Instance 2(participant - p24100_i2)_BSA_indexed_corrected | -2.803935329 | 0.06386229482 | -0.108835344 | 5.81E-13 | 0.387986635 | -0.108835344 | 5.81E-13 | 4359 | 2.63E-11 | TRUE | 8.49E-09 | TRUE | 7.18E-09 | TRUE | FALSE | -2.803935329 |
| **sfrp1** | LV end systolic volume \| Instance 2(participant - p24101_i2)_BSA_indexed_corrected | -1.098613126 | 0.2085955449 | -0.07343828717 | 1.21E-06 | 0.2260238043 | -0.07343828717 | 1.21E-06 | 4359 | 1.70E-05 | TRUE | 0.01772313341 | TRUE | 0.01499022624 | TRUE | FALSE | -1.098613126 |
| **sfrp1** | RV end diastolic volume \| Instance 2_BSA_indexed_corrected | -2.897713665 | -0.06011438419 | -0.1045923109 | 4.44E-12 | 0.4174200904 | -0.1045923109 | 4.44E-12 | 4359 | 1.71E-10 | TRUE | 6.49E-08 | TRUE | 5.49E-08 | TRUE | FALSE | -2.897713665 |
| **sftpd** | RV end diastolic volume \| Instance 2_BSA_indexed_corrected | -1.364881162 | 0.1932625292 | -0.07572093513 | 5.32E-07 | 0.2717943978 | -0.07572093513 | 5.32E-07 | 4375 | 8.03E-06 | TRUE | 0.007789621205 | TRUE | 0.006588461615 | TRUE | FALSE | -1.364881162 |
| **sftpd** | RV end systolic volume \| Instance 2_BSA_indexed_corrected | -0.7642799178 | 0.1890226 | -0.07129936747 | 2.35E-06 | 0.1616852221 | -0.07129936747 | 2.35E-06 | 4375 | 3.12E-05 | TRUE | 0.03439046639 | TRUE | 0.02908745648 | TRUE | FALSE | -0.7642799178 |
| **sit1** | LV end diastolic volume \| Instance 2(participant - p24100_i2)_BSA_indexed_corrected | -2.10505828 | 0.3436958398 | -0.1040966405 | 8.50E-12 | 0.3073535301 | -0.1040966405 | 8.50E-12 | 4284 | 3.13E-10 | TRUE | 1.24E-07 | TRUE | 1.05E-07 | TRUE | FALSE | -2.10505828 |
| **sit1** | LV end systolic volume \| Instance 2(participant - p24101_i2)_BSA_indexed_corrected | -0.9060971344 | 0.3009152067 | -0.07727214192 | 4.11E-07 | 0.1786602781 | -0.07727214192 | 4.11E-07 | 4284 | 6.33E-06 | TRUE | 0.006014745165 | TRUE | 0.00508727146 | TRUE | FALSE | -0.9060971344 |
| **sit1** | RV end diastolic volume \| Instance 2_BSA_indexed_corrected | -1.852449873 | 0.2331409774 | -0.08503060448 | 2.49E-08 | 0.3317203728 | -0.08503060448 | 2.49E-08 | 4284 | 4.81E-07 | TRUE | 0.0003643920177 | TRUE | 0.0003082027686 | TRUE | FALSE | -1.852449873 |
| **skap1** | LV end diastolic volume \| Instance 2(participant - p24100_i2)_BSA_indexed_corrected | -1.599770354 | 0.3028808289 | -0.104525141 | 4.19E-12 | 0.2301768515 | -0.104525141 | 4.19E-12 | 4375 | 1.63E-10 | TRUE | 6.13E-08 | TRUE | 5.18E-08 | TRUE | FALSE | -1.599770354 |
| **skap1** | LV end systolic volume \| Instance 2(participant - p24101_i2)_BSA_indexed_corrected | -0.7338133868 | 0.2889152374 | -0.08259576095 | 4.48E-08 | 0.1338910864 | -0.08259576095 | 4.48E-08 | 4375 | 8.19E-07 | TRUE | 0.0006547541855 | TRUE | 0.0005537910901 | TRUE | FALSE | -0.7338133868 |
| **skap1** | RV end diastolic volume \| Instance 2_BSA_indexed_corrected | -1.466292618 | 0.1974911907 | -0.08905044823 | 3.63E-09 | 0.2480081552 | -0.08905044823 | 3.63E-09 | 4375 | 8.02E-08 | TRUE | 5.31E-05 | TRUE | 4.49E-05 | TRUE | FALSE | -1.466292618 |
| **slc39a5** | LV end diastolic volume \| Instance 2(participant - p24100_i2)_BSA_indexed_corrected | -2.062124785 | 0.1956988515 | -0.107100174 | 1.58E-12 | 0.2909213849 | -0.107100174 | 1.58E-12 | 4332 | 6.52E-11 | TRUE | 2.31E-08 | TRUE | 1.95E-08 | TRUE | FALSE | -2.062124785 |
| **slc39a5** | LV end systolic volume \| Instance 2(participant - p24101_i2)_BSA_indexed_corrected | -1.008443983 | 0.237713971 | -0.09028961823 | 2.63E-09 | 0.1690411849 | -0.09028961823 | 2.63E-09 | 4332 | 5.94E-08 | TRUE | 3.85E-05 | TRUE | 3.26E-05 | TRUE | FALSE | -1.008443983 |
| **slc39a5** | RV end diastolic volume \| Instance 2_BSA_indexed_corrected | -1.958390247 | 0.1001922213 | -0.09455863955 | 4.49E-10 | 0.3133312818 | -0.09455863955 | 4.49E-10 | 4332 | 1.20E-08 | TRUE | 6.57E-06 | TRUE | 5.56E-06 | TRUE | FALSE | -1.958390247 |
| **slitrk2** | LV end diastolic volume \| Instance 2(participant - p24100_i2)_BSA_indexed_corrected | -2.522752512 | 0.2459033252 | -0.09050657114 | 2.01E-09 | 0.4197770807 | -0.09050657114 | 2.01E-09 | 4375 | 4.61E-08 | TRUE | 2.94E-05 | TRUE | 2.49E-05 | TRUE | FALSE | -2.522752512 |
| **slitrk2** | LV end systolic volume \| Instance 2(participant - p24101_i2)_BSA_indexed_corrected | -1.190131618 | 0.2613808265 | -0.07355453745 | 1.11E-06 | 0.2440157008 | -0.07355453745 | 1.11E-06 | 4375 | 1.57E-05 | TRUE | 0.01629519144 | TRUE | 0.01378247292 | TRUE | FALSE | -1.190131618 |
| **slitrk2** | RV end diastolic volume \| Instance 2_BSA_indexed_corrected | -2.830182303 | 0.1232777664 | -0.09437831508 | 3.98E-10 | 0.451449939 | -0.09437831508 | 3.98E-10 | 4375 | 1.07E-08 | TRUE | 5.83E-06 | TRUE | 4.93E-06 | TRUE | FALSE | -2.830182303 |
| **slitrk2** | RV end systolic volume \| Instance 2_BSA_indexed_corrected | -1.520081288 | 0.1525813448 | -0.08523868115 | 1.64E-08 | 0.2686933272 | -0.08523868115 | 1.64E-08 | 4375 | 3.26E-07 | TRUE | 0.0002393165029 | TRUE | 0.0002024138982 | TRUE | FALSE | -1.520081288 |
| **smad5** | LV end diastolic volume \| Instance 2(participant - p24100_i2)_BSA_indexed_corrected | -5.263315361 | 0.08419133033 | -0.1046910396 | 3.87E-12 | 0.7560787329 | -0.1046910396 | 3.87E-12 | 4375 | 1.52E-10 | TRUE | 5.67E-08 | TRUE | 4.79E-08 | TRUE | FALSE | -5.263315361 |
| **smad5** | LV end systolic volume \| Instance 2(participant - p24101_i2)_BSA_indexed_corrected | -2.61755984 | 0.1782199509 | -0.08969242164 | 2.80E-09 | 0.4395387561 | -0.08969242164 | 2.80E-09 | 4375 | 6.29E-08 | TRUE | 4.10E-05 | TRUE | 3.47E-05 | TRUE | FALSE | -2.61755984 |
| **smad5** | RV end diastolic volume \| Instance 2_BSA_indexed_corrected | -5.808804735 | -0.05324214698 | -0.1073962809 | 1.06E-12 | 0.8131837163 | -0.1073962809 | 1.06E-12 | 4375 | 4.53E-11 | TRUE | 1.55E-08 | TRUE | 1.31E-08 | TRUE | FALSE | -5.808804735 |
| **smad5** | RV end systolic volume \| Instance 2_BSA_indexed_corrected | -2.632228107 | 0.08268038499 | -0.08183481148 | 5.95E-08 | 0.4847709478 | -0.08183481148 | 5.95E-08 | 4375 | 1.06E-06 | TRUE | 0.0008699351599 | TRUE | 0.0007357911583 | TRUE | FALSE | -2.632228107 |
| **smoc1** | LV end diastolic volume \| Instance 2(participant - p24100_i2)_BSA_indexed_corrected | -3.80755513 | -0.05816728838 | -0.1169157619 | 8.65E-15 | 0.4890966341 | -0.1169157619 | 8.65E-15 | 4375 | 5.27E-13 | TRUE | 1.27E-10 | TRUE | 1.07E-10 | TRUE | FALSE | -3.80755513 |
| **smoc1** | LV end systolic volume \| Instance 2(participant - p24101_i2)_BSA_indexed_corrected | -1.749276652 | 0.12300533 | -0.09253251669 | 8.69E-10 | 0.2846473262 | -0.09253251669 | 8.69E-10 | 4375 | 2.16E-08 | TRUE | 1.27E-05 | TRUE | 1.07E-05 | TRUE | FALSE | -1.749276652 |
| **smoc1** | RV end diastolic volume \| Instance 2_BSA_indexed_corrected | -3.968097534 | -0.1850770685 | -0.1132560853 | 5.77E-14 | 0.5264143272 | -0.1132560853 | 5.77E-14 | 4375 | 3.11E-12 | TRUE | 8.44E-10 | TRUE | 7.14E-10 | TRUE | FALSE | -3.968097534 |
| **smoc1** | RV end systolic volume \| Instance 2_BSA_indexed_corrected | -2.007751752 | 0.0003019153254 | -0.09636100509 | 1.69E-10 | 0.3136123998 | -0.09636100509 | 1.69E-10 | 4375 | 4.84E-09 | TRUE | 2.48E-06 | TRUE | 2.10E-06 | TRUE | FALSE | -2.007751752 |
| **smoc2** | LV end diastolic volume \| Instance 2(participant - p24100_i2)_BSA_indexed_corrected | -2.117969757 | 0.2540467253 | -0.0762190138 | 5.26E-07 | 0.4215502932 | -0.0762190138 | 5.26E-07 | 4322 | 7.96E-06 | TRUE | 0.007693158366 | TRUE | 0.006506873346 | TRUE | FALSE | -2.117969757 |
| **sncg** | LV end diastolic volume \| Instance 2(participant - p24100_i2)_BSA_indexed_corrected | -1.900938731 | -0.03318982941 | -0.1193951973 | 2.31E-15 | 0.2390417544 | -0.1193951973 | 2.31E-15 | 4375 | 1.58E-13 | TRUE | 3.38E-11 | TRUE | 2.86E-11 | TRUE | FALSE | -1.900938731 |

| | | | | | | | | | | | | | | | | | |
|---|---|---|---|---|---|---|---|---|---|---|---|---|---|---|---|---|---|
| **sncg** | LV end systolic volume \| Instance 2(participant - p24101_i2)_BSA_indexed_corrected | -0.7618482845 | 0.1571305939 | -0.08243207706 | 4.76E-08 | 0.1392842309 | -0.08243207706 | 4.76E-08 | 4375 | 8.70E-07 | TRUE | 0.0006961713935 | TRUE | 0.0005888217646 | TRUE | FALSE | -0.7618482845 |
| **sncg** | RV end diastolic volume \| Instance 2_BSA_indexed_corrected | -2.347479199 | -0.2334840013 | -0.137048042 | 8.60E-20 | 0.2565794652 | -0.137048042 | 8.60E-20 | 4375 | 1.23E-17 | TRUE | 1.26E-15 | TRUE | 1.06E-15 | TRUE | FALSE | -2.347479199 |
| **sncg** | RV end systolic volume \| Instance 2_BSA_indexed_corrected | -1.003428518 | 0.01325947432 | -0.09850757365 | 6.59E-11 | 0.153288526 | -0.09850757365 | 6.59E-11 | 4375 | 2.04E-09 | TRUE | 9.64E-07 | TRUE | 8.16E-07 | TRUE | FALSE | -1.003428518 |
| **sorcs2** | LV cardiac output \| Instance 2_BSA_indexed_corrected | -0.1313233613 | -0.02567994246 | -0.09051490712 | 2.00E-09 | 0.02184971302 | -0.09051490712 | 2.00E-09 | 4375 | 4.61E-08 | TRUE | 2.93E-05 | TRUE | 2.48E-05 | TRUE | FALSE | -0.1313233613 |
| **sorcs2** | LV end diastolic volume \| Instance 2(participant - p24100_i2)_BSA_indexed_corrected | -3.321472765 | 0.2029556396 | -0.102175512 | 1.25E-11 | 0.4890071807 | -0.102175512 | 1.25E-11 | 4375 | 4.44E-10 | TRUE | 1.83E-07 | TRUE | 1.55E-07 | TRUE | FALSE | -3.321472765 |
| **sorcs2** | LV end systolic volume \| Instance 2(participant - p24101_i2)_BSA_indexed_corrected | -1.429888942 | 0.2473114188 | -0.07577526176 | 5.23E-07 | 0.2845343442 | -0.07577526176 | 5.23E-07 | 4375 | 7.92E-06 | TRUE | 0.007644786785 | TRUE | 0.006465960663 | TRUE | FALSE | -1.429888942 |
| **sorcs2** | RV end diastolic volume \| Instance 2_BSA_indexed_corrected | -3.091462768 | 0.1037745484 | -0.08839597523 | 4.72E-09 | 0.5267910294 | -0.08839597523 | 4.72E-09 | 4375 | 1.02E-07 | TRUE | 6.91E-05 | TRUE | 5.84E-05 | TRUE | FALSE | -3.091462768 |
| **sord** | LV end diastolic volume \| Instance 2(participant - p24100_i2)_BSA_indexed_corrected | -1.952796044 | 0.1990090048 | -0.1234592887 | 2.64E-16 | 0.2375505095 | -0.1234592887 | 2.64E-16 | 4368 | 2.14E-14 | TRUE | 3.87E-12 | TRUE | 3.27E-12 | TRUE | FALSE | -1.952796044 |
| **sord** | LV end systolic volume \| Instance 2(participant - p24101_i2)_BSA_indexed_corrected | -0.8622416459 | 0.2487034148 | -0.09394324273 | 4.95E-10 | 0.1382918796 | -0.09394324273 | 4.95E-10 | 4368 | 1.32E-08 | TRUE | 7.24E-06 | TRUE | 6.12E-06 | TRUE | FALSE | -0.8622416459 |
| **sord** | RV end diastolic volume \| Instance 2_BSA_indexed_corrected | -2.023159398 | 0.07869866502 | -0.118969187 | 3.06E-15 | 0.2555396842 | -0.118969187 | 3.06E-15 | 4368 | 2.05E-13 | TRUE | 4.47E-11 | TRUE | 3.78E-11 | TRUE | FALSE | -2.023159398 |
| **sord** | RV end systolic volume \| Instance 2_BSA_indexed_corrected | -0.9128695282 | 0.1442390218 | -0.09023629883 | 2.31E-09 | 0.1524790551 | -0.09023629883 | 2.31E-09 | 4368 | 5.25E-08 | TRUE | 3.38E-05 | TRUE | 2.86E-05 | TRUE | FALSE | -0.9128695282 |
| **sost** | LV end diastolic volume \| Instance 2(participant - p24100_i2)_BSA_indexed_corrected | -3.077781789 | 0.210004088 | -0.1169611943 | 8.51E-15 | 0.3952436415 | -0.1169611943 | 8.51E-15 | 4374 | 5.23E-13 | TRUE | 1.24E-10 | TRUE | 1.05E-10 | TRUE | FALSE | -3.077781789 |
| **sost** | LV end systolic volume \| Instance 2(participant - p24101_i2)_BSA_indexed_corrected | -1.276749406 | 0.2517721319 | -0.08357284942 | 3.11E-08 | 0.2302387448 | -0.08357284942 | 3.11E-08 | 4374 | 5.88E-07 | TRUE | 0.000454553551 | TRUE | 0.0003844613934 | TRUE | FALSE | -1.276749406 |
| **sost** | RV end diastolic volume \| Instance 2_BSA_indexed_corrected | -3.08430471 | 0.09967627351 | -0.1089412656 | 5.03E-13 | 0.4256299914 | -0.1089412656 | 5.03E-13 | 4374 | 2.32E-11 | TRUE | 7.36E-09 | TRUE | 6.23E-09 | TRUE | FALSE | -3.08430471 |
| **sost** | RV end systolic volume \| Instance 2_BSA_indexed_corrected | -1.342617911 | 0.1544827853 | -0.07974090129 | 1.29E-07 | 0.2538318158 | -0.07974090129 | 1.29E-07 | 4374 | 2.17E-06 | TRUE | 0.001883113794 | TRUE | 0.001592737647 | TRUE | FALSE | -1.342617911 |
| **spink5** | LV cardiac output \| Instance 2_BSA_indexed_corrected | -0.1059493566 | -0.01555254051 | -0.0710431552 | 3.03E-06 | 0.02266152279 | -0.0710431552 | 3.03E-06 | 4311 | 3.94E-05 | TRUE | 0.04425604864 | TRUE | 0.03743176594 | TRUE | FALSE | -0.1059493566 |
| **spon1** | LV end diastolic volume \| Instance 2(participant - p24100_i2)_BSA_indexed_corrected | -2.908584806 | 0.1161821195 | -0.08385589699 | 3.53E-08 | 0.5266575979 | -0.08385589699 | 3.53E-08 | 4309 | 6.60E-07 | TRUE | 0.0005170848725 | TRUE | 0.0004373503852 | TRUE | FALSE | -2.908584806 |
| **spon1** | RV end diastolic volume \| Instance 2_BSA_indexed_corrected | -2.759723908 | 0.0260208428 | -0.074024836 | 1.15E-06 | 0.5665099962 | -0.074024836 | 1.15E-06 | 4309 | 1.61E-05 | TRUE | 0.01678950919 | TRUE | 0.01420056688 | TRUE | FALSE | -2.759723908 |
| **srp14** | LV end diastolic volume \| Instance 2(participant - p24100_i2)_BSA_indexed_corrected | -1.773859525 | 0.1870119458 | -0.09747381289 | 1.04E-10 | 0.2738851717 | -0.09747381289 | 1.04E-10 | 4375 | 3.10E-09 | TRUE | 1.52E-06 | TRUE | 1.29E-06 | TRUE | FALSE | -1.773859525 |
| **srp14** | LV end systolic volume \| Instance 2(participant - p24101_i2)_BSA_indexed_corrected | -0.795065126 | 0.2375071815 | -0.07526281287 | 6.23E-07 | 0.1592938447 | -0.07526281287 | 6.23E-07 | 4375 | 9.23E-06 | TRUE | 0.009120775579 | TRUE | 0.007714351985 | TRUE | FALSE | -0.795065126 |
| **srp14** | RV end diastolic volume \| Instance 2_BSA_indexed_corrected | -1.595160537 | 0.09416251938 | -0.08147520681 | 6.79E-08 | 0.2950821318 | -0.08147520681 | 6.79E-08 | 4375 | 1.20E-06 | TRUE | 0.0009940930175 | TRUE | 0.0008408038742 | TRUE | FALSE | -1.595160537 |
| **ssc4d** | LV end diastolic volume \| Instance 2(participant - p24100_i2)_BSA_indexed_corrected | -1.378281939 | 0.4988883014 | -0.1658134444 | 2.46E-28 | 0.1239722178 | -0.1658134444 | 2.46E-28 | 4374 | 1.12E-25 | TRUE | 3.60E-24 | TRUE | 3.04E-24 | TRUE | FALSE | -1.378281939 |
| **ssc4d** | LV end systolic volume \| Instance 2(participant - p24101_i2)_BSA_indexed_corrected | -0.6529911702 | 0.3800286495 | -0.1353145698 | 2.52E-19 | 0.07231191238 | -0.1353145698 | 2.52E-19 | 4374 | 3.12E-17 | TRUE | 3.68E-15 | TRUE | 3.12E-15 | TRUE | FALSE | -0.6529911702 |
| **ssc4d** | RV end diastolic volume \| Instance 2_BSA_indexed_corrected | -1.337384248 | 0.3846316574 | -0.1495440196 | 2.69E-23 | 0.1337321418 | -0.1495440196 | 2.69E-23 | 4374 | 6.26E-21 | TRUE | 3.94E-19 | TRUE | 3.33E-19 | TRUE | FALSE | -1.337384248 |
| **ssc4d** | RV end systolic volume \| Instance 2_BSA_indexed_corrected | -0.6036046593 | 0.2807468464 | -0.1134903653 | 5.15E-14 | 0.07991687156 | -0.1134903653 | 5.15E-14 | 4374 | 2.79E-12 | TRUE | 7.54E-10 | TRUE | 6.37E-10 | TRUE | FALSE | -0.6036046593 |
| **ssc5d** | LV end diastolic volume \| Instance 2(participant - p24100_i2)_BSA_indexed_corrected | -3.481750693 | 0.0683423097 | -0.1397482344 | 2.68E-20 | 0.3752981801 | -0.1397482344 | 2.68E-20 | 4323 | 3.96E-18 | TRUE | 3.92E-16 | TRUE | 3.32E-16 | TRUE | FALSE | -3.481750693 |
| **ssc5d** | LV end systolic volume \| Instance 2(participant - p24101_i2)_BSA_indexed_corrected | -1.419756412 | 0.2001624608 | -0.09813645563 | 1.01E-10 | 0.219023093 | -0.09813645563 | 1.01E-10 | 4323 | 3.00E-09 | TRUE | 1.47E-06 | TRUE | 1.24E-06 | TRUE | FALSE | -1.419756412 |
| **ssc5d** | RV end diastolic volume \| Instance 2_BSA_indexed_corrected | -3.792550968 | -0.06801520126 | -0.1416183769 | 8.29E-21 | 0.4032927005 | -0.1416183769 | 8.29E-21 | 4323 | 1.30E-18 | TRUE | 1.21E-16 | TRUE | 1.03E-16 | TRUE | FALSE | -3.792550968 |
| **ssc5d** | RV end systolic volume \| Instance 2_BSA_indexed_corrected | -1.643797013 | 0.09584413184 | -0.103233253 | 1.02E-11 | 0.2409405298 | -0.103233253 | 1.02E-11 | 4323 | 3.67E-10 | TRUE | 1.49E-07 | TRUE | 1.26E-07 | TRUE | FALSE | -1.643797013 |
| **st3gal1** | LV end diastolic volume \| Instance 2(participant - p24100_i2)_BSA_indexed_corrected | 2.657284831 | 0.2504769191 | 0.1079674145 | 8.05E-13 | 0.3700066437 | 0.1079674145 | 8.05E-13 | 4375 | 3.53E-11 | TRUE | 1.18E-08 | TRUE | 9.96E-09 | TRUE | FALSE | 2.657284831 |
| **st3gal1** | LV end systolic volume \| Instance 2(participant - p24101_i2)_BSA_indexed_corrected | 1.469192229 | 0.2552192232 | 0.1028353016 | 9.23E-12 | 0.2149008215 | 0.1028353016 | 9.23E-12 | 4375 | 3.38E-10 | TRUE | 1.35E-07 | TRUE | 1.14E-07 | TRUE | FALSE | 1.469192229 |
| **st3gal1** | RV end diastolic volume \| Instance 2_BSA_indexed_corrected | 2.717779406 | 0.1385699638 | 0.1026410628 | 1.01E-11 | 0.3982937592 | 0.1026410628 | 1.01E-11 | 4375 | 3.65E-10 | TRUE | 1.48E-07 | TRUE | 1.25E-07 | TRUE | FALSE | 2.717779406 |
| **st3gal1** | RV end systolic volume \| Instance 2_BSA_indexed_corrected | 1.537833106 | 0.1577801544 | 0.09766259115 | 9.58E-11 | 0.2369791502 | 0.09766259115 | 9.58E-11 | 4375 | 2.87E-09 | TRUE | 1.40E-06 | TRUE | 1.19E-06 | TRUE | FALSE | 1.537833106 |
| **st6gal1** | LV end diastolic volume \| Instance 2(participant - p24100_i2)_BSA_indexed_corrected | -2.964258562 | 0.1339138146 | -0.07707236297 | 3.33E-07 | 0.5798744008 | -0.07707236297 | 3.33E-07 | 4375 | 5.20E-06 | TRUE | 0.004865339371 | TRUE | 0.00411510404 | TRUE | FALSE | -2.964258562 |
| **st6gal1** | RV end diastolic volume \| Instance 2_BSA_indexed_corrected | -3.650026911 | -0.02634692698 | -0.08821267698 | 5.08E-09 | 0.6232739723 | -0.08821267698 | 5.08E-09 | 4375 | 1.09E-07 | TRUE | 7.44E-05 | TRUE | 6.29E-05 | TRUE | FALSE | -3.650026911 |
| **stc1** | LV end diastolic volume \| Instance 2(participant - p24100_i2)_BSA_indexed_corrected | -3.480426351 | 0.18539892 | -0.1357208443 | 2.18E-19 | 0.3847296612 | -0.1357208443 | 2.18E-19 | 4363 | 2.80E-17 | TRUE | 3.19E-15 | TRUE | 2.70E-15 | TRUE | FALSE | -3.480426351 |
| **stc1** | LV end systolic volume \| Instance 2(participant - p24101_i2)_BSA_indexed_corrected | -1.435782442 | 0.242617096 | -0.09653456939 | 1.66E-10 | 0.2241711217 | -0.09653456939 | 1.66E-10 | 4363 | 4.76E-09 | TRUE | 2.43E-06 | TRUE | 2.06E-06 | TRUE | FALSE | -1.435782442 |
| **stc1** | RV end diastolic volume \| Instance 2_BSA_indexed_corrected | -3.906077527 | 0.0505989294 | -0.1414733996 | 6.04E-21 | 0.4138879681 | -0.1414733996 | 6.04E-21 | 4363 | 9.93E-19 | TRUE | 8.83E-17 | TRUE | 7.47E-17 | TRUE | FALSE | -3.906077527 |
| **stc1** | RV end systolic volume \| Instance 2_BSA_indexed_corrected | -1.704340874 | 0.1360850991 | -0.1038807616 | 6.06E-12 | 0.2470997318 | -0.1038807616 | 6.06E-12 | 4363 | 2.30E-10 | TRUE | 8.86E-08 | TRUE | 7.50E-08 | TRUE | FALSE | -1.704340874 |
| **stc2** | LV cardiac output \| Instance 2_BSA_indexed_corrected | -0.1551235398 | -0.01951349835 | -0.0823039106 | 6.45E-08 | 0.02864488362 | -0.0823039106 | 6.45E-08 | 4302 | 1.15E-06 | TRUE | 0.0009434623773 | TRUE | 0.0007979804787 | TRUE | FALSE | -0.1551235398 |
| **sult2a1** | LV end diastolic volume \| Instance 2(participant - p24100_i2)_BSA_indexed_corrected | -1.597662584 | 0.3999240167 | -0.077675547 | 3.08E-07 | 0.3116324483 | -0.077675547 | 3.08E-07 | 4332 | 4.85E-06 | TRUE | 0.00450082988 | TRUE | 0.003806801913 | TRUE | FALSE | -1.597662584 |
| **sult2a1** | RV end diastolic volume \| Instance 2_BSA_indexed_corrected | -1.831196353 | 0.3006348407 | -0.08276770517 | 4.89E-08 | 0.3350714061 | -0.08276770517 | 4.89E-08 | 4332 | 8.93E-07 | TRUE | 0.0007149062796 | TRUE | 0.0006046677312 | TRUE | FALSE | -1.831196353 |
| **sumf2** | LV end diastolic volume \| Instance 2(participant - p24100_i2)_BSA_indexed_corrected | -1.506959836 | 0.2110107662 | -0.07165118265 | 2.16E-06 | 0.3176636993 | -0.07165118265 | 2.16E-06 | 4363 | 2.90E-05 | TRUE | 0.03165352514 | TRUE | 0.02677255157 | TRUE | FALSE | -1.506959836 |
| **tafa5** | LV cardiac output \| Instance 2_BSA_indexed_corrected | -0.1545018087 | -0.02787624511 | -0.1022728492 | 1.20E-11 | 0.02272480731 | -0.1022728492 | 1.20E-11 | 4375 | 4.26E-10 | TRUE | 1.75E-07 | TRUE | 1.48E-07 | TRUE | FALSE | -0.1545018087 |
| **tafa5** | LV end diastolic volume \| Instance 2(participant - p24100_i2)_BSA_indexed_corrected | -4.610815972 | 0.1104112197 | -0.1362205354 | 1.43E-19 | 0.5070820318 | -0.1362205354 | 1.43E-19 | 4375 | 1.92E-17 | TRUE | 2.10E-15 | TRUE | 1.77E-15 | TRUE | FALSE | -4.610815972 |
| **tafa5** | LV end systolic volume \| Instance 2(participant - p24101_i2)_BSA_indexed_corrected | -2.137591773 | 0.1994397086 | -0.1087924042 | 5.38E-13 | 0.2953595497 | -0.1087924042 | 5.38E-13 | 4375 | 2.46E-11 | TRUE | 7.87E-09 | TRUE | 6.66E-09 | TRUE | FALSE | -2.137591773 |
| **tafa5** | RV end diastolic volume \| Instance 2_BSA_indexed_corrected | -5.240394538 | -0.03228746131 | -0.1439067362 | 1.12E-21 | 0.5449403568 | -0.1439067362 | 1.12E-21 | 4375 | 2.01E-19 | TRUE | 1.63E-17 | TRUE | 1.38E-17 | TRUE | FALSE | -5.240394538 |
| **tafa5** | RV end systolic volume \| Instance 2_BSA_indexed_corrected | -2.394558455 | 0.09112870073 | -0.1105744229 | 2.23E-13 | 0.3254690258 | -0.1105744229 | 2.23E-13 | 4375 | 1.07E-11 | TRUE | 3.26E-09 | TRUE | 2.76E-09 | TRUE | FALSE | -2.394558455 |
| **tfpi** | LV end diastolic volume \| Instance 2(participant - p24100_i2)_BSA_indexed_corrected | -3.547032033 | 0.1932278168 | -0.09997776971 | 4.69E-11 | 0.5376395726 | -0.09997776971 | 4.69E-11 | 4313 | 1.50E-09 | TRUE | 6.86E-07 | TRUE | 5.80E-07 | TRUE | FALSE | -3.547032033 |
| **tfpi** | LV end systolic volume \| Instance 2(participant - p24101_i2)_BSA_indexed_corrected | -1.513200241 | 0.2444419494 | -0.07339671224 | 1.40E-06 | 0.3131539057 | -0.07339671224 | 1.40E-06 | 4313 | 1.93E-05 | TRUE | 0.02044900651 | TRUE | 0.0172957697 | TRUE | FALSE | -1.513200241 |
| **tfpi** | RV end diastolic volume \| Instance 2_BSA_indexed_corrected | -3.259658098 | 0.08602036417 | -0.08546252661 | 1.90E-08 | 0.5787826187 | -0.08546252661 | 1.90E-08 | 4313 | 3.73E-07 | TRUE | 0.0002773129577 | TRUE | 0.0002345512996 | TRUE | FALSE | -3.259658098 |
| **tgfa** | LV end diastolic volume \| Instance 2(participant - p24100_i2)_BSA_indexed_corrected | -2.94773384 | 0.2677997536 | -0.1180434246 | 6.47E-15 | 0.3768386644 | -0.1180434246 | 6.47E-15 | 4332 | 4.10E-13 | TRUE | 9.47E-11 | TRUE | 8.01E-11 | TRUE | FALSE | -2.94773384 |
| **tgfa** | LV end systolic volume \| Instance 2(participant - p24101_i2)_BSA_indexed_corrected | -1.483344818 | 0.2715681434 | -0.1024015323 | 1.42E-11 | 0.2189791022 | -0.1024015323 | 1.42E-11 | 4332 | 5.00E-10 | TRUE | 2.08E-07 | TRUE | 1.76E-07 | TRUE | FALSE | -1.483344818 |
| **tgfa** | RV end diastolic volume \| Instance 2_BSA_indexed_corrected | -3.323530283 | 0.1510492007 | -0.1237313971 | 3.01E-16 | 0.4050658761 | -0.1237313971 | 3.01E-16 | 4332 | 2.41E-14 | TRUE | 4.41E-12 | TRUE | 3.73E-12 | TRUE | FALSE | -3.323530283 |
| **tgfa** | RV end systolic volume \| Instance 2_BSA_indexed_corrected | -1.591326539 | 0.1756660391 | -0.09951719661 | 5.22E-11 | 0.2417998065 | -0.09951719661 | 5.22E-11 | 4332 | 1.65E-09 | TRUE | 7.64E-07 | TRUE | 6.46E-07 | TRUE | FALSE | -1.591326539 |
| **tgfbr2** | LV end diastolic volume \| Instance 2(participant - p24100_i2)_BSA_indexed_corrected | -4.355134043 | 0.1154109876 | -0.1153696491 | 2.47E-14 | 0.5693187798 | -0.1153696491 | 2.47E-14 | 4340 | 1.42E-12 | TRUE | 3.61E-10 | TRUE | 3.05E-10 | TRUE | FALSE | -4.355134043 |
| **tgfbr2** | LV end systolic volume \| Instance 2(participant - p24101_i2)_BSA_indexed_corrected | -1.856977553 | 0.2092125264 | -0.0847597314 | 2.24E-08 | 0.3314413169 | -0.0847597314 | 2.24E-08 | 4340 | 4.35E-07 | TRUE | 0.0003278008413 | TRUE | 0.0002772539516 | TRUE | FALSE | -1.856977553 |
| **tgfbr2** | RV end diastolic volume \| Instance 2_BSA_indexed_corrected | -5.542848955 | -0.05550530536 | -0.1365206164 | 1.66E-19 | 0.6106673143 | -0.1365206164 | 1.66E-19 | 4340 | 2.21E-17 | TRUE | 2.43E-15 | TRUE | 2.06E-15 | TRUE | FALSE | -5.542848955 |
| **tgfbr2** | RV end systolic volume \| Instance 2_BSA_indexed_corrected | -2.585250099 | 0.07077347112 | -0.1070350811 | 1.55E-12 | 0.3646108077 | -0.1070350811 | 1.55E-12 | 4340 | 6.48E-11 | TRUE | 2.27E-08 | TRUE | 1.92E-08 | TRUE | FALSE | -2.585250099 |
| **thop1** | LV end diastolic volume \| Instance 2(participant - p24100_i2)_BSA_indexed_corrected | -3.409466973 | 0.2352016901 | -0.09920902979 | 5.07E-11 | 0.5177198713 | -0.09920902979 | 5.07E-11 | 4365 | 1.62E-09 | TRUE | 7.42E-07 | TRUE | 6.27E-07 | TRUE | FALSE | -3.409466973 |
| **thop1** | RV end diastolic volume \| Instance 2_BSA_indexed_corrected | -3.082503757 | 0.1288359564 | -0.08337468277 | 3.46E-08 | 0.5577788968 | -0.08337468277 | 3.46E-08 | 4365 | 6.48E-07 | TRUE | 0.0005061051797 | TRUE | 0.000428063761 | TRUE | FALSE | -3.082503757 |
| **thop1** | RV end systolic volume \| Instance 2_BSA_indexed_corrected | -1.537731497 | 0.1595491384 | -0.06993300686 | 3.75E-06 | 0.3320787691 | -0.06993300686 | 3.75E-06 | 4365 | 4.82E-05 | TRUE | 0.05488750497 | FALSE | 0.0464238517 | TRUE | FALSE | -1.537731497 |
| **thy1** | LV end diastolic volume \| Instance 2(participant - p24100_i2)_BSA_indexed_corrected | -4.423355015 | 0.1418884842 | -0.108522346 | 6.60E-13 | 0.6135741253 | -0.108522346 | 6.60E-13 | 4363 | 2.94E-11 | TRUE | 9.66E-09 | TRUE | 8.17E-09 | TRUE | FALSE | -4.423355015 |
| **thy1** | LV end systolic volume \| Instance 2(participant - p24101_i2)_BSA_indexed_corrected | -2.176522883 | 0.2069832073 | -0.09206842955 | 1.11E-09 | 0.3564601158 | -0.09206842955 | 1.11E-09 | 4363 | 2.72E-08 | TRUE | 1.63E-05 | TRUE | 1.37E-05 | TRUE | FALSE | -2.176522883 |
| **thy1** | RV end diastolic volume \| Instance 2_BSA_indexed_corrected | -5.782168893 | -0.03935009148 | -0.1317582979 | 2.36E-18 | 0.6587443724 | -0.1317582979 | 2.36E-18 | 4363 | 2.47E-16 | TRUE | 3.45E-14 | TRUE | 2.92E-14 | TRUE | FALSE | -5.782168893 |
| **thy1** | RV end systolic volume \| Instance 2_BSA_indexed_corrected | -2.565441148 | 0.09470067826 | -0.09837709495 | 7.41E-11 | 0.3929734603 | -0.09837709495 | 7.41E-11 | 4363 | 2.26E-09 | TRUE | 1.08E-06 | TRUE | 9.17E-07 | TRUE | FALSE | -2.565441148 |
| **timp1** | LV end diastolic volume \| Instance 2(participant - p24100_i2)_BSA_indexed_corrected | -3.025586738 | 0.2053866144 | -0.07212199149 | 2.39E-06 | 0.6405427487 | -0.07212199149 | 2.39E-06 | 4269 | 3.17E-05 | TRUE | 0.03500964639 | TRUE | 0.02961115892 | TRUE | FALSE | -3.025586738 |
| **timp1** | RV end diastolic volume \| Instance 2_BSA_indexed_corrected | -3.782620529 | 0.06321505979 | -0.08390954979 | 4.01E-08 | 0.6876789999 | -0.08390954979 | 4.01E-08 | 4269 | 7.41E-07 | TRUE | 0.0005862792131 | TRUE | 0.0004958749584 | TRUE | FALSE | -3.782620529 |

| | | | | | | | | | | | | | | | | | |
|---|---|---|---|---|---|---|---|---|---|---|---|---|---|---|---|---|---|
| **tmprss15** | LV end diastolic volume \| Instance 2(participant - p24100_i2)_BSA_indexed_corrected | -1.087252162 | 0.2968602252 | -0.08928633877 | 3.30E-09 | 0.1834076645 | -0.08928633877 | 3.30E-09 | 4375 | 7.34E-08 | TRUE | 4.83E-05 | TRUE | 4.09E-05 | TRUE | FALSE | -1.087252162 |
| **tmsb10** | LV end diastolic volume \| Instance 2(participant - p24100_i2)_BSA_indexed_corrected | -1.176398835 | 0.1901734179 | -0.08063284225 | 1.15E-07 | 0.2215327528 | -0.08063284225 | 1.15E-07 | 4311 | 1.96E-06 | TRUE | 0.001682117633 | TRUE | 0.001422735094 | TRUE | FALSE | -1.176398835 |
| **tmsb10** | RV end diastolic volume \| Instance 2_BSA_indexed_corrected | -1.391773307 | 0.08346163382 | -0.08861598264 | 5.60E-09 | 0.2383176761 | -0.08861598264 | 5.60E-09 | 4311 | 1.19E-07 | TRUE | 8.20E-05 | TRUE | 6.94E-05 | TRUE | FALSE | -1.391773307 |
| **tnfrsf10a** | LV end diastolic volume \| Instance 2(participant - p24100_i2)_BSA_indexed_corrected | -3.029825081 | 0.09008418533 | -0.08804066067 | 5.44E-09 | 0.5183878635 | -0.08804066067 | 5.44E-09 | 4375 | 1.16E-07 | TRUE | 7.96E-05 | TRUE | 6.74E-05 | TRUE | FALSE | -3.029825081 |
| **tnfrsf10a** | RV end diastolic volume \| Instance 2_BSA_indexed_corrected | -4.350885792 | -0.1341312425 | -0.1175156654 | 6.30E-15 | 0.5559970574 | -0.1175156654 | 6.30E-15 | 4375 | 4.01E-13 | TRUE | 9.22E-11 | TRUE | 7.80E-11 | TRUE | FALSE | -4.350885792 |
| **tnfrsf10a** | RV end systolic volume \| Instance 2_BSA_indexed_corrected | -1.888597666 | 0.05322722304 | -0.08577675824 | 1.33E-08 | 0.3317236845 | -0.08577675824 | 1.33E-08 | 4375 | 2.68E-07 | TRUE | 0.0001942517494 | TRUE | 0.0001642981296 | TRUE | FALSE | -1.888597666 |
| **tnfrsf10b** | LV end diastolic volume \| Instance 2(participant - p24100_i2)_BSA_indexed_corrected | -2.923050294 | 0.1056127721 | -0.09263539852 | 8.32E-10 | 0.4751143245 | -0.09263539852 | 8.32E-10 | 4375 | 2.09E-08 | TRUE | 1.22E-05 | TRUE | 1.03E-05 | TRUE | FALSE | -2.923050294 |
| **tnfrsf10b** | LV end systolic volume \| Instance 2(participant - p24101_i2)_BSA_indexed_corrected | -1.467776684 | 0.1876815048 | -0.08013268256 | 1.11E-07 | 0.2760970156 | -0.08013268256 | 1.11E-07 | 4375 | 1.91E-06 | TRUE | 0.001627746051 | TRUE | 0.00137674761 | TRUE | FALSE | -1.467776684 |
| **tnfrsf10b** | RV end diastolic volume \| Instance 2_BSA_indexed_corrected | -3.753631134 | -0.07425887811 | -0.110571862 | 2.23E-13 | 0.5102065106 | -0.110571862 | 2.23E-13 | 4375 | 1.07E-11 | TRUE | 3.27E-09 | TRUE | 2.76E-09 | TRUE | FALSE | -3.753631134 |
| **tnfrsf10b** | RV end systolic volume \| Instance 2_BSA_indexed_corrected | -1.871245438 | 0.05874217054 | -0.09269064085 | 8.13E-10 | 0.3039705002 | -0.09269064085 | 8.13E-10 | 4375 | 2.06E-08 | TRUE | 1.19E-05 | TRUE | 1.01E-05 | TRUE | FALSE | -1.871245438 |
| **tnfrsf11a** | LV end diastolic volume \| Instance 2(participant - p24100_i2)_BSA_indexed_corrected | -3.572317876 | 0.06020705502 | -0.1149693994 | 2.63E-14 | 0.4675036622 | -0.1149693994 | 2.63E-14 | 4361 | 1.51E-12 | TRUE | 3.85E-10 | TRUE | 3.25E-10 | TRUE | FALSE | -3.572317876 |
| **tnfrsf11a** | LV end systolic volume \| Instance 2(participant - p24101_i2)_BSA_indexed_corrected | -1.699768692 | 0.1707769392 | -0.09417135102 | 4.64E-10 | 0.2721719006 | -0.09417135102 | 4.64E-10 | 4361 | 1.24E-08 | TRUE | 6.78E-06 | TRUE | 5.74E-06 | TRUE | FALSE | -1.699768692 |
| **tnfrsf11a** | RV end diastolic volume \| Instance 2_BSA_indexed_corrected | -4.239464003 | -0.1043430224 | -0.12679555 | 4.28E-17 | 0.5023356925 | -0.12679555 | 4.28E-17 | 4361 | 3.85E-15 | TRUE | 6.27E-13 | TRUE | 5.30E-13 | TRUE | FALSE | -4.239464003 |
| **tnfrsf11a** | RV end systolic volume \| Instance 2_BSA_indexed_corrected | -2.01256057 | 0.04959395304 | -0.1011987448 | 2.11E-11 | 0.2996712561 | -0.1011987448 | 2.11E-11 | 4361 | 7.24E-10 | TRUE | 3.09E-07 | TRUE | 2.61E-07 | TRUE | FALSE | -2.01256057 |
| **tnfrsf12a** | RV end diastolic volume \| Instance 2_BSA_indexed_corrected | -2.402471227 | 0.04868110577 | -0.07255750942 | 1.71E-06 | 0.5014005926 | -0.07255750942 | 1.71E-06 | 4340 | 2.34E-05 | TRUE | 0.02502500642 | TRUE | 0.02116615043 | TRUE | FALSE | -2.402471227 |
| **tnfrsf13b** | LV end diastolic volume \| Instance 2(participant - p24100_i2)_BSA_indexed_corrected | -2.560452376 | 0.2105988784 | -0.07279624413 | 1.72E-06 | 0.5345231352 | -0.07279624413 | 1.72E-06 | 4309 | 2.35E-05 | TRUE | 0.02520007778 | TRUE | 0.02131422579 | TRUE | FALSE | -2.560452376 |
| **tnfrsf13b** | RV end diastolic volume \| Instance 2_BSA_indexed_corrected | -3.272033363 | 0.06867423194 | -0.08655057026 | 1.27E-08 | 0.5738885768 | -0.08655057026 | 1.27E-08 | 4309 | 2.56E-07 | TRUE | 0.0001853489446 | TRUE | 0.0001567681373 | TRUE | FALSE | -3.272033363 |
| **tnfrsf13b** | RV end systolic volume \| Instance 2_BSA_indexed_corrected | -1.672317048 | 0.1151675615 | -0.07442321143 | 1.00E-06 | 0.3414418049 | -0.07442321143 | 1.00E-06 | 4309 | 1.43E-05 | TRUE | 0.01469792794 | TRUE | 0.01243150745 | TRUE | FALSE | -1.672317048 |
| **tnfrsf14** | RV end diastolic volume \| Instance 2_BSA_indexed_corrected | -3.756930189 | 0.07471104388 | -0.0867498524 | 1.17E-08 | 0.6574104681 | -0.0867498524 | 1.17E-08 | 4309 | 2.39E-07 | TRUE | 0.0001715936895 | TRUE | 0.0001451339426 | TRUE | FALSE | -3.756930189 |
| **tnfrsf19** | LV cardiac output \| Instance 2_BSA_indexed_corrected | -0.1305842468 | -0.02544172706 | -0.07574687553 | 5.28E-07 | 0.02599482132 | -0.07574687553 | 5.28E-07 | 4375 | 7.98E-06 | TRUE | 0.007720137382 | TRUE | 0.006529692198 | TRUE | FALSE | -0.1305842468 |
| **tnfrsf19** | RV end diastolic volume \| Instance 2_BSA_indexed_corrected | -3.094886044 | 0.1084742627 | -0.07447473196 | 8.16E-07 | 0.62666901 | -0.07447473196 | 8.16E-07 | 4375 | 1.18E-05 | TRUE | 0.01193925166 | TRUE | 0.01009821905 | TRUE | FALSE | -3.094886044 |
| **tnfrsf1a** | LV end diastolic volume \| Instance 2(participant - p24100_i2)_BSA_indexed_corrected | -4.955643157 | 0.04713179249 | -0.1197870928 | 2.99E-15 | 0.6256959583 | -0.1197870928 | 2.99E-15 | 4311 | 2.02E-13 | TRUE | 4.38E-11 | TRUE | 3.70E-11 | TRUE | FALSE | -4.955643157 |
| **tnfrsf1a** | LV end systolic volume \| Instance 2(participant - p24101_i2)_BSA_indexed_corrected | -2.059345571 | 0.1898623698 | -0.08568888094 | 1.75E-08 | 0.3647675594 | -0.08568888094 | 1.75E-08 | 4311 | 3.47E-07 | TRUE | 0.0002562400137 | TRUE | 0.0002167278036 | TRUE | FALSE | -2.059345571 |
| **tnfrsf1a** | RV end diastolic volume \| Instance 2_BSA_indexed_corrected | -6.687808659 | -0.1353664026 | -0.1501689035 | 3.65E-23 | 0.6707531219 | -0.1501689035 | 3.65E-23 | 4311 | 8.08E-21 | TRUE | 5.34E-19 | TRUE | 4.51E-19 | TRUE | FALSE | -6.687808659 |
| **tnfrsf1a** | RV end systolic volume \| Instance 2_BSA_indexed_corrected | -3.219952409 | 0.03422660743 | -0.1215915515 | 1.14E-15 | 0.4004270636 | -0.1215915515 | 1.14E-15 | 4311 | 8.41E-14 | TRUE | 1.67E-11 | TRUE | 1.41E-11 | TRUE | FALSE | -3.219952409 |
| **tnfrsf1b** | RV end diastolic volume \| Instance 2_BSA_indexed_corrected | -3.922175089 | 0.02740526357 | -0.1025226982 | 1.51E-11 | 0.5797279094 | -0.1025226982 | 1.51E-11 | 4311 | 5.27E-10 | TRUE | 2.20E-07 | TRUE | 1.86E-07 | TRUE | FALSE | -3.922175089 |
| **tnfrsf1b** | RV end systolic volume \| Instance 2_BSA_indexed_corrected | -1.901709557 | 0.1117830476 | -0.08359783234 | 3.86E-08 | 0.3453329695 | -0.08359783234 | 3.86E-08 | 4311 | 7.18E-07 | TRUE | 0.0005654368014 | TRUE | 0.0004782464466 | TRUE | FALSE | -1.901709557 |
| **tnfrsf4** | LV end diastolic volume \| Instance 2(participant - p24100_i2)_BSA_indexed_corrected | -2.349470234 | 0.2614353245 | -0.07319776608 | 1.33E-06 | 0.4852450264 | -0.07319776608 | 1.33E-06 | 4354 | 1.85E-05 | TRUE | 0.01947497037 | TRUE | 0.01647192994 | TRUE | FALSE | -2.349470234 |
| **tnfrsf4** | RV end diastolic volume \| Instance 2_BSA_indexed_corrected | -4.111111252 | 0.06205012597 | -0.1190587357 | 3.23E-15 | 0.5197008623 | -0.1190587357 | 3.23E-15 | 4354 | 2.15E-13 | TRUE | 4.72E-11 | TRUE | 3.99E-11 | TRUE | FALSE | -4.111111252 |
| **tnfrsf4** | RV end systolic volume \| Instance 2_BSA_indexed_corrected | -2.13821118 | 0.120362502 | -0.1040752214 | 5.81E-12 | 0.3097375899 | -0.1040752214 | 5.81E-12 | 4354 | 2.22E-10 | TRUE | 8.51E-08 | TRUE | 7.20E-08 | TRUE | FALSE | -2.13821118 |
| **tnfrsf6b** | LV end diastolic volume \| Instance 2(participant - p24100_i2)_BSA_indexed_corrected | -1.626489024 | 0.1766482684 | -0.07826689207 | 2.18E-07 | 0.3132918076 | -0.07826689207 | 2.18E-07 | 4375 | 3.54E-06 | TRUE | 0.003188545422 | TRUE | 0.002696871718 | TRUE | FALSE | -1.626489024 |
| **tnfrsf6b** | RV end diastolic volume \| Instance 2_BSA_indexed_corrected | -2.32409199 | -0.008568268389 | -0.1039519426 | 5.48E-12 | 0.336257284 | -0.1039519426 | 5.48E-12 | 4375 | 2.10E-10 | TRUE | 8.02E-08 | TRUE | 6.78E-08 | TRUE | FALSE | -2.32409199 |
| **tnfrsf6b** | RV end systolic volume \| Instance 2_BSA_indexed_corrected | -1.109982062 | 0.09676153209 | -0.08348478943 | 3.20E-08 | 0.200354967 | -0.08348478943 | 3.20E-08 | 4375 | 6.05E-07 | TRUE | 0.0004682908131 | TRUE | 0.0003960803697 | TRUE | FALSE | -1.109982062 |
| **tnfsf10** | LV end diastolic volume \| Instance 2(participant - p24100_i2)_BSA_indexed_corrected | -3.356341748 | 0.2952709397 | -0.07764240441 | 2.84E-07 | 0.6527708685 | -0.07764240441 | 2.84E-07 | 4361 | 4.51E-06 | TRUE | 0.004158171913 | TRUE | 0.003516981804 | TRUE | FALSE | -3.356341748 |
| **tnfsf14** | LV end diastolic volume \| Instance 2(participant - p24100_i2)_BSA_indexed_corrected | -1.52234849 | 0.2520382884 | -0.07721208281 | 3.17E-07 | 0.2972628464 | -0.07721208281 | 3.17E-07 | 4375 | 4.98E-06 | TRUE | 0.004632180762 | TRUE | 0.003917898489 | TRUE | FALSE | -1.52234849 |
| **tnfsf14** | RV end diastolic volume \| Instance 2_BSA_indexed_corrected | -1.696224366 | 0.1309325522 | -0.07996626808 | 1.18E-07 | 0.3197375928 | -0.07996626808 | 1.18E-07 | 4375 | 2.01E-06 | TRUE | 0.001729406097 | TRUE | 0.001462731676 | TRUE | FALSE | -1.696224366 |
| **tpp1** | RV end diastolic volume \| Instance 2_BSA_indexed_corrected | -1.910279963 | 0.1603568359 | -0.07211668015 | 2.36E-06 | 0.4041685891 | -0.07211668015 | 2.36E-06 | 4275 | 3.13E-05 | TRUE | 0.03450282793 | TRUE | 0.02918249186 | TRUE | FALSE | -1.910279963 |
| **trem2** | LV end diastolic volume \| Instance 2(participant - p24100_i2)_BSA_indexed_corrected | -1.932778595 | 0.1026994272 | -0.09346106484 | 8.24E-10 | 0.3140603203 | -0.09346106484 | 8.24E-10 | 4300 | 2.08E-08 | TRUE | 1.20E-05 | TRUE | 1.02E-05 | TRUE | FALSE | -1.932778595 |
| **trem2** | RV end diastolic volume \| Instance 2_BSA_indexed_corrected | -2.531652557 | -0.08865902878 | -0.1139159583 | 6.77E-14 | 0.336782875 | -0.1139159583 | 6.77E-14 | 4300 | 3.60E-12 | TRUE | 9.91E-10 | TRUE | 8.38E-10 | TRUE | FALSE | -2.531652557 |
| **trem2** | RV end systolic volume \| Instance 2_BSA_indexed_corrected | -0.995724412 | 0.06711317275 | -0.07542096532 | 7.37E-07 | 0.2008052821 | -0.07542096532 | 7.37E-07 | 4300 | 1.08E-05 | TRUE | 0.01078522266 | TRUE | 0.009122141329 | TRUE | FALSE | -0.995724412 |
| **vasn** | RV end diastolic volume \| Instance 2_BSA_indexed_corrected | -3.275771037 | 0.1518890028 | -0.07085483829 | 3.73E-06 | 0.7072199222 | -0.07085483829 | 3.73E-06 | 4254 | 4.80E-05 | TRUE | 0.05459263874 | FALSE | 0.04617445385 | TRUE | FALSE | -3.275771037 |
| **vmo1** | RV end diastolic volume \| Instance 2_BSA_indexed_corrected | -1.269071463 | 0.04008962028 | -0.07155878095 | 2.16E-06 | 0.2674968134 | -0.07155878095 | 2.16E-06 | 4375 | 2.90E-05 | TRUE | 0.03159414549 | TRUE | 0.02672232826 | TRUE | FALSE | -1.269071463 |
| **vsig4** | LV end diastolic volume \| Instance 2(participant - p24100_i2)_BSA_indexed_corrected | -2.68764083 | 0.08346039535 | -0.09379013255 | 7.12E-10 | 0.4350721127 | -0.09379013255 | 7.12E-10 | 4302 | 1.83E-08 | TRUE | 1.04E-05 | TRUE | 8.81E-06 | TRUE | FALSE | -2.68764083 |
| **vsig4** | RV end diastolic volume \| Instance 2_BSA_indexed_corrected | -3.511875053 | -0.0728874614 | -0.1138215905 | 7.01E-14 | 0.4674644368 | -0.1138215905 | 7.01E-14 | 4302 | 3.70E-12 | TRUE | 1.03E-09 | TRUE | 8.67E-10 | TRUE | FALSE | -3.511875053 |
| **vsig4** | RV end systolic volume \| Instance 2_BSA_indexed_corrected | -1.613029911 | 0.07496158031 | -0.08794972631 | 7.55E-09 | 0.2786041 | -0.08794972631 | 7.55E-09 | 4302 | 1.57E-07 | TRUE | 0.0001105194303 | TRUE | 9.35E-05 | TRUE | FALSE | -1.613029911 |
| **vwa1** | LV end diastolic volume \| Instance 2(participant - p24100_i2)_BSA_indexed_corrected | -2.104698544 | 0.2302006444 | -0.08115743949 | 7.64E-08 | 0.3908740579 | -0.08115743949 | 7.64E-08 | 4375 | 1.35E-06 | TRUE | 0.001117953241 | TRUE | 0.000945564851 | TRUE | FALSE | -2.104698544 |
| **vwa1** | RV end diastolic volume \| Instance 2_BSA_indexed_corrected | -2.381776621 | 0.1044598282 | -0.08536735156 | 1.56E-08 | 0.4203695094 | -0.08536735156 | 1.56E-08 | 4375 | 3.11E-07 | TRUE | 0.0002276960108 | TRUE | 0.0001925852859 | TRUE | FALSE | -2.381776621 |
| **vwc2** | LV end diastolic volume \| Instance 2(participant - p24100_i2)_BSA_indexed_corrected | -2.582883056 | 0.1480732627 | -0.08830092562 | 5.03E-09 | 0.4409081711 | -0.08830092562 | 5.03E-09 | 4369 | 1.08E-07 | TRUE | 7.35E-05 | TRUE | 6.22E-05 | TRUE | FALSE | -2.582883056 |
| **vwc2** | RV end diastolic volume \| Instance 2_BSA_indexed_corrected | -3.728040691 | -0.05718318773 | -0.1184612974 | 3.98E-15 | 0.4728721487 | -0.1184612974 | 3.98E-15 | 4369 | 2.60E-13 | TRUE | 5.82E-11 | TRUE | 4.93E-11 | TRUE | FALSE | -3.728040691 |
| **vwc2** | RV end systolic volume \| Instance 2_BSA_indexed_corrected | -1.6535033 | 0.08262123718 | -0.08837485799 | 4.88E-09 | 0.2820214483 | -0.08837485799 | 4.88E-09 | 4369 | 1.05E-07 | TRUE | 7.14E-05 | TRUE | 6.04E-05 | TRUE | FALSE | -1.6535033 |
| **wfdc2** | LV end diastolic volume \| Instance 2(participant - p24100_i2)_BSA_indexed_corrected | -2.422860857 | 0.1900012825 | -0.07455560809 | 1.99E-06 | 0.5089115871 | -0.07455560809 | 1.99E-06 | 4057 | 2.69E-05 | TRUE | 0.02918510898 | TRUE | 0.02468476518 | TRUE | FALSE | -2.422860857 |
| **wfdc2** | RV end diastolic volume \| Instance 2_BSA_indexed_corrected | -3.317555286 | 0.06422340389 | -0.09482496227 | 1.43E-09 | 0.5469387465 | -0.09482496227 | 1.43E-09 | 4057 | 3.41E-08 | TRUE | 2.10E-05 | TRUE | 1.77E-05 | TRUE | FALSE | -3.317555286 |
| **xcl1** | RV end diastolic volume \| Instance 2_BSA_indexed_corrected | -1.733470137 | 0.2177035935 | -0.08388547634 | 2.75E-08 | 0.3113912703 | -0.08388547634 | 2.75E-08 | 4375 | 5.24E-07 | TRUE | 0.0004021821904 | TRUE | 0.0003401656967 | TRUE | FALSE | -1.733470137 |
| **xg** | RV end diastolic volume \| Instance 2_BSA_indexed_corrected | -2.478705189 | 0.0517523299 | -0.0809637522 | 8.69E-08 | 0.4623407779 | -0.0809637522 | 8.69E-08 | 4358 | 1.52E-06 | TRUE | 0.001272000573 | TRUE | 0.001075858084 | TRUE | FALSE | -2.478705189 |

**Supplemental Table 5. Proteome-wide association analysis of circulating proteins with conventional cardiac parameters.** Significant associations between 1,463 Olink Explore protein levels and 10 conventional cardiac parameters in up to 4,511 UK Biobank participants, after cluster-adjusted Bonferroni correction ($p < 4.04 \times 10^{-6}$; M_eff = 8.5). One thousand one hundred forty-one significant associations across 396 unique proteins are shown, sorted by p-value.

| protein | of_n_associations | trad_n_associations | of_strongest_param | of_strongest_r | of_strongest_p | trad_strongest_param | trad_strongest_r | trad_strongest_p |
|---|---|---|---|---|---|---|---|---|
| ntprobnp | 6 | 3 | late_diastolic_peaks_lv_inner_velocity_corrected | -0.1166870317 | 7.29E-26 | LV end diastolic volume \| Instance 2(participant - p24100_i2)_BSA_indexed_corrected | 0.1318977649 | 1.95E-18 |
| ssc4d | 3 | 4 | late_diastolic_peaks_lv_inner_velocity_corrected | 0.1018882347 | 4.69E-20 | LV end diastolic volume \| Instance 2(participant - p24100_i2)_BSA_indexed_corrected | -0.1658134444 | 2.46E-28 |
| acy1 | 3 | 4 | late_diastolic_peaks_lv_inner_velocity_corrected | 0.09932090672 | 3.80E-19 | LV end diastolic volume \| Instance 2(participant - p24100_i2)_BSA_indexed_corrected | -0.1411937806 | 6.38E-21 |
| hgf | 6 | 4 | late_diastolic_peaks_lv_inner_circumferential_velocity_corrected | 0.09774040021 | 1.60E-18 | RV end diastolic volume \| Instance 2_BSA_indexed_corrected | -0.1896510615 | 1.35E-36 |
| igfbp1 | 4 | 4 | late_diastolic_peaks_lv_inner_velocity_corrected | -0.09551250337 | 8.66E-18 | LV end diastolic volume \| Instance 2(participant - p24100_i2)_BSA_indexed_corrected | 0.1533829127 | 2.07E-24 |
| rarres2 | 5 | 4 | late_diastolic_peaks_lv_inner_radial_velocity_corrected | 0.09458917905 | 2.35E-17 | RV end diastolic volume \| Instance 2_BSA_indexed_corrected | -0.1761006927 | 1.75E-31 |
| rbp5 | 3 | 4 | late_diastolic_peaks_lv_inner_velocity_corrected | 0.09380347335 | 3.08E-17 | RV end diastolic volume \| Instance 2_BSA_indexed_corrected | -0.1545211795 | 8.72E-25 |
| gsta1 | 3 | 4 | late_diastolic_peaks_lv_inner_velocity_corrected | 0.09346923618 | 3.99E-17 | LV end diastolic volume \| Instance 2(participant - p24100_i2)_BSA_indexed_corrected | -0.1508126628 | 1.13E-23 |
| ldlr | 4 | 4 | late_diastolic_peaks_lv_inner_radial_velocity_corrected | 0.09313734164 | 6.59E-17 | LV end diastolic volume \| Instance 2(participant - p24100_i2)_BSA_indexed_corrected | -0.1901795196 | 1.33E-36 |
| adh4 | 3 | 4 | late_diastolic_peaks_lv_inner_velocity_corrected | 0.08898784177 | 1.16E-15 | RV end diastolic volume \| Instance 2_BSA_indexed_corrected | -0.1369032459 | 9.41E-20 |
| col4a1 | 3 | 4 | late_diastolic_peaks_lv_inner_radial_velocity_corrected | -0.08860407235 | 1.54E-15 | LV end diastolic volume \| Instance 2(participant - p24100_i2)_BSA_indexed_corrected | 0.1402863793 | 1.14E-20 |
| ggt1 | 3 | 4 | late_diastolic_peaks_lv_inner_circumferential_velocity_corrected | 0.08885227136 | 2.04E-15 | LV end diastolic volume \| Instance 2(participant - p24100_i2)_BSA_indexed_corrected | -0.1032016499 | 1.12E-11 |
| plat | 3 | 4 | late_diastolic_peaks_lv_inner_velocity_corrected | 0.08752576549 | 3.38E-15 | LV end diastolic volume \| Instance 2(participant - p24100_i2)_BSA_indexed_corrected | -0.179615152 | 4.83E-33 |
| pamr1 | 4 | 4 | late_diastolic_peaks_lv_inner_velocity_corrected | 0.08765151353 | 4.67E-15 | RV end diastolic volume \| Instance 2_BSA_indexed_corrected | -0.134423192 | 7.74E-19 |
| cpm | 3 | 4 | late_diastolic_peaks_lv_inner_velocity_corrected | 0.08679220395 | 7.07E-15 | RV end diastolic volume \| Instance 2_BSA_indexed_corrected | -0.1715057256 | 4.75E-30 |
| f9 | 3 | 4 | late_diastolic_peaks_lv_inner_velocity_corrected | 0.08686338925 | 7.84E-15 | RV end diastolic volume \| Instance 2_BSA_indexed_corrected | -0.1592621432 | 5.99E-26 |
| erbb2 | 5 | 4 | late_diastolic_peaks_lv_inner_circumferential_velocity_corrected | 0.08547548856 | 1.90E-14 | LV end diastolic volume \| Instance 2(participant - p24100_i2)_BSA_indexed_corrected | -0.1423898124 | 4.27E-21 |
| furin | 4 | 4 | late_diastolic_peaks_lv_inner_velocity_corrected | 0.08460284208 | 3.48E-14 | RV end diastolic volume \| Instance 2_BSA_indexed_corrected | -0.1756769777 | 2.02E-31 |
| angptl2 | 3 | 2 | late_diastolic_peaks_lv_inner_velocity_corrected | 0.08355763229 | 8.45E-14 | LV end diastolic volume \| Instance 2(participant - p24100_i2)_BSA_indexed_corrected | -0.08968597548 | 3.66E-09 |
| prcp | 3 | 4 | late_diastolic_peaks_lv_inner_velocity_corrected | 0.08299184443 | 8.61E-14 | RV end diastolic volume \| Instance 2_BSA_indexed_corrected | -0.153204736 | 2.37E-24 |
| f7 | 3 | 4 | late_diastolic_peaks_lv_inner_velocity_corrected | 0.08303328935 | 1.09E-13 | RV end diastolic volume \| Instance 2_BSA_indexed_corrected | -0.1710674085 | 9.54E-30 |
| sele | 3 | 4 | late_diastolic_peaks_lv_inner_velocity_corrected | 0.08249817585 | 1.15E-13 | LV end diastolic volume \| Instance 2(participant - p24100_i2)_BSA_indexed_corrected | -0.1266836809 | 4.07E-17 |
| igfbp2 | 3 | 4 | late_diastolic_peaks_lv_inner_radial_velocity_corrected | -0.08255662944 | 1.18E-13 | LV end diastolic volume \| Instance 2(participant - p24100_i2)_BSA_indexed_corrected | 0.1853416114 | 4.69E-35 |
| asgr1 | 3 | 4 | late_diastolic_peaks_lv_inner_velocity_corrected | 0.0822277261 | 1.39E-13 | RV end diastolic volume \| Instance 2_BSA_indexed_corrected | -0.1750845677 | 1.86E-31 |
| lilra5 | 5 | 3 | late_diastolic_peaks_lv_inner_velocity_corrected | 0.08124631566 | 2.75E-13 | RV end diastolic volume \| Instance 2_BSA_indexed_corrected | -0.09593943665 | 2.04E-10 |
| cdhr2 | 3 | 4 | late_diastolic_peaks_lv_inner_circumferential_velocity_corrected | 0.07947184927 | 8.74E-13 | LV end diastolic volume \| Instance 2(participant - p24100_i2)_BSA_indexed_corrected | -0.1242021299 | 1.65E-16 |
| prss8 | 3 | 4 | late_diastolic_peaks_lv_inner_velocity_corrected | 0.08008864407 | 9.30E-13 | RV end diastolic volume \| Instance 2_BSA_indexed_corrected | -0.1478119161 | 1.99E-22 |
| nppb | 5 | 2 | late_diastolic_peaks_lv_inner_circumferential_velocity_corrected | -0.07895234886 | 1.23E-12 | LV end diastolic volume \| Instance 2(participant - p24100_i2)_BSA_indexed_corrected | 0.09502301709 | 3.02E-10 |
| ces1 | 3 | 4 | late_diastolic_peaks_lv_inner_velocity_corrected | 0.07821141492 | 2.47E-12 | RV end diastolic volume \| Instance 2_BSA_indexed_corrected | -0.1664690572 | 2.36E-28 |
| ccl22 | 8 | 4 | late_diastolic_peaks_lv_inner_velocity_corrected | 0.07887427107 | 2.51E-12 | RV end diastolic volume \| Instance 2_BSA_indexed_corrected | -0.131388651 | 7.88E-18 |
| ca14 | 6 | 4 | late_diastolic_peaks_lv_inner_velocity_corrected | -0.07734275605 | 3.48E-12 | RV end diastolic volume \| Instance 2_BSA_indexed_corrected | 0.1366927736 | 1.07E-19 |
| inhbc | 3 | 4 | late_diastolic_peaks_lv_inner_velocity_corrected | 0.07693585469 | 6.22E-12 | RV end diastolic volume \| Instance 2_BSA_indexed_corrected | -0.1882379085 | 1.12E-35 |
| lep | 6 | 4 | late_diastolic_peaks_lv_inner_radial_velocity_corrected | 0.07636902899 | 6.46E-12 | RV end diastolic volume \| Instance 2_BSA_indexed_corrected | -0.2443739926 | 1.66E-60 |
| gusb | 3 | 4 | late_diastolic_peaks_lv_inner_velocity_corrected | 0.07633752158 | 6.59E-12 | LV end diastolic volume \| Instance 2(participant - p24100_i2)_BSA_indexed_corrected | -0.1567566036 | 1.81E-25 |
| ssc5d | 3 | 4 | late_diastolic_peaks_lv_inner_velocity_corrected | 0.07649160111 | 7.79E-12 | RV end diastolic volume \| Instance 2_BSA_indexed_corrected | -0.1416183769 | 8.29E-21 |
| il1rn | 3 | 4 | late_diastolic_peaks_lv_inner_velocity_corrected | 0.07664283866 | 8.01E-12 | RV end diastolic volume \| Instance 2_BSA_indexed_corrected | -0.2165738297 | 7.07E-47 |
| acp5 | 4 | 4 | late_diastolic_peaks_lv_inner_velocity_corrected | 0.07519867528 | 1.35E-11 | RV end diastolic volume \| Instance 2_BSA_indexed_corrected | -0.1518532038 | 5.53E-24 |
| thop1 | 3 | 3 | late_diastolic_peaks_lv_inner_circumferential_velocity_corrected | 0.07513774588 | 1.52E-11 | LV end diastolic volume \| Instance 2(participant - p24100_i2)_BSA_indexed_corrected | -0.09920902979 | 5.07E-11 |
| dcxr | 3 | 4 | late_diastolic_peaks_lv_inner_velocity_corrected | 0.07511479312 | 1.74E-11 | LV end diastolic volume \| Instance 2(participant - p24100_i2)_BSA_indexed_corrected | -0.1186721423 | 4.38E-15 |
| fgf21 | 3 | 4 | late_diastolic_peaks_lv_inner_velocity_corrected | 0.07450882015 | 2.07E-11 | RV end diastolic volume \| Instance 2_BSA_indexed_corrected | -0.1473655354 | 1.15E-22 |
| scly | 3 | 4 | late_diastolic_peaks_lv_inner_velocity_corrected | 0.07438522571 | 2.28E-11 | LV end diastolic volume \| Instance 2(participant - p24100_i2)_BSA_indexed_corrected | -0.1259356543 | 6.33E-17 |
| ccl16 | 3 | 4 | late_diastolic_peaks_lv_inner_velocity_corrected | 0.07430475727 | 2.34E-11 | LV end diastolic volume \| Instance 2(participant - p24100_i2)_BSA_indexed_corrected | -0.1157163456 | 1.62E-14 |
| gsta3 | 3 | 4 | late_diastolic_peaks_lv_inner_velocity_corrected | 0.07408600973 | 2.68E-11 | LV end diastolic volume \| Instance 2(participant - p24100_i2)_BSA_indexed_corrected | -0.1518399702 | 5.58E-24 |
| il18r1 | 3 | 4 | late_diastolic_peaks_lv_inner_velocity_corrected | 0.07418771993 | 2.75E-11 | LV end diastolic volume \| Instance 2(participant - p24100_i2)_BSA_indexed_corrected | -0.1356337855 | 2.34E-19 |
| kdr | 3 | 4 | late_diastolic_peaks_lv_inner_velocity_corrected | 0.07403812774 | 3.36E-11 | LV end diastolic volume \| Instance 2(participant - p24100_i2)_BSA_indexed_corrected | -0.1095139208 | 4.68E-13 |
| defb4a_defb4b | 3 | 1 | late_diastolic_peaks_lv_inner_velocity_corrected | 0.07353943488 | 4.96E-11 | LV end diastolic volume \| Instance 2(participant - p24100_i2)_BSA_indexed_corrected | -0.07407023257 | 1.11E-06 |
| krt18 | 3 | 4 | late_diastolic_peaks_lv_inner_velocity_corrected | 0.07253302376 | 6.89E-11 | RV end diastolic volume \| Instance 2_BSA_indexed_corrected | -0.1404058322 | 1.05E-20 |

| | | | | | | | | |
|---|---|---|---|---|---|---|---|---|
| **fabp4** | 5 | 4 | late_diastolic_peaks_lv_inner_radial_velocity_corrected | 0.07228100829 | 8.01E-11 | RV end diastolic volume \| Instance 2_BSA_indexed_corrected | -0.2498593612 | 2.94E-63 |
| **kynu** | 3 | 4 | late_diastolic_peaks_lv_inner_velocity_corrected | 0.07240688002 | 8.08E-11 | LV end diastolic volume \| Instance 2(participant - p24100_i2)_BSA_indexed_corrected | -0.1248423643 | 1.29E-16 |
| **dsg2** | 3 | 3 | late_diastolic_peaks_lv_inner_velocity_corrected | -0.07193443405 | 1.31E-10 | LV end diastolic volume \| Instance 2(participant - p24100_i2)_BSA_indexed_corrected | 0.09411197453 | 5.97E-10 |
| **ctsd** | 3 | 4 | late_diastolic_peaks_lv_inner_velocity_corrected | 0.0734126392 | 1.85E-10 | RV end diastolic volume \| Instance 2_BSA_indexed_corrected | -0.1514883176 | 2.34E-22 |
| **cntn3** | 3 | 4 | late_diastolic_peaks_lv_inner_radial_velocity_corrected | 0.07105591348 | 2.18E-10 | LV end diastolic volume \| Instance 2(participant - p24100_i2)_BSA_indexed_corrected | -0.1240896851 | 2.92E-16 |
| **nos3** | 4 | 4 | late_diastolic_peaks_lv_inner_velocity_corrected | 0.07133090802 | 2.25E-10 | RV end diastolic volume \| Instance 2_BSA_indexed_corrected | -0.1039843909 | 9.40E-12 |
| **oxt** | 4 | 4 | late_diastolic_peaks_lv_inner_velocity_corrected | 0.06982267208 | 3.59E-10 | RV end diastolic volume \| Instance 2_BSA_indexed_corrected | -0.1644678253 | 7.84E-28 |
| **ca5a** | 3 | 4 | late_diastolic_peaks_lv_inner_velocity_corrected | 0.06972286505 | 3.62E-10 | LV end diastolic volume \| Instance 2(participant - p24100_i2)_BSA_indexed_corrected | -0.1250655947 | 1.02E-16 |
| **ang** | 3 | 3 | late_diastolic_peaks_lv_inner_velocity_corrected | 0.06984073056 | 4.42E-10 | LV end diastolic volume \| Instance 2(participant - p24100_i2)_BSA_indexed_corrected | -0.1024693875 | 1.53E-11 |
| **baiap2** | 3 | 3 | late_diastolic_peaks_lv_inner_circumferential_velocity_corrected | 0.06919320852 | 4.91E-10 | RV end diastolic volume \| Instance 2_BSA_indexed_corrected | -0.1030643338 | 8.30E-12 |
| **sord** | 3 | 4 | late_diastolic_peaks_lv_inner_circumferential_velocity_corrected | 0.06909170735 | 5.41E-10 | LV end diastolic volume \| Instance 2(participant - p24100_i2)_BSA_indexed_corrected | -0.1234592887 | 2.64E-16 |
| **igsf3** | 3 | 3 | late_diastolic_peaks_lv_inner_velocity_corrected | 0.0686322959 | 6.77E-10 | LV end diastolic volume \| Instance 2(participant - p24100_i2)_BSA_indexed_corrected | -0.1117379322 | 1.25E-13 |
| **nomo1** | 3 | 4 | late_diastolic_peaks_lv_inner_velocity_corrected | 0.06879443397 | 7.98E-10 | RV end diastolic volume \| Instance 2_BSA_indexed_corrected | -0.137877733 | 9.58E-20 |
| **lgals9** | 2 | 4 | late_diastolic_peaks_lv_inner_velocity_corrected | 0.06879015337 | 8.22E-10 | RV end diastolic volume \| Instance 2_BSA_indexed_corrected | -0.1367897373 | 1.84E-19 |
| **pts** | 3 | 4 | late_diastolic_peaks_lv_inner_velocity_corrected | 0.06904724868 | 8.82E-10 | LV end diastolic volume \| Instance 2(participant - p24100_i2)_BSA_indexed_corrected | -0.1215978101 | 1.66E-15 |
| **sema3f** | 2 | 4 | late_diastolic_peaks_lv_inner_circumferential_velocity_corrected | 0.06725910104 | 1.47E-09 | LV end diastolic volume \| Instance 2(participant - p24100_i2)_BSA_indexed_corrected | -0.1427403024 | 2.37E-21 |
| **notch3** | 3 | 4 | late_diastolic_peaks_lv_inner_radial_velocity_corrected | -0.06721999157 | 1.50E-09 | LV end diastolic volume \| Instance 2(participant - p24100_i2)_BSA_indexed_corrected | 0.1121006222 | 1.04E-13 |
| **ifnlr1** | 2 | 3 | late_diastolic_peaks_lv_inner_velocity_corrected | 0.06755569823 | 1.56E-09 | RV end diastolic volume \| Instance 2_BSA_indexed_corrected | -0.09252000127 | 1.06E-09 |
| **pon3** | 3 | 4 | late_diastolic_peaks_lv_inner_velocity_corrected | -0.06725396903 | 1.86E-09 | LV end diastolic volume \| Instance 2(participant - p24100_i2)_BSA_indexed_corrected | 0.152081193 | 8.84E-24 |
| **il6** | 3 | 4 | late_diastolic_peaks_lv_inner_velocity_corrected | 0.06655130028 | 2.18E-09 | RV end diastolic volume \| Instance 2_BSA_indexed_corrected | -0.1156711674 | 1.66E-14 |
| **npy** | 3 | 4 | late_diastolic_peaks_lv_inner_radial_velocity_corrected | 0.06612290794 | 2.76E-09 | LV end diastolic volume \| Instance 2(participant - p24100_i2)_BSA_indexed_corrected | -0.1291761724 | 9.68E-18 |
| **proc** | 3 | 4 | late_diastolic_peaks_lv_inner_velocity_corrected | 0.06481794709 | 6.90E-09 | RV end diastolic volume \| Instance 2_BSA_indexed_corrected | -0.105860934 | 3.01E-12 |
| **hao1** | 3 | 3 | late_diastolic_peaks_lv_inner_velocity_corrected | 0.06424558783 | 7.65E-09 | RV end diastolic volume \| Instance 2_BSA_indexed_corrected | -0.09930788635 | 4.61E-11 |
| **slc39a5** | 3 | 3 | late_diastolic_peaks_lv_inner_velocity_corrected | 0.06455373394 | 7.95E-09 | LV end diastolic volume \| Instance 2(participant - p24100_i2)_BSA_indexed_corrected | -0.107100174 | 1.58E-12 |
| **aldh1a1** | 3 | 4 | late_diastolic_peaks_lv_inner_circumferential_velocity_corrected | 0.06450340952 | 8.36E-09 | LV end diastolic volume \| Instance 2(participant - p24100_i2)_BSA_indexed_corrected | -0.1033575516 | 1.03E-11 |
| **pltp** | 3 | 4 | late_diastolic_peaks_lv_inner_velocity_corrected | -0.06395985256 | 9.25E-09 | LV end diastolic volume \| Instance 2(participant - p24100_i2)_BSA_indexed_corrected | 0.1340516305 | 5.73E-19 |
| **lamp2** | 2 | 4 | late_diastolic_peaks_lv_inner_radial_velocity_corrected | 0.06474785816 | 1.24E-08 | RV end diastolic volume \| Instance 2_BSA_indexed_corrected | -0.1174669786 | 2.34E-14 |
| **crhbp** | 2 | 4 | late_diastolic_peaks_lv_inner_velocity_corrected | 0.06364741223 | 1.41E-08 | RV end diastolic volume \| Instance 2_BSA_indexed_corrected | -0.1202128692 | 2.59E-15 |
| **cdhr5** | 2 | 4 | late_diastolic_peaks_lv_inner_radial_velocity_corrected | 0.0623708475 | 2.06E-08 | LV end diastolic volume \| Instance 2(participant - p24100_i2)_BSA_indexed_corrected | -0.1435992594 | 1.36E-21 |
| **itga5** | 2 | 4 | late_diastolic_peaks_lv_inner_velocity_corrected | 0.06266535317 | 2.19E-08 | RV end diastolic volume \| Instance 2_BSA_indexed_corrected | -0.1365337514 | 2.17E-19 |
| **cd163** | 3 | 2 | late_diastolic_peaks_lv_inner_velocity_corrected | 0.06234474018 | 2.49E-08 | RV end diastolic volume \| Instance 2_BSA_indexed_corrected | -0.091075095 | 1.99E-09 |
| **sult2a1** | 1 | 2 | late_diastolic_peaks_lv_inner_velocity_corrected | 0.06195725656 | 3.08E-08 | RV end diastolic volume \| Instance 2_BSA_indexed_corrected | -0.08276770517 | 4.89E-08 |
| **mfge8** | 2 | 1 | late_diastolic_peaks_lv_inner_radial_velocity_corrected | 0.06193211513 | 3.18E-08 | RV end diastolic volume \| Instance 2_BSA_indexed_corrected | -0.08104629893 | 9.90E-08 |
| **mvk** | 3 | 3 | late_diastolic_peaks_lv_inner_velocity_corrected | 0.06201538142 | 3.49E-08 | RV end diastolic volume \| Instance 2_BSA_indexed_corrected | -0.09443364738 | 5.91E-10 |
| **c19orf12** | 2 | 4 | late_diastolic_peaks_lv_inner_velocity_corrected | 0.06189958612 | 3.74E-08 | LV end diastolic volume \| Instance 2(participant - p24100_i2)_BSA_indexed_corrected | -0.1111062142 | 3.22E-13 |
| **lpl** | 2 | 4 | late_diastolic_peaks_lv_inner_circumferential_velocity_corrected | -0.06073024046 | 4.78E-08 | LV end diastolic volume \| Instance 2(participant - p24100_i2)_BSA_indexed_corrected | 0.1308234412 | 3.69E-18 |
| **spon1** | 1 | 2 | late_diastolic_peaks_rv_inner_radial_velocity_corrected | 0.0611363677 | 4.93E-08 | LV end diastolic volume \| Instance 2(participant - p24100_i2)_BSA_indexed_corrected | -0.08385589699 | 3.53E-08 |
| **esm1** | 3 | 4 | late_diastolic_peaks_lv_inner_circumferential_velocity_corrected | -0.06108876606 | 5.20E-08 | LV end diastolic volume \| Instance 2(participant - p24100_i2)_BSA_indexed_corrected | 0.1197833533 | 3.25E-15 |
| **ambp** | 2 | 4 | late_diastolic_peaks_lv_inner_velocity_corrected | 0.06048547056 | 5.49E-08 | RV end diastolic volume \| Instance 2_BSA_indexed_corrected | -0.1326436099 | 1.27E-18 |
| **plin1** | 2 | 4 | late_diastolic_peaks_lv_inner_radial_velocity_corrected | 0.06102900151 | 5.79E-08 | RV end diastolic volume \| Instance 2_BSA_indexed_corrected | -0.1145011225 | 5.96E-14 |
| **agrn** | 2 | 4 | late_diastolic_peaks_lv_inner_velocity_corrected | 0.06046122725 | 6.62E-08 | RV end diastolic volume \| Instance 2_BSA_indexed_corrected | -0.1710873011 | 9.54E-30 |
| **crip2** | 2 | 4 | late_diastolic_peaks_lv_inner_velocity_corrected | 0.05952554117 | 8.75E-08 | LV end diastolic volume \| Instance 2(participant - p24100_i2)_BSA_indexed_corrected | -0.1021264444 | 1.28E-11 |
| **smoc1** | 2 | 4 | late_diastolic_peaks_lv_inner_velocity_corrected | 0.05929627352 | 9.80E-08 | LV end diastolic volume \| Instance 2(participant - p24100_i2)_BSA_indexed_corrected | -0.1169157619 | 8.65E-15 |
| **tpp1** | 2 | 1 | late_diastolic_peaks_lv_inner_velocity_corrected | 0.05990690798 | 1.04E-07 | RV end diastolic volume \| Instance 2_BSA_indexed_corrected | -0.07211668015 | 2.36E-06 |
| **hnmt** | 1 | 4 | late_diastolic_peaks_lv_inner_velocity_corrected | 0.05917255482 | 1.08E-07 | RV end diastolic volume \| Instance 2_BSA_indexed_corrected | -0.1432537691 | 1.89E-21 |
| **ccl18** | 3 | 2 | late_diastolic_peaks_lv_inner_velocity_corrected | 0.05879749163 | 1.83E-07 | RV end diastolic volume \| Instance 2_BSA_indexed_corrected | -0.08533779762 | 2.51E-08 |
| **fcgr2b** | 3 | 2 | late_diastolic_peaks_lv_inner_velocity_corrected | 0.05799300695 | 1.86E-07 | RV end systolic volume \| Instance 2_BSA_indexed_corrected | -0.07276976371 | 1.45E-06 |
| **cdh2** | 2 | 2 | late_diastolic_peaks_lv_inner_velocity_corrected | 0.05774771915 | 2.09E-07 | RV end diastolic volume \| Instance 2_BSA_indexed_corrected | -0.0813837419 | 7.03E-08 |

| | | | | | | | | |
|---|---|---|---|---|---|---|---|---|
| **rbks** | 2 | 3 | late_diastolic_peaks_lv_inner_circumferential_velocity_corrected | 0.05768834402 | 2.64E-07 | LV end diastolic volume \| Instance 2(participant - p24100_i2)_BSA_indexed_corrected | -0.08987506756 | 3.47E-09 |
| **agrp** | 2 | 1 | late_diastolic_peaks_lv_inner_radial_velocity_corrected | 0.05714547544 | 2.95E-07 | LV end diastolic volume \| Instance 2(participant - p24100_i2)_BSA_indexed_corrected | -0.07383007871 | 1.06E-06 |
| **dkk3** | 3 | 1 | late_diastolic_peaks_lv_inner_circumferential_velocity_corrected | -0.05699315401 | 3.01E-07 | LV cardiac output \| Instance 2_BSA_indexed_corrected | -0.07917990339 | 1.57E-07 |
| **cdcp1** | 1 | 2 | late_diastolic_peaks_lv_inner_velocity_corrected | 0.05672354122 | 3.42E-07 | RV end diastolic volume \| Instance 2_BSA_indexed_corrected | -0.1000241515 | 3.34E-11 |
| **tfpi** | 2 | 3 | late_diastolic_peaks_lv_inner_velocity_corrected | 0.05687458051 | 3.81E-07 | LV end diastolic volume \| Instance 2(participant - p24100_i2)_BSA_indexed_corrected | -0.09997776971 | 4.69E-11 |
| **rgmb** | 1 | 1 | diastolic_peaks_lwave_lv_inner_radial_velocity_corrected | -0.05634710129 | 4.23E-07 | LV cardiac output \| Instance 2_BSA_indexed_corrected | -0.08747807372 | 7.14E-09 |
| **stc1** | 1 | 4 | late_diastolic_peaks_lv_inner_velocity_corrected | 0.05602759491 | 4.91E-07 | RV end diastolic volume \| Instance 2_BSA_indexed_corrected | -0.1414733996 | 6.04E-21 |
| **ccl7** | 1 | 4 | late_diastolic_peaks_lv_inner_velocity_corrected | 0.05622973198 | 5.05E-07 | RV end diastolic volume \| Instance 2_BSA_indexed_corrected | -0.1248923372 | 1.58E-16 |
| **p4hb** | 1 | 2 | late_diastolic_peaks_lv_inner_radial_velocity_corrected | 0.05612635546 | 5.47E-07 | RV end diastolic volume \| Instance 2_BSA_indexed_corrected | -0.09200466967 | 1.41E-09 |
| **cxcl10** | 1 | 2 | late_diastolic_peaks_lv_inner_velocity_corrected | 0.05604010037 | 5.57E-07 | LV end diastolic volume \| Instance 2(participant - p24100_i2)_BSA_indexed_corrected | -0.09860980739 | 8.21E-11 |
| **osm** | 1 | 4 | diastolic_peaks_lwave_lv_inner_circumferential_velocity_corrected | 0.05599770958 | 5.85E-07 | RV end diastolic volume \| Instance 2_BSA_indexed_corrected | -0.1360398927 | 2.93E-19 |
| **ltbp2** | 1 | 1 | late_diastolic_peaks_lv_inner_circumferential_velocity_corrected | -0.05552622547 | 6.01E-07 | LV end systolic volume \| Instance 2(participant - p24101_i2)_BSA_indexed_corrected | 0.07134455504 | 2.32E-06 |
| **il18** | 1 | 2 | late_diastolic_peaks_lv_inner_velocity_corrected | 0.05610086123 | 6.02E-07 | LV end diastolic volume \| Instance 2(participant - p24100_i2)_BSA_indexed_corrected | -0.09053983913 | 2.90E-09 |
| **vwa1** | 1 | 2 | late_diastolic_peaks_lv_inner_velocity_corrected | 0.05552102274 | 6.03E-07 | RV end diastolic volume \| Instance 2_BSA_indexed_corrected | -0.08536735156 | 1.56E-08 |
| **cd4** | 1 | 4 | late_diastolic_peaks_lv_inner_circumferential_velocity_corrected | 0.05584107278 | 6.13E-07 | LV end diastolic volume \| Instance 2(participant - p24100_i2)_BSA_indexed_corrected | -0.1222352644 | 7.00E-16 |
| **fgfr2** | 1 | 3 | late_diastolic_peaks_lv_inner_velocity_corrected | 0.05532418622 | 7.38E-07 | RV end diastolic volume \| Instance 2_BSA_indexed_corrected | -0.1013007069 | 2.25E-11 |
| **mmp9** | 2 | 4 | diastolic_peaks_lwave_lv_inner_circumferential_velocity_corrected | 0.05538185788 | 7.80E-07 | RV end diastolic volume \| Instance 2_BSA_indexed_corrected | -0.1135916275 | 7.77E-14 |
| **il10rb** | 1 | 3 | late_diastolic_peaks_lv_inner_velocity_corrected | 0.05528100443 | 7.91E-07 | RV end diastolic volume \| Instance 2_BSA_indexed_corrected | -0.1083830145 | 9.05E-13 |
| **pbld** | 1 | 2 | late_diastolic_peaks_lv_inner_velocity_corrected | 0.05550006551 | 8.10E-07 | RV end diastolic volume \| Instance 2_BSA_indexed_corrected | -0.08488103106 | 2.73E-08 |
| **gfra1** | 1 | 4 | late_diastolic_peaks_lv_inner_velocity_corrected | 0.05463574864 | 9.07E-07 | RV end diastolic volume \| Instance 2_BSA_indexed_corrected | -0.1334985607 | 7.49E-19 |
| **vmo1** | 1 | 1 | late_diastolic_peaks_lv_inner_velocity_corrected | 0.05445675881 | 9.85E-07 | RV end diastolic volume \| Instance 2_BSA_indexed_corrected | -0.07155878095 | 2.16E-06 |
| **rtn4r** | 1 | 5 | late_diastolic_peaks_lv_inner_velocity_corrected | 0.0543909648 | 1.02E-06 | RV end diastolic volume \| Instance 2_BSA_indexed_corrected | -0.1679927232 | 4.66E-29 |

**Supplemental Table 6. Proteins significantly associated with both optical flow velocity phenotypes and conventional cardiac parameters.** Each of the 117 proteins reaching significance in both analyses is shown with the number of significant associations, strongest phenotype, and corresponding Pearson correlation and p-value in each analysis. Optical flow results reflect cluster-adjusted Bonferroni correction across 1,463 proteins × 32 velocity phenotypes ($p < 1.19 \times 10^{-6}$). Conventional results reflect cluster-adjusted Bonferroni correction across 1,463 proteins × 10 conventional cardiac parameters ($p < 4.04 \times 10^{-6}$). Sorted by optical flow strongest p-value ascending.

| Phenotype | h² (SE) | p-value | Bonferroni significant |
|---|---|---|---|
| LV Inner | | | |
| Systolic LV Inner Circumferential Velocity | 0.0540 (0.0129) | 2.84E-05 | Yes |
| Systolic LV Inner Radial Velocity | 0.0516 (0.0126) | 4.22E-05 | Yes |
| Systolic LV Inner Total Velocity | 0.0519 (0.0128) | 5.02E-05 | Yes |
| Early Diastolic (E-Wave) LV Inner Circumferential Velocity | 0.0726 (0.0134) | 6.04E-08 | Yes |
| Early Diastolic (E-Wave) LV Inner Radial Velocity | 0.0632 (0.0124) | 3.46E-07 | Yes |
| Early Diastolic (E-Wave) LV Inner Total Velocity | 0.0676 (0.0128) | 1.29E-07 | Yes |
| Mid-Diastolic (L-Wave) LV Inner Circumferential Velocity | 0.0961 (0.0132) | 3.36E-13 | Yes |
| Mid-Diastolic (L-Wave) LV Inner Radial Velocity | 0.0882 (0.0126) | 2.58E-12 | Yes |
| Mid-Diastolic (L-Wave) LV Inner Total Velocity | 0.0769 (0.0128) | 1.89E-09 | Yes |
| Late Diastolic (A-Wave) LV Inner Circumferential Velocity | 0.0858 (0.0141) | 1.17E-09 | Yes |
| Late Diastolic (A-Wave) LV Inner Radial Velocity | 0.0889 (0.0135) | 4.57E-11 | Yes |
| Late Diastolic (A-Wave) LV Inner Total Velocity | 0.0961 (0.0143) | 1.82E-11 | Yes |
| LV Myocardium | | | |
| Systolic LV Myocardium Velocity | -0.0094 (0.0109) | 0.388 | No |
| Early Diastolic (E-Wave) LV Myocardium Velocity | 0.0257 (0.0115) | 0.025 | No |
| Mid-Diastolic (L-Wave) LV Myocardium Velocity | 0.0290 (0.0121) | 0.017 | No |
| Late Diastolic (A-Wave) LV Myocardium Velocity | 0.0384 (0.0122) | 0.002 | No |
| RV Inner | | | |
| Systolic RV Inner Circumferential Velocity | 0.0794 (0.0152) | 1.76E-07 | Yes |
| Systolic RV Inner Radial Velocity | 0.0656 (0.0146) | 7.02E-06 | Yes |
| Systolic RV Inner Total Velocity | 0.0732 (0.0150) | 1.06E-06 | Yes |
| Early Diastolic (E-Wave) RV Inner Circumferential Velocity | 0.0502 (0.0129) | 9.97E-05 | Yes |
| Early Diastolic (E-Wave) RV Inner Radial Velocity | 0.0652 (0.0142) | 4.40E-06 | Yes |
| Early Diastolic (E-Wave) RV Inner Total Velocity | 0.0493 (0.0139) | 3.90E-04 | Yes |
| Mid-Diastolic (L-Wave) RV Inner Circumferential Velocity | 0.0998 (0.0128) | 6.41E-15 | Yes |
| Mid-Diastolic (L-Wave) RV Inner Radial Velocity | 0.0873 (0.0140) | 4.52E-10 | Yes |
| Mid-Diastolic (L-Wave) RV Inner Total Velocity | 0.0821 (0.0146) | 1.88E-08 | Yes |
| Late Diastolic (A-Wave) RV Inner Circumferential Velocity | 0.0816 (0.0136) | 1.98E-09 | Yes |
| Late Diastolic (A-Wave) RV Inner Radial Velocity | 0.0503 (0.0141) | 3.61E-04 | Yes |
| Late Diastolic (A-Wave) RV Inner Total Velocity | 0.0658 (0.0150) | 1.15E-05 | Yes |

| | | | |
|---|---|---|---|
| RV Myocardial Free Wall | | | |
| Systolic RV Myocardial Free Wall Velocity | 0.0169 (0.0107) | 0.114 | No |
| Early Diastolic (E-Wave) RV Myocardial Free Wall Velocity | 0.0116 (0.0100) | 0.246 | No |
| Mid-Diastolic (L-Wave) RV Myocardial Free Wall Velocity | 0.0113 (0.0110) | 0.304 | No |
| Late Diastolic (A-Wave) RV Myocardial Free Wall Velocity | 0.0360 (0.0109) | 9.58E-04 | Yes |

**Supplemental Table 7. SNP-based heritability of optical flow velocity phenotypes.** Heritability ($h^2SNP$) was estimated using linkage disequilibrium score regression applied to GWAS summary statistics from up to 73,978 European-ancestry UK Biobank participants. Phenotypes are grouped by cardiac structure. LV = left ventricular; RV = right ventricular; SE = standard error.

| | S LVi Tot | S LVi Circ | S LVi Rad | E LVi Tot | E LVi Circ | E LVi Rad | L LVi Tot | L LVi Circ | L LVi Rad | A LVi Tot | A LVi Circ | A LVi Rad | S LVm | E LVm | L LVm | A LVm | S RVi Tot | S RVi Circ | S RVi Rad | E RVi Tot | E RVi Circ | E RVi Rad | L RVi Tot | L RVi Circ | L RVi Rad | A RVi Tot | A RVi Circ | A RVi Rad | S RVfw | E RVfw | L RVfw | A RVfw |
|---|---|---|---|---|---|---|---|---|---|---|---|---|---|---|---|---|---|---|---|---|---|---|---|---|---|---|---|---|---|---|---|---|
| **S LVi Tot** | 1 | 0.9579 | 0.9759 | 0.3871 | 0.4595 | 0.1851 | 0.4352 | 0.5683 | 0.3007 | 0.4322 | 0.5823 | 0.4008 | | 0.1034 | 0.3913 | 0.841 | 0.4797 | 0.5452 | 0.3479 | 0.7099 | 0.5431 | 0.6588 | 0.559 | 0.5603 | 0.5921 | 0.5687 | 0.4725 | 0.6029 | -0.3761 | -0.1981 | 0.2071 | 0.37 |
| **S LVi Circ** | 0.9579 | 1 | 0.8914 | 0.3965 | 0.4679 | 0.1615 | 0.3137 | 0.4778 | 0.1903 | 0.3232 | 0.4589 | 0.312 | | 0.1071 | 0.3749 | 0.6916 | 0.4296 | 0.4492 | 0.344 | 0.566 | 0.3548 | 0.5565 | 0.4322 | 0.4585 | 0.4949 | 0.5126 | 0.3996 | 0.5761 | -0.0699 | 0.0287 | 0.0327 | 0.2309 |
| **S LVi Rad** | 0.9759 | 0.8914 | 1 | 0.3457 | 0.4125 | 0.166 | 0.541 | 0.6853 | 0.4014 | 0.5013 | 0.6543 | 0.4722 | | -0.0245 | 0.5351 | 0.8113 | 0.5756 | 0.6664 | 0.4404 | 0.7389 | 0.6306 | 0.6695 | 0.6779 | 0.6603 | 0.6886 | 0.6338 | 0.5825 | 0.6377 | -0.4556 | -0.2173 | 0.2149 | 0.4156 |
| **E LVi Tot** | 0.3871 | 0.3965 | 0.3457 | 1 | 0.931 | 0.9526 | 0.0788 | 0.1359 | 0.0378 | -0.1124 | -0.024 | -0.0723 | | 0.6322 | -0.1875 | 0.3077 | 0.0882 | 0.1293 | 0.0662 | 0.5493 | 0.4093 | 0.5279 | 0.1627 | 0.1527 | 0.2243 | 0.0762 | 0.0864 | 0.0679 | 0.3132 | 0.3692 | -0.5812 | -0.1628 |
| **E LVi Circ** | 0.4595 | 0.4679 | 0.4125 | 0.931 | 1 | 0.7976 | 0.0654 | 0.1904 | -0.0546 | -0.0822 | 0.0374 | -0.0782 | | 0.5671 | -0.1181 | 0.1914 | 0.0751 | 0.1105 | 0.0342 | 0.537 | 0.3496 | 0.5335 | 0.1446 | 0.1583 | 0.1811 | 0.1573 | 0.1437 | 0.1664 | 0.3388 | 0.2296 | -0.5526 | -0.0724 |
| **E LVi Rad** | 0.1851 | 0.1615 | 0.166 | 0.9526 | 0.7976 | 1 | 0.0985 | 0.0605 | 0.156 | -0.1684 | -0.1472 | -0.1089 | | 0.5954 | -0.1663 | 0.1455 | 0.0591 | 0.0905 | 0.0073 | 0.5107 | 0.3966 | 0.4482 | 0.1228 | 0.1054 | 0.1359 | -0.0025 | 0.0181 | -0.0504 | 0.1979 | 0.1233 | -0.8882 | -0.2446 |
| **L LVi Tot** | 0.4352 | 0.3137 | 0.541 | 0.0788 | 0.0654 | 0.0985 | 1 | 0.9669 | 1.0011 | 0.7138 | 0.7249 | 0.7327 | | -0.3374 | 1.0671 | 1.0556 | 0.4394 | 0.4449 | 0.3506 | 0.573 | 0.5229 | 0.5279 | 0.9711 | 1.0079 | 0.8811 | 0.6075 | 0.5636 | 0.6566 | -0.3713 | -0.1199 | 0.7332 | 0.2853 |
| **L LVi Circ** | 0.5683 | 0.4778 | 0.6853 | 0.1359 | 0.1904 | 0.0605 | 0.9669 | 1 | 0.94 | 0.7164 | 0.795 | 0.6831 | | -0.3514 | 1.0561 | 0.9577 | 0.4547 | 0.4874 | 0.4217 | 0.6924 | 0.5808 | 0.6666 | 0.9452 | 0.9138 | 0.923 | 0.6546 | 0.6158 | 0.7272 | -0.4847 | -0.0256 | 0.7597 | 0.3748 |
| **L LVi Rad** | 0.3007 | 0.1903 | 0.4014 | 0.0378 | -0.0546 | 0.156 | 1.0011 | 0.94 | 1 | 0.5772 | 0.6038 | 0.5737 | | -0.3308 | 1.0514 | 0.8916 | 0.3451 | 0.357 | 0.2627 | 0.5221 | 0.4256 | 0.5181 | 0.9962 | 0.9624 | 0.9168 | 0.5128 | 0.4812 | 0.595 | -0.3526 | -0.1534 | 0.6472 | 0.5454 |
| **A LVi Tot** | 0.4322 | 0.3232 | 0.5013 | -0.1124 | -0.0822 | -0.1684 | 0.7138 | 0.7164 | 0.5772 | 1 | 1.0047 | 1.0017 | | -0.2092 | 0.3394 | 0.7987 | 0.2862 | 0.2551 | 0.211 | 0.3866 | 0.3047 | 0.3448 | 0.7018 | 0.6704 | 0.6132 | 0.7152 | 0.6373 | 0.7731 | -0.5808 | -0.2395 | 0.0709 | 0.4026 |
| **A LVi Circ** | 0.5823 | 0.4589 | 0.6543 | -0.024 | 0.0374 | -0.1472 | 0.7249 | 0.795 | 0.6038 | 1.0047 | 1 | 0.9859 | | -0.1747 | 0.5212 | 0.8463 | 0.373 | 0.352 | 0.3342 | 0.4958 | 0.3876 | 0.4468 | 0.828 | 0.7883 | 0.7873 | 0.8291 | 0.747 | 0.9216 | -0.6052 | -0.2188 | 0.4825 | 0.4794 |
| **A LVi Rad** | 0.4008 | 0.312 | 0.4722 | -0.0723 | -0.0782 | -0.1089 | 0.7327 | 0.6831 | 0.5737 | 1.0017 | 0.9859 | 1 | | -0.281 | 0.3811 | 0.7571 | 0.3277 | 0.2864 | 0.2632 | 0.4309 | 0.3517 | 0.3723 | 0.7101 | 0.6444 | 0.6135 | 0.6905 | 0.6366 | 0.7439 | -0.6386 | -0.4574 | 0.0066 | 0.4138 |
| **S LVm** | | | | | | | | | | | | | 1 | | | | | | | | | | | | | | | | | | | |
| **E LVm** | 0.1034 | 0.1071 | -0.0245 | 0.6322 | 0.5671 | 0.5954 | -0.3374 | -0.3514 | -0.3308 | -0.2092 | -0.1747 | -0.281 | | 1 | -0.2076 | -0.3269 | -0.272 | -0.2626 | -0.3144 | 0.1285 | 0.0498 | 0.1923 | -0.0958 | -0.1757 | 0.0164 | -0.3348 | -0.5007 | -0.3217 | 0.8381 | 0.3575 | -0.283 | -0.2327 |
| **L LVm** | 0.3913 | 0.3749 | 0.5351 | -0.1875 | -0.1181 | -0.1663 | 1.0671 | 1.0561 | 1.0514 | 0.3394 | 0.5212 | 0.3811 | | -0.2076 | 1 | 0.8803 | 0.2613 | 0.2911 | 0.3255 | 0.8233 | 0.5461 | 0.9485 | 0.9763 | 0.889 | 0.9568 | 0.5384 | 0.5923 | 0.5209 | -0.3507 | -0.1905 | 1.0669 | 0.5124 |
| **A LVm** | 0.841 | 0.6916 | 0.8113 | 0.3077 | 0.1914 | 0.1455 | 1.0556 | 0.9577 | 0.8916 | 0.7987 | 0.8463 | 0.7571 | | -0.3269 | 0.8803 | 1 | 0.474 | 0.4582 | 0.4094 | 0.8434 | 0.7973 | 0.7929 | 1.0131 | 0.8709 | 1.0284 | 0.5889 | 0.5278 | 0.6464 | -0.8933 | -0.1973 | 0.6989 | 0.3051 |
| **S RVi Tot** | 0.4797 | 0.4296 | 0.5756 | 0.0882 | 0.0751 | 0.0591 | 0.4394 | 0.4547 | 0.3451 | 0.2862 | 0.373 | 0.3277 | | -0.272 | 0.2613 | 0.474 | 1 | 0.9554 | 0.9823 | 0.5233 | 0.4728 | 0.3658 | 0.5759 | 0.6091 | 0.6045 | 0.7803 | 0.7035 | 0.7745 | -0.3442 | 0.0974 | 0.3337 | 0.3374 |
| **S RVi Circ** | 0.5452 | 0.4492 | 0.6664 | 0.1293 | 0.1105 | 0.0905 | 0.4449 | 0.4874 | 0.357 | 0.2551 | 0.352 | 0.2864 | | -0.2626 | 0.2911 | 0.4582 | 0.9554 | 1 | 0.9049 | 0.5595 | 0.538 | 0.3859 | 0.5644 | 0.6188 | 0.5885 | 0.7892 | 0.6866 | 0.7848 | -0.416 | 0.1263 | 0.377 | 0.3749 |
| **S RVi Rad** | 0.3479 | 0.344 | 0.4404 | 0.0662 | 0.0342 | 0.0073 | 0.3506 | 0.4217 | 0.2627 | 0.211 | 0.3342 | 0.2632 | | -0.3144 | 0.3255 | 0.4094 | 0.9823 | 0.9049 | 1 | 0.45 | 0.3992 | 0.3207 | 0.5161 | 0.5685 | 0.5481 | 0.7698 | 0.682 | 0.7661 | -0.1769 | 0.0668 | 0.2275 | 0.2918 |
| **E RVi Tot** | 0.7099 | 0.566 | 0.7389 | 0.5493 | 0.537 | 0.5107 | 0.573 | 0.6924 | 0.5221 | 0.3866 | 0.4958 | 0.4309 | | 0.1285 | 0.8233 | 0.8434 | 0.5233 | 0.5595 | 0.45 | 1 | 0.8742 | 0.968 | 0.8636 | 0.8252 | 0.8922 | 0.7705 | 0.6672 | 0.8443 | -0.077 | 0.3564 | 0.3072 | 0.7381 |
| **E RVi Circ** | 0.5431 | 0.3548 | 0.6306 | 0.4093 | 0.3496 | 0.3966 | 0.5229 | 0.5808 | 0.4256 | 0.3047 | 0.3876 | 0.3517 | | 0.0498 | 0.5461 | 0.7973 | 0.4728 | 0.538 | 0.3992 | 0.8742 | 1 | 0.7067 | 0.7515 | 0.7645 | 0.7346 | 0.6324 | 0.5757 | 0.6327 | -0.098 | 0.2807 | 0.3884 | 0.6523 |
| **E RVi Rad** | 0.6588 | 0.5565 | 0.6695 | 0.5279 | 0.5335 | 0.4482 | 0.5279 | 0.6666 | 0.5181 | 0.3448 | 0.4468 | 0.3723 | | 0.1923 | 0.9485 | 0.7929 | 0.3658 | 0.3859 | 0.3207 | 0.968 | 0.7067 | 1 | 0.7561 | 0.7331 | 0.8262 | 0.6138 | 0.5072 | 0.7079 | 0.1428 | 0.4058 | 0.376 | 0.5614 |
| **L RVi Tot** | 0.559 | 0.4322 | 0.6779 | 0.1627 | 0.1446 | 0.1228 | 0.9711 | 0.9452 | 0.9962 | 0.7018 | 0.828 | 0.7101 | | -0.0958 | 0.9763 | 1.0131 | 0.5759 | 0.5644 | 0.5161 | 0.8636 | 0.7515 | 0.7561 | 1 | 0.9944 | 1.0441 | 0.8076 | 0.7905 | 0.8036 | -0.6135 | -0.3221 | 0.4948 | 0.5468 |
| **L RVi Circ** | 0.5603 | 0.4585 | 0.6603 | 0.1527 | 0.1583 | 0.1054 | 1.0079 | 0.9138 | 0.9624 | 0.6704 | 0.7883 | 0.6444 | | -0.1757 | 0.889 | 0.8709 | 0.6091 | 0.6188 | 0.5685 | 0.8252 | 0.7645 | 0.7331 | 0.9944 | 1 | 0.983 | 0.8689 | 0.8426 | 0.8522 | -0.6297 | -0.35 | 0.4167 | 0.4478 |
| **L RVi Rad** | 0.5921 | 0.4949 | 0.6886 | 0.2243 | 0.1811 | 0.1359 | 0.8811 | 0.923 | 0.9168 | 0.6132 | 0.7873 | 0.6135 | | 0.0164 | 0.9568 | 1.0284 | 0.6045 | 0.5885 | 0.5481 | 0.8922 | 0.7346 | 0.8262 | 1.0441 | 0.983 | 1 | 0.8152 | 0.7961 | 0.858 | -0.3992 | -0.5055 | 0.4721 | 0.3465 |
| **A RVi Tot** | 0.5687 | 0.5126 | 0.6338 | 0.0762 | 0.1573 | -0.0025 | 0.6075 | 0.6546 | 0.5128 | 0.7152 | 0.8291 | 0.6905 | | -0.3348 | 0.5384 | 0.5889 | 0.7803 | 0.7892 | 0.7698 | 0.7705 | 0.6324 | 0.6138 | 0.8076 | 0.8689 | 0.8152 | 1 | 0.985 | 1 | -0.7529 | -0.1045 | 0.0937 | 0.4658 |
| **A RVi Circ** | 0.4725 | 0.3996 | 0.5825 | 0.0864 | 0.1437 | 0.0181 | 0.5636 | 0.6158 | 0.4812 | 0.6373 | 0.747 | 0.6366 | | -0.5007 | 0.5923 | 0.5278 | 0.7035 | 0.6866 | 0.682 | 0.6672 | 0.5757 | 0.5072 | 0.7905 | 0.8426 | 0.7961 | 0.985 | 1 | 0.9567 | -0.5424 | -0.1268 | 0.1604 | 0.4421 |
| **A RVi Rad** | 0.6029 | 0.5761 | 0.6377 | 0.0679 | 0.1664 | -0.0504 | 0.6566 | 0.7272 | 0.595 | 0.7731 | 0.9216 | 0.7439 | | -0.3217 | 0.5209 | 0.6464 | 0.7745 | 0.7848 | 0.7661 | 0.8443 | 0.6327 | 0.7079 | 0.8036 | 0.8522 | 0.858 | 1 | 0.9567 | 1 | -0.9438 | -0.1881 | 0.2271 | 0.477 |
| **S RVfw** | -0.3761 | -0.0699 | -0.4556 | 0.3132 | 0.3388 | 0.1979 | -0.3713 | -0.4847 | -0.3526 | -0.5808 | -0.6052 | -0.6386 | | 0.8381 | -0.3507 | -0.8933 | -0.3442 | -0.416 | -0.1769 | -0.077 | -0.098 | 0.1428 | -0.6135 | -0.6297 | -0.3992 | -0.7529 | -0.5424 | -0.9438 | 1 | 0.3882 | -0.8598 | -1.1952 |
| **E RVfw** | -0.1981 | 0.0287 | -0.2173 | 0.3692 | 0.2296 | 0.1233 | -0.1199 | -0.0256 | -0.1534 | -0.2395 | -0.2188 | -0.4574 | | 0.3575 | -0.1905 | -0.1973 | 0.0974 | 0.1263 | 0.0668 | 0.3564 | 0.2807 | 0.4058 | -0.3221 | -0.35 | -0.5055 | -0.1045 | -0.1268 | -0.1881 | 0.3882 | 1 | 0.1668 | -0.0754 |
| **L RVfw** | 0.2071 | 0.0327 | 0.2149 | -0.5812 | -0.5526 | -0.8882 | 0.7332 | 0.7597 | 0.6472 | 0.0709 | 0.4825 | 0.0066 | | -0.283 | 1.0669 | 0.6989 | 0.3337 | 0.377 | 0.2275 | 0.3072 | 0.3884 | 0.376 | 0.4948 | 0.4167 | 0.4721 | 0.0937 | 0.1604 | 0.2271 | -0.8598 | 0.1668 | 1 | 0.447 |
| **A RVfw** | 0.37 | 0.2309 | 0.4156 | -0.1628 | -0.0724 | -0.2446 | 0.2853 | 0.3748 | 0.5454 | 0.4026 | 0.4794 | 0.4138 | | -0.2327 | 0.5124 | 0.3051 | 0.3374 | 0.3749 | 0.2918 | 0.7381 | 0.6523 | 0.5614 | 0.5468 | 0.4478 | 0.3465 | 0.4658 | 0.4421 | 0.477 | -1.1952 | -0.0754 | 0.447 | 1 |

**Supplemental Table 8. Pairwise genetic correlation matrix for optical flow-derived cardiac velocity phenotypes.** CSV file containing genetic correlations (rg) estimated by cross-trait LD score regression for all 32 phenotypes. GWAS summary statistics were derived from up to 73,978 European-ancestry UK Biobank participants. Rows and columns are labeled by phenotype abbreviation: S/E/L/A = cardiac phase (systolic, early diastolic, mid-diastolic, late diastolic); LVi/LVm/RVi/RVfw = cardiac structure; Tot/Circ/Rad = velocity component.

| Phenotype | Phase | Region | λ | GW-Significant | N Significant SNPs | Top SNP | Top P-value |
|---|---|---|---|---|---|---|---|
| S-wave LV Inner Circumferential | S | LV Inner | 1.1 | Yes | 82 | 6:118691445:G:C | 4.70E-15 |
| S-wave LV Inner Radial | S | LV Inner | 1.08 | Yes | 36 | 6:118711303:G:C | 4.80E-14 |
| S-wave LV Inner Total | S | LV Inner | 1.09 | Yes | 72 | 6:118691445:G:C | 1.00E-13 |
| S-wave RV Inner Circumferential | S | RV Inner | 1.12 | PAR only | 3 | rs2259750 | 2.20E-12 |
| S-wave RV Inner Radial | S | RV Inner | 1.11 | No | 0 | 5:122481928:T:C | 4.50E-09 |
| S-wave RV Inner Total | S | RV Inner | 1.11 | PAR only | 3 | rs2259750 | 2.20E-10 |
| S-wave LV Myocardium Total | S | Myocardial | 1.02 | No | 0 | 17:64193824:T:C | 1.40E-06 |
| S-wave RV Myocardial Free Wall Total | S | Myocardial | 1.01 | No | 0 | 2:31204607:CA:C | 2.00E-08 |
| E-wave LV Inner Circumferential | E | LV Inner | 1.1 | Yes | 2 | 6:36645968:G:C | 3.20E-09 |
| E-wave LV Inner Radial | E | LV Inner | 1.1 | No | 0 | 10:19842386:C:G | 2.80E-07 |
| E-wave LV Inner Total | E | LV Inner | 1.09 | No | 0 | 3:153462759:G:A | 5.20E-07 |
| E-wave RV Inner Circumferential | E | RV Inner | 1.08 | No | 0 | rs11205277 | 3.00E-07 |
| E-wave RV Inner Radial | E | RV Inner | 1.07 | Yes | 1 | 6:118684824:A:T | 2.70E-09 |
| E-wave RV Inner Total | E | RV Inner | 1.08 | No | 0 | 22:40877004:T:C | 3.90E-07 |
| E-wave LV Myocardium Total | E | Myocardial | 1.01 | No | 0 | 4:39520527:T:C | 2.40E-07 |
| E-wave RV Myocardial Free Wall Total | E | Myocardial | 1.01 | No | 0 | 5:146712282:C:G | 6.10E-07 |
| L-wave LV Inner Circumferential | L | LV Inner | 1.1 | Yes | 257 | rs11153730 | 2.20E-17 |
| L-wave LV Inner Radial | L | LV Inner | 1.08 | Yes | 19 | 12:24760456:A:G | 8.70E-18 |
| L-wave LV Inner Total | L | LV Inner | 1.08 | Yes | 132 | 12:24760456:A:G | 6.60E-18 |
| L-wave RV Inner Circumferential | L | RV Inner | 1.09 | Yes | 123 | 12:24760456:A:G | 9.20E-15 |
| L-wave RV Inner Radial | L | RV Inner | 1.09 | Yes | 45 | 12:24760456:A:G | 5.70E-14 |
| L-wave RV Inner Total | L | RV Inner | 1.09 | Yes | 63 | rs17287293 | 4.50E-17 |
| L-wave LV Myocardium Total | L | Myocardial | 1.03 | No | 0 | rs17180489 | 3.90E-09 |
| L-wave RV Myocardial Free Wall Total | L | Myocardial | 1.01 | No | 0 | rs115749119 | 5.50E-07 |
| A-wave LV Inner Circumferential | A | LV Inner | 1.08 | Yes | 215 | rs11153730 | 4.10E-22 |
| A-wave LV Inner Radial | A | LV Inner | 1.09 | Yes | 49 | rs12210810 | 5.90E-12 |
| A-wave LV Inner Total | A | LV Inner | 1.09 | Yes | 89 | 6:118691445:G:C | 1.10E-14 |
| A-wave RV Inner Circumferential | A | RV Inner | 1.07 | Yes | 5 | rs10497529 | 1.80E-10 |
| A-wave RV Inner Radial | A | RV Inner | 1.05 | Yes | 7 | rs12210810 | 4.10E-10 |
| A-wave RV Inner Total | A | RV Inner | 1.07 | Yes | 9 | rs12210810 | 4.70E-10 |
| A-wave LV Myocardium Total | A | Myocardial | 1.03 | No | 0 | 3:38777554:T:G | 2.80E-08 |

| | | | | | | | |
|---|---|---|---|---|---|---|---|
| A-wave RV Myocardial Free Wall Total | A | Myocardial | 1.03 | No | 0 | 10:4237930:T:C | 2.90E-07 |

**Supplemental Table 9. GWAS summary for all 32 optical flow phenotypes.** Nineteen of 32 optical flow velocity phenotypes yielded at least one genome-wide significant association after cluster-corrected Bonferroni correction ($p < 3.85 \times 10^{-9}$; see Methods), including 17 phenotypes with autosomal hits and 2 phenotypes (S-wave RV inner circumferential and S-wave RV inner total) with associations only in the pseudoautosomal region. The 17 phenotypes with autosomal hits comprised 6 mid-diastolic (L-wave) inner cavity phenotypes, 6 late diastolic (A-wave) inner cavity phenotypes, 3 systolic (S-wave) inner cavity phenotypes, and 2 early diastolic (E-wave) inner cavity phenotypes (Supplemental Figure 7). No associations with myocardial velocity phenotypes reached genome-wide significance, consistent with their lower heritability. Phase: S = systolic, E = early diastolic (E-wave), L = mid-diastolic (L-wave), A = late diastolic (A-wave). Region: LV = left ventricle, RV = right ventricle. λ = genomic inflation factor. GW-Significant = phenotypes with at least one SNP reaching genome-wide significance after cluster-corrected Bonferroni correction for multiple testing across correlated phenotypes (see Methods). PAR only = associations in the pseudoautosomal region only. N Significant SNPs = count at cluster-corrected threshold. Top SNP and Top P-value = lead SNP with the lowest p-value for each phenotype and its corresponding p-value, reported regardless of whether genome-wide significance was reached. Sample sizes ranged from 73,590 to 73,978 depending on phenotype availability. PAR = pseudoautosomal region.

| Table_1_Locus | Table_1_Mapped_Gene | GWAS_Catalog_Trait | PMID | PMID_Link | First_Author | Year | GWAS_Catalog_Mapped_Gene |
|---|---|---|---|---|---|---|---|
| 1 | TTN | Cardiorespiratory fitness (vo2 max) | 34753499 | 34753499 | Hanscombe KB | 2021 | CCDC141 |
| 1 | TTN | Diastolic blood pressure | 35762941 | 35762941 | Plotnikov D | 2022 | CCDC141 |
| 1 | TTN | Electrocardiogram morphology (amplitude at temporal datapoints) | 32916098 | 32916098 | Verweij N | 2020 | CCDC141 |
| 1 | TTN | Heart rate | 23583979 | 23583979 | den Hoed M | 2013 | CCDC141 |
| 1 | TTN | Heart rate response to exercise | 29769521 | 29769521 | Ramirez J | 2018 | CCDC141 |
| 1 | TTN | Heart rate response to recovery post exercise (10 sec) | 29497042 | 29497042 | Verweij N | 2018 | CCDC141 |
| 1 | TTN | Hypertrophic cardiomyopathy (sarcomere negative) | 33495597 | 33495597 | Harper AR | 2021 | CCDC141 |
| 1 | TTN | Predicted heart recovery rate | 36046430 | 36046430 | Diamant N | 2022 | CCDC141 |
| 1 | TTN | Pulse pressure | 34594039 | 34594039 | Sakaue S | 2021 | CCDC141 |
| 1 | TTN | QRS duration | 36050321 | 36050321 | Young WJ | 2022 | CCDC141 |
| 1 | TTN | QTc interval | 35389749 | 35389749 | Nauffal V | 2022 | CCDC141 |
| 1 | TTN | Resting heart rate | 27798624 | 27798624 | Eppinga RN | 2016 | CCDC141 |
| 2 | SCN10A | Atrial fibrillation | 30061737 | 30061737 | Nielsen JB | 2018 | SCN10A |
| 2 | SCN10A | Atrial fibrillation (MTAG) | 35872910 | 35872910 | Carcel-Marquez J | 2022 | SCN10A |
| 2 | SCN10A | Atrial fibrillation/atrial flutter | 34594039 | 34594039 | Sakaue S | 2021 | SCN10A |
| 2 | SCN10A | Atrioventricular conduction | 21041692 | 21041692 | Denny JC | 2010 | SCN10A |
| 2 | SCN10A | Brugada syndrome | 35210625 | 35210625 | Barc J | 2022 | SCN10A |
| 2 | SCN10A | Cardiorespiratory fitness (vo2 max) | 34753499 | 34753499 | Hanscombe KB | 2021 | SCN10A |
| 2 | SCN10A | Electrocardiogram morphology (amplitude at temporal datapoints) | 32916098 | 32916098 | Verweij N | 2020 | SCN10A |
| 2 | SCN10A | Electrocardiographic conduction measures | 23463857 | 23463857 | Ritchie MD | 2013 | SCN10A |
| 2 | SCN10A | Electrocardiographic traits | 20062061 | 20062061 | Chambers JC | 2010 | SCN10A |
| 2 | SCN10A | Electrocardiographic traits (multivariate) | 32602732 | 32602732 | Baldassari AR | 2020 | SCN10A |
| 2 | SCN10A | Heart rate response to exercise | 29769521 | 29769521 | Ramirez J | 2018 | SCN10A |
| 2 | SCN10A | Heart rate response to recovery post exercise | 29769521 | 29769521 | Ramirez J | 2018 | SCN10A |
| 2 | SCN10A | P wave duration | 28794112 | 28794112 | Christophersen IE | 2017 | SCN10A |
| 2 | SCN10A | PR interval | 30046033 | 30046033 | van Setten J | 2018 | SCN10A |
| 2 | SCN10A | PR segment duration | 24850809 | 24850809 | Verweij N | 2014 | SCN10A |
| 2 | SCN10A | Pulse pressure | 30224653 | 30224653 | Evangelou E | 2018 | SCN10A |

| | | | | | | | |
|---|---|---|---|---|---|---|---|
| 2 | SCN10A | QRS duration | 30012220 | 30012220 | Prins BP | 2018 | SCN10A |
| 2 | SCN10A | QT interval | 24952745 | 24952745 | Arking DE | 2014 | SCN10A |
| 2 | SCN10A | Resting heart rate | 27798624 | 27798624 | Eppinga RN | 2016 | SCN10A |
| 2 | SCN10A | Sick sinus syndrome | 33580673 | 33580673 | Thorolfsdottir RB | 2021 | SCN10A |
| 2 | SCN10A | T wave morphology restitution during exercise | 31607149 | 31607149 | Ramirez J | 2019 | SCN10A |
| 2 | SCN10A | T wave morphology restitution during exercise (MTAG) | 31607149 | 31607149 | Ramirez J | 2019 | SCN10A |
| 3 | NKX2-5 | Electrocardiogram morphology (amplitude at temporal datapoints) | 32916098 | 32916098 | Verweij N | 2020 | BNIP1 - RPL7AP33 |
| 3 | NKX2-5 | JT interval | 36050321 | 36050321 | Young WJ | 2022 | BNIP1 - RPL7AP33 |
| 3 | NKX2-5 | QT interval | 36050321 | 36050321 | Young WJ | 2022 | BNIP1 - RPL7AP33 |
| 3 | NKX2-5 | Resting heart rate | 27798624 | 27798624 | Eppinga RN | 2016 | BNIP1 - RPL7AP33 |
| 4 | NUDT3 | Ascending aorta maximum area | 34968759 | 34968759 | Benjamins JW | 2021 | DINOL, CDKN1A |
| 4 | NUDT3 | Ascending thoracic aortic diameter | 34837083 | 34837083 | Pirruccello JP | 2021 | RNU1-88P - Y_RNA |
| 4 | NUDT3 | Atrial fibrillation | 30061737 | 30061737 | Nielsen JB | 2018 | CDKN1A |
| 4 | NUDT3 | Atrial fibrillation (MTAG) | 35872910 | 35872910 | Carcel-Marquez J | 2022 | CDKN1A |
| 4 | NUDT3 | Atrial fibrillation/atrial flutter | 34594039 | 34594039 | Sakaue S | 2021 | CDKN1A, DINOL |
| 4 | NUDT3 | Cardioembolic stroke (MTAG) | 35872910 | 35872910 | Carcel-Marquez J | 2022 | CDKN1A |
| 4 | NUDT3 | Electrocardiogram morphology (amplitude at temporal datapoints) | 32916098 | 32916098 | Verweij N | 2020 | CDKN1A, DINOL |
| 4 | NUDT3 | Electrocardiographic conduction measures | 23463857 | 23463857 | Ritchie MD | 2013 | RNU1-88P - Y_RNA |
| 4 | NUDT3 | Electrocardiographic traits | 20062063 | 20062063 | Holm H | 2010 | RNU1-88P - Y_RNA |
| 4 | NUDT3 | Electrocardiographic traits (multivariate) | 32602732 | 32602732 | Baldassari AR | 2020 | CDKN1A |
| 4 | NUDT3 | Hypertrophic cardiomyopathy | 33495596 | 33495596 | Tadros R | 2021 | RNU1-88P - Y_RNA |
| 4 | NUDT3 | Hypertrophic cardiomyopathy (MTAG) | 33495596 | 33495596 | Tadros R | 2021 | RNU1-88P - Y_RNA |
| 4 | NUDT3 | Hypertrophic cardiomyopathy (sarcomere negative) | 33495597 | 33495597 | Harper AR | 2021 | CDKN1A |
| 4 | NUDT3 | Ischemic stroke | 29531354 | 29531354 | Malik R | 2018 | CDKN1A, DINOL |
| 4 | NUDT3 | JT interval | 36050321 | 36050321 | Young WJ | 2022 | CDKN1A |
| 4 | NUDT3 | Left ventricle wall thickness | 33495596 | 33495596 | Tadros R | 2021 | RNU1-88P - Y_RNA |
| 4 | NUDT3 | Left ventricular fractional shortening | 28394258 | 28394258 | Wild PS | 2017 | RNU1-88P - Y_RNA |
| 4 | NUDT3 | Left ventricular global circumferential strain | 33495596 | 33495596 | Tadros R | 2021 | RNU1-88P - Y_RNA |

| | | | | | | | |
|---|---|---|---|---|---|---|---|
| 4 | NUDT3 | Left ventricular global radial strain | 33495596 | 33495596 | Tadros R | 2021 | RNU1-88P - Y_RNA |
| 4 | NUDT3 | Left ventricular mass to end-diastolic volume ratio | 33495596 | 33495596 | Tadros R | 2021 | RNU1-88P - Y_RNA |
| 4 | NUDT3 | PR interval | 32439900 | 32439900 | Ntalla I | 2020 | CDKN1A, DINOL |
| 4 | NUDT3 | QRS complex (Cornell) | 27659466 | 27659466 | van der Harst P | 2016 | RNU1-88P - Y_RNA |
| 4 | NUDT3 | QRS duration | 36050321 | 36050321 | Young WJ | 2022 | CDKN1A |
| 4 | NUDT3 | QT dynamics during exercise | 32527199 | 32527199 | van Duijvenboden S | 2020 | RNU1-88P - Y_RNA |
| 4 | NUDT3 | QT interval | 36050321 | 36050321 | Young WJ | 2022 | CDKN1A |
| 5 | PLN | Atrial fibrillation | 30061737 | 30061737 | Nielsen JB | 2018 | SLC35F1 |
| 5 | PLN | Atrial fibrillation (MTAG) | 35872910 | 35872910 | Carcel-Marquez J | 2022 | SLC35F1 |
| 5 | PLN | Atrial fibrillation/atrial flutter | 34594039 | 34594039 | Sakaue S | 2021 | SLC35F1 - CEP85L |
| 5 | PLN | Cardiac structure and function | 19584346 | 19584346 | Vasan RS | 2009 | SLC35F1 |
| 5 | PLN | Diastolic blood pressure | 35762941 | 35762941 | Plotnikov D | 2022 | SLC35F1 - CEP85L |
| 5 | PLN | Electrocardiogram morphology (amplitude at temporal datapoints) | 32916098 | 32916098 | Verweij N | 2020 | SLC35F1 - CEP85L |
| 5 | PLN | Electrocardiographic conduction measures | 23463857 | 23463857 | Ritchie MD | 2013 | CEP85L |
| 5 | PLN | Electrocardiographic traits | 20062063 | 20062063 | Holm H | 2010 | SLC35F1 - CEP85L |
| 5 | PLN | Electrocardiographic traits (multivariate) | 32602732 | 32602732 | Baldassari AR | 2020 | NA - CEP85L |
| 5 | PLN | Heart rate | 23583979 | 23583979 | den Hoed M | 2013 | SLC35F1 - CEP85L |
| 5 | PLN | Hypertrophic cardiomyopathy | 33495596 | 33495596 | Tadros R | 2021 | SLC35F1 - CEP85L |
| 5 | PLN | Hypertrophic cardiomyopathy (MTAG) | 33495596 | 33495596 | Tadros R | 2021 | SLC35F1 - CEP85L |
| 5 | PLN | Hypertrophic cardiomyopathy (sarcomere negative) | 33495597 | 33495597 | Harper AR | 2021 | SLC35F1 - NA |
| 5 | PLN | JT interval | 36050321 | 36050321 | Young WJ | 2022 | SLC35F1 - CEP85L |
| 5 | PLN | Left ventricular global longitudinal strain | 33495596 | 33495596 | Tadros R | 2021 | SLC35F1 - NA |
| 5 | PLN | Longitudinal peak diastolic strain rate | 35479509 | 35479509 | Thanaj M | 2022 | SLC35F1 - CEP85L |
| 5 | PLN | Myocardial fractal dimension (slice 5) | 32814899 | 32814899 | Meyer HV | 2020 | SLC35F1 - CEP85L |
| 5 | PLN | Myocardial fractal dimension (slice 6) | 32814899 | 32814899 | Meyer HV | 2020 | SLC35F1 - CEP85L |
| 5 | PLN | Myocardial fractal dimension (slice 7) | 32814899 | 32814899 | Meyer HV | 2020 | SLC35F1 - CEP85L |
| 5 | PLN | Myocardial fractal dimension (slice 8) | 32814899 | 32814899 | Meyer HV | 2020 | SLC35F1 - CEP85L |
| 5 | PLN | PR interval | 32439900 | 32439900 | Ntalla I | 2020 | SLC35F1 - CEP85L |
| 5 | PLN | Pulse rate | 36329257 | 36329257 | Lee CJ | 2022 | SLC35F1 - CEP85L |
| 5 | PLN | QRS complex (Sokolow-Lyon) | 27659466 | 27659466 | van der Harst P | 2016 | SLC35F1 - CEP85L |

| | | | | | | | |
|---|---|---|---|---|---|---|---|
| 5 | PLN | QRS duration | 36050321 | 36050321 | Young WJ | 2022 | SLC35F1 - CEP85L |
| 5 | PLN | QT dynamics during exercise | 32527199 | 32527199 | van Duijvenboden S | 2020 | SLC35F1 - NA |
| 5 | PLN | QT interval | 36050321 | 36050321 | Young WJ | 2022 | SLC35F1 - CEP85L |
| 5 | PLN | QTc interval | 35389749 | 35389749 | Nauffal V | 2022 | SLC35F1 - CEP85L |
| 5 | PLN | RR interval (heart rate) | 30679814 | 30679814 | van Setten J | 2019 | CEP85L |
| 5 | PLN | Resting heart rate | 27798624 | 27798624 | Eppinga RN | 2016 | SLC35F1 |
| 5 | PLN | Right ventricular stroke volume | 35697868 | 35697868 | Aung N | 2022 | SLC35F1 |
| 5 | PLN | TPE interval (resting) | 32386560 | 32386560 | Ramirez J | 2020 | SLC35F1 - CEP85L |
| 6 | GJA1 | Atrial fibrillation | 30061737 | 30061737 | Nielsen JB | 2018 | RPL23AP48 - HMGB3P18 |
| 6 | GJA1 | Descending aorta distensibility (MTAG) | 35922433 | 35922433 | Francis CM | 2022 | RPL23AP48 - HMGB3P18 |
| 6 | GJA1 | Diastolic blood pressure | 30578418 | 30578418 | Giri A | 2018 | NA - HMGB3P18 |
| 6 | GJA1 | Heart rate | 23583979 | 23583979 | den Hoed M | 2013 | NA - HMGB3P18 |
| 6 | GJA1 | PR interval | 32439900 | 32439900 | Ntalla I | 2020 | NA - HMGB3P18 |
| 6 | GJA1 | Pulse pressure | 34594039 | 34594039 | Sakaue S | 2021 | HMGB3P18 - RNU1-18P |
| 6 | GJA1 | Pulse rate | 36329257 | 36329257 | Lee CJ | 2022 | RPL23AP48 - HMGB3P18 |
| 6 | GJA1 | RR interval (heart rate) | 30679814 | 30679814 | van Setten J | 2019 | NA - HMGB3P18 |
| 6 | GJA1 | Radial peak diastolic strain rate | 35479509 | 35479509 | Thanaj M | 2022 | HMGB3P18 - RNU1-18P |
| 6 | GJA1 | Resting heart rate | 27798624 | 27798624 | Eppinga RN | 2016 | NA - HMGB3P18 |
| 7 | GATA4* | Diastolic blood pressure | 35762941 | 35762941 | Plotnikov D | 2022 | OR7E161P - DEFB136 |
| 7 | GATA4* | Dilated cardiomyopathy (MTAG) | 33495596 | 33495596 | Tadros R | 2021 | OR7E158P - OR7E161P |
| 7 | GATA4* | Electrocardiogram morphology (amplitude at temporal datapoints) | 32916098 | 32916098 | Verweij N | 2020 | OR7E161P - DEFB136 |
| 7 | GATA4* | Essential hypertension (time to event) | 32589924 | 32589924 | Bi W | 2020 | OR7E161P - DEFB136 |
| 7 | GATA4* | Pulse pressure | 35762941 | 35762941 | Plotnikov D | 2022 | OR7E161P - DEFB136 |
| 7 | GATA4* | Systolic blood pressure | 35762941 | 35762941 | Plotnikov D | 2022 | OR7E161P - DEFB136 |
| 8 | SOX5 | Atrial fibrillation | 29892015 | 29892015 | Roselli C | 2018 | KNOP1P1 - RN7SL38P |
| 8 | SOX5 | Atrial fibrillation (MTAG) | 35872910 | 35872910 | Carcel-Marquez J | 2022 | KNOP1P1 - RN7SL38P |
| 8 | SOX5 | Atrial fibrillation/atrial flutter | 34594039 | 34594039 | Sakaue S | 2021 | KNOP1P1 - RN7SL38P |
| 8 | SOX5 | Cardiorespiratory fitness slope | 34753499 | 34753499 | Hanscombe KB | 2021 | KNOP1P1 - RN7SL38P |
| 8 | SOX5 | Diastolic blood pressure | 33230300 | 33230300 | Surendran P | 2020 | KNOP1P1 - RN7SL38P |
| 8 | SOX5 | Electrocardiogram morphology (amplitude at temporal datapoints) | 32916098 | 32916098 | Verweij N | 2020 | KNOP1P1 - RN7SL38P |
| 8 | SOX5 | Electrocardiographic traits (multivariate) | 32602732 | 32602732 | Baldassari AR | 2020 | KNOP1P1 - RN7SL38P |

| | | | | | | | |
|---|---|---|---|---|---|---|---|
| 8 | SOX5 | Heart rate | 23583979 | 23583979 | den Hoed M | 2013 | KNOP1P1 - RN7SL38P |
| 8 | SOX5 | Heart rate increase in response to exercise | 29497042 | 29497042 | Verweij N | 2018 | KNOP1P1 - RN7SL38P |
| 8 | SOX5 | Heart rate response to exercise | 29769521 | 29769521 | Ramirez J | 2018 | KNOP1P1 - RN7SL38P |
| 8 | SOX5 | Heart rate response to recovery post exercise | 29769521 | 29769521 | Ramirez J | 2018 | KNOP1P1 - RN7SL38P |
| 8 | SOX5 | Heart rate response to recovery post exercise (10 sec) | 29497042 | 29497042 | Verweij N | 2018 | KNOP1P1 - RN7SL38P |
| 8 | SOX5 | Heart rate response to recovery post exercise (20 sec) | 29497042 | 29497042 | Verweij N | 2018 | KNOP1P1 - RN7SL38P |
| 8 | SOX5 | Heart rate response to recovery post exercise (30 sec) | 29497042 | 29497042 | Verweij N | 2018 | KNOP1P1 - RN7SL38P |
| 8 | SOX5 | Heart rate response to recovery post exercise (40 sec) | 29497042 | 29497042 | Verweij N | 2018 | KNOP1P1 - RN7SL38P |
| 8 | SOX5 | Heart rate response to recovery post exercise (50 sec) | 29497042 | 29497042 | Verweij N | 2018 | KNOP1P1 - RN7SL38P |
| 8 | SOX5 | Heart rate variability traits (RMSSD) | 28613276 | 28613276 | Nolte IM | 2017 | KNOP1P1 - RN7SL38P |
| 8 | SOX5 | Heart rate variability traits (SDNN) | 28613276 | 28613276 | Nolte IM | 2017 | KNOP1P1 - RN7SL38P |
| 8 | SOX5 | Heart rate variability traits (pvRSA/HF) | 28613276 | 28613276 | Nolte IM | 2017 | KNOP1P1 - RN7SL38P |
| 8 | SOX5 | PR interval | 32439900 | 32439900 | Ntalla I | 2020 | KNOP1P1 - RN7SL38P |
| 8 | SOX5 | Predicted heart recovery rate | 36046430 | 36046430 | Diamant N | 2022 | KNOP1P1 - RN7SL38P |
| 8 | SOX5 | Pulse pressure | 33230300 | 33230300 | Surendran P | 2020 | KNOP1P1 - RN7SL38P |
| 8 | SOX5 | Pulse rate | 36329257 | 36329257 | Lee CJ | 2022 | KNOP1P1 - RN7SL38P |
| 8 | SOX5 | Resting heart rate | 27798624 | 27798624 | Eppinga RN | 2016 | KNOP1P1 - RN7SL38P |
| 9 | PKP2 | Cardiorespiratory fitness slope | 34753499 | 34753499 | Hanscombe KB | 2021 | SYT10 - RNU6-400P |
| 9 | PKP2 | Descending aorta distensibility (MTAG) | 35922433 | 35922433 | Francis CM | 2022 | SYT10 - RNU6-400P |
| 9 | PKP2 | Diastolic blood pressure | 33230300 | 33230300 | Surendran P | 2020 | ASS1P14 - SYT10 |
| 9 | PKP2 | Electrocardiogram morphology (amplitude at temporal datapoints) | 32916098 | 32916098 | Verweij N | 2020 | SYT10 |
| 9 | PKP2 | Heart rate | 23583979 | 23583979 | den Hoed M | 2013 | SYT10 |
| 9 | PKP2 | Heart rate increase in response to exercise | 29497042 | 29497042 | Verweij N | 2018 | SYT10 - NA |
| 9 | PKP2 | Heart rate response to exercise | 29769521 | 29769521 | Ramirez J | 2018 | SYT10 |
| 9 | PKP2 | Heart rate response to recovery post exercise | 29769521 | 29769521 | Ramirez J | 2018 | RNU6-400P - NA |
| 9 | PKP2 | Heart rate response to recovery post exercise (10 sec) | 29497042 | 29497042 | Verweij N | 2018 | SYT10 - NA |

| | | | | | | | |
|---|---|---|---|---|---|---|---|
| 9 | PKP2 | Heart rate response to recovery post exercise (20 sec) | 29497042 | 29497042 | Verweij N | 2018 | SYT10 - NA |
| 9 | PKP2 | Heart rate response to recovery post exercise (30 sec) | 29497042 | 29497042 | Verweij N | 2018 | SYT10 - NA |
| 9 | PKP2 | Heart rate response to recovery post exercise (40 sec) | 29497042 | 29497042 | Verweij N | 2018 | SYT10 - NA |
| 9 | PKP2 | Heart rate response to recovery post exercise (50 sec) | 29497042 | 29497042 | Verweij N | 2018 | SYT10 - NA |
| 9 | PKP2 | Heart rate variability traits (RMSSD) | 28613276 | 28613276 | Nolte IM | 2017 | SYT10 |
| 9 | PKP2 | Heart rate variability traits (SDNN) | 28613276 | 28613276 | Nolte IM | 2017 | SYT10 - NA |
| 9 | PKP2 | Heart rate variability traits (pvRSA/HF) | 28613276 | 28613276 | Nolte IM | 2017 | SYT10 - NA |
| 9 | PKP2 | Longitudinal peak diastolic strain rate | 35479509 | 35479509 | Thanaj M | 2022 | SYT10 - RNU6-400P |
| 9 | PKP2 | Pulse pressure | 33230300 | 33230300 | Surendran P | 2020 | ASS1P14 - SYT10 |
| 9 | PKP2 | RR interval (heart rate) | 30679814 | 30679814 | van Setten J | 2019 | SYT10 |
| 9 | PKP2 | Radial peak diastolic strain rate | 35479509 | 35479509 | Thanaj M | 2022 | SYT10 - RNU6-400P |
| 9 | PKP2 | Resting heart rate | 27798624 | 27798624 | Eppinga RN | 2016 | SYT10 - RNU6-400P |
| 10 | MYH6/MYH7 | Atrial fibrillation | 30061737 | 30061737 | Nielsen JB | 2018 | MYH6 |
| 10 | MYH6/MYH7 | Atrial fibrillation/atrial flutter | 34594039 | 34594039 | Sakaue S | 2021 | MYH6 |
| 10 | MYH6/MYH7 | Cardiac dysrhythmias (time to event) | 32589924 | 32589924 | Bi W | 2020 | MYH6 |
| 10 | MYH6/MYH7 | Descending thoracic aortic diameter | 34837083 | 34837083 | Pirruccello JP | 2021 | MYH6 |
| 10 | MYH6/MYH7 | Diastolic blood pressure | 30578418 | 30578418 | Giri A | 2018 | MYH6 |
| 10 | MYH6/MYH7 | Electrocardiogram morphology (amplitude at temporal datapoints) | 32916098 | 32916098 | Verweij N | 2020 | MYH6 |
| 10 | MYH6/MYH7 | Electrocardiographic traits | 20062063 | 20062063 | Holm H | 2010 | MYH6 |
| 10 | MYH6/MYH7 | Electrocardiographic traits (multivariate) | 32602732 | 32602732 | Baldassari AR | 2020 | MYH6 |
| 10 | MYH6/MYH7 | Heart rate | 23583979 | 23583979 | den Hoed M | 2013 | MYH6 |
| 10 | MYH6/MYH7 | P wave duration | 28794112 | 28794112 | Christophersen IE | 2017 | MYH6 |
| 10 | MYH6/MYH7 | Predicted heart recovery rate | 36046430 | 36046430 | Diamant N | 2022 | MYH6 |
| 10 | MYH6/MYH7 | Pulse pressure | 34594039 | 34594039 | Sakaue S | 2021 | MYH6 |
| 10 | MYH6/MYH7 | Pulse rate | 36329257 | 36329257 | Lee CJ | 2022 | MYH6 |
| 10 | MYH6/MYH7 | Resting heart rate | 27798624 | 27798624 | Eppinga RN | 2016 | MYH6 |
| 10 | MYH6/MYH7 | Systolic blood pressure | 34594039 | 34594039 | Sakaue S | 2021 | MYH6 |
| 11 | HCN4 | Atrial fibrillation | 30061737 | 30061737 | Nielsen JB | 2018 | HCN4 - REC114 |
| 11 | HCN4 | Atrial fibrillation (MTAG) | 35872910 | 35872910 | Carcel-Marquez J | 2022 | HCN4 - REC114 |

| | | | | | | | |
|---|---|---|---|---|---|---|---|
| 11 | HCN4 | Atrial fibrillation/atrial flutter | 34594039 | 34594039 | Sakaue S | 2021 | HCN4 - REC114 |
| 11 | HCN4 | Cardioembolic stroke (MTAG) | 35872910 | 35872910 | Carcel-Marquez J | 2022 | HCN4 - REC114 |
| 11 | HCN4 | Electrocardiogram morphology (amplitude at temporal datapoints) | 32916098 | 32916098 | Verweij N | 2020 | HCN4 |
| 11 | HCN4 | Heart rate | 23583979 | 23583979 | den Hoed M | 2013 | HCN4 - REC114 |
| 11 | HCN4 | Heart rate variability traits (RMSSD) | 28613276 | 28613276 | Nolte IM | 2017 | HCN4 |
| 11 | HCN4 | Heart rate variability traits (SDNN) | 28613276 | 28613276 | Nolte IM | 2017 | HCN4 |
| 11 | HCN4 | Heart rate variability traits (pvRSA/HF) | 28613276 | 28613276 | Nolte IM | 2017 | HCN4 |
| 11 | HCN4 | PR interval | 32439900 | 32439900 | Ntalla I | 2020 | HCN4 - REC114 |
| 11 | HCN4 | Resting heart rate | 27798624 | 27798624 | Eppinga RN | 2016 | HCN4 - REC114 |
| 12 | VSTM2L | Cardioembolic stroke (MTAG) | 35872910 | 35872910 | Carcel-Marquez J | 2022 | KIAA1755 |
| 12 | VSTM2L | Cardiorespiratory fitness (vo2 max) | 34753499 | 34753499 | Hanscombe KB | 2021 | KIAA1755 |
| 12 | VSTM2L | Diastolic blood pressure | 33230300 | 33230300 | Surendran P | 2020 | KIAA1755 |
| 12 | VSTM2L | Electrocardiogram morphology (amplitude at temporal datapoints) | 32916098 | 32916098 | Verweij N | 2020 | KIAA1755 |
| 12 | VSTM2L | Heart rate | 23583979 | 23583979 | den Hoed M | 2013 | KIAA1755 |
| 12 | VSTM2L | Heart rate variability traits (RMSSD) | 28613276 | 28613276 | Nolte IM | 2017 | KIAA1755 |
| 12 | VSTM2L | Heart rate variability traits (SDNN) | 28613276 | 28613276 | Nolte IM | 2017 | KIAA1755 |
| 12 | VSTM2L | PR interval | 32439900 | 32439900 | Ntalla I | 2020 | KIAA1755 |
| 12 | VSTM2L | Predicted heart recovery rate | 36046430 | 36046430 | Diamant N | 2022 | KIAA1755 |
| 12 | VSTM2L | Pulse pressure | 30224653 | 30224653 | Evangelou E | 2018 | KIAA1755 |
| 12 | VSTM2L | QT dynamics during exercise | 32527199 | 32527199 | van Duijvenboden S | 2020 | TGM2 - KIAA1755 |
| 12 | VSTM2L | RR interval (heart rate) | 30679814 | 30679814 | van Setten J | 2019 | KIAA1755 |
| 12 | VSTM2L | Resting heart rate | 27798624 | 27798624 | Eppinga RN | 2016 | KIAA1755 |

**Supplemental Table 10. Previously reported GWAS Catalog associations at genomic risk loci identified in the optical flow velocity phenotype GWAS.** For each of the 12 genomic risk loci in Table 2, variants in linkage disequilibrium with the locus lead SNP were queried against the GWAS Catalog using FUMA. Associations were filtered to cardiovascular-relevant traits including heart rate, blood pressure, cardiac conduction intervals, atrial fibrillation, cardiac structure and function, and related electrophysiological and haemodynamic phenotypes. Table 2 Mapped Gene reflects the prioritized gene at each locus based on positional mapping. *GATA4 is approximately 160 kb downstream of the locus lead SNP, the nearest mapped transcript is AF131215.9. eQTL = expression quantitative trait locus; GWAS = genome wide association study; SNP = single nucleotide polymorphism.

| Exposure | Outcome | n SNPs | IVW β | IVW SE | IVW P | Weighted median β | Weighted median SE | Weighted median P | Egger β | Egger SE | Egger P | Egger intercept | Egger intercept SE | Egger intercept P | MR-PRESSO global P | MR-PRESSO n outliers | MR-PRESSO corrected β | MR-PRESSO corrected P |
|---|---|---|---|---|---|---|---|---|---|---|---|---|---|---|---|---|---|---|
| RABGAP1L | E-wave RV Inner Circumferential | 15 | -0.255 | 0.026 | 5.50E-22 | -0.261 | 0.035 | 7.00E-14 | -0.265 | 0.096 | 0.017 | 0.001 | 0.007 | 0.92 | 0.98 | 0 | NA | NA |
| HCG22 | A-wave RV Inner Circumferential | 74 | -0.061 | 0.007 | 7.90E-18 | -0.064 | 0.011 | 4.60E-09 | -0.005 | 0.019 | 0.77 | -0.011 | 0.004 | 0.0029 | 0.084 | 0 | NA | NA |
| RABGAP1L | E-wave RV Inner Total | 15 | -0.225 | 0.026 | 1.70E-17 | -0.239 | 0.033 | 9.40E-13 | -0.173 | 0.096 | 0.095 | -0.004 | 0.007 | 0.58 | 1 | 0 | NA | NA |
| RABGAP1L | E-wave RV Inner Radial | 15 | -0.206 | 0.026 | 8.00E-15 | -0.206 | 0.034 | 2.00E-09 | -0.2 | 0.096 | 0.059 | 0 | 0.007 | 0.95 | 1 | 0 | NA | NA |
| RABGAP1L | E-wave LV Inner Radial | 15 | -0.204 | 0.026 | 8.50E-15 | -0.236 | 0.036 | 5.60E-11 | -0.106 | 0.099 | 0.3 | -0.007 | 0.007 | 0.32 | 0.52 | 0 | NA | NA |
| HLA-E | S-wave RV Inner Radial | 43 | 0.043 | 0.006 | 2.20E-14 | 0.059 | 0.008 | 1.50E-14 | 0.081 | 0.014 | 2.20E-06 | -0.01 | 0.004 | 0.01 | 0.1 | 0 | NA | NA |
| HCG22 | A-wave RV Inner Total | 74 | -0.054 | 0.007 | 2.30E-14 | -0.054 | 0.011 | 1.00E-06 | 0.002 | 0.018 | 0.91 | -0.011 | 0.003 | 0.0022 | 0.12 | 0 | NA | NA |
| HCG22 | A-wave RV Inner Radial | 74 | -0.051 | 0.007 | 4.80E-13 | -0.043 | 0.011 | 0.00017 | -0.023 | 0.02 | 0.26 | -0.006 | 0.004 | 0.14 | 0.072 | 0 | NA | NA |
| HLA-E | A-wave RV Inner Radial | 43 | 0.037 | 0.006 | 3.10E-11 | 0.056 | 0.008 | 2.70E-12 | 0.088 | 0.017 | 1.40E-05 | -0.014 | 0.005 | 0.0045 | NA | 1 | NA | NA |
| ITIH1 | S-wave RV Inner Circumferential | 12 | -0.132 | 0.02 | 4.10E-11 | -0.138 | 0.029 | 2.10E-06 | -0.026 | 0.247 | 0.92 | -0.011 | 0.028 | 0.69 | 0.74 | 0 | NA | NA |
| ITIH1 | S-wave RV Inner Total | 12 | -0.131 | 0.02 | 6.80E-11 | -0.138 | 0.029 | 1.40E-06 | 0.152 | 0.309 | 0.64 | -0.031 | 0.035 | 0.39 | 0.21 | 0 | NA | NA |
| PKLR | A-wave LV Inner Circumferential | 13 | 0.108 | 0.017 | 1.50E-10 | 0.123 | 0.025 | 1.20E-06 | 0.091 | 0.043 | 0.062 | 0.002 | 0.006 | 0.79 | 0.48 | 0 | NA | NA |
| RABGAP1L | L-wave RV Inner Total | 15 | -0.169 | 0.027 | 1.70E-10 | -0.179 | 0.035 | 2.40E-07 | -0.062 | 0.097 | 0.53 | -0.008 | 0.007 | 0.27 | 1 | 0 | NA | NA |
| AGRP | S-wave LV Inner Radial | 13 | -0.258 | 0.041 | 3.90E-10 | -0.253 | 0.067 | 0.00016 | -0.37 | 0.185 | 0.074 | 0.008 | 0.016 | 0.63 | 0.06 | 0 | NA | NA |
| ITGAV | L-wave RV Inner Radial | 37 | -0.085 | 0.014 | 4.20E-10 | -0.077 | 0.026 | 0.0027 | -0.031 | 0.048 | 0.52 | -0.008 | 0.006 | 0.19 | 0.03 | 1 | -0.107 | 3.80E-06 |
| MAP4K5 | E-wave RV Inner Total | 13 | 0.096 | 0.015 | 4.50E-10 | 0.091 | 0.023 | 9.70E-05 | 0.09 | 0.038 | 0.041 | 0 | 0.008 | 0.96 | 0.43 | 0 | NA | NA |
| PTH1R | S-wave LV Inner Total | 9 | 0.193 | 0.031 | 5.60E-10 | 0.209 | 0.039 | 7.00E-08 | 0.316 | 0.3 | 0.33 | -0.014 | 0.034 | 0.69 | 0.4 | 0 | NA | NA |
| RABGAP1L | E-wave LV Inner Total | 15 | -0.162 | 0.026 | 7.00E-10 | -0.18 | 0.033 | 4.60E-08 | -0.034 | 0.096 | 0.72 | -0.009 | 0.007 | 0.19 | 0.93 | 0 | NA | NA |
| RNF5 | A-wave RV Inner Circumferential | 6 | -0.42 | 0.069 | 9.70E-10 | -0.41 | 0.102 | 5.50E-05 | -0.688 | 0.604 | 0.34 | 0.013 | 0.028 | 0.66 | 0.99 | 0 | NA | NA |
| PTH1R | S-wave RV Inner Total | 9 | 0.19 | 0.031 | 1.10E-09 | 0.218 | 0.042 | 2.00E-07 | -0.008 | 0.32 | 0.98 | 0.022 | 0.036 | 0.55 | 0.28 | 0 | NA | NA |
| CRYZL1 | S-wave LV Myocardium Total | 9 | 0.156 | 0.026 | 1.10E-09 | 0.18 | 0.035 | 1.70E-07 | 0.234 | 0.04 | 0.00065 | -0.015 | 0.006 | 0.04 | 0.22 | 0 | NA | NA |
| PTPRC | S-wave RV Myocardial Free Wall Total | 10 | -0.087 | 0.015 | 2.00E-09 | -0.091 | 0.017 | 8.80E-08 | -0.105 | 0.018 | 0.001 | 0.008 | 0.005 | 0.13 | 0.84 | 0 | NA | NA |
| MAP4K5 | E-wave RV Inner Radial | 13 | 0.091 | 0.015 | 3.70E-09 | 0.082 | 0.022 | 0.00027 | 0.075 | 0.034 | 0.056 | 0.002 | 0.008 | 0.78 | 0.84 | 0 | NA | NA |
| PRKAR2A | A-wave RV Inner Circumferential | 6 | 0.305 | 0.052 | 4.10E-09 | 0.377 | 0.078 | 1.50E-06 | 1.606 | 0.755 | 0.12 | -0.067 | 0.038 | 0.17 | NA | 1 | 0.395 | 0.00092 |
| LTB | A-wave RV Inner Circumferential | 32 | 0.07 | 0.012 | 4.10E-09 | 0.075 | 0.02 | 0.00025 | 0.072 | 0.048 | 0.14 | -0.002 | 0.007 | 0.78 | 0.012 | 1 | 0.077 | 5.70E-05 |
| PRKAR2A | E-wave RV Inner Total | 6 | 0.303 | 0.052 | 4.80E-09 | 0.368 | 0.079 | 3.20E-06 | 1.528 | 0.661 | 0.1 | -0.063 | 0.033 | 0.15 | 0.001 | 1 | 0.386 | 0.00035 |
| RABGAP1L | L-wave RV Inner Circumferential | 15 | -0.155 | 0.027 | 5.30E-09 | -0.147 | 0.034 | 1.80E-05 | 0.032 | 0.097 | 0.75 | -0.014 | 0.007 | 0.065 | 0.97 | 0 | NA | NA |
| AOC1 | A-wave LV Myocardium Total | 9 | 0.095 | 0.016 | 5.60E-09 | 0.068 | 0.02 | 0.00056 | 0.047 | 0.025 | 0.11 | 0.009 | 0.004 | 0.086 | 0.42 | 0 | NA | NA |
| ASPN | A-wave RV Inner Total | 43 | -0.054 | 0.009 | 6.70E-09 | -0.062 | 0.014 | 8.00E-06 | -0.079 | 0.029 | 0.011 | 0.005 | 0.006 | 0.41 | 0.58 | 0 | NA | NA |
| PTH1R | S-wave LV Inner Radial | 9 | 0.18 | 0.031 | 7.80E-09 | 0.195 | 0.039 | 4.70E-07 | 0.301 | 0.25 | 0.27 | -0.014 | 0.028 | 0.64 | 0.62 | 0 | NA | NA |
| FBN2 | A-wave LV Inner Radial | 16 | 0.057 | 0.01 | 1.20E-08 | 0.057 | 0.013 | 1.60E-05 | 0.061 | 0.02 | 0.0098 | -0.001 | 0.005 | 0.83 | 0.24 | 0 | NA | NA |
| PRKAR2A | A-wave LV Inner Total | 6 | 0.295 | 0.052 | 1.20E-08 | 0.378 | 0.076 | 7.70E-07 | 1.759 | 0.724 | 0.093 | -0.075 | 0.036 | 0.13 | NA | 1 | 0.39 | 0.00013 |
| MAP4K5 | E-wave RV Inner Circumferential | 13 | 0.087 | 0.015 | 1.50E-08 | 0.086 | 0.022 | 0.00013 | 0.083 | 0.036 | 0.046 | -0.001 | 0.008 | 0.9 | 0.52 | 0 | NA | NA |
| HLA-E | A-wave RV Inner Circumferential | 43 | 0.032 | 0.006 | 1.50E-08 | 0.052 | 0.008 | 3.00E-10 | 0.08 | 0.018 | 0.00011 | -0.013 | 0.005 | 0.0084 | NA | 1 | 0.035 | 8.90E-05 |
| RNF5 | A-wave RV Inner Radial | 6 | -0.386 | 0.069 | 1.80E-08 | -0.382 | 0.097 | 8.40E-05 | -0.645 | 0.603 | 0.36 | 0.013 | 0.028 | 0.68 | 1 | 0 | NA | NA |
| PRKAR2A | A-wave RV Myocardial Free Wall Total | 6 | 0.29 | 0.052 | 2.10E-08 | 0.347 | 0.079 | 1.20E-05 | 1.351 | 0.637 | 0.12 | -0.054 | 0.032 | 0.19 | 0.008 | 1 | 0.37 | 0.00091 |
| PRKAR2A | S-wave LV Inner Radial | 6 | 0.287 | 0.052 | 2.70E-08 | 0.352 | 0.078 | 6.50E-06 | 1.411 | 0.526 | 0.075 | -0.058 | 0.026 | 0.12 | 0.008 | 1 | 0.357 | 2.40E-05 |
| PTH1R | S-wave RV Inner Radial | 9 | 0.173 | 0.031 | 2.90E-08 | 0.198 | 0.041 | 1.30E-06 | -0.023 | 0.29 | 0.94 | 0.022 | 0.033 | 0.52 | 0.37 | 0 | NA | NA |
| DPEP2 | S-wave RV Inner Radial | 26 | -0.066 | 0.012 | 3.10E-08 | -0.087 | 0.018 | 2.50E-06 | -0.095 | 0.027 | 0.0022 | 0.005 | 0.005 | 0.33 | 0.92 | 0 | NA | NA |
| ITIH1 | S-wave RV Inner Radial | 12 | -0.111 | 0.02 | 3.20E-08 | -0.124 | 0.029 | 1.40E-05 | 0.25 | 0.328 | 0.47 | -0.04 | 0.037 | 0.3 | 0.11 | 0 | NA | NA |
| PRKAR2A | A-wave RV Inner Total | 6 | 0.284 | 0.052 | 4.00E-08 | 0.355 | 0.078 | 5.60E-06 | 1.503 | 0.671 | 0.11 | -0.063 | 0.033 | 0.16 | 0.002 | 1 | 0.366 | 0.00052 |
| RNF5 | A-wave RV Inner Total | 6 | -0.373 | 0.069 | 5.60E-08 | -0.376 | 0.099 | 0.00016 | -0.711 | 0.604 | 0.32 | 0.017 | 0.028 | 0.59 | 0.98 | 0 | NA | NA |
| HLA-E | A-wave RV Inner Total | 43 | 0.03 | 0.006 | 6.50E-08 | 0.052 | 0.009 | 3.40E-09 | 0.083 | 0.017 | 2.80E-05 | -0.015 | 0.004 | 0.0025 | 0.001 | 3 | 0.039 | 1.10E-06 |
| APCS | A-wave RV Myocardial Free Wall Total | 11 | 0.097 | 0.018 | 8.30E-08 | 0.114 | 0.023 | 9.90E-07 | 0.105 | 0.026 | 0.0051 | -0.003 | 0.005 | 0.63 | 0.44 | 0 | NA | NA |
| CTSB | A-wave RV Inner Circumferential | 27 | -0.059 | 0.011 | 1.20E-07 | -0.054 | 0.02 | 0.0082 | -0.002 | 0.026 | 0.95 | -0.013 | 0.006 | 0.038 | 0.032 | 1 | -0.069 | 9.20E-05 |
| TNFAIP8L2 | E-wave RV Myocardial Free Wall Total | 5 | -0.307 | 0.058 | 1.20E-07 | -0.365 | 0.082 | 7.70E-06 | 1.143 | 0.441 | 0.081 | -0.132 | 0.04 | 0.045 | 0.061 | 0 | NA | NA |
| CLEC10A | A-wave LV Myocardium Total | 20 | -0.046 | 0.009 | 1.20E-07 | -0.044 | 0.011 | 9.50E-05 | -0.045 | 0.019 | 0.034 | -0.001 | 0.006 | 0.86 | 0.4 | 0 | NA | NA |
| PTH1R | S-wave LV Inner Circumferential | 9 | 0.164 | 0.031 | 1.20E-07 | 0.183 | 0.041 | 7.70E-06 | 0.32 | 0.346 | 0.39 | -0.018 | 0.039 | 0.66 | 0.19 | 0 | NA | NA |
| CTSB | A-wave RV Inner Total | 27 | -0.059 | 0.011 | 1.30E-07 | -0.028 | 0.021 | 0.18 | -0.004 | 0.028 | 0.88 | -0.012 | 0.006 | 0.056 | 0.008 | 2 | -0.057 | 0.00089 |
| RNF5 | S-wave RV Inner Radial | 6 | -0.362 | 0.069 | 1.30E-07 | -0.345 | 0.107 | 0.0012 | -0.47 | 0.604 | 0.49 | 0.005 | 0.028 | 0.86 | 1 | 0 | NA | NA |
| PTPRC | S-wave LV Myocardium Total | 10 | 0.075 | 0.014 | 1.80E-07 | 0.071 | 0.017 | 3.50E-05 | 0.081 | 0.018 | 0.0039 | -0.002 | 0.004 | 0.62 | 0.89 | 0 | NA | NA |
| FBN2 | A-wave RV Inner Radial | 16 | 0.052 | 0.01 | 2.20E-07 | 0.05 | 0.014 | 0.00034 | 0.066 | 0.018 | 0.0042 | -0.003 | 0.004 | 0.47 | 0.3 | 0 | NA | NA |
| AGRP | S-wave LV Inner Total | 13 | -0.213 | 0.041 | 2.30E-07 | -0.227 | 0.063 | 0.00033 | -0.269 | 0.148 | 0.099 | 0.003 | 0.013 | 0.81 | 0.33 | 0 | NA | NA |
| RECK | S-wave LV Inner Radial | 16 | 0.079 | 0.015 | 2.50E-07 | 0.058 | 0.023 | 0.012 | 0.071 | 0.028 | 0.028 | 0.004 | 0.006 | 0.57 | 0.25 | 0 | NA | NA |
| SCG3 | E-wave RV Inner Total | 78 | 0.031 | 0.006 | 2.50E-07 | 0.038 | 0.009 | 5.90E-05 | 0.055 | 0.013 | 5.80E-05 | -0.009 | 0.004 | 0.026 | 0.32 | 0 | NA | NA |
| FCER1A | S-wave LV Myocardium Total | 20 | -0.106 | 0.021 | 3.20E-07 | -0.118 | 0.029 | 4.20E-05 | -0.301 | 0.085 | 0.0026 | 0.028 | 0.012 | 0.032 | 0.43 | 0 | NA | NA |
| DPEP2 | S-wave RV Inner Total | 26 | -0.061 | 0.012 | 3.30E-07 | -0.076 | 0.017 | 5.70E-06 | -0.078 | 0.027 | 0.0093 | 0.002 | 0.005 | 0.62 | 0.89 | 0 | NA | NA |
| PTH1R | S-wave RV Inner Circumferential | 9 | 0.158 | 0.031 | 3.70E-07 | 0.172 | 0.041 | 2.60E-05 | 0.161 | 0.25 | 0.54 | 0 | 0.028 | 0.99 | 0.83 | 0 | NA | NA |

| | | | | | | | | | | | | | | | | | |
|---|---|---|---|---|---|---|---|---|---|---|---|---|---|---|---|---|---|
| PRKAR2A | A-wave RV Inner Radial | 6 | 0.262 | 0.052 | 3.90E-07 | 0.332 | 0.076 | 1.10E-05 | 1.339 | 0.515 | 0.08 | -0.055 | 0.026 | 0.12 | 0.017 | 1 | 0.328 | 7.30E-05 |
| C2 | S-wave LV Inner Total | 39 | 0.034 | 0.007 | 5.10E-07 | 0.034 | 0.011 | 0.0023 | 0.076 | 0.017 | 9.00E-05 | -0.01 | 0.004 | 0.015 | 0.4 | 0 | NA | NA |
| LTBP3 | L-wave RV Inner Radial | 37 | 0.05 | 0.01 | 6.00E-07 | 0.051 | 0.015 | 0.00048 | 0.066 | 0.021 | 0.0032 | -0.005 | 0.004 | 0.29 | 0.83 | 0 | NA | NA |
| RBP7 | A-wave RV Inner Circumferential | 24 | 0.053 | 0.011 | 6.20E-07 | 0.064 | 0.016 | 4.20E-05 | 0.056 | 0.023 | 0.028 | 0 | 0.006 | 0.97 | 0.93 | 0 | NA | NA |
| PKLR | A-wave LV Inner Total | 13 | 0.084 | 0.017 | 7.10E-07 | 0.096 | 0.024 | 6.90E-05 | 0.071 | 0.043 | 0.13 | 0.001 | 0.006 | 0.88 | 0.68 | 0 | NA | NA |
| HLA-E | S-wave RV Inner Total | 43 | 0.028 | 0.006 | 8.20E-07 | 0.037 | 0.008 | 1.10E-06 | 0.05 | 0.014 | 0.00082 | -0.006 | 0.004 | 0.097 | 0.84 | 0 | NA | NA |
| GSN | A-wave LV Inner Radial | 17 | 0.103 | 0.021 | 8.60E-07 | 0.131 | 0.028 | 4.00E-06 | 0.146 | 0.033 | 0.00066 | -0.006 | 0.004 | 0.19 | 0.63 | 0 | NA | NA |
| WWP2 | A-wave LV Inner Radial | 6 | -0.097 | 0.02 | 8.60E-07 | -0.127 | 0.023 | 1.90E-08 | -0.252 | 0.032 | 0.0043 | 0.038 | 0.006 | 0.0095 | NA | 1 | -0.118 | 0.034 |
| WWP2 | A-wave LV Inner Total | 6 | -0.096 | 0.02 | 9.00E-07 | -0.127 | 0.023 | 2.70E-08 | -0.254 | 0.032 | 0.0041 | 0.038 | 0.006 | 0.0089 | NA | 2 | -0.127 | 0.0026 |
| HLA-A | S-wave RV Inner Radial | 87 | 0.011 | 0.002 | 9.10E-07 | 0.015 | 0.004 | 0.00026 | 0.025 | 0.007 | 0.00087 | -0.009 | 0.004 | 0.031 | NA | 1 | NA | NA |
| RABGAP1L | L-wave RV Inner Radial | 15 | -0.13 | 0.026 | 9.20E-07 | -0.121 | 0.036 | 0.00063 | 0.013 | 0.096 | 0.89 | -0.011 | 0.007 | 0.15 | 0.98 | 0 | NA | NA |
| LGALS3 | A-wave RV Myocardial Free Wall Total | 33 | 0.023 | 0.005 | 9.30E-07 | 0.021 | 0.007 | 0.0038 | 0.022 | 0.011 | 0.043 | 0.002 | 0.005 | 0.74 | 0.64 | 0 | NA | NA |
| MAP4K5 | S-wave RV Inner Circumferential | 13 | 0.075 | 0.015 | 1.00E-06 | 0.072 | 0.023 | 0.0014 | 0.05 | 0.034 | 0.18 | 0.005 | 0.008 | 0.51 | 0.91 | 0 | NA | NA |
| HLA-A | A-wave RV Inner Radial | 87 | 0.011 | 0.002 | 1.10E-06 | 0.022 | 0.004 | 3.20E-09 | 0.023 | 0.008 | 0.003 | -0.008 | 0.004 | 0.061 | NA | 1 | 0.01 | 0.0025 |
| IST1 | A-wave RV Myocardial Free Wall Total | 7 | 0.181 | 0.037 | 1.10E-06 | 0.201 | 0.047 | 2.10E-05 | 0.219 | 0.052 | 0.0086 | -0.005 | 0.005 | 0.35 | 0.92 | 0 | NA | NA |
| HYAL1 | A-wave LV Myocardium Total | 19 | -0.055 | 0.011 | 1.30E-06 | -0.058 | 0.015 | 7.20E-05 | -0.051 | 0.024 | 0.045 | -0.001 | 0.004 | 0.84 | 0.007 | 2 | -0.052 | 0.00082 |
| CES2 | A-wave LV Myocardium Total | 6 | -0.197 | 0.041 | 1.30E-06 | -0.202 | 0.052 | 9.90E-05 | -0.233 | 0.136 | 0.16 | 0.005 | 0.019 | 0.8 | 0.66 | 0 | NA | NA |
| SEMA3F | S-wave LV Inner Radial | 22 | 0.075 | 0.016 | 1.30E-06 | 0.072 | 0.024 | 0.0026 | 0.151 | 0.046 | 0.0041 | -0.011 | 0.006 | 0.1 | 0.68 | 0 | NA | NA |
| RBP7 | S-wave LV Inner Circumferential | 24 | 0.051 | 0.011 | 1.40E-06 | 0.058 | 0.017 | 0.00049 | 0.029 | 0.023 | 0.23 | 0.007 | 0.006 | 0.26 | 0.48 | 0 | NA | NA |
| ROBO1 | E-wave LV Myocardium Total | 45 | 0.076 | 0.016 | 1.40E-06 | 0.1 | 0.029 | 0.00051 | 0.109 | 0.038 | 0.0071 | -0.004 | 0.005 | 0.43 | 0.16 | 0 | NA | NA |
| PRKAR2A | A-wave LV Myocardium Total | 6 | 0.249 | 0.052 | 1.60E-06 | 0.265 | 0.078 | 0.00068 | 0.816 | 0.499 | 0.2 | -0.029 | 0.025 | 0.33 | 0.16 | 0 | NA | NA |
| ACRV1 | A-wave LV Myocardium Total | 8 | -0.27 | 0.057 | 1.80E-06 | -0.307 | 0.073 | 2.80E-05 | -0.781 | 0.176 | 0.0044 | 0.029 | 0.01 | 0.022 | 0.15 | 0 | NA | NA |
| HCG22 | S-wave LV Inner Circumferential | 74 | -0.034 | 0.007 | 1.80E-06 | -0.03 | 0.011 | 0.006 | -0.001 | 0.018 | 0.93 | -0.007 | 0.003 | 0.049 | 0.36 | 0 | NA | NA |

**Supplemental Table 11. Mendelian randomization sensitivity analyses for 81 protein-phenotype associations.** Two-sample MR results for all protein-phenotype associations reaching IVW significance at Bonferroni-corrected threshold ($p < 1.86\times10^{-6}$) and passing MR-PRESSO outlier correction. Genetic instruments were cis-pQTLs from the UK Biobank Pharma Proteomics Project (UKB-PPP). Outcome GWAS were performed in 65,162-65,496 participants after excluding 9,480 individuals with overlapping optical flow and UKB-PPP proteomics data. IVW: inverse variance-weighted estimate using fixed-effects model. Weighted median: robust to up to 50% invalid instruments. MR-Egger: allows for directional pleiotropy; Egger intercept $p > 0.05$ indicates absence of directional pleiotropy. MR-PRESSO: global $p < 0.05$ indicates horizontal pleiotropy; corrected estimates shown where outlier removal resolved pleiotropy; β values are per unit NPX (log2-normalized protein expression). cis-pQTL = cis-protein quantitative trait locus; IVW = inverse variance weighted; LV = left ventricular; MR = Mendelian randomization; RV = right ventricular; UKB-PPP = UK Biobank Pharma Proteomics Project.

| Protein | Phenotype | GTEx Tissue | N Shared SNPs | PP.H3 | PP.H4 |
|---|---|---|---|---|---|
| RABGAP1L | E-wave RV Inner Circumferential | Artery Aorta | 392 | 0.091 | 0.428 |
| RABGAP1L | E-wave RV Inner Circumferential | Heart Atrial Appendage | 628 | 0.192 | 0.089 |
| RABGAP1L | E-wave RV Inner Circumferential | Heart Left Ventricle | 36 | 0 | 0.011 |
| RABGAP1L | E-wave RV Inner Total | Artery Aorta | 392 | 0.035 | 0.201 |
| RABGAP1L | E-wave RV Inner Total | Heart Atrial Appendage | 628 | 0.108 | 0.116 |
| RABGAP1L | E-wave RV Inner Total | Heart Left Ventricle | 36 | 0.001 | 0.019 |
| RABGAP1L | E-wave RV Inner Radial | Artery Aorta | 392 | 0.034 | 0.238 |
| RABGAP1L | E-wave RV Inner Radial | Heart Atrial Appendage | 628 | 0.046 | 0.047 |
| RABGAP1L | E-wave RV Inner Radial | Heart Left Ventricle | 36 | 0 | 0.005 |
| RABGAP1L | E-wave LV Inner Radial | Artery Aorta | 392 | 0.024 | 0.115 |
| RABGAP1L | E-wave LV Inner Radial | Heart Atrial Appendage | 628 | 0.311 | 0.032 |
| RABGAP1L | E-wave LV Inner Radial | Heart Left Ventricle | 36 | 0.001 | 0.019 |
| RABGAP1L | L-wave RV Inner Total | Artery Aorta | 392 | 0.006 | 0.024 |
| RABGAP1L | L-wave RV Inner Total | Heart Atrial Appendage | 628 | 0.011 | 0.019 |
| RABGAP1L | L-wave RV Inner Total | Heart Left Ventricle | 36 | 0 | 0.004 |
| RABGAP1L | E-wave LV Inner Total | Heart Atrial Appendage | 628 | 0.022 | 0.025 |
| RABGAP1L | E-wave LV Inner Total | Artery Aorta | 392 | 0.007 | 0.016 |
| RABGAP1L | E-wave LV Inner Total | Heart Left Ventricle | 36 | 0 | 0.011 |
| RABGAP1L | L-wave RV Inner Circumferential | Heart Atrial Appendage | 628 | 0.012 | 0.055 |
| RABGAP1L | L-wave RV Inner Circumferential | Artery Aorta | 392 | 0.011 | 0.011 |
| RABGAP1L | L-wave RV Inner Circumferential | Heart Left Ventricle | 36 | 0 | 0.008 |
| RABGAP1L | L-wave RV Inner Radial | Heart Atrial Appendage | 628 | 0.007 | 0.031 |
| RABGAP1L | L-wave RV Inner Radial | Artery Aorta | 392 | 0.006 | 0.008 |
| RABGAP1L | L-wave RV Inner Radial | Heart Left Ventricle | 36 | 0 | 0.005 |
| HCG22 | A-wave RV Inner Circumferential | Artery Aorta | 438 | 0.087 | 0.014 |
| HCG22 | A-wave RV Inner Circumferential | Heart Atrial Appendage | 3539 | 0.188 | 0.009 |
| HCG22 | A-wave RV Inner Circumferential | Heart Left Ventricle | 1681 | 0.118 | 0.009 |
| HCG22 | A-wave RV Inner Circumferential | Artery Coronary | 699 | 0.085 | 0.009 |
| HCG22 | A-wave RV Inner Total | Heart Left Ventricle | 1681 | 0.052 | 0.013 |
| HCG22 | A-wave RV Inner Total | Artery Aorta | 438 | 0.028 | 0.013 |
| HCG22 | A-wave RV Inner Total | Artery Coronary | 699 | 0.03 | 0.013 |

| | | | | | |
|---|---|---|---|---|---|
| HCG22 | A-wave RV Inner Total | Heart Atrial Appendage | 3539 | 0.113 | 0.011 |
| HCG22 | A-wave RV Inner Radial | Artery Aorta | 438 | 0.038 | 0.012 |
| HCG22 | A-wave RV Inner Radial | Artery Coronary | 699 | 0.041 | 0.01 |
| HCG22 | A-wave RV Inner Radial | Heart Atrial Appendage | 3539 | 0.17 | 0.009 |
| HCG22 | A-wave RV Inner Radial | Heart Left Ventricle | 1681 | 0.106 | 0.009 |
| HCG22 | S-wave LV Inner Circumferential | Heart Atrial Appendage | 3539 | 0.051 | 0.004 |
| HCG22 | S-wave LV Inner Circumferential | Heart Left Ventricle | 1681 | 0.014 | 0.004 |
| HCG22 | S-wave LV Inner Circumferential | Artery Aorta | 438 | 0.002 | 0.004 |
| HCG22 | S-wave LV Inner Circumferential | Artery Coronary | 699 | 0.004 | 0.004 |
| HLA-E | S-wave RV Inner Radial | Heart Atrial Appendage | 335 | 0.062 | 0.039 |
| HLA-E | S-wave RV Inner Radial | Artery Coronary | 193 | 0.005 | 0.032 |
| HLA-E | S-wave RV Inner Radial | Heart Left Ventricle | 88 | 0.002 | 0.028 |
| HLA-E | A-wave RV Inner Radial | Heart Left Ventricle | 88 | 0 | 0.005 |
| HLA-E | A-wave RV Inner Radial | Heart Atrial Appendage | 335 | 0.026 | 0.004 |
| HLA-E | A-wave RV Inner Radial | Artery Coronary | 193 | 0.001 | 0.004 |
| HLA-E | A-wave RV Inner Circumferential | Heart Atrial Appendage | 335 | 0.034 | 0.004 |
| HLA-E | A-wave RV Inner Circumferential | Heart Left Ventricle | 88 | 0 | 0.004 |
| HLA-E | A-wave RV Inner Circumferential | Artery Coronary | 193 | 0.001 | 0.004 |
| HLA-E | A-wave RV Inner Total | Heart Atrial Appendage | 335 | 0.015 | 0.004 |
| HLA-E | A-wave RV Inner Total | Heart Left Ventricle | 88 | 0 | 0.004 |
| HLA-E | A-wave RV Inner Total | Artery Coronary | 193 | 0.001 | 0.004 |
| HLA-E | S-wave RV Inner Total | Heart Atrial Appendage | 335 | 0.007 | 0.01 |
| HLA-E | S-wave RV Inner Total | Artery Coronary | 193 | 0.001 | 0.008 |
| HLA-E | S-wave RV Inner Total | Heart Left Ventricle | 88 | 0.001 | 0.007 |
| ITIH1 | S-wave RV Inner Circumferential | Heart Atrial Appendage | 148 | 0.072 | 0.481 |
| ITIH1 | S-wave RV Inner Circumferential | Artery Coronary | 25 | 0.008 | 0.401 |
| ITIH1 | S-wave RV Inner Total | Artery Coronary | 25 | 0.011 | 0.555 |
| ITIH1 | S-wave RV Inner Total | Heart Atrial Appendage | 148 | 0.072 | 0.533 |
| ITIH1 | S-wave RV Inner Radial | Heart Atrial Appendage | 148 | 0.027 | 0.218 |
| ITIH1 | S-wave RV Inner Radial | Artery Coronary | 25 | 0.004 | 0.178 |
| ITGAV | L-wave RV Inner Radial | Heart Atrial Appendage | 134 | 0.005 | 0.017 |

| | | | | | |
|---|---|---|---|---|---|
| ITGAV | L-wave RV Inner Radial | Heart Left Ventricle | 97 | 0.001 | 0.01 |
| ITGAV | L-wave RV Inner Radial | Artery Aorta | 20 | 0 | 0.003 |
| MAP4K5 | E-wave RV Inner Total | Artery Aorta | 21 | 0 | 0.015 |
| MAP4K5 | E-wave RV Inner Total | Heart Atrial Appendage | 33 | 0 | 0.009 |
| MAP4K5 | E-wave RV Inner Radial | Artery Aorta | 21 | 0 | 0.01 |
| MAP4K5 | E-wave RV Inner Radial | Heart Atrial Appendage | 33 | 0 | 0.008 |
| MAP4K5 | E-wave RV Inner Circumferential | Artery Aorta | 21 | 0 | 0.008 |
| MAP4K5 | E-wave RV Inner Circumferential | Heart Atrial Appendage | 33 | 0 | 0.006 |
| MAP4K5 | S-wave RV Inner Circumferential | Heart Atrial Appendage | 33 | 0 | 0.006 |
| MAP4K5 | S-wave RV Inner Circumferential | Artery Aorta | 21 | 0 | 0.004 |
| PTH1R | S-wave LV Inner Total | Heart Atrial Appendage | 44 | 0 | 0.005 |
| PTH1R | S-wave RV Inner Total | Heart Atrial Appendage | 44 | 0.001 | 0.014 |
| PTH1R | S-wave LV Inner Radial | Heart Atrial Appendage | 44 | 0 | 0.006 |
| PTH1R | S-wave RV Inner Radial | Heart Atrial Appendage | 44 | 0.001 | 0.011 |
| PTH1R | S-wave LV Inner Circumferential | Heart Atrial Appendage | 44 | 0 | 0.004 |
| PTH1R | S-wave RV Inner Circumferential | Heart Atrial Appendage | 44 | 0.001 | 0.007 |
| RNF5 | A-wave RV Inner Circumferential | Artery Aorta | 452 | 0.119 | 0.452 |
| RNF5 | A-wave RV Inner Circumferential | Heart Atrial Appendage | 602 | 0.096 | 0.449 |
| RNF5 | A-wave RV Inner Circumferential | Heart Left Ventricle | 839 | 0.132 | 0.287 |
| RNF5 | A-wave RV Inner Radial | Heart Atrial Appendage | 602 | 0.025 | 0.11 |
| RNF5 | A-wave RV Inner Radial | Artery Aorta | 452 | 0.039 | 0.109 |
| RNF5 | A-wave RV Inner Radial | Heart Left Ventricle | 839 | 0.036 | 0.08 |
| RNF5 | A-wave RV Inner Total | Heart Atrial Appendage | 602 | 0.04 | 0.247 |
| RNF5 | A-wave RV Inner Total | Artery Aorta | 452 | 0.053 | 0.227 |
| RNF5 | A-wave RV Inner Total | Heart Left Ventricle | 839 | 0.052 | 0.184 |
| RNF5 | S-wave RV Inner Radial | Heart Left Ventricle | 839 | 0.028 | 0.271 |
| RNF5 | S-wave RV Inner Radial | Heart Atrial Appendage | 602 | 0.024 | 0.235 |
| RNF5 | S-wave RV Inner Radial | Artery Aorta | 452 | 0.021 | 0.2 |
| PRKAR2A | A-wave RV Inner Circumferential | Heart Atrial Appendage | 166 | 0.088 | 0.494 |
| PRKAR2A | A-wave RV Inner Circumferential | Artery Aorta | 20 | 0 | 0.001 |
| PRKAR2A | E-wave RV Inner Total | Heart Atrial Appendage | 166 | 0.074 | 0.429 |

| | | | | | |
|---|---|---|---|---|---|
| PRKAR2A | E-wave RV Inner Total | Artery Aorta | 20 | 0 | 0.001 |
| PRKAR2A | A-wave LV Inner Total | Heart Atrial Appendage | 166 | 0.089 | 0.495 |
| PRKAR2A | A-wave LV Inner Total | Artery Aorta | 20 | 0 | 0.001 |
| PRKAR2A | A-wave RV Myocardial Free Wall Total | Heart Atrial Appendage | 166 | 0.062 | 0.354 |
| PRKAR2A | A-wave RV Myocardial Free Wall Total | Artery Aorta | 20 | 0 | 0.001 |
| PRKAR2A | S-wave LV Inner Radial | Heart Atrial Appendage | 166 | 0.029 | 0.186 |
| PRKAR2A | S-wave LV Inner Radial | Artery Aorta | 20 | 0 | 0.001 |
| PRKAR2A | A-wave RV Inner Total | Heart Atrial Appendage | 166 | 0.047 | 0.268 |
| PRKAR2A | A-wave RV Inner Total | Artery Aorta | 20 | 0 | 0.001 |
| PRKAR2A | A-wave RV Inner Radial | Heart Atrial Appendage | 166 | 0.016 | 0.103 |
| PRKAR2A | A-wave RV Inner Radial | Artery Aorta | 20 | 0 | 0.001 |
| PRKAR2A | A-wave LV Myocardium Total | Heart Atrial Appendage | 166 | 0.029 | 0.16 |
| PRKAR2A | A-wave LV Myocardium Total | Artery Aorta | 20 | 0 | 0.001 |
| AOC1 | A-wave LV Myocardium Total | Heart Atrial Appendage | 193 | 0.003 | 0.005 |
| AOC1 | A-wave LV Myocardium Total | Heart Left Ventricle | 138 | 0.001 | 0.005 |
| ASPN | A-wave RV Inner Total | Heart Atrial Appendage | 40 | 0.001 | 0.015 |
| ASPN | A-wave RV Inner Total | Artery Aorta | 244 | 0.003 | 0.013 |
| FBN2 | A-wave LV Inner Radial | Artery Aorta | 580 | 0.068 | 0.421 |
| FBN2 | A-wave LV Inner Radial | Artery Coronary | 531 | 0.077 | 0.343 |
| FBN2 | A-wave LV Inner Radial | Heart Atrial Appendage | 814 | 0.122 | 0.014 |
| FBN2 | A-wave LV Inner Radial | Heart Left Ventricle | 493 | 0.004 | 0.01 |
| FBN2 | A-wave RV Inner Radial | Artery Coronary | 531 | 0.074 | 0.33 |
| FBN2 | A-wave RV Inner Radial | Artery Aorta | 580 | 0.078 | 0.308 |
| FBN2 | A-wave RV Inner Radial | Heart Atrial Appendage | 814 | 0.085 | 0.015 |
| FBN2 | A-wave RV Inner Radial | Heart Left Ventricle | 493 | 0.003 | 0.005 |
| DPEP2 | S-wave RV Inner Radial | Artery Aorta | 133 | 0.006 | 0.04 |
| DPEP2 | S-wave RV Inner Radial | Heart Atrial Appendage | 20 | 0 | 0.01 |
| DPEP2 | S-wave RV Inner Total | Artery Aorta | 133 | 0.004 | 0.028 |
| DPEP2 | S-wave RV Inner Total | Heart Atrial Appendage | 20 | 0 | 0.006 |
| CTSB | A-wave RV Inner Circumferential | Heart Left Ventricle | 78 | 0.002 | 0.02 |
| CTSB | A-wave RV Inner Circumferential | Artery Aorta | 81 | 0.003 | 0.02 |

| | | | | | |
|---|---|---|---|---|---|
| CTSB | A-wave RV Inner Circumferential | Heart Atrial Appendage | 90 | 0.006 | 0.019 |
| CTSB | A-wave RV Inner Circumferential | Artery Coronary | 28 | 0.001 | 0.006 |
| CTSB | A-wave RV Inner Total | Heart Left Ventricle | 78 | 0.08 | 0.057 |
| CTSB | A-wave RV Inner Total | Heart Atrial Appendage | 90 | 0.022 | 0.056 |
| CTSB | A-wave RV Inner Total | Artery Aorta | 81 | 0.081 | 0.056 |
| CTSB | A-wave RV Inner Total | Artery Coronary | 28 | 0.002 | 0.01 |
| FCER1A | S-wave LV Myocardium Total | Heart Left Ventricle | 20 | 0.056 | 0.43 |
| FCER1A | S-wave LV Myocardium Total | Heart Atrial Appendage | 39 | 0.082 | 0.171 |
| FCER1A | S-wave LV Myocardium Total | Artery Aorta | 55 | 0.149 | 0.106 |
| C2 | S-wave LV Inner Total | Artery Aorta | 82 | 0 | 0.005 |
| RBP7 | A-wave RV Inner Circumferential | Heart Atrial Appendage | 29 | 0 | 0.145 |
| RBP7 | S-wave LV Inner Circumferential | Heart Atrial Appendage | 29 | 0.003 | 0.059 |
| WWP2 | A-wave LV Inner Radial | Artery Coronary | 22 | 0.014 | 0.878 |
| WWP2 | A-wave LV Inner Radial | Artery Aorta | 263 | 0.28 | 0.029 |
| WWP2 | A-wave LV Inner Total | Artery Coronary | 22 | 0.019 | 0.893 |
| WWP2 | A-wave LV Inner Total | Artery Aorta | 263 | 0.404 | 0.034 |
| HLA-A | S-wave RV Inner Radial | Heart Left Ventricle | 2838 | 0.086 | 0.031 |
| HLA-A | S-wave RV Inner Radial | Artery Coronary | 1185 | 0.072 | 0.014 |
| HLA-A | S-wave RV Inner Radial | Heart Atrial Appendage | 2750 | 0.218 | 0.011 |
| HLA-A | S-wave RV Inner Radial | Artery Aorta | 4113 | 0.436 | 0.003 |
| HLA-A | A-wave RV Inner Radial | Heart Left Ventricle | 2838 | 0.058 | 0.007 |
| HLA-A | A-wave RV Inner Radial | Artery Coronary | 1185 | 0.035 | 0.007 |
| HLA-A | A-wave RV Inner Radial | Heart Atrial Appendage | 2750 | 0.206 | 0.003 |
| HLA-A | A-wave RV Inner Radial | Artery Aorta | 4113 | 0.263 | 0.003 |
| LGALS3 | A-wave RV Myocardial Free Wall Total | Artery Aorta | 547 | 0.055 | 0.281 |
| IST1 | A-wave RV Myocardial Free Wall Total | Heart Atrial Appendage | 79 | 0 | 0.004 |
| IST1 | A-wave RV Myocardial Free Wall Total | Heart Left Ventricle | 131 | 0.001 | 0.004 |
| HYAL1 | A-wave LV Myocardium Total | Heart Left Ventricle | 394 | 0.014 | 0.013 |
| HYAL1 | A-wave LV Myocardium Total | Heart Atrial Appendage | 628 | 0.067 | 0.006 |
| CES2 | A-wave LV Myocardium Total | Heart Left Ventricle | 68 | 0.011 | 0.154 |
| CES2 | A-wave LV Myocardium Total | Heart Atrial Appendage | 32 | 0.003 | 0.127 |

| | | | | | |
|---|---|---|---|---|---|
| SEMA3F | S-wave LV Inner Radial | Heart Atrial Appendage | 288 | 0.01 | 0.308 |
| SEMA3F | S-wave LV Inner Radial | Heart Left Ventricle | 215 | 0.009 | 0.17 |
| SEMA3F | S-wave LV Inner Radial | Artery Aorta | 466 | 0.021 | 0.02 |
| ROBO1 | E-wave LV Myocardium Total | Artery Aorta | 84 | 0.001 | 0.005 |
| ROBO1 | E-wave LV Myocardium Total | Heart Atrial Appendage | 66 | 0 | 0.004 |

**Supplemental Table 12. eQTL colocalization of MR-significant proteins with optical flow GWAS signals in cardiovascular tissues.** Bayesian colocalization between optical flow GWAS summary statistics and eQTL data from GTEx v10 for 25 of 38 MR-PRESSO-validated proteins with available eQTL data in at least one cardiovascular tissue. Tissues tested: Heart Atrial Appendage, Heart Left Ventricle, Artery Aorta, and Artery Coronary. eQTL = expression quantitative trait locus; GWAS = genome-wide association study; PP.H3 = posterior probability of independent causal variants; PP.H4 = posterior probability of shared causal variant.

| Supplemental Table 13A — Input proteins | | | |
|---|---|---|---|
| **Index** | **Gene symbol** | **STRING identifier** | **Note** |
| 1 | ACRV1 | 9606.ENSP00000432816 | |
| 2 | AGRP | 9606.ENSP00000290953 | |
| 3 | AOC1 | 9606.ENSP00000411613 | |
| 4 | APCS | 9606.ENSP00000255040 | |
| 5 | ASPN | 9606.ENSP00000364694 | |
| 6 | C2 | 9606.ENSP00000299367 | |
| 7 | CES2 | 9606.ENSP00000499140 | |
| 8 | CLEC10A | 9606.ENSP00000254868 | |
| 9 | CRYZL1 | 9606.ENSP00000370966 | |
| 10 | CTSB | 9606.ENSP00000345672 | |
| 11 | DPEP2 | 9606.ENSP00000458977 | |
| 12 | FBN2 | 9606.ENSP00000424571 | |
| 13 | FCER1A | 9606.ENSP00000357097 | |
| 14 | GSN | 9606.ENSP00000362924 | |
| 15 | HCG22 | | Excluded; non-coding RNA gene, not mapped in STRING |
| 16 | HLA-A | 9606.ENSP00000379873 | |
| 17 | HLA-E | 9606.ENSP00000365817 | |
| 18 | HYAL1 | 9606.ENSP00000266031 | |

| | | |
|---|---|---|
| 19 | IST1 | 9606.ENSP00000438399 |
| 20 | ITGAV | 9606.ENSP00000261023 |
| 21 | ITIH1 | 9606.ENSP00000273283 |
| 22 | LGALS3 | 9606.ENSP00000254301 |
| 23 | LTB | 9606.ENSP00000410481 |
| 24 | LTBP3 | 9606.ENSP00000301873 |
| 25 | MAP4K5 | 9606.ENSP00000013125 |
| 26 | PKLR | 9606.ENSP00000339933 |
| 27 | PRKAR2A | 9606.ENSP00000265563 |
| 28 | PTH1R | 9606.ENSP00000402723 |
| 29 | PTPRC | 9606.ENSP00000411355 |
| 30 | RABGAP1L | 9606.ENSP00000251507 |
| 31 | RBP7 | 9606.ENSP00000294435 |
| 32 | RECK | 9606.ENSP00000367202 |
| 33 | RNF5 | 9606.ENSP00000364235 |
| 34 | ROBO1 | 9606.ENSP00000420321 |
| 35 | SCG3 | 9606.ENSP00000220478 |
| 36 | SEMA3F | 9606.ENSP00000002829 |
| 37 | TNFAIP8L2 | 9606.ENSP00000357906 |
| 38 | WWP2 | 9606.ENSP00000352069 |

| **Supplemental Table 13B — PPI network summary statistics** | |
|---|---|
| **Metric** | **Value** |
| Number of nodes | 37 |
| Number of edges | 19 |
| Expected number of edges | 7 |
| PPI enrichment P-value | 1.18E-04 |

| **Supplemental Table 13C — GO Cellular Component enrichment** | | | | | |
|---|---|---|---|---|---|
| **Term ID** | **Description** | **Observed gene count** | **FDR** | **Strength** | **Proteins in term** |
| GO:0005615 | Extracellular space | 22 | 9.21E-06 | 0.56 | SEMA3F, LGALS3, APCS, ITGAV, PRKAR2A, HYAL1, ITIH1, AGRP, C2, LTBP3, PKLR, CTSB, WWP2, GSN, ASPN, HLA-E, HLA-A, LTB, PTPRC, AOC1, FBN2, IST1 |
| GO:0005576 | Extracellular region | 24 | 1.34E-05 | 0.49 | SEMA3F, SCG3, LGALS3, APCS, ITGAV, PRKAR2A, HYAL1, ITIH1, AGRP, C2, LTBP3, PKLR, CTSB, WWP2, GSN, ASPN, HLA-E, RECK, HLA-A, LTB, PTPRC, AOC1, FBN2, IST1 |

| | | | | | |
|---|---|---|---|---|---|
| GO:0070062 | Extracellular exosome | 17 | 3.97E-05 | 0.64 | LGALS3, APCS, ITGAV, PRKAR2A, HYAL1, ITIH1, C2, LTBP3, PKLR, CTSB, WWP2, GSN, HLA-E, HLA-A, PTPRC, AOC1, IST1 |
| GO:0031982 | Vesicle | 20 | 0.0015 | 0.43 | SCG3, RABGAP1L, LGALS3, APCS, ITGAV, PRKAR2A, HYAL1, ITIH1, C2, LTBP3, PKLR, CTSB, WWP2, GSN, HLA-E, HLA-A, PTPRC, AOC1, ACRV1, IST1 |
| GO:0062023 | Collagen-containing extracellular matrix | 7 | 0.0026 | 0.96 | LGALS3, APCS, ITIH1, LTBP3, CTSB, ASPN, FBN2 |
| GO:0030141 | Secretory granule | 9 | 0.0062 | 0.74 | SCG3, LGALS3, ITGAV, CTSB, GSN, PTPRC, AOC1, ACRV1, IST1 |
| GO:0009986 | Cell surface | 9 | 0.0068 | 0.73 | LGALS3, CLEC10A, ITGAV, CTSB, FCER1A, HLA-E, HLA-A, PTPRC, ROBO1 |
| GO:0071944 | Cell periphery | 23 | 0.0129 | 0.31 | SEMA3F, LGALS3, CLEC10A, APCS, ITGAV, PRKAR2A, ITIH1, C2, LTBP3, CTSB, FCER1A, GSN, RNF5, ASPN, HLA-E, RECK, HLA-A, PTH1R, LTB, PTPRC, AOC1, ROBO1, FBN2 |
| GO:0042612 | MHC class I protein complex | 2 | 0.0198 | 2.12 | HLA-E, HLA-A |
| GO:0045335 | Phagocytic vesicle | 4 | 0.0198 | 1.19 | ITGAV, GSN, HLA-E, HLA-A |

| | | | | | |
|---|---|---|---|---|---|
| GO:0031410 | Cytoplasmic vesicle | 13 | 0.0457 | 0.45 | SCG3, RABGAP1L, LGALS3, ITGAV, HYAL1, CTSB, GSN, HLA-E, HLA-A, PTPRC, AOC1, ACRV1, IST1 |
| **Supplemental Table 13D — Reactome pathway enrichment** | | | | | |
| **Term ID** | **Description** | **Observed gene count** | **FDR** | **Strength** | **Proteins in term** |
| HSA-168249 | Innate Immune System | 12 | 0.00061 | 0.79 | LGALS3, CLEC10A, ITGAV, C2, CTSB, FCER1A, GSN, HLA-E, HLA-A, PTPRC, AOC1, IST1 |
| HSA-6798695 | Neutrophil degranulation | 8 | 0.0029 | 0.95 | LGALS3, ITGAV, CTSB, GSN, HLA-A, PTPRC, AOC1, IST1 |
| HSA-168256 | Immune System | 13 | 0.0287 | 0.54 | LGALS3, CLEC10A, ITGAV, C2, CTSB, FCER1A, GSN, HLA-E, HLA-A, LTB, PTPRC, AOC1, IST1 |
| HSA-2129379 | Molecules associated with elastic fibres | 3 | 0.0326 | 1.64 | ITGAV, LTBP3, FBN2 |
| **Supplemental Table 13E — KEGG pathway enrichment** | | | | | |

| Term ID | Description | Observed gene count | FDR | Strengt h | Proteins in term |
|---|---|---|---|---|---|
| hsa04514 | Cell adhesion molecules | 4 | 0.0474 | 1.19 | ITGAV, HLA-E, HLA-A, PTPRC |
| hsa04612 | Antigen processing and presentation | 3 | 0.0474 | 1.4 | CTSB, HLA-E, HLA-A |

**Supplemental Table 13 (A-E). STRING-DB protein-protein interaction and functional enrichment analysis of MR-significant proteins.** Protein-protein interaction network and functional enrichment analysis for 38 proteins with MR-PRESSO-validated causal associations with optical flow-derived cardiac motion phenotypes (37 mapped to STRING; HCG22 excluded as a non-coding RNA gene). STRING v[12.0] was used with default settings. Only terms with FDR < 0.05 (Benjamini-Hochberg corrected) are shown. (A) Input proteins with gene symbols and STRING identifiers. (B) Protein-protein interaction network summary statistics. (C) GO Cellular Component enrichment. (D) Reactome pathway enrichment. (E) KEGG pathway enrichment. Observed gene count: number of input proteins annotated to each term. FDR: false discovery rate (Benjamini-Hochberg corrected). Strength: $log_{10}$(observed/expected enrichment ratio). Proteins in term: input proteins annotated to each enrichment term.
